# General practitioner workforce sustainability to maximise effective and equitable patient care: a realist review

**DOI:** 10.1101/2025.01.26.25321129

**Authors:** Emily Owen-Boukra, Bryan Burford, Tanya Cohen, Claire Duddy, Harry Dunn, Vacha Fadia, Claire Goodman, Cecily Henry, Elizabeth I Lamb, Margaret Ogden, Tim Rapley, Eliot L Rees, Etienne Royer-Gray, Gillian Vance, Geoff Wong, Sophie Park

## Abstract

**Background:** Global and United Kingdom (UK) primary care face significant General Practitioner (GP) workforce shortages. Worldwide strategies to address this issue include the introduction of additional healthcare professionals and increasing technology utilisation, to reduce GP workload. However, whether these strategies can sustain the GP workforce remains unclear. Our review examines the factors that sustain and enable GPs to flourish.

**Aim:** To examine how general practice work and healthcare systems support GP workforce sustainability and effective and equitable patient care.

**Design & setting:** A realist review of existing empirical and grey literature. The search strategy encompassed six electronic databases.

**Method:** Realist synthesis involved (1) finding existing theories, (2) searching for evidence, (3) selecting articles, (4) extracting data, and (5) synthesising evidence/drawing conclusions. Context-Mechanism-Outcome Configurations were developed using extracted data and patient and public involvement/stakeholder suggestions to refine our programme theory.

**Results:** 168 documents were included. Findings underscore the importance of meaningful work and engagement; relationships across individuals, organisations, and communities; and learning and development. We emphasise the need for congruence between GPs’ core values and their work; cumulative-knowledge building; system agility; psychological safety; and direct human connections.

**Conclusion:** General practice structures, policies, and practices, and the interactions they facilitate, are crucial for the sustainability of the workforce. Collaboration among GPs, the public, and national policymakers is essential for implementing the principles from this review. Future systems should enable personalised care; sustain meaningful work and relationships; facilitate meaning-making; and promote agency, agility, and flexibility to enable GPs to utilise, adapt, and cultivate expertise.

**How this fits in:** Global and UK primary care are experiencing a crisis due to GP workforce shortages. Previous UK GP workforce research has emphasised individual-level factors and solutions, including well-being, self-efficacy, training readiness, resilience, and professional identity shifts. This realist review examines the organisational and system-level factors affecting GPs in their work and proposes recommendations to support a sustainable GP workforce and equitable patient care.

## Introduction

Improving healthcare access, quality, and efficiency in general practice requires multiple factors including the availability, expertise, and distribution of healthcare professionals (HCP) (1). Global and United Kingdom (UK) primary care is experiencing a crisis characterised by substantial General Practitioner (GP) workforce shortages (2–6). UK GPs report low job satisfaction and high workload burden (1). Many GPs have left or are considering leaving their profession (7–9). The GP shortage crisis is critical for the affordability and sustainability of future healthcare systems and patient care; GPs provide safe, high-quality, holistic, and comprehensive care (10, 11). They balance gate-keeping (e.g. limiting patient medicalisation and investigation) and gate-opening (e.g. advocacy) activities, enhancing patient safety, health system efficiency, and health equity (10, 12–14).

Worldwide policies and strategies aimed at addressing GP shortages and workloads encompass additional funding schemes, investments in technology and infrastructure, the introduction of additional HCPs, and multidisciplinary teams (15–17). These strategies have the potential to enhance workforce sustainability (1, 18). However, evidence to guide decisions regarding key causal factors and the effectiveness of specific strategies in diverse contexts remains limited.

This review examines the factors that support GPs to flourish in their work, focusing on the organisational- and system-level characteristics that influence workforce sustainability. Previous UK GP workforce research has conceptualised challenges as individual-level factors or ‘choices’ producing individual-focused perspectives and solutions. These include wellbeing, GP self-efficacy, training readiness, resilience, and professional identity (5, 19, 20). Campbell et al. (21) and Sturmberg et al. (11) emphasised the need to explore GP workforce sustainability as part of a social system and the related social interactions that shape GP work. Our review addresses this gap by identifying and analysing the wider system-level factors that enable (or inhibit) GPs to thrive and continue their work in general practice. In essence, this review explored which broader system factors contribute to or detract from ‘joy’ in a GP’s role, informing recommendations to sustain a GP workforce capable of delivering effective and equitable patient care.

## Method

Realist review is a theory-driven approach to synthesising diverse and complex evidence (22, 23). It formulates causal explanations of what works, for whom, and under what circumstances by identifying interactions between contexts, mechanisms, and outcomes, forming Context-Mechanism-Outcome Configurations (CMOCs). These configurations iteratively shape an emerging programme theory, or an overall explanation of the topic and research question(s). Realist reviews draw on diverse literature to understand complex interventions from multiple perspectives (23), incorporating experiential expertise through patient and public involvement (PPI) and stakeholders, enhancing the research team’s critical engagement with data during analysis. Our review, registered on PROSPERO (CRD42023395583), adheres to the Realist and Meta-narrative Evidence Synthesis: Evolving Standards (RAMESES) quality and reporting standards (24, 25), with methods explained in our published protocol (26).

Initial programme theory (IPT) development involved a literature scoping review in conjunction with individual and small group PPI and stakeholder discussions regarding factors that contribute to ‘joy’ in general practice and facilitate (or create conditions conducive to) effective and equitable patient care (26). These discussions identified numerous challenges while also elucidating important positive factors for further investigation (e.g. meaningful interactions, connections, kindness, and collaboration). Formal searches conducted in April 2023 (Supplementary Information 1) used six electronic databases (MEDLINE, Embase, PsycINFO, CINAHL, HMIC, and Web of Science Core Collection (SCIE, SSCI, AHCI)), encompassing academic and grey literature from health-focused and broader disciplines. These searches initially yielded 1,463 documents. The inclusion and exclusion criteria are summarised in Box 1. Documents were assessed based on their relevance (whether they contained data related to context, mechanisms, and outcomes and contributed to the IPT refinement) and rigour (whether the methods used to generate the relevant data were credible, plausible, and trustworthy) (23, 24).

### Box 1. Inclusion and exclusion criteria

Inclusion:

- Publications from 2013 onward
- UK publication
- GP or UK general practice focus
- Content pertains to relationship between GPs and work, or meaning attributed to work

Exclusion:

- Only trainee participants
- Recruitment only (i.e. nothing about retention)
- No rich or detailed information relevant to IPT

Programme theory refinement involved a synthesis of the literature and PPI/stakeholder consultations, to scrutinise the data sources and formulate explanatory CMOCs. Document characteristics were extracted (HD, VF, and EOB) into a Microsoft word document (see Supplementary Table 1). Relevant data interpreted by the authors (EOB, SP, and GW) were coded and synthesised using a realist logic of analysis. CMOCs were formulated with narrative summaries and direct quotations relating to each configuration (EOB and ERG). The emerging analytical findings and CMOCs were then critically discussed by the research team (SP, EOB, BB, CD, HD, VF, CG, EIL, TR, ER, ERG, GV, and GW), including our PPI co-applicants (TC, MO, and CH), to support CMOC modification, expansion, and refinement (see Supplementary Table 2 for CMOCs and supporting evidence).

## Results

A total of 168 documents published between 2013 and 2023 were included, comprising 106 published research articles, 2 conference abstracts, and 60 other types (e.g. policy reports, guidance articles, editorials, and books) (see Figure 1). Supplementary Table 1 presents the main characteristics of the included documents. Our findings encompass three overarching and interrelated categories: meaningful work and engagement (contributing and mitigating factors); relationships across individuals, organisations, and communities (knowledge accumulation: long-term patient-GP relationships; connection-rich contexts; relationships with allied HCPs), and learning and development (enabling cultures and organisations). Each category is influenced by opportunities for direct interactions and connections within work. The CMOCs and selected excerpts are presented in Table 1, with narrative summaries below.

**Figure 1.**
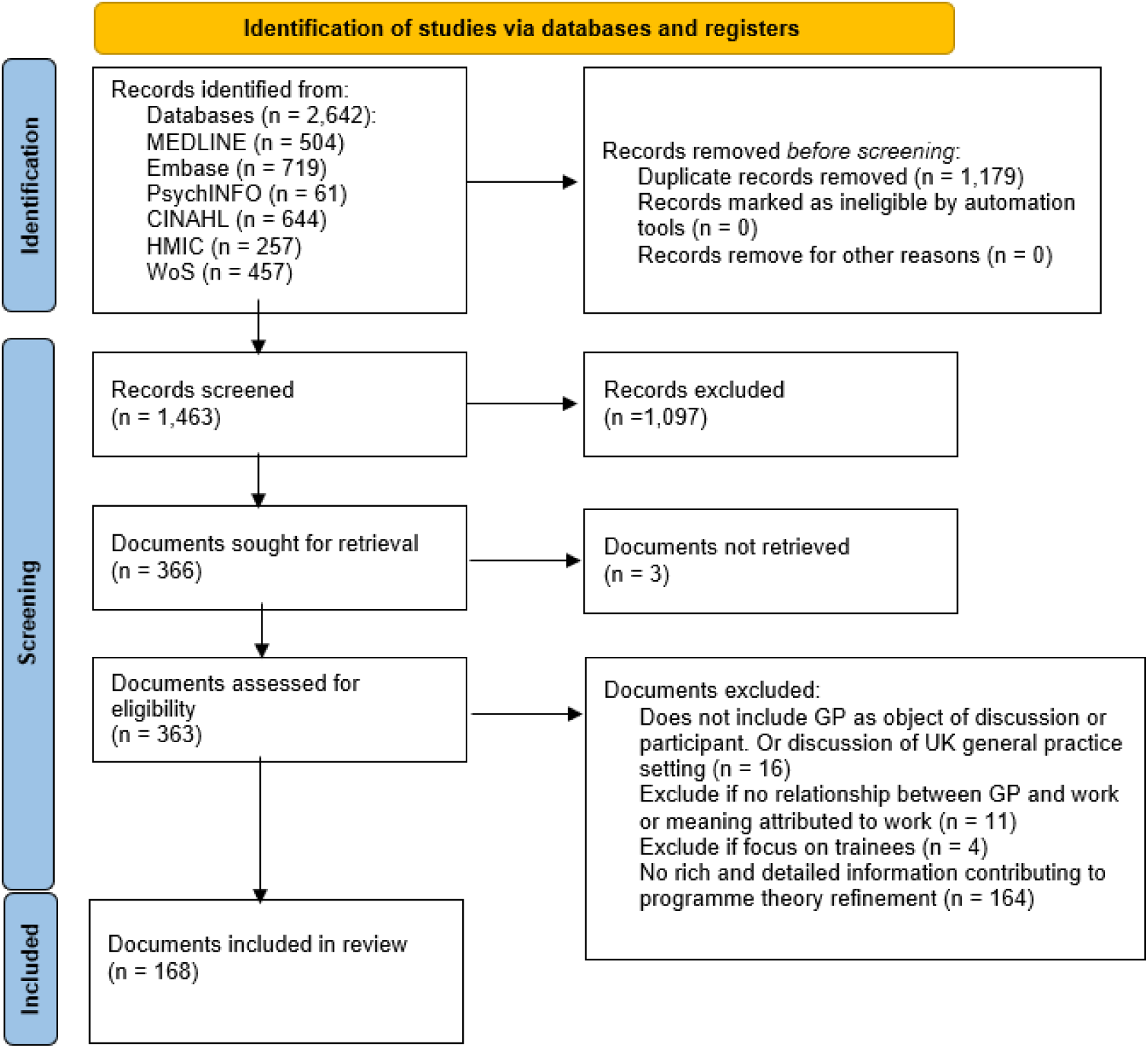
PRISMA summary of searching and selection processes.

**Table 1.**
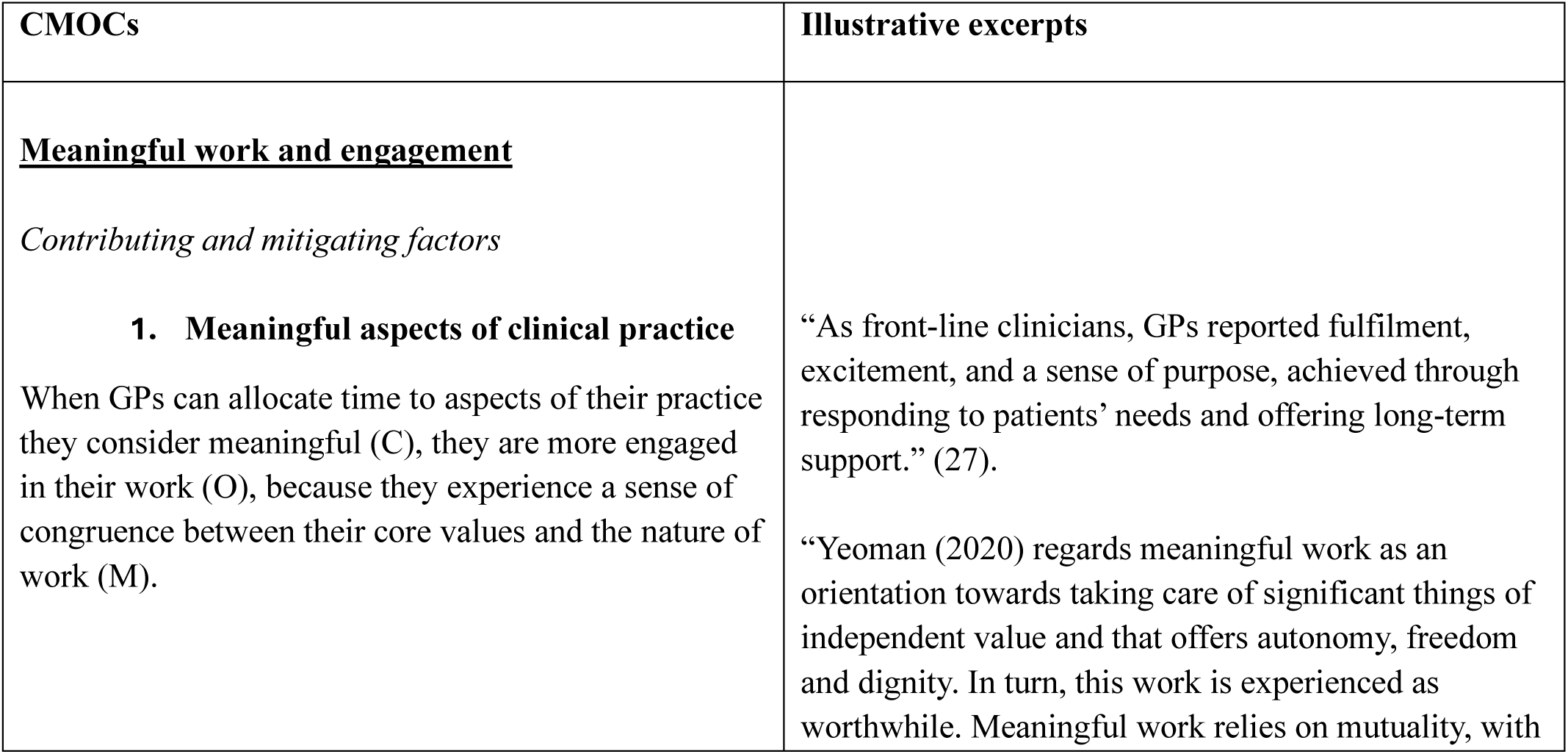
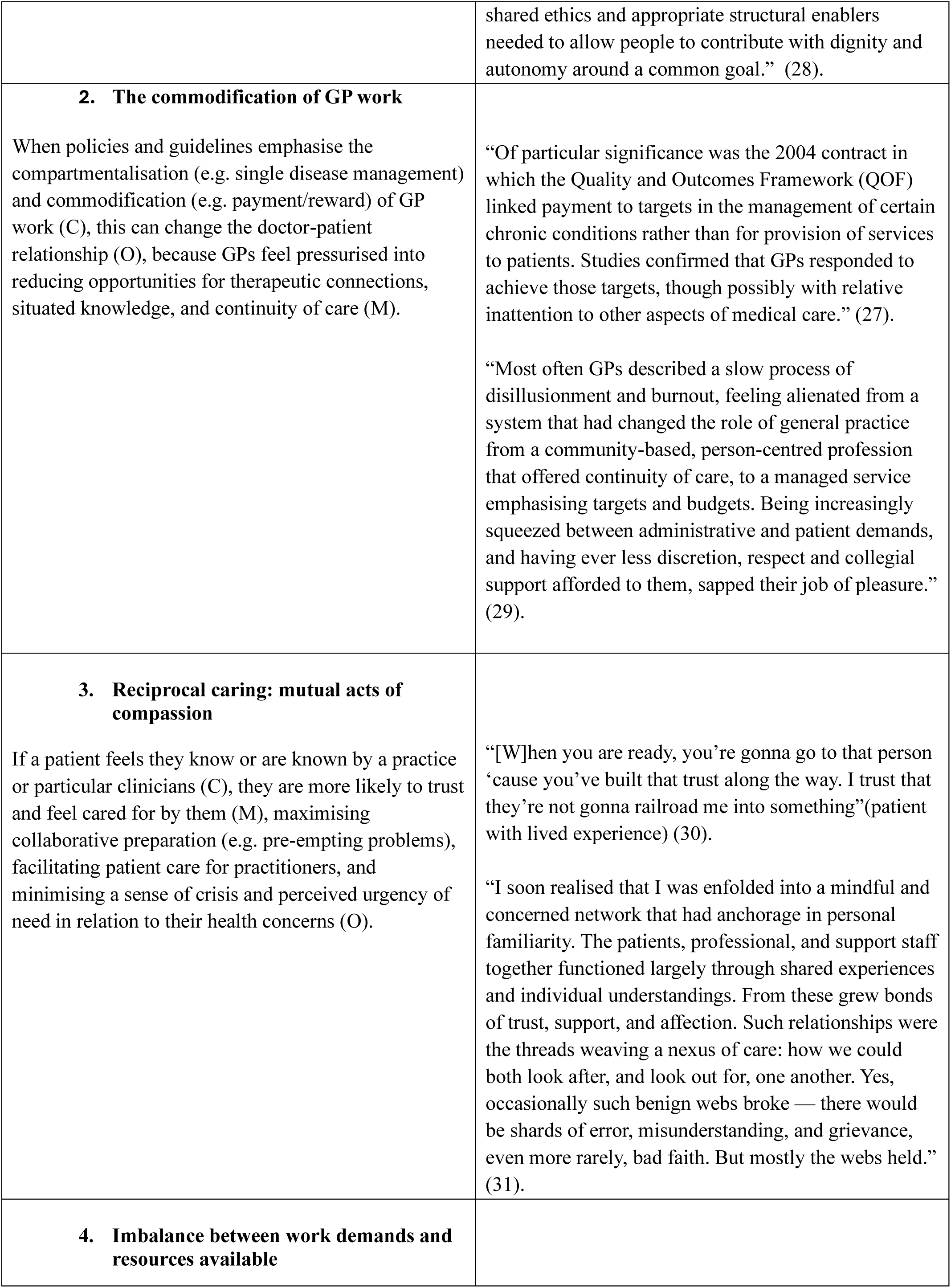
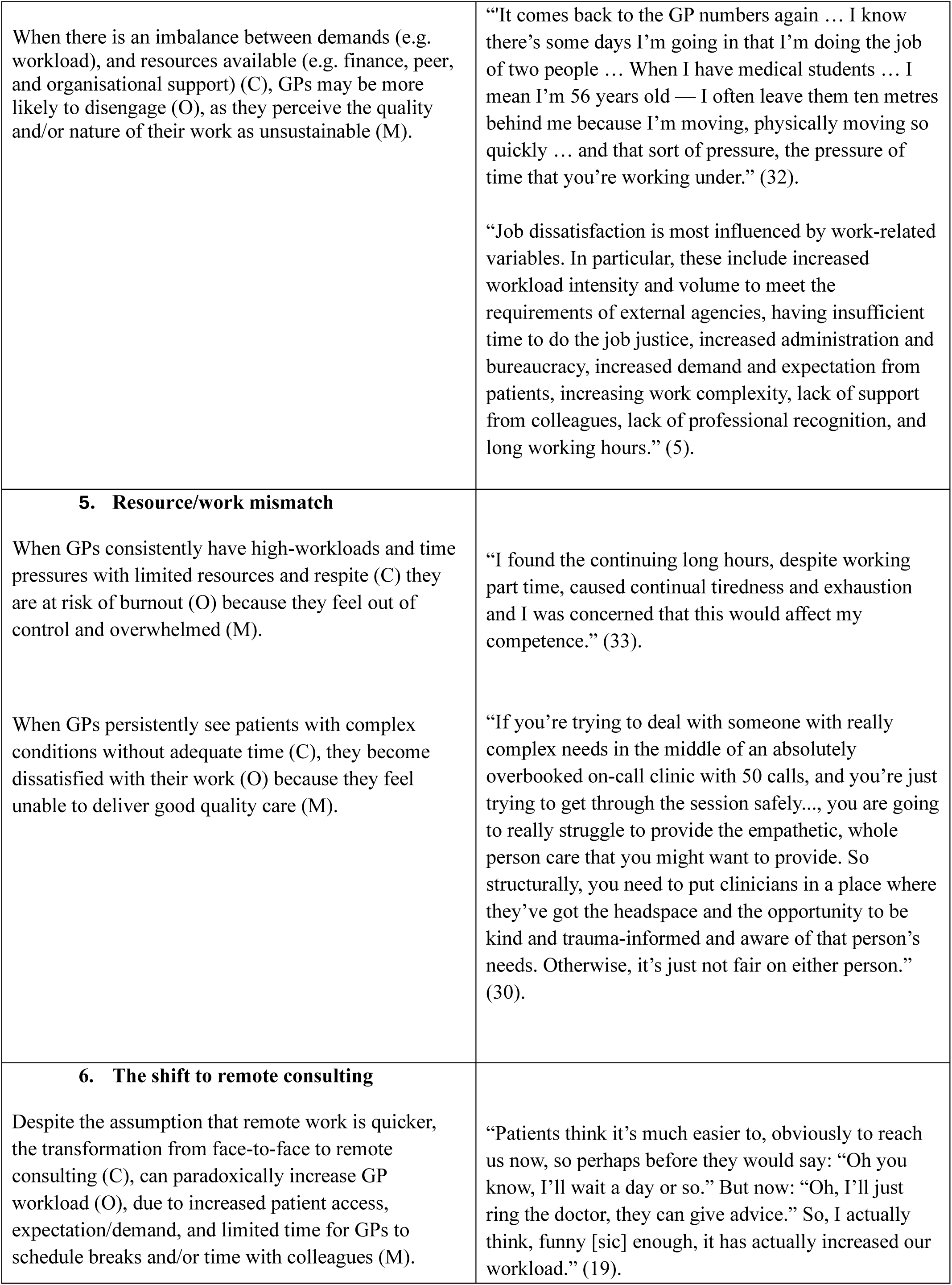
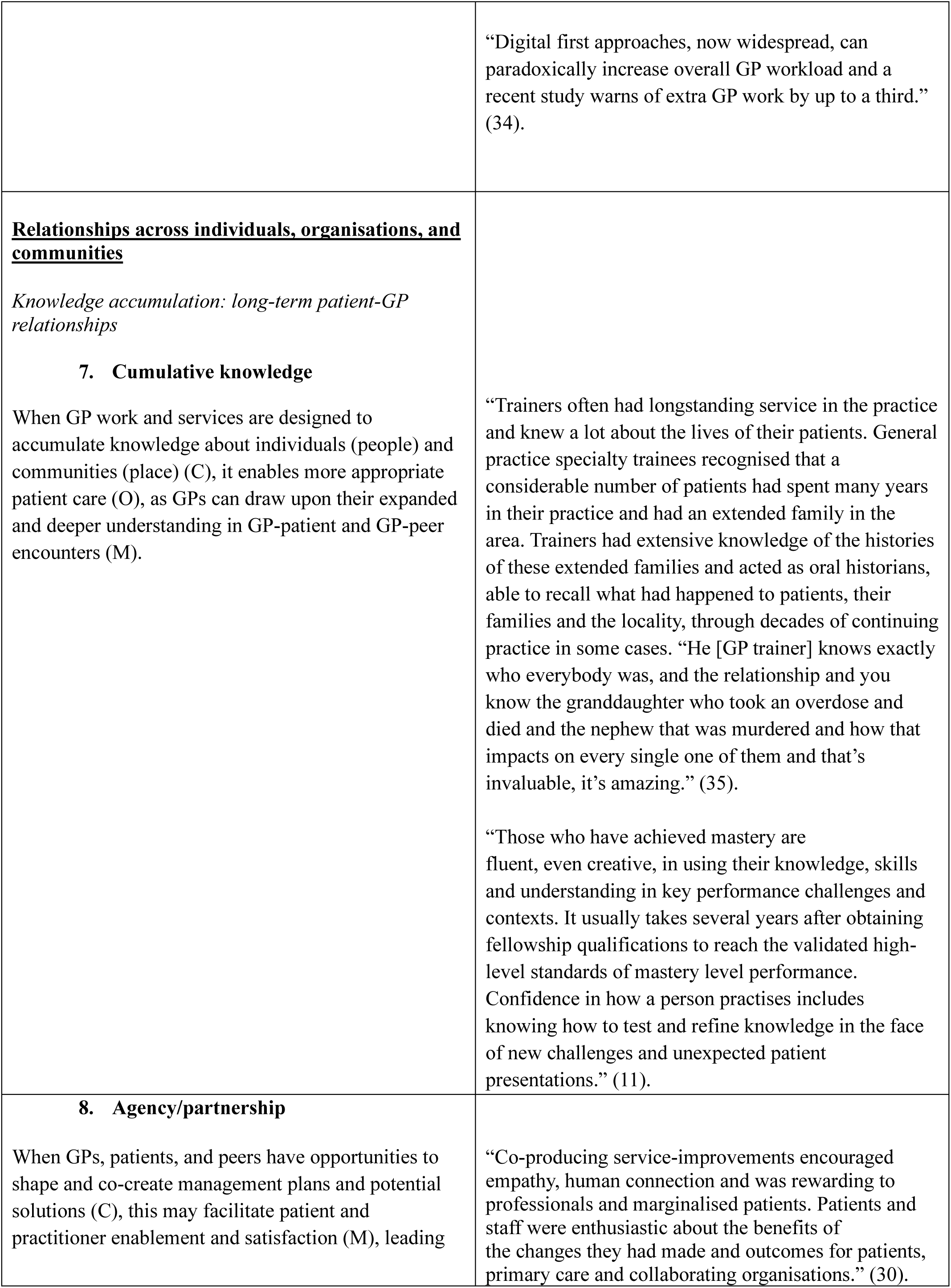
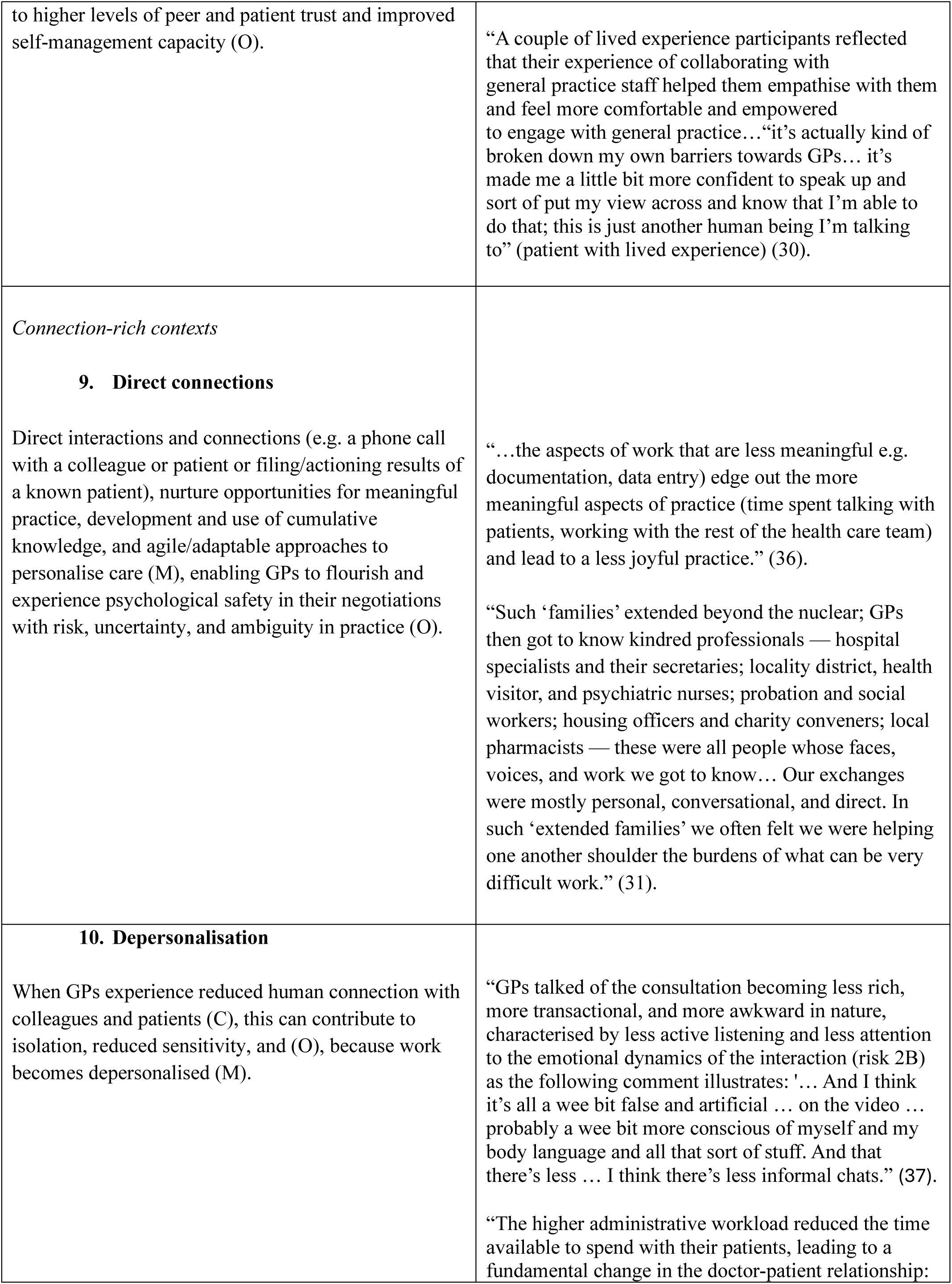
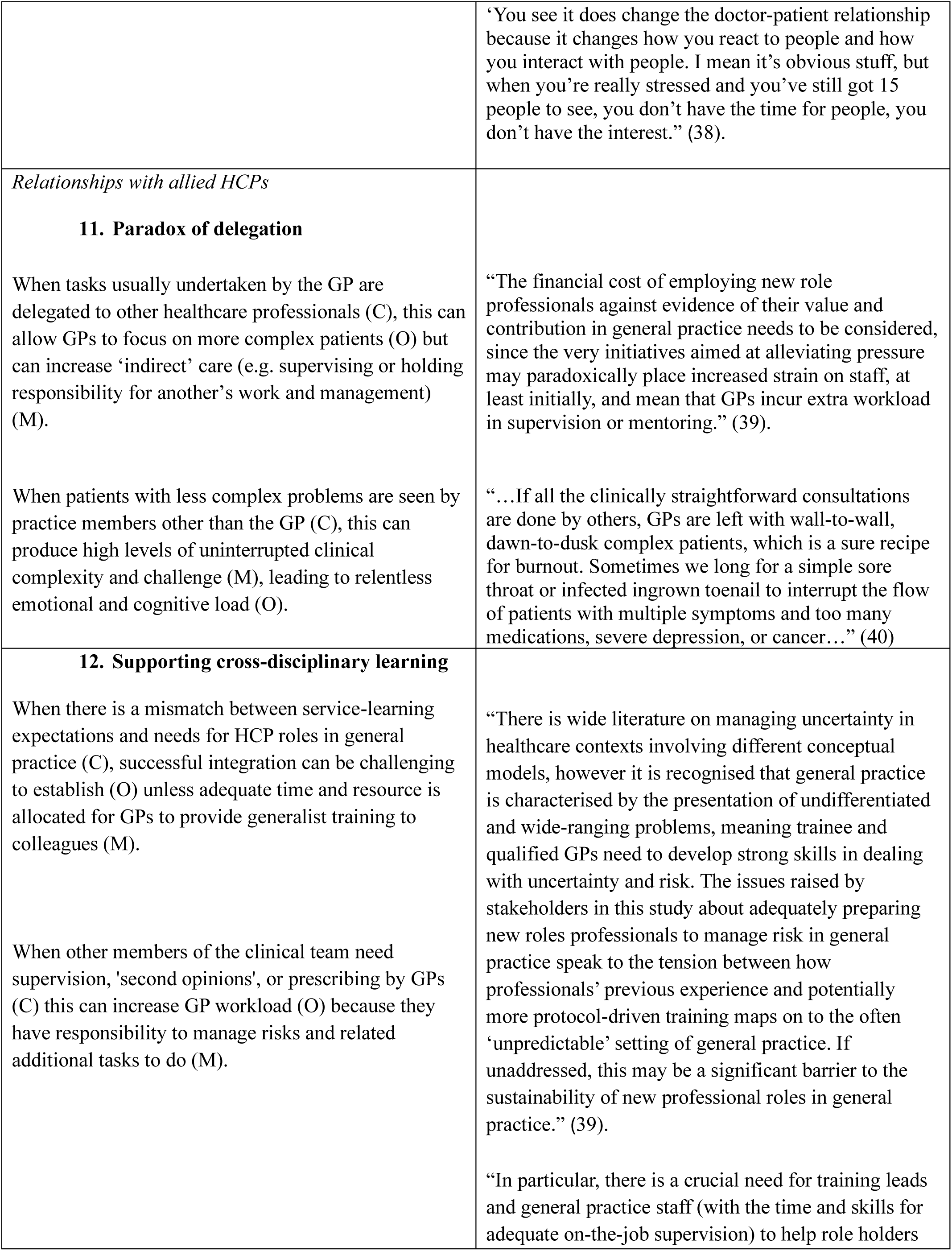
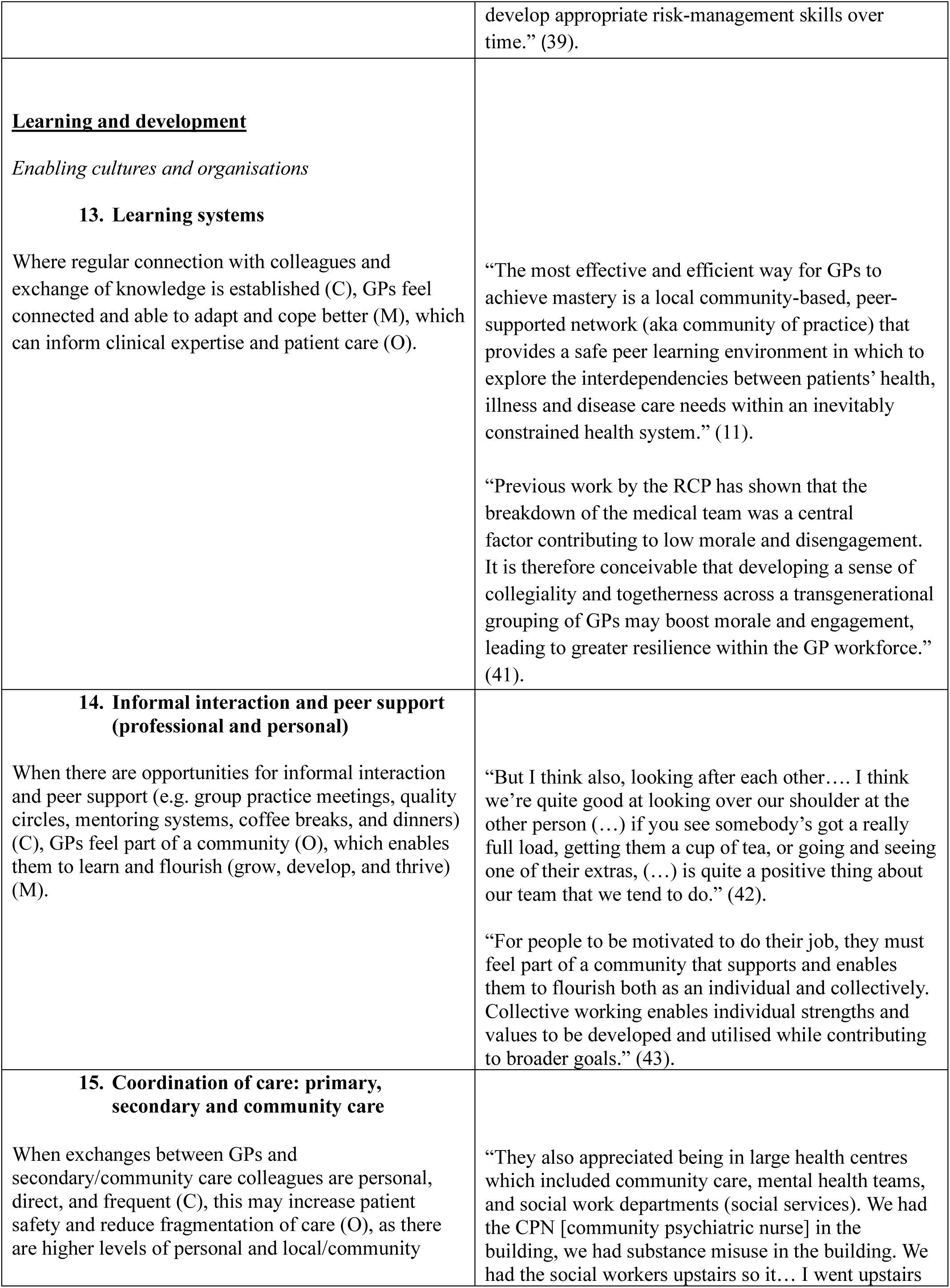
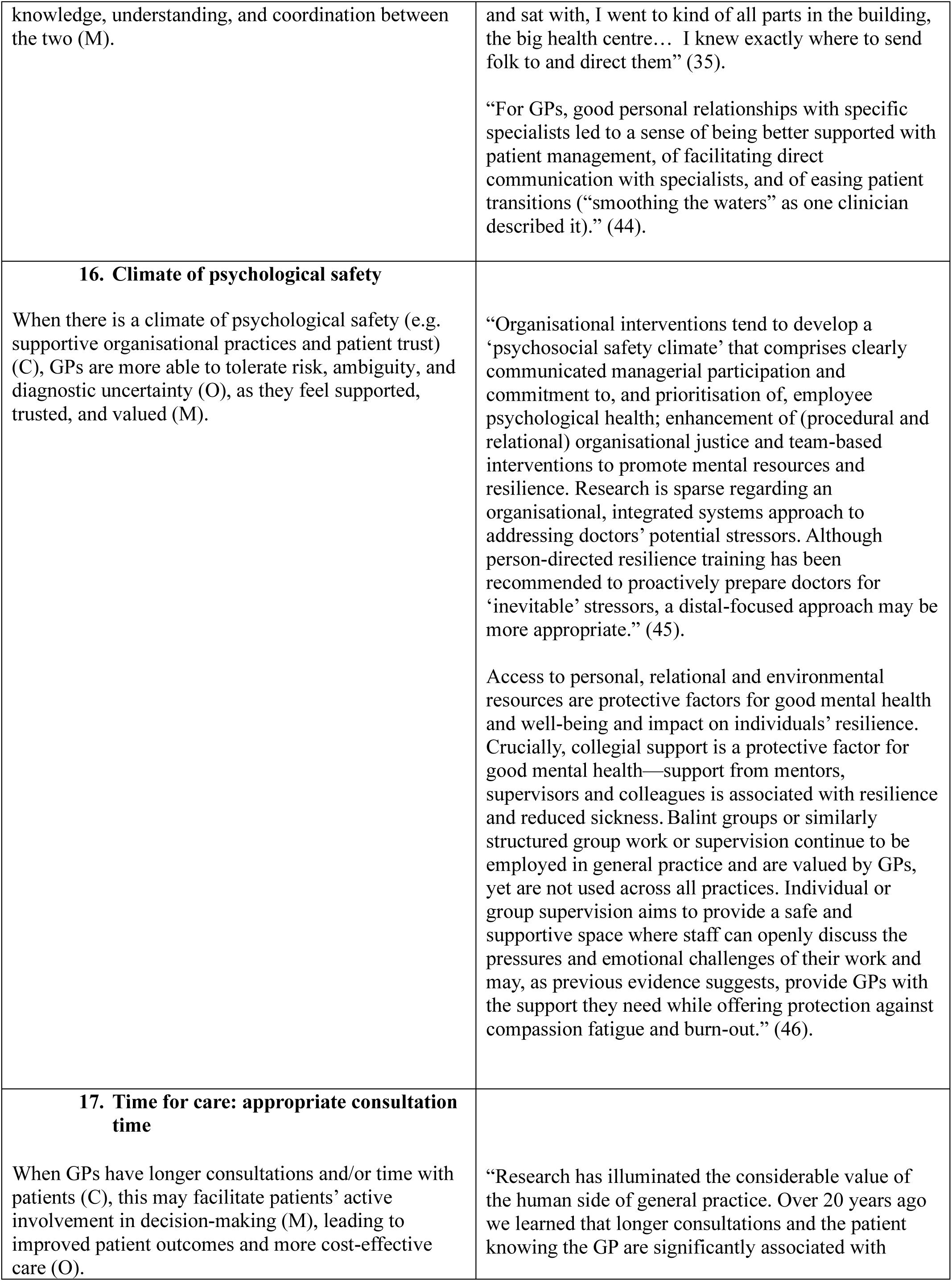
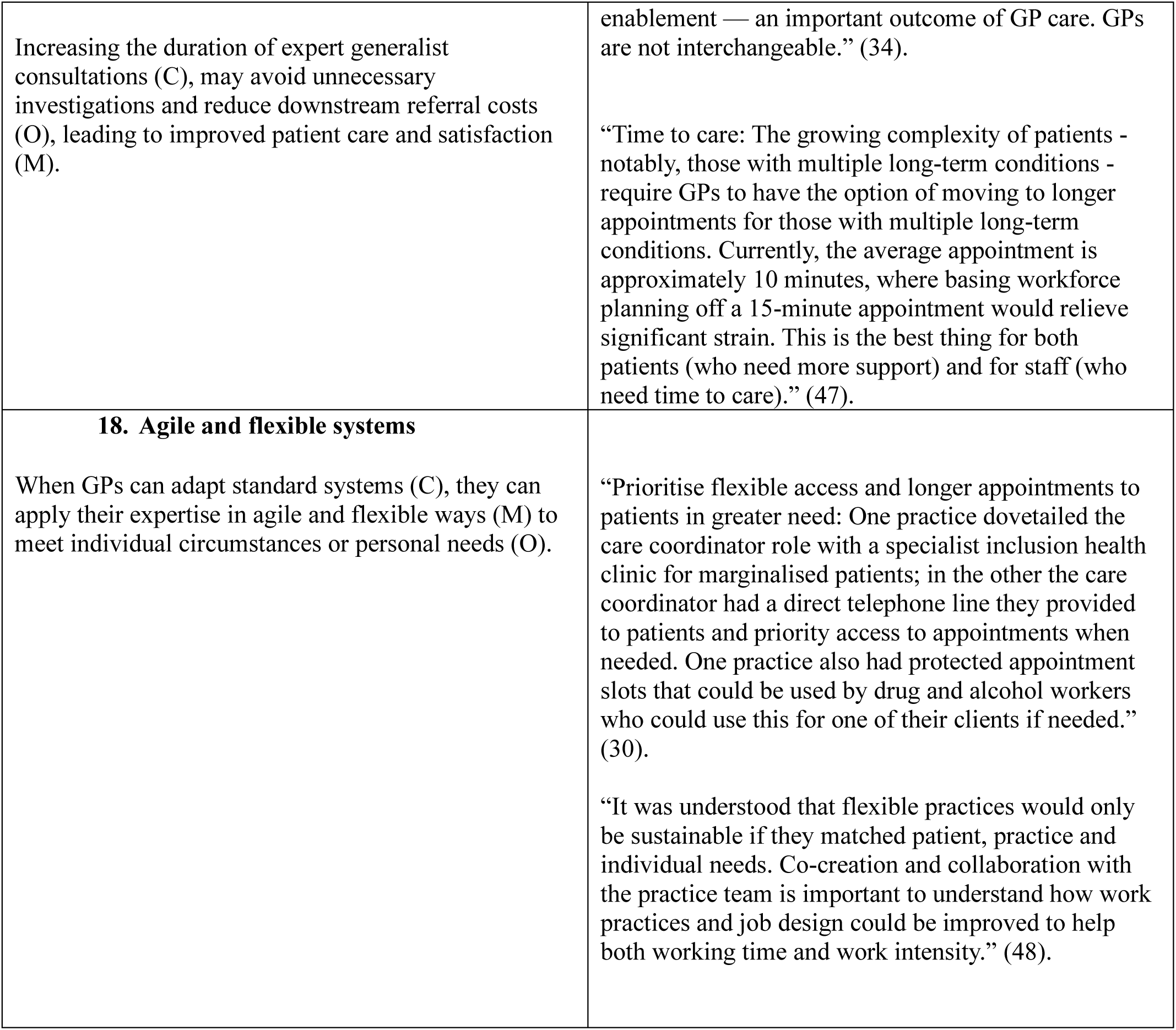
CMOCs and illustrative excerpts.

| CMOCs | Illustrative excerpts |
| --- | --- |
| **<u>Meaningful work and engagement</u>**<br>*Contributing and mitigating factors*<br>**1. Meaningful aspects of clinical practice**<br>When GPs can allocate time to aspects of their practice they consider meaningful (C), they are more engaged in their work (O), because they experience a sense of congruence between their core values and the nature of work (M). | “As front-line clinicians, GPs reported fulfilment, excitement, and a sense of purpose, achieved through responding to patients’ needs and offering long-term support.” (27).<br>“Yeoman (2020) regards meaningful work as an orientation towards taking care of significant things of independent value and that offers autonomy, freedom and dignity. In turn, this work is experienced as worthwhile. Meaningful work relies on mutuality, with |
|  | shared ethics and appropriate structural enablers needed to allow people to contribute with dignity and autonomy around a common goal.” (28). |
| **2. The commodification of GP work**<br>When policies and guidelines emphasise the compartmentalisation (e.g. single disease management) and commodification (e.g. payment/reward) of GP work (C), this can change the doctor-patient relationship (O), because GPs feel pressurised into reducing opportunities for therapeutic connections, situated knowledge, and continuity of care (M). | “Of particular significance was the 2004 contract in which the Quality and Outcomes Framework (QOF) linked payment to targets in the management of certain chronic conditions rather than for provision of services to patients. Studies confirmed that GPs responded to achieve those targets, though possibly with relative inattention to other aspects of medical care.” (27).<br>“Most often GPs described a slow process of disillusionment and burnout, feeling alienated from a system that had changed the role of general practice from a community-based, person-centred profession that offered continuity of care, to a managed service emphasising targets and budgets. Being increasingly squeezed between administrative and patient demands, and having ever less discretion, respect and collegial support afforded to them, sapped their job of pleasure.” (29). |
| **3. Reciprocal caring: mutual acts of compassion**<br>If a patient feels they know or are known by a practice or particular clinicians (C), they are more likely to trust and feel cared for by them (M), maximising collaborative preparation (e.g. pre-empting problems), facilitating patient care for practitioners, and minimising a sense of crisis and perceived urgency of need in relation to their health concerns (O). | “[W]hen you are ready, you’re gonna go to that person ‘cause you’ve built that trust along the way. I trust that they’re not gonna railroad me into something”(patient with lived experience) (30).<br>“I soon realised that I was enfolded into a mindful and concerned network that had anchorage in personal familiarity. The patients, professional, and support staff together functioned largely through shared experiences and individual understandings. From these grew bonds of trust, support, and affection. Such relationships were the threads weaving a nexus of care: how we could both look after, and look out for, one another. Yes, occasionally such benign webs broke — there would be shards of error, misunderstanding, and grievance, even more rarely, bad faith. But mostly the webs held.” (31). |
| **4. Imbalance between work demands and resources available** |  |
| When there is an imbalance between demands (e.g. workload), and resources available (e.g. finance, peer, and organisational support) (C), GPs may be more likely to disengage (O), as they perceive the quality and/or nature of their work as unsustainable (M). | “It comes back to the GP numbers again ... I know there’s some days I’m going in that I’m doing the job of two people ... When I have medical students ... I mean I’m 56 years old — I often leave them ten metres behind me because I’m moving, physically moving so quickly ... and that sort of pressure, the pressure of time that you’re working under.” (32).<br>“Job dissatisfaction is most influenced by work-related variables. In particular, these include increased workload intensity and volume to meet the requirements of external agencies, having insufficient time to do the job justice, increased administration and bureaucracy, increased demand and expectation from patients, increasing work complexity, lack of support from colleagues, lack of professional recognition, and long working hours.” (5). |
| **5. Resource/work mismatch**<br>When GPs consistently have high-workloads and time pressures with limited resources and respite (C) they are at risk of burnout (O) because they feel out of control and overwhelmed (M).<br>When GPs persistently see patients with complex conditions without adequate time (C), they become dissatisfied with their work (O) because they feel unable to deliver good quality care (M). | “I found the continuing long hours, despite working part time, caused continual tiredness and exhaustion and I was concerned that this would affect my competence.” (33).<br>“If you’re trying to deal with someone with really complex needs in the middle of an absolutely overbooked on-call clinic with 50 calls, and you’re just trying to get through the session safely..., you are going to really struggle to provide the empathetic, whole person care that you might want to provide. So structurally, you need to put clinicians in a place where they’ve got the headspace and the opportunity to be kind and trauma-informed and aware of that person’s needs. Otherwise, it’s just not fair on either person.” (30). |
| **6. The shift to remote consulting**<br>Despite the assumption that remote work is quicker, the transformation from face-to-face to remote consulting (C), can paradoxically increase GP workload (O), due to increased patient access, expectation/demand, and limited time for GPs to schedule breaks and/or time with colleagues (M). | “Patients think it’s much easier to, obviously to reach us now, so perhaps before they would say: “Oh you know, I’ll wait a day or so.” But now: “Oh, I’ll just ring the doctor, they can give advice.” So, I actually think, funny [sic] enough, it has actually increased our workload.” (19). |
|  | “Digital first approaches, now widespread, can paradoxically increase overall GP workload and a recent study warns of extra GP work by up to a third.” (34). |
| **<u>Relationships across individuals, organisations, and communities</u>**<br>*Knowledge accumulation: long-term patient-GP relationships*<br>**7. Cumulative knowledge**<br>When GP work and services are designed to accumulate knowledge about individuals (people) and communities (place) (C), it enables more appropriate patient care (O), as GPs can draw upon their expanded and deeper understanding in GP-patient and GP-peer encounters (M). | “Trainers often had longstanding service in the practice and knew a lot about the lives of their patients. General practice specialty trainees recognised that a considerable number of patients had spent many years in their practice and had an extended family in the area. Trainers had extensive knowledge of the histories of these extended families and acted as oral historians, able to recall what had happened to patients, their families and the locality, through decades of continuing practice in some cases. “He [GP trainer] knows exactly who everybody was, and the relationship and you know the granddaughter who took an overdose and died and the nephew that was murdered and how that impacts on every single one of them and that’s invaluable, it’s amazing.” (35).<br>“Those who have achieved mastery are fluent, even creative, in using their knowledge, skills and understanding in key performance challenges and contexts. It usually takes several years after obtaining fellowship qualifications to reach the validated high-level standards of mastery level performance. Confidence in how a person practises includes knowing how to test and refine knowledge in the face of new challenges and unexpected patient presentations.” (11). |
| **8. Agency/partnership**<br>When GPs, patients, and peers have opportunities to shape and co-create management plans and potential solutions (C), this may facilitate patient and practitioner enablement and satisfaction (M), leading | “Co-producing service-improvements encouraged empathy, human connection and was rewarding to professionals and marginalised patients. Patients and staff were enthusiastic about the benefits of the changes they had made and outcomes for patients, primary care and collaborating organisations.” (30). |
| to higher levels of peer and patient trust and improved self-management capacity (O). | “A couple of lived experience participants reflected that their experience of collaborating with general practice staff helped them empathise with them and feel more comfortable and empowered to engage with general practice... “it’s actually kind of broken down my own barriers towards GPs... it’s made me a little bit more confident to speak up and sort of put my view across and know that I’m able to do that; this is just another human being I’m talking to” (patient with lived experience) (30). |
| *Connection-rich contexts*<br>**9. Direct connections**<br>Direct interactions and connections (e.g. a phone call with a colleague or patient or filing/actioning results of a known patient), nurture opportunities for meaningful practice, development and use of cumulative knowledge, and agile/adaptable approaches to personalise care (M), enabling GPs to flourish and experience psychological safety in their negotiations with risk, uncertainty, and ambiguity in practice (O). | “...the aspects of work that are less meaningful e.g. documentation, data entry) edge out the more meaningful aspects of practice (time spent talking with patients, working with the rest of the health care team) and lead to a less joyful practice.” (36).<br>“Such ‘families’ extended beyond the nuclear; GPs then got to know kindred professionals — hospital specialists and their secretaries; locality district, health visitor, and psychiatric nurses; probation and social workers; housing officers and charity conveners; local pharmacists — these were all people whose faces, voices, and work we got to know... Our exchanges were mostly personal, conversational, and direct. In such ‘extended families’ we often felt we were helping one another shoulder the burdens of what can be very difficult work.” (31). |
| **10. Depersonalisation**<br>When GPs experience reduced human connection with colleagues and patients (C), this can contribute to isolation, reduced sensitivity, and (O), because work becomes depersonalised (M). | “GPs talked of the consultation becoming less rich, more transactional, and more awkward in nature, characterised by less active listening and less attention to the emotional dynamics of the interaction (risk 2B) as the following comment illustrates: ‘... And I think it’s all a wee bit false and artificial ... on the video ... probably a wee bit more conscious of myself and my body language and all that sort of stuff. And that there’s less ... I think there’s less informal chats.’ (37).<br>“The higher administrative workload reduced the time available to spend with their patients, leading to a fundamental change in the doctor-patient relationship: |
|  | ‘You see it does change the doctor-patient relationship because it changes how you react to people and how you interact with people. I mean it’s obvious stuff, but when you’re really stressed and you’ve still got 15 people to see, you don’t have the time for people, you don’t have the interest.’ (38). |
| *Relationships with allied HCPs*<br>**11. Paradox of delegation**<br>When tasks usually undertaken by the GP are delegated to other healthcare professionals (C), this can allow GPs to focus on more complex patients (O) but can increase ‘indirect’ care (e.g. supervising or holding responsibility for another’s work and management) (M).<br>When patients with less complex problems are seen by practice members other than the GP (C), this can produce high levels of uninterrupted clinical complexity and challenge (M), leading to relentless emotional and cognitive load (O). | “The financial cost of employing new role professionals against evidence of their value and contribution in general practice needs to be considered, since the very initiatives aimed at alleviating pressure may paradoxically place increased strain on staff, at least initially, and mean that GPs incur extra workload in supervision or mentoring.” (39).<br>“...If all the clinically straightforward consultations are done by others, GPs are left with wall-to-wall, dawn-to-dusk complex patients, which is a sure recipe for burnout. Sometimes we long for a simple sore throat or infected ingrown toenail to interrupt the flow of patients with multiple symptoms and too many medications, severe depression, or cancer...” (40) |
| **12. Supporting cross-disciplinary learning**<br>When there is a mismatch between service-learning expectations and needs for HCP roles in general practice (C), successful integration can be challenging to establish (O) unless adequate time and resource is allocated for GPs to provide generalist training to colleagues (M).<br>When other members of the clinical team need supervision, 'second opinions', or prescribing by GPs (C) this can increase GP workload (O) because they have responsibility to manage risks and related additional tasks to do (M). | “There is wide literature on managing uncertainty in healthcare contexts involving different conceptual models, however it is recognised that general practice is characterised by the presentation of undifferentiated and wide-ranging problems, meaning trainee and qualified GPs need to develop strong skills in dealing with uncertainty and risk. The issues raised by stakeholders in this study about adequately preparing new roles professionals to manage risk in general practice speak to the tension between how professionals’ previous experience and potentially more protocol-driven training maps on to the often ‘unpredictable’ setting of general practice. If unaddressed, this may be a significant barrier to the sustainability of new professional roles in general practice.” (39).<br>“In particular, there is a crucial need for training leads and general practice staff (with the time and skills for adequate on-the-job supervision) to help role holders |
|  | develop appropriate risk-management skills over time.” (39). |
| **<u>Learning and development</u>**<br>*Enabling cultures and organisations*<br>**13. Learning systems**<br>Where regular connection with colleagues and exchange of knowledge is established (C), GPs feel connected and able to adapt and cope better (M), which can inform clinical expertise and patient care (O). | “The most effective and efficient way for GPs to achieve mastery is a local community-based, peer-supported network (aka community of practice) that provides a safe peer learning environment in which to explore the interdependencies between patients’ health, illness and disease care needs within an inevitably constrained health system.” (11).<br>“Previous work by the RCP has shown that the breakdown of the medical team was a central factor contributing to low morale and disengagement. It is therefore conceivable that developing a sense of collegiality and togetherness across a transgenerational grouping of GPs may boost morale and engagement, leading to greater resilience within the GP workforce.” (41). |
| **14. Informal interaction and peer support (professional and personal)**<br>When there are opportunities for informal interaction and peer support (e.g. group practice meetings, quality circles, mentoring systems, coffee breaks, and dinners) (C), GPs feel part of a community (O), which enables them to learn and flourish (grow, develop, and thrive) (M). | “But I think also, looking after each other.... I think we’re quite good at looking over our shoulder at the other person (...) if you see somebody’s got a really full load, getting them a cup of tea, or going and seeing one of their extras, (...) is quite a positive thing about our team that we tend to do.” (42).<br>“For people to be motivated to do their job, they must feel part of a community that supports and enables them to flourish both as an individual and collectively. Collective working enables individual strengths and values to be developed and utilised while contributing to broader goals.” (43). |
| **15. Coordination of care: primary, secondary and community care**<br>When exchanges between GPs and secondary/community care colleagues are personal, direct, and frequent (C), this may increase patient safety and reduce fragmentation of care (O), as there are higher levels of personal and local/community | “They also appreciated being in large health centres which included community care, mental health teams, and social work departments (social services). We had the CPN [community psychiatric nurse] in the building, we had substance misuse in the building. We had the social workers upstairs so it... I went upstairs |
| knowledge, understanding, and coordination between the two (M). | and sat with, I went to kind of all parts in the building, the big health centre... I knew exactly where to send folk to and direct them” (35).<br>“For GPs, good personal relationships with specific specialists led to a sense of being better supported with patient management, of facilitating direct communication with specialists, and of easing patient transitions (“smoothing the waters” as one clinician described it).” (44). |
| **16. Climate of psychological safety**<br>When there is a climate of psychological safety (e.g. supportive organisational practices and patient trust) (C), GPs are more able to tolerate risk, ambiguity, and diagnostic uncertainty (O), as they feel supported, trusted, and valued (M). | “Organisational interventions tend to develop a ‘psychosocial safety climate’ that comprises clearly communicated managerial participation and commitment to, and prioritisation of, employee psychological health; enhancement of (procedural and relational) organisational justice and team-based interventions to promote mental resources and resilience. Research is sparse regarding an organisational, integrated systems approach to addressing doctors’ potential stressors. Although person-directed resilience training has been recommended to proactively prepare doctors for ‘inevitable’ stressors, a distal-focused approach may be more appropriate.” (45).<br>Access to personal, relational and environmental resources are protective factors for good mental health and well-being and impact on individuals’ resilience. Crucially, collegial support is a protective factor for good mental health—support from mentors, supervisors and colleagues is associated with resilience and reduced sickness. Balint groups or similarly structured group work or supervision continue to be employed in general practice and are valued by GPs, yet are not used across all practices. Individual or group supervision aims to provide a safe and supportive space where staff can openly discuss the pressures and emotional challenges of their work and may, as previous evidence suggests, provide GPs with the support they need while offering protection against compassion fatigue and burn-out.” (46). |
| **17. Time for care: appropriate consultation time**<br>When GPs have longer consultations and/or time with patients (C), this may facilitate patients’ active involvement in decision-making (M), leading to improved patient outcomes and more cost-effective care (O). | “Research has illuminated the considerable value of the human side of general practice. Over 20 years ago we learned that longer consultations and the patient knowing the GP are significantly associated with |
| Increasing the duration of expert generalist consultations (C), may avoid unnecessary investigations and reduce downstream referral costs (O), leading to improved patient care and satisfaction (M). | enablement — an important outcome of GP care. GPs are not interchangeable.” (34).<br>“Time to care: The growing complexity of patients - notably, those with multiple long-term conditions - require GPs to have the option of moving to longer appointments for those with multiple long-term conditions. Currently, the average appointment is approximately 10 minutes, where basing workforce planning off a 15-minute appointment would relieve significant strain. This is the best thing for both patients (who need more support) and for staff (who need time to care).” (47). |
| **18. Agile and flexible systems**<br>When GPs can adapt standard systems (C), they can apply their expertise in agile and flexible ways (M) to meet individual circumstances or personal needs (O). | “Prioritise flexible access and longer appointments to patients in greater need: One practice dovetailed the care coordinator role with a specialist inclusion health clinic for marginalised patients; in the other the care coordinator had a direct telephone line they provided to patients and priority access to appointments when needed. One practice also had protected appointment slots that could be used by drug and alcohol workers who could use this for one of their clients if needed.” (30).<br>“It was understood that flexible practices would only be sustainable if they matched patient, practice and individual needs. Co-creation and collaboration with the practice team is important to understand how work practices and job design could be improved to help both working time and work intensity.” (48). |

### Meaningful work and engagement

#### Contributing and mitigating factors

For many GPs, it is essential that their work is purposeful, significant, and aligns with their core values (11, 30, 49–51). When GPs are able to allocate time to aspects of their practice they consider meaningful (e.g. direct doctor-patient interactions, or administrative tasks pertaining to known patients), they are more engaged in their work, because they experience a sense of congruence or alignment between their core values and the nature of work (CMOC1) (5, 27, 43, 49–55). The experiences and perceptions of meaningful work varied among GPs. GPs derive unity, purpose, and meaning through long-term therapeutic relationships with patients and families, in addition to performing patient advocacy, health promotion work, community participation, and improving local service provision (5, 11, 27, 30, 35, 46, 49, 54, 56–62).

GPs find intellectual stimulation in managing ill-defined illnesses, chronic complex multimorbidity, and accurate diagnoses (5, 27, 38, 49, 56, 62–64). GPs indicated the highest level of satisfaction and meaning when experiencing feelings of competency and mastery (5, 11, 56), and from consultations in which they perceived their personal contributions to have resulted in successful patient outcomes (27, 49). The act of mentoring and teaching medical students and junior colleagues through involvement in undergraduate and postgraduate training schemes is also particularly meaningful (43, 49, 52, 62, 65). Many GPs value the opportunity to contribute to the professional development of others and to reciprocate the support they received during their own training (27).

Various challenges prevent GPs from engaging in meaningful work. To address these challenges, work must be proportionate and structured to enable GPs to maintain meaning. In contexts where policies and guidelines emphasise GP work as compartmentalised (e.g. single disease management) and/or commodified (e.g. payment/reward), this can distort the doctor-patient relationship, as GPs experience pressure to reduce opportunities for therapeutic connections, situated knowledge, and continuity of care (CMOC2) (27, 29, 66). This can compromise GPs’ ability to address diverse patient needs, disproportionately affecting the most vulnerable and socioeconomically disadvantaged patients (19, 27, 50, 54, 57, 65). Many GPs find aspects of their work intellectually stimulating, fulfilling, and meaningful. However, most report pressure to balance patient/carer needs against target-driven accountability, stringent bureaucratic monitoring, and a standards-driven reward system (5, 27–29, 38, 50, 65, 67, 68). While larger practice sizes do not inhibit meaningful work, certain common at-scale organisational approaches (e.g. task delegation) can disintegrate and/or compartmentalise work (e.g. repeat prescribing, filing results, and acute provision of clinical care), minimising personal continuity and reducing opportunities for meaningful connections (36).

Media and policy attention has recently oriented general practice systems towards prioritising rapid access, superseding many key elements for sustainable, meaningful, and equitable general practice (19, 69). A critical factor that our PPI co-applicants identified and supported in our literature analysis was the importance of mutual and reciprocal care (CMOC3): “What about patient care for their GP?”. Current policies tend to position ‘patient demand’ as a fixed entity (and often expanding/overwhelming). Our analysis demonstrated that demand can be viewed as dynamic: constructed and negotiated through social interactions between patients and their GPs and practices. When systems become depersonalised (e.g. prioritising the speed of triage/access), the individual relationships between clinicians and patients can diminish (19, 50). Patients may be (re)positioned as consumers and experience reduced personal connections with their GP or practice. This can limit opportunities for GP ‘holding work’ (also identified in primary care link worker roles (70)) and follow-up appointments to help patients manage anxieties, concerns, and healthcare navigation. Without a sense of connection, patients may feel overwhelmed and perceive a need to seek more frequent or urgent attention.

When familiarity with a practitioner exists, patients are more likely to establish trust and feel cared for by the GP. This familiarity facilitates opportunities for collaborative preparation or pre-emptive planning regarding ‘what to look out for’ or ‘when to worry’, as well as a mutual understanding of temporal expectations (e.g. previous patterns for these symptoms and/or this patient, plus likely duration self-limiting, short- or long-term). Patients can reciprocate care and empathy for their practitioners and can (when safe) moderate their help-seeking behaviours where they feel they have adequate information and trust their practitioner, minimising a sense of crisis and perceived urgency of healthcare needs (19, 30, 60, 62).

Impersonal and consumerist systems benefit patients skilled in commanding attention, navigating complex systems, and possessing high health literacy (71). Technological advancements may disadvantage certain patient populations, increasing their distance from general practice support and exacerbating inequality gaps (19). Contemporary policies that shape healthcare access and services often assume that patients present with pre-formed and predetermined problems (72). This overlooks ‘problem-setting’, an essential step in GP-patient interactions for collaborative identification and prioritisation of issues. Active patient participation in problem-setting promotes collaboration with GPs, fostering sustainable care approaches, and reducing feelings of hopelessness, apprehension, and disempowerment (35).

Workload (demand, nature, and quantity) and resource imbalances (e.g. finance, peer, and organisational support), can impede meaningful work (5, 19, 28, 35, 43, 46, 50, 53, 65, 73–77) (CMOC4). When GPs experience consistently high workloads with limited resources and respite, they are at risk of burnout, as they feel out of control and overwhelmed (5, 16, 19, 30, 35, 50, 73–75) (CMOC5). Despite the assumption that remote work enhances efficiency, the transformation from face-to-face to remote consulting can paradoxically increase GP workload due to increased patient access, expectation/demands, and limited opportunities for GPs to schedule breaks and/or time with colleagues (19, 34) (CMOC6).

Numerous studies identify administrative work as problematic (19, 28, 29, 38, 43, 45, 50, 75). It is frequently categorised as ’hidden’ or ‘invisible’, reflecting a lack of allocated, explicit, or scheduled time (in contrast to a hospital consultant who might have a combination of clinical and administrative sessions) (65). Administrative work was more acceptable when it could be attributed meaning, such as when it was requested by the GP, or involved reviewing the results and considering management options for a known patient. When work organisation resulted in GPs seeing fewer patients (e.g. delegated to HCPs) to perform more administrative tasks and/or these tasks became disconnected from known patients, administrative work was perceived as more challenging, risky, and burdensome (36).

However, in contexts where direct human connections and face-to-face interactions with patients and colleagues are present, paperwork/administrative work can be perceived as a means to a larger end and become an accepted part of GP’s work (43, 62).

### Relationships across individuals, organisations, and communities

#### Knowledge accumulation: long-term patient-GP relationships

While some assume that learning is acontextual, irrespective of educational, social, and cultural circumstances, this review demonstrates the centrality of situated learning and expertise (78). By nurturing situated learning in general practice (emphasising the importance of social interactions with patients and peers, and conceptualising learning and knowledge accumulation as a dynamic process), GPs describe more meaningful work (11, 30, 49, 50, 57–59). For instance, when GP work and services are designed to accumulate knowledge about individuals (people) and communities (place), it facilitates more appropriate patient care, as GPs can draw upon and adapt their expanded and deeper understanding in GP-patient and GP-peer encounters (11, 16, 27, 30, 35, 43, 49, 57, 79) (CMOC7). GPs demonstrated a deep contextual understanding of patients’ personal circumstances informing and shaping medical care (35). Familiarity with the local area and wide knowledge of community needs could enhance how and when GPs implement knowledge to inform patient care and enable community advocacy (11, 35, 49, 60).

The growth and implementation of cumulative knowledge can flourish through interactions with patients and peers (both within and beyond the institution), cultivating knowledge of local systems and community preferences (27, 28, 49, 57, 69, 79, 80). This facilitates opportunities for patient partnership and advocacy (‘gate-opening’), as well as the development and use of adaptive expertise to address the needs of specific circumstances (30, 35, 69, 81). In contexts where GPs, patients, and peers have opportunities to shape and co-create management plans and potential solutions, this may promote patient and practitioner enablement and satisfaction, resulting in higher levels of peer and patient trust, and improved self-management capacity (CMOC8) (27, 30, 35, 57). From a patient’s perspective, mutual trust and respect was deemed essential (30).

#### Connection-rich contexts

Direct interactions and connections (e.g. a phone call with a colleague/patient or filing and actioning the results of a known patient) facilitate opportunities for meaningful practice, development and utilisation of cumulative knowledge, and agile/adaptable approaches to personalise care (35, 57). This enables GPs to flourish and experience psychological safety in their negotiations with risk, uncertainty, and ambiguity in practice (CMOC9). Excessive shifts towards indirect interactions (e.g. managing substantial risk indirectly through supervision of allied HCP patient interactions, or reviewing results for unknown patients and making disembodied decisions without patient interaction or knowledge of why a test was conducted), increase work-related risk, leading to diminished GP experiences of connection and engagement (19, 36). GPs support many patients with undifferentiated illnesses, complex needs, and multiple conditions. Protocol-driven or standardised care is, in these circumstances, often less safe or desirable for patients, and generates inappropriate and additional (expensive) healthcare service demands (14). Managing these compromises and adaptations with patients, involves complex and dynamic negotiation of risk. GPs mitigate this risk not by standardising, but by rapid exchange, adaptation, and development of management plans with patients. Indirect (rather than direct) interactions can make the associated risks and navigation of uncertainty overwhelming and unmanageable, impacting patient care and GP wellbeing (19). In contexts where GPs experience reduced human connections with colleagues and patients, this can contribute to isolation, reduced sensitivity, and motivation because work becomes depersonalised (28, 37, 38, 46, 50, 53, 54, 57, 68) (CMOC10).

#### Relationships with allied HCPs

Effective collaborative relationships with allied HCPs may contribute to meaningful work (67, 82, 83). However, the reviewed literature identifies a potential paradox in delegating work to allied HCPs. Managing risk and uncertainty can be *more* complex when interactions between patients and colleagues are indirect (39, 84, 85). This has significant implications for the support, time, and nature of supervision that GPs need to provide for allied HCPs working with GPs to ensure effective and equitable patient care (86). When tasks typically undertaken by the GP are delegated to allied HCPs, this can allow GPs to focus on more complex patients but can increase ‘indirect’ care (e.g. supervising or holding responsibility for another’s work and management), and may erode continuity of care (39, 74, 86) (CMOC11.1). Triaging ‘simpler’ or ‘appropriate’ patients for staff can be challenging in the context of undifferentiated illnesses. However, when patients with potentially less complex problems are seen by practice members other than the GP, this can produce high levels of uninterrupted clinical complexity and challenges, leading to relentless emotional and cognitive load (CMOC11.2) (40). In contexts where there is a discrepancy between service-learning expectations and needs for HCP roles in general practice, successful integration can be challenging unless sufficient time and resources are allocated for GPs to provide generalist training to colleagues (CMOC12).

### Learning and development

#### Enabling cultures and organisations

Regular interactions with colleagues and established systems and routines for knowledge exchange can facilitate GPs’ sense of connection, adaptability, and coping mechanisms, thereby informing clinical expertise and patient care (CMOC13) (31, 41, 43, 46, 49, 52, 58, 74, 80). Such interactions include opportunities for informal engagement and peer support (e.g. group practice meetings, quality circles, team ‘huddles’, mentoring systems, coffee breaks, and dinners) that may enable GPs to experience a sense of community, which, in turn, allows them to learn and flourish (grow, develop, and thrive) (CMOC14) (46, 53, 77, 87). Additional examples include personal, direct, and/or frequent exchanges between GPs and secondary/community care colleagues; increasing patient safety; and reducing fragmentation of care, as there are higher levels of personal and local/community knowledge, understanding, and coordination between the two (CMOC15) (30, 35, 44, 49, 53, 88, 89).

A climate of psychological safety is essential to facilitate learning and regular interactions with colleagues (28, 43, 45, 55, 73). In contexts where psychological safety exists (e.g. supportive organisational practices and patient trust), GPs can use their expertise to support patient care in the presence of uncertainty (CMOC16). Trust is a crucial element in psychological safety. This is particularly important when GP work involves frequent negotiation of risk, ambiguity, and diagnostic uncertainty (11, 39, 84). While guidelines are beneficial in certain circumstances, rigid expectations that all clinical work should be possible to procotolise causes many GPs to feel disempowered due to a lack of autonomy and authenticity to adapt to patient needs and, consequently, to disengage from their work (11, 28, 43, 52). One unintended consequence of protocol over-reliance was an increasing discrepancy between ‘imagined work’ as a series of single disease management pathways, and patient interactions requiring navigation of complex and interrelated problems that were either at the boundaries of biomedical pathways, multiple in nature, or interconnected with additional complex issues such as social or ecological factors (e.g. housing, fleeing domestic violence).

## Discussion

### Summary

Numerous synergies exist between achieving effective and equitable patient care, and ensuring a sustainable future GP workforce. Social interactions in general practice, such as those between patients and GPs, significantly influence healthcare delivery, shape the proportionate support necessary for universal care, and affect GP’s job satisfaction and workforce sustainability (37, 55, 90). GP work and patient engagement are dynamic and socially negotiated. Chronic underinvestment in UK general practice has resulted in overwhelming workloads and understaffing, negatively affecting workforce morale (54, 66, 67). Consequently, increased investment in the GP workforce is crucial. Contrary to prevailing narratives, it is not inevitable for modern general practice to entail impersonal GP-patient interactions, overwhelming workloads, and excessive documentation at the expense of patient contact. Our findings elucidate the nature of ‘GP workforce problems’ requiring urgent attention, instead of the current emphasis on access to any practitioner, additional triage, indirect supervision, and risk-escalating workload. These short-term measures neglect sustaining GP-patient connections, which help manage ambiguity and risk effectively and equitably (34, 37, 54). Although there is no universal approach, this review outlines the key *principles* that maximise opportunities for joy in GP work and the provision of effective and equitable patient care.

Organisational and financial models are critical for GP workforce sustainability. Financial incentives influence the nature of work and GP engagement in general practice (27, 29, 66, 91). While meaning-making is not solely related to financial factors, it significantly impacts team composition, patient needs addressed, appointment systems, and shapes the opportunities and challenges experienced within and outside work (28). Previous research (92–94) has emphasised the importance of prioritising collective outcome goals over individual financial incentives to promote collaboration and service integration. This review demonstrates how commodification affects GP-patient interactions, often prioritising short-term financial objectives over meaningful discussions or spontaneous patient-clinician engagement (27, 29, 30, 68).

Based on the findings of our review and extensive stakeholder and patient engagement, we propose the following priorities for the organisation and delivery of general practice:

- Support GPs in tolerating and negotiating risk through patient and peer interactions
- Enable flexibility and agility to implement personalised care
- Empower GPs to work with patients to align person-centred values with work requirements and activities

These objectives can be achieved by addressing three key areas:

1. Meaningful work and engagement
2. Relationships across individuals, organisations, and communities
3. Learning and development

Our refined programme theory summaries our findings, as shown in Table 2.

**Table 2:** Refined programme theory.

| Facilitators |  |
| --- | --- |
| Meaningful work and engagement | a. Congruence between GPs' core values and the nature of their work<br>b. Opportunities for reciprocal care and mutual acts of compassion |
|  | c. Balance work demands and available resources (e.g. appropriate consultation time)<br>d. Support GP roles as advocates and enable GP/patient agency<br>e. Recognise and enable intellectual stimulation in GP work (e.g. agile and flexible expertise to personalise/contextualise care) |
| **Relationships across individuals, organisations, and communities** | a. Connection-rich contexts (direct interactions and connections within work activities)<br>b. Cultivate and use cumulative knowledge (regarding local people and place) to inform the organisation and delivery of care<br>c. Facilitate direct connections and cross-disciplinary learning opportunities between peers and organisations<br>d. Enable informal learning, engagement, and peer support |
| **Learning and development** | a. Climate of psychological safety that enables opportunities for care and on-going practice-based learning with patients and peers<br>b. Embed spaces for learning and exchanging cumulative knowledge into learning structures and systems<br>c. Promote enabling cultures and dynamic learning systems to facilitate the negotiation of risk, ambiguity, and uncertainty |
| **Barriers** |  |
| a. De-personalisation and commodification of GP work<br>b. Lack of recognition/planning to support potential disconnection and additional work inherent in remote consulting<br>c. Paradox of delegation to supervise and support allied healthcare professionals |  |

### Strengths and limitations

This realist review represents the first exploration of GP workforce sustainability, enhancing our understanding of the factors that maintain and sustain GPs, addressing challenges, and offering recommendations for future primary care. Our research extends previous individual-orientated perspectives to a systemic view of organisational characteristics and the role of the social environment. Included documents encompass a diverse range of materials, including grey literature, conference materials, and policy reports, maximising the breadth and depth of the review. To enhance transferability, we examined documents on GP workforce sustainability across various geographic settings. Through collaborative discussions and reflective dialogue with PPI and stakeholder groups, we verified our approach, expanded analytical possibilities, and ensured recommendations were relevant and applicable to policy, practice, and patient care. However, our review is limited by the current evidence base. Given the ongoing uncertainty and rapidly changing healthcare environment, new information on GP workforce sustainability may have emerged after the completion of our review in response to the NHS Long-Term Plan and Lord Darzi’s report.

### Comparison with existing literature

Prior UK GP workforce studies have predominantly focused on individual-level factors, such as wellbeing, GP self-efficacy, training readiness, resilience, and professional identity (5, 19). This review presents a broader organisational- and system-level analysis of the factors that facilitate or impede GP workforce sustainability. We formulated recommendations (see Table 2) to inform future GP workforce sustainability and effective and equitable patient care.

### Implications for research and/or practice

The development of a sustainable GP workforce to deliver effective and equitable patient care requires examination of the interrelationship between systemic and individual factors, and how these shape the nature of GP work and patient care. Despite current pressures to privatise elements of NHS general practice, there is widespread recognition of the clinical and economic value of general practice as a crucial component of UK primary care. This recognition underscores the need for sustainable financial investment with an emphasis on long-term outcomes to facilitate well-planned and collaborative approaches to support the general practice workforce. Alignment with meaning-making requires organisational and financial support to maximise sustained and direct connections with patients, peers, and communities, thereby fostering ongoing learning systems and cumulative knowledge development. This long-term approach enables GPs and patients to shape and maximise the forward planning of contacts, rather than treating each encounter and practice interaction as a discrete entity.

## Data Availability

Our CMOC analysis and supporting evidence are available in the online supplement.

## Ethical approval

Not applicable.

## Acknowledgements

The authors express their sincere gratitude for the support and contributions provided by stakeholder members and PPI contributors.

