## Supplementary material for "General practitioner workforce sustainability to maximise effective and equitable patient care: a realist review": Online Supplement

Supplementary Information S1: Search strategies

Supplementary Table S1: Included document characteristics

Supplementary Table S2: CMOCs and supporting evidence

Supplementary Information S1: Search strategies

Main searches (April 2023)

MEDLINE

Host: Ovid

Data parameters: MEDLINE® Epub Ahead of Print, In-Process & Other Non-Indexed Citations, Ovid MEDLINE® Daily and Ovid MEDLINE® 1946 to present

Date range searched: 2013 – current (Daily update)

Date searched: 20th April 2023

Searcher: CD

Hits: n=504

| | | |
|---|---|---|
| 1 | general practi*.ti,ab,kw. | 92437 |
| 2 | *General Practitioners/ | 8488 |
| 3 | exp *General Practice/ | 51683 |
| 4 | or/1-3 | 121611 |
| 5 | exp *Work/ | 43272 |
| 6 | exp *Workplace/ | 14,019 |
| 7 | *Job Satisfaction/ | 12818 |
| 8 | exp *Occupational Stress/ | 16249 |
| 9 | exp *"Personnel Staffing and Scheduling"/ | 24174 |
| 10 | (work* or staffing or career* or job*).ti,ab,kw. | 2041372 |
| 11 | (stress* or burn out or burnout).ti,ab,kw. | 1066414 |
| 12 | or/5-11 | 3013398 |
| 13 | exp *Workforce/ | 7820 |
| 14 | *Retirement/ | 6784 |
| 15 | *Personnel Turnover/ | 3003 |
| 16 | *Work Engagement/ | 559 |
| 17 | *Career Mobility/ | 6434 |
| 18 | (workforce* or staff* level*).ti,ab,kw. | 37788 |
| 19 | (part?time* or "less than full time" or LTFT).ti,ab,kw. | 239 |
| 20 | (retire* or retention* or retain* or sustain* or stability or turnover).ti,ab,kw. | 1509240 |
| 21 | (work* adj1 pattern*).ti,ab,kw. | 1433 |
| 22 | (flexible* adj1 work*).ti,ab,kw. | 1028 |
| 23 | ((reason* or rationale* or decision* or intention*) adj1 (leav* or stay*)).ti,ab,kw. | 313 |
| 24 | (career* adj1 (break* or end*)).ti,ab,kw. | 312 |
| 25 | ((participat* or engag*) adj1 (work* or job*)).ti,ab,kw. | 6597 |
| 26 | or/13-25 | 1560462 |
| 27 | 4 and 12 and 26 | 2544 |
| 28 | exp United Kingdom/ | 389181 |
| 29 | (national health service* or nhs*).ti,ab,in. | 265557 |
| 30 | (english not ((published or publication* or translat* or written or language* or speak* or literature* or citation*) adj5 english)).ti,ab. | 48488 |

| | | |
|---|---|---|
| 31 | (gb or "g.b." or britain* or (british* not "british columbia") or uk or "u.k." or united kingdom* or (england* not "new england") or northern ireland* or northern irish* or scotland* or scottish* or ((wales or "south wales") not "new south wales") or welsh*).ti,ab,jw,in. | 2421175 |
| 32 | (bath or "bath's" or ((birmingham not alabama*) or ("birmingham's" not alabama*) or bradford or "bradford's" or brighton or "brighton's" or bristol or "bristol's" or carlisle* or "carlisle's" or (cambridge not (massachusetts* or boston* or harvard*)) or ("cambridge's" not (massachusetts* or boston* or harvard*)) or (canterbury not zealand*) or ("canterbury's" not zealand*) or chelmsford or "chelmsford's" or chester or "chester's" or chichester or "chichester's" or coventry or "coventry's" or derby or "derby's" or (durham not (carolina* or nc)) or ("durham's" not (carolina* or nc)) or ely or "ely's" or exeter or "exeter's" or gloucester or "gloucester's" or hereford or "hereford's" or hull or "hull's" or lancaster or "lancaster's" or leeds* or leicester or "leicester's" or (lincoln not nebraska*) or ("lincoln's" not nebraska*) or (liverpool not (new south wales* or nsw)) or ("liverpool's" not (new south wales* or nsw)) or ((london not (ontario* or ont or toronto*)) or ("london's" not (ontario* or ont or toronto)) or manchester or "manchester's" or (newcastle not (new south wales* or nsw)) or ("newcastle's" not (new south wales* or nsw)) or norwich or "norwich's" or nottingham or "nottingham's" or oxford or "oxford's" or peterborough or "peterborough's" or plymouth or "plymouth's" or portsmouth or "portsmouth's" or preston or "preston's" or ripon or "ripon's" or salford or "salford's" or salisbury or "salisbury's" or sheffield or "sheffield's" or southampton or "southampton's" or st ablans or stoke or "stoke's" or sunderland or "sunderland's" or truro or "truro's" or wakefield or "wakefield's" or wells or westminster or "westminster's" or winchester or "winchester's" or wolverhampton or "wolverhampton's" or (worcester not (massachusetts* or boston* or harvard*)) or ("worcester's" not (massachusetts* or boston* or harvard*)) or (york not ("new york" or ny or ontario* or ont or toronto*)) or ("york's" not ("new york" or ny or ontario* or ont or toronto*))))).ti,ab,in. | 1720005 |
| 33 | (bangor* or "bangor's" or cardiff or "cardiff's" or newport or "newport's" or st asaph or "st asaph's" or st davids or swansea or "swansea's").ti,ab,in. | 69203 |
| 34 | (aberdeen or "aberdeen's" or dundee or "dundee's" or edinburgh or "edinburgh's" or glasgow or "glasgow's" or inverness or (perth not australia*) or ("perth's" not australia*) or stirling or "stirling's").ti,ab,in. | 253603 |
| 35 | (armagh or "armagh's" or belfast or "belfast's" or lisburn or "lisburn's" or londonderry or "londonderry's" or derry or "derry's" or newry or "newry's").ti,ab,in. | 33231 |
| 36 | or/28-35 | 3037247 |
| 37 | (exp africa/ or exp americas/ or exp antarctic regions/ or exp arctic regions/ or exp asia/ or exp oceania/) not (exp United Kingdom/ or europe/) | 3308914 |
| 38 | 36 not 37 | 2875391 |
| 39 | 27 and 38 | 793 |
| 40 | limit 39 to (english language and yr="2013 -Current") | 504 |

Embase
Host: Ovid

Data parameters: Embase 1974 to present

Date range searched: 2013 – current (Daily update)

Date searched: 21st April 2023

Searcher: CD

Hits: n=719

| | | |
|---|---|---|
| 1 | general practi*.ti,ab,kw. | 116652 |
| 2 | *general practitioner/ | 28183 |
| 3 | *general practice/ | 39852 |
| 4 | or/1-3 | 151341 |
| 5 | exp *work/ | 161091 |
| 6 | (work* or staffing or career* or job*).ti,ab,kw. | 2,612,756 |
| 7 | (stress* or burn out or burnout).ti,ab,kw. | 1343279 |
| 8 | or/5-7 | 3867649 |
| 9 | exp *workforce/ | 4028 |
| 10 | *retirement/ | 6756 |
| 11 | *personnel management/ | 32919 |
| 12 | *work engagement/ | 1060 |
| 13 | *career mobility/ | 5235 |
| 14 | (workforce* or staff* level*).ti,ab,kw. | 46188 |
| 15 | (part?time* or "less than full time" or LTFT).ti,ab,kw. | 548 |
| 16 | (retire* or retention* or retain* or sustain* or stability or turnover).ti,ab,kw. | 1843534 |
| 17 | (work* adj1 pattern*).ti,ab,kw. | 2077 |
| 18 | (flexible* adj1 work*).ti,ab,kw. | 1228 |
| 19 | ((reason* or rationale* or decision* or intention*) adj1 (leav* or stay*)).ti,ab,kw. | 471 |
| 20 | (career* adj1 (break* or end*)).ti,ab,kw. | 372 |
| 21 | ((participat* or engag*) adj1 (work* or job*)).ti,ab,kw. | 7548 |
| 22 | or/9-21 | 1925069 |
| 23 | 4 and 8 and 22 | 3636 |
| 24 | United Kingdom/ | 407760 |
| 25 | (national health service* or nhs*).ti,ab,in,ad. | 469261 |
| 26 | (english not ((published or publication* or translat* or written or language* or speak* or literature or citation*) adj5 english)).ti,ab. | 59307 |
| 27 | (gb or "g.b." or britain* or (british* not "british columbia") or uk or "u.k." or united kingdom* or (england* not "new england") or northern ireland* or northern irish* or scotland* or scottish* or ((wales or "south wales") not "new south wales") or welsh*).ti,ab,jw,in,ad. | 3727597 |
| 28 | (bath or "bath's" or ((birmingham not alabama*) or ("birmingham's" not alabama*) or bradford or "bradford's" or brighton or "brighton's" or bristol or "bristol's" or carlisle* or "carlisle's" or (cambridge not (massachusetts* or boston* or harvard*)) or ("cambridge's" not (massachusetts* or boston* or harvard*)) or (canterbury not zealand*) or ("canterbury's" not | 2903989 |

| | | |
|---|---|---|
| | zealand*) or chelmsford or "chelmsford's" or chester or "chester's" or chichester or "chichester's" or coventry or "coventry's" or derby or "derby's" or (durham not (carolina* or nc)) or ("durham's" not (carolina* or nc)) or ely or "ely's" or exeter or "exeter's" or gloucester or "gloucester's" or hereford or "hereford's" or hull or hull's or lancaster or "lancaster's" or leeds* or leicester or "leicester's" or (lincoln not nebraska*) or ("lincoln's" not nebraska*) or (liverpool not (new south wales* or nsw)) or ("liverpool's" not (new south wales* or nsw)) or ((london not (ontario* or ont or toronto*)) or ("london's" not (ontario* or ont or toronto)) or manchester or manchester's or (newcastle not (new south wales* or nsw)) or ("newcastle's" not (new south wales* or nsw)) or norwich or "norwich's" or nottingham or "nottingham's" or oxford or "oxford's" or peterborough or "peterborough's" or plymouth or "plymouth's" or portsmouth or "portsmouth's" or preston or "preston's" or ripon or "ripon's" or salford or salford's or salisbury or "salisbury's" or sheffield or "sheffield's" or southampton or "southampton's" or st ablans or stoke or "stoke's" or sunderland or "sunderland's" or truro or "truro's" or wakefield or "wakefield's" or wells or westminster or "westminster's" or winchester or "winchester's" or wolverhampton or "wolverhampton's" or (worcester not (massachusetts* or boston* or harvard*)) or ("worcester's" not (massachusetts* or boston* or harvard*)) or (york not ("new york" or ny or ontario* or ont or toronto*)) or ("york's" not ("new york" or ny or ontario* or ont or toronto*))))).ti,ab,in,ad. | |
| 29 | (bangor* or "bangor's" or cardiff or "cardiff's" or newport or "newport's" or st asaph or "st asaph's" or st davids or swansea or "swansea's").ti,ab,in,ad. | 119607 |
| 30 | (aberdeen or "aberdeen's" or dundee or "dundee's" or edinburgh or "edinburgh's" or glasgow or "glasgow's" or inverness or (perth not australia*) or ("perth's" not australia*) or stirling or "stirling's").ti,ab,in,ad. | 399773 |
| 31 | (armagh or "armagh's" or belfast or "belfast's" or lisburn or "lisburn's" or londonderry or "londonderry's" or derry or "derry's" or newry or "newry's").ti,ab,in,ad. | 55571 |
| 32 | or/24-31 | 4546440 |
| 33 | (exp "arctic and antarctic"/ or exp oceanic regions/ or exp western hemisphere/ or exp africa/ or exp asia/) not (united kingdom/ or europe/) | 3504690 |
| 34 | 32 not 33 | 4286376 |
| 35 | 23 and 34 | 1275 |
| 36 | limit 35 to (english language and yr="2013 -Current") | 719 |

PsycINFO
Host: Ovid

Data parameters: PsycINFO 1806 to present

Date range searched: 2013 – current (Weekly update)

Date searched: 21st April 2023

Searcher: CD

Hits: n=61

| | | |
|---|---|---|
| 1 | general practi*.ti,ab,id. | 14680 |
| 2 | *general practitioners/ | 4235 |
| 3 | 1 or 2 | 15534 |
| 4 | exp *working conditions/ | 26262 |
| 5 | *"quality of work life"/ | 1345 |
| 6 | exp *job characteristics/ | 10587 |
| 7 | *work-life balance/ or exp *family work relationship/ | 5533 |
| 8 | exp *occupational stress/ or *employee well being/ | 22320 |
| 9 | (work* or staffing or career* or job*).ti,ab,id. | 941313 |
| 10 | (stress* or burn out or burnout).ti,ab,id. | 315931 |
| 11 | or/4-10 | 1187651 |
| 12 | *employee retention/ | 1293 |
| 13 | *employee turnover/ | 5075 |
| 14 | *retirement/ | 4320 |
| 15 | *occupational mobility/ or *career change/ | **1918** |
| 16 | exp *employee engagement/ or exp *employee attitudes/ or *organizational commitment/ or *career commitment/ or *occupational attitudes/ or *"work (attitudes toward)"/ or *job involvement/ or *employee motivation/ | 44203 |
| 17 | (workforce* or staff* level*).ti,ab,id. | 20177 |
| 18 | (part?time* or "less than full time" or LTFT).ti,ab,id. | 104 |
| 19 | (retire* or retention* or retain* or sustain* or stability or turnover).ti,ab,id. | 244306 |
| 20 | (work* adj1 pattern*).ti,ab,id. | 744 |
| 21 | (flexible* adj1 work*).ti,ab,id. | 1169 |
| 22 | ((reason* or rationale* or decision* or intention*) adj1 (leav* or stay*)).ti,ab,id. | 282 |
| 23 | (career* adj1 (break* or end*)).ti,ab,id. | 159 |
| 24 | ((participat* or engag*) adj1 (work* or job*)).ti,ab,id. | 6111 |
| 25 | or/12-24 | 302299 |
| 26 | 3 and 11 and 25 | 436 |
| 27 | united kingdom.ti,ab,id,ca,in,lo. | 353262 |
| 28 | national health service*.mp. or nhs*.ti,ab,id,ca,in,lo. [mp=title, abstract, heading word, table of contents, key concepts, original title, tests & measures, mesh word] | 28447 |
| 29 | (english not ((published or publication* or translat* or written or language* or speak* or literature or citation*) adj5 english)).ti,ab. | 101007 |
| 30 | (gb or "g.b." or britain* or (british* not "british columbia") or uk or "u.k." or united kingdom* or (england* not "new england") or northern ireland* or northern irish* or scotland* or scottish* or ((wales or "south wales") not "new south wales") or welsh*).ti,ab,id,ca,in,lo. | 504922 |
| 31 | (bath or "bath's" or ((birmingham not alabama*) or ("birmingham's" not alabama*) or bradford or "bradford's" or brighton or "brighton's" or bristol or "bristol's" or carlisle* or "carlisle's" or (cambridge not (massachusetts* or boston* or harvard*)) or ("cambridge's" not (massachusetts* or boston* or harvard*)) or (canterbury not zealand*) or ("canterbury's" not zealand*) or chelmsford or "chelmsford's" or chester or "chester's" or chichester or "chichester's" or coventry or | 404232 |

| | | |
|---|---|---|
| | "coventry's" or derby or "derby's" or (durham not (carolina* or nc)) or ("durham's" not (carolina* or nc)) or ely or "ely's" or exeter or "exeter's" or gloucester or "gloucester's" or hereford or "hereford's" or hull or hull's or lancaster or "lancaster's" or leeds* or leicester or "leicester's" or (lincoln not nebraska*) or ("lincoln's" not nebraska*) or (liverpool not (new south wales* or nsw)) or ("liverpool's" not (new south wales* or nsw)) or ((london not (ontario* or ont or toronto*)) or ("london's" not (ontario* or ont or toronto)) or manchester or manchester's or (newcastle not (new south wales* or nsw)) or ("newcastle's" not (new south wales* or nsw)) or norwich or "norwich's" or nottingham or "nottingham's" or oxford or "oxford's" or peterborough or "peterborough's" or plymouth or "plymouth's" or portsmouth or "portsmouth's" or preston or "preston's" or ripon or "ripon's" or salford or salford's or salisbury or "salisbury's" or sheffield or "sheffield's" or southampton or "southampton's" or st ablans or stoke or "stoke's" or sunderland or "sunderland's" or truro or "truro's" or wakefield or "wakefield's" or wells or westminster or "westminster's" or winchester or "winchester's" or wolverhampton or "wolverhampton's" or (worcester not (massachusetts* or boston* or harvard*)) or ("worcester's" not (massachusetts* or boston* or harvard*)) or (york not ("new york" or ny or ontario* or ont or toronto*)) or ("york's" not ("new york" or ny or ontario* or ont or toronto*))))).ti,ab,id,ca,in,lo. | |
| 32 | (bangor* or "bangor's" or cardiff or "cardiff's" or newport or "newport's" or st asaph or "st asaph's" or st davids or swansea or "swansea's").ti,ab,id,ca,in,lo. | 21156 |
| 33 | (aberdeen or "aberdeen's" or dundee or "dundee's" or edinburgh or "edinburgh's" or glasgow or "glasgow's" or inverness or (perth not australia*) or ("perth's" not australia*) or stirling or "stirling's").ti,ab,id,ca,in,lo. | 49991 |
| 34 | (armagh or "armagh's" or belfast or "belfast's" or lisburn or "lisburn's" or londonderry or "londonderry's" or derry or "derry's" or newry or "newry's").ti,ab,id,ca,in,lo. | 6758 |
| 35 | or/27-34 | 661109 |
| 36 | 26 and 35 | 158 |
| 37 | limit 36 to (english language and yr="2013 -Current") | 61 |

CINAHL
Host: EbscoHost

Date range searched: 2013 – current (Update unknown)

Date searched: 21st April 2023

Searcher: CD

Hits: n=644

| | | |
|---|---|---|
| S1 | TI general practi* OR AB general practi* OR SU general practi* | 40,633 |
| S2 | (MM "Physicians, Family") | 13,183 |
| S3 | (MM "Family Practice") | 17,451 |

| | | |
|---|---|---|
| S4 | S1 OR S2 OR S3 | 58,482 |
| S5 | (MM "Work") | 4,339 |
| S6 | (MM "Work Environment") | 17,856 |
| S7 | (MM "Quality of Working Life") | 1,402 |
| S8 | (MM "Job Characteristics") | 1,718 |
| S9 | (MM "Job Satisfaction") | 9,788 |
| S10 | (MM "Personnel Staffing and Scheduling") | 11,041 |
| S11 | (MM "Stress, Occupational+") | 19,847 |
| S12 | (MM "Work-Life Balance") | 1,095 |
| S13 | (MM "Workload") | 5,362 |
| S14 | TI ( (work* or staffing or career* or job*) ) OR AB ( (work* or staffing or career* or job*) ) OR SU ( (work* or staffing or career* or job*) ) | 771,742 |
| S15 | TI ( (stress* or "burn out" or burnout) ) OR AB ( (stress* or "burn out" or burnout) ) OR SU ( (stress* or "burn out" or burnout) ) | 281,935 |
| S16 | S5 OR S6 OR S7 OR S8 OR S9 OR S10 OR S11 OR S12 OR S13 OR S14 OR S15 | 997,106 |
| S17 | (MM "Workforce") | 5,983 |
| S18 | (MM "Personnel Retention") | 4,825 |
| S19 | (MM "Personnel Turnover") | 2,813 |
| S20 | (MM "Retirement") | 5,291 |
| S21 | (MM "Career Mobility") | 2,767 |
| S22 | (MM "Work Engagement") | 918 |
| S23 | (MM "Employee Attitudes") | 2,849 |
| S24 | TI ( (workforce* or "staff* level*") ) OR AB ( (workforce* or "staff* level*") ) OR SU ( (workforce* or "staff* level*") ) | 38,327 |
| S25 | TI ( (part?time* or "less than full time" or LTFT) ) OR AB ( (part?time* or "less than full time" or LTFT) ) OR SU ( (part?time* or "less than full time" or LTFT) ) | 70 |
| S26 | TI ( (retire* or retention* or retain* or sustain* or stability or turnover) ) OR AB ( (retire* or retention* or retain* or sustain* or stability or turnover) ) OR SU ( (retire* or retention* or retain* or sustain* or stability or turnover) ) | 238,439 |
| S27 | TI (work* N2 pattern*) OR AB (work* N2 pattern*) OR SU (work* N2 pattern*) | 1,551 |
| S28 | TI (flexible* N2 work*) OR AB (flexible* N2 work*) OR SU (flexible* N2 work*) | 954 |
| S29 | TI ( ((reason* or rationale* or decision* or intention*) N2 (leav* or stay*)) ) OR AB ( ((reason* or rationale* or decision* or intention*) N2 (leav* or stay*)) ) OR SU ( ((reason* or rationale* or decision* or intention*) N2 (leav* or stay*)) ) | 2,107 |
| S30 | TI ( (career* N2 (break* or end*)) ) OR AB ( (career* N2 (break* or end*)) ) OR SU ( (career* N2 (break* or end*)) ) | 432 |
| S31 | TI ( ((participat* or engag*) N2 (work* or job*)) ) OR AB ( ((participat* or engag*) N2 (work* or job*)) ) OR SU ( ((participat* or engag*) N2 (work* or job*)) ) | 9,748 |
| S32 | S17 OR S18 OR S19 OR S20 OR S21 OR S22 OR S23 OR S24 OR S25 OR S26 OR S27 OR S28 OR S29 OR S30 OR S31 | 285,364 |
| S33 | S4 AND S16 AND S32 | 2,226 |
| S34 | (MH "United Kingdom+") | 321,698 |

| | | |
|---|---|---|
| S35 | TI ( ("national health service*" or nhs*) ) OR AB ( ("national health service*" or nhs*) ) OR SU ( ("national health service*" or nhs*) ) | 44,031 |
| S36 | TI ( (english not ((published or publication* or translat* or written or language* or speak* or literature* or citation*) N5 english)) ) OR AB ( (english not ((published or publication* or translat* or written or language* or speak* or literature* or citation*) N5 english)) ) | 20,520 |
| S37 | TI ( (gb or "g.b." or britain* or (british* not "british columbia") or uk or "u.k." or "united kingdom*" or (england* not "new england") or "northern ireland*" or "northern irish*" or scotland* or scottish* or ((wales or "south wales") not "new south wales") or welsh*) ) OR AB ( (gb or "g.b." or britain* or (british* not "british columbia") or uk or "u.k." or "united kingdom*" or (england* not "new england") or "northern ireland*" or "northern irish*" or scotland* or scottish* or ((wales or "sou ... | 728,634 |
| S38 | TI ( (bath or "bath's" or ((birmingham not alabama*) or ("birmingham's" not alabama*) or bradford or "bradford's" or brighton or "brighton's" or bristol or "bristol's" or carlisle* or "carlisle's" or (cambridge not (massachusetts* or boston* or harvard*)) or ("cambridge's" not (massachusetts* or boston* or harvard*)) or (canterbury not zealand*) or ("canterbury's" not zealand*) or chelmsford or "chelmsford's" or chester or "chester's" or chichester or "chichester's" or coventry or "coventry's" o ... | 1,079,244 |
| S39 | TI ( (bangor* or "bangor's" or cardiff or "cardiff's" or newport or "newport's" or st asaph or "st asaph's" or st davids or swansea or "swansea's") ) OR AB ( (bangor* or "bangor's" or cardiff or "cardiff's" or newport or "newport's" or st asaph or "st asaph's" or st davids or swansea or "swansea's") ) OR AF ( (bangor* or "bangor's" or cardiff or "cardiff's" or newport or "newport's" or st asaph or "st asaph's" or st davids or swansea or "swansea's") ) | 25,400 |
| S40 | TI ( (aberdeen or "aberdeen's" or dundee or "dundee's" or edinburgh or "edinburgh's" or glasgow or "glasgow's" or inverness or (perth not australia*) or ("perth's" not australia*) or stirling or "stirling's") ) OR AB ( (aberdeen or "aberdeen's" or dundee or "dundee's" or edinburgh or "edinburgh's" or glasgow or "glasgow's" or inverness or (perth not australia*) or ("perth's" not australia*) or stirling or "stirling's") ) OR AF ( (aberdeen or "aberdeen's" or dundee or "dundee's" or edinburgh or " ... | 69,749 |
| S41 | TI ( (armagh or "armagh's" or belfast or "belfast's" or lisburn or "lisburn's" or londonderry or "londonderry's" or derry or "derry's" or newry or "newry's") ) OR AB ( (armagh or "armagh's" or belfast or "belfast's" or lisburn or "lisburn's" or londonderry or "londonderry's" or derry or "derry's" or newry or "newry's") ) OR AF ( (armagh or "armagh's" or belfast or "belfast's" or lisburn or "lisburn's" or londonderry or "londonderry's" or derry or "derry's" or newry or "newry's") ) | 11,013 |
| S42 | S34 OR S35 OR S36 OR S37 OR S38 OR S39 OR S40 OR S41 | 1,551,334 |
| S43 | (MH "Africa+") OR (MH "America+") OR (MH "Antarctic Regions") OR (MH "Arctic Regions") OR (MH "Asia+") OR (MH "Atlantic Islands+") OR (MH "Australia+") | 1,465,138 |
| S44 | (MH "Europe+") OR (MH "United Kingdom+") | 645,520 |
| S45 | S43 NOT S44 | 1,424,349 |

| | | |
|---|---|---|
| S46 | S42 NOT S45 | 1,371,907 |
| S47 | S33 AND S46 | 923 |
| | Limiters - Published Date: 20130101-20230431; English Language | 644 |

HMIC
Host: Ovid

Data parameters: Health Management Information Consortium 1979 – March 2023

Date range searched: 2013 – current (March update)

Date searched: 21st April 2023

Searcher: CD

Hits: n=257

| | | |
|---|---|---|
| 1 | general practi*.ti,ab. | 16475 |
| 2 | exp general practice/ | 9456 |
| 3 | exp general practices/ | 394 |
| 4 | general practitioners/ | 9701 |
| 5 | or/1-4 | 25317 |
| 6 | exp employment/ | 4,518 |
| 7 | exp "conditions of employment"/ | **15797** |
| 8 | exp job development/ | 9 |
| 9 | job satisfaction/ | 1163 |
| 10 | occupational stress/ | 1354 |
| 11 | staff working life/ | 86 |
| 12 | (work* or staffing or career* or job*).ti,ab. | 79646 |
| 13 | (stress* or burn out or burnout).ti,ab. | 7220 |
| 14 | or/6-13 | 96505 |
| 15 | staff retention/ | 1636 |
| 16 | staff turnover/ | 272 |
| 17 | exp retirement/ | 952 |
| 18 | occupational mobility/ | 19 |
| 19 | (workforce* or staff* level*).ti,ab. | 6301 |
| 20 | (part?time* or "less than full time" or LTFT).ti,ab. | 32 |
| 21 | (retire* or retention* or retain* or sustain* or stability or turnover).ti,ab. | 11386 |
| 22 | (work* adj1 pattern*).ti,ab. | 350 |
| 23 | (flexible* adj1 work*).ti,ab. | 308 |
| 24 | ((reason* or rationale* or decision* or intention*) adj1 (leav* or stay*)).ti,ab. | 17 |
| 25 | (career* adj1 (break* or end*)).ti,ab. | 75 |
| 26 | ((participat* or engag*) adj1 (work* or job*)).ti,ab. | 158 |
| 27 | or/15-26 | 18283 |
| 28 | 5 and 14 and 27 | 835 |
| 29 | limit 28 to (yr="2013 -Current" and english) | 257 |

Web of Science
Host: Clarivate

Data parameters: Core Collection indexes SCIE, SSCI, AHCI

Date range searched: 2013 – current (Update unknown)

Date searched: 21st April 2023

Searcher: CD

Hits: n=457

| | | |
|---|---|---|
| 1 | TS=("general practi*") | 82,220 |
| 2 | TS=(work* or staffing or career* or job*) | 4,558,155 |
| 3 | TS=(stress* or "burn out" or burnout) | 2,246,505 |
| 4 | #3 OR #2 | 6,536,599 |
| 5 | TS=((workforce* or "staff* level*")) | 47,843 |
| 6 | TS=(part-time or "part time" or "less than full time" or LTFT) | 9.307 |
| 7 | TS=((retire* or retention* or retain* or sustain* or stability or turnover)) | 3,435,391 |
| 8 | TS=((work near/1 pattern*)) | 3732 |
| 9 | TS=((flexible* near/1 work*)) | 3477 |
| 10 | TS=((reason* or rationale* or decision* or intention*) near/1 (leav* or stay*)) | 4217 |
| 11 | TS=(career near/1 (break* or end*)) | 607 |
| 12 | TS=((participat* or engag*) near/1 (work* or job*)) | 17046 |
| 13 | #12 OR #11 OR #10 OR #9 OR #8 OR #7 OR #6 OR #5 | 3502986 |
| 14 | #13 AND #4 AND #1 | 2280 |
| 15 | TS=("united kingdom" OR "national health service*" or nhs* ) | 115067 |
| 16 | TS=((english not ((published or publication* or translat* or written or language* or speak* or literature* or citation*) near/5 english))) | 152827 |
| 17 | (((TS=((gb or "g.b." or britain* or (british* not "british columbia") or uk or "u.k." or united kingdom* or (england* not "new england") or northern ireland* or northern irish* or scotland* or scottish* or ((wales or "south wales") not "new south wales") or welsh*))) OR SO=((gb or "g.b." or britain* or (british* not "british columbia") or uk or "u.k." or united kingdom* or (england* not "new england") or northern ireland* or northern irish* or scotland* or scottish* or ((wales or "south wales") not "new south wales") or welsh*).)) OR OG=((gb or "g.b." or britain* or (british* not "british columbia") or uk or "u.k." or united kingdom* or (england* not "new england") or northern ireland* or northern irish* or scotland* or scottish* or ((wales or "south wales") not "new south wales") or welsh*))) OR AD=((gb or "g.b." or britain* or (british* not "british columbia") or uk or "u.k." or united kingdom* or (england* not "new england") or northern ireland* or northern irish* or scotland* or scottish* or ((wales or "south wales") not "new south wales") or welsh*)) | 6506362 |
| 18 | #16 OR #15 OR #17 | 6631631 |
| 19 | #18 AND #14 | 851 |

| | | |
|---|---|---|
| 20 | Publication Years: 2023 or 2022 or 2021 or 2020 or 2019 or 2018 or 2017 or 2016 or 2015 or 2014 or 2013 AND Languages: English | 457 |

Supplementary Table S1: Included document characteristics

| | |
|---|---|
| Paper Number | 1 |
| Author | Andah et al |
| Year | 2021 |
| Title | Understanding the impact of professional motivation on the workforce crisis in medicine: a rapid review |
| Journal | BJGP Open |
| Region (England, Scotland, Wales, and Northern Ireland) | England |
| Study methodology | Rapid review |
| Type of document | Secondary research |
| Topic or focus of paper | Impact of motivation on medical workforce retention in NHS. |
| Research objectives and focus | 1. Describe factors stated as reasons for leaving the medical profession and the variables that predict and/or explain variation in responses<br>2. Analyse whether the 3Ms model can explain these trends in retention |
| Theoretical framework/theories/ underpinnings | Kanter's three Ms model of workforce motivation: Meaning (repeat and reinforce a larger purpose), Mastery (help people develop deep skills) and Membership (create community by honouring individuality). |
| Description of intervention | First conducted a rapid review to identify common reasons for leaving the medical profession. Then performed a thematic analysis on the resultant dataset to identify core themes and compared these to Kanter's three Ms model of workforce motivation. |
| Rationale for paper/review | Current workforce crisis in the NHS – noted that although many studies examine recruitment, few studies examine retention. |
| What creates meaning that sustains GPs engagement with, and involvement in, work? | Linked the findings of rapid review to 'Meaning' in Kanter's model. Core themes of (dis-) connection and (un-) manageable change create a sense of meaning. Connection is created by a match between doctors and patient expectations of what is possible, and corresponding public respect of healthcare professionals (portrayal of doctors in media plays a big role in influencing this). This connection is influenced at the level of health systems, by governments providing appropriate resources to match patient expectations. Manageable change is characterised by adequate resourcing and the perception of control over the change. With regard to resourcing: if services are going to be moved into primary care, they need to be matched by appropriate resource provision. Likewise, the availability and duration of appointments needs to be increased to match the needs of an ageing population. With regard to control over change: doctors need to be involved in decisions regarding changes to their role or to the structure of the system. |
| What solutions/recommendations are proposed? | Use the 3 Ms model to redesign policy and practice to encourage workforce retention: e.g. deprioritising working weeks to enable employees to undertake (lead) their own quality improvement projects; buddying and mentoring programmes, which also support outside-of-work activities and broader personal development; and active engagement during work time with community service activities. |
| From which stakeholders' perspective? (General | Doctors in primary care (n=47), secondary care (n=14) and both (n=21). |

| | |
|---|---|
| practitioners, patients, other HCPs) | |
| How is work-related joy constructed? (Diversity of ways individuals may think about joy) | Feeling of meaning, mastery and membership; continuity of care; autonomy in clinical practice. |
| What prevents/removes work-related joy? | Burnout; increased workload; long working hours; lack of resources; demands associated with revalidation, appraisals and exams. |
| Organisational characteristics (small, large) | |
| Managing ill-defined illnesses and chronic complex multimorbidity | |
| Human connection with patients | Linked the findings of rapid review to 'Membership' in Kanter's model. Core themes of connection and manageable change create a sense of membership with patients. Mostly covered above: increased time with patients also noted to be an important influence on connection with patients (which is affected by levels of administrative work). |
| Human connection with peers | Linked the findings of rapid review to 'Membership' in Kanter's model. Core theme of manageable change creates a sense of membership with peers. Specifically, less restructuring and certainty about the future nurtures work communities. Core theme of personal and professional support also creates a sense of membership with peers. This can take the form of support from management, and supervision and mentorship. |
| Human connection across other organisations (e.g., secondary care) | |
| Learning systems (e.g., competence, capability, mastery, socially situated) | Linked the findings of rapid review to 'Mastery' in Kanter's model. Core themes of manageable change and connection support the development of mastery. Manageable change: Rapidly changing systems make it difficult for doctors to exercise expertise and develop new areas of expertise. Connection: Doctors feel unconfident in using expertise for fear of missing expectations of patients or health systems. |
| Additional useful information | Of note, some parts of the workforce feel more (or less) supported than others: men reported less job satisfaction than women, although women were shown to have more mental health symptoms. Poor work–life balance and the demands of family commitments has also been shown to affect women more than men. Lower job satisfaction in doctors from minority ethnic backgrounds and those serving urban and deprived populations is also observed |
| Summary of findings | Thematic analysis identified four core themes that explain retention problems: 1. Low morale 2. Disconnect 3. Unmanageable change 4. Lack of personal and professional support. These were linked to Kanter's three Ms model of workforce motivation. |
| Strengths, limitations, and future research directions | The review introduces the three Ms themes of motivating a workforce: meaning, mastery, and membership. The review highlights how these themes might be utilised to aid in further discussions aimed at effectively tackling recruitment and retention in the NHS. The use of a systematic search and theory-informed analysis, as well as its resonance with the results of the Marchand and Peckham8 review, are strengths of this article. |

| | |
|---|---|
| | Limitations included a narrow database search (Embase, MEDLINE, and HMIC) owing to the time constraint on completing this piece of work. Most of the included studies were questionnaires or surveys with no clear validation tools highlighted in some of the articles, possibly introducing a risk of bias.<br><br>New research is now needed to examine the enablers and barriers to development of each element of meaning, mastery, and membership in the current workplace. Within the general practice setting, the present authors propose the need for multiprofessional and patient input into systematically describing what changes are needed, and developing, implementing and evaluating their effects. |

| | |
|---|---|
| Paper Number | 2 |
| Author | Croxson et al |
| Year | 2017 |
| Title | GPs' perceptions of workload in England: a qualitative interview study |
| Journal | BJGP |
| Region (England, Scotland, Wales, and Northern Ireland) | England |
| Study methodology | Semi-structured, qualitative interviews with GPs - followed by thematic analysis of responses (or just qualitative?) |
| Type of document | Primary research |
| Topic or focus of paper | GP workload |
| Research objectives and focus | Gather an in-depth understanding of GPs' perceptions and attitudes towards workload |
| Theoretical framework/theories/ underpinnings | |
| Description of intervention | |
| Rationale for paper/review | GPs report the lowest levels of morale amongst doctors, which impacts retention of the GP workforce. Workload is frequently cited as a major contributor to this. |
| What creates meaning that sustains GPs engagement with, and involvement in, work? | A manageable workload. Four main themes were found to have increased workload to unmanageable levels in recent years: (1) increased patient needs and expectations, (2) a changing relationship between primary and secondary care, (3) bureaucracy and resources, (4) the balance of workload within a practice. |

| | |
|---|---|
| What solutions/recommendations are proposed? | Above all, this study highlights an urgent need to address perceived lack of investment and clinical capacity, already acknowledged by the NHS, and which the General Practice Forward View published in April 2016, may go some way to addressing. It also suggests that influencing, or rather reducing, patient expectations of what primary care can deliver, and reducing bureaucracy, have become key issues, at least until the capacity issues are resolved. |
| From which stakeholders' perspective? (General practitioners, patients, other HCPs) | 34 GPs |
| How is work-related joy constructed? (Diversity of ways individuals may think about joy) | |
| What prevents/removes work-related joy? | Long, intense working days (10-14 hours with little or no break); increase in bureaucracy, particularly with the introduction of the QOF, enhanced services, Care Quality Commission (CQC) inspections, and care planning. |
| Organisational characteristics (small, large) | Patient list size was reported to impact workload due to the number of consultations and more difficulty in establishing continuity. A few GPs described smaller teams as being more affected by increases in workload. |
| Managing ill-defined illnesses and chronic complex multimorbidity | |
| Human connection with patients | Patient needs and patient expectations have increased. This means that GPs cannot form a human connection (or indeed properly address medical needs) within the confines of a 10-minute consultation. Medical factors contributing to increased patient needs are increasing complexity due to an ageing population, chronic disease, mental illness, dementia, multimorbidity, and polypharmacy. With regard to socioeconomic factors, patients in more deprived areas were less well educated about health, and requested more consultations for minor illness, and patients in more affluent areas had higher expectations, and perhaps higher levels of anxiety. Patient expectations within consultations (and also demand for consultations) have increased because of a growing lack of self-management and reduced tolerance for illness. Three main factors were felt to contribute to this: (1) breakdown in society, resulting in patients having less medical and social support from other sources, (2) increasing public access to information about health and illness, (3) inflation of patient expectations, which some participants believed was partially politically and media driven.<br><br>GPs noted that a curter consultation style was needed to address the large volume of information needing to be covered in 10 minutes. However, this may impede human connection with patients.<br><br>Having an ongoing relationship with patients was generally felt to improve workload and enhance patient care, particularly for chronic conditions, because patients would not have to explain their condition to multiple GPs. Continuity was felt by some participants to be disappearing, due to part-time working and employing locums (often as a strategy for dealing with workload), and this had a negative impact on patient care. Paradoxically, a GP also reported that continuity of care could increase workload. |

| | |
|---|---|
| Human connection with peers | The importance of a good team was frequently cited as a positive influence on workload and job satisfaction. Some experienced GPs felt communication among their practice team had diminished due to increased workload intensity, which negatively impacted support. |
| Human connection across other organisations (e.g., secondary care) | Some GPs, particularly those who were more experienced, felt that personal communication with colleagues has been eroded by electronic systems. |
| Learning systems (e.g., competence, capability, mastery, socially situated) | |
| Additional useful information | Participants described a shift in tasks from secondary to primary care, much of which was felt to be appropriate, but which was not accompanied by sufficient resources. GPs now manage conditions previously managed by secondary care, such as palliative care and chronic disease, and patients are discharged to their GP more quickly. However, hospital doctors also increasing request inappropriate tasks of GPs, such as further investigations. GPs described arranging referrals and follow-up as difficult and time consuming, and inefficient communication particularly increased workload – eg not receiving discharge information. Availability, waiting times, and communication with other local health services all led to patients being managed more, and for longer, in primary care.<br><br>Running the practice was a major contributor to workload for partner GPs.<br><br>Consultation style, age and experience and sex (e.g. female GP with disproportionate gynaecological consultations) all influence workload.<br><br>Other members of the practice team (nursing and administrative staff, practice manager) were acknowledged as important to many GPs, with some staff members seen as invaluable to managing workload, and staff absences being very problematic.<br><br>In areas with a significant ethnic minority population, the time needed to organise and utilise translation services, and explain the pathways through and constraints of the NHS, considerably added to workload.<br><br>Increase in telephone consultations has increased ease of patient access to doctors and therefore demand for consultations. |
| Summary of findings | The workload of GPs has increased to unmanageable levels in recent years, the reasons for which can be divided into four main themes: (1) increased patient needs and expectations, (2) a changing relationship between primary and secondary care, (3) bureaucracy and resources, (4) the balance of workload within a practice. |
| Strengths, limitations, and future research directions | This qualitative study provides a more in-depth assessment of GPs' perceptions and concerns regarding workload compared with quantitative methods. Interviews were conducted by a non-GP, so participants may have felt able to speak more freely about feelings on workload.<br><br>Limitations include those associated with qualitative research, such as the small, diverse sample, which is not necessarily representative of GPs in England. It is possible that there was natural self-selection, resulting in over-reporting of workload problems. |

| | |
|---|---|
| Paper Number | 3 |
| Author | Eley |
| Year | 2018 |
| Title | Professional resilience in GPs working in areas of socioeconomic deprivation: a qualitative study in primary care |
| Journal | BJGP |
| Region (England, Scotland, Wales, and Northern Ireland) | England |
| Study methodology | Interviews and focus groups with GPs who work in areas of high socioeconomic deprivation, and subsequent thematic analysis of responses. |
| Type of document | Primary research |
| Topic or focus of paper | Professional resilience |
| Research objectives and focus | To understand how GPs working in areas of high socioeconomic deprivation consider professional resilience. |
| Theoretical framework/theories/ underpinnings | |
| Description of intervention | |
| Rationale for paper/review | GPs working in areas of high socioeconomic deprivation are at increased risk of professional burnout. An understanding of this is necessary to improve workforce retention. |
| What creates meaning that sustains GPs engagement with, and involvement in, work? | Three major themes arose relating to resilience, therefore meaning and engagement: (1) flexibility and adaptability, (2) resilience is dependent on others and the system, (3) resilience at work requires integration between work and life – both in terms of activities and values. |
| What solutions/recommendations are proposed? | Future interventions to target practitioner resilience, particularly in highly-deprived areas, must appreciate the multidimensional nature of resilience and nurture teams.<br><br>Efforts to protect practitioners must allow professionals flexibility rather than enforcing conformity, support teams to support themselves, and foster the integration of personal and professional values rather than enforcing systems that set them against each other. |
| From which stakeholders' perspective? (General practitioners, patients, other HCPs) | |
| How is work-related joy constructed? (Diversity of | Ability to work flexibility – e.g. half days because get tired in the afternoon; ability to combine GP with other activities – e.g. teaching at a university; fulfilment in personal life alongside fulfilment in professional life; values and beliefs aligning with GP work. |

| | |
|---|---|
| ways individuals may think about joy) | |
| What prevents/removes work-related joy? | |
| Organisational characteristics (small, large) | Resilience can be context dependent - “One participant had experienced feelings of burnout and an inability to cope in a previous practice, but in a new environment felt able to function and maintain their wellbeing” |
| Managing ill-defined illnesses and chronic complex multimorbidity | |
| Human connection with patients | |
| Human connection with peers | Having a supportive team was seen as a buffer and support for practitioners, who otherwise might not cope on their own. Routines that bring GPs together were seen as beneficial in fostering this. |
| Human connection across other organisations (e.g., secondary care) | |
| Learning systems (e.g., competence, capability, mastery, socially situated) | Resilience increased with experience, and perceived increase in ability to cope with adversity (e.g. very sick patients). |
| Additional useful information | Ability to work flexibility – e.g. half days because get tired in the afternoon – noted as important to improving resilience. |
| Summary of findings | The present study found that GPs working in areas of high socioeconomic deprivation had a view of resilience that was more complex than the simplistic notion of personal strength and bouncing back from adversity. Resilience was seen as requiring flexibility and adaptability, it was enacted through teams rather than by individuals, and involved integration between work and personal values. |
| Strengths, limitations, and future research directions | This study is the first to explore resilience exclusively among GPs working in areas of high deprivation.<br><br>Several of the interviewees were female or had <5 years’ experience. |

| | |
|---|---|
| Paper Number | 4 |
| Author | Humphrey & Cleaver |
| Year | 2018 |
| Title | General practitioners’ perceptions on their role in light of the NHS five year forward view: a qualitative study |

| | |
|---|---|
| Journal | London Journal of Primary Care |
| Region (England, Scotland, Wales, and Northern Ireland) | England |
| Study methodology | Interviews and focus groups with GPs who work(ed) in CCGs, and subsequent thematic analysis of responses. |
| Type of document | Primary research |
| Topic or focus of paper | GPs perceptions of changing role |
| Research objectives and focus | Produce a description of how GP's construct their current and future general practice, professional status and identify within the context of the current NHS transformation agenda. |
| Theoretical framework/theories/ underpinnings | |
| Description of intervention | |
| Rationale for paper/review | Lack of academic literature on General Practitioners' perceptions on their role in light of the new models of care proposed by the NHS Five Year Forward View. |
| What creates meaning that sustains GPs engagement with, and involvement in, work? | Number of changes to the role of a traditional GP noted in this paper. Although GPs felt these were necessary, some of these changes erode work-related joy, human connection with patients and mastery (see below). |
| What solutions/recommendations are proposed? | |
| From which stakeholders' perspective? (General practitioners, patients, other HCPs) | GPs |
| How is work-related joy constructed? (Diversity of ways individuals may think about joy) | |
| What prevents/removes work-related joy? | Volume of work, lack of adequate resources, burden of administrative work (especially in the context of changing role expressed in NHS Five Year Forward View). |
| Organisational characteristics (small, large) | |

| | |
|---|---|
| Managing ill-defined illnesses and chronic complex multimorbidity | |
| Human connection with patients | Increased patient demand alongside increased patient expectations in recent years (a major contributing factor of which is population change).<br><br>The development of multiprofessional teams was perceived to be eroding continuity of care. |
| Human connection with peers | |
| Human connection across other organisations (e.g., secondary care) | |
| Learning systems (e.g., competence, capability, mastery, socially situated) | Concern that changes expressed in Five Year Forward View might deskill GPs from a clinical perspective. |
| Additional useful information | GPs reported frustrations with the unequal balance of funding between primary care and secondary care.<br><br>There was a consensus that the GP of the future will need more leadership and managerial qualities. This paper did not examine GPs' perceptions of this, however. |
| Summary of findings | Although much of this study examined GPs perspectives on proposed changes to the system as opposed to their work, a few factors which influence GPs *changing* relationship with work reported. |
| Strengths, limitations, and future research directions | First qualitative study that examines how GP's are responding to and engaging with new care models and associated agenda arising from 5YFV.<br><br>Small sample size. |

| | |
|---|---|
| Paper Number | 5 |
| Author | Nelson et al |
| Year | 2019 |
| Title | Skill-mix change in general practice: a qualitative comparison of three 'new' non-medical roles in English primary care |
| Journal | BJGP |
| Region (England, Scotland, Wales, and Northern Ireland) | England |
| Study methodology | Interviews and focus groups with stakeholders involved in the implementation of advanced practitioners, physician associates and practice pharmacists, and subsequent thematic analysis of responses. |

| | |
|---|---|
| Type of document | Primary research. |
| Topic or focus of paper | Multiprofessional teams in GP. |
| Research objectives and focus | To compare how three non-medical roles were being established in general practice, understand common implementation barriers, and identify measurable impacts or unintended consequences. |
| Theoretical framework/theories/ underpinnings | |
| Description of intervention | |
| Rationale for paper/review | Introducing new non-medical roles is a common solution to the GP workforce challenge. There are few previous studies that investigate the implementation of these roles and offer lessons for stakeholders. |
| What creates meaning that sustains GPs engagement with, and involvement in, work? | |
| What solutions/recommendations are proposed? | The financial cost of employing new role professionals against evidence of their value and contribution in general practice needs to be considered, since the very initiatives aimed at alleviating pressure may paradoxically place increased strain on staff, at least initially, and mean that GPs incur extra workload in supervision or mentoring.<br><br>Improved dialogue between training organisations and general practice could facilitate this process, helping to set realistic expectations, harness the commitment of individual practices, and prepare them to support practitioner training. |
| From which stakeholders' perspective? (General practitioners, patients, other HCPs) | Variety of stakeholders, including GPs themselves, involved in implementing new non-medical roles within the GP setting. |
| How is work-related joy constructed? (Diversity of ways individuals may think about joy) | Non-medical roles like PAs can reduce the workload for GPs. |
| What prevents/removes work-related joy? | In some cases, non-medical roles like PAs can increase the workload for GPs due to need for training; GPs feel responsibility for clinical practice of non-medical roles like PAs, potentially increasing stress. |
| Organisational characteristics (small, large) | |
| Managing ill-defined illnesses and chronic complex multimorbidity | |

| | |
|---|---|
| Human connection with patients | |
| Human connection with peers | Perception amongst GPs that new roles are ‘taking our jobs’. |
| Human connection across other organisations (e.g., secondary care) | |
| Learning systems (e.g., competence, capability, mastery, socially situated) | |
| Additional useful information | |
| Summary of findings | This study focussed on the implementation of non-medical staff in a GP setting. However, a few points were relevant to GP work. |
| Strengths, limitations, and future research directions | The study’s key strength was the in-depth, in-context, multistakeholder comparison of the introduction of three non-medical roles in general practice, informed by an extensive literature review.<br><br>The main study limitation was the missing patient perspective on new roles. The number of GP host staff in the study was relatively low, which could have limited conclusions, however the views of these GP participants varied and may be reflective of wider GP experiences in the context of new roles. |

| | |
|---|---|
| Paper Number | 6 |
| Author | Roger Jones |
| Year | 2021 |
| Title | General practice in the years ahead: relationships will matter more than ever |
| Journal | BJGP |
| Region (England, Scotland, Wales, and Northern Ireland) | N/A |
| Study methodology | Editorial |
| Type of document | Secondary research |
| Topic or focus of paper | Relationship-based care in GP |

| | |
|---|---|
| Research objectives and focus | N/A |
| Theoretical framework/theories/ underpinnings | |
| Description of intervention | |
| Rationale for paper/review | The rapid, radical changing to GP working patterns posed by COVID-19 constitutes an important opportunity to rethink priorities for the future. |
| What creates meaning that sustains GPs engagement with, and involvement in, work? | Relationship based care, which has two main components: patient-centred care and continuity of care. |
| What solutions/recommendations are proposed? | Creating organisational arrangements in which relationship-based medicine can be practised is likely to be a major step in the right direction. |
| From which stakeholders' perspective? (General practitioners, patients, other HCPs) | GP |
| How is work-related joy constructed? (Diversity of ways individuals may think about joy) | |
| What prevents/removes work-related joy? | |
| Organisational characteristics (small, large) | |
| Managing ill-defined illnesses and chronic complex multimorbidity | |
| Human connection with patients | Relationship based care often incorporates continuity of care but goes beyond it, signifying patient care that is enhanced by a high-quality relationship between doctor and patient, and is characterised by trust, mutual respect, and the sharing of power. This has been shown to increase the wellbeing of doctors. |
| Human connection with peers | |

| | |
|---|---|
| Human connection across other organisations (e.g., secondary care) | |
| Learning systems (e.g., competence, capability, mastery, socially situated) | |
| Additional useful information | |
| Summary of findings | Makes the case for relationship-based care more from a patient perspective than a GP perspective. |
| Strengths, limitations, and future research directions | |

| | |
|---|---|
| Paper Number | 7 |
| Author | Jackson et al |
| Year | 2017 |
| Title | Barriers and facilitators to integration of physician associates into the general practice workforce: a grounded theory approach |
| Journal | BGJP |
| Region (England, Scotland, Wales, and Northern Ireland) | Suggests England |
| Study methodology | Themes generated from interviews with stakeholders were used to derive a theoretical framework and define search terms for a focused scoping review of the literature. This framework was then tested in stakeholder focus groups to create a conceptual model describing key barriers and facilitators to the integration of the PA role into general practice teams within the region. |
| Type of document | Primary & secondary research |
| Topic or focus of paper | |
| Research objectives and focus | To investigate the barriers and facilitators to the integration of PAs into the general practice workforce. |
| Theoretical framework/theories/ underpinnings | |
| Description of intervention | |
| Rationale for paper/review | Physician associates are increasingly described as part of the solution to workforce shortages in general practice, but there is little evidence on how easily this might be achieved. |

| | |
|---|---|
| What creates meaning that sustains GPs engagement with, and involvement in, work? | Workload – felt to have increased to unsustainable levels in recent years, and GPs expressed mixed feelings about whether the introduction of PAs would be able to respond to this. On one hand, they are filling a gap in the workforce. On the other hand, the levels of supervision required might increase GP workload (especially because PAs cannot prescribe and with complex patients). |
| What solutions/recommendations are proposed? | There was concern about how prepared PAs would be to manage healthcare presentations in a primary care setting, particularly around medical complexity. This was most strongly voiced by GPs but echoed by ANPs and recognised by patients. Professional and educational institutions involved in postgraduate education and training of PAs should reflect on curricular design and delivery in seeking to address this issue.<br><br>Additional barriers relate to regulatory factors, particularly with respect to prescribing rights and indemnity. A strategic approach to addressing this has been an important factor in supporting integration of the role internationally. It is unlikely that significant numbers of PAs will integrate into primary care teams until this is addressed. |
| From which stakeholders' perspective? (General practitioners, patients, other HCPs) | Stakeholders involved in the implementation of PAs: 30 GPs, 11 ANPs and 10 patients. |
| How is work-related joy constructed? (Diversity of ways individuals may think about joy) | |
| What prevents/removes work-related joy? | Perception that PAs are undermining GPs and 'stealing their jobs'. |
| Organisational characteristics (small, large) | |
| Managing ill-defined illnesses and chronic complex multimorbidity | |
| Human connection with patients | |
| Human connection with peers | |
| Human connection across other organisations (e.g., secondary care) | |
| Learning systems (e.g., competence, capability, mastery, socially situated) | |

| | |
|---|---|
| Additional useful information | |
| Summary of findings | The paper highlights key concerns of GPs about the recent introduction of PAs into multiprofessional teams. |
| Strengths, limitations, and future research directions | The modified grounded theory methodology provides credibility to this study.8 This allowed themes generated directly from the community of primary care to shape the initial theoretical framework, and the focus of the literature search, with these themes then being formally tested through focus groups.<br><br>Limitations in the study also need to be recognised. Many of the GPs were already involved in training and supervision of general practice specialty trainees, and therefore, as a group, will be more familiar with clinical supervision in the workplace than the wider workforce. The general lack of knowledge regarding the role among participants will have naturally influenced some of the opinions voiced. |

| | |
|---|---|
| Paper Number | 8 |
| Author | Jeffers & Baker |
| Year | 2016 |
| Title | Continuity of care: still important in modern-day general practice |
| Journal | BJGP |
| Region (England, Scotland, Wales, and Northern Ireland) | N/A |
| Study methodology | Editorial |
| Type of document | Secondary research |
| Topic or focus of paper | Continuity of care |
| Research objectives and focus | |
| Theoretical framework/theories/ underpinnings | |
| Description of intervention | |
| Rationale for paper/review | |
| What creates meaning that sustains GPs engagement with, and involvement in, work? | The ability to develop relationships over time with a patient. 43% of GPs say that insufficient time with patients negatively impacts on their commitment to general practice; 70% state that implementing longer consultation times would resolve the pressures they were under in relation to continuity. |

| | |
|---|---|
| What solutions/recommendations are proposed? | An increased workforce; greater funding for practice-based pharmacists, mental health therapists, practice-based nurses, and physician associates; a reduction in practice burdens that will release time to care for patients; greater use of technology; longer consultations; identifying and prioritising the patients who will benefit from continuity of care most; formation of GP federations, which enable practices to share back-office functions, reduce cost, and broaden skill-mix while retaining the local nature of general practice; a greater emphasis on managerial and informational continuity where traditional relational continuity cannot be provided. |
| From which stakeholders' perspective? (General practitioners, patients, other HCPs) | GP |
| How is work-related joy constructed? (Diversity of ways individuals may think about joy) | |
| What prevents/removes work-related joy? | |
| Organisational characteristics (small, large) | |
| Managing ill-defined illnesses and chronic complex multimorbidity | |
| Human connection with patients | |
| Human connection with peers | |
| Human connection across other organisations (e.g., secondary care) | |
| Learning systems (e.g., competence, capability, mastery, socially situated) | |
| Additional useful information | Importance factors currently impacting continuity of care in GP include:<br>1. Current lack of GPs<br>2. More GPs choosing locum or part-time work<br>3. New models of care and new working structure |

| | |
|---|---|
| | 4. Successive governments prioritising access over continuity (i.e. more important to see *a* GP rather than the *right* GP) |
| Summary of findings | Highlights the importance of continuity of care for GPs engagement with work (and, indeed, for patients). Discusses the main factors which are limiting continuity of care and explores solutions. |
| Strengths, limitations, and future research directions | |

| | |
|---|---|
| Paper Number | 9 |
| Author | Lyon-Maris et al |
| Year | 2015 |
| Title | GP workload: time for a rethink of the generalist model of care to promote retention |
| Journal | BJGP |
| Region (England, Scotland, Wales, and Northern Ireland) | England |
| Study methodology | Survey of and interviews with GPs, followed by thematic analysis of responses. |
| Type of document | Primary research |
| Topic or focus of paper | How we can change the 'traditional' role of a GP to promote workforce retention |
| Research objectives and focus | To seek the experiences of GPs and their views of working in general practice today, and to ask them to look ahead to the future. |
| Theoretical framework/theories/ underpinnings | |
| Description of intervention | |
| Rationale for paper/review | Although much has been written about the workforce crisis in GP, few solutions have been proposed. |
| What creates meaning that sustains GPs engagement with, and involvement in, work? | Professional autonomy – GP today was perceived as 'one size fits all' (10 minutes for every patient with little scope for GPs to take on varied roles and/or develop their expertise in certain areas). This was perceived as eroding professional autonomy and leaving GPs feeling overwhelmed. Informants thought that for a career in general practice to become attractive again there should be an emphasis on developing a flexible workforce structure, with portfolio roles and interests (such as education/training, mentoring, special interest, appraising, and research) to sit alongside clinical work. |

| | |
|---|---|
| What solutions/recommendations are proposed? | Re-model the traditional, 'generalist' model of GP. Divide primary care into four main groups: acute and chronic conditions, which are subdivided on the basis of whether the patient is mobile (ambulatory and non-ambulatory). Each of these types of care offers the opportunity to provide services in different ways, by different groups of healthcare practitioners, depending on demand and supply in a locality; further, it allows the GP workforce to be allocated to each sector depending on experience and career preferences. In addition to providing career structure and professional autonomy for GPs, this would be better for patients (i.e. lots of complex patients need longer than 10 minutes).<br><br>On a system level, resourcing for general practice needs to keep up with service delivery costs. |
| From which stakeholders' perspective? (General practitioners, patients, other HCPs) | GPs |
| How is work-related joy constructed? (Diversity of ways individuals may think about joy) | |
| What prevents/removes work-related joy? | Intense workload without breaks; rising levels of administration within a culture of target management and reporting. |
| Organisational characteristics (small, large) | |
| Managing ill-defined illnesses and chronic complex multimorbidity | |
| Human connection with patients | |
| Human connection with peers | |
| Human connection across other organisations (e.g., secondary care) | |
| Learning systems (e.g., competence, capability, mastery, socially situated) | |
| Additional useful information | |
| Summary of findings | This paper highlights the importance of professional autonomy and career structure, and proposes policies to increase this. |

| | |
|---|---|
| Strengths, limitations, and future research directions | The key point to note about this study is that the model of GP proposed – namely, dividing primary care into four areas – was not discussed with the GPs consulted in the surveys and interviews. It was merely flagged as a potential solution to some of the issues raised by GPs. |

| | |
|---|---|
| Paper Number | 10 |
| Author | Jefferson et al |
| Year | 2022 |
| Title | GP wellbeing during the COVID-19 pandemic: a systematic review |
| Journal | BJGP |
| Region (England, Scotland, Wales, and Northern Ireland) | N/A |
| Study methodology | Systematic review including thematic analysis of studies which met the inclusion criteria |
| Type of document | Secondary research |
| Topic or focus of paper | Primary care doctors' mental health and wellbeing |
| Research objectives and focus | To synthesise international evidence exploring the impact of COVID-19 on primary care doctors' mental health and wellbeing, and identify risk factors associated with their psychological wellbeing during this time. |
| Theoretical framework/theories/ underpinnings | |
| Description of intervention | |
| Rationale for paper/review | Doctors' organisations in the UK have reported worrying levels of work-related stress and burnout in the GP workforce for some time, and the COVID-19 pandemic has presented clear new challenges. |
| What creates meaning that sustains GPs engagement with, and involvement in, work? | Intense workload can erode meaning – which GPs in most studies have reported to have risen due to phone/video consultations (however, there are a few examples of studies reporting that phone/video consultations have decreased workload). |
| What solutions/recommendations are proposed? | No specific policy changes proposed, only that policymakers need to support GPs in the context of COVID-19. |

| | |
|---|---|
| From which stakeholders' perspective? (General practitioners, patients, other HCPs) | |
| How is work-related joy constructed? (Diversity of ways individuals may think about joy) | |
| What prevents/removes work-related joy? | Inadequate PPE and/or exposure to COVID-19; information overload in the context of conflicting guidelines early in the COVID-19 pandemic; (lack of) organisational preparedness for COVID waves; |
| Organisational characteristics (small, large) | |
| Managing ill-defined illnesses and chronic complex multimorbidity | |
| Human connection with patients | |
| Human connection with peers | Studies describe the sources of support used by GPs, with positive sentiment described around social support, but less positivity with regards state organisations or employers |
| Human connection across other organisations (e.g., secondary care) | Studies reported challenges in the communication and lack of cohesion across primary and other healthcare sectors increasing stress. |
| Learning systems (e.g., competence, capability, mastery, socially situated) | |
| Additional useful information | Gender and age differences found in primary care doctors' mental health. Studies found gender differences, with female GPs reporting worse outcomes on all facets of psychological wellbeing. Experiences according to age varied across studies, with higher stress reported in older groups but more anxiety and depression in younger groups. |
| Summary of findings | This review found that COVID-19 adversely affected primary care doctors' wellbeing on an international scale. It does not go into great detail on the sources of this stress, although there are relevant data in supplementary table 2. |
| Strengths, limitations, and future research directions | To the authors' knowledge, this is the first systematic review exploring GPs' psychological wellbeing during the COVID-19 pandemic. A rigorous methodology was used, and the combination of qualitative and quantitative literature generates an in-depth understanding of stressors and outcomes.<br><br>Although the quality of the evidence was generally good, there were some limitations in consideration of confounders and in reporting across studies, with results pertaining to GPs often not disentangled from other healthcare workers, limiting the pool of research. Furthermore, most studies used cross-sectional survey designs so there may be selection bias in the types of GPs responding. |

| | |
|---|---|
| Paper Number | 11 |
| Author | Jefferson et al |
| Year | 2022 |
| Title | GP workforce crisis: what can we do now? |
| Journal | BJGP |
| Region (England, Scotland, Wales, and Northern Ireland) | N/A |
| Study methodology | Editorial |
| Type of document | Secondary research |
| Topic or focus of paper | |
| Research objectives and focus | Consider potential solutions to support general practice in the immediate term and retain those doctors we have |
| Theoretical framework/theories/ underpinnings | The ABC of needs that has previously been shown to be integral to promoting the wellbeing of the health workforce in general: Autonomy, Belonging and Contribution. |
| Description of intervention | |
| Rationale for paper/review | To address the workforce crisis in GP, in addition to recruiting new doctors we need to retain the doctors we do have, as increasing the pool of doctors will take time. |
| What creates meaning that sustains GPs engagement with, and involvement in, work? | 1. Autonomy and control: GPs feeling able to exert control over their working lives.<br>2. Belonging: GPs feeling valued, respected and supported in their work.<br>3. Contribution: GPs experiencing effectiveness and feeling they are delivering valued outcomes. |
| What solutions/recommendations are proposed? | Appropriate of technology, multiprofessional teams and The Additional Roles Reimbursement Scheme to address workload challenges and increase GP autonomy and control<br><br>Greater understanding of how the implementation of ICSs can enable general practice to really belong and contribute in an equitable way across the health system. |

| | |
|---|---|
| | Progress has been made during the pandemic to develop a culture that promotes and nurtures staff wellbeing; however, there remains issues of presenteeism and a perceived tokenism to these measures. All staff in general practice need to feel it is ok to not be ok. Support is being offered from multiple avenues and schemes such as ‘Looking after you too’ have been met with demonstrable success. A more overt shift towards a culture that allows time for and normalises the proactive pursuit of wellbeing is perhaps necessary if the profession’s sense of belonging is not to be eroded further.<br><br>Build on the civic partnerships seen during the COVID-19 vaccination rollout to strengthen communities and the public perception of primary care.<br><br>The importance of patient participation and involvement in the development of Integrated Care Systems has been highlighted as an opportunity to reset relationships between the public and health services.<br><br>Integration across all sectors of health and social care has an important role to play in not only delivering effective outcomes, but also in creating a wider sense of mutual understanding of what each specialty does. Potential for general practice rotations as standard for all post-graduate doctors.<br><br>In order to allow the development of mastery, in North Yorkshire, workload is protected allowing space to grow into the profession, develop interests, continue learning, and prevent burnout. |
| From which stakeholders’ perspective? (General practitioners, patients, other HCPs) | GP |
| How is work-related joy constructed? (Diversity of ways individuals may think about joy) | |
| What prevents/removes work-related joy? | Increased demand alongside a continued national emphasis on quicker and longer access erodes autonomy and control; culture of fear and blame erodes belonging |
| Organisational characteristics (small, large) | Highly functioning and effective teams, effective IT systems and buildings that are fit for purpose all increase perception of autonomy and control. |
| Managing ill-defined illnesses and chronic complex multimorbidity | |

| | |
|---|---|
| Human connection with patients | The political and media-driven narrative that described the role of UK general practice during the pandemic did much to demoralise the profession and threatened their all so important relationship with patients; sense of belonging can be eroded by remote working |
| Human connection with peers | |
| Human connection across other organisations (e.g., secondary care) | |
| Learning systems (e.g., competence, capability, mastery, socially situated) | |
| Additional useful information | |
| Summary of findings | This editorial highlight some of the workforce issues and opportunities for change in general practice, using the lens of clinicians' need for autonomy, belonging, and contribution in their working lives. |
| Strengths, limitations, and future research directions | |

| | |
|---|---|
| Paper Number | 12 |
| Author | Jefferson et al |
| Year | 2023 |
| Title | General practitioner well-being during the COVID-19 pandemic: a qualitative interview study |
| Journal | BMJ Open |
| Region (England, Scotland, Wales, and Northern Ireland) | All |
| Study methodology | In-depth qualitative interviews, conducted remotely by telephone or video call, with UK National Health Service GPs – followed by thematic analysis of resulting dataset. |
| Type of document | Primary research |
| Topic or focus of paper | GP wellbeing |
| Research objectives and focus | This study sought to explore the lived experience of UK GPs during COVID-19, and the pandemic's impact on their psychological well-being. |

| | |
|---|---|
| Theoretical framework/theories/ underpinnings | |
| Description of intervention | |
| Rationale for paper/review | The COVID-19 pandemic presented new challenges for general practitioners' (GPs') mental health and well-being, with growing international evidence of its negative impact. While there has been a wide UK commentary on this topic, research evidence from a UK setting is lacking. |
| What creates meaning that sustains GPs engagement with, and involvement in, work? | |
| What solutions/recommendations are proposed? | To address worsening public perception of GPs: Communication about delays in outpatient appointments and routine surgery was seen as vital, as were government campaigns encouraging health awareness about common illnesses and more signposting to appropriate specialists.<br><br>Flagged formal support structures; ranging from coaching and mentoring support (which several participants had used) to more formal mental health support.<br><br>Some GPs developed portfolio careers to manage work pressure and support wellbeing.<br><br>Phased retirement schemes can reduce workload and therefore retain GPs. |
| From which stakeholders' perspective? (General practitioners, patients, other HCPs) | GPs |
| How is work-related joy constructed? (Diversity of ways individuals may think about joy) | GPs described working additional hours to manage vaccinations, but with a sense of teamwork and pride. |
| What prevents/removes work-related joy? | Uncertainty in the context of COVID-19 caused stress, particularly with regard to future demand; increasing workload; anxiety about missing something in the context of overwhelming workload; GPs reported fear of putting themselves and family members at risk in the context of COVID-19; Changing guidance around implementation of 'hot' sites, use of and access to Personal Protective Equipment (PPE) heightened anxiety; large amounts of administration which is often unpaid; feeling that trainees were used as 'cannon fodder' during COVID-19 and had missed out on training; negative financial impacts of the pandemic were described by some GPs, mostly due to reduced availability of locum work |
| Organisational characteristics (small, large) | Adaptations to organisational characteristics in the context of COVID-19 caused stress (eg remote working and hot sites); E-consultation systems were perceived to increase demand due to greater accessibility; Telephone and video consultations were noted to decrease workload when used appropriately; some GPs suggested that well-being support was perhaps more easily adopted by larger practices with greater |

| | |
|---|---|
| | infrastructure; team ‘huddles’ were used to debrief on complex cases, provide social support and share anxieties, but small rooms and safe distancing in some practices prohibited in-person staff meetings. |
| Managing ill-defined illnesses and chronic complex multimorbidity | |
| Human connection with patients | In the context of COVID-19, GPs described emotional strain from patients presenting late with serious conditions; Telephone and video consultations are making some GPs feel isolated (GPs noted that personal contact with patients had encouraged their career choice); GPs noted that public perception of GPs had been eroded during the pandemic by negative portrayals by government, NHS England and the media (indeed, this was noted to be upsetting given the amount GPs had worked) |
| Human connection with peers | Low motivation, dissatisfaction with work, frustration and anger were caused by social isolation in the context of COVID-19; many GPs described reluctance to seek support because of stigma and guilt from taking time off as this would burden their colleagues without a ‘buffer in the system’; during the pandemic, a greater sense of camaraderie and improved working across PCNs was reported; isolation from teams was problematic particularly for early-career GPs who lacked support and found it difficult to integrate; shared breaks provided opportunities to raise difficulties informally, which was important to some who felt less inclined to seek formal support either due to workload pressures or stigma; interviewees sought informal support through family and friends, colleagues and peers. They described the benefits of talking to other medics who could relate to their experiences |
| Human connection across other organisations (e.g., secondary care) | Some GPs described conflict between primary and secondary care, relating to lengthy hospital waiting lists and some service closures increased workload for GPs, who felt they were the only support for some high-risk patients. |
| Learning systems (e.g., competence, capability, mastery, socially situated) | |
| Additional useful information | Even before COVID-19, GPs highlighted that treatment advances and shifting care out of hospitals had significantly increased workload. During COVID-19, workload was increased by increased patient demand (particularly mental health problems) alongside increased administrative load due to vaccine role out.<br><br>Some participants were disenchanted with initiatives that sought to improve ‘resilience’ as they felt that this placed the onus of responsibility at an individual rather than structural level. |
| Summary of findings | Highlights a range of factors which impacted the wellbeing of GPs during the pandemic – many of which remain. |
| Strengths, limitations, and future research directions | This research provides rich and contextualised understanding of the experiences of a varied sample of GPs during the pandemic, which our recent systematic review16 identified as lacking from a UK setting. While there may be selection bias in the views expressed by GPs willing to share experiences, for example, GPs experiencing particular difficulties may have been more willing to participate, our interview findings are consistent with other international research and wider commentary on this topic.<br><br>Further research could explore these findings more widely through quantitative methods, preferably with some comparison with prepandemic well-being scores. E-consultation systems, which appear to have increased demand, could be further evaluated, as should |

| | |
|---|---|
| | schemes to supplement the GP workforce with other non-medical staff through the Additional Roles Reimbursement Scheme that formed part of UK GP contract revisions. |

| | |
|---|---|
| Paper Number | 13 |
| Author | Le Floch et al |
| Year | 2016 |
| Title | Which positive factors determine the GP satisfaction in clinical practice? A systematic literature review |
| Journal | BMC Family Practice |
| Region (England, Scotland, Wales, and Northern Ireland) | International – 17 articles met this systematic review's inclusion criteria with 1 pertaining to the UK. However, we opted to include because many of the findings are applicable to a UK context. |
| Study methodology | Systematic review |
| Type of document | Secondary research |
| Topic or focus of paper | GP satisfaction |
| Research objectives and focus | Systematically studying the literature to identify what is already available in terms of GP profession satisfaction factors. |
| Theoretical framework/theories/ underpinnings | |
| Description of intervention | |
| Rationale for paper/review | Examining what makes GPs happy in their profession, may be important in increasing the GP workforce in the future. |
| What creates meaning that sustains GPs engagement with, and involvement in, work? | Autonomy was noted as one of the most relevant topics found in the literature - GPs wanted to have the freedom to choose their work method, their payment method, and have flexible working hours. Furthermore, GPs wated vocational choice and the ability to choose the kind of doctor they want to be. |
| What solutions/recommendations are proposed? | Within general practice, aim to maximise the 'satisfaction' factors described by this review. |
| From which stakeholders' perspective? (General | GPs |

| | |
|---|---|
| practitioners, patients, other HCPs) | |
| How is work-related joy constructed? (Diversity of ways individuals may think about joy) | Teaching and academic responsibilities. |
| What prevents/removes work-related joy? | Working long hours. However, what constitutes 'long hours' was noted to be subjective, therefore flexibility in work hours was also noted to be an important influence on GP satisfaction. |
| Organisational characteristics (small, large) | |
| Managing ill-defined illnesses and chronic complex multimorbidity | One of the most important aspects of satisfaction was intellectual stimulation and the ability to use one's abilities. Intellectual stimulation was often provided by dealing with a wide variety of patients and problems. |
| Human connection with patients | GPs enjoy playing an important role in their local community.<br><br>An efficient doctor-patient relationship was also considered an important factor in GP job satisfaction |
| Human connection with peers | Human connection with peers was emphasised as important. |
| Human connection across other organisations (e.g., secondary care) | Human connection across organisations was emphasised as important. |
| Learning systems (e.g., competence, capability, mastery, socially situated) | Successful medical management of patients and the subsequent feelings of being competent were associated with a higher degree of satisfaction. In a Scottish qualitative study, GPs derived the greatest satisfaction from consultations where GPs perceived that they personally had contributed to a successful outcome for the patient. |
| Additional useful information | Payment systems had differing effects on GP satisfaction: rural physicians preferred the fee-for-service method, while urban physicians supported blended or fixed payment schemes.<br><br>The influence of GPs' physical health was examined in the literature. A good health is associated with GP satisfaction. Not smoking and not being overweight is correlated with greater GP satisfaction. |
| Summary of findings | This paper describes a broad range of factors that GPs consider satisfying in relation to their job. |
| Strengths, limitations, and future research directions | One of the results of this literature review is a broad overview of the positive factors which keep GPs in their clinical practice. This comprehensive view on satisfaction factors cannot be found by reading the individual articles.<br><br>Confounding factors or interpretation bias could come from differences between social health systems and linguistic understanding. |

| | |
|---|---|
| Paper Number | 14 |
| Author | Marchand & Peckham |
| Year | 2016 |
| Title | Addressing the crisis of GP recruitment and retention: a systematic review |
| Journal | BJGP |
| Region (England, Scotland, Wales, and Northern Ireland) | This study synthesised evidence from the UK and countries with similar health systems to the UK (e.g. Canada and Australia) |
| Study methodology | Systematic review |
| Type of document | Secondary research |
| Topic or focus of paper | GP recruitment and retention |
| Research objectives and focus | To identify evidence on different approaches to retention and recruitment of GPs, such as intrinsic versus extrinsic motivational determinants. |
| Theoretical framework/theories/ underpinnings | |
| Description of intervention | |
| Rationale for paper/review | The numbers of GPs and training places in general practice are declining, and retaining GPs in their practices is an increasing problem. |
| What creates meaning that sustains GPs engagement with, and involvement in, work? | Motivating factors for medical students to pursue a career in GP include: support from colleagues, autonomy, flexibility and independence, proximity with patients, the continuity of care, health promotion, compatibility with family life and the medical breadth of the discipline.<br><br>Job satisfaction and dissatisfaction are related to three factors: job stressors (for example, workload), job characteristics and attributes (for example, job autonomy), and other conditions (for example, practice geographical location). |
| What solutions/recommendations are proposed? | Mentorship schemes, opportunities to develop portfolio careers and providing learning and development activities (eg management skills) would increase GP recruitment and retention.<br><br>An increase in satisfaction of intellectual and altruistic needs, and functional flexibility within their practice, could improve satisfaction and fulfilment, and consequently GPs' retention |
| From which stakeholders' perspective? (General practitioners, patients, other HCPs) | 1. GPs<br>2. Medical students who are interested in GP |
| How is work-related joy constructed? (Diversity of | Job autonomy, the feeling of doing an important job and social support positively affect job satisfaction.<br><br>Flexibility and part-time working have always been seen as factors that make general practice a more attractive working environment, |

| | |
|---|---|
| ways individuals may think about joy) | |
| What prevents/removes work-related joy? | Low pay is a source of dissatisfaction but, interestingly, the evidence suggests that increases in income would not compensate for other sources of job dissatisfaction, like workload.<br><br>Increased administration and bureaucracy.<br><br>Adverse publicity from the media, changes imposed from local primary care organisations, and insufficient resources within the practice.<br><br>Job satisfaction is also influenced by expectations about future events.45 If doctors perceive that their workload will not reduce, and that demands will always increase, it is likely that they will feel more overwhelmed and less satisfied with their job. |
| Organisational characteristics (small, large) | |
| Managing ill-defined illnesses and chronic complex multimorbidity | Increased work complexity was noted as causing job dissatisfaction.<br><br>Doing a variety of work increases job satisfaction. |
| Human connection with patients | Increased demand and expectation from patients causes job dissatisfaction.<br><br>Emotional impact of working as a GP can decrease job satisfaction. |
| Human connection with peers | Lack of support from colleagues causes job dissatisfaction.<br><br>A good practice environment positively increases job satisfaction. |
| Human connection across other organisations (e.g., secondary care) | Lack of professional recognition causes job dissatisfaction |
| Learning systems (e.g., competence, capability, mastery, socially situated) | Having insufficient time to do the job justice (which can be linked to workload) was a cause of job dissatisfaction.<br><br>Personal development increases job satisfaction. |
| Additional useful information | The literature made clear that expectations about the future affect recruitment and retention. |
| Summary of findings | Factors that seemed to influence retention were subspecialisation and portfolio careers, and job satisfaction. The most important determinants of recruitment and retention were intrinsic and idiosyncratic factors, such as recognition, rather than extrinsic factors, such as income. |
| Strengths, limitations, and future research directions | Overall, the published evidence in relation to GP recruitment and retention is limited and mostly focuses on attracting GPs to rural areas — particularly in Australia. |

| | |
|---|---|
| Paper Number | 15 |
| Author | Razai & Majeed |
| Year | 2022 |
| Title | General Practice in England The Current Crisis, Opportunities, and Challenges |
| Journal | Journal of Ambulatory Care Management |
| Region (England, Scotland, Wales, and Northern Ireland) | England |
| Study methodology | Commentary |
| Type of document | Secondary Research |
| Topic or focus of paper | General practice 'crisis' |
| Research objectives and focus | Describe the evolving nature of general practice and the current crisis, as well as potential solutions and opportunities going forward |
| Theoretical framework/theories/ underpinnings | |
| Description of intervention | |
| Rationale for paper/review | NHS primary care has recently come under unprecedented political and media spotlight instigated by the pressures of the current pandemic on the NHS. This coupled with collapsing morale among general practitioners (GPs), a shrinking GP workforce, inexorable demands, increasing workload, and decreasing real-terms per capita funding have caused many to sound alarm on a general practice in "crisis". |
| What creates meaning that sustains GPs engagement with, and involvement in, work? | |
| What solutions/recommendations are proposed? | There needs to an honest, transparent dialogue between the politicians, professionals, and the public about the kind of primary care that could be delivered and funded (Razai & Majeed, 2021). |
| From which stakeholders' perspective? (General practitioners, patients, other HCPs) | |
| How is work-related joy constructed? (Diversity of | Remote consultations can reduce workload. |

| | |
|---|---|
| ways individuals may think about joy) | |
| What prevents/removes work-related joy? | GP workload has continued to increase over the last decade, with higher rates of patient consultations, increasing roles and responsibilities, and a greater administrative burden, which has worsened during the COVID-19 pandemic. A consequence of such high workload is increasing emotional distress, anxiety, burnout, and depression among GPs.<br><br>Alongside this workload, the authors noted that patients expectations have increased over the last few decades. Patients were characterised as wanting more responsive GP practices with better coordination and more relationship-based care with continuity with the same GP. Furthermore, many patients want appointment times that suit them with wider and more convenient range of services. Furthermore, patients increasingly act as informed consumers of health care and no longer regard a doctor's opinion as "sacrosanct and a new dialogue is developing between healthcare consumers and providers". These increased expectations have been caused by the work of patient advocacy groups, single-issue campaigns, and the idealism of regulatory bodies such as the GMC. Pressure from politicians who have made ambitious promises without a matching resolve to provide funding, and resources have also contributed to this problem. |
| Organisational characteristics (small, large) | |
| Managing ill-defined illnesses and chronic complex multimorbidity | |
| Human connection with patients | Despite GPs have one of the highest public satisfaction ratings of any public service in the UK, individual cases of patients feeling let down are often magnified by the media without context or more awareness of the causes and fair minded solutions. It is crucial that GPs feel valued, respected, and given the mental space to work. Feeling denigrated and called out for systemic failures will likely worsen the mental health of a workforce on whom the NHS depends (Gerada, 2021a, 2021b). |
| Human connection with peers | |
| Human connection across other organisations (e.g., secondary care) | |
| Learning systems (e.g., competence, capability, mastery, socially situated) | |
| Additional useful information | The authors noted that the changing socioeconomic landscape of Britain, funding pressures and poor workforce planning are three general factors that are ultimately responsible for the crisis in GP. |
| Summary of findings | This paper discusses the wider crisis in general practice; therefore, some points pertain to GPs relationship with work. |
| Strengths, limitations, and future research directions | |

| | |
|---|---|
| Paper Number | 16 |
| Author | Reeve et al |
| Year | 2022 |
| Title | The CATALYST programme: can a knowledge work intervention help tackle the UK GP workforce crisis? |
| Journal | Australian Journal of Primary Health (conference abstract) |
| Region (England, Scotland, Wales, and Northern Ireland) | England |
| Study methodology | 80 GPs underwent complex intervention (CATALYST), followed by Normalisation Process Theory informed evaluation of the delivery and impact of programme using observation, interview, focus groups and survey data. |
| Type of document | Primary research |
| Topic or focus of paper | Expert generalism |
| Research objectives and focus | To understand if and how CATALYST impacts on professional confidence and skills for everyday practice for new-to-practice GPs in northeast England. |
| Theoretical framework/theories/ underpinnings | |
| Description of intervention | CATALYST is an 11-month programme that aims to address professional barriers to expert generalist care; namely, a lack of confidence and skills in the knowledge work of beyond-disease-protocol and whole-person medicine. GPs attend 2 days a month, and CATALYST delivers 6 modules on knowledge work alongside a 5-month advanced QI project. |
| Rationale for paper/review | UK healthcare recognises 'the future doctor is a generalist'. Yet research highlights both professional and service/system barriers to the practice of expert generalist care. |
| What creates meaning that sustains GPs engagement with, and involvement in, work? | CATALYST increased understanding of and confidence in the knowledge work of generalist practice; through the practice and outputs of clinician scholarship - formation of new communities of practice, supporting generation of mindlines. Participants described new perception of value of their role (as consultant in primary care medicine); confidence and skills in whole-person-medicine; and extended network to learn together. |
| What solutions/recommendations are proposed? | |
| From which stakeholders' perspective? (General practitioners, patients, other HCPs) | |
| How is work-related joy constructed? (Diversity of | |

| | |
|---|---|
| ways individuals may think about joy) | |
| What prevents/removes work-related joy? | |
| Organisational characteristics (small, large) | |
| Managing ill-defined illnesses and chronic complex multimorbidity | |
| Human connection with patients | |
| Human connection with peers | |
| Human connection across other organisations (e.g., secondary care) | |
| Learning systems (e.g., competence, capability, mastery, socially situated) | |
| Additional useful information | |
| Summary of findings | This paper suggests that the CATALYST programme increased GPs' confidence in – and their value placed on – generalist practice. In general, this suggests that teaching GPs about the value of their role as an expert generalist may contribute to increased perceptions of meaning. |
| Strengths, limitations, and future research directions | Scale and test if the CATALYST approach can enhance professional practice elsewhere. |

| | |
|---|---|
| Paper Number | 17 |
| Author | Riley et al |
| Year | 2018 |
| Title | What are the sources of stress and distress for general practitioners working in England? A qualitative study |
| Journal | BMJ Open |

| | |
|---|---|
| Region (England, Scotland, Wales, and Northern Ireland) | England |
| Study methodology | Interviews with GPs followed by thematic analysis of resultant dataset. |
| Type of document | Primary research |
| Topic or focus of paper | Stress in GP |
| Research objectives and focus | Report the sources of stress and distress experienced by GPs |
| Theoretical framework/theories/ underpinnings | |
| Description of intervention | |
| Rationale for paper/review | Compared with the general population, general practitioners experience high levels of workforce stress. This can lead to burnout and early retirement, compounding pre-existing workforce shortages. There is also a clear link between the experiences and well-being of NHS staff, including doctors, and the experiences of patients and the quality of care they receive. Therefore, it is important to understand the sources of the increased levels of stress and distress, in order to address them more effectively. |
| What creates meaning that sustains GPs engagement with, and involvement in, work? | Three main themes relating to stress in general practice:<br>1. Emotion work—the work invested and required in managing and responding to the psychosocial component of GPs' work and dealing with abusive or confrontational patients.<br>2. Practice culture—practice dynamics and collegial conflict, bullying, isolation and lack of support.<br>3. Work role and demands—fear of making mistakes, complaints and inquests, revalidation, appraisal, inspections and financial worries. |
| What solutions/recommendations are proposed? | Providing a safe space for GPs to process the emotional and clinical content of their work and the potential stressors related to the organisational culture (eg, bullying in the workplace) and relationships at work (eg, collegial conflict) is imperative.<br><br>The ability to respond effectively to the emotional demands and anxiety often expressed by patients in the consultation without feeling overwhelmed needs to be addressed in GP training as well as in ongoing support and supervision. Crucially, collegial support is a protective factor for good mental health—support from mentors, supervisors and colleagues is associated with resilience and reduced sickness.<br><br>Tackling the culture of invulnerability early on in medical training is also key. Schwartz Centre Rounds, for instance, are currently being piloted in medical schools with early evidence supporting their value. First 5, buddy systems and access to regular supervision or mentorship can also offer support and a reflective space—talking, sharing and having one's feelings normalised, understood and validated are critical in maintaining good mental health. |

| | |
|---|---|
| From which stakeholders' perspective? (General practitioners, patients, other HCPs) | GPs |
| How is work-related joy constructed? (Diversity of ways individuals may think about joy) | |
| What prevents/removes work-related joy? | GPs frequently reported stress due to workload and long hours – paper highlighted how these have already been extensively covered in the literature.<br><br>A significant source of palpable anxiety and distress for many GP participants was the fear of making a clinical mistake and the resulting repercussions. Many participants were concerned that the escalating workload directly increased the risk of making a mistake.<br><br>Stress was caused by managing complaints from patients.<br><br>The additional workload and stress associated with undertaking revalidation, appraisal and care quality commission (CQC) inspections was a dominant theme among the participants, some of whom found the process to be punitive, unhelpful and at times overwhelming.<br><br>Stress was caused by the financial burdens associated with being a GP partner – which has been exacerbated recently by practices being expected to do more with fewer resources. |
| Organisational characteristics (small, large) | |
| Managing ill-defined illnesses and chronic complex multimorbidity | |
| Human connection with patients | Emotion work was caused by managing patients with difficult problems and lots of sadness. Emotion work was also caused by managing confrontational, physically and verbally abusive patients.<br><br>However, many GPs also indicated that positive patient contact was the most enjoyable and meaningful part of their job: "My job satisfaction was patients coming in and going out having got their needs attended to and happy". |
| Human connection with peers | The stress and anxiety associated with the responsibilities related to staffing issues, practice dynamics and collegial conflict was a key source of stress among GP participants.<br><br>Many GPs experienced bullying by colleagues and/or partners. |

| | |
|---|---|
| | Feeling isolated and unsupported in their work was a dominant theme among the majority of the participants, and contributed to participants' existing stress and distress. The loneliness and isolation was also compounded by the burden of responsibility and increasing workload, which left GPs with little time to connect with their colleagues.<br><br>A lack of support and acknowledgement among other colleagues or GP partners for those who were experiencing mental illness was also observed and experienced by many participants.<br><br>In contrast, some GP participants clearly benefited from being part of a team and working alongside colleagues who have invested in cultivating a supportive culture over time. |
| Human connection across other organisations (e.g., secondary care) | |
| Learning systems (e.g., competence, capability, mastery, socially situated) | |
| Additional useful information | |
| Summary of findings | This study highlights that the sources of stress and distress cannot solely be attributed to increases in workload and occupational stress linked to the work role demands of being a GP. Sources of stress and distress are also linked to unhealthy practice cultures, which may be characterised by bullying or the absence of collegial support and the opportunity to discuss the emotional and clinical demands of GPs' work. Such a workplace climate can cause and add to stress and distress, and leave GPs feeling isolated. Promoting compassionate and supportive work cultures is therefore imperative in addressing the dynamic interplay between the personal, professional and organisational sources of stress and distress for GPs. |
| Strengths, limitations, and future research directions | Few studies employing qualitative methods, using in-depth interviews, have been used to examine this topic.<br><br>While recruiting individuals with experience of mental illness and burnout, we also included the perspectives of GPs who had no disclosed history of mental illness, which enables the data to be more widely generalisable to the GP population.<br><br>Participants were self-selecting, which may be perceived as a limitation; however, the sampling of participants ensured that the sample was varied in terms of age, gender, number of years as practising GPs, level of seniority/employment status in the practice, and geographical location. |

| | |
|---|---|
| Paper Number | 18 |
| Author | Riley et al |
| Year | 2018 |

| | |
|---|---|
| Title | 'Treading water but drowning slowly': what are GPs' experiences of living and working with mental illness and distress in England? A qualitative study |
| Journal | BMJ Open |
| Region (England, Scotland, Wales, and Northern Ireland) | England |
| Study methodology | Interviews with GPs followed by thematic analysis of resultant dataset. |
| Type of document | Primary research |
| Topic or focus of paper | GPs with mental illnesses |
| Research objectives and focus | Provide an in-depth account of general practitioners' (GPs) experiences of living and working with mental illness and distress |
| Theoretical framework/theories/ underpinnings | |
| Description of intervention | |
| Rationale for paper/review | To date, few studies have explored the experiences and accounts of GPs living and working with enduring levels of stress, mental ill health and psychological distress using in-depth qualitative methods. |
| What creates meaning that sustains GPs engagement with, and involvement in, work? | |
| What solutions/recommendations are proposed? | 1. Greater recognition of the components and cumulative effect of the occupational stressors for doctors, such as increasing workload and the clinical and emotional demands of the job.<br>2. A culture change within medicine, with a shift to more supportive and compassionate work cultures.<br>3. The destigmatising of mental ill health and the recognition of GPs' experiences and emotional responses as symptomatic of occupational stress as opposed to any perceived failure to cope in the individual. |
| From which stakeholders' perspective? (General practitioners, patients, other HCPs) | GPs |
| How is work-related joy constructed? (Diversity of | |

| | |
|---|---|
| ways individuals may think about joy) | |
| What prevents/removes work-related joy? | Intense workload and burnout were noted as the major contributor to much of the mental ill health amongst participants. |
| Organisational characteristics (small, large) | |
| Managing ill-defined illnesses and chronic complex multimorbidity | |
| Human connection with patients | |
| Human connection with peers | Perhaps symptomatic of the stigma attached to mental illness and the culture of invulnerability within medicine, some GP participants described how they felt ashamed, embarrassed, humiliated and a sense of having failed, due to their perceived lack of resilience and inability to cope.<br><br>The participant below highlights how the denial and intolerance of mental ill health among some colleagues can enter into an individual's internal dialogue, thus compounding existing feelings of shame. |
| Human connection across other organisations (e.g., secondary care) | |
| Learning systems (e.g., competence, capability, mastery, socially situated) | |
| Additional useful information | |
| Summary of findings | This study focusses on characterising the mental ill health experienced by some GPs (e.g. low self-esteem, anger, suicidal thoughts). With a few exceptions (see above) it does not analyse the parts of a GPs working life that contribute to this mental ill health. |
| Strengths, limitations, and future research directions | Participants were self-selecting, which may be perceived as a limitation; however, the recruitment of participants ensured that the sample was varied in terms of age, gender, number of years as practising GPs, level of seniority/employment status in the practice and geographical location.<br><br>In recruiting individuals with experience of mental illness and burn-out, we also included the perspectives of GPs who had no disclosed history of mental illness which enables the data to be more widely transferable to the UK GP population. |

| | |
|---|---|
| Paper Number | 19 |

| | |
|---|---|
| Author | Sansom et al |
| Year | 2016 |
| Title | Understanding quit decisions in primary care: a qualitative study of older GPs |
| Journal | BMJ Open |
| Region (England, Scotland, Wales, and Northern Ireland) | England |
| Study methodology | Interviews with senior GPs who had either retired from patient care or intended to retire within 5 years. Then, thematic analysis of the responses. |
| Type of document | Primary research |
| Topic or focus of paper | GP early retirement |
| Research objectives and focus | Explore and understand an individual GPs early retirement intentions, and the factors influencing these decisions. |
| Theoretical framework/theories/ underpinnings | |
| Description of intervention | |
| Rationale for paper/review | Workforce crisis in GP: the National Institute of Health Research—Health Services Delivery strand—has recently commissioned a call for research into understanding and ameliorating these workforce issues. |
| What creates meaning that sustains GPs engagement with, and involvement in, work? | By and large, the GPs reported that seeing patients was the part of the job they enjoyed the most, however, there were several factors that made the job 'on the cusp of undoable at times' (GP5, Partner, female, age 55 years), and these factors were all cited as influencing decisions about when to retire/quit direct patient care. Key issues identified by the GPs were: workload; change; NHS structures and systems; concerns about the future of general practice; partnership issues; politics; the impact of ageing and ill health; and reduced morale and emotional resilience. |
| What solutions/recommendations are proposed? | Giving GPs dedicated time for administrative tasks; sharing workload with other practices; and introducing a GP buddy system to help manage incoming queries if one GP was on annual leave or off sick.<br><br>GPs also outlined a wish for more emotional, practical and organisational support. These 'hypothetical solutions' included: creating greater peer support among GPs; provision of mandatory study days to help GPs prioritise their own health and well-being; making it easier for GPs to reduce their hours and/or become a salaried or locum GP; improving access to information to help GPs stay up-to-date; giving GPs one email account solely for patient matters, and another account for all non-patient matters; improving the integration of health and social care to make it easier for GPs to access community beds for their patients (as an example); and either taking the NHS out of Government control or making it a crossparty issue with commitment to longer term planning to ensure greater consistency and less party-specific changes. |

| | |
|---|---|
| From which stakeholders' perspective? (General practitioners, patients, other HCPs) | Senior (nearing retirement) GPs. |
| How is work-related joy constructed? (Diversity of ways individuals may think about joy) | Locum work is viewed positively because it offers less involvement with the administrative side of the practice work and it allows more time for GPs to spend on professional (and indeed personal) areas of interest. |
| What prevents/removes work-related joy? | Some GPs take early retirement out of fear of being viewed as an 'old doctor' who is 'out of touch'.<br><br>Demand for care therefore workload significantly increased in recent years – contributing factors include unrealistic patient expectations; an increase in the number of patient contacts without a corresponding increase in the number of GPs; and additional workload from secondary care. This problem is compounded by external factors including the need to meet targets, to stay up-to-date with new guidelines, and to prepare for Care Quality Commission (CQC) visits.<br><br>Several of the GPs felt they were chasing targets all the time and 'government interference' and micromanagement were seen by one GP as a lack of trust by the Government. CQC checks were viewed as a burden of additional paperwork, bureaucracy and stress by many of the GPs.<br><br>GPs felt stressed and demoralised by Government proposals and changes (eg, 7-day access to primary care).<br><br>Many GPs were concerned about the future of general practice and their jobs, which was a source of stress.<br><br>GPs identified ageing as having an impact on their confidence and ability and, consequently, their perceived capacity to continue working in direct patient care. The length and pace of the working day were felt to be exhausting, stamina decreased with age, and this was compounded for some female GPs who reported experiencing sleep disturbance which they attributed to the menopause. Deteriorating cognitive (eg, memory) and physical (eg, eyesight) functioning led to fear, anxiety, loss of confidence and stress. GPs described the fear of 'unconscious incompetence'.<br><br>Morale was described as being very low among GPs, and key reasons for this included 'media bashing', medicine/doctors being used for political gain, being worn down by change. |
| Organisational characteristics (small, large) | GPs described a 'drip, drip' of organisational changes over the years, which they have little control over and cannot see the value of: 'you never feel that you've quite got your feet on solid ground and it's always constantly changing'. |
| Managing ill-defined illnesses and chronic complex multimorbidity | Negative perception that GP workload had become more complex in recent years. |
| Human connection with patients | Morale was affected by not being valued by politicians and consequently patients. |

| | |
|---|---|
| Human connection with peers | GPs felt there was a cultural division between younger and older GPs, with the younger GPs having a different style of work. This led to a situation that, when older GP colleagues retired, GPs felt out of place.<br><br>GPs expressed mixed views about the appraisal and revalidation system. Some found appraisals helpful while others felt they were an additional burden and ineffective.<br><br>Good working relationships with other partners in the practice were cited as helping to mitigate some of the difficulties around managing workload, change, etc (the converse of this was also noted to be true when good relationships had not developed). |
| Human connection across other organisations (e.g., secondary care) | GPs felt that referral pathways had become more complex and time-consuming. They received more referrals and were required to do more follow-ups due to unrealistic expectations of patients, and hospital doctors lacking resources. There was also a complaint from one GP that GPs are working in 'a vacuum of information' owing to delays in communication from hospitals. |
| Learning systems (e.g., competence, capability, mastery, socially situated) | |
| Additional useful information | GPs taking early retirement can have a knock-on effect on the practice – eg increasing the workload for other GPs and/or requiring the appointment of additional staff. |
| Summary of findings | This paper examines GPs' reasons for early retirement. Many of these reasons relate to GPs' relationship with work. |
| Strengths, limitations, and future research directions | There was limited success in recruiting GPs who were already retired, and also those currently working as salaried GPs, however, the final sample size was felt to adequately answer the research question.<br><br>Interviews were conducted around the time of the 2015 general election: this may have influenced GPs' perceptions of some of the reported issues. |

| | |
|---|---|
| Paper Number | 20 |
| Author | Sansom et al |
| Year | 2017 |
| Title | Why do GPs leave direct patient care and what might help to retain them? A qualitative study of GPs in South West England |
| Journal | BMJ Open |
| Region (England, Scotland, Wales, and Northern Ireland) | England |
| Study methodology | Interviews with GPs intending to leave patient care for a variety of reasons, followed by thematic analysis of responses. |
| Type of document | Primary research |
| Topic or focus of paper | GP workforce retention |

| | |
|---|---|
| Research objectives and focus | The aim of this study was to gain insight into the lived experiences of GPs in today's National Health Service (NHS), with the intention of identifying factors which, if addressed, might facilitate retention in direct patient care. |
| Theoretical framework/theories/ underpinnings | |
| Description of intervention | |
| Rationale for paper/review | Recent survey of all GPs in South West England revealed that 37% of GPs reported a high likelihood of quitting direct patient care within 5 years. Few qualitative studies provide a contemporary view of decision-making in this regard, or include GPs of more than one age group or decision outcome. There is also a lack of evidence to support the view that recent policies and strategies developed to address GP workforce retention (eg, as outlined in the General Practice Forward View) will help to reverse the GP shortage. |
| What creates meaning that sustains GPs engagement with, and involvement in, work? | Identity and professionalism were important to GPs. However, many felt compromised in their ability to practise in their preferred ways (eg, in the length of time they were able to spend with patients) and this impacted negatively on morale, professionalism and identity. This was partly due to: unrealistic demands; concerns about complaints; targets and guidelines; complexity of cases and lack of time to address them; lack of continuity and loss of professional autonomy. GPs felt compromised in striking a balance between delivering high-quality care and doing this within the constraints and burdens they experienced. |
| What solutions/recommendations are proposed? | Future policies and strategies aimed at retaining GPs in direct patient care should clarify the role and expectations of general practice and align with GPs' perception of their own roles and identity; demonstrate to GPs that they are valued and listened to in planning delivery of the UK healthcare; target GPs' concerns regarding fear and risk, seeking to reduce these to manageable levels and give GPs viable options to support them to remain in direct patient care |
| From which stakeholders' perspective? (General practitioners, patients, other HCPs) | |
| How is work-related joy constructed? (Diversity of ways individuals may think about joy) | |
| What prevents/removes work-related joy? | Fear and anxiety were experienced regarding different aspects of risk that had to be managed within the GP role – in some cases this has led to GPs taking sick leave. Risk was related to 'unmanageable' workloads, the complexity of cases, the large number of decisions that had to be made and the impact of cumulative decision-making throughout the day. Patients were perceived to have easy routes to complain about their GP/practice, but there was felt to be little support or recourse for GPs in actively managing those concerns. Where GPs had experienced complaints or being sued, they described the process as being drawn out and stressful.<br><br>Buying into a practice was associated with the risk of personal debt and therefore increased stress.<br><br>Older GPs who had invested were experiencing stress and anxiety due to concerns about changes to practice mortgages, the threat of having to make staff redundant or practice closure and responsibilities arising from joint civil liability for a practice. |

| | |
|---|---|
| Organisational characteristics (small, large) | |
| Managing ill-defined illnesses and chronic complex multimorbidity | |
| Human connection with patients | Feeling valued on account of their work was important to GPs. However, they reported often feeling undervalued by the general public, the NHS, the media and the government. |
| Human connection with peers | A stigma exists around GPs accessing help—particularly mental health support—and GPs expressed concerns about confidentiality. |
| Human connection across other organisations (e.g., secondary care) | Tensions were identified between the primary and secondary care interface: general practice tended to ‘pick up’ and manage aspects of care that were felt to be the remit of other services and there was a clear sense that the ‘buck stops’ with general practice. |
| Learning systems (e.g., competence, capability, mastery, socially situated) | |
| Additional useful information | There was frustration over a perceived lack of ability to determine the future of general practice and the lack of a unified model that could be implemented.<br><br>Many of the factors noted above, relating to why GPs are quitting patient care, would be manageable in isolation. However, GPs noted that the factors compounded and reinforced each other to create an unsustainable situation.<br><br>GPs’ resilience has been eroded in recent years, which was linked to a perceived lack of control over their future. |
| Summary of findings | Reasons for leaving direct patient care were complex and based on a range of job-related and individual factors. Three key themes underpinned the interviewed GPs’ thinking and rationale: issues relating to their personal and professional identity and the perceived value of general practice-based care within the healthcare system; concerns regarding fear and risk, for example, in respect of medical litigation and managing administrative challenges within the context of increasingly complex care pathways and environments; and issues around choice and volition in respect of personal social, financial, domestic and professional considerations. |
| Strengths, limitations, and future research directions | A maximum variation general practitioner (GP) sample was achieved through use of the workforce census survey returns.<br><br>GPs were self-selecting and represented mainly those indicating the intention to leave (or who had already left) direct patient care. |

| | |
|---|---|
| Paper Number | 21 |
| Author | Schofield et al |
| Year | 2020 |
| Title | Exploring how paramedics are deployed in general practice and the perceived benefits and drawbacks: a mixedmethods scoping study |

| | |
|---|---|
| Journal | BJGP Open |
| Region (England, Scotland, Wales, and Northern Ireland) | England |
| Study methodology | Three phases. First, a literature review and survey into the deployment of paramedics in general practice. Second, meetings with key informants in CCGs to ascertain preexisting theories about the intended outcomes of deploying paramedics in general practice. Third, interviews with stakeholders involved with the deployment of paramedics in general practice, followed by thematic analysis of the responses. |
| Type of document | Both primary and secondary research |
| Topic or focus of paper | Paramedics in general practice |
| Research objectives and focus | To understand how paramedics are deployed in general practice, and to investigate the theories and drivers that underpin this service development. |
| Theoretical framework/theories/ underpinnings | |
| Description of intervention | |
| Rationale for paper/review | General practice in the UK faces continuing challenges to balance a workforce shortage against rising demand. The NHS England GP Forward View proposes development of the multidisciplinary, integrated primary care workforce to support frontline service delivery, including the employment of paramedics. However, very little is known about the safety, clinical effectiveness, or cost-effectiveness of paramedics working in general practice. Research is needed to understand the potential benefits and drawbacks of this model of workforce organisation. |
| What creates meaning that sustains GPs engagement with, and involvement in, work? | |
| What solutions/recommendations are proposed? | |
| From which stakeholders' perspective? (General practitioners, patients, other HCPs) | Stakeholders involved with paramedic deployment in general practice – including, but not limited to, GPs. |
| How is work-related joy constructed? (Diversity of ways individuals may think about joy) | In general, paramedics are seen as a positive addition to the general practice team; reducing demand on GP time and ensuring more patients are seen sooner. This reduces stress level amongst GPs. |

| | |
|---|---|
| What prevents/removes work-related joy? | Support and supervision of paramedics can increase workload for GPs. |
| Organisational characteristics (small, large) | |
| Managing ill-defined illnesses and chronic complex multimorbidity | The tendency for paramedics to see low-complexity cases can result in GPs primarily seeing patients with multimorbidity, chronic illness, and, often, social complexities. This has the potential to impact on the job satisfaction and mental wellbeing of GPs: 'But if the paramedics see all of these "easy stuff" ... you're left with only the really complicated stuff ... I found that emotionally very hard work and draining.' |
| Human connection with patients | |
| Human connection with peers | |
| Human connection across other organisations (e.g., secondary care) | |
| Learning systems (e.g., competence, capability, mastery, socially situated) | |
| Additional useful information | |
| Summary of findings | There is very little evidence on the safety and cost-effectiveness of paramedics working in general practice and significant variation in the ways that paramedics are deployed, particularly in terms of the patients seen and conditions treated. Nonetheless, there is a largely positive view of this development and a perceived reduction in GP workload. However, some concerns centre on the time needed from GPs to train and supervise paramedic staff. |
| Strengths, limitations, and future research directions | These findings provide new information about the current provision of paramedics in general practice and what the intended benefits and unintended consequences of this workforce organisation might be. However, there are a number of limitations; the survey used a convenience sample, and it is not possible to determine the response rate or whether the views expressed were representative of a wider group. The interviews were conducted within two neighbouring CCGs, and this may limit the generalisability of the findings. |

| | |
|---|---|
| Paper Number | 22 |
| Author | Shen et al |
| Year | 2022 |
| Title | The global prevalence of burnout among general practitioners: a systematic review and meta-analysis |
| Journal | Family practice |

| | |
|---|---|
| Region (England, Scotland, Wales, and Northern Ireland) | International – this systematic review included 16 studies, one of which was from the UK. |
| Study methodology | First, a systematic review was conducted to identify cross-sectional studies investigating the prevalence of burnout amongst GPs. Then, a meta-analysis was conducted using a random-effects model. Random-effects model attempted to generalise findings beyond the included studies by assuming that the selected studies are random samples from a larger population. |
| Type of document | Secondary research |
| Topic or focus of paper | Burnout amongst GPs |
| Research objectives and focus | Systematically measure the global prevalence of burnout amongst GPs |
| Theoretical framework/theories/ underpinnings | |
| Description of intervention | |
| Rationale for paper/review | Burnout among general practitioners (GPs) has attracted the attention of more and more researchers. An adequate understanding the prevalence and related factors of burnout to prevent and reduce burnout is necessary. This study systematically measured the global prevalence of burnout among GPs. |
| What creates meaning that sustains GPs engagement with, and involvement in, work? | |
| What solutions/recommendations are proposed? | |
| From which stakeholders' perspective? (General practitioners, patients, other HCPs) | GPs |
| How is work-related joy constructed? (Diversity of ways individuals may think about joy) | |
| What prevents/removes work-related joy? | |
| Organisational characteristics (small, large) | |

| | |
|---|---|
| Managing ill-defined illnesses and chronic complex multimorbidity | |
| Human connection with patients | |
| Human connection with peers | |
| Human connection across other organisations (e.g., secondary care) | |
| Learning systems (e.g., competence, capability, mastery, socially situated) | |
| Additional useful information | The pooled burnout rate showed 37%, 28%, and 26% of general GPs suffer from high emotional exhaustion (EE), high depersonalization (DP), and low personal accomplishment (PA), respectively |
| Summary of findings | Globally, there is a high prevalence of burnout within general practice. Health managers need to tailor their strategies to retain this community. Targeted initiatives are needed to provide adequate GPs' well-being and maintain primary health care. |
| Strengths, limitations, and future research directions | There were still some studies that used other tools to measure burnout that were not included in the analysis.<br><br>Another major limitation is that different studies have reported large variations in prevalence rates. This variation may be attributed to the type of population included in the study. Finally, most of the studies included in this study are from Europe, and there is a lack of studies from the other regions, especially in America. This would leads to regional differences in the studies, which causes the conclusions of this study lack of generality. |

| | |
|---|---|
| Paper Number | 23 |
| Author | Sidhu et al |
| Year | 2022 |
| Title | Vertical integration of GP practices with acute hospitals in England and Wales: rapid evaluation |
| Journal | Health and Social Care Delivery Research |
| Region (England, Scotland, Wales, and Northern Ireland) | England and Wales |
| Study methodology | Rapid qualitative cross-comparative case study evaluation at three sites in England (n = 2) and Wales (n = 1), which comprised three work packages: (1) a rapid review of literature, telephone scoping interviews and a stakeholder workshop; (2) interviews with stakeholders across |

| | |
|---|---|
| | case study sites, alongside observations of strategic meetings and analysis of key documents from the sites; and (3) development of a theory of change for each site and for vertical integration overall. |
| Type of document | Primary research |
| Topic or focus of paper | Vertical integration between primary care and secondary care |
| Research objectives and focus | Our aim was to (1) understand the rationale for, and early impact of, vertical integration in the NHS in England and Wales and (2) develop a theory of change for vertical integration. |
| Theoretical framework/theories/ underpinnings | |
| Description of intervention | |
| Rationale for paper/review | |
| What creates meaning that sustains GPs engagement with, and involvement in, work? | |
| What solutions/recommendations are proposed? | |
| From which stakeholders' perspective? (General practitioners, patients, other HCPs) | NHS staff, policy experts and stakeholders involved in integrated care |
| How is work-related joy constructed? (Diversity of ways individuals may think about joy) | Partner GPs becoming employees of a trust in a process of vertical integration, and therefore having a regular salary, was perceived positively by GPs.<br><br>Integration was perceived to be creating more career opportunities for GPs within hospitals<br><br>Vertical integration can have a very positive impact, especially for GP services that were previously struggling: "I know that morale in our team is good. People feel listened to. We've had, the practice I've had, hasn't been [vertical integration company] for a huge amount of time, but going from being a practice that was really struggling to getting very positive patient feedback now is very nice. And to have people happy where they're working, feel that they can actually make a difference, can develop things themselves, I think that's a sign of success." |
| What prevents/removes work-related joy? | GPs were concerned that vertical integration of services would deprive them of autonomy.<br><br>The initial shock associated with integration and subsequent loss of independence can cause stress and push GPs out of the workforce.<br><br>Vertical integration was perceived as stressful by some GPs: "Other GPs in the patch, obviously, became very nervous. Where a big organisation starts to throw their weight around primary care, you know, it's kind of worrying for their business model". |

| | |
|---|---|
| | According to our interviewees, it was clear by that time, and has become clearer in the years since, that the traditional (partner) model of general practice was unattractive to some GPs. This is due to the responsibilities and financial risks of running a small business. |
| Organisational characteristics (small, large) | NHS staff and policy experts recognised benefits of vertical integration, especially for younger GPs, as younger GPs could be given greater opportunities to acquire specialist training across the primary and secondary interface and would be more likely to have a set/defined workload and greater back office support (e.g. legal, indemnity, procurement), creating more time for clinical contribution.<br><br>Although vertical integration did often provide greater back office support, GPs also noted that it can create bureaucracy and reporting requirements: "You know, with GPs you're quite slick and quick and everything happens quickly, you know. We're running out of rubber gloves, that's absolutely fine: 'Can you order 50 boxes of rubber gloves?' 'They'll be here tomorrow [and] we get 20% discount.' Couldn't do that with the trust, so they've got a procurement process." |
| Managing ill-defined illnesses and chronic complex multimorbidity | |
| Human connection with patients | |
| Human connection with peers | Vertical integration led to more salaried GPs and locums in the place of partners. This was perceived as leading to a lack of cohesiveness within GP teams: "I mean, none of the GPs here have been a partner before but they behave like partners, if you like. So, like, with holiday, we all say: 'Unwritten rule: no one has time off at Christmas, because it's not fair for one or the other'. They do what they like and they book time off, which upsets everybody else, which upsets the girls, because I'm saying, 'You can't have time off'. They're, like, 'Well, they're salaried like I'm salaried, so why do they get it and not me?'" |
| Human connection across other organisations (e.g., secondary care) | Integration across primary and secondary care was often associated with tensions between primary care and acute trust colleagues. |
| Learning systems (e.g., competence, capability, mastery, socially situated) | |
| Additional useful information | |
| Summary of findings | Although not the main focus of this paper, this study sets out a number of ways in which GPs' relationship with work will change as the NHS moves towards a model of integrated care. These changes are viewed in an overall positive light by GPs and other stakeholders involve in vertical integration: with vertical integration removing many financial responsibilities from GPs, lessening administrative work and creating career opportunities (both within general practice and the wider trust). However, some GPs find this new loss of autonomy stressful, with integration increasing administration and reporting in some cases whilst introducing tensions between primary and secondary care colleagues. |

| | |
|---|---|
| Strengths, limitations, and future research directions | Across all three case study sites, the study team was unable to complete the desired number of non-participant observations. The pace of data collection during early interviews and documentary analysis varied. Owing to the circumstances of the COVID-19 pandemic during project write-up, the team was unable to undertake site-specific workshops during data analysis and an overall workshop with policy experts. |

| | |
|---|---|
| Paper Number | 24 |
| Author | Simon et al |
| Year | 2018 |
| Title | What is the root cause of the GP workforce crisis? |
| Journal | BJGP |
| Region (England, Scotland, Wales, and Northern Ireland) | UK |
| Study methodology | Commentary |
| Type of document | Secondary research |
| Topic or focus of paper | GP workforce crisis |
| Research objectives and focus | Suggest a possible root cause for the workforce crisis in GP |
| Theoretical framework/theories/ underpinnings | |
| Description of intervention | |
| Rationale for paper/review | |
| What creates meaning that sustains GPs engagement with, and involvement in, work? | GPs in training ranked their top 5 priorities for an ‘ideal GP job’: Appropriate remuneration, supportive team, flexible working hours and patterns, happy working environment, manageable workload.<br><br>Newly qualified GPs ranked their top 5 priorities for an ‘ideal GP job’: supportive team, flexible working hours and patterns, appropriate remuneration, manageable workload, fair contract including maternity cover.<br><br>This paper highlights that problems with retention (and indeed retention) are not unique to general practice. They are occurring in other medical specialties and other careers (eg teaching). The authors suggest that a root cause may be the different career aspirations held by Generation Y (or ‘Millennials’). Generation Y have been shown to want more versatility and flexibility in the workplace. They move for increased pay and innovative working environments, and strive for a strong work–life balance. They are more detached from institutions and networked with friends; they use social media and embrace technology. Generation Y often want to have parallel or ‘portfolio’ careers. |

| | |
|---|---|
| What solutions/recommendations are proposed? | We suggest that GPs show similar traits to their Generation Y peers and the workforce challenges within general practice are not unique to our profession. If we want to improve recruitment and retention in general practice, we need to understand how an increasingly salaried and locum workforce can be accommodated. We need to account for generational traits, do more to create flexible career paths to enable 'career hopping' between different roles, and carefully evaluate the effectiveness of existing initiatives to support greater GP career diversity/flexibility. Urgent research is also needed to establish if the variety offered by portfolio careers is protective or if managing a 'work–work–life balance' ultimately becomes too demanding. |
| From which stakeholders' perspective? (General practitioners, patients, other HCPs) | GPs |
| How is work-related joy constructed? (Diversity of ways individuals may think about joy) | Upon surveying GP trainees in Dorset, portfolio working was very common: 42% currently had more than one role, including private online GP work, GP with special interest posts, fellowships, and educational roles; 50% anticipated having more than one role in 5 years. |
| What prevents/removes work-related joy? | GPs in the UK have reported finding their job stressful for a number of well rehearsed reasons, including management of patients with increasingly complex needs and expectations, increasing numbers of face to-face consultations, work that is more intense, and longer working days.<br><br>Few GP trainees would recommend general practice to junior doctors, and reasons included workload, job-related stress, government policies, long hours, bureaucracy and administrative burden, high patient demand, and threat of litigation.<br><br>The King's Fund found that GPs in training intended to work as locums (44%) or salaried doctors (48%) after qualification, with only 3% intending to work as partners. |
| Organisational characteristics (small, large) | |
| Managing ill-defined illnesses and chronic complex multimorbidity | |
| Human connection with patients | |
| Human connection with peers | |
| Human connection across other organisations (e.g., secondary care) | |

| | |
|---|---|
| Learning systems (e.g., competence, capability, mastery, socially situated) | |
| Additional useful information | GPs in the UK report finding their job more stressful than GPs in other countries. |
| Summary of findings | In addition to highlighting some well-rehearsed factors which influence the relationship between GPs and their work, this paper proposes that the root cause of the GP workforce crisis (in particular, the finding that very few trainee GPs aspire to partnership) is the different career intentions and aspirations of 'Generation Y'. |
| Strengths, limitations, and future research directions | |

| | |
|---|---|
| Paper Number | 25 |
| Author | Adelekan-Kamara et al |
| Year | 2023 |
| Title | Factors underpinning an improved menopausal experience in the workplace for doctors: a UK-based qualitative study |
| Journal | BMJ Open |
| Region (England, Scotland, Wales, and Northern Ireland) | UK |
| Study methodology | |
| Type of document | Primary research |
| Topic or focus of paper | Menopausal experience of doctors |
| Research objectives and focus | Determine the factors underpinning an improved menopausal experience for UK doctors. |
| Theoretical framework/theories/ underpinnings | Interviews with menopausal doctors (n=21) and nonmenopausal (n=20) doctors, then subsequent thematic analysis of responses. |
| Description of intervention | |
| Rationale for paper/review | An improved menopausal experience (IME) in the workplace has been associated with increased job satisfaction, increased economic participation and reduced absenteeism. Currently, existing literature fails to explore menopausal doctors' experiences, and none factors in non-menopausal colleagues' perspectives. |
| What creates meaning that sustains GPs engagement with, and involvement in, work? | Four overarching themes underpinning an IME were identified: menopausal knowledge and awareness, openness to discussion, organisational culture, and supported personal autonomy. The levels of knowledge held by menopausal participants themselves, their colleagues and their superiors were identified as crucial in determining menopausal experiences. Likewise, the ability to openly discuss menopause was also identified as an important factor. The NHS culture, gender dynamics and an adopted superhero mentality—where |

| | |
|---|---|
| | doctors feel compelled to prioritise work over personal well-being—further impacted under the umbrella of Organisational culture. Personal autonomy at work was considered important in improving menopausal experiences at work for doctors. The superhero mentality, lack of organisational support and a lack of open discussion were identified as novel themes not found in current literature, particularly in the healthcare context. |
| What solutions/recommendations are proposed? | The potential benefits of an IME for doctors in the NHS are considerable. NHS leaders can address these challenges by using pre-existing training materials and resources for their employees if menopausal doctors are to feel supported and retained. |
| From which stakeholders' perspective? (General practitioners, patients, other HCPs) | Doctors including GPs. |
| How is work-related joy constructed? (Diversity of ways individuals may think about joy) | |
| What prevents/removes work-related joy? | |
| Organisational characteristics (small, large) | |
| Managing ill-defined illnesses and chronic complex multimorbidity | |
| Human connection with patients | |
| Human connection with peers | |
| Human connection across other organisations (e.g., secondary care) | |
| Learning systems (e.g., competence, capability, mastery, socially situated) | |
| Additional useful information | |
| Summary of findings | This study highlights that doctors' factors underpinning an IME in the workplace are comparable to other sectors. |
| Strengths, limitations, and future research directions | Voluntary participants are likely to have an underlying interest in the experience of menopause in the workplace, therefore, some perspectives, such as disinterest, may not have been captured. |

| | |
|---|---|
| Paper Number | 26 |
| Author | Cheshire et al |
| Year | 2017 |
| Title | GPs' perceptions of resilience training: a qualitative study |
| Journal | BJGP |
| Region (England, Scotland, Wales, and Northern Ireland) | England |
| Study methodology | Interviews with GPs followed by subsequent analysis of responses. |
| Type of document | Primary research |
| Topic or focus of paper | Resilience amongst GPs |
| Research objectives and focus | To explore GPs' perspectives on the content, context, and acceptability of resilience training programmes in general practice, in order to build more effective GP resilience programmes. |
| Theoretical framework/theories/ underpinnings | |
| Description of intervention | |
| Rationale for paper/review | GPs are reporting increasing levels of burnout, stress, and job dissatisfaction, and there is a looming GP shortage. Promoting resilience is a key strategy for enhancing the sustainability of the healthcare workforce and improving patient care. |
| What creates meaning that sustains GPs engagement with, and involvement in, work? | Participants perceived resilience training to be potentially valuable in ameliorating workplace stresses – however this can only go so far, given the extent of work pressures GPs currently face.<br><br>Key components of resilience training included: mindfulness/meditation or yoga/breathing exercises, lifestyle advice (including exercise and dietary advice), general stress management advice (including relaxation/self-care techniques), and better understanding of the physiology of the stress response |
| What solutions/recommendations are proposed? | None relevant to this review. |
| From which stakeholders' perspective? (General practitioners, patients, other HCPs) | GPs |

| | |
|---|---|
| How is work-related joy constructed? (Diversity of ways individuals may think about joy) | |
| What prevents/removes work-related joy? | |
| Organisational characteristics (small, large) | Some participants highlighted that it would be beneficial to include practical approaches to reducing stressors in the GP workplace in resilience training. Training here could include advice to address some of the challenges faced in a practice and/or at local level, including improving communication and support among work colleagues, and simple, practical approaches to improving workplace efficiency. |
| Managing ill-defined illnesses and chronic complex multimorbidity | |
| Human connection with patients | |
| Human connection with peers | Others highlighted that being able to share experiences with peers was particularly therapeutic, engendering support and problem solving among colleagues. There were suggestions, however, that skilled facilitation could ensure that forums did not become a detrimental 'moan fest'.<br><br>Feeling part of a team within your own practice and offering mutual support was seen as bolstering GP resilience<br><br>Some highlighted that the inclusion of mentoring from more senior colleagues as part of resilience training or a 'buddy' could be beneficial to the long-term resilience of GPs. |
| Human connection across other organisations (e.g., secondary care) | |
| Learning systems (e.g., competence, capability, mastery, socially situated) | |
| Additional useful information | |
| Summary of findings | This paper reports the positive perceptions of resilience training amongst GPs, and discusses a number of ways in which it can be useful in ameliorating GP stress relating to their work. It also discusses modes of delivery of resilience training, which is less relevant to our review. |
| Strengths, limitations, and future research directions | Sampling methods may have attracted GPs with an interest in resilience and time to participate. Interviews and focus groups provided a helpful combination of data collection methods. |

| | |
|---|---|
| Paper Number | 27 |
| Author | Royal College of General Practitioners |

| | |
|---|---|
| Year | Not reported |
| Title | General practice in the post Covid world |
| Journal | Report published by the RCGP |
| Region (England, Scotland, Wales, and Northern Ireland) | UK |
| Study methodology | Commentary |
| Type of document | Secondary research |
| Topic or focus of paper | Future of general practice |
| Research objectives and focus | Discuss the three key features of the COVID-19 response which the RCGP believes have the most potential for transforming general practice so that it is equipped to meet the health challenges of the 21st century. |
| Theoretical framework/theories/ underpinnings | |
| Description of intervention | |
| Rationale for paper/review | The Covid-19 pandemic has the potential to change general practice radically and permanently. Across the UK, practice teams have had to reconfigure their operating model overnight. The pace of change has demonstrated the remarkable adaptability of general practice. Now, as we start to transition towards a 'new normal', we have an historic opportunity to embed or accelerate those changes which have been for the better, while discarding those which are detrimental to the profession, to the NHS and to the communities we serve. |
| What creates meaning that sustains GPs engagement with, and involvement in, work? | The ability to deliver consultations from home or to schedule 'call back' appointments around their other commitments has enabled GPs to work more flexibly. Remote working could, therefore, make partnership or salaried roles more attractive to GPs, particularly those with caring responsibilities.<br><br>Clinicians talked about the positive impacts of COVID-19 on their well-being, including the ability to contribute to the Covid-19 response. Initially reduced workload pressures and fewer administrative burdens, coupled with more flexibility in appointment length, enabled many to devote more time for patients without having to work unsustainable hours. Similarly, many GPs, particularly earlier career GPs and students, said that being able to work from home and in more flexible ways were a significant improvement. The NHS Practitioner Health Service was also valued. |
| What solutions/recommendations are proposed? | |
| From which stakeholders' perspective? (General practitioners, patients, other HCPs) | GPs |

| | |
|---|---|
| How is work-related joy constructed? (Diversity of ways individuals may think about joy) | "Members consistently tell us that the [appraisal and revalidation] requirements are too burdensome." |
| What prevents/removes work-related joy? | The Covid-19 pandemic placed a significant amount of pressure on staff in general practice, both professionally and personally, due to a range of factors including the demands of implementing new ways of working, cancellation of planned closures, and staff not taking leave. Many staff were also working in more high-risk settings such as 'hot sites', caring for patients with Covid-19 symptoms, and remote working appears to have increased feelings of isolation among some clinicians. |
| Organisational characteristics (small, large) | The ability to deliver consultations from home or to schedule 'call back' appointments around their other commitments has enabled GPs to work more flexibly. Remote working could, therefore, make partnership or salaried roles more attractive to GPs, particularly those with caring responsibilities. |
| Managing ill-defined illnesses and chronic complex multimorbidity | |
| Human connection with patients | However, we should not lose sight of the centrality of the relationship between the GP and patient which lies at the heart of general practice. Remote consultations have clear benefits – such as convenience and ease of access – for some patients and certain types of consultations, e.g. minor health problems, but they cannot be a substitute for faceto-face consultations, which are better suited to fostering trust and empathy between doctors and patients. |
| Human connection with peers | One of the challenges faced by GPs and practice staff during lockdown is the potential impact of remote working on team cohesion. This challenge has forced practices to adopt new ways for teams to talk and bond and to ensure staff do not feel isolated, for example through morning 'virtual huddles'. In some practices, this has actually helped to improve team working.<br><br>The introduction of virtual 'tea breaks', or WhatsApp groups, have provided space for staff discussions. Some GPs felt that the widespread wearing of scrubs, which was necessary for infection control, helped to create a sense of being part of a team. |
| Human connection across other organisations (e.g., secondary care) | |
| Learning systems (e.g., competence, capability, mastery, socially situated) | |
| Additional useful information | |
| Summary of findings | This paper summarises how the RCGP believes general practice should change in the wake of the COVID-19 pandemic. Many of these changes centre around increasing efficiency and achieving better outcomes for patients, however some relate to the relationship between GPs and their work, and I have extracted these above. |
| Strengths, limitations, and future research directions | |

| | |
|---|---|
| Paper Number | 28 |
| Author | Checkland & Spooner |
| Year | 2015 |
| Title | The future of primary care? Reflections on the Primary Care Workforce Commission report |
| Journal | BJGP |
| Region (England, Scotland, Wales, and Northern Ireland) | UK |
| Study methodology | Editorial |
| Type of document | Secondary research |
| Topic or focus of paper | |
| Research objectives and focus | Reflect on the Primary Care Workforce Commission report |
| Theoretical framework/theories/ underpinnings | |
| Description of intervention | |
| Rationale for paper/review | Health Education England commissioned Professor Martin Roland to chair a Primary Care Workforce Commission to gather evidence and make recommendations not only to address the current crisis, but also to project forward and attempt to 'future proof' the profession. |
| What creates meaning that sustains GPs engagement with, and involvement in, work? | The report implies the development of a GP consultant role, with senior GPs taking a more coordinating and consultative approach, overseeing a team of healthcare providers including nurses, pharmacists, and PAs. Although some GPs may embrace this new role, others may regret the loss of their place as the first port of call where skill, insight, and expertise can be vital; most GPs can recall examples of apparently simple and routine consultations that uncovered something more serious. |
| What solutions/recommendations are proposed? | |
| From which stakeholders' perspective? (General practitioners, patients, other HCPs) | GPs |
| How is work-related joy constructed? (Diversity of | |

| | |
|---|---|
| ways individuals may think about joy) | |
| What prevents/removes work-related joy? | |
| Organisational characteristics (small, large) | |
| Managing ill-defined illnesses and chronic complex multimorbidity | |
| Human connection with patients | |
| Human connection with peers | |
| Human connection across other organisations (e.g., secondary care) | |
| Learning systems (e.g., competence, capability, mastery, socially situated) | |
| Additional useful information | |
| Summary of findings | All but one (see above) of the reflections in this paper do not relate to the relationship between GPs and their work. |
| Strengths, limitations, and future research directions | |

| | |
|---|---|
| Paper Number | 29 |
| Author | John Glasspool |
| Year | 2015 |
| Title | GP recruitment and retention |
| Journal | BJGP |
| Region (England, Scotland, Wales, and Northern Ireland) | UK |
| Study methodology | Letter |

| | |
|---|---|
| Type of document | Secondary research |
| Topic or focus of paper | |
| Research objectives and focus | |
| Theoretical framework/theories/ underpinnings | |
| Description of intervention | |
| Rationale for paper/review | |
| What creates meaning that sustains GPs engagement with, and involvement in, work? | “My wife and I both took early retirement from general practice, in our late 50s. The strain of trying to be ‘all things to all people’ was just getting too much. We were worried that we would start to make mistakes by just having too many balls to juggle.”<br><br>Scrapping seniority payments is a disincentive for older GPs to stay on. |
| What solutions/recommendations are proposed? | |
| From which stakeholders’ perspective? (General practitioners, patients, other HCPs) | |
| How is work-related joy constructed? (Diversity of ways individuals may think about joy) | |
| What prevents/removes work-related joy? | “Introducing a contract with open-ended workload for no extra money [is] unacceptable”<br><br>The CQC is a burden.<br><br>“The complaints culture takes up too much time and effort for doctors: the GMC is an organisation out of control, which we now find, according to its own report, has been involved in the deaths of at least 28 doctors under investigation.” |
| Organisational characteristics (small, large) | |
| Managing ill-defined illnesses and chronic complex multimorbidity | |

| | |
|---|---|
| Human connection with patients | |
| Human connection with peers | |
| Human connection across other organisations (e.g., secondary care) | |
| Learning systems (e.g., competence, capability, mastery, socially situated) | |
| Additional useful information | |
| Summary of findings | This short letter highlights a few major factors which are eroding joy and meaning in general p |
| Strengths, limitations, and future research directions | |

| | |
|---|---|
| Paper Number | 30 |
| Author | Hall et al |
| Year | 2017 |
| Title | Strategies to improve general practitioner well-being: findings from a focus group study |
| Journal | Family Practice |
| Region (England, Scotland, Wales, and Northern Ireland) | England |
| Study methodology | Focus groups with GPs. Discussions were centered on the workplace factors that they perceived to influence their well-being, along with strategies that they use either personally, or as a practice, to try and prevent burnout. Furthermore, strategies that could feasibly be implemented by individuals and practices to improve well-being, as well as changes that are needed by groups or organizations that are external to their practice (e.g., the government) to improve the working conditions, were explored. Subsequent thematic analysis of transcripts. |
| Type of document | Primary research |
| Topic or focus of paper | GP wellbeing |
| Research objectives and focus | To explore strategies that could improve GP well-being and reduce or prevent burnout, based on GP perceptions of the workplace factors that affect their levels of well-being and burnout. |

| | |
|---|---|
| Theoretical framework/theories/ underpinnings | |
| Description of intervention | |
| Rationale for paper/review | There are no qualitative studies that specifically investigated the best ways to improve well-being and prevent burnout in primary care physicians. |
| What creates meaning that sustains GPs engagement with, and involvement in, work? | Internal influencers of well-being comprised Team support, Variety (within their roles, practices, or patients), Control (over their work environment and/or timetable) and an Intense and unmanageable workload.<br><br>External influencers of well-being were discussed in negative terms. These consisted of Increases in pressures and workload (particuarly administrative work), Increases in patients' expectations and complaints, the Negative portrayal of general practice (in the media, by patients and the government) and a Lack of support (from the public, patients, the government and the media). |
| What solutions/recommendations are proposed? | Scheduling a coffee and/or lunch break into the working day was viewed as a feasible strategy that would be very beneficial to their well-being. Participants voiced that even if GPs only briefly left their office to make a cup of tea and take it back to their office, this very short respite and chance of interaction could be enough to make a big change to their well-being.<br><br>Having social support within the practice, peer-to-peer and from both medics and nonmedics outside of their practice was found to be useful for preventing burnout. To improve support at the practice level, buddying and mentoring systems were suggested, along with regular meetings to 'check in' with how team members are doing. A suggestion for improving support from patients was to communicate the state of the surgery with them and ask for their patience and support.<br><br>Participants discussed the need to make time for exercise to support physical and psychological well-being.<br><br>Strategies that participants used to deal with the emotional toll of patient contact included being emotionally guarded or setting boundaries and isolating themselves. The latter approach, however, was acknowledged to be unhealthy and did indeed worsen one participant's ability to cope. Maintaining awareness of the risk of burnout was voiced as a useful strategy that some participants used, and this could be implemented internally within practices and externally at the training stage.<br><br>Control over how much, where, and when they worked was seen as a positive strategy that some GPs (mostly locums) used to prevent burnout.<br><br>Participants voiced the need for support from the government, their patients, the healthcare organization as a whole and the wider public and press through a reduction in negative media portrayal. Additionally, a need for support from other services was discussed, for example, from social services, to ensure that patients only presented with medical problems and could go to social services in other instances.<br><br>Participants stated the need for a reduction in the tasks that decrease their time that should be spent on direct patient care, such as administrative work, quality assessment exercises and additional work pushed onto them from secondary care. |

| | |
|---|---|
| | Increasing resources for primary care was seen as an ideal solution that would help us to improve all the previous factors mentioned, such as reducing pressures and enabling time for breaks. |
| From which stakeholders' perspective? (General practitioners, patients, other HCPs) | GPs |
| How is work-related joy constructed? (Diversity of ways individuals may think about joy) | |
| What prevents/removes work-related joy? | |
| Organisational characteristics (small, large) | |
| Managing ill-defined illnesses and chronic complex multimorbidity | |
| Human connection with patients | |
| Human connection with peers | |
| Human connection across other organisations (e.g., secondary care) | |
| Learning systems (e.g., competence, capability, mastery, socially situated) | |
| Additional useful information | |
| Summary of findings | Based on the contributors to burnout and workplace well-being that the participants identified, the following feasible strategies were suggested: compulsory daily coffee breaks, increasing self- and organizational awareness of the risks of burnout and mentoring or buddy systems. System-level organizational changes were voiced as vital, however, to improve the wellbeing of all primary care physicians. Increasing resources seemed to be the ideal solution, to allow for more administrative staff and GPs. |
| Strengths, limitations, and future research directions | The primary strength of this study is the practical and feasible strategies that could be implemented within practices immediately to improve workplace well-being in the interim before organizational change can be implemented. |

| | |
|---|---|
| Paper Number | 31 |
| Author | Hamilton-West et al |
| Year | 2018 |
| Title | Does a modified mindfulness-based cognitive therapy (MBCT) course have the potential to reduce stress and burnout in NHS GPs? Feasibility study |
| Journal | Primary Health Care Research & Development |
| Region (England, Scotland, Wales, and Northern Ireland) | England |
| Study methodology | Participants recorded their perceived stress levels before and after undertaking a modified mindfulness-based cognitive therapy (MBCT) course. Qualitative data was also obtained on participants' experience of the course. |
| Type of document | Primary research |
| Topic or focus of paper | Mindfulness amongst GPs |
| Research objectives and focus | The purpose of this study was to explore, for the first time, whether a modified MBCT course has the potential to reduce stress and burnout in NHS GPs. |
| Theoretical framework/theories/ underpinnings | |
| Description of intervention | The course was a modified version of the MBCT course approved by National Institute for Health and Care Excellence (NICE) for prevention of depressive relapse. The eight weekly 2-h sessions adhered closely to the MBCT protocol, with one main difference: the homework (completed between sessions) was less time-consuming to facilitate compliance among the GP cohort and used the Mindfulness: A Practical Guide to Finding Peace in a Frantic World book (Williams and Penman, 2011) as the homework text (i.e. designed for a non-clinical cohort, in contrast to the NICE approved MBCT course, which is designed for patients with recurrent depression. The course was offered to GPs in two Clinical Commissioning Group areas in South East England as an eight-week evening course in May and June 2016. |
| Rationale for paper/review | There is a crisis of low morale among NHS GPs, with most describing their workload as 'unmanageable'. MBCT has been demonstrated to improve stress and burnout in other populations, but has not yet been evaluated in a cohort of NHS GPs. |
| What creates meaning that sustains GPs engagement with, and involvement in, work? | Following the MBCT course, burnout scores of emotional exhaustion and depersonalisation were significantly lower. GPs noted that the course taught them to 'notice how [their] mind works', helping them deal with stress. |
| What solutions/recommendations are proposed? | Mindfulness training may be useful in reducing stress amongst GPs - although further research is needed before conclusions are definitively drawn. |

| | |
|---|---|
| From which stakeholders' perspective? (General practitioners, patients, other HCPs) | GPs |
| How is work-related joy constructed? (Diversity of ways individuals may think about joy) | Learning how to prioritise own needs at work was noted as a key benefit of the MBCT course.<br><br>Following the course, one GP noted that they were taking shorts breaks throughout the day, instead of working from 8am to 6:30pm with no break.<br><br>GPs noted increased enjoyment of job following MBCT course and a feeling that they have more time: eg "even the routine things are more enjoyable". |
| What prevents/removes work-related joy? | The course taught GPs techniques to manage stress as work: eg "Learning to do three-minute meditation when stressed at work". |
| Organisational characteristics (small, large) | |
| Managing ill-defined illnesses and chronic complex multimorbidity | |
| Human connection with patients | |
| Human connection with peers | Meeting others and sharing experiences was noted as a key benefit of the MBCT course. |
| Human connection across other organisations (e.g., secondary care) | |
| Learning systems (e.g., competence, capability, mastery, socially situated) | |
| Additional useful information | |
| Summary of findings | This study provides support for benefits of mindfulness in an NHS GP cohort. |
| Strengths, limitations, and future research directions | It is important to acknowledge that this is a small preliminary evaluation and a larger scale randomized-controlled trial will be needed before definitive conclusions can be drawn.<br><br>A further potential limitation of the current study is that GPs were recruited from only two Clinical Commissioning Group areas in South East England. |

| | |
|---|---|
| Paper Number | 32 |
| Author | Hayes et al |
| Year | 2020 |
| Title | Working lives of GPs in Scotland and England: cross-sectional analysis of national surveys |
| Journal | BMJ Open |
| Region (England, Scotland, Wales, and Northern Ireland) | England & Scotland105 |
| Study methodology | Comparison of cross-sectional analysis of survey responses of GPs in England and Scotland in 2017 and 2018, respectively, using linear regression to adjust the differences for gender, age, ethnicity, urbanicity and deprivation. |
| Type of document | Primary research |
| Topic or focus of paper | Reformed Scottish GP contract |
| Research objectives and focus | The UK faces major problems in retaining general practitioners (GPs). Scotland introduced a new GP contract in April 2018, intended to better support GPs. This study compares the career intentions and working lives of GPs in Scotland with GPs in England, shortly after the new Scotland contract was introduced. |
| Theoretical framework/theories/ underpinnings | |
| Description of intervention | |
| Rationale for paper/review | [See ‘research objectives and focus’] |
| What creates meaning that sustains GPs engagement with, and involvement in, work? | GPs in Scotland reported higher levels of satisfaction (measured across the domains of: colleagues, variety, responsibility, physical, use abilities, freedom, overall how you feel about job, pay, recognition, hours of work), lower levels of pressure (measured across the domains of: increasing workloads, patient demands, insufficient time, problem patients, paperwork, finding a locum, earlier hospital discharges, long working hours, adverse media publicity, high expectations, patient complaints, insufficient resources, emergency calls), similar positive job attributes (measured across the domains of: variety interesting, know responsibilities, choice how I do job, involved in changes, choice what I do, consulted about changes, flexible, clear feedback, changes better patient care) and less agreement in response to negative job attributes (measured across the domains of: work intensively, work fast, unimportant tasks, not enough time). This suggests the 2018 GP contract in Scotland has engendered meaning and GP involvement with work. This moved away from the principle of competition, abolishing the QOF and establishing GP clusters to promote collaboration and integration. |
| What solutions/recommendations are proposed? | Lessons for general practice in England are offered by the new Scottish GP contract. |

| | |
|---|---|
| From which stakeholders' perspective? (General practitioners, patients, other HCPs) | GPs |
| How is work-related joy constructed? (Diversity of ways individuals may think about joy) | |
| What prevents/removes work-related joy? | |
| Organisational characteristics (small, large) | |
| Managing ill-defined illnesses and chronic complex multimorbidity | |
| Human connection with patients | |
| Human connection with peers | |
| Human connection across other organisations (e.g., secondary care) | |
| Learning systems (e.g., competence, capability, mastery, socially situated) | |
| Additional useful information | |
| Summary of findings | Following the introduction of the new contract in Scotland, GPs in Scotland reported significantly better working lives and lower intention to reduce work participation than England. |
| Strengths, limitations, and future research directions | This is the first study to compare the working lives of GPs in Scotland and England using a set of common, validated questions on satisfaction, job characteristics and job pressures using a similar data collection method.<br><br>Some of the differences reported in this study may be longstanding and unrelated to the recent, new Scottish GP contract – cannot prove causality. |

| | |
|---|---|
| Paper Number | 33 |

| | |
|---|---|
| Author | Hampson & Ruane |
| Year | 2019 |
| Title | The value of pharmacists in general practice: perspectives of general practitioners—an exploratory interview study |
| Journal | International Journal of Clinical Pharmacy |
| Region (England, Scotland, Wales, and Northern Ireland) | England |
| Study methodology | Interviews with GPs, followed by thematic analysis of responses. |
| Type of document | Primary research |
| Topic or focus of paper | Pharmacists in general practice |
| Research objectives and focus | This study aimed to explore the perspectives of GPs with experience of fully funding a pharmacist in general practice, focussing on the value that GPs place on the role of the pharmacist. |
| Theoretical framework/theories/ underpinnings | |
| Description of intervention | |
| Rationale for paper/review | NHS England has recently invested on a large scale in the development of the role of pharmacists in general practice. Despite this initial funding, the perpetuation of pharmacists' roles ultimately relies on the funding and co-operation of GPs. |
| What creates meaning that sustains GPs engagement with, and involvement in, work? | GPs described how the medication expertise of the pharmacist enabled them to delegate tasks and roles to the pharmacist which would otherwise be the remit of the GP. The majority of GPs also specifically mentioned the personal value and reassurance they felt in having easy and direct access to a medication expert within the team whom they could approach about prescribing-related decisions in specific patients. |
| What solutions/recommendations are proposed? | The views expressed in this study broadly support the movement to increase the employment of pharmacists within GP practices. |
| From which stakeholders' perspective? (General practitioners, patients, other HCPs) | GPs |
| How is work-related joy constructed? (Diversity of ways individuals may think about joy) | |

| | |
|---|---|
| What prevents/removes work-related joy? | Some GPs talked about pharmacists increasing their workload, due to time required to supervise and train. |
| Organisational characteristics (small, large) | |
| Managing ill-defined illnesses and chronic complex multimorbidity | |
| Human connection with patients | |
| Human connection with peers | GPs valued their pharmacist as an integral member of the team, someone who understood the ethos and intricacies of the practice and worked within that to achieve the best outcome possible: "So there's no formal process of being able to say that (having a pharmacist) it's positive, but I know that generally everyone is much happier for having her". |
| Human connection across other organisations (e.g., secondary care) | |
| Learning systems (e.g., competence, capability, mastery, socially situated) | |
| Additional useful information | |
| Summary of findings | This study provides an initial insight into what GPs value in the role of pharmacists in general practice. Positive findings relevant to our study include the value GPs place on the medication-related expertise which pharmacists bring to the team and the role pharmacists play within the practice team. Negative findings include the added workload associated with training and supervising pharmacists. |
| Strengths, limitations, and future research directions | |

| | |
|---|---|
| Paper Number | 34 |
| Author | Verma et al |
| Year | 2016 |
| Title | A systematic review of strategies to recruit and retain primary care doctors |
| Journal | BMC Health Services Research |
| Region (England, Scotland, Wales, and Northern Ireland) | 51 studies included: have only extracted data from the 12 studies that were conducted in the UK. |

| | |
|---|---|
| Study methodology | Systematic review. Articles assessing interventions aimed at recruiting or retaining doctors in high income countries, applicable to primary care doctors were included. |
| Type of document | Secondary research |
| Topic or focus of paper | A systematic review of strategies to recruit and retain primary care doctors |
| Research objectives and focus | The aim of this systematic review is to evaluate interventions and strategies used to recruit and retain primary care doctors internationally. |
| Theoretical framework/theories/ underpinnings | |
| Description of intervention | |
| Rationale for paper/review | There is a workforce crisis in primary care. Previous research has looked at the reasons underlying recruitment and retention problems, but little research has looked at what works to improve recruitment and retention. |
| What creates meaning that sustains GPs engagement with, and involvement in, work? | The London Initiative Zone Educational Incentives Scheme (LIZEI) aimed to improve recruitment, retention and refreshment of London GPs (1994–1999) via various schemes which focused on increasing the academic aspect of training through academic/research associate schemes. The scheme reported high levels of retention (75 % from the London Academic Training Scheme (LATS) cohort continued to practice in London), but the lack of a comparison group makes this difficult to interpret. |
| What solutions/recommendations are proposed? | |
| From which stakeholders' perspective? (General practitioners, patients, other HCPs) | |
| How is work-related joy constructed? (Diversity of ways individuals may think about joy) | |
| What prevents/removes work-related joy? | |
| Organisational characteristics (small, large) | |
| Managing ill-defined illnesses and chronic complex multimorbidity | |

| | |
|---|---|
| Human connection with patients | |
| Human connection with peers | |
| Human connection across other organisations (e.g., secondary care) | |
| Learning systems (e.g., competence, capability, mastery, socially situated) | |
| Additional useful information | |
| Summary of findings | Most studies included in this systematic review were either international or address issues relating to recruitment and retainers, therefore are not relevant to this study. Three studies were conducted in London focussing on the benefits of the LIZEI scheme on retention, and I have extracted data from these above. |
| Strengths, limitations, and future research directions | |

| | |
|---|---|
| Paper Number | 35 |
| Author | Zhou et al |
| Year | 2022 |
| Title | Investigating the links between diagnostic uncertainty, emotional exhaustion, and turnover intention in General Practitioners working in the United Kingdom |
| Journal | Frontier in Psychiatry |
| Region (England, Scotland, Wales, and Northern Ireland) | England |
| Study methodology | Questionnaire sent to GP practices, followed by regression analysis investigating associations between diagnostic uncertainty, emotional exhaustion, depersonalization, job satisfaction, and turnover intention. |
| Type of document | Primary research |
| Topic or focus of paper | Diagnostic uncertainty |
| Research objectives and focus | To examine associations between diagnostic uncertainty, emotional exhaustion, depersonalization, job satisfaction, and turnover intention in GPs. |
| Theoretical framework/theories/ underpinnings | |

| | |
|---|---|
| Description of intervention | |
| Rationale for paper/review | General Practitioners (GPs) report high levels of burnout, job dissatisfaction, and turnover intention. The complexity of presenting problems to general practice makes diagnostic uncertainty a common occurrence that has been linked to burnout. The interrelationship between diagnostic uncertainty with other factors such as burnout, job satisfaction and turnover intention have not been previously examined. |
| What creates meaning that sustains GPs engagement with, and involvement in, work? | |
| What solutions/recommendations are proposed? | |
| From which stakeholders' perspective? (General practitioners, patients, other HCPs) | GPs |
| How is work-related joy constructed? (Diversity of ways individuals may think about joy) | GPs reporting a good work life balance had decreased odds for experiencing emotional exhaustion. |
| What prevents/removes work-related joy? | Almost one-third of GPs surveyed reported experiencing >10% of diagnostic uncertainty in their day-to-day practice over the past year. GPs reporting greater diagnostic uncertainty had higher levels of emotional exhaustion, job dissatisfaction and turnover intention.<br><br>GPs working longer hours had over increased odds for higher depersonalization and increased odds of being dissatisfied with their job. |
| Organisational characteristics (small, large) | |
| Managing ill-defined illnesses and chronic complex multimorbidity | |
| Human connection with patients | |
| Human connection with peers | |
| Human connection across other organisations (e.g., secondary care) | |

| | |
|---|---|
| Learning systems (e.g., competence, capability, mastery, socially situated) | |
| Additional useful information | GPs who did not report any episodes of sickness presenteeism (or reported only one episode) over the past 12 month had decreased odds for emotional exhaustion, decreased odds for job dissatisfaction and increased odds of turnover intention compared to GPs who reported >1 episode. |
| Summary of findings | Diagnostic uncertainty was found to be associated with EE, turnover intention and job dissatisfaction. |
| Strengths, limitations, and future research directions | The results of this study suggest the need to understand not just how diagnostic uncertainty impacts individual GPs but also the wider negative implications of diagnostic uncertainty in the workforce planning and service delivery of primary care.<br><br>However, this study has several limitations. First, this was a cross-sectional study and therefore causation cannot be determined. Secondly, although the survey was intended to reach over 700 GPs, however, we only received 348 complete responses which gives an overall response rate of <50%. The response rate in this study is similar to previous studies involving GPs (50–52), however, our results may only reflect the perspectives of participating GPs rather than all GPs working in practices across the UK. In our study, we assessed EE and DP as separate dependent variables, however, another potential consideration for future research could be to consider EE and DP together as a dependent variable. |

| | |
|---|---|
| Paper Number | 36 |
| Author | Wedderburn et al |
| Year | 2013 |
| Title | The views and experiences of female GPs on professional practice and career support |
| Journal | Education for Primary Care |
| Region (England, Scotland, Wales, and Northern Ireland) | England |
| Study methodology | Female GPs were surveyed and completed a semi-structured questionnaire. Resulting dataset was analysed qualitatively and quantitatively. |
| Type of document | Primary research |
| Topic or focus of paper | 'Feminisation' of GP workforce |
| Research objectives and focus | The aim of this work was to capture female GPs' experiences of working in general practice and their expectations concerning their career and educational development. |
| Theoretical framework/theories/ underpinnings | |
| Description of intervention | |

| | |
|---|---|
| Rationale for paper/review | With female GP specialty trainees outnumbering males, there is continuing 'feminisation' of the GP workforce. The changing composition of the workforce presents challenges in terms of how best to support the long-term career and educational development needs of this sector. |
| What creates meaning that sustains GPs engagement with, and involvement in, work? | Flexible working (95% of under 34 age group, 47% of over 50 age group), cutting down number of hours worked (86% of under 34 age group, 60% and 66% of the 45-49 and over 50 age groups respectively), careers advice (48% of the under 34 age group, 12% of the over 50 age group) and developing a new interest (73% of under 34 age group, 42% of over 50 group) were all reported to encourage later retirement.<br><br>Difficulties in career progression would motivate some GPs under the age of 34 to retire. |
| What solutions/recommendations are proposed? | The needs of this sector of the workforce are clear: female GPs require support in terms of their education, training and childcare needs in order to facilitate increased representation in senior practice, healthcare and education roles in readiness for times ahead when they will be critical to service delivery. In order to retain this sector of the workforce and prevent early retirement of female GPs, such planning needs to be prospective, based upon greater job flexibility and provision, and on robust data. Consideration should be given to the promotion of roles in healthcare planning, education and developing special interests early on in a career, if not in training, in order to sow seeds for future career aspiration and to invest in its development. |
| From which stakeholders' perspective? (General practitioners, patients, other HCPs) | Female GPs. |
| How is work-related joy constructed? (Diversity of ways individuals may think about joy) | Working excessive hours – relating to this, family commitments were a motivating factor for early retirement. |
| What prevents/removes work-related joy? | Childcare was a significant problem for female GPs – with many reporting experiencing problems, or having had to take time off work, due to childcare responsibilities. Notably, these were compounded when there were significant perceived workplace changes in job role and/or new government policy changes that directly impacted on clinical practice. GPs who reported fewer challenges tended to have significant levels of social support (e.g. home-based husband, close proximity of extended family) to 'buffer' childcare demands and workplace stress. Furthermore, nearly 75% of GPs who were experiencing difficulties with childcare reported that flexible working would encourage later retirement. |
| Organisational characteristics (small, large) | Changes in medical technology would motivate some GPs over the age of 50 to retire. |
| Managing ill-defined illnesses and chronic complex multimorbidity | A perceived lack of decision making would encourage 50% of GPs under the age of 34 to retire (notably, this decreases to 7% in the over 50 age group). |
| Human connection with patients | |
| Human connection with peers | |

| | |
|---|---|
| Human connection across other organisations (e.g., secondary care) | |
| Learning systems (e.g., competence, capability, mastery, socially situated) | |
| Additional useful information | |
| Summary of findings | Working excessive hours, family commitments, childcare, changes in medical technology and a perceived lack of decision making were all motivating factors for female GPs to retire early. Flexible working, cutting down number of hours worked, careers advice and developing a new interest were all reported to encourage later retirement. The influence of these factors over retirement intentions were different in different age groups (see above). |
| Strengths, limitations, and future research directions | More research is needed to explore the stability of career aspirations over time. |

| | |
|---|---|
| Paper Number | 37 |
| Author | Laverty et al |
| Year | 2023 |
| Title | Unpromising Futures: Early Career GPs' Narrative Accounts of Meaningful Work during a Professional Workforce Crisis |
| Journal | Work, Employment and Society |
| Region (England, Scotland, Wales, and Northern Ireland) | England |
| Study methodology | Interviews and focus groups with early career GPs. Subsequent thematic analysis of dataset using Adam and Groves' approach to futures as a conceptual lens (see below). |
| Type of document | Primary research |
| Topic or focus of paper | Meaningful work for GPs |
| Research objectives and focus | 1. How do past and present experiences of meaningful work shape the imagined future of early-career GPs?<br>2. In a time of crisis, how is meaningful work understood and experienced by earlycareer GPs? |
| Theoretical framework/theories/ underpinnings | Utilises Adam and Groves' approach to futures as a conceptual lens: the future is often conceptualised in contemporary western societies as an abstract and open space that is full of limitless possibilities. This has been critically analysed by Adam and Groves (2007) who argue that this perspective of the future as empty and disconnected from the present can result in the future being exploited in a variety of ways. They describe how empty futures can be commodified for the benefit of the present, a process they call present futures, that strips away |

| | |
|---|---|
| | responsibility for the potential outcomes (Adam and Groves, 2007). A present futures approach is more likely in times of uncertainty, as the future becomes ever more distant, making it difficult to invest in long-term futures (Leccardi, 2008). An alternative orientation, one of future presents, treats the future as real and already in motion (although it may not have yet materialised) by past and present action that motivates an ethic of responsibility. This links to Yeoman's (2014) focus on having an ethic of care for meaningful work – that active engagement must require thinking about and acting on futures. If futures are disregarded and uncertain, some individuals will likely struggle to find meaningfulness in the present. Indeed, Berlant (2011) proposes that when an expected and hoped-for future becomes unachievable it forces individuals to adapt, stall, or give up in the present. |
| Description of intervention | |
| Rationale for paper/review | Over the past few decades, the intensification and reorganisation of work have led to growing precarity, insecurity and uncertainty for employees, affecting even professionals tied to traditionally model employers. Doctors, in particular, have seen substantial changes to their work: having to work harder, longer and more intensely with reductions in expected autonomy, deference and respect. |
| What creates meaning that sustains GPs engagement with, and involvement in, work? | The trainees and new GPs (mainly in their late 20s and early 30s) frequently drew on their perceptions of what general practice used to be when describing what they expected from their current and future work. General practice of the past was seen as desirable and meaningful predominantly because the work environment was perceived as contained, focused and morally significant. In terms of containment, this was characterised by the size of practices, viewing small or single-handed local practices as enabling a volume of work that could be managed within the confines of a 9-to-5 job. The new GPs believed that older GPs were familiar with all of their patients and had autonomy and freedom from many of the bureaucratic restraints they now faced. It was the belief of participants that contained and focused work in the past allowed GPs to see the outcome of their clinical work in the relationships and gratitude from patients. The new GPs, in contrast, reported that the relationship with patients has become less reciprocal. Many participants were concerned about a rising complaint culture and demands out with traditional doctoring work, as expectations of GPs have shifted. In choosing general practice as a specialism, then, this 'old' idea of GP is what the participants signed up for and expected in their future work. Instead, what they are observing is that components of general practice that they imagined would lead to meaningful work, such as autonomy and shared values, are evaporating.<br><br>For the participants, desired imagined futures came from the perceived work of past generations. At the same time, there was a recognition that the current crisis was a result of a present future approach that prioritised short-term activities rather than protecting the long-term future of the profession. The discounting of the future also featured in accounts of episodic experiences of meaninglessness that arose from situations in which the present needs of the system were prioritised over the doctors' long-term future needs and well-being. These most frequently occurred within their foundation training where, as junior doctors, they had little freedom and say over where, what and when their work was. GPs often felt they were only there to fill the rota – to bulk up the workforce to meet the current demands on services – which was often to the detriment of their training needs. The participants described feeling like the sacrifices they had had to make were not valued or cared about. Their dignity had been compromised. Yeoman (2020) refers to dignity as a core aspect of meaningfulness – that individuals have to be recognised as people with lives to lead and that to care for others they need to have the capacity to care for themselves. Instead, in these hospital settings, they were treated as a commodity for the current needs of the organisation rather than for their benefit as the next generation of doctors.<br><br>The participants generally talked about the future of general practice as uncertain, with some holding stronger anxieties that it was going to change beyond all recognition. The doctors described a strong commitment to the public values of the NHS as a source of meaningfulness that was a strong motivator for both deciding to become a doctor and to stay. The participants perceived that there was overwhelming |

| | |
|---|---|
| | uncertainty about the future. When this occurs, the future can press onto the present, making it difficult to see the long-term outcomes (Adam and Groves, 2007), and in turn, can make decision-making about the future challenging.<br><br>When discussing their plans, the participants could be split into three groups. The first group were actively seeking to leave the profession to regain autonomy, freedom and dignity (Yeoman, 2014) in their work, recognising that the sacrifice of their time and well-being was not likely to be acknowledged in general practice. The second group were interested in actively changing general practice for the future. This group saw themselves as having a different outlook on general practice and wanting to bring a more hopeful collective approach that would encourage others to join the profession. The third, and the largest, group were planning on continuing in general practice with an important caveat: to continue in general practice they had to get sovereignty over their time and prioritise their well-being. Part-time flexible work was seen as the only viable working option for all participants, men and women, as it allowed them to: (a) repay the time debt accumulated during training, by having some free time to connect with friends and family; (b) reduce the pressure of GP clinical work, in particular – occupying other days with breaks or other types of work such as teaching or working in a specialist area; and (c) protect their future self from burnout and to be able to continue long-term. It could be argued then that these doctors are treating the lived future as one in which the current workforce crisis will continue and get worse, and thus is in need of action in the present. |
| What solutions/recommendations are proposed? | |
| From which stakeholders' perspective? (General practitioners, patients, other HCPs) | Trainee GPs and new GPs |
| How is work-related joy constructed? (Diversity of ways individuals may think about joy) | |
| What prevents/removes work-related joy? | |
| Organisational characteristics (small, large) | |
| Managing ill-defined illnesses and chronic complex multimorbidity | |
| Human connection with patients | |

| | |
|---|---|
| Human connection with peers | Low morale within practices can erode meaning and enjoyment. |
| Human connection across other organisations (e.g., secondary care) | |
| Learning systems (e.g., competence, capability, mastery, socially situated) | |
| Additional useful information | The participants recognised that the current crisis in general practice was the result of multiple past failings in the system: decades of political disinvestment, policy and contract changes, population variations and patient expectations. However, some went further by explicitly linking how their present and future work has been made precarious by previous generations of GPs. Several participants felt that previous generations had taken their share of benefits in their present but did not consider the longer-term future of the profession. |
| Summary of findings | The findings show that there is a mismatch between imagined futures and lived futures for early career GPs. In the past, general practice comprised morally significant activities, a work environment that allowed autonomy, freedom and dignity, in a profession that was respected. These are components of Yeoman's (2014) definitions of meaningful work. The lived future these GPs were faced with, however, was a work environment characterised by accelerating work pressures and intensity, uncontained and changing work environments, alongside a loss of public respect. Upon realising their imagined future is not possible, participants responded in different ways: (a) by leaving and seeking that future elsewhere, (b) working to transform the present to achieve that future, or (c) making do by altering their present future and hoping things turn out better in the end. Present future solutions (e.g. reducing working days) then, make more sense to individuals than investing in future present solutions (such as taking up partnerships) given the uncertainty. These solutions reflect what Chaudhry et al. (2021) have referred to as depersonalisation and detachment strategies in medical work during crises. |
| Strengths, limitations, and future research directions | It is, currently, unclear whether individual strategies to increase meaning – like flexible working - can properly support meaningful work and further research exploring these temporal clashes could prove fruitful.<br><br>Further research might look to examine if and how new imagined futures can be made and how meaningfulness may be recovered by different orientations towards the present and future. |

| | |
|---|---|
| Paper Number | 38 |
| Author | David Zigmond |
| Year | 2022 |
| Title | Manufacturing doctors is one thing; sustaining working communities is quite another |
| Journal | BGJP |
| Region (England, Scotland, Wales, and Northern Ireland) | N/A |

| | |
|---|---|
| Study methodology | Opinion piece on website |
| Type of document | Grey literature |
| Topic or focus of paper | Human connections between GPs, patients and peers. |
| Research objectives and focus | |
| Theoretical framework/theories/ underpinnings | |
| Description of intervention | |
| Rationale for paper/review | |
| What creates meaning that sustains GPs engagement with, and involvement in, work? | In the past, GPs found deep, personal work satisfactions in the long-term, stable relationships they developed with patients, peers and communities. Recent reforms and changes to the job of a GP have eroded this: "known faces, voices, and stories; shared jests, joys, and sorrows; nuanced consolations and affections … all become rare as the people we know and the communities we organically grew, and which nourished us, disappear…". |
| What solutions/recommendations are proposed? | We need the restoration of the 'pastoral' healthcare of old. |
| From which stakeholders' perspective? (General practitioners, patients, other HCPs) | One retired GP and his discussions with colleagues throughout his career. |
| How is work-related joy constructed? (Diversity of ways individuals may think about joy) | |
| What prevents/removes work-related joy? | |
| Organisational characteristics (small, large) | |
| Managing ill-defined illnesses and chronic complex multimorbidity | |
| Human connection with patients | GPs of the past found great meaning in the long-term relationships they developed with patients: "With those individuals he shared much beyond their current sufferings and reliefs — the substratum of their dreads and hopes; what brought them aspiration, inspiration, love, or |

| | |
|---|---|
| | hatred; what triggered grief or laughter; their hauntings and dreams; what they (wanted to?) get up for with daylight; and what they feared when darkness came.” |
| Human connection with peers | GPs of the past also found meaning and happiness forming close relationships with peers: “Yet it was also true in another sense: our practices usually functioned much like happier families — with high levels of personal knowledge, understanding, trust, loyalty, and intelligent flexibility. I felt that this family doctor surgery where I now worked provided me with a secondary family, a home from home.” |
| Human connection across other organisations (e.g., secondary care) | In the past, GPs got to know kindred professionals – eg hospital specialists and their secretaries; locality district, health visitor, and psychiatric nurses; probation and social workers; housing officers and charity conveners; local pharmacists. In such ‘extended families’ we often felt we were helping one another shoulder the burdens of what can be very difficult work. |
| Learning systems (e.g., competence, capability, mastery, socially situated) | |
| Additional useful information | |
| Summary of findings | |
| Strengths, limitations, and future research directions | |

| | |
|---|---|
| Paper Number | 39 |
| Author | Kathy Oxtoby |
| Year | 2021 |
| Title | GP life now: problems to tackle, opportunities to grasp, and changes to absorb |
| Journal | BMJ |
| Region (England, Scotland, Wales, and Northern Ireland) | N/A |
| Study methodology | Commentary |
| Type of document | Secondary research |
| Topic or focus of paper | The changing role of GPs |
| Research objectives and focus | After a “torrid year” for general practitioners, Kathy Oxtoby examines how their role is developing, what these changes mean to them, and what being a GP could soon look like |
| Theoretical framework/theories/ underpinnings | |
| Description of intervention | |

| | |
|---|---|
| Rationale for paper/review | The role and working life of GPs is rapidly changing. |
| What creates meaning that sustains GPs engagement with, and involvement in, work? | Preeti Shukla, a GP at the Richmond Hill Practice in Colne, Lancashire, says that GPs are leaving “in droves” because the role, particularly regarding covid-19, is exhausting. “Less bureaucracy, less ‘media bashing’ of doctors, and having less of a regulatory burden from bodies such as the Care Quality Commission and NHS England” could help, she says. But, if working conditions don’t improve, “people will continue to leave.”<br><br>Some GPs reduce hours for positive reasons, like wanting a portfolio career or spending time with families. However, others do it because they are finding the job undoable. |
| What solutions/recommendations are proposed? | The RCGP’s priorities relevant to our paper include:<br>1. Longer appointments<br>2. Higher trust in the profession, with less arguous regulation<br>3. Reduced workload by greatly expanding the GP and practice team workforce<br>4. Promotion of relationship-based care<br><br>The BMA’s GP priorities relevant to our paper include:<br>1. Trusting GPs to do their job<br>2. Enabling flexibility of work<br>3. Reducing bureaucracy and regulation<br>4. Supporting practices to prioritise workload |
| From which stakeholders’ perspective? (General practitioners, patients, other HCPs) | GPs |
| How is work-related joy constructed? (Diversity of ways individuals may think about joy) | Vautrey notes that “doctors didn’t train to work in call centres. They want to have that regular face-to-face interaction with their patients, which is one of the joys of general practice.” |
| What prevents/removes work-related joy? | 47% of GPs surveyed by the BMA reported increased levels of stress, anxiety and emotional distress since the pandemic began. |

| | |
|---|---|
| Organisational characteristics (small, large) | |
| Managing ill-defined illnesses and chronic complex multimorbidity | |
| Human connection with patients | The relationship that develops between GPs and their patients is the “reason why most GPs have chosen to do this job”. |
| Human connection with peers | |
| Human connection across other organisations (e.g., secondary care) | |
| Learning systems (e.g., competence, capability, mastery, socially situated) | |
| Additional useful information | |
| Summary of findings | This paper highlights a number of recent changes to the role of a GP, including increased part time working, increased salaried GPs, telemedicine, rise in self-care, PCNs and GP-patient relationships. Of particular relevance to our review, the GP-patient relationship (including seeing patients face-to-face) is an important source of meaning. Furthermore, bureaucracy, the pandemic, ‘media bashing’, workload and regulatory burden have all eroded joy and meaning. |
| Strengths, limitations, and future research directions | |

| | |
|---|---|
| Paper Number | 40 |
| Author | Park et al |
| Year | 2019 |
| Title | Reorganisation of general practice: be careful what you wish for |
| Journal | BJGP |
| Region (England, Scotland, Wales, and Northern Ireland) | N/A |
| Study methodology | Commentary |
| Type of document | Secondary research |

| | |
|---|---|
| Topic or focus of paper | General practice reorganisation |
| Research objectives and focus | The naming of a general practice 'crisis' has led to a number of initiatives attempting to address this. Policies have promoted IT, mixing of skills, role expansion, and delegation of tasks to allied healthcare professionals. While not undermining or devaluing the value many of these initiatives provide for patients, this article seeks to explore some of the unintended consequences and underpinning assumptions of these changes. |
| Theoretical framework/theories/ underpinnings | |
| Description of intervention | |
| Rationale for paper/review | Recent publication of the NHS Long Term Plan and the renewal of the GP contract |
| What creates meaning that sustains GPs engagement with, and involvement in, work? | Traditionally, general practice has challenged the 'disease-based' organisation of healthcare systems – encounters between GPs and patients have typically been oriented towards experiences and problems. Many recent policy changes within general practice have changed the nature of this work, shifting the GP role from one of an expert generalist to one who engages with a more selective patient group with a potentially higher incidence of pathology and complexity. This might erode job satisfaction and meaning for GPs who value high-quality comprehensive care with a continuous relationship. |
| What solutions/recommendations are proposed? | |
| From which stakeholders' perspective? (General practitioners, patients, other HCPs) | |
| How is work-related joy constructed? (Diversity of ways individuals may think about joy) | |
| What prevents/removes work-related joy? | |
| Organisational characteristics (small, large) | |
| Managing ill-defined illnesses and chronic complex multimorbidity | |
| Human connection with patients | |

| | |
|---|---|
| Human connection with peers | |
| Human connection across other organisations (e.g., secondary care) | |
| Learning systems (e.g., competence, capability, mastery, socially situated) | |
| Additional useful information | |
| Summary of findings | This paper describes the potential unintended consequences of the recent changes to the role of a GP, which move away from the traditional model of an expert generalist. Some of these consequences might affect patients (e.g. increased health inequalities), however of particular relevance to our review these changes might also decrease meaning for doctors no longer able to provide comprehensive, person-centred and continuous care. |
| Strengths, limitations, and future research directions | |

| | |
|---|---|
| Paper Number | 41 |
| Author | Bevan et al |
| Year | 2018 |
| Title | Job crafting and flexible working in general practice |
| Journal | Institute for Employment Studies report |
| Region (England, Scotland, Wales, and Northern Ireland) | England |
| Study methodology | Interviews with stakeholders, including GPs, in six GP practices in England which have implemented flexible work designs and job crafting. |
| Type of document | Grey literature |
| Topic or focus of paper | Flexible working |
| Research objectives and focus | This study has been designed to explore how six GP practices have approached the challenge of meeting increasing demands from patients and ensuring that the needs of GPs and other staff for flexible working and reconfigured job roles can be balanced. |

| | |
|---|---|
| Theoretical framework/theories/ underpinnings | |
| Description of intervention | |
| Rationale for paper/review | Although there is evidence of new and innovative approaches to flexible job design and flexible working patterns in the NHS as whole (a promise contained in the NHS people plan), there are few examples of how this can be applied within general practice. Previous research has suggested reconfiguring roles, whether through introducing workplace and role flexibility and job crafting in ways that fosters job satisfaction and reduce potential burnout in employees. With evidence accumulating reporting the high levels of stress, work intensity and burnout among GPs (exacerbated by the Covid-19 pandemic), and concerns about both recruiting and retaining GPs, now may be the time to understand if improved work flexibility, and the ability for GPs to craft their roles can protect the mental health of GPs, whilst maintaining high patient safety and service quality standards. |
| What creates meaning that sustains GPs engagement with, and involvement in, work? | Flexibility was regarded by many GPs as important (and maybe even necessary) in preventing burnout, as it makes work patterns more sustainable: “The reason why I am still working here and have not given up GP practice forever is the flexibility that was given to me. I did think about giving up, but I felt that would have been a waste as I have a lot to give. If they had said no to the accommodations, I would not have had a choice but to resign. But what they get from me is my loyalty.”<br><br>“The four-day working week has certainly had an impact on my wellbeing, it has been a game changer as you can have a day off without worrying about anything. It can be tough work, but you always have that light at the end of the tunnel. I actually think it has had a massive impact on everyone’s mental health.” |
| What solutions/recommendations are proposed? | |
| From which stakeholders’ perspective? (General practitioners, patients, other HCPs) | GPs themselves, practice managers, practice nurses and other clinical and support staff. |
| How is work-related joy constructed? (Diversity of ways individuals may think about joy) | |
| What prevents/removes work-related joy? | Increasing patient expectations, alongside the assumption that GPs were not working hard enough, has led to confrontations with patients that erodes moral within general practice.<br><br>Isolated working – maybe seeing 40 patients a day – without the wider support of the team was noted as unsustainable. |
| Organisational characteristics (small, large) | Flexible working is much easier to administrate in large primary care organisations like PCNs. |

| | |
|---|---|
| Managing ill-defined illnesses and chronic complex multimorbidity | "The GPs were feeling the burden of seeing elderly patients, and their time was being taken up with issues that were frequently not clinical or medical issues, and the work was just spiralling for them. This also meant that other patients with clinical issues were not having the opportunity to see a GP"<br><br>"Meeting the complex needs of patients in a relatively deprived community seems to be a strong unifying theme which binds together the staff in the Grange practice". |
| Human connection with patients | |
| Human connection with peers | Allowing flexible working patterns engendered loyalty from GPs towards their practice, therefore creating team spirit. This may relate to a larger point about the importance of permissive, kind primary care management.<br><br>The staff interviewed commented how the practice has a positive culture which had developed over the last few years, aided by a sense of staff cohesion, where practice staff pitch in to help where they could, and where open discussions about practice matters are welcomed and encouraged. |
| Human connection across other organisations (e.g., secondary care) | |
| Learning systems (e.g., competence, capability, mastery, socially situated) | One tactic that was seen to work was providing role variety, to reduce the intensity of constant face-to-face patients that 'have become more demanding'. This allowed for staff to undertake clinic work (which meant that they were able to use any specialist training they had) and feel like they had a little more control over their patient demands. |
| Additional useful information | |
| Summary of findings | This study provides a detailed overview of how six GP practices have implemented flexible working. Of particular relevance to our study, flexible working was highlighted to prevent burnout from the intense GP workload whilst also providing an opportunity for GPs to explore areas of special interest. |
| Strengths, limitations, and future research directions | |

| | |
|---|---|
| Paper Number | 42 |
| Author | Naureen Bhatti |
| Year | 2017 |
| Title | Transforming the lives and careers of senior doctors: retaining and harnessing skills and enthusiasm in difficult times |
| Journal | BJGP |

| | |
|---|---|
| Region (England, Scotland, Wales, and Northern Ireland) | England |
| Study methodology | Commentary |
| Type of document | Secondary research |
| Topic or focus of paper | GP retainer scheme |
| Research objectives and focus | Explore what is being done – and what is needed – to retain senior GPs |
| Theoretical framework/theories/ underpinnings | |
| Description of intervention | |
| Rationale for paper/review | Senior GPs have worked hard and been committed to our roles, often at great personal sacrifice. As we have got older many are in senior positions with added responsibilities, some have health issues, many of us are sandwiched between caring roles for elderly parents and children: 80% report stress and contemplate leaving early. But we have a workforce crisis that needs them to stay. |
| What creates meaning that sustains GPs engagement with, and involvement in, work? | Senior doctors, who are nearing conventional retirement age, still enjoy the practice of medicine. However, they often want a change of pace or role, a move to part time working, being nearer to home, and more time with family and friends. The system needs to have the flexibility to allow this, so these GPs are retained within the workforce.<br><br>Other initiatives are badly needed. Coaching and mentoring are highly valued by senior doctors. We need meaningful appraisals with less onerous paperwork and more supportive discussion. We need confidential health services for everyone, not just doctors in London or GPs across England. And we need initiatives for increased peer support, reduced partnership stresses for GPs, greater educational initiatives, and more flexible working and part-time options for established doctors. |
| What solutions/recommendations are proposed? | |
| From which stakeholders' perspective? (General practitioners, patients, other HCPs) | |
| How is work-related joy constructed? (Diversity of ways individuals may think about joy) | |
| What prevents/removes work-related joy? | |

| | |
|---|---|
| Organisational characteristics (small, large) | |
| Managing ill-defined illnesses and chronic complex multimorbidity | |
| Human connection with patients | |
| Human connection with peers | |
| Human connection across other organisations (e.g., secondary care) | |
| Learning systems (e.g., competence, capability, mastery, socially situated) | |
| Additional useful information | |
| Summary of findings | This short commentary make the case for flexibility in the working lives of senior GPs – amongst other initiatives – in order to retain this important component of the workforce. |
| Strengths, limitations, and future research directions | |

| | |
|---|---|
| Paper Number | 43 |
| Author | Blease et al |
| Year | 2019 |
| Title | Artificial Intelligence and the Future of Primary Care: Exploratory Qualitative Study of UK General Practitioners' Views |
| Journal | Journal of Medical Internet Research |
| Region (England, Scotland, Wales, and Northern Ireland) | UK |
| Study methodology | Web-based survey of 720 UK GPs' opinions about the impact of future technology (including AI) on primary care, and subsequent thematic analysis of responses. |

| | |
|---|---|
| Type of document | Primary research |
| Topic or focus of paper | AI in general practice |
| Research objectives and focus | This study aimed to explore general practitioners' (GPs') opinions about the potential impact of future technology on key tasks in primary care. |
| Theoretical framework/theories/ underpinnings | |
| Description of intervention | |
| Rationale for paper/review | The potential for machine learning to disrupt the medical profession is the subject of ongoing debate within biomedical informatics and related fields. |
| What creates meaning that sustains GPs engagement with, and involvement in, work? | Overall, GPs were sceptical about the scope of future technology (particularly AI) to encroach on the traditional role of a GP. Major reasons for this included the belief that communication and empathy are exclusively human competencies and that clinical reasoning and the ability to provide value-based care as necessitating physicians' judgments. However, some GPs did suggest that AI will lead to major disruptions within general practice, although they were not specific about the nature of these disruptions. This uncertainty may be a source of anxiety for some GPs. "Medicine will be unrecognizable compared to its present form in 25 years.".<br><br>Some GPs expressed the idea that AI would reduce workload burdens, particularly administrative work: "Be useful to develop AI to do analyses of pathology returns, and read all the letters, to provide another presence in the consulting room, and to write the referral letters, organize investigations and the like, ie, act like a personal assistant might do." |
| What solutions/recommendations are proposed? | |
| From which stakeholders' perspective? (General practitioners, patients, other HCPs) | GPs |
| How is work-related joy constructed? (Diversity of ways individuals may think about joy) | |
| What prevents/removes work-related joy? | |
| Organisational characteristics (small, large) | |

| | |
|---|---|
| Managing ill-defined illnesses and chronic complex multimorbidity | |
| Human connection with patients | |
| Human connection with peers | |
| Human connection across other organisations (e.g., secondary care) | |
| Learning systems (e.g., competence, capability, mastery, socially situated) | |
| Additional useful information | |
| Summary of findings | This study presents timely information on physicians' views about the scope of artificial intelligence (AI) in primary care. Overwhelmingly, GPs considered the potential of AI to be limited, which may suggest that they are not worried about significant changes in their working lives going forward. Notably, these views differ from those working in the AI field.<br><br>However, GPs did express optimism about a potential reduction in administrative workload which may stem from AI. |
| Strengths, limitations, and future research directions | Comments were often brief, and because of the restrictions of online surveys, it was not possible to probe participants' responses to obtain a richer understanding of their views. |

| | |
|---|---|
| Paper Number | 44 |
| Author | Sola et al |
| Year | 2018 |
| Title | Predicting GPs' engagement with artificial intelligence |
| Journal | British Journal of Healthcare Management |
| Region (England, Scotland, Wales, and Northern Ireland) | UK |

| | |
|---|---|
| Study methodology | Survey of 26 GPs about perceived self-efficacy and view of artificial intelligence |
| Type of document | Primary research |
| Topic or focus of paper | AI in general practice |
| Research objectives and focus | Investigate how measures of self-efficacy influence the perception of AI as an opportunity rather than as a threat |
| Theoretical framework/theories/ underpinnings | |
| Description of intervention | |
| Rationale for paper/review | Artificial intelligence (AI) can provide GPs with a solution for two key issues. It can support improved efficiencies by helping GPs to see more patients with improved quality and reduced error. It can also improve diagnostic capabilities by allowing more data to be processed simultaneously. However, technology alone is not sufficient. GPs must be willing to adopt the technology, to trust it and to feel capable of using it for patient care. Lack of proactive engagement could be a hindrance to AI adoption, as a major change in professional behaviours may be required. Prior work suggested that higher measures of self-efficacy would correlate with the view that artificial intelligence is an opportunity. |
| What creates meaning that sustains GPs engagement with, and involvement in, work? | |
| What solutions/recommendations are proposed? | |
| From which stakeholders' perspective? (General practitioners, patients, other HCPs) | GPs |
| How is work-related joy constructed? (Diversity of ways individuals may think about joy) | |
| What prevents/removes work-related joy? | 50–69% of respondents in the groups considered AI as an opportunity, with the proportion increasing as the GSE (which measures perceived self-efficacy) score increased. Consistently, the vast majority of respondents in each group did not view AI as a threat, while the degree of intensity of threat perceptions increased with the increase of GSE score. Consistently, the vast majority of respondents in each group did not view AI as a threat, while the degree of intensity of threat perceptions increased with the increase of GSE score. This suggests that GPs in general do not view AI as a threat, and therefore that this is not a major source of anxiety for the future, but an important context relating to this is perceived self-efficacy. |

| | |
|---|---|
| Organisational characteristics (small, large) | |
| Managing ill-defined illnesses and chronic complex multimorbidity | |
| Human connection with patients | |
| Human connection with peers | |
| Human connection across other organisations (e.g., secondary care) | |
| Learning systems (e.g., competence, capability, mastery, socially situated) | |
| Additional useful information | |
| Summary of findings | A survey of GPs suggested that most do not view AI as a threat, although a lower perceived self-efficacy is associated with a greater likelihood of seeing AI as a threat (and vice-versa). |
| Strengths, limitations, and future research directions | The findings of this research are hampered by the low response rate and small sample size which limits speculation to the wider GP group. We recognise that further investigation with a larger group of GPs, perhaps segmented by the size of their practice, would be valuable. |

| | |
|---|---|
| Paper Number | 45 |
| Author | Potter et al |
| Year | 2023 |
| Title | Bridging Gaps: Improving access to general practice for and with marginalised patients- “it’s quite joyful for us, it’s really improved our work” |
| Journal | Pre-print: https://www.medrxiv.org/content/10.1101/2023.05.24.23290453v1 |
| Region (England, Scotland, Wales, and Northern Ireland) | England |
| Study methodology | Bridging Gaps is a collaboration between healthcare staff, researchers, women with lived experience of SMD and a charity that supports them in a UK city. We co-produced a project to improve access to general practice for marginalised patients, that was further developed with |

| | |
|---|---|
| | 3 inner city general practices. This study observed six collaborative service improvement meetings at three general practices and conducted documentary analysis of minutes of a further three meetings. Furthermore, interviews and focus groups with stakeholders were conducted. Data were subsequently analysed thematically. |
| Type of document | Primary research (pre-print) |
| Topic or focus of paper | To explore perceptions and experiences of improving access to general practice for people with SMD in healthcare staff and people with lived experience. |
| Research objectives and focus | To explore perceptions and experiences of improving access to general practice for people with SMD in healthcare staff and people with lived experience. |
| Theoretical framework/theories/ underpinnings | |
| Description of intervention | |
| Rationale for paper/review | People with severe and multiple disadvantage (SMD- combinations of homelessness, substance misuse, violence, abuse and poor mental health) have high health needs and poor access to primary care. |
| What creates meaning that sustains GPs engagement with, and involvement in, work? | Staff were enthusiastic about the benefits of the changes they had made and outcomes for patients, primary care and collaborating organisations: "there is people behind these labels and there is humans and human behaviour and I think we have made the difference. I think we made the difference like in Practice2, I think we made a difference in Area8... I love that and I think that's a massive thing" |
| What solutions/recommendations are proposed? | Similar co-produced interventions, like that examined in this study, may be implemented elsewhere. |
| From which stakeholders' perspective? (General practitioners, patients, other HCPs) | Stakeholders – including GPs - involved in implementing a project to improve access to general practice for patients with severe and multiple disadvantage. |
| How is work-related joy constructed? (Diversity of ways individuals may think about joy) | One GP who delivered the inclusion health clinic developed during the project in addition to mainstream general practice highlighted how enjoyable it was to have protected time to 'do a good job' for patients who really need it: "I think as well as hopefully being good for the patients. It's quite joyful for us, it's really improved our work". GPs also made the point that these complex patients were then 'offloaded' from standard clinics, reducing stress (see below) for other GPs as well. |
| What prevents/removes work-related joy? | Complex patients with SMD can be very difficult to manage properly, in an empathetic and patient centred manner, within the confines of a normal GP consultation. This can lead to overwork and burnout for GPs. This might also lead to moral distress: the experience of knowing the right thing to do while being in a situation in which it is nearly impossible to do it. |
| Organisational characteristics (small, large) | |

| | |
|---|---|
| Managing ill-defined illnesses and chronic complex multimorbidity | |
| Human connection with patients | |
| Human connection with peers | Both the process of co-production and the service-improvements that were developed initiated or strengthened teamwork and connectedness both within and between general practice teams. Some participants noted the value of relationships and teams within general practice who were focused on delivering better care for marginalised patients. One participant described relief at feeling part of a wider team of practices also motivated to tackle health inequalities. |
| Human connection across other organisations (e.g., secondary care) | |
| Learning systems (e.g., competence, capability, mastery, socially situated) | |
| Additional useful information | |
| Summary of findings | This study examined the views of a range of stakeholders to demonstrate the value of a co-produced intervention to improve access to general practice for marginalised patients. Of particular relevance to our study, professional participants found joy in doing ‘a good job’ for patients who really need it, the process of co-production strengthened teams within and between practices and prioritising continuity for patients who really need it may ease pressure on other general practice clinics. |
| Strengths, limitations, and future research directions | Men were not included in the lived experience co-production team (this was the decision of the team and organisation that supports them). |

| | |
|---|---|
| Paper Number | 46 |
| Author | Lynch et al |
| Year | 2023 |
| Title | Holding the complex whole: generalist philosophy, priorities and practice that facilitate whole-person care |
| Journal | Australian Journal of General Practice |
| Region (England, Scotland, Wales, and Northern Ireland) | N/A |
| Study methodology | Commentary |

| | |
|---|---|
| Type of document | Secondary research |
| Topic or focus of paper | Generalism |
| Research objectives and focus | Generalist philosophy, priorities and clinical skills are outlined in this paper as part of the ongoing effort to help GPs value, hone and protect the often-misunderstood complexity of their work. |
| Theoretical framework/theories/ underpinnings | The four priorities of the Craft of Generalism within general practice: whole-person scope, relational process, healing orientation and integrative wisdom. The five practical skills associated with these priorities: broad awareness, respectful connection, capable engagement, calm-sense making and owning yourself. |
| Description of intervention | |
| Rationale for paper/review | Generalist care is undermined by systemic and bureaucratic devaluing of GP time and expertise, primary care policy that fragments care and encourages short transactional encounters and an excessive focus on disease or procedures. Clinical decision making in the face of uncertainty, undifferentiated symptoms, conflicted goals or chronic disease and grief is also cognitively, emotionally and morally difficult. In order to protect GPs from doubting the value of their work, it is important to understand generalism as so much more than biomedical service provision: it requires philosophical attitudes, overarching priorities and practical skills that are often unnamed and unnoticed. |
| What creates meaning that sustains GPs engagement with, and involvement in, work? | This paper argues that, in recent years, there has been an increasing focus on a narrow, biomedical approach in medicine. This (alongside other changes to the work of GPs) has demoralised GPs. This paper attempts to clarify the value of generalism, which it argues will increasing the meaning GPs associate with their jobs.<br><br>The biomedical approach is often blind to the whole person in their context and relationships – general practice can bring generalist innovation to the whole healthcare system. Others have highlighted the importance to generalist care of stories, the personhood of the patient, the relational ways of ‘being’ of the clinician, the complexity of people and systems, practical goals that help people live their lives, scholarly wisdom and ‘knowing’ that comes through experience (none of these are elaborated on by the paper). |
| What solutions/recommendations are proposed? | |
| From which stakeholders’ perspective? (General practitioners, patients, other HCPs) | GPs |
| How is work-related joy constructed? (Diversity of ways individuals may think about joy) | |
| What prevents/removes work-related joy? | |
| Organisational characteristics (small, large) | |

| | |
|---|---|
| Managing ill-defined illnesses and chronic complex multimorbidity | |
| Human connection with patients | |
| Human connection with peers | |
| Human connection across other organisations (e.g., secondary care) | |
| Learning systems (e.g., competence, capability, mastery, socially situated) | |
| Additional useful information | |
| Summary of findings | Greater awareness of generalist philosophy and priorities amongst GPs may help them value their work more, in the context of a system built on biomedical service provision. |
| Strengths, limitations, and future research directions | |

| | |
|---|---|
| Paper Number | 47 – This comprehensive report on the ReGROUP study (Campbell et al., 2019) comprises a collection of mostly self-contained workstreams and contains too much information for extraction into a single form. Consequently, separate data extraction forms were generated for each workstream at the end of this document. |

| | |
|---|---|
| Paper Number | 48 |
| Author | Agius et al |
| Year | 2015 |
| Title | How can a postgraduate professional education and development course benefit general practitioners?: A qualitative study |
| Journal | Journal of Educational Evaluation for Health Professions |

| | |
|---|---|
| Region (England, Scotland, Wales, and Northern Ireland) | England |
| Study methodology | Thematic analysis of the assessed coursework portfolios of GPs who had undertaken a 'professional education and development course'. These detailed the perceived impact of the course on personal and practice development. |
| Type of document | Primary research |
| Topic or focus of paper | Postgraduate medical education |
| Research objectives and focus | |
| Theoretical framework/theories/ underpinnings | |
| Description of intervention | |
| Rationale for paper/review | |
| What creates meaning that sustains GPs engagement with, and involvement in, work? | Seven major thematic categories were extrapolated from the data: leadership, resilience, quality improvement, change management, development of new services, educational expertise, and patient safety. In each category, we found evidence that the course enabled development of practitioners by enhancing knowledge and skills which had a positive impact upon their self-perceived effectiveness and motivation (link to mastery here). |
| What solutions/recommendations are proposed? | Postgraduate medical education – like PED courses – may serve a valuable purpose for current trainees and existing GPs. |
| From which stakeholders' perspective? (General practitioners, patients, other HCPs) | GPs |
| How is work-related joy constructed? (Diversity of ways individuals may think about joy) | |
| What prevents/removes work-related joy? | |
| Organisational characteristics (small, large) | |

| | |
|---|---|
| Managing ill-defined illnesses and chronic complex multimorbidity | |
| Human connection with patients | |
| Human connection with peers | GPs who undertook the PED believed their resilience was increased when confronted by difficult patients and colleagues. Recurrent themes attributed to PED included improved positivity; enthusiasm; flexibility, particularly for problem solving; personal and team resilience; team building, with the ideas prompted from the course material and discussions facilitating a happier and more effective practice team; peer support, including its beneficial role in burnout prevention; and a greater awareness of maintaining work-life balance.<br><br>Discussing patient safety issues with the team at practice meetings was thought to significantly assist in ensuring positive outcomes and reduce the possibility of staff burnout – in instances where GPs were not part of a team, the potential stress of these incidences was particularly noted. |
| Human connection across other organisations (e.g., secondary care) | |
| Learning systems (e.g., competence, capability, mastery, socially situated) | |
| Additional useful information | |
| Summary of findings | This paper suggests that postgraduate education for GPs can increase their effectiveness in key areas of practice. Relevant to our review, this may contribute to the development of mastery and motivation. |
| Strengths, limitations, and future research directions | Our case study relates to a small sample of participants who have undertaken a PED course in a single region, so there are inevitable limitations in terms of generalizability in the quantitative sense. A further limitation relates to the fact that the study was based on analysing portfolio entries that have been completed as part of an assessment process. There is a possibility that the entries were completed by students in a way that met learning outcomes and satisfied tutors rather than reflect their own views, |

| | |
|---|---|
| Paper Number | 49 |
| Author | Baird et al |
| Year | 2022 |
| Title | Levers for change in primary care: a review of the literature |
| Journal | Report authored by The King’s Fund |

| | |
|---|---|
| Region (England, Scotland, Wales, and Northern Ireland) | England |
| Study methodology | Literature review |
| Type of document | Secondary research |
| Topic or focus of paper | Programmes of change in general practice |
| Research objectives and focus | Consider the available literature on the processes and levers used in programmes of change, improvement, innovation and transformation in primary care, with a particular focus on general practice. |
| Theoretical framework/theories/ underpinnings | |
| Description of intervention | |
| Rationale for paper/review | Commissioned by NHS England to inform the Fuller Stocktake. |
| What creates meaning that sustains GPs engagement with, and involvement in, work? | |
| What solutions/recommendations are proposed? | |
| From which stakeholders' perspective? (General practitioners, patients, other HCPs) | |
| How is work-related joy constructed? (Diversity of ways individuals may think about joy) | |
| What prevents/removes work-related joy? | The implementation of financial levers can have adverse effects on staff morale. Best practice suggests working with staff to design any system, so that they do not end up feeling disempowered by the incentive structures, feel like they are compromising ethical standards or feel like the extra administrative tasks they are being asked to carry out to demonstrate that they are achieving targets are more trouble than they are worth.<br><br>GPs are "not supportive" of CCGs' use of performance management mechanisms. |
| Organisational characteristics (small, large) | |

| | |
|---|---|
| Managing ill-defined illnesses and chronic complex multimorbidity | |
| Human connection with patients | |
| Human connection with peers | |
| Human connection across other organisations (e.g., secondary care) | |
| Learning systems (e.g., competence, capability, mastery, socially situated) | |
| Additional useful information | 'Soft' levers of culture and leadership are critical for successful initiatives, creating an organisation that offers a safe environment for people to learn and experiment. |
| Summary of findings | This paper considers the processes and levers of change within general practice, mainly focussing on how to achieve efficiency and better outcomes. The only relevant finding to our study is that financial incentives (pay for performance) can have an adverse impact on staff moral. This paper also reiterates the importance of co-design to staff moral. |
| Strengths, limitations, and future research directions | |

| | |
|---|---|
| Paper Number | 50 |
| Author | Dale et al |
| Year | 2015 |
| Title | Retaining the general practitioner workforce in England: what matters to GPs? A cross-sectional study |
| Journal | BMC Family Practice |
| Region (England, Scotland, Wales, and Northern Ireland) | England |
| Study methodology | Survey of GPs exploring career plans and experiences of work-related pressures. Quantitative data were analysed using logistic regression analysis and principal components analysis. Qualitative data were analysed thematically. |
| Type of document | Primary research |

| | |
|---|---|
| Topic or focus of paper | GP retention |
| Research objectives and focus | Investigate factors underlying GP early retirement and how these might be mitigated. |
| Theoretical framework/theories/ underpinnings | |
| Description of intervention | |
| Rationale for paper/review | The general practice (GP) workforce in England is in crisis, reflected in increasing rates of early retirement and intentions to reduce hours of working. |
| What creates meaning that sustains GPs engagement with, and involvement in, work? | Quantitative analysis indicated five key 'push' factors for GP early retirement (ranked by 488 GPs from 1, 'not important', to 5, 'very important'): intensity of workload (mean = 4.6), volume of workload (4.5), time spent on unimportant tasks (4.4), introduction of a seven-day working week (4.2) and job satisfaction (4.2).<br><br>With regards to workplace related factors, two principle components emerged: overall workload (weighted combination of all work-related factors) and working conditions (the importance given to flexibility and job satisfaction relative to volume and intensity of work and the other items). Overall workload is more important to GPs aged under 50 years and more important to GP partners. Working conditions became less important with increasing age and are more important for males than females. Furthermore, the number of GPs within a practice is inversely correlated with working conditions.<br><br>With regards to non-workforce related factors, two principle components emerged: work-life flexibility and personal development. Work-life flexibility is a weighted mean of all non-work related factors and personal development represents the difference in importance of scores between those factors related to training and education opportunities and other nonwork related factors. Work-life flexibility is less important with increasing age, more important for female GPs, less important for increased length of time in practice, less important for GPs working in large practices than small practices, and more important for GPs with portfolio careers. Personal development is more important for female GPs, GPs with over 10 years' service, and for GP principals than non-principal GPs. |
| What solutions/recommendations are proposed? | Quantitative analysis indicated a number of measures that might reverse intention to leave general practice: reduced workload intensity (mean = 4.5), reduced workload volume (4.4), less administration (4.3), greater time spent with patients (4.2), no out-of-hour commitments (3.9), more flexible working conditions (3.5) and greater clinical autonomy (3.3). Individual motivators included incentive payments (3.7), increased pay (3.1), protected time for education and training (3.6), additional annual leave (2.9) and opportunity for a sabbatical (2.7).<br><br>Participants' comments emphasised the importance of the NHS, government and representative professional bodies adopting a more 'realistic' approach in order to reduce workload, administrative activity and patients' expectations, and increasing the funding available to general practice: "...change in attitude from politicians and press blaming all the NHS problems at the door of general practice. No more unrealisable promises to the public!". |

| | |
|---|---|
| | Addressing the costs of indemnity and professional registration was seen as important. In addition, the need for more supportive structures and networks was also emphasised, together with a more flexible and developmental approach to GP appraisal and revalidation: “Local support for struggling GPs i.e. access to counselling and CBT locally, face to face and confidential. RCGP should stop adding hoops to GMC revalidation supporting information requirements. All GP's should have access to a network not just '20 plus' and 'first 5'. Do not forget the 1 in 4 GP's who are locums. Allow GP's to take career breaks and fund/ support their return to general practice”. |
| From which stakeholders’ perspective? (General practitioners, patients, other HCPs) | GPs |
| How is work-related joy constructed? (Diversity of ways individuals may think about joy) | |
| What prevents/removes work-related joy? | Free text comments allowed participants to expand on the key components of their increased workload. These were: growth in patient demand and expectations (which may reflect an increasingly consumerist approach to healthcare), recruitment and retention difficulties, “exponential” increases in administration and bureaucracy (with little perceived benefit to patients), growth in additional roles and responsibilities (e.g. CCG-related responsibilities), transfer of work from secondary care without commensurate resources, increasing complexity and chronic ill health (unmanageable within 10-minute consultations), revalidation and regulatory assessment which were perceived to be of little benefit and political pressure for a seven-day working week in general practice. These work-related pressures significantly increased GPs’ stress and led to burn out: “...I have recently negotiated with my practice to reduce my hours from 3 to 2 days per week. While I know it sounds pathetic I really don't feel I can cope working 3 days anymore. The pressure is tremendous. I get home after a 10 h day and feel as if I have been hit round the head with a brick...I am only 40 yet feel burnt out. It is a miserable situation”.<br><br>GPs felt that the ‘constant’ and ‘relentless’ negative portrayal of general practice by the government and media was damaging their morale and professional identity. |
| Organisational characteristics (small, large) | |
| Managing ill-defined illnesses and chronic complex multimorbidity | |
| Human connection with patients | |
| Human connection with peers | |

| | |
|---|---|
| Human connection across other organisations (e.g., secondary care) | |
| Learning systems (e.g., competence, capability, mastery, socially situated) | |
| Additional useful information | The quantitative analysis highlights just how bad GP retention is: of 1,192 GPs who participated, 978 (82.0 %) stated that they intend to leave general practice, take a career break and/or reduce clinical hours of work within the next five years. This included 488 (41.9 %) who intend to leave practice, and almost a quarter (279; 23.2 %) intending to take a career break. Only 67 (5.6 %) planned to increase their hours of clinical work. |
| Summary of findings | For participants planning to leave practice, quantitative analysis indicated that the issues that most influenced intentions were volume and intensity of workload, time spent on "unimportant tasks", introduction of seven-day working and lack of job satisfaction. Participants expanded on these qualitatively, with the following themes identified: cumulative impact of work-related pressures, the changing and growing nature of the workload, and the consequent stress.<br><br>Reducing workload intensity, workload volume, administrative activities, with increased time for patient care, no out-of-hour commitments, more flexible working conditions and greater clinical autonomy were identified as the most important requirements to address the workforce crisis. In addition, incentive payments, increased pay and protected time for education and training were also rated as important. |
| Strengths, limitations, and future research directions | A major strength is the diversity of GPs that participated, from those early in their career and undertaking portfolio careers to those approaching retirement, from practices of all sizes, and from settings ranging from inner city to rural. A further strength is that our study explored not only intentions to leave general practice or reduce working hours, but also to take a career break.<br><br>A key limitation is the uncertainty over the response rate. Owing to governance issues, we faced difficulties in obtaining GPs' contact details and hence relied on emailed invitations to practices being forwarded by managers to potential participants, and that the intended recipients read the email. The survey was undertaken around the Christmas/New Year period and seasonal factors, such as workload pressures prevalent at this time of year, may have influenced responses. It is possible that those who felt most over-worked or disillusioned may have been less likely to notice or feel motivated to respond to the invitation to the survey. |

| | |
|---|---|
| Paper Number | 51 |
| Author | Baird et al |
| Year | 2016 |

| | |
|---|---|
| Title | Understanding pressures in general practice |
| Journal | King’s Fund Report |
| Region (England, Scotland, Wales, and Northern Ireland) | UK |
| Study methodology | Quantitative analysis:<br>1. A study of 30 million individual contacts with patients from 177 practices over five years provided by ResearchOne<br>2. A survey of 43 practices for a sample week in October 2015 examining activity and workload.<br>3. A survey of 318 GP trainees, examining workload and future career intentions.<br><br>Qualitative analysis<br>1. Interviews with 60 staff at four practices of varying sizes in Plymouth, Shrewsbury, Sheffield and London<br>2. Literature search<br>3. Scoping conversations with a range of stakeholders, including national bodies and leaders of CCGs<br>4. Survey of 318 GP trainees and subsequent thematic analysis of responses |
| Type of document | Secondary research |
| Topic or focus of paper | Crisis in general practice |
| Research objectives and focus | Provide the most detailed analysis to date (2016) about how and why the crisis in general practice has happened |
| Theoretical framework/theories/ underpinnings | |
| Description of intervention | |
| Rationale for paper/review | General practice is in crisis. Workload has increased substantially in recent years and has not been matched by growth in either funding or in workforce. A lack of nationally available, real-time data means that this crisis has been until recently largely invisible to commissioners and policy-makers. |
| What creates meaning that sustains GPs engagement with, and involvement in, work? | Of 318 GP trainees surveyed, 31 per cent intended to do full-time clinical work one year after qualification, falling to 11 per cent five years after qualification and just 10 per cent 10 years after qualification. Part-time work appears to be a more attractive option at earlier career stages (preferred by 46 per cent of respondents at one year after qualification, 37 per cent at five years after qualification and 24 per cent at 10 years after qualification). |

| | |
|---|---|
| What solutions/recommendations are proposed? | Based on our research, our 'prescription' for the future of general practice that can deliver the care people need has five items, which we explain in detail below: understanding data at a system level; improving the current system of care; developing and promoting new models of general practice; developing and sustaining a workforce for the future; and ensuring that capacity and funding match changing workloads (the report expands on these in detail, but they are not directly relevant to our review). |
| From which stakeholders' perspective? (General practitioners, patients, other HCPs) | |
| How is work-related joy constructed? (Diversity of ways individuals may think about joy) | |
| What prevents/removes work-related joy? | Activity in general practice increased significantly over the five years examined: total direct face-to-face and telephone contacts with patients increased by 15.4 per cent across all clinical staff groups between 2010/11 and 2014/15. During the same period, the average patient list size increased by 10 per cent. Qualitative analysis suggested a number of reasons for this, such as patients' desire for both rapid access and continuity of care (other factors, which are not directly relevant to our review, are expanded on in the report).<br><br>In 2015, in real terms, income for GPs fell to its lowest level since 2002/3.<br><br>The raising referral threshold has left GPs to manage difficult cases without appropriate support, and holding a level of risk that was adding to feelings of burnout.<br><br>Finally, a fear of litigation came across strongly in our survey of trainees, who pointed to higher indemnity costs, increased incidence of complaints or being sued, and shifting public views about doctors. |
| Organisational characteristics (small, large) | The results of the trainee survey show that partnership is no longer an attractive option for many. Reasons for this included: increased pressures, reduced work–life balance, reduced income, extra responsibility with reducing benefit, uncertainty, changes in the NHS and general practice funding crisis. |
| Managing ill-defined illnesses and chronic complex multimorbidity | The increasing numbers of complex patients with multiple, long-term conditions presented challenges to GPs working within a 10 minute model of consultation: "The way the system is set up is, it is set up to enable you to have a 10-minute consultation and within that 10-minute consultation you deal with the reactive stuff. But then you are thinking, 'Oh my gosh, they've got all of those long-term conditions, I'm not really doing anything. I need to...' So you know, you can see that things aren't being addressed but they've not come for that problem.". Of note, the multidisciplinary practice team was noted by GPs as being very useful in helping support some of this work.<br><br>Due to increasing delegation in general practice, theanalysis found that GPs are increasingly seeing patients with multiple long-term conditions, which requires them to manage complex processes and make difficult decisions in a short time with little respite, resulting in a very intense working day: "You rarely see anybody who's just got one problem. You don't see really any just basic chest infections, exacerbation of asthma, sore throats. The nurses see all those things." |

| | |
|---|---|
| Human connection with patients | There was a strong consensus among our interviewees and survey respondents that patients' expectations of care have increased over time. Some interviewees reported a widespread expectation among patients that they should be prescribed antibiotics for minor illnesses or should receive diagnostic tests or be referred for specialist care when not clinically indicated. This might harm the doctor-patient relationship.<br><br>Studies have found numerous benefits of continuity, including increased patient and staff satisfaction, greater accountability, increased trust within the doctorpatient relationship (with positive effects on treatment compliance), improved disease management and better health outcomes (Freeman and Hughes 2010). Loss of continuity is challenging for GPs as well as patients; it can be extremely difficult for a GP to understand and address the needs of a patient with complex problems within a single 10-minute consultation. |
| Human connection with peers | The nature of general practice means that, while relationships within the primary care team may be good, it is an isolated occupation for clinicians. Unlike in a hospital setting – where there is a team of consultants, junior doctors, nursing and other staff all sharing the care of a patient – that sense of a team approach is much less evident in primary care. GPs do not have another doctor to hand over to at the end of a shift |
| Human connection across other organisations (e.g., secondary care) | GPs believed that referral management schemes had eroded their relationship with secondary care – and therefore their ability to ask for support, guidance and opinion from secondary care colleagues: "I think relationships with the hospital are really poor. I think they are far poorer now than they were 10 years ago. I think the days of being able to phone up a consultant and ask for some advice seem to be long gone and there are one or two who you can get hold of – email seems to be the preferred way – and often you don't get the advice that you want." |
| Learning systems (e.g., competence, capability, mastery, socially situated) | |
| Additional useful information | Studies of reforms in the early 2000s designed to improve access did find that they had reduced continuity of care (Campbell et al 2010).<br><br>The growth of part-time working in general practice has been widely attributed to feminisation of the workforce. However, our survey showed that intentions for part-time or portfolio working are common across both sexes. Although a greater proportion of female than male respondents intend to work part-time and fewer intend to work full-time, these differences diminish over progressive career points. The most commonly cited reason for not pursuing full time clinical work was 'intensity of the working day' rather than 'family commitments'. |
| Summary of findings | This report is a very detailed analysis of how and why general practice has got busier. Much of this is not directly relevant to our review, although is a useful resource if we wish to explore workload pressures further. However, scattered throughout the report, there are individual points which relate to GPs' relationship with work, and I have extracted these above. |
| Strengths, limitations, and future research directions | |

| | |
|---|---|
| Paper Number | 52 |
| Author | Grut et al |

| | |
|---|---|
| Year | 2023 |
| Title | Primary health care during the COVID-19 pandemic: A qualitative exploration of the challenges and changes in practice experienced by GPs and GP trainees |
| Journal | PLOS One |
| Region (England, Scotland, Wales, and Northern Ireland) | UK |
| Study methodology | Interviews with GPs and GP trainees. Subsequent thematic analysis of responses. |
| Type of document | Primary research |
| Topic or focus of paper | Changes to general practice during COVID-19 |
| Research objectives and focus | Explore the experiences and perspectives of GPs and GP trainees during the pandemic. |
| Theoretical framework/theories/ underpinnings | |
| Description of intervention | |
| Rationale for paper/review | |
| What creates meaning that sustains GPs engagement with, and involvement in, work? | |
| What solutions/recommendations are proposed? | Drawing on lessons for the pandemic, GPs made four main suggestions for the future:<br>1. Telemedicine hybrid (balancing remote and face-to-face consulting) with effective triaging<br>2. Faster specialist advice for GPs, preventing unnecessary referrals<br>3. Public education on accessing GP care, reinforcing practices were open but consulting mainly via telephone and that this was an appropriate model of care<br>4. Professional education, including clinical leadership and guidance |
| From which stakeholders' perspective? (General practitioners, patients, other HCPs) | GPs & GP trainees. |
| How is work-related joy constructed? (Diversity of | Telemedicine was noted to have both positive and negative effects on workload. It increased workload in some cases, by increasing patients' ease of access to diagnostic care. In other cases, it saved time by increasing efficiency, especially on consultations that could easily be dealt |

| | |
|---|---|
| ways individuals may think about joy) | with over the phone. Interestingly, GP trainees often felt more comfortable during remote consultations as they could more easily refer to resources like NICE CKS. |
| What prevents/removes work-related joy? | GPs thought they were 'thrown in at the deep end' during the COVID-19 pandemic. They found the rapid changes associated with COVID-19 – including social distancing, telemedicine and managing an unfamiliar condition – isolating and frightening. They perceived a lack of support and guidance from governing bodies for these changes.<br><br>Changes to teaching and assessment was a major source of stress for GP trainees during the pandemic. Trainees felt like they were missing out on non-pandemic clinical experiences and teaching: "I'm pretty sure I'm going to quality in August having never done a full face-to-face GP clinic. Which I find kind of horrifying". Some trainees also described reduced support in their clinical decision making during the pandemic. Exam formats were changed during the pandemic, which resulted in trainees having to restart and rethink their preparation.<br><br>GPs were worried about future demand for general practice caused by the pandemic – in particular, due to the elective backlog and missed diagnoses. |
| Organisational characteristics (small, large) | |
| Managing ill-defined illnesses and chronic complex multimorbidity | GPs found it difficult to safely risk stratify patients over the phone, which may contribute to anxiety.<br><br>Delayed referrals to secondary care during the pandemic led to GPs managing more complex patients. GPs interviewed had mixed feelings about this. Some described themselves to be "quite excited to be taking on more responsibilities from the hospitals", whilst others were unhappy with the additional burdens. |
| Human connection with patients | Participants expressed concerns regarding their ability to develop strong and reassuring doctor-patient relationships on the phone. Trainees described the difficulties they faced trying to 'build up the trust. .. as a new doctor in that surgery', reporting that they instead felt like a faceless 'call centre' or 'convenience store of general practice' for their patients.<br><br>The delayed referrals to secondary care during the pandemic were emotionally difficult for GPs: "I've had letters back from secondary care saying... patients who have been on a 2-Week Wait or a Rapid Access Cancer Pathway, you should not tell them they'll be seen soon because they won't be. Which is really hard, to speak to cancer patients" |
| Human connection with peers | Given the uncertainty shared amongst practice staff during the pandemic, participants valued camaraderie and the reassurance they felt navigating the pandemic 'as a team, supporting each other'.<br><br>During COVID-19, GP trainees felt "lonely" following the loss of networking with other trainees, whilst having to distance from their usual support networks outside of work. |
| Human connection across other organisations (e.g., secondary care) | |

| | |
|---|---|
| Learning systems (e.g., competence, capability, mastery, socially situated) | |
| Additional useful information | |
| Summary of findings | Thematic analysis of GPs' experiences during the pandemic revealed five main themes: 'Thrown in at the deep end', Telemedicine: 'It needs to be a happy balance', Delayed referrals and 'holding' patients, The Covid Cohort–training in Covid and Time for change. |
| Strengths, limitations, and future research directions | Recruitment yielded a cohort varied by both participant and practice characteristics, with participants assuming a variety of roles within practices and key decision-making groups, producing a rich and novel dataset. A limitation of recruitment was the potential for self-selection bias, with those expressing interest in the study potentially having had a more extreme experience. Furthermore, most participants were recruited via social media, resulting in a relatively young cohort. Findings may therefore not reflect the views of more established GPs; however, they were in line with a recent unpublished study investigating video-consulting, which sampled an older cohort of GPs. |

| | |
|---|---|
| Paper Number | 53 |
| Author | Martin & McDowall |
| Year | 2022 |
| Title | The professional resilience of mid-career GPs in the UK: a qualitative study |
| Journal | BJGP |
| Region (England, Scotland, Wales, and Northern Ireland) | UK |
| Study methodology | Interview with GPs, followed by thematic analysis of responses. |
| Type of document | Primary research |
| Topic or focus of paper | GP resilience |
| Research objectives and focus | Explore how GPs perceive professional resilience, and what workplace factors influence it? |
| Theoretical framework/theories/ underpinnings | |
| Description of intervention | |
| Rationale for paper/review | With a continued crisis of increasing workload and reduced workforce in general practice, supporting resilience is a key strategy for sustaining the profession into the future. |

| | |
|---|---|
| What creates meaning that sustains GPs engagement with, and involvement in, work? | Relationships out of work and corresponding support, hobbies and interests increased resilience.<br><br>Personal factors contributed to reduced resilience included: life events, a poor work–life balance, caring responsibilities outside of work, financial outgoings, isolation at work, a tendency to be self-critical, and the emotional burden of absorbing patient distress repeatedly, day after day.<br><br>In order to increase resilience and prevent burnout, many GPs reported refusing to do certain aspects of work (sometimes with patients being negatively impacted as a result) and reducing hours. In many cases, this involved portfolio work to maintain income whilst reducing clinical work. One GP argued that this is undermining the essence and meaning of general practice, and might also change how GPs are perceived by colleagues and the public. However, others noted that taking on other roles outside of clinical is interesting and valuable got GPs' professional development and resilience: "Senior partners used to do 5 days a week, nine sessions full time...you are working too much and you need to reduce that. It's a different job intensity-wise since they started". |
| What solutions/recommendations are proposed? | That GPs feel to improve resilience they need to work fewer clinical hours may have huge implications for a workforce already in crisis, and ultimately, for the health care of the UK population. Urgent consideration must be given to this by GP leaders, NHS England, and the Department of Health and Social Care to understand how the GP role could be made more attractive to newly qualified doctors and how the health service can retain its mid-career GPs. In addition, any lessons learned not only regarding future workforce planning but also a strategic approach to career development and support must be applied. |
| From which stakeholders' perspective? (General practitioners, patients, other HCPs) | GPs |
| How is work-related joy constructed? (Diversity of ways individuals may think about joy) | |
| What prevents/removes work-related joy? | Strongly reflected in most of the interviews was that external factors reduced resilience. Top-down change imposed by government and excessive, regular changes to the GP contract, excessive regulation, an overly target-driven culture, a hostile public perception of the profession, and a perceived lack of support from the Royal College of General Practitioners were all identified as significant negative influences, which left GPs with a sense of having lost their voice and identity.<br><br>Work intensification was also highlighted by most participants, which includes aspects such as long working hours, few breaks, time pressures, and constant interruptions as being the norm for a working day.<br><br>Being too caring was linked to emotional distress and decreased resilience: "I know some GPs who are incredibly caring; the make great GPs but it is not good for their resilience and at the other end you meet [GPs] who couldn't care less but they go home happy every night".<br><br>Some GPs suggested that social media can reduce their resilience at work, due to it feeling hostile and difficult. |

| | |
|---|---|
| Organisational characteristics (small, large) | Well-run practice, good staff, good leadership, and stable finances increased resilience.<br><br>Organisational factors such as long-term absences of GPs and other staff, problems with recruitment, personal and partnership financial difficulties, complex pension and tax issues, poorly functioning IT systems, rising practice list sizes, increasing patient demand, and a need to provide high levels of patient satisfaction were all identified as factors that reduce resilience at work |
| Managing ill-defined illnesses and chronic complex multimorbidity | |
| Human connection with patients | High levels of continuity of care increased resilience |
| Human connection with peers | Relationships in (and out) of work, sharing of responsibility, good communication and social time with colleagues increased resilience. |
| Human connection across other organisations (e.g., secondary care) | |
| Learning systems (e.g., competence, capability, mastery, socially situated) | |
| Additional useful information | This paper discusses the varied definitions of the term 'resilience' amongst GPs. Most of the GP participants admitted that they felt unsure about the definition of resilience but acknowledged its current wide use in the NHS. Every participant talked about how they had felt different levels of resilience at different times in their lives and careers. Importantly, resilience was perceived to be about a positive sense of thriving at work, and more than just getting by – which links to mastery. |
| Summary of findings | This study discusses the definition of resilience amongst GPs then presents a number of factors which increase or decrease the resilience of GPs at work. Of particular notability, this study highlights that GPs feel they need to be 'obstructive' at work to increase their resilience, which means working fewer clinical hours and saying no to additional tasks. |
| Strengths, limitations, and future research directions | Conducting a qualitative study in the early months of a world pandemic may influence opinions on resilience. It cannot be discounted that the participants may have taken part because of a history of difficulties with resilience, constituting a potential self-selection bias. |

| | |
|---|---|
| Paper Number | 54 |
| Author | Mitchell et al |
| Year | 2018 |
| Title | Recruitment, retention and returning to General Practice: A rapid scoping review to inform the Greater Manchester Workforce Strategy |
| Journal | N/A (NIHR Collaboration for Leadership in Applied Health Research and Care) |

| | |
|---|---|
| Region (England, Scotland, Wales, and Northern Ireland) | UK (with "some international studies where relevant") |
| Study methodology | Rapid scoping review which included papers looking at the recruitment or retention of GPs, mainly from the UK |
| Type of document | Secondary research |
| Topic or focus of paper | GP recruitment, retention and return |
| Research objectives and focus | Understand contextual factors and pressures affecting the GP workforce |
| Theoretical framework/theories/ underpinnings | |
| Description of intervention | |
| Rationale for paper/review | General Practitioner numbers are falling and not keeping pace with workload demands, with fewer newly qualified doctors choosing to enter primary care and GPs leaving the profession early |
| What creates meaning that sustains GPs engagement with, and involvement in, work? | Existing studies report a complex range of factors influencing GPs' decisions to leave the profession, including: organisational change, clash of values, increasing workload, negative portrayal in the media, workplace issues, lack of support, perceived value as a profession, feelings of autonomy over their work and perceived lack of support from leaders. There is also some suggestion from the research that the lack of funding for UK general practice may make working abroad more appealing<br><br>One qualitative study examining GP retention found that GPs benefitted from clarified roles and expectations within general practice, wishing to feel valued and listened to when planning healthcare delivery and be more supported with risk management and maintaining patient contact.<br><br>The most likely factors reported to encourage later retirement were reduced hours and administrative duties as well as protected pension rights rather than purely financial incentives.<br><br>Whether or not GPs are having families there is a growing demand for better work-life balance with an increased trend for both men and women to work part-time and be salaried. |
| What solutions/recommendations are proposed? | Reducing GP workload is critical. Changes to day-to-day GP workload by using administrative staff for various tasks such as correspondence, prescriptions and other tasks showed promise in some of the vanguard sites. Other actions such as reception staff, websites and other professionals directing patients to the most appropriate professional when a GP isn't needed (active signposting), developing the team, supporting self-care and social prescribing also showed positive signs to increasing capacity and were broadly welcomed by GPs<br><br>'GP Career Plus', is a new strategy being implemented as a pilot project for twelve months in 10 areas nationally to offer flexibility and support to GPs who are about to leave general practice; this scheme will be evaluated in 2018. The idea of 'GP Career Plus' is to increase |

| | |
|---|---|
| | clinical capacity by offering clinical support, carrying out a specific and limited clinical role, as well as offering more strategic leadership, mentoring and training. |
| From which stakeholders' perspective? (General practitioners, patients, other HCPs) | N/A |
| How is work-related joy constructed? (Diversity of ways individuals may think about joy) | Studies examining GP retention have suggested enabling GPs to develop a portfolio career or a sub-specialisation, echoing qualitative findings that job satisfaction is more important than income to GPs |
| What prevents/removes work-related joy? | A qualitative study carried out in England of GPs aged 50-60 years also highlighted other reasons for early retirement. One of the key messages reported by GPs was the feeling of doing an (almost) 'undoable' job, pessimism about the future of general practice as well as being financially able to retire. |
| Organisational characteristics (small, large) | |
| Managing ill-defined illnesses and chronic complex multimorbidity | |
| Human connection with patients | Studies examining GP retention have suggested slowing the pace of administrative change to protect face-to-face patient contact. |
| Human connection with peers | |
| Human connection across other organisations (e.g., secondary care) | |
| Learning systems (e.g., competence, capability, mastery, socially situated) | |
| Additional useful information | |
| Summary of findings | This scoping review highlights that a range of factors are leading to GPs leaving the profession, including an unmanageable workload with poor support and constant organisational change, a perception that the profession not valued and a perceived lack of autonomy and support. Strategies to improve retention relate to trying to increase capacity and reduce workload, encourage variation in working life through portfolio careers and subspecialisms as well as greater support for those wishing to change their clinical workload. I have not extracted data on recruitment as this is not relevant to our review. |
| Strengths, limitations, and future research directions | |

| | |
|---|---|
| Paper Number | 55 |
| Author | Rammya Mathew |
| Year | 2021 |
| Title | Rammya Mathew: Making general practice a sustainable career again |
| Journal | BMJ |
| Region (England, Scotland, Wales, and Northern Ireland) | UK |
| Study methodology | Commentary |
| Type of document | Secondary research |
| Topic or focus of paper | Sustainability of the partnership model in general practice |
| Research objectives and focus | Explain why the partnership model is currently unsustainable within general practice |
| Theoretical framework/theories/ underpinnings | |
| Description of intervention | |
| Rationale for paper/review | |
| What creates meaning that sustains GPs engagement with, and involvement in, work? | Partnership has traditionally been viewed as the pinnacle of a career in general practice. It brings autonomy: a sense of control over how you run your practice, see your patients, and work with your colleagues. Many GPs also strongly believe that partnership fosters a unique commitment to one’s population and provides one of the most cost-effective models of primary care delivery.<br><br>However, there are currently two major barriers to the partnership model. Firstly, GPs who mostly have no training beyond clinical medicine become owners of a small to medium sized business overnight. They’re in charge of a budget, accountable for the safe running of a practice, and responsible for employing a range of staff, both clinical and non-clinical (from the perspective of our review, it is also easy to see how this might be a source of stress and uncertainty). Secondly, it is no longer financially viable to have an all partner model, which was how most practices traditionally operated. With successive years of underinvestment in general practice, the only way for partners to now make a profit is to have a lean management structure with a few profit sharing partners and many more salaried GPs on a fixed income, who have no financial stake in the practice. |

| | |
|---|---|
| What solutions/recommendations are proposed? | |
| From which stakeholders' perspective? (General practitioners, patients, other HCPs) | |
| How is work-related joy constructed? (Diversity of ways individuals may think about joy) | |
| What prevents/removes work-related joy? | "The work is relentless" |
| Organisational characteristics (small, large) | |
| Managing ill-defined illnesses and chronic complex multimorbidity | |
| Human connection with patients | |
| Human connection with peers | We have ended up in a situation where partners and salaried GPs are pitted against each other, with the progression of one group not in the best interests of the other. |
| Human connection across other organisations (e.g., secondary care) | |
| Learning systems (e.g., competence, capability, mastery, socially situated) | |
| Additional useful information | |
| Summary of findings | This short commentary emphasises how partnership is a source of joy and meaning in general practice, as it provides autonomy and a commitment to one's local population. However, it explains that partnership is current unsustainable due to the lack of funding of primary care and because GPs are not trained in management. From the perspective of our review, these factors may lead to less GPs opting for this potentially meaningful career route. |
| Strengths, limitations, and future research directions | |

| | |
|---|---|
| Paper Number | 56 |
| Author | Owen et al |
| Year | 2019 |
| Title | GP retention in the UK: a worsening crisis. Findings from a cross-sectional survey |
| Journal | BMJ Open |
| Region (England, Scotland, Wales, and Northern Ireland) | England |
| Study methodology | Online questionnaire survey with qualitative and quantitative questions. Responses to the qualitative questions were subsequently analysed thematically. |
| Type of document | Primary research |
| Topic or focus of paper | GP retention |
| Research objectives and focus | To investigate how recent national policy-led workforce interventions are affecting intentions to remain working as a general practitioner (GP) |
| Theoretical framework/theories/ underpinnings | |
| Description of intervention | |
| Rationale for paper/review | The general practice workforce in England has been recognised as being at crisis point for several years. Despite a government commitment in 2015 to create 5000 additional general practitioner (GP) posts by 2020,4 recent figures suggest that a further deficit of 1300 full-time equivalent GPs has developed. |
| What creates meaning that sustains GPs engagement with, and involvement in, work? | GPs were asked to rate on a Likert scale (1=not important, 5=very important) factors that might be contributing to their intention to leave GP and remain working in GP. Factors influencing decision to leave were: intensity of workload (mean =4.4), volume of workload, (4.3) Too much time spent on unimportant tasks (4.0), Lack of time for patient contact (3.8), Potential introduction of 7 days a week working (3.8), Reduced job satisfaction (3.6), Poor flexibility of hours (2.8), Revalidation (2.6), Age (3.5), Medical indemnity payments (3.4), Increased risk of litigation (3.0), Changes to pension taxation (2.7), Family commitments (2.6), Ill health (1.8), Embarking on career outside general practice (1.6) and planned career break (1.4). Factors that might retain GPs within general practice were: Reduced intensity of workload (4.1), Longer appointment times/more time to spend with patients (4.0), Reduced volume of workload (3.9), Less administration (3.9), No out of hours commitments (3.6), Incentive payment (3.5), Protected time for education and training (3.3), More flexible working conditions (3.2), Greater clinical autonomy (3.0), Increased pay (2.9), Improved skill mix in the practice (2.8), Additional annual leave (2.8), Shorter practice opening times (2.7), Opportunity for a sabbatical (2.6), Introduction of ‘Twenty Plus’ (2.3), Expansion of GP retainer scheme (2.1), Extended interests; for example, Clinical Commissioning Group (CCG) role (2.0), Reintroduction of the flexible careers scheme (2.0), Option to work term time only (1.6). |

| | |
|---|---|
| | Thematic analysis of the responses to the open question ‘What is the greatest problem within general practice at the current time?’ yielded five key themes: increasing demands, expectations and complexity of patients; workload; GP recruitment and retention; inadequate funding; and bureaucratic and administrative burden.<br><br>Respondents were asked to rate whether they thought about the nationally led initiatives that had been recently introduced to address workforce issues in general practice, specifically whether the initiatives would have a positive, negative or no impact. A net rating was calculated by subtracting the percentage who rated the initiative as negative from the percentage who rated it as a positive: Investment in practice nursing +91.3%, Closer working with specialists; for example, phone and email advice lines +85.3%, Investment in technology +85.3%, Expansion of GP workforce +76.1%, Streamlining CQC, reduced inspection for good and outstanding practices +73.1%, Investment in primary care infrastructure +70.3%, Releasing time for patients +60.6%, Releasing time for patients +60.6%, Paramedics in primary care +44.5%, Practice resilience programme +41.2%, Multispecialty community provider projects +25.3%, Federation of GP practices +19.3%, Better care fund +13.2%, Physicians associates −0.2%, Local sustainability and transformation plans −21.3%, Video and e-consultations −26.6%. Free-text comments suggested that these initiatives were a distraction from the need for significant investment in general practice and tackling key issues affecting the workforce. An additional theme suggested that some initiatives could be further undermining GP morale: “I object to the term 'resilience' and any resources invested into it. We should be focusing all our intentions on making the job better rather than coaching GPs to be more robust against the stress. The very term makes it sound to me like it is somehow the GPs fault in the first place for not coping with the stains and demands of the job.” |
| What solutions/recommendations are proposed? | Responses to the open question ‘What intervention would help general practice the most?’ revealed eight themes: greater funding, more GPs, educate patients and the public (making patients aware of the costs and limitations of primary care, in addition to lessons on how to manage their own health), increase clinical and support staff (which might allow GPs to focus on complex issues that they are trained to deal with), reduce bureaucracy and administration, more time per patient (but without increased working hours or reduced remuneration), protection from financial risk (government or CCG paying indemnity), enhanced public image reputation of general practice. |
| From which stakeholders’ perspective? (General practitioners, patients, other HCPs) | GPs |
| How is work-related joy constructed? (Diversity of ways individuals may think about joy) | |
| What prevents/removes work-related joy? | Comparing current workload with 2 years previously, 51.0% (470) reported working longer hours with almost all (94.4%; 423) giving increased workload as the predominant reason; 26.6% had reduced their hours of work, with most (72.3%; 172) stating this was due to increasing intensity of workload and for many (29.8%; 71) it was related to stress and mental health. This contrasts with the intentions stated in the previous survey where 21.5% of GPs wished to reduce their hours worked and only 2.7% wished to increase.<br><br>The complexity of patients seen by GPs is increasing, alongside increasing patient demands and expectations driven by the media and government. However, there are insufficient health service resources to deal with this. |

| | |
|---|---|
| | Many GPs perceived inadequate funding to provide the staff and services to meet patient's needs – which might lead to guilt and stress.<br><br>"Excessive bureaucracy that is, CQC, CCG, NHS England, appraisal. We are grossly over managed, this prevents us seeing patients or developing services for our patients and employs an army of managers (some clinical)" |
| Organisational characteristics (small, large) | |
| Managing ill-defined illnesses and chronic complex multimorbidity | |
| Human connection with patients | |
| Human connection with peers | |
| Human connection across other organisations (e.g., secondary care) | |
| Learning systems (e.g., competence, capability, mastery, socially situated) | |
| Additional useful information | Morale was reported as having reduced over the past 2 years for 59.4% (510) of respondents and increased for 14.1%.<br><br>When asked to think about their career plans compared with 2 years ago, 409 (48.5%) said they had brought forward their plans to leave GP, with just 47 (5.6%) planning to remain longer. Intention to leave/retire in the next 2 years has increased from 13% in 2014 to 18% (p=0.02), while 63.9% reported an intention to continue working for at least the next 5 years in 2014 compared with only 48.5% in 2017. |
| Summary of findings | The survey in this study demonstrates that the workforce crisis in general practice is worsening, with half of GPs surveyed reporting bringing forward their plans to leave GP. The main contributing factors to this are increasing demands, expectations and complexity of patients; workload; GP recruitment and retention; inadequate funding; and bureaucratic and administrative burden. GPs suggested increased funding, more GPs, better education of the public and expanding non-clinical and support staff as interventions to improve GP retention. National initiatives that aligned with these priorities, such as funding to expand practice nursing were viewed positively, but low numbers of GPs had seen evidence of their roll-out. Conversely, national initiatives that did not align, such as video consulting, were viewed negatively. |
| Strengths, limitations, and future research directions | This is the first survey to report general practitioners' (GPs) views and experience of national initiatives which have been introduced in England to address the workforce crisis in general practice.<br><br>The survey was conducted in the same region as a similar survey in 2014, so allowing some analysis of how views are changing over time.<br><br>The free-text qualitative data added depth to the findings. |

| | |
|---|---|
| Paper Number | 57 |
| Author | Peckham et al |
| Year | 2016 |
| Title | General practitioner recruitment and retention: An evidence synthesis |
| Journal | N/A (report commissioned by NHS England and DHSC) |
| Region (England, Scotland, Wales, and Northern Ireland) | N/A (included papers were mainly relevant to the UK although selected international studies were also included) |
| Study methodology | Literature search to identify studies relating to GP recruitment and retention, followed by evidence synthesis to summarise the key findings of these papers relating to elements of the 10 Point Plan. |
| Type of document | Secondary research |
| Topic or focus of paper | GP recruitment and retention |
| Research objectives and focus | We conducted a synthesis of the research evidence and wider literature on the key objectives of the 10 Point Plan (see 'rationale' below) and the use of incentives to identify:<br>1. Evidence to support the specific actions set out in the plan.<br>2. Evidence on other approaches to retention and recruitment of GPs |
| Theoretical framework/theories/ underpinnings | |
| Description of intervention | |
| Rationale for paper/review | In January 2015 NHS England published the 10 Point Plan to support the recruitment, retention and re-employment of GPs. In order to support a review of the 10 Point Plan in 2016, NHS England and the Department of Health commissioned the Policy Research Unit in Commissioning and the Healthcare System (PRUComm) to undertake an evidence synthesis on GP recruitment, retention and re-employment. |
| What creates meaning that sustains GPs engagement with, and involvement in, work? | Landry et al (2011) found that training doctors locally increased the odds of their retention, from which the authors of this report inferred that socialisation of junior doctors to the primary care practice and more precisely to the general practice could have an important effect on retention since they would have a *realistic view* of the profession, its challenges and implications.<br><br>Humphreys et al's (2001) review of the rural medical workforce identified three main factors pertinent to retention and turnover: professional issues, social factors, and external contextual factors. Professional issues are related to the work content, vocational satisfaction, |

| | |
|---|---|
| | support and remuneration. The social factors are related to personal characteristics and family circumstances. Lastly, the external factors are related to the community and its geographical location. While these factors might be important for rural practices, they may also influence GPs retention of all contexts.<br><br>Flexibility and part-time working have always been seen as factors that make general practice a more attractive working environment although this is increasingly seen to be less relevant (CFWI 2014, Evans et al 2000, Wordsworth et al 2004).<br><br>Van Ham et al (2006) suggest that job satisfaction is influenced by job autonomy (including work diversity and variety), social support (including relationship and collaborative partnerships with colleagues and patients), and the practice environment such as whether or not it is linked to academic hospitals or centres, and whether there is the opportunity to teach medical students and advanced students. The factors influencing dissatisfaction are: number of working hours, compensation and income, workload, high work demands, lack of support or colleagues, lack of professional recognition, and increased bureaucracy and practice administration.<br><br>Job satisfaction is also related to Humphreys et al's identification of the factors that lead doctors to remain in rural practice although professional satisfaction (variety of work, autonomy of practice, and a feeling of doing an important job) were identified as the main reasons.<br><br>Groenewegen & Hutten (1991) have highlighted some sources of stress that have an effect on satisfaction. These include the interruption of daily routine, emotional involvement, administrative workload, and routine work.<br><br>Dale et al (2015) found that the work-related factors most likely to influence intention to leave are the intensity of workload, volume of workload, the time spent on unimportant tasks, introduction of the seven-day working week and job satisfaction. As for the non-work-related factors, they are the work-life flexibility and personal development. |
| What solutions/recommendations are proposed? | The elements that most likely to increase and influence retention in general practice include:<br>1. Supporting intrinsic factors of the job<br>2. Strategies to improve job satisfaction<br>3. Reducing job stressors such as work overload; lack of support and high demands increases the likelihood of quitting the practice and/or profession. |
| From which stakeholders' perspective? (General practitioners, patients, other HCPs) | N/A |
| How is work-related joy constructed? (Diversity of ways individuals may think about joy) | At a more micro level, GP accessibility to sub-specialisation and portfolio careers could have an effect on retention. It is also suggested that increased learning (satisfaction of intellectual and altruistic needs) and functional flexibility (adaptable and transferable within their practice) could improve satisfaction, morale and fulfilment and, as a result, GP retention. |

| | |
|---|---|
| | Positive job attributes of general practice that declined in general practice from 2012 – 2015 included: choice in deciding how to do job, choice in deciding what to do at work, involvement in deciding changes that affect work, flexibility of working time, interesting variety of the job, and being consulted about changes that affect work.<br><br>GPs completing the survey seemed to be satisfied with colleagues and fellow workers, physical working conditions, the amount of variety in the job. |
| What prevents/removes work-related joy? | Young and Leese (1999) suggested that reducing the income differential between general practice and hospital work could increase retention. More precisely, appropriate remuneration schemes could have a key role in GP recruitment and retention but the current arrangements are inadequate to compensate for the increasing workload. In fact, while low pay might be a source of dissatisfaction toward the job, the evidence suggests that increases in income or salary would not compensate for other sources of job dissatisfaction such as workload (Sibbald et al 2000). Similarly, when interviewing older GPs about retainer schemes, many GPs commented that money was not an issue but workload was.<br><br>The job stressors with the highest scores are: increased workload, changes to meet requirements of external bodies, having insufficient time to do the job justice, paperwork, increased demand from patients and long working hours. Key job stressors that increased between 2012 and 2015 are: adverse publicity by the media, finding a locum, changes imposed from the primary care organisation, and insufficient resources within the practice.<br><br>GPs completing the survey seemed to be satisfied with colleagues and fellow workers, physical working conditions, the amount of variety in the job<br><br>Dale et al (2015) included a qualitative component where they asked their respondent to answer freely on factors contributing to work-related pressures. Responses included the growth in patient expectations and demand, recruitment and retention difficulties, burgeoning administration and bureaucracy, growth in additional roles, responsibilities and meetings, transfer of work from secondary care, increasing complexity and chronic ill health, and the introduction of seven-day working in general practice. They also found an unexpected theme: the emotional impact of working as a GP. In fact, respondents felt stressed, exhausted, disillusioned, frustrated, burnt out, and overwhelmed and mentioned that the constant negative portrayal of GPs in the media and by the government was affecting their spirit and professional identity.<br><br>A study of the relationship between job strain, blood pressure and heart rate found physiological signs of stress during non-working days and nights, indicating “a failure to relax following the termination of demands”. |
| Organisational characteristics (small, large) | Groenewegen & Hutten (1991) suggest a relationship between the workload (consequences of the list size and practice composition) and job satisfaction. |
| Managing ill-defined illnesses and chronic complex multimorbidity | While the job provides a variety of interesting things, GPs report that they do not have time to carry out all work and are required to do unimportant tasks, preventing them from completing more important ones. They also report having to work very fast without any clear feedback about how well they are doing their job. These negative job attributes increased from 2012 – 2015. |
| Human connection with patients | Wordsworth et al (2004) suggest that enhancing patient care aspects of GPs work is most likely to provide a positive view of practice and act as a key for retention. |

| | |
|---|---|
| Human connection with peers | Mentorship schemes would be welcome at every stage during GPs' careers.<br><br>Saxén et al (2008) showed that the shortage of physicians had very little negative effects on health care staff job satisfaction, and explained their results suggesting that shortage could increase solidarity and "... force the staff to work together more closely to get the work done." |
| Human connection across other organisations (e.g., secondary care) | |
| Learning systems (e.g., competence, capability, mastery, socially situated) | |
| Additional useful information | The NHS England 10 Point Plan identified four strategies to increase retention in general practice: 1) Investment in retainer schemes 2) Improving the training capacity in general practice 3) Incentives to remain in practice 4) New ways of working.<br><br>Some of the reported studies use job satisfaction as a synonym for retention and job dissatisfaction as a synonym for turnover despite the fact that the wider literature shows that both job satisfaction and job dissatisfaction are variables that influence the retention – turnover process but are distinct from it. |
| Summary of findings | This review flagged a multitude of factors relevant to GP retention. Positive factors as viewed by students and GPs about general practice as a profession – such as patient contact, variety, continuity of care – are intrinsic to what it means for them to be a GP. Feeley (2003) has however highlighted the importance of expectation versus reality. What junior doctors expected when they became a GP and the real life of a GP is likely to impact on retention. The evidence does suggest that tackling key aspects of job stress are important but supporting the key factors of how GPs view the essential nature of general practice in terms of patient contact may be critical alongside developing new opportunities for diversity of practice through sub-specialities and broader portfolio careers. As for the new ways of working, it is likely that the inclusion of nurses, pharmacists, and even social workers might help reduce the strain of the workload and burnout symptoms of GPs. GPs leave both for reasons of job dissatisfaction – possibly reflecting a frustration or a disappointment toward the changing roles in their practice - and also to retire before 60 years old, even if not discontent. Reasons may include lack of resilience to deal with stress but also a simple view that they have undertaken sufficient lifetime service.<br><br>I did not extract data relating to recruitment or from other countries as not relevant to our study. |
| Strengths, limitations, and future research directions | |

| | |
|---|---|
| Paper Number | 58 |
| Author | Sharon Spooner |
| Year | 2016 |

| | |
|---|---|
| Title | Unfashionable tales: narratives about what is (still) great in NHS general practice |
| Journal | BJGP |
| Region (England, Scotland, Wales, and Northern Ireland) | UK (does not say specific region) |
| Study methodology | Interviews with 8 senior GPs, followed by thematic analysis of responses |
| Type of document | Primary research |
| Topic or focus of paper | Meaning in general practice |
| Research objectives and focus | To understand which aspects of work continue to motivate and engage senior GPs by exploring their narrative accounts. |
| Theoretical framework/theories/ underpinnings | |
| Description of intervention | |
| Rationale for paper/review | There is clear evidence that general practice has become a less popular career choice and among GPs there are high levels of dissatisfaction and demotivation. Little empirical evidence has emerged to indicate which factors contribute intrinsic value to the working lives of GPs and sustain their ongoing commitment. |
| What creates meaning that sustains GPs engagement with, and involvement in, work? | |
| What solutions/recommendations are proposed? | |
| From which stakeholders' perspective? (General practitioners, patients, other HCPs) | GPs |
| How is work-related joy constructed? (Diversity of ways individuals may think about joy) | Drawing on their experienced early on in their training, when they worked in hospitals, many GPs regarded general practice as a more comfortable place to work, with greater flexibility and a more appealing lifestyle, an escape from the demands and discomforts of hospital on-call work where, as junior doctors, they could feel like 'just a tool in a machine' and caught in a competitive environment with 'challenging, hierarchical people'.<br><br>In response to initiatives to reduce prescribing costs and informed by regular monitoring updates, GPs spoke of achieving satisfaction by successfully maintaining a good budgetary position when compared with neighbouring practices. |

| | |
|---|---|
| What prevents/removes work-related joy? | |
| Organisational characteristics (small, large) | |
| Managing ill-defined illnesses and chronic complex multimorbidity | |
| Human connection with patients | Long-term engagement with patients was highly valued: “I saw a patient today who … was already struggling as a teenager when I first met her 22 years ago … psychiatry has bounced around saying she is just personality disorder and so … impossible to treat, and I have seen her through my whole career … she comes back time and time and time and time again. It's got to be a privilege hasn’t it because you know that’s something that no one else will have with that person, and she obviously respects me”<br><br>One GP noted, that for some patients, he could be the “second most important person in their life” |
| Human connection with peers | Team building through finding time for interpersonal communication was highly valued. Making sure that all GPs shared a coffee break provided ‘very supportive’ opportunities to allocate tasks, discuss any problems, and efficiently decide on any necessary action. They could also pass on information, talk through complex cases, or simply consolidate a sense of working together. |
| Human connection across other organisations (e.g., secondary care) | In addition to teams within their practices, GPs were members of other teams, networks, and forums through involvement with undergraduate education and postgraduate training roles. These could be particularly stimulating: “The teaching gives me a high, it’s just being with young students and going back to basics again ... teaching examination, looking at interesting patients, so it’s exactly back to basic clinical skills.” |
| Learning systems (e.g., competence, capability, mastery, socially situated) | |
| Additional useful information | |
| Summary of findings | This paper adds a counter-perspective to many articles, papers, and reports that focus on negative aspects of GPs’ working lives. It talks about how the 8 senior GPs interviewed find meaning in responding to patients’ needs and offering long term support, in teaching medical students and junior colleagues, working alongside peers both in practices and other organisations, and adapting to (and thriving under) new regulatory mechanisms.<br><br>The paper concludes with the line: “The nuanced narratives of these GPs provide evidence to confirm that, however unfashionable it is to say so, general practice can still be a great place to work.” |
| Strengths, limitations, and future research directions | It must be acknowledged that a limited study such as this cannot encompass all opinions or convey how widely held each position may be across this or another group of GPs. Further, since all had continued in practice for 25 years after graduation they had, by definition, achieved some success in negotiating workplace changes. |

| | |
|---|---|
| Paper Number | 59 |
| Author | Abrahamson et al |
| Year | 2020 |
| Title | An evaluation of the clinical microsystems approach in general practice quality improvement |
| Journal | Primary Health Care Research & Development |
| Region (England, Scotland, Wales, and Northern Ireland) | England |
| Study methodology | First, a review of all CCG clinical microsystems projects using pre-existing data. Subsequently, interviews and focus groups with stakeholders involved in implementing the clinical microsystems approach in GP practices. |
| Type of document | Primary research |
| Topic or focus of paper | Clinical microsystems in general practice |
| Research objectives and focus | To explore experiences of GP staff participating in a clinical microsystems programme. |
| Theoretical framework/theories/ underpinnings | A clinical microsystem is defined as a ‘small group of people who work together on a regular basis to provide care to discrete subpopulation of patients’. It is a functioning unit which produces services that can be measured as performance outcomes. GPs are distinct clinical practice units with a designated purpose and function, fitting this definition well. The CMS process involves identifying a problem and developing a systematic approach to address it using a ‘toolkit’ of activities and a workbook to lead the team through a process. |
| Description of intervention | |
| Rationale for paper/review | Changes to the general practice (GP) contract in England (April 2019) introduced a new quality improvement (QI) domain. The clinical microsystems programme is an approach to QI with limited evidence in primary care. |
| What creates meaning that sustains GPs engagement with, and involvement in, work? | In order to facilitate continued participation with CMS, practice staff needed to feel that they were in control of the agenda. Conversely, participation was limited if staff felt ‘coerced’ into accepting the programme or had prior unsuccessful attempts at solving the corresponding problem. This might relate to a general point about the importance of co-designing policy interventions with GPs. |
| What solutions/recommendations are proposed? | |
| From which stakeholders’ perspective? (General practitioners, patients, other HCPs) | 11 coaches and 16 practice staff (including 10 GPs) involved in implementing the clinical microsystems approach within GP practices. |

| | |
|---|---|
| How is work-related joy constructed? (Diversity of ways individuals may think about joy) | Compensating practices for their time spent participating in the CMS program was seen as essential for continued engagement. This may relate to a general point about adequately resourcing and financing GPs for ‘extra’ projects. |
| What prevents/removes work-related joy? | |
| Organisational characteristics (small, large) | |
| Managing ill-defined illnesses and chronic complex multimorbidity | |
| Human connection with patients | |
| Human connection with peers | Where there was discord, the CMS approach was regarded as an opportunity to redress internal politics and rebuild fragmented relationships: “The last practice manager had gone, there’d been a period with … no real leadership and the staff team was very fragmented … this was a chance to come together and work on something as a group”<br><br>The CMS approach appeared to benefit relationships within the practice, challenged hierarchies and allowed frontline staff to feel valued and listened to. |
| Human connection across other organisations (e.g., secondary care) | Successful projects helped foster positive working relationships between the CCG and the practice and this supported ongoing and mutually beneficial communication. |
| Learning systems (e.g., competence, capability, mastery, socially situated) | |
| Additional useful information | |
| Summary of findings | Mainly, this paper evaluates the efficacy of the CMS approach to quality improvement in English general practices. Some points are relevant to our review: the importance of staff feeling in control of practice/policy changes, the importance of compensating practices for time spent on their CMS project, and the CMS approach improving working relationships within practices and externally with the CCG. |
| Strengths, limitations, and future research directions | This was a small evaluation, and the main study limitation was recruiting sufficient GPs particularly those who had not participated or withdrew. It was difficult to engage more than one person per practice which limited the range of perspectives represented in the data and may have resulted in selection bias. |

| | |
|---|---|
| Paper Number | 60 |
| Author | Allen et al |
| Year | 2017 |
| Title | Does the proportion of pay linked to performance affect the job satisfaction of general practitioners? |
| Journal | Social Science & Medicine |
| Region (England, Scotland, Wales, and Northern Ireland) | UK |
| Study methodology | GPs surveyed on job satisfaction and related variables (see below) before and after the introduction of the QOF. Exposure of these GPs to P4P then calculated, and a continuous difference-in-differences model used to assess the impact of this job satisfaction variables. |
| Type of document | Primary research |
| Topic or focus of paper | Pay-for-performance |
| Research objectives and focus | Examine whether changes in GPs' job satisfaction before and after the introduction of the QOF in 2004 were correlated with the proportion of their income that became exposed to P4P. |
| Theoretical framework/theories/ underpinnings | |
| Description of intervention | |
| Rationale for paper/review | There is concern that pay-for-performance (P4P) can negatively affect general practitioners (GPs) by reducing their autonomy, increasing their wage dispersion or eroding their intrinsic motivation. This is especially a concern for the Quality and Outcomes Framework (QOF), a highly powered P4P scheme for UK GPs. |
| What creates meaning that sustains GPs engagement with, and involvement in, work? | This study found no significant effects of P4P exposure on overall job satisfaction or 12 additional measures of working lives in either the short or longer term:<br>1. Working hours<br>2. Intentions to quit<br>3. Life satisfaction<br>4. Satisfaction with physical working conditions<br>5. Satisfaction with freedom to choose own method of working<br>6. Satisfaction with colleagues & fellow workers<br>7. Satisfaction with recognition you get for good work<br>8. Satisfaction with amount of responsibility you are given |

| | |
|---|---|
| | 9. Satisfaction with remuneration<br>10. Satisfaction with opportunity to use abilities<br>11. Satisfaction with hours of work<br>12. Satisfaction with amount of variety in job |
| What solutions/recommendations are proposed? | |
| From which stakeholders' perspective? (General practitioners, patients, other HCPs) | GPs. |
| How is work-related joy constructed? (Diversity of ways individuals may think about joy) | |
| What prevents/removes work-related joy? | |
| Organisational characteristics (small, large) | |
| Managing ill-defined illnesses and chronic complex multimorbidity | |
| Human connection with patients | |
| Human connection with peers | |
| Human connection across other organisations (e.g., secondary care) | |
| Learning systems (e.g., competence, capability, mastery, socially situated) | |
| Additional useful information | |

| | |
|---|---|
| Summary of findings | This quantitative study finds that P4P has no effect on GPs' job satisfaction (provided overall income remains the same). |
| Strengths, limitations, and future research directions | Strengths: We have a large sample of GPs who could not self-select into or out of P4P. We have a survey which asks a range of questions about job satisfaction and working lives. This survey is linked to administrative data to create a unique linked dataset which has enabled us to measure the amount of P4P exposure for each GP in our sample. We employed an econometric method which estimates the effect of increased P4P exposure. In addition to job satisfaction, we also model the effects of P4P exposure on intentions to quit, hours worked, life satisfaction and nine sub-domains of jobs satisfaction. We analysed the immediate effect one year after the introduction of P4P and the effects four years after P4P was introduced.<br><br>Limitations: We had to estimate P4P exposure based on two assumptions. Firstly, that the sharing of QOF income within a practice is related only to a GP's FTE. This assumption is likely to oversimplify a complicated set of income sharing rules. Our second assumption is that our predictions of GP income had the QOF not been introduced are accurate. |

| | |
|---|---|
| Paper Number | 61 |
| Author | Barker et al |
| Year | 2021 |
| Title | Future-proofing the primary care workforce: A qualitative study of home visits by emergency care practitioners in the UK |
| Journal | European Journal of General Practice |
| Region (England, Scotland, Wales, and Northern Ireland) | England |
| Study methodology | Three practices in England piloted an ECP (extended-scope practitioners with a paramedic or nursing background) home visiting service. Following the pilot, focus groups were conducted with each of the three primary healthcare teams and one with ECPs, alongside nine individual patient interviews. Data were analysed using a modified framework approach. |
| Type of document | Primary research |
| Topic or focus of paper | Multidisciplinary team in general practice |
| Research objectives and focus | This study sought to explore patients' and professionals' experiences of a primary care home visiting service delivered by emergency care practitioners (ECPs), in place of GPs; to determine positive impacts/unintended consequences and establish whether interdisciplinary working was achieved. |
| Theoretical framework/theories/ underpinnings | |
| Description of intervention | |

| | |
|---|---|
| Rationale for paper/review | Broadening the skill-mix in general practice is advocated to build resilience into the primary care workforce. However, there is little understanding of how extended-scope practitioners from different disciplines, such as paramedicine and nursing, embed into roles traditionally ascribed to general practitioners (GPs). |
| What creates meaning that sustains GPs engagement with, and involvement in, work? | |
| What solutions/recommendations are proposed? | All interviewees supported a wider roll out of the ECP home visiting pilot. |
| From which stakeholders' perspective? (General practitioners, patients, other HCPs) | Stakeholders (including GPs) involved in implementing an ECP home visiting service. |
| How is work-related joy constructed? (Diversity of ways individuals may think about joy) | ECPs were perceived to reduce GPs' workload: "Initially obviously we were really positive about it because [ ... ] we get a lot of home visits and anything that could be done to try and reduce that workload" |
| What prevents/removes work-related joy? | Initially, some practice staff had reservations about ECPs, thinking patients would prefer 'traditional' GPs and that ECPs might have a narrower skillset than GPs. In particular, GPs felt they were particularly capable in managing medical complexity and feared that ECPs might be more inclined to admit patients to hospital. However, initial fears and preconceptions of GPs about the capabilities of the ECPs were resolved throughout the pilot: "It was apparent – the quality of assessments people were getting [from the ECPs] was really high– and the judgements, it all sounded appropriate". |
| Organisational characteristics (small, large) | |
| Managing ill-defined illnesses and chronic complex multimorbidity | |
| Human connection with patients | Collaboration between GPs and ECPs was noted to contribute towards maintaining continuity of care. |
| Human connection with peers | |
| Human connection across other organisations (e.g., secondary care) | |

| | |
|---|---|
| Learning systems (e.g., competence, capability, mastery, socially situated) | |
| Additional useful information | |
| Summary of findings | This study evaluated the views of a range of stakeholders on the implementation of an ECP scheme in three GP practices. A few findings are relevant to our review. Firstly, although GPs were initially apprehensive about ECPs due to concerns about competence and how patients would react, these fears were quickly alleyed when they began working with ECPs. Secondly, as emphasised by other studies, ECPs had a positive effect on GPs' workload. |
| Strengths, limitations, and future research directions | The findings presented in this study relate to three neighbouring practices in northern England so the applicability of findings to different primary care settings may vary |

| | |
|---|---|
| Paper Number | 62 |
| Author | Barry & Greenhalgh |
| Year | 2019 |
| Title | General practice in UK newspapers: an empirical analysis of over 400 articles |
| Journal | BJGP |
| Region (England, Scotland, Wales, and Northern Ireland) | UK |
| Study methodology | A thematic analysis of all newspaper articles mentioning GPs or general practice published in the UK from late October 2016 to early October 2017 was undertaken, along with a sample of articles on hospital medicine. |
| Type of document | Primary research |
| Topic or focus of paper | General practice in the media |
| Research objectives and focus | To explore how general practice and GPs are depicted in UK national newspapers. |
| Theoretical framework/theories/ underpinnings | |
| Description of intervention | |
| Rationale for paper/review | In the context of the biggest GP workforce crisis since the NHS began, the alleged negative portrayal of UK general practice in the media is often cited as a reason for falling recruitment. |

| | |
|---|---|
| What creates meaning that sustains GPs engagement with, and involvement in, work? | |
| What solutions/recommendations are proposed? | Efforts to influence the media to provide a more balanced perspective of general practice should continue. |
| From which stakeholders' perspective? (General practitioners, patients, other HCPs) | Newspapers – Thematic analysis conducted by GPs |
| How is work-related joy constructed? (Diversity of ways individuals may think about joy) | |
| What prevents/removes work-related joy? | GPs were portrayed very negatively in the media, across five main themes:<br>1. The workforce crisis: Newspapers noted that older GPs were able to retire early due to high salaries that had 'filled their pension pots', whilst younger GPs opting for part-time positions were depicted as lazier than their older colleagues.<br>2. Patient access to general practice: Many newspapers reported GP practices closing down, and this was sometimes framed in the context of GPs' lack of commitment or professionalism.<br>3. Organisation of care: GPs were typically depicted as seeking to personally profit from new, privatised (or part-privatised) service models. Articles describing the reassignment of professional tasks from GPs to other health professionals tended to depict these changes negatively.<br>4. GPs' poor clinical performance: Only one article described a prompt diagnosis by a GP, which saved the patient's life; the remainder constructed an overriding narrative of clinical failure characterised by missed diagnoses (particularly cancer), incorrect prescriptions, failure to prevent suicide, and refusal to treat patients who subsequently die<br>5. GPs' personal vices (and absence of virtues): One in seven stories in the sample of articles on general practice covered GPs' alleged vices (negative character traits); virtues were not a focus. Such stories included GPs perpetrating acts that were both illegal and unprofessional, including domestic violence, sexual harassment, assault, fraud, murder, and conducting sexual relationships with patients. |

| | |
|---|---|
| | Furthermore, there was a striking lack of balance in the news articles, with a near-absence of positive portrayals of the specialty and no articles examining the effect of government economic policies on general practice or offering positive coverage of extended-hours surgeries or 7-day access.<br><br>Interestingly, in equivalent articles on hospital specialties presented doctors very positively, with pressures and recent negative changes depicted as being the fault of the government. |
| Organisational characteristics (small, large) | |
| Managing ill-defined illnesses and chronic complex multimorbidity | |
| Human connection with patients | |
| Human connection with peers | |
| Human connection across other organisations (e.g., secondary care) | |
| Learning systems (e.g., competence, capability, mastery, socially situated) | |
| Additional useful information | |
| Summary of findings | This thematic analysis of newspaper articles mentioning GPs explicitly demonstrates that GPs were depicted in an overwhelmingly negative manner by the media, across 5 themes: workforce, patient access, care organisation, clinical performance and personal vices. In contrast, in equivalent articles, hospital specialties were presented very positively. In the context of our review, this reinforces the findings of other studies which report that negative media portrayals erode meaning for GPs. |
| Strengths, limitations, and future research directions | This study was novel in that it directly and systematically analysed a corpus of over 400 news articles on general practice spanning a 12-month period. Those articles were compared with a smaller (but still extensive) sample of 100 articles on hospital medicine, allowing a novel comparison to be made of the language used to describe similar scenarios in different settings. |

| | |
|---|---|
| Paper Number | 63 |
| Author | Blane et al |

| | |
|---|---|
| Year | 2017 |
| Title | A Change Model for GPs Serving Deprived Areas |
| Journal | Annals of Family Medicine |
| Region (England, Scotland, Wales, and Northern Ireland) | Scotland |
| Study methodology | Implementation of the Deep End Pioneer Scheme (see below). |
| Type of document | Primary research |
| Topic or focus of paper | Professional and service development |
| Research objectives and focus | Describe the Deep End Pioneer scheme and report earliest results |
| Theoretical framework/theories/ underpinnings | |
| Description of intervention | Fellows participating in the Deep End Pioneer Scheme work 4 days a week, with 3 days in practice and 1 day of protected time to be used in alternate weeks for service development work (collaborating with the lead GP for the practice) and professional development, via a program of day-release sessions with a curriculum based on identified learning needs for Deep End practitioners.1 An academic coordinator organizes these day-release sessions and supports the fellows in producing written outputs for wider dissemination. The lead GPs for each practice meet every 6 to 8 weeks and share documents online |
| Rationale for paper/review | In 2016, The Scottish Government launched a GP recruitment and retention fund in response to the "GP crisis" brought about by increasing demands on general practice (aging population, more complex multimorbidity) and difficulties recruiting and retaining GPs, particularly in rural areas and areas of high socioeconomic deprivation. |
| What creates meaning that sustains GPs engagement with, and involvement in, work? | The Pioneer scheme is still at an early stage, but the host GPs involved already report a decrease in stress and renewed enthusiasm for their work. |
| What solutions/recommendations are proposed? | |
| From which stakeholders' perspective? (General practitioners, patients, other HCPs) | GPs |
| How is work-related joy constructed? (Diversity of | |

| | |
|---|---|
| ways individuals may think about joy) | |
| What prevents/removes work-related joy? | |
| Organisational characteristics (small, large) | |
| Managing ill-defined illnesses and chronic complex multimorbidity | |
| Human connection with patients | |
| Human connection with peers | |
| Human connection across other organisations (e.g., secondary care) | |
| Learning systems (e.g., competence, capability, mastery, socially situated) | |
| Additional useful information | |
| Summary of findings | This very short paper describes how a scheme that provides protected time for GPs' professional and service development has decreased stress and increased enthusiasm for GPs involved. |
| Strengths, limitations, and future research directions | This paper is not a rigorous evaluation of the scheme – therefore does not explain on how they have assessed GPs' experience of the scheme (indeed, the findings could just be what the authors have informally heard). |

| | |
|---|---|
| Paper Number | 64 |
| Author | Blane & Basu |
| Year | 2023 |
| Title | Tackling climate change and health inequalities in primary care |
| Journal | Family Practice |

| | |
|---|---|
| Region (England, Scotland, Wales, and Northern Ireland) | Scotland |
| Study methodology | A group of 10 GP colleagues from a variety of Deep End settings (which have the greatest percentage of registered patients living in the most socioeconomically deprived postcodes in Scotland) contributed to an online roundtable event to explore climate change in the context of health inequalities. This report was a brief summary of the discussion. |
| Type of document | Primary research |
| Topic or focus of paper | Climate change |
| Research objectives and focus | Explore the impacts of climate change on Deep End Patients and practices in particular, but also discuss the urgent need for system-wide solutions to tackle this burden. |
| Theoretical framework/theories/ underpinnings | |
| Description of intervention | |
| Rationale for paper/review | The Climate Emergency is now widely accepted as the biggest public health crisis facing humanity. Previous research has highlighted how social and health inequalities shape the health impacts of climate change in the UK, but there has been little attention to the role of general practice in deprived areas. |
| What creates meaning that sustains GPs engagement with, and involvement in, work? | |
| What solutions/recommendations are proposed? | |
| From which stakeholders' perspective? (General practitioners, patients, other HCPs) | GPs |
| How is work-related joy constructed? (Diversity of ways individuals may think about joy) | |
| What prevents/removes work-related joy? | Climate change was highlighted to be worsening health outcomes through a variety of mechanisms: patients are malnourished because of damaging food systems; patients struggle with weight, inactivity, and chronic respiratory and cardiovascular conditions because of unhealthy transport systems and environments; stressed patients use cigarettes, alcohol, drugs, and violence to cope with economic and social |

| | |
|---|---|
| | injustices. This suggests that climate change is contributing to increasing workload for GPs, which other papers have highlighted to be an importance influence on meaning. |
| Organisational characteristics (small, large) | |
| Managing ill-defined illnesses and chronic complex multimorbidity | |
| Human connection with patients | |
| Human connection with peers | |
| Human connection across other organisations (e.g., secondary care) | |
| Learning systems (e.g., competence, capability, mastery, socially situated) | |
| Additional useful information | |
| Summary of findings | This short paper reports how GPs who work in areas with high degrees of socioeconomic deprivation perceive the impact of the climate crisis on their patients – and how GPs can work to mitigate these impacts. The only finding relevant to our review is that the effects of climate change might be contributing to the increasing workload of GPs. |
| Strengths, limitations, and future research directions | |

| | |
|---|---|
| Paper Number | 65 |
| Author | Blease et al |
| Year | 2018 |
| Title | Computerization and the future of primary care: Asurvey of general practitioners in the UK |
| Journal | PLOS One |

| | |
|---|---|
| Region (England, Scotland, Wales, and Northern Ireland) | UK |
| Study methodology | Via an online survey, GPs rated the likelihood of future technology replacing GPs at core six primary care tasks using a Likert scale. Furthermore, if GPs considered replacement for a particular task likely, the survey measured opinions about how many years from now this technological capacity might emerge. A second set of items asked participants about their perceptions of the GP workforce (including patient demand and career satisfaction). |
| Type of document | Primary research |
| Topic or focus of paper | AI in primary care |
| Research objectives and focus | To describe the opinions of British general practitioners regarding the potential of future technology (AI) to replace key tasks carried out in primary care |
| Theoretical framework/theories/ underpinnings | |
| Description of intervention | |
| Rationale for paper/review | Within the medical community there has also been an upsurge in debate about the potential of big data and machine learning to disrupt healthcare professions (including obviate the need for physicians altogether). Despite this, to the authors' knowledge, there is no exploration of the views of practicing physicians on the impact of AI on health professions. |
| What creates meaning that sustains GPs engagement with, and involvement in, work? | |
| What solutions/recommendations are proposed? | |
| From which stakeholders' perspective? (General practitioners, patients, other HCPs) | GPs |
| How is work-related joy constructed? (Diversity of ways individuals may think about joy) | Our data show that the overwhelming majority of UK GPs studied (94%) believed it was unlikely or extremely unlikely that technology will be able to provide empathic care to patients as well as or better than the average GP. In contrast with opinions about other tasks, most GPs were optimistic about the potential for technology to fully replace them in performing tasks related to patient documentation. Eight in ten expected that future technology will be able to undertake this role, and 79% of these respondents estimated that the wherewithal will emerge up to ten years from now. |
| What prevents/removes work-related joy? | Most respondents to this survey (61–68%) considered it unlikely or extremely unlikely that technology will be able to fully replace humans in performing key primary care tasks including diagnosis, referral to other specialists, and the formulation of personalized treatment plans; though, as these figures reflect, a significant minority of participants disagreed with these opinions. GPs were evenly split over prognostics |

| | |
|---|---|
| | with just over half (53%) considering it likely or extremely likely that technology will be able to replace them in performing this task. Nearly one in two of these respondents estimated that the technological capacity will arise in the next decade.<br><br>In the second part of the survey, an overwhelming majority of participants (97%, 701/720) believed that the number of GPs in the UK is much less or somewhat less than patient demand. Nearly 4 in 10 GPs (38%, 272/720) reported that they were somewhat dissatisfied or very dissatisfied with their career, and 38% (269/720) reported burnout. |
| Organisational characteristics (small, large) | |
| Managing ill-defined illnesses and chronic complex multimorbidity | |
| Human connection with patients | |
| Human connection with peers | |
| Human connection across other organisations (e.g., secondary care) | |
| Learning systems (e.g., competence, capability, mastery, socially situated) | |
| Additional useful information | |
| Summary of findings | The survey conducted by this study reveals that, although the majority of GPs consider it unlikely that core aspects of their job will be replaced by AI, a significant minority consider it likely – and of these respondents, one in two believe the technological capacity will arise in the next decade. This might be a source of anxieties about the (near) future for these GPs. However, alongside this, a large majority of GPs surveyed believe that AI will soon be able to undertake administrative tasks for GPs – which might be a source of hope for the future. |
| Strengths, limitations, and future research directions | This is the first survey to investigate physicians' opinions about the future impact of technology on amedical specialty. While the majority of UK GPs are members of Doctors.net.org, a potential limitation of the study isthat the participant response rate depended on individuals being logged into the network. The decision to complete the survey may have been influenced by the extent to which the responder was interested in the topic, and this, in turn, may have affected findings.<br><br>The study investigated GPs' opinions about whether technology will have the *capacity* to fully replace physicians on various tasks and not whether this will, in fact, happen. This latter, distinctive research question is also valuable and worth exploring. |

| | |
|---|---|
| Paper Number | 66 |
| Author | Burn et al |
| Year | 2022 |
| Title | A longitudinal qualitative study of the UK general practice workforce experience of COVID-19 |
| Journal | Primary Health Care Research & Development |
| Region (England, Scotland, Wales, and Northern Ireland) | England and Scotland |
| Study methodology | 17 primary care practitioners and managers submitted narrative accounts in the first year of the COVID-19 pandemic either by writing, self-recording or interview. These were subsequently analysed using a grounded theory approach, with thematic coding used to construct common themes. |
| Type of document | Primary research |
| Topic or focus of paper | General practice during COVID-19 |
| Research objectives and focus | The aim of this project was to describe the changes in the delivery of general practice in the United Kingdom in the first year of the COVID-19 pandemic. Furthermore, to explore primary care practitioners' and managers' experiences of change within general practice during this time and investigate shifts in perceptions of professional identities. |
| Theoretical framework/theories/ underpinnings | |
| Description of intervention | |
| Rationale for paper/review | The COVID-19 pandemic has led to significant reorganisation of general practice in the United Kingdom and around the world. We know very little about how those working in general practice experienced the service reorganisation introduced in the first year of the COVID-19 pandemic. |
| What creates meaning that sustains GPs engagement with, and involvement in, work? | Throughout the study, GPs repeatedly emphasised the importance of feeling they are delivering high-quality care. |
| What solutions/recommendations are proposed? | At the initial stage of the pandemic, when practices were thrust into developing a response to COVID-19, participants spoke of the sense of almost liberation and increased autonomy for practices and this initial increased autonomy appeared to re-energise some participants in our research. Investigating how an increased sense of autonomy could be embedded within training and regulatory frameworks may offer a further avenue to explore how to address burnout and exhaustion within general practice. |
| From which stakeholders' perspective? (General practitioners, patients, other HCPs) | 13 GPs, 2 nurses, 2 practice managers |

| | |
|---|---|
| How is work-related joy constructed? (Diversity of ways individuals may think about joy) | Participants often described an initial sense of liberation and freedom, as the usual bureaucracy was swept away to allow service changes to be made quickly: 'So it was just a huge change really and it was no good kind of waiting for national guidance, from NHS England really, about what to do. I think it really was individual practices and maybe practices speaking within their Primary Care Networks, to sort of come up with a plan really and think how to work it'. For example, general practitioner appraisals and routine inspections by the Care Quality Commission (the independent regulator of health and adult social care services in England) were suspended in the early phase of the pandemic. Notably, the 'old' bureaucracy crept back in during the pandemic.<br><br>Some general practitioner participants reflected that the introduction of remote working had made it easier to schedule multidisciplinary team meetings and had reduced the amount of time spent travelling to meetings and events.<br><br>GPs spoke very positively of the vaccination programme, feeling a "real sense of satisfaction": "it feels like a hopeful, positive thing to do. As GPs and a primary care team who work really closely with a community, I think it feels like a lovely thing to do with, and for, that community and I think in policy terms it makes a lot of sense for general practice to be involved in delivering the vaccine." |
| What prevents/removes work-related joy? | During the initial stages of the pandemic, participants spoke of the difficulty of keeping up with constantly changing guidelines on the management of COVID-19 patients and the delivery of services for non COVID-19 patients.<br><br>The increased autonomy experienced during the pandemic met by feelings of great uncertainty from some participants as ways of working and modes of consulting changed.<br><br>Participants reflected frequently that there was an underlying uncertainty as to what the future of general practice would look like and that they and colleagues were dealing with great weariness and unease.<br><br>Some GPs reflected that the introduction of remote working had blurred the boundaries between their home and professional life, with the feeling of being "on call all the time".<br><br>Workload increased during the pandemic – bringing with it pressure and stress. Some GPs suggested that the increased availability of remote consultations might be responsible for this.<br><br>Telephone consultations during COVID-19 deprived GPs of cognitive shortcuts developed through their experience of consulting face-to-face: for example, the patient's gait as they walk into the room.<br><br>Initially during the pandemic, participants described being in "crisis" mode. This receded as time went on, leaving behind day-to-day work that was perceived as not as fulfilling as before the pandemic, leading to GPs feeling "flat". |
| Organisational characteristics (small, large) | |
| Managing ill-defined illnesses and chronic complex multimorbidity | |

| | |
|---|---|
| Human connection with patients | Practitioner participants often identified with general practice's strong roots within the community. This emphasis on the local community was evident when participants spoke about the positive relationship patients had with the practice and the value of continuity of care. As the pandemic progressed, and the pressures of social distancing became more acute, participants noted that the relationship between practice and patient could become strained at times – leading to anger from patients. Some participants noted an increasing tendency for patients to perceive that the practice was 'closed' and discussed negative portrayals of general practice in the media.<br><br>Remote consultations were described by some participants as feeling transactional and in opposition to the relationship-based care of general practice which they valued highly as a core feature of general practitioner and practice nurse roles. The relational aspects of general practice were also noted to be eroded by the efforts made on a daily basis to respond to COVID-19.<br><br>Interestingly, in a few cases, telephone consultations seemed to benefit patients who appeared to feel more comfortable talking over the telephone rather than discussing their concerns face-to-face.<br><br>One GP reflected that, during the pandemic, they felt they held more risk on behalf of patients and were more concerned about those in their care: "And I'm not depressed and I'm not angry, I'm just sadder. And I come home from work and I know I've done the best job I can, but I'm often more worried about my patients on average than I used to be, because I haven't satisfied my professional enquiry that I've done – have I ruled everything out? Have I missed anything, you know? And that just takes its toll over time". |
| Human connection with peers | |
| Human connection across other organisations (e.g., secondary care) | |
| Learning systems (e.g., competence, capability, mastery, socially situated) | |
| Additional useful information | |
| Summary of findings | This study describes the experiences of 17 primary care practitioners (predominantly GPs) during COVID-19. They describe an initial sense of autonomy but uncertainty, that was replaced by stability, reemerging bureaucracy and increased demand (thus workload), with patients feeling increasingly frustrated due to the pressures of social distancing and negative portrayals of GPs in the media. Although they helped reduce workload, remote consultations were described as transactional and detracting from relationship-based care. GPs reported taking on more risk and feeling more concerned about patients. Working alongside local communities to coordinate the vaccine role out was described very positively. |
| Strengths, limitations, and future research directions | Findings from this study reflect the *in-depth* experience of 17 general practice staff recorded over time. While the sample is relatively small, our findings nonetheless have wider relevance for the NHS general practice workforce.<br><br>Our study demonstrates the urgent need to build on the findings of previous studies and explore how the primary care workforce is responding to the new ways of working brought about by remote-first consulting. This exploration should consider what would support |

| | |
|---|---|
| | primary care practitioners in their work and wider well-being and ensure sustainability of the general practice and primary care nursing workforce in the face of increasing demand for services. |

| | |
|---|---|
| Paper Number | 67 |
| Author | Chilvers et al |
| Year | 2019 |
| Title | Identifying policies and strategies for general practitioner retention in direct patient care in the United Kingdom: a RAND/UCLA appropriateness method panel study |
| Journal | BMC Family Practice |
| Region (England, Scotland, Wales, and Northern Ireland) | England |
| Study methodology | 12 GPs participated in a RAND/UCLA Appropriateness Method panel. Panellists were asked to read an evidence briefing summary, and then complete an online survey on two occasions. During each round, participants rated the appropriateness of policies and strategies aimed at improving GP retention using a nine point scale (1 ‘extremely inappropriate’ to 9 ‘extremely appropriate’). Fifty-four potential policies and strategies (equating to 100 statements) were tested, focusing on factors influencing job satisfaction (e.g. well-being, workload, incentives and remuneration, flexible working, human resources systems). Ratings were analysed for panel consensus and categorised based on appropriateness (‘appropriate’, ‘uncertain’, ‘inappropriate’). |
| Type of document | Primary research |
| Topic or focus of paper | GP retention |
| Research objectives and focus | Identify policies and strategies that might be potentially appropriate at facilitating the retention of GPs in direct patient care in the UK. |
| Theoretical framework/theories/ underpinnings | |
| Description of intervention | |
| Rationale for paper/review | The United Kingdom (UK) is experiencing a general practitioner (GP) workforce retention crisis. Research has focused on investigating why GPs intend to quit, but less is known about the acceptability and effectiveness of policies and strategies to improve GP retention. |
| What creates meaning that sustains GPs engagement with, and involvement in, work? | |

| | |
|---|---|
| What solutions/recommendations are proposed? | 24/54 areas were deemed appropriate after round two (median score presented alongside each statement – see methodology above):<br>*Consensus was defined as ≤ 2 panellists' ratings outside the 'appropriate' range band (7–9).*<br><br>For implementation at a national/regional level:<br>1. In order to assess 'at-risk of GP shortages' status in a commissioning/planning area and taking into account confidentiality GP practices should be able to self-register their organisation's 'at-risk' status. 8<br>2. GP practices identified as being 'at-risk' of GP shortages should be provided with a toolkit to manage recruitment and retention. 8.5<br>3. New incentive and support packages should be available to GPs and other organisations setting up new practices or new ways of working in under-doctored areas. 7.5<br>4. There should be a publicity campaign focussing on managing expectations of patients in line with the resources and constraints of GP-based primary care services. 9<br>5. GP practices identified as being 'at-risk of GP shortages' should be managed with an appropriate and sensitive supportive arrangement – for (i) optional implementation. 8<br>6. GP practices identified as being 'at-risk of GP shortages' should be allocated a specialist team for managing recruitment and retention – for (i) optional implementation. 9<br><br>For implementation at a practice level:<br>1. GPs who are returning to work after a period of absence or after a career break should have access to 'Health and Wellbeing programmes' to help them manage their re-entry into the workforce – for (i) optional implementation. 8.5<br>2. GPs who are returning to work after a period of absence or after a career break should have access to schemes that have a range of routes and options that can be combined in a personal package for re-entry. 9<br>3. GPs who are returning to work after a period of absence or after a career break should have access to schemes that use a mix of online education and face-to-face meetings to ensure timely access to induction and refresher courses. 9<br>4. GP practices should implement strategically planned exits for retiring GPs. 7<br><br>For implementation at a GP level: |

| | |
|---|---|
| | 1. Peer support initiatives should be made available to GPs aimed specifically at health and well-being - for (i) GPs who are not reaching retirement age. 8.5<br>2. GPs should have access to their own specialised health care service to ensure a quick and confidential occupational healthcare service – for (i) all GPs, (ii) GPs reaching retirement and who could take their pensions, or (iii) GPs not reaching retirement. 9, 9, 9<br>3. A structured programme of training and support should be made available to all GPs in their first 5 years following qualification as an independent GP to help them establish healthy, productive careers – for (i) optional implementation. 7<br>4. GPs should consider portfolio working as part of their career pathway and this should be optional - for (i) all GPs, (ii) GPs reaching retirement and who could take their pensions, or (iii) GPs not reaching retirement. 9, 7, 7 **(however, notably, portfolio working was rate 'extremely inappropriate if compulsory').**<br>5. Career support should be available to GPs to enable portfolio opportunities to be identified and taken up in a strategic way to inform their future ambitions - for (i) all GPs, or (ii) GPs not reaching retirement. 8, 7.5<br>6. Incentives and support packages should be available for those GPs developing portfolio careers who are linking their portfolio activities to specialisms/areas that are directly beneficial to local clinical priorities - for (i) all GPs, (ii) GPs reaching retirement and who could take their pensions, or (iii) GPs not reaching retirement. 8, 8.5, 8.5<br>7. Where a strong case can be made that there is a financial risk directly relating to the work of the practice (e.g. ownership of premises), GPs should have access to schemes to reduce financial burden (e.g. buy back schemes for premises) – for (i) all GPs or (ii) GPs reaching retirement and who could take their pensions 9, 9<br>8. There should be an agreed maximum in the number of consultations that a GP should be allowed to conduct in a working day in order to protect patient safety as well as the health of the GP - for (i) all GPs, (ii) GPs reaching retirement and who could take their pensions, or (iii) GPs not reaching retirement. 9, 9, 9<br>9. There should be contractual changes to encourage longer consultations where appropriate - for (i) all GPs, (ii) GPs reaching retirement and who could take their pensions, or (iii) GPs not reaching retirement. 9, 9, 9<br>10. The working hours of GPs should routinely include fully-funded, dedicated time to accommodate the full range of roles (administrative, clinical, training, management, CPD, business undertaken as part of care professional activity – for (i) all GPs or (ii) GPs reaching retirement and who could take their pensions. 9, 9, 9 |

| | |
|---|---|
| | 11. Contracts based on specified programmed activities should be available to GPs to work across several GP practices and on other health related activities – for (i) all GPs, (ii) GPs reaching retirement and who could take their pensions, or (iii) GPs not reaching retirement. 7, 8, 8<br>12. For such GPs a comprehensive flexible careers scheme should be introduced with a view to supporting annualised hours, part-time working, and/or ad-hoc contributions to direct patient care. 9<br>13. For such GPs there should be financial incentives for such GPs who have maintained a prolonged/sustained period of direct patient care. 8.5<br>14. The annual appraisal and revalidation process should be reviewed with a view to streamlining and simplifying the process - for (i) GPs who have not encountered any concerns in the previous revalidation/appraisal processes, or (ii) GPs who would like to work with a specified and limited scope of practice. 8.5, 8.5 |
| From which stakeholders' perspective? (General practitioners, patients, other HCPs) | GP partners and GPs involved in national level activities for recruitment and retention. |
| How is work-related joy constructed? (Diversity of ways individuals may think about joy) | |
| What prevents/removes work-related joy? | |
| Organisational characteristics (small, large) | |
| Managing ill-defined illnesses and chronic complex multimorbidity | |
| Human connection with patients | |
| Human connection with peers | |
| Human connection across other organisations (e.g., secondary care) | |

| | |
|---|---|
| Learning systems (e.g., competence, capability, mastery, socially situated) | |
| Additional useful information | |
| Summary of findings | The panel of experienced GPs identified a wide range of practical ways to improve GP retention through interventions that might enhance job satisfaction and work-life balance. |
| Strengths, limitations, and future research directions | Only a quarter of GPs approached completed both rounds of voting – 12 GPs, which is a small number.<br><br>The panel was limited to GP Partners from two geographical areas, or GPs involved in national level activities for GP recruitment and retention, and their views may not be representative of other groups of GPs.<br><br>Despite providing a carefully worded definition of 'appropriateness', differences in interpretation amongst participants might have remained. |

| | |
|---|---|
| Paper Number | 68 |
| Author | Coventry, Coventry & Coventry |
| Year | 2017 |
| Title | Multidisciplinary team meetings in primary care: could they help to attract the GPs of tomorrow? |
| Journal | BJGP |
| Region (England, Scotland, Wales, and Northern Ireland) | UK |
| Study methodology | Commentary |
| Type of document | Secondary research |
| Topic or focus of paper | Multidisplinary teams |
| Research objectives and focus | Present the arguments for increased multidisciplinary teams within general practice – most notably in the context of GP recruitment. |
| Theoretical framework/theories/ underpinnings | |

| | |
|---|---|
| Description of intervention | |
| Rationale for paper/review | It is well recognised that there is currently a crisis in recruiting medical students and doctors to a career in general practice. |
| What creates meaning that sustains GPs engagement with, and involvement in, work? | |
| What solutions/recommendations are proposed? | |
| From which stakeholders' perspective? (General practitioners, patients, other HCPs) | |
| How is work-related joy constructed? (Diversity of ways individuals may think about joy) | |
| What prevents/removes work-related joy? | |
| Organisational characteristics (small, large) | |
| Managing ill-defined illnesses and chronic complex multimorbidity | |
| Human connection with patients | |
| Human connection with peers | This paper suggests that multidisplinary teams within general practice might increase support and team cohesion, reducing well-documented feelings of isolation – this has been shown to be the case for hospital doctors. |
| Human connection across other organisations (e.g., secondary care) | |
| Learning systems (e.g., competence, capability, mastery, socially situated) | |
| Additional useful information | |

| | |
|---|---|
| Summary of findings | This paper makes the case for multidisplinary teams in the contexts of patient care, medical education, recruitment, resilience and retention. The main point relevant to our review is that multidisplinary teams might increase team support and reduce feelings of isolation for GPs. |
| Strengths, limitations, and future research directions | |

| | |
|---|---|
| Paper Number | 69 |
| Author | Dale et al |
| Year | 2016 |
| Title | The general practitioner workforce crisis in England: a qualitative study of how appraisal and revalidation are contributing to intentions to leave practice |
| Journal | BMC Family Practice |
| Region (England, Scotland, Wales, and Northern Ireland) | England |
| Study methodology | GPs were surveyed about career intentions. All comments that included mention of appraisal or revalidation were extracted and analysed thematically. |
| Type of document | Primary research |
| Topic or focus of paper | GP appraisal and revalidation |
| Research objectives and focus | Identify how the experience of appraisal and revalidation might be influencing intentions to leave general practice. |
| Theoretical framework/theories/ underpinnings | |
| Description of intervention | |
| Rationale for paper/review | The general practice (GP) workforce in England is in crisis, with declining morale and job satisfaction, increasing early retirement and declining interest in training to become a GP. We recently reported on factors that are influencing this, with appraisal and revalidation emerging as an unexpected finding; 28.6 % of GPs stating an intention to leave general practice within the next 5 years included this as ‘very important’ or ‘important’ to their decision. |
| What creates meaning that sustains GPs engagement with, and involvement in, work? | In a previous study, 28.6 % of GPs stating an intention to leave general practice within the next 5 years listed appraisal and/or revalidation as an important factor in their decision. Thematic analysis of free text comments relating to this revealed five main themes:<br>1. Time-consuming and bureaucratic nature of the process |

| | |
|---|---|
| | 2. Relevance and validity to different work patterns (such as working as a locum)<br>3. Emotional impact<br>4. The culminative impact of many factors with appraisals being the "final straw"<br>5. The need for the process to be improved<br><br>Many respondents felt that appraisal and revalidation lacked educational and developmental value, and hence served little purpose for the public or the profession. Collecting the required evidence was seen as having become a tick box exercise, and some respondents questioned the experience and skills of their appraiser to provide meaningful feedback: "Complete waste of time. Everyone I speak to just does it to fulfil the legal requirements rather than for self-improvement which is what it should be for"<br><br>Many comments reflected the way that appraisal, revalidation and other inspection, such as CQC, were experienced as irrelevant to general practice and demeaned their professionalism: "Am constantly having to prove that I am not a criminal or an incompetent to the CQC, GMC, NHS England—all treat us as the enemy, or as naughty children who cannot be trusted".<br><br>The perceived lack of flexibility in the requirements for appraisal and revalidation were seen as disadvantaging part-time, portfolio and locum GPs – impacting GPs who wanted to reduce their hours to part time but could not face the appraisal burden. |
| What solutions/recommendations are proposed? | Several respondents highlighted the importance of changing appraisal and revalidation in order to make it less onerous, more flexible and more relevant to individual GPs' pattern of work. In addition, it was felt that the processes of appraisal, revalidation, practice inspections by the CQC and the CCG could all be bundled into a single process which would reduce the cost and time involved. Specific elements of appraisal and revalidation, such as the expectation that an audit had been conducted, were seen as barriers to remaining in general practice. There was also a view that the extent of evidence needed to support appraisal and revalidation should be relevant to the stage of career that the doctor was at.<br><br>GPs are known to have more positive attitudes towards appraisal that has local ownership, is educationally oriented, and where the GP feels they have more control over the process . Hence, appraisal could offer a mechanism to provide support and direction which might help to delay retirement. However, this might require a substantial loosening of its link with performance management. |
| From which stakeholders' perspective? (General practitioners, patients, other HCPs) | GPs |
| How is work-related joy constructed? (Diversity of ways individuals may think about joy) | |
| What prevents/removes work-related joy? | Although some GPs understood the principle of appraisal and revalidation, in reality it was regarded as unnecessarily complex, time-consuming and a "bureaucratic nightmare". Several respondents linked this to the Care Quality Commission (CQC) and other inspections |

| | |
|---|---|
| | which were also seen as being overly complex. Together with other managerial and administrative activities faced by GPs, this was adding to the workload and work-related stress that many respondents described.<br><br>Preparing for appraisals was regarded as a significant emotional burden: “...one of the most depressing parts of general practice is revalidation. It is so time-consuming and yet so unnecessary. It does not in any way improve my practice but it hugely negatively impacts on my time and my morale. I feel as though I’m being punished for one other doctor’s conduct in an inappropriate way”<br><br>Appraisal was regarded as the final straw for many GPs: “APPRAISAL AT END OF CAREER IS A SURE FIRED WAY OF GPS THROWING THEIR TOWEL IN when they are still capable of helping in various clinical ways .........so I am getting out NOW and collecting my pension ASAP” |
| Organisational characteristics (small, large) | |
| Managing ill-defined illnesses and chronic complex multimorbidity | |
| Human connection with patients | |
| Human connection with peers | |
| Human connection across other organisations (e.g., secondary care) | |
| Learning systems (e.g., competence, capability, mastery, socially situated) | |
| Additional useful information | |
| Summary of findings | This paper analysed how GPs’ views on appraisal and revalidation were influencing intentions to leave general practice. It found that appraisal and/or revalidation was regarded as time-consuming and bureaucratic, inflexible, demeaning to the profession and emotionally draining. For senior GPs, appraisal was often the final straw leading to their retirement. It was highlighted that the process needs to be less onerous, more flexible and more relevant – and possible bundled together with practice inspections by the CQC and CCG. |
| Strengths, limitations, and future research directions | Uncertainty over the representativeness of respondents; responders may have been more dissatisfied or intent on leaving general practice than nonresponders, although it is possible that those who felt the most over-worked or disillusioned may have been less motivated to respond and hence been under-represented. |

| | |
|---|---|
| | Finally, there is a need for further research into GPs' views as part of an overall evaluation of the costs and consequences of appraisal and revalidation. Such research should explore the aspects of appraisal/revalidation which are most problematic for GPs, investigate appraisers' views and be aimed at identifying an approach that would be trusted and have greater effectiveness and applicability across the workforce. |

| | |
|---|---|
| Paper Number | 70 |
| Author | Dhanni & Blane |
| Year | 2022 |
| Title | The Deep End GP Pioneer Scheme: a qualitative evaluation |
| Journal | Australian Journal of Primary Health |
| Region (England, Scotland, Wales, and Northern Ireland) | Scotland |
| Study methodology | Interviews with GPs stakeholders involved in the Scottish Deep End Project (see below), followed by thematic analysis of responses |
| Type of document | Primary research |
| Topic or focus of paper | Retention of GPs working in areas of socio-economic deprivation. |
| Research objectives and focus | Explore the views and experiences of the GP fellows and lead GPs from participating practices, as a qualitative evaluation of the Pioneer Scheme. |
| Theoretical framework/theories/ underpinnings | |
| Description of intervention | The aim of the Deep End Pioneer Scheme was to develop a change model for general practices serving areas of socio-economic deprivation (the Deep End), involving the recruitment of younger GPs, the retention of experienced GPs, and their joint engagement in strengthening the role of general practice as the natural hub of local health systems. This included a bespoke academic program of learning related to 'Deep End' issues such as chronic pain, trauma, child protection and more. Early career GP 'fellows' worked either four or six sessions in Deep End practices and had one session of protected time, which alternated between the academic program of learning and working on service development projects with the lead GP from their practice. The practices involved met every 6–8 weeks to share ideas and learning, supported by an overall lead GP for the project. |
| Rationale for paper/review | Challenges for healthcare teams working in areas of severe socio-economic deprivation in Scotland are well documented and include premature multi-morbidity; increased GP stress and lower patient enablement; and a relatively older cohort of GPs approaching retirement, such that the GP workforce 'crisis'–traditionally associated with remote and rural areas – is also felt particularly acutely in urban deprived practices. |

| | |
|---|---|
| What creates meaning that sustains GPs engagement with, and involvement in, work? | Senior GPs felt a duty to their patients to pass on their knowledge and learning to the next generation of Deep End GPs: "I was very interested to be involved in order to ... free up some time for myself and also share learning with another colleague … you know. I do feel there is that obligation on me to want to share that before, you know, I'm off."<br><br>GP fellows reported feeling an increased sense of meaning in their work, which they ascribed to having an overarching goal (the Deep End GP vision of improving general practice in deprived areas) and the resources (time and learning) to facilitate efforts towards that goal, including advocacy work.<br><br>All fellows highlighted lifestyle factors such as balancing family commitments, commuting time, and risk of burnout with full-time work as key considerations for future work plans. |
| What solutions/recommendations are proposed? | The Pioneer Scheme has inspired a similar program called the Fairhealth Trailblazer Scheme across England. There is growing support to re-launch the scheme in Scotland too. |
| From which stakeholders' perspective? (General practitioners, patients, other HCPs) | |
| How is work-related joy constructed? (Diversity of ways individuals may think about joy) | Lead GPs explained that they were attracted to the protected time that was created by having this additional clinical capacity, as it allowed experienced GPs the 'headspace' to think 'in a more creative way and therefore in a more proactive and preventative way'.<br><br>Both early career and lead GPs involved in the scheme reported feeling greater autonomy, and a greater ability to implement ideas and initiatives for their practice populations and professional interests: "When I moved into the practice I was in most of the time, in meetings I could make a suggestion and actually people would change things which hadn't happened before."<br><br>Many lead GPs spoke of how the Pioneer Scheme had given them a renewed sense of purpose and enjoyment in their work, in the context of feeling previously worn out. |
| What prevents/removes work-related joy? | Similar to other papers in the literature, GP fellows nearing the end of the scheme expressed negative views about partnership: "I wouldn't have any intention to … become a partner because at the moment I think that the payoffs of that aren't [great]. It's unnecessary stress that the system is under pressure and it's going through a lot of changes with uncertainty. So it's high risk with what I see at the moment as very little reward."<br><br>There was recognition amongst participants that current workloads in Deep End practices – particularly those with a higher percentage of patients living in the most deprived postcodes – was unsustainable: "So I think one thing that I've realised by the specific practice that I'm in, and I'm in one of the … most deprived, so it's around 80 plus percent deprived, is that I don't think that I could sustain a GP career in that level of deprivation. I'm very interested in [Deep End GP] but I think even dropping down to 40/50/60 percent of level of deprivation I think, I could imagine [would be more sustainable]. This is what I want to go and taste through locuming." |

| | |
|---|---|
| Organisational characteristics (small, large) | |
| Managing ill-defined illnesses and chronic complex multimorbidity | |
| Human connection with patients | |
| Human connection with peers | Early career GPs were attracted to the idea of support and mentoring from more senior GPs, as a transition from their GP training to progressively more independent practice. The access to a peer support network was an additional attraction for fellows, particularly for those who had moved from elsewhere and were newly practicing as GPs in Glasgow: "[prior to the Scheme] I started working in a Deep End practice and the work was really interesting and the work was really challenging but I had no one to bounce ideas off of. In the way of peer support, people were really nice and everyone was doing their best, but it was really official firefighting. Everyone was just firefighting and I felt like there wasn't much connectivity between people".<br><br>Some fellows expressed the desire to seek out like-minded peers interested in addressing social exclusion and health inequalities.<br><br>Both GP fellows and lead GPs agreed that the Pioneer Scheme had increased their sense of relatedness and connection. Participants described feeling part of something bigger, facilitated by the round table discussions that took place at both the fortnightly academic sessions (for fellows) and the 6–8 weekly wider-team meetings (for fellows and lead GPs). This was set in contrast to their prior experience of feeling increasingly isolated and disconnected from colleagues due to increasing work pressures. |
| Human connection across other organisations (e.g., secondary care) | |
| Learning systems (e.g., competence, capability, mastery, socially situated) | Many early career GPs expressed a pre-existing interest in addressing health inequalities and were attracted to the Pioneer Scheme as a mechanism by which they could feel more equipped to address these issues. Relating to this, access to high-quality continuing professional development (CPD), which focused on health inequalities and subjects not covered in postgraduate training, with protected time, was also attractive.<br><br>GP fellows were particularly vocal about the increased clinical confidence, or mastery, that they had developed as a result of the Pioneer Scheme. They described different mechanisms underpinning this, including the dedicated academic sessions for professional development, the mentoring relationship with their lead GP, sharing of learning within and between practices (via team meetings and an online platform), and peer support. Increased knowledge of local resources and third sector organisations, and greater familiarity with referral processes, were other elements of mastery that participants felt improved patient care. For lead GPs, too, there was a feeling of increased mastery, but this was less about clinical competence and more about having the time and headspace to feel present and compassionate during consultations. |
| Additional useful information | |

| | |
|---|---|
| Summary of findings | This qualitative study of lead GP and GP fellow experiences of the Deep End GP Pioneer Scheme found strong support for the Pioneer Scheme as a mechanism to improve GP recruitment and retention in areas of high socio-economic disadvantage (and, indeed, to improve quality of care in these areas). Key ingredients of the Pioneer Scheme were the additional clinical capacity (addressing the inverse care law), protected time for both GP fellows and experienced GPs to lead on service development initiatives and to share learning within and between practices, and the shared ethos and values of the Pioneer Scheme.<br><br>For our review, I think the really important takeaway of this study is that GP work in areas of high socio-economic deprivation can be very meaningful and enjoyable, provided appropriate support and capacity is in place. |
| Strengths, limitations, and future research directions | Strengths of the study include the high response rate, with 12 out of 14 participating practices taking part. A rigorous approach to qualitative analysis was followed, with the use of theory underpinning the section on GP work motivation. A potential limitation of this study is that it took place before the second cohort had completed the Pioneer Scheme, but over a year after the first cohort had completed, so there may be issues of social desirability bias and recall bias, respectively. |

| | |
|---|---|
| Paper Number | 71 |
| Author | John Frey |
| Year | 2018 |
| Title | Professional Loneliness and the Loss of the Doctors' Dining Room |
| Journal | Annals of Family Medicine |
| Region (England, Scotland, Wales, and Northern Ireland) | N/A (Commentary written by a doctor from the USA) |
| Study methodology | Commentary |
| Type of document | Secondary research |
| Topic or focus of paper | Professional connections and relationships |
| Research objectives and focus | Highlight how professional relationships have been eroded in medicine in recent years – particularly for family doctors – and describe the impact of this on physicians. |
| Theoretical framework/theories/ underpinnings | |
| Description of intervention | |

| | |
|---|---|
| Rationale for paper/review | Historically, family physicians moved among all the venues of medical care office, hospital, community—and were a part of a connected professional community. The current fragmentation of medicine into narrowly defined, boundaried workspaces and job descriptions, as well as the increasing size of practices has negatively affected the professional culture in which physicians work. |
| What creates meaning that sustains GPs engagement with, and involvement in, work? | This paper explains that family physicians are becoming increasingly isolated from peers both in and outside of their practices (see below). This impacts upon life satisfaction and energy levels. A Canadian study found that increasing social interactions and friendships among medical students decreases the tendency to experience emotional and physical exhaustion. Another Canadian study found that the more related physicians felt to each other, the higher the level of professional well-being, professional satisfaction, and energy in their work. Furthermore, this makes it more difficult for doctors to support each other, as struggles are normally conveyed through unconscious cues: More often than not, the "help" is in a colleague's demeanour, voice, eyes, giving us a chance to put a hand on a shoulder and ask, "Are you all right?". |
| What solutions/recommendations are proposed? | The paper suggests that systems need to create safe spaces for time and conversation and connections between colleagues. On an individual level, doctors need to make an effort to form relationships – like calling a colleague or getting a meal together. |
| From which stakeholders' perspective? (General practitioners, patients, other HCPs) | |
| How is work-related joy constructed? (Diversity of ways individuals may think about joy) | |
| What prevents/removes work-related joy? | |
| Organisational characteristics (small, large) | The author highlighted that big practices are more likely to lead to isolation amongst family doctors, due to large teams: "But large buildings in large health systems in big cities are among the loneliest and most alienated places to practice these days." "Big clinics offer anonymity". |
| Managing ill-defined illnesses and chronic complex multimorbidity | |
| Human connection with patients | |
| Human connection with peers | The author explained that, in modern family medicine, doctors spend less time forming relationships with colleagues, leading to isolation. This is in contrast to the past, when doctors might do things like get lunch together or have informal conversations. In turn, this was ascribed to healthcare systems not valuing physicians spending time with one another.<br><br>"Today, lunches are solitary, eaten at desks from plastic containers from home or takeout from local restaurants while catching up on email, writing charts before the afternoon begins, or answering online patient inquiries. Even in common spaces and nurses' stations, everyone sits |

| | |
|---|---|
| | silently in front of a computer screen. No talking of Cabbages and Kings or pigs with wings these days. Meetings are functional, organizational, and informational. People come late and leave early. “Send me an email” we say.” |
| Human connection across other organisations (e.g., secondary care) | Generalists nowadays are less engaged in hospital care and, as a result, have much less personal contact with the consultants they use. The times spent with colleagues in labor and delivery, in the ED, or morning rounds are being replaced by e-consults and wordy electronic progress notes more filled with facts than information. Primary care clinicians don’t get personal notes or phone calls back—they are told that notes in the EHR should suffice. This leads to isolation amongst primary care doctors. |
| Learning systems (e.g., competence, capability, mastery, socially situated) | |
| Additional useful information | |
| Summary of findings | Not valuing time with other physicians or allowing for informal conversations leads to a soulless efficiency and professional isolation that drains physicians of our ability to help ourselves, help each other, and help patients. |
| Strengths, limitations, and future research directions | |

| | |
|---|---|
| Paper Number | 72 |
| Author | Shah et al |
| Year | 2021 |
| Title | A survey of the retention of GPs following the induction and refresher scheme in London |
| Journal | Education for Primary Care |
| Region (England, Scotland, Wales, and Northern Ireland) | England |
| Study methodology | A questionnaire survey (including two free text questions) was sent out to all 116 GPs who completed the London I&R scheme between January 2015 and April 2020. Survey results were analysed by the authors. Furthermore, a search was carried out using Primary Care Support England to determine whether these candidates are still available to work as a GP. |
| Type of document | Primary research |
| Topic or focus of paper | Retention of GPs who have taken time out of practice |
| Research objectives and focus | Report on whether participants found the I&R scheme useful and whether they have continued to work as GPs. |

| | |
|---|---|
| Theoretical framework/theories/ underpinnings | |
| Description of intervention | The I&R Scheme is mandated by NHS England after an absence from general practice of two years or more, in order to regain entry to the Medical Performer's List (MPL). It is also open to GPs who qualified abroad. Doctors joining the Scheme are required to pass entry assessments consisting of two multiple-choice question (MCQ) papers designed to test clinical knowledge and situational judgement. Following successful completion of the assessments, a supervised placement is organised for doctors on the Scheme. Whilst on the London I&R Scheme doctors have access to the full range of services offered by the London Professional Support Unit (PSU), are granted Associate Membership of the Royal College of GPs and have free access to the Fourteen Fish portfolio and learning materials. They are also invited to participate in monthly learning sets (lasting 2–3 hours) through which they can form social networks with their peers |
| Rationale for paper/review | The NHS faces challenges in recruiting and retaining GPs. The NHS Long Term Plan aims to increase NHS staffing levels as one of its objectives. The Induction and Refresher (I&R) Scheme supports this aim by providing a safe induction or return to UK practice for GPs who are new to UK general practice or who have been away from it for 2 years or more. |
| What creates meaning that sustains GPs engagement with, and involvement in, work? | This paper found that 94% of doctors surveyed who have completed the I&R scheme are still available to work as GPs. UK 'returners' explained that the scheme supported them in building their confidence and making a gradual scheme to general practice: "Very supportive times with colleagues in I&R scheme. Excellent training practice which allowed me to familiarise myself with UK general practice and allowed me to regain my confidence working in a different health setting".<br><br>Non-UK qualified GPs made similar positive comments, noting how the scheme helped them to understand NHS systems: "Getting to know how the NHS works, referral pathways, local guidance. It was also a good cultural immersion to understand patient expectations and the general attitude of patients as well as doctors in the UK". |
| What solutions/recommendations are proposed? | |
| From which stakeholders' perspective? (General practitioners, patients, other HCPs) | GPs |
| How is work-related joy constructed? (Diversity of ways individuals may think about joy) | |
| What prevents/removes work-related joy? | |
| Organisational characteristics (small, large) | |

| | |
|---|---|
| Managing ill-defined illnesses and chronic complex multimorbidity | |
| Human connection with patients | |
| Human connection with peers | Both UK returners and non-UK qualified GPs highlighted the value of monthly meeting with peers and resultant building of support networks. |
| Human connection across other organisations (e.g., secondary care) | |
| Learning systems (e.g., competence, capability, mastery, socially situated) | |
| Additional useful information | |
| Summary of findings | This paper emphasised the success of the I&R scheme in retaining UK GPs who have taken time out of practice and non-UK qualified GPs. Of note to our review, it suggests that GPs who have taken time out of practice benefit from a gradual, supported return to help them build up confidence. |
| Strengths, limitations, and future research directions | |

| | |
|---|---|
| Paper Number | 73 |
| Author | Atherton et al |
| Year | 2013 |
| Title | Experiences of using email for general practice consultations: a qualitative study |
| Journal | BJGP |
| Region (England, Scotland, Wales, and Northern Ireland) | England |
| Study methodology | Interviews with patients and GPs about email consultations, as well as experts in this field. Subsequent thematic analysis of responses. |
| Type of document | Primary research |
| Topic or focus of paper | Email consultations |

| | |
|---|---|
| Research objectives and focus | To understand the use of email consultation in general practice by investigating the experiences of existing users and views of experts. |
| Theoretical framework/theories/ underpinnings | |
| Description of intervention | |
| Rationale for paper/review | |
| What creates meaning that sustains GPs engagement with, and involvement in, work? | |
| What solutions/recommendations are proposed? | GPs attempted to place boundaries around their email use, using email only with selected patients and setting ground rules. The implication was that the chosen patients were less likely to abuse it.<br><br>GPs created rules to enable them to control safety concerns and manage risk. For example, they informed patients that they may not receive a response immediately.<br><br>GPs attempted to apply previous experience with in-person consultations and usual professional standards to email consultations.<br><br>Overall, both GPs and patients agreed that email consultation is not suitable for all patients, or for every situation. There was recognition of the need for regulation and guidance in using email consultation. |
| From which stakeholders' perspective? (General practitioners, patients, other HCPs) | 14 patients, 10 GPs, 6 experts |
| How is work-related joy constructed? (Diversity of ways individuals may think about joy) | Both groups were positive about the convenience that email offers, with advantages ranging from its 'efficiency' and 'speed' to specific characteristics like asynchronicity and the written format. Furthermore, for GPs, email allows them to avoid 'telephone tag' with working patients. |
| What prevents/removes work-related joy? | Workload difficulties arose because email is not integrated into practice, time is not allowed for 'extra' consultations and, in some cases, GPs reported that email was generating more work and additional consultations. This was supported by patient accounts that they were consulting about concerns for which they would not otherwise have contacted the practice.<br><br>Some GPs experienced inappropriate patient behaviour when using email: "And one of my colleagues gave him my email address and that wasn't something I would [have done]. He wasn't someone I would necessarily have chosen, and I then got probably about 10 emails from him every week, and, we had to sort of make some very strong ground rules about those." |

| | |
|---|---|
| | Some GPs reported email consultations leading to anxiety: "And ... and ... there's this is kind of which I'm sure has come up in your other conversations—but this kind of feeling that, in some way, what if they're lost, or missed, or someone doesn't pick it up, or if I'm not here, who will manage it?'<br><br>GPs reported uncertainty about email consultations stemming from a lack of agreed rules of engagement. |
| Organisational characteristics (small, large) | |
| Managing ill-defined illnesses and chronic complex multimorbidity | |
| Human connection with patients | In cases whereby patients had a relationship with a specific doctor, email was seen as promoting and maintaining that relationship, allowing GPs to deliver personalised care. |
| Human connection with peers | |
| Human connection across other organisations (e.g., secondary care) | |
| Learning systems (e.g., competence, capability, mastery, socially situated) | |
| Additional useful information | |
| Summary of findings | This study analysed the views of patients, GPs and experts on email consultations. Relevant to our study, it found that email consultations are more convenient and efficient for some patients and problems. However, they can also increased GPs' workload, lead to inappropriate behaviour from patients and increase anxiety and uncertainty amongst GPs. To combat this, GPs need to define boundaries and rules for email consultations, such that they are used safely and appropriately. |
| Strengths, limitations, and future research directions | This study included a wide range of participants who reported varying frequencies and duration of email use. A major strength was the coherence of findings between the three participant groups. Saturation was reached in the sample. The study was set in London and so the transferability of the findings outside of this setting may be questionable. Patient participants were largely educated to higher level (degree level and above) and only one was aged ≥65 years. As there is speculation that older adults and those in lower socioeconomic groups are more likely to be left behind in relation to the uptake and use of new technologies, this may reduce the transferability of the findings.<br><br>Key areas that warrant further research include:<br>1. Understanding with which patients email is best used and what for<br>2. The effect of email consultation on workload |

| | |
|---|---|
| | 3. The safety of email consultation |

| | |
|---|---|
| Paper Number | 74 |
| Author | Baker et al |
| Year | 2014 |
| Title | Primary care quality and safety systems in the English National Health Service: a case study of a new type of primary care provider |
| Journal | Journal of Health Services Research |
| Region (England, Scotland, Wales, and Northern Ireland) | England |
| Study methodology | Interviews with practice staff, senior staff and owners. Ethnographic observation of interactions between staff and staff and between staff and patients in non-clinical areas. Analysis was based on the constant comparative method. |
| Type of document | Primary research |
| Topic or focus of paper | Practice size and organisation |
| Research objectives and focus | Characterise the quality and safety systems and processes of a large, privately owned company providing primary care through a chain of general practices in England. |
| Theoretical framework/theories/ underpinnings | |
| Description of intervention | |
| Rationale for paper/review | Although the predominant model of general practice in the UK National Health Service (NHS) remains the small partnership owned and run by general practitioners (GPs), new types of provider are emerging. Despite concerns about the potential impact of new types of provider, particularly in the form of corporate chains of practices, on personal care and other issues, studies of such providers remain limited. |
| What creates meaning that sustains GPs engagement with, and involvement in, work? | |
| What solutions/recommendations are proposed? | |

| | |
|---|---|
| From which stakeholders' perspective? (General practitioners, patients, other HCPs) | Senior managers, GPs, practice administrative staff and nurses. |
| How is work-related joy constructed? (Diversity of ways individuals may think about joy) | |
| What prevents/removes work-related joy? | The company performance managed practices and individuals through various indicators, including QOF indicators, recruitment and exit of registered patients, numbers of appointments offered and unused, together with feedback and benchmarking. Staff, including GPs, who did not meet the requirements of Primer were "performance managed out of the organization". Although not explcitely stated by the paper, it is easy to see how this could create stress for GPs.<br><br>There were some suggestions from our observations and interviews that being part of a large organization, where no-one locally owned the practice, meant that staff felt less valued. |
| Organisational characteristics (small, large) | The large scale of the organisation (50 practices) was highlighted to have benefits for GPs, including a reduced bureaucratic burden, a simplification of certain tasks and enhanced training. |
| Managing ill-defined illnesses and chronic complex multimorbidity | |
| Human connection with patients | |
| Human connection with peers | The study noted that it was difficult to socialise the high number of temporary, locum GPs into the culture of practices and the company. |
| Human connection across other organisations (e.g., secondary care) | |
| Learning systems (e.g., competence, capability, mastery, socially situated) | |
| Additional useful information | |
| Summary of findings | This study qualitatively investigates a large primary care provider, focussing mainly on quality and safety issues. Relevant to our review, the authors note that a large-scale organisation can reduce administrative tasks for clinical staff and enhance training. However, it can also lead to staff feeling less valued, and the high levels of performance management may lead to stress amongst GPs. |

| | |
|---|---|
| Strengths, limitations, and future research directions | We have only looked at one type of new provider, and at a particular stage in that organization's history. The provider had been initiated by a small number of GPs who continued to be leaders in the organization, and the findings may not, therefore, reflect approaches to quality and safety in organizations established from the start by corporate organizations.<br><br>We did not undertake a 'before and after study' of transformation in the three practices we studied in depth, nor did we make a direct comparison with more traditional GP practices.<br><br>We interviewed staff and conducted observations in three practices only, and although we have no reason to suspect that these practices were atypical, it is possible that our impression of the quality and safety systems and culture is incomplete.<br><br>The multi-unit chain model is proliferating in health care. Future research should seek to investigate the characteristics of different, larger organizational forms in primary care, including ways in which commissioners can reap the benefits of larger organizations without requiring the formal merger of practices. |

| | |
|---|---|
| Paper Number | 75 |
| Author | Beech et al |
| Year | 2019 |
| Title | Closing the gap: key areas for action on the health and care workforce |
| Journal | Joint report from The King's Fund, The Health Foundation and The Nuffield Trust |
| Region (England, Scotland, Wales, and Northern Ireland) | England |
| Study methodology | Collection of policy analyses by experts from the three tanks. These have 'drawn on published literature and data' and '[the authors] have also spoken to national and professional leaders' |
| Type of document | Secondary research |
| Topic or focus of paper | Workforce crisis in the NHS |
| Research objectives and focus | Lay out a set of high-impact interventions that, if put into action now, could help to ameliorate the current workforce crisis within the English NHS. We focus on the areas where severe national problems are having an immediate impact. |
| Theoretical framework/theories/ underpinnings | |
| Description of intervention | |

| | |
|---|---|
| Rationale for paper/review | The NHS workforce crisis: NHS hospitals and mental health and community providers are currently reporting a shortage of more than 100,000 FTE staff. |
| What creates meaning that sustains GPs engagement with, and involvement in, work? | The report highlights that the range of clinical and non-clinical staff working alongside GPs will have to be expanded to address workforce shortages and increasing patient demand (an approach echoed by the NHS Long Term Plan). Examples include advanced practice roles (e.g. ANP and PAs) and clinical support roles (e.g. social prescribing link workers). These new roles in general practice may reduce workload for GPs and lead to more appropriate care for patients, however will also change the nature of work conducted by GPs who will likely focus on more complex cases. Indeed, the report emphasises how “all frontline staff will need to acquire new skills and adopt new ways of working to support changing models of care”.<br><br>Furthermore, with new roles in general practice, the report emphasised the importance of “regulation, clarity of purpose and a defined, safe remit” alongside “significant investment in training, mentorship and support”.<br><br>The general practice estate was highlighted as a significant barrier to expanding the multidisplinary team within general practice. The British Medical Association reports that seven out of ten GPs in England feel that their facilities are too small to deliver extra or additional services to patients, while four out of ten GPs say that their practice currently does not have adequate facilities to deliver services to patients. A review of GP premises has been commissioned to find solutions to problems with the current general practice estate. Due consideration must be given to the ease with which general practice (and other parts of the NHS's estate seeing similar 'mixed use') can be expected to co-locate professionals in the same place as teams expand. Significant investment in the primary care estate will be needed.<br><br>Both in the UK and internationally, there is a shift away from 'doctor-driven' to team-based ways of working within primary care. In these models, non-medical staff are not there to be 'delegated' to but to form part of a wider and enriched health care team with a different skill mix |
| What solutions/recommendations are proposed? | |
| From which stakeholders’ perspective? (General practitioners, patients, other HCPs) | |
| How is work-related joy constructed? (Diversity of ways individuals may think about joy) | New technologies are also noted to be decreasing the workload for GPs. |
| What prevents/removes work-related joy? | For both hospital doctors and GPs, the combination of changes to the NHS pension scheme and lowering the thresholds of the pensions annual and lifetime allowances appears to have incentivised experienced doctors to retire early or to reduce their working hours.<br><br>GPs are subject to ongoing pressures and a high workload. There are multiple causes of this including the workforce crisis and increasing patient demand and complexity. These pressures, in-and-of-themself, lead to problems with GP recruitment and retention. |

| | |
|---|---|
| Organisational characteristics (small, large) | |
| Managing ill-defined illnesses and chronic complex multimorbidity | |
| Human connection with patients | |
| Human connection with peers | |
| Human connection across other organisations (e.g., secondary care) | |
| Learning systems (e.g., competence, capability, mastery, socially situated) | |
| Additional useful information | |
| Summary of findings | This report summarises key recommendations for addressing the workforce crisis in the English NHS. Much of it is not specific to general practice, however chapter 5 focusses heavily on primary care. In particular, it stresses the importance of expanding the multidisciplinary team in general practice to respond to patient demand and decrease workload for GPs. Of relevance to our review, it highlights how this will change the nature of work conducted by GPs who will likely focus on more complex patients. Furthermore, these new roles will require regulation, clarity and investment in training and estates – failure to do so may place new burdens and anxieties on GPs. Interestingly, the report discusses these changes in the context of an international shift away from ‘’doctor-driven’ ways of working within primary care. |
| Strengths, limitations, and future research directions | This is not a comprehensive programme and there are many important aspects of workforce policy we have not addressed, including, for example, geographical inequalities. As explained previously, our focus is narrowly on the staff directly employed in NHS providers and general practice. However, national workforce policy needs to be more comprehensive, understanding the NHS's role in the wider health and care sector.<br><br>Our policy recommendations are based on the current evidence but in many areas this is limited, so many of our recommendations are not based on as robust an evidence base as we would wish. The problem is that the workforce issues are too acute and too critical to wait for better information and research. |

| | |
|---|---|
| Paper Number | 76 |
| Author | Borek et al |

| | |
|---|---|
| Year | 2020 |
| Title | Social and Contextual Influences on Antibiotic Prescribing and Antimicrobial Stewardship: A Qualitative Study with Clinical Commissioning Group and General Practice Professionals |
| Journal | Antibiotics |
| Region (England, Scotland, Wales, and Northern Ireland) | England |
| Study methodology | Interviews with CCG professionals and GPs, followed by thematic analysis of responses. |
| Type of document | Primary research |
| Topic or focus of paper | Antimicrobial stewardship |
| Research objectives and focus | Assess social and contextual factors affecting antibiotic prescribing and engagement with antimicrobial stewardship (AMS) initiatives. |
| Theoretical framework/theories/ underpinnings | |
| Description of intervention | |
| Rationale for paper/review | Antibiotic prescribing in England varies considerably between Clinical Commissioning Groups (CCGs) and general practices. |
| What creates meaning that sustains GPs engagement with, and involvement in, work? | |
| What solutions/recommendations are proposed? | AMS strategies need to fit within the constraints imposed by the immense pressure upon general practice, for example, by being easily accessible and adaptable, not requiring (much) additional work or effort, and being concise and with clear benefits to practitioners. CCG professionals also reported the need for more tailoring of AMS approaches to practices and local contexts to account for the different reasons for high prescribing and variations in scope and areas for improvement. |
| From which stakeholders' perspective? (General practitioners, patients, other HCPs) | 22 CCG professionals and 19 GPs. |
| How is work-related joy constructed? (Diversity of ways individuals may think about joy) | |
| What prevents/removes work-related joy? | Reducing antibiotic prescribing was just seen as 'another challenge' in general practices under large amounts of pressure (stable staff and patients, allowing for continuity of care, were seen as factors enabling more appropriate prescribing). |

| | |
|---|---|
| | Some higher prescribers were seen as more concerned about risks or having experience of adverse effects, and potential malpractice accusations and legal consequences, of not prescribing antibiotics.<br><br>There was a sense that professionals are increasingly “tired” of the constant pressure to change practice but without the necessary changes in the context that affects it: “The thing that everyone recognizes is that it takes so much longer to explain why you’re not prescribing than it does to just churn out a script for an antibiotic so time is key and over-pressurized practices with insufficient staff for the workload might be predicted to prescribe more or more often inappropriately, possibly.”<br><br>Participants questioned how certain antibiotic prescribing targets and thresholds were identified and what evidence guided this process, describing them as “arbitrary”. Yet, they were then used to benchmark CCGs’ and practices’ prescribing, leading to defining some as “high” prescribers: “Why have they chosen 10%’? And why this and why that and, ‘They just plucked these figures out of the air’, so it did create some negative feedback.” |
| Organisational characteristics (small, large) | |
| Managing ill-defined illnesses and chronic complex multimorbidity | |
| Human connection with patients | GPs refusing antibiotics to patients often led to conflict when patients expected antibiotics. A number of factors increased patients’ expectations for antibiotics including: greater deprivation, affluent rural communities, cultural/ethnic background and language barriers. |
| Human connection with peers | |
| Human connection across other organisations (e.g., secondary care) | |
| Learning systems (e.g., competence, capability, mastery, socially situated) | |
| Additional useful information | |
| Summary of findings | This qualitative study mainly focuses on the enables and barriers to antimicrobial stewardship initiatives within general practice. A few findings are indirectly relevant to our study – including the added burden of antimicrobial stewardship to practices under immense pressure, the anxiety experienced by GPs who choose not to prescribe antibiotics, ‘arbitrary’ targets set by the system which don’t account for local considerations and pressure to change practice without corresponding support from the system. |
| Strengths, limitations, and future research directions | Purposeful sampling to ensure variation was planned but not possible; eventually, the final pragmatic sample was still relatively diverse and included CCGs and practices with a range of antibiotic prescribing levels, deprivation, and size. We included in the study all participants who expressed interest and agreed to participate; their views might have differed from those who did not volunteer to participate. As with all qualitative studies, the findings are time and context specific. |

| | |
|---|---|
| Paper Number | 77 |
| Author | Borek et al |
| Year | 2022 |
| Title | Role of locum GPs in antibiotic prescribing and stewardship: a mixed-methods study |
| Journal | BJGP |
| Region (England, Scotland, Wales, and Northern Ireland) | England |
| Study methodology | First, quantitative data on antibiotic prescribing, diagnoses, and patient and prescriber characteristics were extracted from The Health Improvement Network database. A mixed-effects logistic model was used to compare locums' and other prescribers' antibiotic prescribing for conditions that do not usually benefit from antibiotics. Second, locum GPs were interviewed about antibiotic prescribing, and their responses analysed thematically. |
| Type of document | Primary research |
| Topic or focus of paper | Antimicrobial stewardship |
| Research objectives and focus | To identify how locums' antibiotic prescribing compares with other general practice prescribers, and how they perceive their role in antibiotic prescribing and AMS. |
| Theoretical framework/theories/ underpinnings | |
| Description of intervention | |
| Rationale for paper/review | Most antibiotics are prescribed in primary care. Locum or sessional GPs (locums) are perceived as contributing to higher prescribing and may face barriers to engaging with antimicrobial stewardship (AMS). |
| What creates meaning that sustains GPs engagement with, and involvement in, work? | Locum GPs described control and flexibility in being able to choose the practices where they work. For example, they reported avoiding practices perceived as being disorganised and struggling with demand and insufficient staff (and therefore having more staff turnover and less control over prescribing quality). They perceived appropriate prescribing and good-quality care as being more difficult in such practices. |
| What solutions/recommendations are proposed? | Participants discussed how practices and commissioners should better integrate and support locums; for example, by circulating information, updates, and training opportunities to all GPs registered on Performers Lists. Some suggested that AMS-related training should be mandatory, and that local peer groups for locums should be encouraged. Finally, some suggested that locums should be better recognised as a considerable professional group, and involved in wider policy development |
| From which stakeholders' perspective? (General | Locum GPs |

| | |
|---|---|
| practitioners, patients, other HCPs) | |
| How is work-related joy constructed? (Diversity of ways individuals may think about joy) | Some participants described how locums have more control and flexibility over their work. Thus, some requested longer appointments or catch-up slots to ensure sufficient time to provide good-quality care |
| What prevents/removes work-related joy? | Other patients perceived locums as being under more pressure from patients and time owing to hourly payment. This could lead to quickly 'closing' consultations with antibiotic prescriptions.<br><br>Locums were described to receive little communication from commissioners, being less connected to professional groups and networks, and having to participate in professional training and meetings during unpaid time. Consequently, locums found keeping up to date with guidelines, evidence, and training more challenging, and having fewer opportunities for peer learning. |
| Organisational characteristics (small, large) | Participants worked across different practices and areas, which varied considerably in IT systems, workflows, and prescribing guidelines. This variation was challenging for locums who, as a result, might not always follow the local guidelines or workflows. For example, some participants reported following familiar prescribing guidelines from a different area to where they currently worked, and relied on IT prompts to indicate non-concordance with local guidelines. |
| Managing ill-defined illnesses and chronic complex multimorbidity | |
| Human connection with patients | Participants described a potential negative impact on the doctor-patient relationship of not prescribing antibiotics (which was noted to be less of a problem for locums as they are unlikely to see the same patient multiple times). |
| Human connection with peers | Participants described how locums were generally perceived, and felt to be 'just to see patients' and not a part of practice teams: "you're not part of the team and they don't make you feel part of a team, you're just there to come in and cover the session and that's all you're there to do."<br><br>Participants described locums as even more professionally isolated than practice-based GPs. |
| Human connection across other organisations (e.g., secondary care) | |
| Learning systems (e.g., competence, capability, mastery, socially situated) | |
| Additional useful information | Acute sore throat and acute cough were the two main conditions for which antibiotics were prescribed. The number of patients receiving antibiotics for these conditions was 4% higher (on an absolute scale) when consulting with locums compared with when consulting with other GPs |

| | |
|---|---|
| Summary of findings | Quantitative analysis demonstrated that locums were more likely to prescribe antibiotics than other GPs. Qualitative analysis explored the reasons behind this. Several findings are relevant to our study. Echoing other studies, not prescribing antibiotics was noted to harm the doctor-patient relationship. Locums have significant control over the practices they work in and how they work within these practices, which is viewed positively. However, some locums report greater time pressures, and working across several practices can lead to difficulties in keeping track of workflows and guidelines. Unsurprisingly, locums report feeling isolated from colleagues. |
| Strengths, limitations, and future research directions | It is possible that locums who volunteered to be interviewed had different views and experiences to non-volunteers. Finally, the findings are limited to locums practising in England, and their relevance to other parts of the UK may not necessarily be transferable. |

| | |
|---|---|
| Paper Number | 78 |
| Author | Brunton et al |
| Year | 2022 |
| Title | The challenges of integrating signposting into general practice: qualitative stakeholder perspectives on care navigation and social prescribing in primary care |
| Journal | BMC Primary Care |
| Region (England, Scotland, Wales, and Northern Ireland) | England |
| Study methodology | Interviews and a focus group with stakeholders involved in implementing active signposting staff into general practice. Data were analysed using a Template Analysis approach. |
| Type of document | Primary research |
| Topic or focus of paper | Active signposting |
| Research objectives and focus | Examine how active signposting is being integrated into general practice. |
| Theoretical framework/theories/ underpinnings | |
| Description of intervention | |
| Rationale for paper/review | A national policy focus in England to address general practice workforce issues has led to a commitment to employ significant numbers of non-general practitioner (GP) roles to redistribute workload. This paper focuses on two such roles: the care navigation (CN) and social prescribing link worker (SPLW) roles, which both aim to introduce ‘active signposting’ into primary care, to direct patients to the right professional/services at the right time and free up GP time. There is a lack of research exploring staff views of how these roles are being planned and operationalised into general practice and how signposting is being integrated into primary care. |

| | |
|---|---|
| What creates meaning that sustains GPs engagement with, and involvement in, work? | |
| What solutions/recommendations are proposed? | |
| From which stakeholders' perspective? (General practitioners, patients, other HCPs) | Stakeholders involved in CN and SPLW roles including those in service lead roles, role holders (e.g. receptionists undertaking care navigation; SPLWs) and host general practice staff (such as GPs and practice managers). |
| How is work-related joy constructed? (Diversity of ways individuals may think about joy) | Active signposting staff helped reduce GPs' workload by directing patients to other sources of help or ensuring patients saw the correct clinician. In particular, CNs were considered a 'safe' form of triage by GPs as their work is non-clinical. |
| What prevents/removes work-related joy? | Some GPs were sceptical about the usefulness of active signposting staff: "Some [colleagues] just were very sceptical. I know a GP colleague in my own health centre would say, "Are you going off singing? Some of us are practising proper medicine"... So, I think, there is some scepticism... there is some cynicism [about setting up social prescribing roles in general practice]."<br><br>Some practices described how active signposting roles and staff were forced upon them by external organisations, like CCGs.<br><br>Some stakeholders (including CNs themselves) regarded the CN role as a form of clinical triage, therefore leading to concerns regarding patient safety – this might lead to anxiety on the part of GPs. |
| Organisational characteristics (small, large) | |
| Managing ill-defined illnesses and chronic complex multimorbidity | |
| Human connection with patients | Some patients felt that signposting staff were trying to steer them away from GP appointments – which might harm the doctor-patient relationship. |
| Human connection with peers | |
| Human connection across other organisations (e.g., secondary care) | |

| | |
|---|---|
| Learning systems (e.g., competence, capability, mastery, socially situated) | |
| Additional useful information | |
| Summary of findings | This study examined the perspectives of stakeholders involved in implementing active signposting roles into general practice. A few findings are relevant to GPs' relationship with work. Although active signposting roles reduced GPs' workload, some GPs were sceptical about the usefulness and safety of these staff. Practices describes how active signposting roles could be forced on them by CCGs. Patients felt that signposting staff were trying to steer them away from GP appointments. |
| Strengths, limitations, and future research directions | This study took place in one metropolitan region within the UK and this may limit the transferability of findings to other (for example, more rural) settings. We were able to recruit SPLW stakeholder staff from five of six areas across the region where the role was operational, however our CN staff views come from only one area of five across the region where the role was operating and may be different from experiences in other Greater Manchester areas<br><br>In addition, we only managed to recruit three GPs into the study sample. We highlight this as a limitation given that the reduction of GP workload is one of the key drivers for the introduction of these signposting roles. |

| | |
|---|---|
| Paper Number | 79 |
| Author | Budd et al |
| Year | 2022 |
| Title | Can a new role, the (Trainee) Associate Psychological Practitioner (T/APP), add value in General Practice? Results from the pilot year evaluation |
| Journal | Primary Health Care Research & Development |
| Region (England, Scotland, Wales, and Northern Ireland) | England |
| Study methodology | Mixed-methods. First, to evaluate clinical outcomes, patients completed measures of wellbeing, depression, anxiety and resilience at the first session, final session and at a 4–6 week follow-up. Paired-samples t-tests were conducted comparing scores from session 1 and session 4, and session 1 and follow-up for each of the four outcome measures. Second, to evaluate acceptability, questionnaires were sent to General Practice staff, T/APPs and patients to gather qualitative and quantitative feedback on their views of the T/APP role. Quantitative responses were collated and summarised. Qualitative responses were analysed using inductive summative content analysis to identify themes. |
| Type of document | Primary research |
| Topic or focus of paper | Associate psychological practitioner role |

| | |
|---|---|
| Research objectives and focus | To evaluate the clinical impact and efficacy of the mental health prevention and promotion service, provided by the T/APPs and the acceptability of the role from the perspective of the workforce and the role to T/APPs, patients and services. |
| Theoretical framework/theories/ underpinnings | |
| Description of intervention | |
| Rationale for paper/review | The deployment of (Trainee) Associate Psychological Practitioners (T/APPs) to deliver brief psychological interventions focusing on preventing mental health deterioration and promoting emotional wellbeing in General Practice settings is a novel development in the North West of England. As the need and demand for psychological practitioners increases, new workforce supply routes are required to meet this growth. |
| What creates meaning that sustains GPs engagement with, and involvement in, work? | Thirty-two (97%) General Practice staff stated the addition of a T/APP practitioner had a positive impact on the service they worked in and the remaining 1 (3%) staff member stated they were unsure.<br><br>T/APP practitioner intervened quickly with psychiatric patients, before their problems escalated, which was viewed positively by GPs and contributed to improving clinical outcomes. These are patients that, otherwise, practices would have struggled to help – indeed, GPs described how T/APP practitioners received great feedback from patients. |
| What solutions/recommendations are proposed? | Overall, this paper suggests that T/APPs should be implemented more widely within – and outside of – the Northwest of England. |
| From which stakeholders' perspective? (General practitioners, patients, other HCPs) | Patients, GPs, and other general practice staff including practice managers. |
| How is work-related joy constructed? (Diversity of ways individuals may think about joy) | "I have somewhere to signpost for immediate access to psychoeducation/brief therapy which is invaluable" - GP<br><br>T/APP practitioners reduced workload for GPs. |
| What prevents/removes work-related joy? | Initially, there was confusion about the role of T/APP practitioners and what services they could offer.<br><br>"Some of our patients have long established mental health or serious MH issues that they were not suitable for our T/APP" |
| Organisational characteristics (small, large) | Some practices struggled to find physical space for the T/APP practitioners. |
| Managing ill-defined illnesses and chronic complex multimorbidity | |

| | |
|---|---|
| Human connection with patients | |
| Human connection with peers | T/APP practitioners were “quick to integrate” with the practice team.<br><br>An ‘in-house’ mental health expert was viewed positively: “The whole team has also benefitted from sharing good practice with, and learning from, our T/APP” |
| Human connection across other organisations (e.g., secondary care) | |
| Learning systems (e.g., competence, capability, mastery, socially situated) | |
| Additional useful information | Although not very relevant to our study, the quantitative analysis demonstrated the introduction of T/APPs to be a success: for each outcome measure, there were clinical improvements in wellbeing from session 1 to session 4 and at follow-up. |
| Summary of findings | This paper demonstrates that the introduction of T/APPs is clinically effective and acceptable to patients, practice staff and psychology graduates. Relevant to our review, it suggests that T/APPs reduced workload for GPs, successfully integrated with the practice team and provided a quick, in-house service for psychiatric patients before their problems escalated. Negative points included initial confusion about the role of T/APPs and the fact that some complex patients are unsuitable for T/APPs. |
| Strengths, limitations, and future research directions | To strengthen this evaluation further, further research is needed. In particular, a comparison against a control group would be beneficial. |

| | |
|---|---|
| Paper Number | 80 |
| Author | NHS England (prepared by Rachel Snow-Miller) |
| Year | 2015 |
| Title | Document Title: Building the Workforce – the New Deal for General Practice |
| Journal | [Official publication] |
| Region (England, Scotland, Wales, and Northern Ireland) | England |
| Study methodology | Policy document |
| Type of document | Grey literature |
| Topic or focus of paper | Workforce crisis |

| | |
|---|---|
| Research objectives and focus | Produce a ten-point action plan to address immediate issues with workforce in general practice, and to take initial steps in building the workforce for the future and new models of care. |
| Theoretical framework/theories/ underpinnings | |
| Description of intervention | |
| Rationale for paper/review | NHS England, HEE, RCGP and the BMA GPs committee (GPC) are working closely together to ensure that we have a skilled, trained and motivated workforce in general practice. |
| What creates meaning that sustains GPs engagement with, and involvement in, work? | |
| What solutions/recommendations are proposed? | There are three key strands to the ten-point plan: (1) Improving recruitment into general practice, (2) Retaining doctors within general practice and (3) Supporting those who wish to return to general practice. (2) and (3) are relevant to our study.<br><br>Retention:<br>1. Investment in retainer schemes<br>2. Improving the training capacity in general practice (to support a positive training experience for GPs)<br>3. Incentives to remain in practice (which may include a funded mentorship scheme, opportunities to develop a portfolio career towards the end of your working life, and a clearer range of career pathways)<br>4. New ways of working (identify key workforce initiatives that are known to support general practice – including e.g. physician associates, medical assistants, clinical pharmacists and advanced practitioners)<br><br>Return:<br>1. Easy return to practice (NHSE and HEE to publish a new induction and returner scheme, recognising the different needs of those returning from work overseas or from a career break)<br>2. Targeted investment in returners (offer resources to help with both the costs of returning and the cost of employing these staff) |
| From which stakeholders' perspective? (General practitioners, patients, other HCPs) | N/A |

| | |
|---|---|
| How is work-related joy constructed? (Diversity of ways individuals may think about joy) | |
| What prevents/removes work-related joy? | |
| Organisational characteristics (small, large) | |
| Managing ill-defined illnesses and chronic complex multimorbidity | |
| Human connection with patients | |
| Human connection with peers | |
| Human connection across other organisations (e.g., secondary care) | |
| Learning systems (e.g., competence, capability, mastery, socially situated) | |
| Additional useful information | |
| Summary of findings | This policy document published by NHSE outlines ten priorities for addressing workforce problems within general practice. The latter two strands (retention and return) are relevant to our review. |
| Strengths, limitations, and future research directions | |

| | |
|---|---|
| Paper Number | 81 |
| Author | Fisher et al |
| Year | 2022 |
| Title | Tackling the inverse care law: Analysis of policies to improve general practice in deprived areas since 1990 |

| | |
|---|---|
| Journal | The Health Foundation report |
| Region (England, Scotland, Wales, and Northern Ireland) | England |
| Study methodology | To identify policies addressing the inverse care law in general practice, we analysed government policy documents, reviews of policy developments, and consulted relevant experts. To identify evidence on the impact and implementation of these and other policies, we searched relevant bibliographic databases of published literature and government archives. |
| Type of document | Grey literature |
| Topic or focus of paper | Inverse care law |
| Research objectives and focus | Analyse national policies to reduce inequities in the supply of GP services in England since 1990, and evidence on their impact. |
| Theoretical framework/theories/ underpinnings | |
| Description of intervention | |
| Rationale for paper/review | The inverse care law persists in the NHS today. GP practices in more deprived areas of England are relatively underfunded, under-doctored, and perform less well on a range of quality indicators compared with practices in wealthier areas. |
| What creates meaning that sustains GPs engagement with, and involvement in, work? | |
| What solutions/recommendations are proposed? | 1. Addressing the inverse care law in general practice should become an explicit national priority, set by government. This should form part of a broader national strategy to reduce health inequalities, with clear goals for improvement and measures for tracking progress.<br>2. New policies for general practice in England should be subject to an ‘equity test’. This should consider how the policy may impact on the equitable provision of general practice, and how it will interact with other policies in place or being introduced.<br>3. An independent review of general practice funding allocations should be established to examine all income streams into general practice, how these differ between more and less deprived areas, and alternative approaches to allocating resources more fairly based on health care needs. Government should act on recommendations from the review.<br>4. Government should work with national NHS bodies to develop a medium and long-term workforce strategy for general practice, as part of a broader workforce strategy for the NHS in England. This should include plans and projections for GP retention and |

| | |
|---|---|
| | recruitment, and consider mechanisms to more equitably distribute the GP and allied health professional workforce. As part of this strategy, government should consider stronger central coordination and oversight of GP distribution in England.<br>5. Integrated care boards should be required to develop plans for reducing inequities in the provision of high-quality GP services. This should include clear goals for improvement and how they will be measured. Integrated care boards should also produce plans for data collection and analysis to effectively monitor general practice provision relative to need.<br>6. Government must commit to more rigorous testing and evaluation of policies affecting general practice – including current policies to recruit GPs in under-doctored areas. Evaluations should explicitly consider equity and be made publicly available. |
| From which stakeholders' perspective? (General practitioners, patients, other HCPs) | N/A |
| How is work-related joy constructed? (Diversity of ways individuals may think about joy) | |
| What prevents/removes work-related joy? | Practices within deprived areas are underfunded. There is wide acknowledgement that the formula used to distribute most funding to general practice does not sufficiently account for workload associated with the additional needs of people living in deprived areas. In 2020, practices serving more deprived populations received around 7% less funding per need adjusted registered patient than those serving less deprived populations.<br><br>Practices within deprived areas have a lack of doctors. After accounting for different levels of need, a GP working in a practice serving the most deprived patients will on average be responsible for the care of almost 10% more patients than a GP serving patients in more affluent areas. Inequities in GP supply widened between 2015 and 2020. By 2020, there were 1.4 fewer full-time equivalent GPs per 10,000 patients in the most deprived areas compared with the most affluent areas. This has not always been the case. Between 2004/05 and 2013/14, the number of GPs working in more deprived areas rose.<br><br>The demands made on GPs working in deprived areas are different to those working in more affluent areas – for instance, because of the complexity of patients' needs.<br><br>GPs are also under pressure from government and national NHS bodies to increase face-to-face appointments – and risk being 'named and shamed' for not doing so. |
| Organisational characteristics (small, large) | |

| | |
|---|---|
| Managing ill-defined illnesses and chronic complex multimorbidity | |
| Human connection with patients | |
| Human connection with peers | GP turnover – the proportion of staff who leave a practice in a given time period – is also higher in areas of high deprivation. |
| Human connection across other organisations (e.g., secondary care) | |
| Learning systems (e.g., competence, capability, mastery, socially situated) | |
| Additional useful information | Both QoF scores and CQC inspections suggest that practices in more deprived areas provide a worse standard of care. |
| Summary of findings | This report explains that general practices within deprived areas are under-funded and under-doctored (the inverse care law, as these populations have the greatest need for healthcare). From the perspective of our review, it is easy to see how this might erode meaning and joy for GPs currently working within these areas. To address these problems, the principle of proportionate universalism should be applied to NHS resource (funding and workforce) allocation. |
| Strengths, limitations, and future research directions | First, we focused only on policies that explicitly aimed to reduce inequities in the supply of general practice services, such as central controls on GP distribution. This enabled us to identify a small set of interventions that directly focused on tackling the inverse care law. But it means that we did not focus on how policies with broader aims – for example, changes to pay-for-performance schemes for GPs affected equity in general practice. As a result, we did not cover all policies that shape how high-quality care is distributed. We also focused on national policy changes rather than local programmes.<br><br>Second, data on the detail and implementation of policy were limited. Unclear terminology in some policy papers – for example, terms such as ‘inner-city’ or ‘under-served’ – can also make it difficult to understand which population groups were targeted by initiatives.<br><br>Third, our analysis was limited by the quantity and quality of evidence. Our literature searches identified some relevant evidence related to key policies. But many policies that we identified in our analysis were not evaluated or their evaluations were weak.<br><br>Finally, our analysis focused primarily on general practice – for example, the distribution of the number of GPs between areas of England – rather than primary care as a whole. This means that some relevant policy changes in other areas of primary care that affect quality in general practice, such as the development of new job roles, are not fully covered by our analysis. |

| | |
|---|---|
| Paper Number | 82 |
| Author | Fletcher et al |

| | |
|---|---|
| Year | 2017 |
| Title | Quitting patient care and career break intentions among general practitioners in South West England: findings of a census survey of general practitioners |
| Journal | BMJ Open |
| Region (England, Scotland, Wales, and Northern Ireland) | England |
| Study methodology | Questionnaire sent to GPs. GPs rated likelihood of leaving patient care, taking a career break and/or reducing hours on a four-point scale from 'very likely' to 'very unlikely'. Also asked about current level of morale and information on background (e.g. gender, age, GP employment status). Logistic regression was used to investigate the association between responses to questions regarding future career intentions. Initially, unadjusted associations were examined for effects attributable to the explanatory factors of gender, age, country of qualification, ethnicity, role/ position and rating of current morale. Subsequently, regression models adjusting simultaneously for all explanatory factors were used to examine adjusted associations. |
| Type of document | Primary research |
| Topic or focus of paper | GPs' career intentions |
| Research objectives and focus | Describe GPs' career intentions, especially those which might impact on GP workforce availability over the next 5 years. |
| Theoretical framework/theories/ underpinnings | |
| Description of intervention | |
| Rationale for paper/review | UK general practice is facing major problems regarding maintaining the GP workforce, with imminent GP shortages and a concomitant potential risk to patient care. |
| What creates meaning that sustains GPs engagement with, and involvement in, work? | |
| What solutions/recommendations are proposed? | Retaining the direct patient care capacity of established GPs represents an immediate challenge for healthcare planners. Recognising and addressing issues of low GP morale is an area of importance, and addressing the issue may improve how GPs feel about their work and potentially be associated with improvements in patients' experience of the care received. |
| From which stakeholders' perspective? (General practitioners, patients, other HCPs) | 2177 GPs |
| How is work-related joy constructed? (Diversity of | |

| | |
|---|---|
| ways individuals may think about joy) | |
| What prevents/removes work-related joy? | Of the 2177 GP participants included in the analysis, 473 reported a high likelihood of quitting direct patient care (‘likely’ or ‘very likely’) within the next 2 years and 863 within the next 5 years. There were 1252 participants who reported being likely or very likely to reduce hours, and 770 who reported the intention to take a career break within the next 5 years. The majority of participants also had low morale, with a substantially greater proportion of participants (1195) reporting ‘low’ or ‘very low’ levels of morale compared with only 305 reporting ‘high’ or ‘very high’ morale.<br><br>Reported quit intentions are strongly related to age, remaining low (<20%) among younger GPs (45 years and under) for quitting direct patient care in both of the next 2 and 5 years. Both of these outcomes show a sharp rise from the age of 52 years, with the proportion stating they were likely to quit in 5 years rising to almost 90% by the age of 56 years. Age was also a strong predictor of the intention to reduce hours. The proportion anticipating taking a career break as ‘likely’ was highest for the youngest GPs, especially among younger women where nearly 9 out of 10 female GPs aged 30 years reported the intention to take a career break, presumably on account of anticipated maternity leave.<br><br>After adjustment for other variables, women were substantially less likely to report intending to reduce hours spent in direct patient care, and locum GPs remained more likely to report intending to take a career break when compared with GPs in other roles.<br><br>After adjustment for other variables, GPs with ‘very low’ morale who are most likely to report intending to leave direct patient care, reduce hours in patient care or to take a career break.<br><br>Only age and practice role show any evidence of being associated with reported morale (p<0.001 for both). Those aged 50–54 years old are most likely to report low morale. In respect of role, GP partners are the most likely to report low morale, followed by salaried GPs and with locums and other GPs the least likely groups to report low morale. |
| Organisational characteristics (small, large) | |
| Managing ill-defined illnesses and chronic complex multimorbidity | |
| Human connection with patients | |
| Human connection with peers | |
| Human connection across other organisations (e.g., secondary care) | |

| | |
|---|---|
| Learning systems (e.g., competence, capability, mastery, socially situated) | |
| Additional useful information | |
| Summary of findings | The questionnaire sent to GPs in this study found that a significant proportion intend to leave direct patient care, reduce hours and/or take a career break. These career intentions were strongly related to age: older GPs were more likely to intend to leave direct patient care or reduce hours, whereas younger GPs were more likely to take a career break. After adjusting for other variables, women were less likely to report intending to reduce hours and locum GPs were more likely to intend to take a career break than GPs in other roles. GPs aged 50-54 and partner GPs were most likely to report low morale (after adjusting for other variables). |
| Strengths, limitations, and future research directions | Unlike most other similar surveys which tend to survey samples of GPs, we undertook a census survey of all GPs currently eligible to provide patient care across a large area, providing a cross-sectional overview of the quit intentions of all GPs in South West England. The response rate was high (67%), reflecting both rigorous planning and implementation of the survey and interest among GPs in the subject matter of workforce challenges. Further, we have reduced the potential for non-response bias by weighting our estimates to account for differential response rates by age, gender and practice role.<br><br>The study has a number of limitations. The survey was conducted among GPs in a single region of England which may not be entirely typical of England as a whole, for example in respect of population mix and practice setting. The region is often informally regarded as having desirable living and working conditions. It seems likely that other regions of England, with more inner-city areas with higher levels of deprivation, are likely to face even greater challenges in respect of the GP workforce than in the findings we have presented here. |

| | |
|---|---|
| Paper Number | 83 |
| Author | Forde & Collins |
| Year | 2022 |
| Title | GPs’ understanding of the wider workforce in primary care |
| Journal | BJGP |
| Region (England, Scotland, Wales, and Northern Ireland) | England |
| Study methodology | 21 newly qualified GPs on a fellowship programme were asked to rate their understanding of 16 roles and summarise their views on the key responsibilities for GPs. |
| Type of document | Primary research |
| Topic or focus of paper | Multidisciplinary teams |

| | |
|---|---|
| Research objectives and focus | Explore GPs' understanding of the wider workforce in primary care |
| Theoretical framework/theories/ underpinnings | |
| Description of intervention | |
| Rationale for paper/review | NHS England has allocated national funding for a wider multidisciplinary team with a forecast of 26 000 additional staff by 2024. |
| What creates meaning that sustains GPs engagement with, and involvement in, work? | "GPs perceived their key responsibilities to be leadership, giving advice, coordinating patient care, holistic management, and managing complexity." |
| What solutions/recommendations are proposed? | Further education is urgently needed to upskill the workforce so that they understand each others roles – for example, GPs valued listening to short talks from colleagues with other roles and, following an afternoon workshop, felt more equipped to work in wide multidisciplinary teams. |
| From which stakeholders' perspective? (General practitioners, patients, other HCPs) | |
| How is work-related joy constructed? (Diversity of ways individuals may think about joy) | |
| What prevents/removes work-related joy? | GPs' understanding of other roles was highly variable with reasonable understanding of some (for example, advanced nurse practitioners, first contact physiotherapists) but poor for others (for example, physician associates, pharmacy technicians, health and wellbeing coaches, care coordinators). |
| Organisational characteristics (small, large) | |
| Managing ill-defined illnesses and chronic complex multimorbidity | |
| Human connection with patients | |
| Human connection with peers | |

| | |
|---|---|
| Human connection across other organisations (e.g., secondary care) | |
| Learning systems (e.g., competence, capability, mastery, socially situated) | |
| Additional useful information | |
| Summary of findings | The survey conducted by this very short paper emphasised that, while GPs were enthusiastic about the diversification of the primary care workforce, they lacked understanding about some new roles (which short talks from colleagues helped address). |
| Strengths, limitations, and future research directions | |

| | |
|---|---|
| Paper Number | 84 |
| Author | Cunningham & Yeoman |
| Year | 2019 |
| Title | Recently-qualified general practitioners' perceptions and experiences of General Practice Specialty Training (GPST) in deprived areas of NHS Scotland – a qualitative study |
| Journal | Education for Primary Care |
| Region (England, Scotland, Wales, and Northern Ireland) | Scotland |
| Study methodology | Grounded theory approach. Focus groups and interviews with 14 GPs who trained in DAPs, followed by thematic analysis of responses. |
| Type of document | Primary research |
| Topic or focus of paper | General practice in deprived areas |
| Research objectives and focus | Identify the perceptions and experiences of recently-qualified GPs who had trained in Deprived Area Practices (DAPs) in Scotland |
| Theoretical framework/theories/ underpinnings | |
| Description of intervention | |

| | |
|---|---|
| Rationale for paper/review | GP recruitment and retention are problematic for deprived area practices (DAPs). Recently-qualified GPs are more likely to join practices in areas where they have trained and so it is important to encourage GP training in deprived areas |
| What creates meaning that sustains GPs engagement with, and involvement in, work? | Some GPs expressed ethical motivations for requesting training in a DAP. |
| What solutions/recommendations are proposed? | Two to three-month attachments in DAPs by GPSTs who are based in affluent areas may dispel the myths of deprived area training, and GPSTs would benefit from a wider training experience. |
| From which stakeholders' perspective? (General practitioners, patients, other HCPs) | Recently qualified GPs who had trained in DAPs |
| How is work-related joy constructed? (Diversity of ways individuals may think about joy) | |
| What prevents/removes work-related joy? | Some participants were apprehensive about starting training in a DAP. Apprehensions concerned their personal safety, with anxieties about visiting patients at home on their own, while others remembered their concerns about the challenging behaviours of patients with drug dependency. A number reflected that these safety concerns were not realised during their attachment. Their apprehensions were caused by a lack of information about the training practice and by narratives relayed to them through professional networks of friends and colleagues.<br><br>A few held reservations about working in very affluent areas where they might find challenges from some patients: those with strong health beliefs, those who wanted immediate referral to private health care appointments, and those patients who wanted to strongly influence the outcome of their consultations.<br><br>Many participants emphasised the higher workload in deprived practices: "I mean it was very hard work I think. Having worked in different practices during my rural fellowship, ******** [deprived area practice] was by far the highest workload and the patients that I saw generated much more work than the standard I would say". This heavy workload was also noted to make consultations necessarily more 'doctor-centred' than 'patient-centred', which might in turn detract from human connection with patients in these areas.<br><br>Participants believed that a persisting and heavy clinical workload would lead to fatigue and burnout. They considered that an additional secondary career could help them continue practising medicine in deprived areas. |
| Organisational characteristics (small, large) | Smaller (training) practices were perceived to be more vulnerable to fluctuations in daily workload.<br><br>Those participants in small practices, typically in a small practice-owned building, felt more vulnerable and less supported than their colleagues in larger teams accommodated in purpose-built health centres. A large primary healthcare team, consisting of several professions |

| | |
|---|---|
| | including attached community teams of district nurses and health visitors, was an asset for GPSTs. They also appreciated being in large health centres which included community care, mental health teams, and social work departments (social services). |
| Managing ill-defined illnesses and chronic complex multimorbidity | |
| Human connection with patients | Trainers (ie senior GPs) often had longstanding service in the practice and knew a lot about the lives of their patients. Trainers had extensive knowledge of the histories of patients' extended families and acted as oral historians, able to recall what had happened to patients, their families and the locality, through decades of continuing practice in some cases: "He knows exactly who everybody was, and the relationship and you know the granddaughter who took an overdose and died and the nephew that was murdered and how that impacts on every single one of them and that's invaluable, it's amazing". As is obvious from this quote, there was significant admiration amongst the trainees for senior GPs who were dedicated to their practice and worked hard for their patients.<br><br>Senior GPs were regarded as assets to the local area, that they understood life there, and were long-standing champions for patients and the services needed by them.<br><br>A dominant concern expressed by participants was that consultations with patients were different in DAPs. Patients were seen as being reluctant to share their ideas, concerns and health beliefs and also reluctant or unable to be involved in shared decision-making. Participants alluded to significant numbers of their patients being asylum-seekers who required interpreters to accompany them during consultations and that patients from abroad were used to more doctor-centred healthcare (ie all of this might reduce the ability of GPs in these areas to form a human connection with their patients). |
| Human connection with peers | Participants saw themselves as a necessary part of the working team, that they were not super-numerary, and that practices would struggle greatly without their contributions. |
| Human connection across other organisations (e.g., secondary care) | |
| Learning systems (e.g., competence, capability, mastery, socially situated) | A few participants had requested to train in a deprived area, giving a variety of reasons for their request. One felt that the practice would give a good grounding in clinical care because of the high prevalence of acute and chronic disease affecting patients. This contrasted with the perceptions held by participants that GP training in affluent areas would have higher levels of 'the worried well' and patients would consider they were ill when participants may consider otherwise (both of these points emphasise the importance of mastery to GPs).<br><br>Most participants considered that DAPs had a good reputation for the quality of their training and this attracted them.<br><br>Participants described a certain amount of pride from having trained in a deprived practice: "I kind of felt 'I have done the **** [abbreviated name for deprived area of Glasgow] I can go anywhere!" |
| Additional useful information | |

| | |
|---|---|
| Summary of findings | Although this study focusses on the experiences of GP trainees in DAPs, many of its findings are generalisable to the work of qualified GPs. In particular, participants described initial anxieties about DAPs (which were later dispelled), reservations about challenging patients in more affluent areas, workload challenges in DAPs, the benefits of larger practices, the strong human connection senior GPs in DAPs formed with their patients, the different nature of the GP consultation in DAPs and how they sought out DAPs due to better training opportunities (mastery). |
| Strengths, limitations, and future research directions | One limitation was that data was not collected from remote and rural areas in Scotland that may have problems with rural deprivation and GP training experiences may have been different as a consequence. |

| | |
|---|---|
| Paper Number | 85 |
| Author | Helen Salisbury |
| Year | 2023 |
| Title | Helen Salisbury: Playing to our strengths |
| Journal | BMJ |
| Region (England, Scotland, Wales, and Northern Ireland) | N/A |
| Study methodology | Commentary |
| Type of document | Secondary research |
| Topic or focus of paper | New roles in general practice |
| Research objectives and focus | Highlight some problems and opportunities associated with the recent diversification of the general practice team |
| Theoretical framework/theories/ underpinnings | |
| Description of intervention | |
| Rationale for paper/review | In an effort to reduce the burden on GPs, the Additional Roles Reimbursement Scheme funds non-medical staff to work alongside doctors in their surgeries, and this is specifically meant to be patient facing work. |
| What creates meaning that sustains GPs engagement with, and involvement in, work? | |

| | |
|---|---|
| What solutions/recommendations are proposed? | What would really help is not an allied health professional doing some of my consultations but relief from the tedious paperwork. A recent role added to the reimbursement scheme is that of general practice assistant. Some of the tasks in the role specification would normally be done by a healthcare assistant (phlebotomy, ECGs), but much of it is administrative. This, at last, seems to be a step in the right direction: funding and training for staff to ease the burden of paperwork that keeps many doctors so busy late into the night. |
| From which stakeholders' perspective? (General practitioners, patients, other HCPs) | GP |
| How is work-related joy constructed? (Diversity of ways individuals may think about joy) | |
| What prevents/removes work-related joy? | In many practices most of the simpler patient presentations are seen by paramedics, nurse practitioners, or physician associates. This has consequences. If all the clinically straightforward consultations are done by others, GPs are left with wall-to-wall, dawn-to-dusk complex patients, which is a sure recipe for burnout. |
| Organisational characteristics (small, large) | |
| Managing ill-defined illnesses and chronic complex multimorbidity | |
| Human connection with patients | Diverting simpler cases to other staff also disrupts continuity and our attempts to establish relationships. If I don't see the teenagers when they come about their acne, will they later trust me with their eating disorders, unplanned pregnancies, or substance misuse? |
| Human connection with peers | |
| Human connection across other organisations (e.g., secondary care) | |
| Learning systems (e.g., competence, capability, mastery, socially situated) | |
| Additional useful information | |
| Summary of findings | This short commentary explains that the recent introduction of 'new' clinical roles into general practice has left GPs with all the complex cases, and makes it more difficult for GPs to build relationships with their patients. Instead, the author explains that new roles should focus on relieving GPs of their administrative burden. |
| Strengths, limitations, and future research directions | |

| | |
|---|---|
| Paper Number | 86 |
| Author | Caesar et al |
| Year | 2013 |
| Title | Medical appraisal and revalidation: can it improve patient care? |
| Journal | Education for Primary Care |
| Region (England, Scotland, Wales, and Northern Ireland) | England |
| Study methodology | Interviews with appraisers involved in setting up a medical appraisal system in Jersey. Thematic analysis of GPs' personal summaries of appraisal and PDPs. |
| Type of document | Primary research |
| Topic or focus of paper | Appraisal and revalidation |
| Research objectives and focus | Demonstrate the impact of the introduction of an appraisal system to a community of GPs which had not previously had any opportunity to reflect on their practice through a formal appraisal with a trained peer. |
| Theoretical framework/theories/ underpinnings | |
| Description of intervention | Expert appraisers from the Wessex Deanery Appraisal and Revalidation Service travelled to Jersey to introduce a medical appraisal system. |
| Rationale for paper/review | Revalidation for all doctors in England, following the process defined in the Medical Appraisal Guide,1 finally started in December 2012 after ten years' experience of developmental appraisal in the NHS. Although the benefits of appraisal for individual doctors are recognised, there is a pressing need to look at the outcomes of appraisal not only from the individual's perspective, but also in terms of the wider context of a practice, locality and the healthcare system. |
| What creates meaning that sustains GPs engagement with, and involvement in, work? | From the appraisers' perspective, as well as the self-reported perceptions of change found in the existing literature, we found that the doctors being appraised valued the appraisers' expertise in contributing to the process of reflection on practice. It was reported that they expressed a sense of relief and reassurance at having an external perspective that could validate their practice, and that the learning needs identified were given greater credibility through recognition by the appraiser. |
| What solutions/recommendations are proposed? | |

| | |
|---|---|
| From which stakeholders' perspective? (General practitioners, patients, other HCPs) | |
| How is work-related joy constructed? (Diversity of ways individuals may think about joy) | |
| What prevents/removes work-related joy? | Appraisers undertake the professional conversation with the doctor, and in doing so they are also well placed to recognise early signs of stress, illness and performance issues, all of which can ultimately impact on patient care, the GP and the functioning of a practice. |
| Organisational characteristics (small, large) | Analysis of GPs' feedback indicated that appraisal often prompted improvements to practice infrastructure through adopting IT and patient information management systems. |
| Managing ill-defined illnesses and chronic complex multimorbidity | |
| Human connection with patients | |
| Human connection with peers | Analysis of GPs' feedback indicated that appraisal often improved practice relationships through the development of practice agreements, and in some cases their introduction. |
| Human connection across other organisations (e.g., secondary care) | |
| Learning systems (e.g., competence, capability, mastery, socially situated) | |
| Additional useful information | |
| Summary of findings | This short paper describes how GPs were reassured by external validation of their practice, and how appraisers can recognise early signs of stress amongst GPs (therefore allowing steps to be taken to mitigate this). |
| Strengths, limitations, and future research directions | There is a need to broaden the focus of research into appraisal in order to identify the outcomes of appraisal within the wider context of a practice, locality and the healthcare system. |

| | |
|---|---|
| Paper Number | 87 – appendix 5 |

| | |
|---|---|
| Author | Chadborn et al |
| Year | 2021 |
| Title | GPs' involvement to improve care quality in care homes in the UK: a realist review |
| Journal | Health Services and Delivery Research |
| Region (England, Scotland, Wales, and Northern Ireland) | UK |
| Study methodology | Realist review: evolving Standards (RAMESES) guidelines were followed, comprising literature scoping, interviews with general practitioners, iterative searches of academic databases and grey literature, and synthesis and development of overarching programme theories. |
| Type of document | Primary research |
| Topic or focus of paper | GPs & care homes |
| Research objectives and focus | 1. To develop a programme theory describing contexts in which GPs can improve care in UK care homes and in international settings similar to UK care homes.<br>2. To describe the causative mechanisms whereby GP involvement with care homes results in outcomes of service development, implementation of evidence and improved quality of care. |
| Theoretical framework/theories/ underpinnings | |
| Description of intervention | |
| Rationale for paper/review | Organising health-care services for residents living in care homes is an important area of development in the UK and elsewhere. Medical care is provided by general practitioners in the UK, and the unique arrangement of the NHS means that general practitioners are also gatekeepers to other health services. Despite recent focus on improving health care for residents, there is a lack of knowledge about the role of general practitioners. |
| What creates meaning that sustains GPs engagement with, and involvement in, work? | |
| What solutions/recommendations are proposed? | |
| From which stakeholders' perspective? (General | |

| | |
|---|---|
| practitioners, patients, other HCPs) | |
| How is work-related joy constructed? (Diversity of ways individuals may think about joy) | The ‘Difficult Conversations’ programme, as evaluated by Brighton et al, was highlighted in support of programme theory 2. This aimed to train GPs (and, indeed, other members of the care team) in communicating about end-of-life care with patients and carers. The evaluation suggested that it increased GPs’ confidence and their understanding of their role in the team. This was evidence in support of the following context and mechanism: specific GP-focused learning outcomes and training delivered as part of a multidisciplinary training programme -> GPs can understand and feel confident in their role in the team.<br><br>“. . . if you’ve got a very good pharmacist, I think GPs are only too happy for them to do some of the work” - Participant in initial GP interviews |
| What prevents/removes work-related joy? | Many of the improvement projects that participants in initial GP interviews had seen undertaken around care homes had been led by non-GP health-care professionals. This was attributed to work pressures, how clinical work was prioritised and the limited capacity of GPs to take on these kind of roles. Some respondents felt, however, that this left GPs exposed to risks associated with improvement projects and without any control over their content or progress. This respondent described an experience of working with pharmacist-led improvement, and felt that GPs had been left to ‘pick up the pieces’: “Unless the pharmacist is working at the highest level of their competency, then either they are unsafe, or the workload falls back on primary care anyway.”<br><br>Context highlighted in programme theory 1: “GP fear of adverse events and responsibility for these” |
| Organisational characteristics (small, large) | |
| Managing ill-defined illnesses and chronic complex multimorbidity | |
| Human connection with patients | “. . . when I first started in general practice, I really knew all my patients and their families and there was continuity of care and currently continuity of care is diminishing rapidly and that in itself is a potential real problem” - Participant in initial GP interviews |
| Human connection with peers | |
| Human connection across other organisations (e.g., secondary care) | |
| Learning systems (e.g., competence, capability, mastery, socially situated) | One participant in initial GP interviews described the development of a special interest group for GPs as part of the British Geriatrics Society. The group wanted to develop a GP with special interest model, in which some GPs would step away from their other responsibilities to focus on care of older people, including in care homes. The group expressed frustration that training and accreditation mechanisms did not currently enable them to fully pursue this approach in their working: “...we’ve had challenges about whether or not, in order to remain accredited, we have to do what is seen as standard general practice or Geri-GP [GP with extended role in geriatrics] work, and there is inconsistency across the country. So some appraisers are saying that GPs have to do sessions in normal primary care and other assessors are saying we don’t.” |

| | |
|---|---|
| Additional useful information | |
| Summary of findings | The findings of this realist review are predominantly focussed on how GPs can contribute to quality improvement projects in care homes. A few points in the theory-gleaning interviews are relevant to our review. GPs highlighted how they are "only too happy" for clinical pharmacists to undertake some of their work – provided they are competent. Furthermore, GPs are often left to pick up the pieces from QI programs in care homes that fail despite having little input into setting them up. Some GPs are frustrated at barriers to them developing their expertise in geriatrics and, similar to other studies, GPs highlight how continuity of care is diminishing rapidly. |
| Strengths, limitations, and future research directions | One reviewer screened all search results, but two reviewers conducted selection and data extraction steps.<br><br>Future work should describe the specific role, capacity and expertise of general practitioners, as well as the diversity of relationships between general practitioners and care homes. |

| | |
|---|---|
| Paper Number | 88 |
| Author | Cleland et al |
| Year | 2021 |
| Title | Won't you stay just a little bit longer? A discrete choice experiment of UK doctors' preferences for delaying retirement |
| Journal | Health policy |
| Region (England, Scotland, Wales, and Northern Ireland) | Scotland |
| Study methodology | We used a Discrete Choice Experiment (DCE) to estimate the relative importance of job characteristics in doctors' willingness to delay retirement, and the number of extra years they were willing to delay retirement when job characteristics improved. Interviews with doctors and a literature review were used to identify characteristics. During the DCE, respondents were first asked at what age they planned to stop working for the NHS. They were then asked to imagine they were approaching that age; each choice task asked if they would consider continuing to work beyond their intended retirement age. The choice sets were presented as a binary choice: respondents were asked if they would accept the job. |
| Type of document | Primary research |
| Topic or focus of paper | Retention |
| Research objectives and focus | Determine the relative importance of, and trade-offs between, job-related factors that influence UK doctors' decisions to retire from clinical work. |
| Theoretical framework/theories/ underpinnings | |

| | |
|---|---|
| Description of intervention | |
| Rationale for paper/review | Health systems around the world face difficulties retaining their workforce, which is exacerbated by the early retirement of experienced clinicians. Influences on retirement decisions are multi-faceted - individual (e.g., poor health, retirement of a spouse, age-related deterioration) and/or job-related (e.g., work autonomy, work flexibility, provision of rewards/recognition, relationships with work colleagues). However, to the best of our knowledge, there are no studies directly examining the relative importance of these different factors, and the trade-offs that doctors are willing to make when considering retiring or staying in the workforce. |
| What creates meaning that sustains GPs engagement with, and involvement in, work? | For GPs, the number of extra years expected to work was the most important factor in the decision to delay retirement (Relative Importance (RI)=37%) followed by job intensity (RI=25%). Both these attributes are above the percentage we would expect if all attributes were equally important. Working hours fall slightly below the equi-weight line (RI=17%) with personalised working conditions the least important (RI=8%) followed by taxation of pension (RI=13%). |
| What solutions/recommendations are proposed? | |
| From which stakeholders' perspective? (General practitioners, patients, other HCPs) | 788 GPs and hospital doctors who were British Medical Association members aged between 50 and 70 years and who had not started to draw a pension yet. |
| How is work-related joy constructed? (Diversity of ways individuals may think about joy) | |
| What prevents/removes work-related joy? | |
| Organisational characteristics (small, large) | |
| Managing ill-defined illnesses and chronic complex multimorbidity | |
| Human connection with patients | |
| Human connection with peers | |
| Human connection across other organisations (e.g., secondary care) | |

| | |
|---|---|
| Learning systems (e.g., competence, capability, mastery, socially situated) | |
| Additional useful information | For hospital doctors, the number of extra years expected to work was again the most important factor in the decision to continue working (RI=23%), followed closely by working hours (RI=22%) and on-call commitment (RI=21%) with all three of these attributes being above the equi-weight line. Personalised working conditions were least important (RI=8%, Fig. 1) followed by taxation of pension (RI=11%) and job intensity (RI=15%).<br><br>Setting all attributes at their best levels, GPs and HDs would be willing to delay their retirement plans by four and seven years, respectively. |
| Summary of findings | Relevant to our review, this paper describes the relative importance of five factors that might influence GPs' decisions to delay retirement: number of extra years, job intensity, working hours, taxation of pension and personalised working conditions. Interestingly, job intensity was a more important factor for GPs than hospital doctors, which suggests workload in general practice is uniquely intense. Furthermore, characteristics related to the nature of the job were more important than simple financial incentives. |
| Strengths, limitations, and future research directions | The DCE focused solely on job-related attributes since no job focused intervention can control for the influence of personal factors, such as personal health or a spouse that has retired, on retirement decisions. Talking to those who have retired may provide insight into how job- and personal- factors are weighted in retirement decision-making.<br><br>Response bias may be possible. |

| | |
|---|---|
| Paper Number | 89 |
| Author | Clinch & Benson |
| Year | 2013 |
| Title | Making information 'relevant': General Practitioner judgments and the production of patient involvement |
| Journal | Social Science & Medicine |
| Region (England, Scotland, Wales, and Northern Ireland) | UK |
| Study methodology | Following consultations at their practices with simulated patients about taking statins, GPs were interviewed about their underlying perceptions, values and feelings, as well as the practical and structural issues that they felt they encountered when attempting to involve patients in decisions about starting statins for the primary prevention of cardiovascular disease. They were also asked to reflect on the issue of patient involvement in healthcare decisions more generally. Subsequent thematic analysis of responses. |
| Type of document | Primary research |
| Topic or focus of paper | Patient involvement in decision-making |

| | |
|---|---|
| Research objectives and focus | Explore the ways in which General Practitioners (GPs) in the United Kingdom evaluate and construct what information they think is relevant when attempting to involve patients in decisions about their health. |
| Theoretical framework/theories/ underpinnings | Drawing on the work of Entwistle, Cribb, and Watt (2012) and models of relational autonomy (Mackenzie & Stoljar, 2000; Sherwin, 1998; Sherwin, 2000) the study takes seriously how a clinician's own values, beliefs and professional judgements, specifically with regard to the management and communication of healthcare information, can and should affect a patient's treatment preferences, choices, and critically, capabilities for autonomy. |
| Description of intervention | |
| Rationale for paper/review | Sociological work that has engaged with the issue of patient involvement in health care suggests it needs to be recognised that decision-making is not simply an individual cognitive act contained in a single consultation, but a process that is distributed across multiple encounters in relation to a range of agents and non-human actors. |
| What creates meaning that sustains GPs engagement with, and involvement in, work? | The belief in a patient's right to choose and yet still be supported in making their choice was considered to be important by many of the GPs interviewed. However, time constraints (in particular, those associated with the 10-minute consultation) were highlighted as a barrier to ensuring that patients were fully informed. Despite this, many GPs interviewed made it a priority to ensure that patients had enough time to understand the information that was being communicated to them. Using what many GPs identified as 'the beauty of general practice', GPF1 would often bring patients back for subsequent appointments until they felt informed enough to make a decision about the course of action to take. |
| What solutions/recommendations are proposed? | |
| From which stakeholders' perspective? (General practitioners, patients, other HCPs) | |
| How is work-related joy constructed? (Diversity of ways individuals may think about joy) | |
| What prevents/removes work-related joy? | One GP noted that guidelines and targets for primary prevention of cardiovascular disease were decided by specialists (e.g. cardiologists), who have a narrow view of the subject. Therefore, these guidelines did not necessarily match his views or concerns as a GP. Relating to this, a number of GPs who were interviewed were uncomfortable with the medicalising potential brought about by the primary prevention of cardiovascular disease. |
| Organisational characteristics (small, large) | |
| Managing ill-defined illnesses and chronic complex multimorbidity | |

| | |
|---|---|
| Human connection with patients | |
| Human connection with peers | |
| Human connection across other organisations (e.g., secondary care) | |
| Learning systems (e.g., competence, capability, mastery, socially situated) | |
| Additional useful information | |
| Summary of findings | This paper mainly focusses on *how* GPs' involve patients in decision making as opposed to the relationship between GPs and their work. However, relevant to our study it highlights how GPs believe strongly in supporting a patient's right to choose, although this is often constrained by time and resources. Furthermore, it notes how many guidelines are decided by specialists who do not take into account primary care considerations. |
| Strengths, limitations, and future research directions | The data relates to what the GPs say they do in practice and cannot be read as an account of what they do in practice, which could only be picked up by other research methods such as participant observation. |

| | |
|---|---|
| Paper Number | 90 |
| Author | Alba Pereira |
| Year | 2023 |
| Title | The question is: to GP or not to GP? |
| Journal | RCGP blog |
| Region (England, Scotland, Wales, and Northern Ireland) | UK |
| Study methodology | Commentary |
| Type of document | Grey literature |
| Topic or focus of paper | Reasons to be a GP |
| Research objectives and focus | Describe some reasons why the author decided to pursue general practice instead of surgery |

| | |
|---|---|
| Theoretical framework/theories/ underpinnings | |
| Description of intervention | |
| Rationale for paper/review | When moving from Brazil to England, the author switched from a surgical specialty to general practice |
| What creates meaning that sustains GPs engagement with, and involvement in, work? | |
| What solutions/recommendations are proposed? | |
| From which stakeholders' perspective? (General practitioners, patients, other HCPs) | GP |
| How is work-related joy constructed? (Diversity of ways individuals may think about joy) | Not having to work night shifts and weekends |
| What prevents/removes work-related joy? | |
| Organisational characteristics (small, large) | |
| Managing ill-defined illnesses and chronic complex multimorbidity | " GPs are the modern-day detectives. Piecing together medical puzzles that stump even the savviest search engines, from deciphering mysterious symptoms that Dr Google could never crack to playing referee in family disputes over who gets the remote control. The enigmas are endless, and the rewards of decoding them are beyond satisfying." |
| Human connection with patients | " The joy of being a GP lies in the tapestry of patient interactions. As I embarked on this unexpected journey, I discovered the unparalleled satisfaction of providing continuity of care. Building long-term relationships with patients allowed me to witness their health journey unfold, celebrate their victories, and offer support during setbacks. While the challenges are real and the days can be long, these encounters forge bonds that make GPs a cherished member of their extended family. General Practice isn't just about prescriptions; it's about being constant in their lives, an ally in their journey." |
| Human connection with peers | |

| | |
|---|---|
| Human connection across other organisations (e.g., secondary care) | |
| Learning systems (e.g., competence, capability, mastery, socially situated) | A significant motivating factor for the author to pursue general practice was the centralised GP referral system in the UK. The author recognised that GPs see the bigger picture and can consider all a patient's problems together (generalism), which is valuable both for the patient and the system. For example, the author gives the example of a Brazilian patient self-referring to a range of specialist teams for a cough, when the cough was simply a side effect of a heart medication they were taking. |
| Additional useful information | |
| Summary of findings | In this short blog, the author highlights three motivating factors for a career in general practice: solving interesting medical puzzles, long term relationships with patients and the value of generalism. |
| Strengths, limitations, and future research directions | |

| | |
|---|---|
| Paper Number | 91 |
| Author | Sturmberg et al |
| Year | 2023 |
| Title | A sustainable vision for general practice: addressing the challenges |
| Journal | Australian Journal of General Practice |
| Region (England, Scotland, Wales, and Northern Ireland) | Australia |
| Study methodology | Commentary |
| Type of document | Secondary research |
| Topic or focus of paper | Mastery |
| Research objectives and focus | Suggest a model for general practice that allows the full scope of practice to be developed while creating seamlessly integrated general practice colleges that support general practitioners on their journey to 'mastery' in their chosen discipline. |
| Theoretical framework/theories/ underpinnings | |

| | |
|---|---|
| Description of intervention | |
| Rationale for paper/review | The systemic problems and challenges of general practice within the health system require systemic solutions. |
| What creates meaning that sustains GPs engagement with, and involvement in, work? | |
| What solutions/recommendations are proposed? | |
| From which stakeholders' perspective? (General practitioners, patients, other HCPs) | |
| How is work-related joy constructed? (Diversity of ways individuals may think about joy) | |
| What prevents/removes work-related joy? | General practice work often operates in the realms of complexity and uncertainty, which can be very challenging and requires aptitude and emotional coping skills to become a resilient clinician. |
| Organisational characteristics (small, large) | |
| Managing ill-defined illnesses and chronic complex multimorbidity | |
| Human connection with patients | |
| Human connection with peers | |
| Human connection across other organisations (e.g., secondary care) | |
| Learning systems (e.g., competence, capability, mastery, socially situated) | This paper explains that general practice is often demeaned and oversimplified by non-general practitioner (GP) colleagues, as well as members of the community and politicians. In reality, general practice work is highly complex, dealing with poorly definable illness experiences and/or chronic complex multimorbidity. Neither of these problems are amenable to standardised guideline- or protocol-based approaches which have been developed for the management of single conditions by specialists. They require a generalist approach and an ability to establish a therapeutic relationship with patients. Indeed, the paper highlights that patient-centred, generalist care is equally effective as disease specialist care for common chronic conditions, and, most importantly, more cost effective. |

| | |
|---|---|
| | The paper explains that we need to support GPs on their journey from competence to capability to mastery. Capability is when GPs are able to adapt their learning to new, unforeseen circumstance. Mastery means GPs are fluent and creative, in using their skills, knowledge, and understanding in key performance challenges and contexts. Furthermore, it highlights that the most effective and efficient way for GPs to achieve mastery is a local community-based, peer-supported network (aka community of practice) that provides a safe peer learning environment in which to explore the interdependencies between patients' health, illness and disease care needs within an inevitably constrained health system.<br><br>To better support GPs in developing mastery (defined as those who are fluent, even creative, in using their knowledge, skills and understanding in key performance challenges and contexts) this paper makes several system-wide suggestions:<br>1. Implement a teaching unit that enables GPs to apply complexity understandings of health and disease to the generalist approach of patient care.<br>2. Revise all educational activities based on heutagogical principles to build competencies and capabilities for context-sensitive generalist care.<br>3. Implement the assessment-based learning framework of skills development across the career trajectory.<br>4. Establish an advocacy agency consisting of health professionals, community leaders and patient representatives charged to highlight the systemic deficiencies in primary care to the public and policy makers.<br>5. Re-establish local and regional GP networks that provide interprofessional support and assist in the equitable distribution of scarce resources.<br>6. Involve and engage interested medical students with all of the above goals, including observership on the college boards. |
| Additional useful information | |
| Summary of findings | This paper highlights the importance of the complex, patient-centred, generalist care GPs provide. In order to support GPs in attaining mastery in this area, it puts forward a number of recommendations for healthcare systems. |
| Strengths, limitations, and future research directions | |

| | |
|---|---|
| Paper Number | 92 |
| Author | Cottrell et al |

| | |
|---|---|
| Year | 2021 |
| Title | Acceptability of physician associate interns in primary care: results from a service evaluation |
| Journal | BMC Family Practice |
| Region (England, Scotland, Wales, and Northern Ireland) | England |
| Study methodology | Interviews with GPs, practice managers and PA interns at the start, mid-point and end of the SPAI scheme. Surveys were also sent to practices to obtain more detail about intern and practice characteristics. Patient acceptability data was collected by practices at one time-point using a practice delivered patient survey. Data were gathered and subsequently analysed according to domains within Lau et al.'s conceptual framework for implementing interventions in primary care. |
| Type of document | Primary research |
| Topic or focus of paper | Physician associates |
| Research objectives and focus | Establish the acceptability of PA interns within primary care. |
| Theoretical framework/theories/ underpinnings | |
| Description of intervention | The novel 1 year Staffordshire PA Internship (SPAI) scheme, introduced in 2017, was established to support the integration of PAs into primary care. PA interns concurrently worked in primary and secondary care posts, with protected weekly primary care focussed education sessions. Practices were provided with published literature available at the time to support the integration of the PA into the primary care workforce, however, they were free to utilise the PA in any of the described role that best matched their practice's needs. |
| Rationale for paper/review | Whilst PAs have been introduced as a rapid and relatively inexpensive way to grow the UK primary care workforce, little is known about their clinical effectiveness, potential for return on investment, the integration of newly qualified PAs ('new PAs') into the UK workforce nor their acceptability to patients and primary care healthcare professionals. |
| What creates meaning that sustains GPs engagement with, and involvement in, work? | |
| What solutions/recommendations are proposed? | Host-practices saw PAs as an acceptable addition to the primary care workforce. They stressed that, to optimise the primary care PA value and acceptability, a continued commitment from NHSE and HEE and accurate, realistic information for practices was needed. This should include recognition of, and support for, high-intensity GP supervision for new PAs. A commitment to a national scheme was requested, to provide standardised education, practical and financial support to make the integration of PAs into primary care a success. This was felt to be necessary for at least a few years, until a critical mass of experienced primary care PAs is realised. Host-practices wanted a collaborative network to develop a shared understanding and standardised approaches to supporting their interns. |
| From which stakeholders' perspective? (General | GPs, practice managers and PAs. |

| | |
|---|---|
| practitioners, patients, other HCPs) | |
| How is work-related joy constructed? (Diversity of ways individuals may think about joy) | |
| What prevents/removes work-related joy? | |
| Organisational characteristics (small, large) | Smaller practices struggled most to provide ‘blocked-out’ appointments to accommodate ‘just-in-time’ supervision, due to the relative impact on appointment capacity. |
| Managing ill-defined illnesses and chronic complex multimorbidity | |
| Human connection with patients | |
| Human connection with peers | While individual PAs integrated as well-liked team members, both PAs and practices lacked clarity about what a PA is, and/or how best to communicate this to patients and colleagues. Practices often recognised that PAs were dependent on GPs for supervision, but lacked clarity about the wider implications of this (e.g. the process for administration of influenza vaccinations).<br><br>Whilst PAs had some gaps in their capabilities, compared with trained GPs (the lack of prescribing capability was mentioned repeatedly), their flexibility to perform elements of different healthcare professionals’ roles was valued by GPs and practice managers. “...there’s lots of things that they can assist us with...things like phoning patients on our behalf...rather than just seeing their own patients – helping us with our patients...” (General Practitioner).<br><br>This evaluation uncovered a significant expectation-preparedness gap. All PAs expected to undertake book-on-day (BOD) appointments in primary care, though only 44% came into the internship with this experience. Similarly, 89% of interns had expected to do long-term condition reviews but only 22% had prior experience.<br><br>Upon internship completion, most host-practices were overwhelmingly positive about their PA; they demonstrated good clinical skills and performed well in a variety of scenarios. Acceptability of PAs grew as initial reservations from some primary care team members eased. Reservations often arose from nursing staff and were related to potential implications of unclear role boundaries. Other concerns, from GPs, included issues regarding prescribing, accountability and supervision time pressures. “I mean [the GP] and myself often refer to the silent workload...GP’s have got a silent workload of the prescribing...the referrals, etc....there’s a huge silent workload for the practice...all these clinical audits and stuff that requires clinical input but not particularly a GP...that’s where [the intern has] been so useful” (Practice Manager)<br><br>PA acceptability among practices was not always driven by increased clinical contact capacity. Pressure in GPs’ working days includes the ‘silent workload’; the administrative workload undertaken around full clinics, often unseen by patients and some staff. Introduction of the |

| | |
|---|---|
| | intern, and resultant increased requirement for supervision and blocked appointments, provided some additional time for GPs to undertake this workload. Further, the PAs undertook some of the GPs' silent workload.<br><br>High-intensity, close supervision was necessary for responsible GPs to build trust in the interns' capabilities. Single salaried GP in a host-practice refused to provide clinical supervision for the intern due to perceived risks. Some GPs reviewed all intern patients for an extended period but felt overburdened by this. Conversely, after a short induction period (a few weeks), other GPs allowed supervision to be determined by the intern and case complexity. Practices that had invested a lot of highlevel supervision early on appeared more satisfied at internship end. Over the year, GP face-to-face reviews reduced, and practices and interns developed efficient and sustainable supervision and support methods which, in turn, increased acceptability of supervision. "at the moment, it's not really working for the GPs because so much of their time is going in training...At the moment it's more work. .." (Practice Manager)<br><br>The inability of interns to prescribe was a commonly cited barrier to integration and certainty of the relative benefit of PAs compared with other professional groups (e.g. Nurse Practitioners). |
| Human connection across other organisations (e.g., secondary care) | |
| Learning systems (e.g., competence, capability, mastery, socially situated) | |
| Additional useful information | |
| Summary of findings | This study evaluated the acceptability of PAs within general practices. Relevant to our review, it found that although PAs can be well-liked and effective primary care team members, there are two main barriers to their implementation: role ambiguity and supervision requirements. As national bodies push to increase the numbers of PAs working in general practices, they need to recognise these barriers and provide appropriate guidance and funding. |
| Strengths, limitations, and future research directions | Limitations are that participants were not required to complete every element of data collection, that patient satisfaction data were obtained by only five of the eight practices, and that the pragmatic design (adopted to maximise engagement) meant that exact resource use (both time and financial) could not be evaluated. |

| | |
|---|---|
| Paper Number | 93 |
| Author | Cunningham et al |
| Year | 2021 |
| Title | Learning needs, preferred learning methods and learning challenges of first five general practitioners in NHS Scotland: a qualitative study |
| Journal | BMJ Open |

| | |
|---|---|
| Region (England, Scotland, Wales, and Northern Ireland) | Scotland |
| Study methodology | Focus groups and interviews with GPs within the first five years of completion of their training, followed by thematic analysis of responses. |
| Type of document | Primary research |
| Topic or focus of paper | Learning needs and methods of GPs |
| Research objectives and focus | To identify the learning needs and preferred learning methods of First5 general practitioners (GPs) in National Health Service (NHS) Scotland. |
| Theoretical framework/theories/ underpinnings | |
| Description of intervention | |
| Rationale for paper/review | |
| What creates meaning that sustains GPs engagement with, and involvement in, work? | |
| What solutions/recommendations are proposed? | First5s need additional investment in the provision and support for CPD particularly in R & R areas. This could include the development of small group learning and other resources that encourage discussion and professional socialisation—key needs for First5s. PLT is an important issue for First5s who are keen to separate learning from work and family life. NHS boards need to invest more in this area.<br><br>Those who are charged with providing CPD may come from a different generation from First5 GPs and need to take into account that learning experiences and preferences may be different. |
| From which stakeholders' perspective? (General practitioners, patients, other HCPs) | GPs |
| How is work-related joy constructed? (Diversity of ways individuals may think about joy) | Self-care, emotional intelligence and maintaining good mental health were important. Participants were conscious they were beginning a long career and would be practising for decades. |
| What prevents/removes work-related joy? | GPs perceived gaps in their training with a wide range of factors contributing to this. Participants perceived the focus of training was to cover the curriculum and pass the Member of the Royal College of General Practitioners (MRCGP) examination, which is different to the information GPs need day-to-day. Several referred to the short duration of training and the difficulties of learning everything that GPs may encounter. Some recommended that training should be extended with more time spent in general practice. Others suggested that increasing the number, quality and relevance of secondary care posts would be beneficial. "I feel, probably, that the bare minimum of GP training is done, in terms of actually time in a GP practice and learning what a GP does." Most participants perceived that training was focused on |

| | |
|---|---|
| | clinical learning, leaving knowledge and skills of business, leadership and practice management as important learning needs for the first 5 years.<br><br>Appraisal and revalidation were important learning topics not covered during training and were common learning needs for newly qualified First5s. Experienced First5s were more familiar with appraisal and revalidation, but considered the processes often detracted from useful learning, with the challenges of documenting evidence of learning activity for appraisal.<br><br>The transition from supervised trainee to independent practitioner was a time of challenge. Loss of dedicated supervision from their trainer and wider training practice, isolation from training peers, and adapting to work in a new practice or unfamiliar working environment, were common concerns. Participants recognised their limited opportunities to ask colleagues for advice compared with training. “You may even be single-handed in a practice which is such a steep learning curve coming out of ST3. You might be the only person in a practice, doing home visits completely on your own and not really having anyone to talk to, it’s like: ‘Argh!’”<br><br>Resilience was considered by many participants, who discussed factors reducing their ability to remain resilient, including heavy workloads and professional isolation. Several discussed their lack of experience of clinical presentations despite having finished training and judged clinically competent. Learning to be resilient and avoiding burnout were considered difficult, as learning needs varied from person to person. Common themes included practising realistic medicine, the management of challenging patients with complex social and mental health problems, and managing time and patient demand: “How to say no to patients, how to... We’re taught as GP trainees to be so lovely and understanding and listen to every single problem but in real life that is gonna burn you out so a way to sort of be nice but be firm.”<br><br>Maintaining a work–life balance, and incorporating continued professional development (CPD) into this was important to participants, particularly those with families. Most considered that CPD required protected learning time (PLT) within the working day, separate from clinical duties and home life.<br><br>This suggested that R & R practice was more challenging for First5s due to their inexperience. First5s sought out learning to counter this and reduce concerns about clinical uncertainty. Concerns were raised about providing services to Police Scotland particularly assessing patients in police custody with no prior training or supervision in this work. “I just think it’s a national scandal really. There’s so little regulation and there’s so little training. I really don’t think we should be able to be a police surgeon without having done at least a weekend competency course or something like that.”<br><br>Cost was the most cited barrier to attending learning events. Most found that costs were high with some arguing that specific funding should be available. This related to event fees and travel, subsistence, accommodation and additional childcare costs. Locum GPs cited potential loss of earnings when attending weekday courses. Timing and accessibility were major issues with participants divided on the optimum timing for learning events. Some viewed weekends or evening meetings as the most convenient as it did not involve taking time out of practice. |
| Organisational characteristics (small, large) | Some concerns were specific to remote and rural (R&R) practitioners. CPD access was a particular concern, and R & R First5s identified inequity in CPD provision, feeling disadvantaged and unsupported by national GP organisations. Access issues centred around cost, including travel and accommodation, and time away from practice and family. |

| | |
|---|---|
| Managing ill-defined illnesses and chronic complex multimorbidity | |
| Human connection with patients | |
| Human connection with peers | Intergenerational learning was beneficial for some participants. Contrasting learning approaches were attributed to different ages of practitioners and seen as a barrier to effective interactions between First5s and older GPs. Older GPs were considered less likely to learn using technology and more reliant on printed resources like books and journals. Older GPs were considered confident to learn through experience. First5s valued experiential learning but expressed reservations and preferred more formal learning.<br><br>First5s preferred to learn with their peers, particularly in settings such as Practice-Based Small Group Learning (PBSGL), where professional socialisation and group learning were valued. More experienced First5s valued CPD with senior colleagues as they gained confidence.<br><br>Professional socialisation of attending courses was viewed positively by R & R participants, with the exchange of ideas valued by those who were relatively isolated.<br><br>Locum GPs had fewer opportunities to learn from practice and from colleagues and had to spend time learning outside working hours to compensate. "I think that's where it's a little bit more difficult being a locum, because I will go back to the practice and actually see myself, going back to patients I've seen before to try and work out what's happened. Whereas someone that may be salaried, would be seeing these people back and reflecting on what you've done last and what you should do next, so it's definitely more difficult in that respect being a locum." |
| Human connection across other organisations (e.g., secondary care) | |
| Learning systems (e.g., competence, capability, mastery, socially situated) | Participants identified learning needs arising from reflections about patient encounters and perceived gaps in knowledge. Some had a personal interest in a topic and a desire to remain up to date. Fear, clinical uncertainty and lack of confidence were motivators for learning. |
| Additional useful information | |
| Summary of findings | Participants recognised that there were gaps in their GP training that became apparent in independent practice. They were interested in learning from an older generation of GPs but perceived that preferred learning methods differed between them. There were considerations about resilience and of the CPD challenges in R & R areas of NHS Scotland. Participants appreciated collective learning and commented about the logistics and costs of learning.<br><br>I have not extracted *most* of the data relating to preferred learning methods as it is not relevent to our study. |
| Strengths, limitations, and future research directions | Participants were diverse and held contrasting opinions producing rich data. Data saturation was achieved. |

| | |
|---|---|
| | There were limitations to the research. No participant took part from the three island boards in NHS Scotland and they may have held different views from mainland GPs. Researchers were based in NES which provides GP education and this may have biased the recruitment of participants and data analysis. Furthermore, as three of the four researchers were First5 GPs and acquainted with some of the potential participants, this may have influenced who responded to the invitation to participate, and what was said at interview. |

| | |
|---|---|
| Paper Number | 94 |
| Author | Curtis et al |
| Year | 2018 |
| Title | Written reflection in NHS appraisal: time for an alternative |
| Journal | BJGP |
| Region (England, Scotland, Wales, and Northern Ireland) | UK |
| Study methodology | Editorial |
| Type of document | Secondary research |
| Topic or focus of paper | Written reflection in appraisal |
| Research objectives and focus | Set out the problems with written reflection in NHS appraisal and suggest some alternatives |
| Theoretical framework/theories/ underpinnings | |
| Description of intervention | |
| Rationale for paper/review | Reflective writing became an obligatory part of licensing and revalidation in the UK because it is thought to provide evidence of reflective thinking and show that doctors are continuing to learn. Reflections on learning activities are verified at a yearly appraisal, helping to provide the evidence for the 5-yearly revalidation that allows an individual to continue to work as a doctor. |
| What creates meaning that sustains GPs engagement with, and involvement in, work? | Most GPs do find reflection valuable, reporting that it is embedded into their daily routines — at work, on the way to and from work, and at home. Some feel that reflection can help with processing thoughts and feelings, describing it as ‘therapeutic’ and ‘cathartic’ in helping to process emotionally difficult situations. |
| What solutions/recommendations are proposed? | The survey gave GPs a list of methods that could be used to assess their performance and learning over the year, and asked them how strongly they would favour each as a component of NHS appraisal: discussion of cases with an assessor (85%), a simple list of educational events attended (70%), submission of a video of my consultations for external assessment (55%), records of discussions in peer groups |

| | |
|---|---|
| | (52%), a multiple-choice questionnaire marked externally (42%), external assessors in my practice to watch me consulting (38%), a simulated surgery with actors (38%), trust me and rely on my professionalism (31%) and an assessment of my written reflection (28%).<br><br>The authors recommend that GPs and their appraisers should agree and choose an option for the following year that would be integral to the individual doctor's personal development plan. These options could include: the current model of a reflective learning diary, appropriate multiple-choice question tests, an observation of the GP's video-recorded consultations, an observation of the GP's video-recorded consultations, a selection of cases to discuss with the appraiser at the next appraisal. Over a 5-year cycle, a GP could choose a different option each year, culminating in a standard 'revalidation-ready' appraisal in the fifth year. This would introduce a wide variety of styles of appraisal, each year tailored to the individual GP's particular circumstances and developmental needs, but still keeping a sufficient standard of quality and consistency. |
| From which stakeholders' perspective? (General practitioners, patients, other HCPs) | GPs |
| How is work-related joy constructed? (Diversity of ways individuals may think about joy) | |
| What prevents/removes work-related joy? | The authors reference a previous UK-wide study to seek and quantify the views of GPs on the role of written reflection in learning and assessment. 1000 GPs answered an anonymous online questionnaire. In both the established and trainee GP groups, a majority stated that they did not find mandatory written reflection helpful or valuable. Respondents reported that written reflection was time consuming, with an adverse impact on other learning opportunities and their work–life balance. In both the established and trainee GP groups, a majority stated that they did not find mandatory written reflection helpful or valuable. Respondents reported that written reflection was time consuming, with an adverse impact on other learning opportunities and their work–life balance. Most worryingly, the anger caused by this process was cited by some GPs as a reason for them considering leaving their careers early. Notably, this survey took place before the Bawa-Garba case. There was widespread dismay that a reflective document from her e-portfolio, which she filled in 7 days after the incident, was fed into her trial and, more recently, concerns were raised that the GMC's approach to her case may promote a regressive move from an open culture, focused on learning from errors, to a blame culture. The authors describe how they expect this case to have increased negative feelings about written reflection in their portfolios. |
| Organisational characteristics (small, large) | |
| Managing ill-defined illnesses and chronic complex multimorbidity | |
| Human connection with patients | |

| | |
|---|---|
| Human connection with peers | |
| Human connection across other organisations (e.g., secondary care) | |
| Learning systems (e.g., competence, capability, mastery, socially situated) | |
| Additional useful information | The recent Pearson review of medical revalidation called for an increase in the quality of appraisal and a reduction in the administrative demands on doctors. It recommended that the GMC should 'hear the voice of those doctors that find revalidation to be more difficult, more time consuming and perhaps more arduous than it should be'. In its response the GMC has stated that it needs to reduce unnecessary burdens and bureaucracy for doctors. It acknowledges that some doctors feel they have to spend too long preparing for appraisal, and that, particularly to reduce the burden on GPs, the guidance on appraisal should be clarified, while at the same time increasing the value of appraisal |
| Summary of findings | The survey discussed by this editorial highlights that, for many GPs, the written reflection mandated for their annual appraisal is an onerous process rather than beneficial to their learning. The authors propose that, for at least some of the 5-year revalidation cycle, doctors should be given a choice of methods (see above) to demonstrate their learning and competence. |
| Strengths, limitations, and future research directions | |

| | |
|---|---|
| Paper Number | 95 |
| Author | Dale et al |
| Year | 2017 |
| Title | Extended training to prepare GPs for future workforce needs: a qualitative investigation of a 1-year fellowship in urgent care |
| Journal | BJGP |
| Region (England, Scotland, Wales, and Northern Ireland) | England |
| Study methodology | Interviews with GPs undertaking the fellowship on three occasions: twice during the programme and at 12 months post-completion. In addition, observational data were collected at each of the clinical settings of fellows' activities and interactions in order to contextualise the interview data. Data were analysed using a framework approach. |
| Type of document | Primary research |
| Topic or focus of paper | Extended GP training |

| | |
|---|---|
| Research objectives and focus | To investigate the experience of recently trained GPs undertaking a 1-year full-time fellowship programme designed to provide advanced skills training in urgent care, integrated care, leadership, and academic practice; and its impact on subsequent career development. |
| Theoretical framework/theories/ underpinnings | |
| Description of intervention | The year-long fellowship was delivered through three complementary elements:<br>1. 2 days a week clinical attachments, each of 4 months' duration in an emergency department, a medical admissions unit, and an ambulance service.<br>2. 2 days a week within a GP training practice<br>3. 1 day a week undertaking academic study, which included a bespoke postgraduate certificate in Urgent and Acute Care, and participation in an action learning set. |
| Rationale for paper/review | It has been argued that UK general practice specialist training should be extended to better prepare GPs for the challenges facing 21st-century health care. Evidence is needed to inform how this should occur. |
| What creates meaning that sustains GPs engagement with, and involvement in, work? | |
| What solutions/recommendations are proposed? | The fellowship scheme provides a template for advanced training and professional development combined with enriching the GPs' clinical experience that could be applied to other key interface clinical areas, such as mental health. The findings also provide evidence to inform discussion about extending general practice training to 4 years. The Shape of Training report supported by the Royal College of General Practitioners recommended that all specialist training should be a minimum of 4 years, and newly qualified GPs are reported to feel underprepared for independent practice. An extra year of training is felt necessary to ensure the increasingly complex demands of the NHS are met by a workforce with the skills and attributes to meet them. |
| From which stakeholders' perspective? (General practitioners, patients, other HCPs) | GPs. |
| How is work-related joy constructed? (Diversity of ways individuals may think about joy) | The fellows felt that they were benefiting patient care and contributing to service improvement and integration in several ways: through the impact of their clinical work, the varied interaction with colleagues in urgent/emergency care and primary care settings, and by undertaking service improvement projects. They felt that the fellowship was changing the way that they worked, their understanding of the healthcare system, and in particular their capacity to help patients receive care in the community and avoid hospital admission. "It has had a huge impact on my practice. You see the total care. If you are just working in isolation you don't see it. [As a result of the fellowship] you get a better perspective on the services and the care, and what you can do". |
| What prevents/removes work-related joy? | |

| | |
|---|---|
| Organisational characteristics (small, large) | |
| Managing ill-defined illnesses and chronic complex multimorbidity | |
| Human connection with patients | |
| Human connection with peers | |
| Human connection across other organisations (e.g., secondary care) | The opportunity to facilitate more integrated care by applying their knowledge about community resources, and encouraging communication and working relationships across organisational and professional boundaries, was viewed as a significant benefit. It was observed on several occasions that medical staff in urgent care environments approached fellows for advice about community and primary care.<br><br>Challenging the negative attitudes about general practice that are held in secondary care was viewed as an unanticipated benefit of the scheme. The leadership training was felt to prepare them for this, and their presence in acute care settings had led to secondary care colleagues becoming more appreciative of the skillset of general practice. "Everybody is working in silos and we are actually just trying to bridge that gap...you need people to act as the ambassadors of each side to go to them and say, 'well, this is what we do, do you want to know more? We don't bite, you can come as ask us questions, you know'". |
| Learning systems (e.g., competence, capability, mastery, socially situated) | All participants described having been attracted to the fellowship scheme as an early career opportunity to gain experience and skills that went beyond those obtained in vocational training, particularly in relation to understanding the roles and expertise of primary care professionals working across the urgent and emergency care system. Generally, this reflected personal ambitions to develop a portfolio career within which urgent care would be a key aspect.<br><br>The fellowship supported GPs in the development of mastery: "last night as an out-of-hours GP I had a confused old lady, lives on her own, no family around, and a GP's mind is, "Oh we've got no choice, we've got to admit the patient". But having gone through the fellowship, it made me think laterally and, with access to all this knowledge, I was able to get an emergency social worker, speak to the community emergency response team, we were able to keep the patient at home".<br><br>The weekly academic days were felt to complement the clinical skills development and were valued as providing practical, evidence-based learning opportunities and peer support. They provided an opportunity to consolidate on experiences and build confidence.<br><br>The fellows described numerous ways in which they had found that the programme was attracting interest from those undertaking vocational training: "we went there [Vocational Training Services training days] and did a talk about clinical teachings and all that and there were so many ST1s and ST2s who said they were interested in it and they said "this is new, this is so interesting, I would like to do that, it is exciting". |
| Additional useful information | There is also a need to consider the impact of such schemes on the future GP workforce. Although undertaking the fellowship may support integration of care and open up career opportunities for GPs, so making vocational training in general practice a more attractive option for |

| | |
|---|---|
| | newly qualified doctors, there is a risk that in the short term such portfolio and interface roles will exacerbate the workforce crisis facing general practice. |
| Summary of findings | Overall, this study found a high level of satisfaction with the fellowship scheme. Several points are relevant to our review. GPs were attracted to the scheme because of the opportunity to gain experience and improve their skills, emphasising the importance of mastery. When undertaking the scheme, GPs talked positively about benefitting patient care and contributing to service improvement and integration in several ways. The fellowship scheme improved their understanding of the importance of their work, as they could see the impact of GPs' decisions down the line in A&E. Furthermore, it helped GPs form relationships across organisational boundaries and challenge negative perceptions of general practice in secondary care. |
| Strengths, limitations, and future research directions | A strength of the evaluation is that all the participating GPs agreed to fully participate in interviews, so allowing the collection of longitudinal data. This enabled description of fellows' experience of the scheme at different points in the year. However, the findings need to be interpreted in the context of a relatively small cohort of GPs undertaking what was a pilot year of the scheme. The scheme was only open to a small number of individuals, and it is possible that the seven who were appointed may have been atypical in terms of interest, aptitude, and commitment. |

| | |
|---|---|
| Paper Number | 96 |
| Author | de Lusignan & Pearce |
| Year | 2018 |
| Title | Time to mandate systems that promote collaborative working with computerised medical record systems, at a time of general practitioner shortage |
| Journal | Journal of Innovation in Health Informatics |
| Region (England, Scotland, Wales, and Northern Ireland) | N/A |
| Study methodology | Editorial |
| Type of document | Secondary research |
| Topic or focus of paper | Collaborative working with computerised medical record (CMR) systems |
| Research objectives and focus | Explain that the working lives of modern GPs contain many isolating GP-computer hours, and make the case for collaborative working via CMR systems. |
| Theoretical framework/theories/ underpinnings | |

| | |
|---|---|
| Description of intervention | |
| Rationale for paper/review | Primary care internationally is computerising and the UK and Australia provide examples of countries with high levels of computer use and the challenges of recruiting and retaining the GP workforce. Insufficient thought has been given to promoting collaborative working with computerised medical record (CMR) systems. |
| What creates meaning that sustains GPs engagement with, and involvement in, work? | |
| What solutions/recommendations are proposed? | Computers have been designed to support the patient/doctor dyad – but this should now be extended to take into account the full scope of clinical-computer interactions. Our computer systems ‘know’ how long we are using them and they could record how many hours of use. They could be a platform for collaborative interactions, including off-site working, by a clinical workforce that did not need to be within our building to complete these tasks. |
| From which stakeholders’ perspective? (General practitioners, patients, other HCPs) | |
| How is work-related joy constructed? (Diversity of ways individuals may think about joy) | |
| What prevents/removes work-related joy? | Across the globe, GPs are working harder, for a combination of reasons, including patient factors (ageing population with more multimorbidity), health system factors (more information flowing into general practice), supply-side factors (including shortages in key roles) and even the demands of computers themselves. |
| Organisational characteristics (small, large) | |
| Managing ill-defined illnesses and chronic complex multimorbidity | |
| Human connection with patients | |
| Human connection with peers | When we started as general practitioners (GPs), we did much more collaboratively. We sat as a group of clinicians for morning coffee or lunch, recounted about clinical consultations and worked through the similar data as now, albeit smaller volumes. The letters we worked through could be categorised into blood and other test results, prescriptions, letters and other miscellaneous requests. How to deal with a problem took into account who the person was and your colleague’s heuristics about clinical management. This was collaborative medical work. At that time it was also easier to phone consultant colleagues and we even went into hospitals to visit patients admitted to the hospital, though admission duration was much longer then. |

| | |
|---|---|
| | Our typical practices days involve extensive work that can be isolating: they involve a lot of GP-computer hours and tasks previously done collectively are now done in isolation. Activities that once could and were maybe better carried out in a collaborative environment include: (1) Reviewing and taking action on receipt of laboratory results, (2) Authorising prescriptions for people on long-term medicines, (3) Reading, highlighting for coding and creating actions that arise from the hospital, specialist and other letters and (4) Other administrative tasks that aren't in one of the three categories above, these might include response to requests for advice from specialists (in England via formal 'Advice and Guidance' electronic link), medical reports for third parties and other ad hoc requests.<br><br>By design, many CMR systems block the collaborative affordances of the paper record, for example, simultaneous viewing or superimposed synchronous or asynchronous discussion or annotation, which has the status of exploratory discussion |
| Human connection across other organisations (e.g., secondary care) | When we started as GPs it was also easier to phone consultant colleagues and we even went into hospitals to visit patients admitted to the hospital, though admission duration was much longer then. |
| Learning systems (e.g., competence, capability, mastery, socially situated) | |
| Additional useful information | |
| Summary of findings | This editorial explains that general practice has become increasingly computerised in recent years, which means that activities like reviewing laboratory results that were previously carried out collaboratively are now carried out in isolation. To address this and increase workforce sustainability it suggests promoting collaborative working via CMR systems. Furthermore, CMR systems could be leveraged to allow more flexible, remote working – although the authors note that we need to be careful to preserve continuity of care. |
| Strengths, limitations, and future research directions | |

| | |
|---|---|
| Paper Number | 97 |
| Author | Dominic et al |
| Year | 2021 |
| Title | 'It's like juggling fire daily': Well-being, workload and burnout in the British NHS – A survey of 721 physicians |
| Journal | Work |
| Region (England, Scotland, Wales, and Northern Ireland) | UK |

| | |
|---|---|
| Study methodology | Questionnaire circulated to physicians via the Doctors Association UK website and social media. This asked 11 mandatory closed questions about their discipline, sector, grade, stress/ mental wellbeing, sources of help sought in the past, sources of help to use in future, ability to raise concerns, culture about workload/stress, future sources of help. This also asked 2 non-mandatory open questions with free text for open thoughts about ability to raise concerns about workload and culture about workload/stress. Qualitative data were analysed thematically. |
| Type of document | Primary research |
| Topic or focus of paper | Burnout |
| Research objectives and focus | Understand the causes of work-related burnout, identify what supportive resources physicians utilise, and to propose solutions. |
| Theoretical framework/theories/ underpinnings | |
| Description of intervention | |
| Rationale for paper/review | Physicians are at higher risk for burnout than workers in other fields. Burnout negatively impacts physician health, care delivery and healthcare cost. Existing studies quantify the workforce affected by burnout whilst qualitative studies use specific specialty groups limiting generalisability of solutions. This is important given increased stress during the COVID-19 pandemic. |
| What creates meaning that sustains GPs engagement with, and involvement in, work? | |
| What solutions/recommendations are proposed? | An opportunity exists for an independent, non-affiliated entity to drive change to improve uptake of support organisations through working with others to address real and perceived barriers and signpost doctors to resources. There is a need to tackle negative professional culture, inadequate resourcing and high workloads with multi-level intervention beyond individual physicians: at work unit, organisation and national level. This will require significant investment in our healthcare system to support doctors as well as healthcare staff as they manage this pandemic and future pandemics which are yet to come. |
| From which stakeholders' perspective? (General practitioners, patients, other HCPs) | 721 doctors of whom 293 were GPs. |
| How is work-related joy constructed? (Diversity of ways individuals may think about joy) | |
| What prevents/removes work-related joy? | 41% of respondents felt their wellbeing was regularly affected by job stress, and 43% felt that it was sometimes affected.<br><br>One in two (53%) respondents felt unable to raise concerns in the workplace regarding wellbeing and stress or workload: many of those who indicated they were able to raise concerns stated that they felt no solutions would be implemented. The overwhelming majority (97%, n = 695) of respondents felt the NHS has a culture of viewing excessive stress and workload as the norm. |

| | |
|---|---|
| | Respondents stated that they felt the current culture of the NHS was not conducive to positive mental wellbeing with great emphasis on building personal resilience rather than dealing with lack of system or workplace resilience such as inadequate staffing or resourcing: “Doctors are constantly being told to work longer, harder, do more, be more resilient when it is the system that is actually broken”. Some mentioned that discussing stress may be perceived as a lack of resilience, which could be detrimental to doctors’ careers.<br><br>Issues of fear of failure or innate perfectionism were prevalent amongst doctors: “No back up for clinicians who are struggling combined with perfectionist tendencies and high expectations of meeting responsibilities in doctors leads to large stress burdens.”<br><br>“The NHS only runs today on the goodwill of its staff. Every member of staff works copious hours above those paid through a duty of care and compassion for patients, but nobody extends that duty of care and compassion to the staff.” (Participant 38, GP)<br><br>Many survey participants highlighted inadequate resourcing including staff, funding, and time for providing healthcare leading to high workloads with inadequate support. Some doctors mentioned that management were or would be unsupportive in responding to poor resourcing.<br><br>“The NHS has a culture of presenteeism as well as being designed & set up with zero flexibility in staffing numbers, such that it isn’t working safely when ‘100% manned’, so even worse when staff are away for any reason.” |
| Organisational characteristics (small, large) | |
| Managing ill-defined illnesses and chronic complex multimorbidity | |
| Human connection with patients | |
| Human connection with peers | Respondents felt that senior colleagues would judge them for raising concerns regarding workplace safety which would in turn affect career progression: “How can we raise concerns when our own supervisors . . . happen to be the ones supervising you, writing your reports and your references?”<br><br>Respondents felt there was an expectation to deal with emotionally traumatic events at work and unable to talk this at work: “There is pressure not to mention your own struggle with workload, emotional trauma and exhaustion.”<br><br>Respondents raised the issue of ‘bravado’ about how much work was being done and rewarding those who prioritised efficiency at work over mental or physical health: “People bragging about how theirs was the most difficult on-call, how they got the least amount of sleep, how they did the most shifts back-to-back.” |

| | |
|---|---|
| | "There was a sense of guilt felt for putting extra work onto other colleagues if they had to take time off or seek help . . . huge levels of guilt when off sick due to colleagues having to pick up the slack, lists being cancelled and patients being disadvantaged therefore we don't take time off." Relating to this, respondents noted a culture of presenteeism in the NHS: "The NHS has a culture of presenteeism as well as being designed & set up with zero flexibility in staffing numbers, such that it isn't working safely when '100% manned', so even worse when staff are away for any reason."<br><br>A common theme was that senior doctors held unrealistic expectations of medicine forged by their difficult training and seemed to believe that all juniors must undergo difficult experience to become competent doctors. This may have been compounded by psychosocial aspects indicated in theme (a) such as stigma and bravado.<br><br>"Short rotations through training mean that you never get to know your bosses really well and they often only invest in you superficially." (Participant 217, GP)<br><br>"Things which are compulsory, such as keeping up to date with mandatory training modules, doing compulsory audits as part of training etc do not have any time allocated for them to be done, so they have to be done in 'free time'. In other careers if the company expects you to do an audit you would be allocated work time to do it." |
| Human connection across other organisations (e.g., secondary care) | |
| Learning systems (e.g., competence, capability, mastery, socially situated) | |
| Additional useful information | Of all participants, not just those who experienced burnout, most (57%, n = 409) stated that they would seek help from family or friends alongside 31% (n = 224) going to their GP and one in four (28%, n = 201) dealing with it alone. Much smaller proportions would seek help from occupational health (14%, n = 98) and the BMA (7%, n = 54). In the future, the majority (76%, n = 530 and 57%, n = 399) would seek help from family and friends and their own GP respectively if they encountered difficulty. |
| Summary of findings | This survey of NHS doctors found that the majority were affected by workplace stress. An overwhelming majority also felt that the NHS has a culture of viewing excessive workload and stress as the norm. Responses to the open questions revealed that doctors perceive NHS leaders to focus on increasing personal resilience rather than fixing system problems. Doctors felt unable to raise concerns with peers or take time off work due to stigma (and the resulting potential for perceived weakness to affect their career progression). Relating to this, doctors described a culture of presenteeism in the NHS. |
| Strengths, limitations, and future research directions | The lack of mandatory participation in the open questions could have skewed the responses towards specific recommendations. Furthermore, though we have aimed to comment on the various organisational and individual factors affecting burnout within our discussion, we are aware it may not have been possible to every factor available as these will be context dependent. |

| | |
|---|---|
| Paper Number | 98 |
| Author | Donaghy et al |
| Year | 2023 |
| Title | Primary care transformation in Scotland: qualitative evaluation of the views of national senior stakeholders and cluster quality leads |
| Journal | BJGP |
| Region (England, Scotland, Wales, and Northern Ireland) | Scotland |
| Study methodology | |
| Type of document | Primary research |
| Topic or focus of paper | GP clusters and expanded multidisciplinary teams |
| Research objectives and focus | To explore progress in the implementation of the GP contract in Scotland in terms of the MDT and cluster working. |
| Theoretical framework/theories/ underpinnings | Interviews with GP cluster quality leads (CQLs) and national primary care stakeholders (PCSs) involved in supporting the new GP contract in Scotland. Responses were analysed thematically. |
| Description of intervention | |
| Rationale for paper/review | Primary care transformation in Scotland aims to improve population health, reduce health inequalities, and reduce GP workload. Two key strategies (formalised in April 2018 in the new Scottish GP contract) are the expansion of the multidisciplinary team (MDT) and GP cluster working. These follow the abolition of the Quality and Outcomes Framework (QOF) in 2016. |
| What creates meaning that sustains GPs engagement with, and involvement in, work? | There was general support for the original aims of the primary care reforms, and the majority of PCSs and CQLs believed that ending the QOF was a positive step forward as it was seen as too numbers driven, a tick box exercise, and was too focused on single diseases. The reforms were seen as an opportunity to deliver more person centred, holistic care.<br><br>The introduction of GP clusters was seen positively by most: "...replacing QOF with GP clusters, driving quality according to the priorities of their local populations, flexibility of practices being able to develop their MDTs according to the needs of their local population—are all a really good idea.'" |
| What solutions/recommendations are proposed? | Interviewees were asked to reflect on the key learning from the introduction of the GP contract and wider primary care reforms to date. Four key issues emerged. These were, first, the need for a re-assessment of the new Scottish GP contract to promote a faster pace of change in primary care transformation. Second, there was improved public/ patient engagement about the contract/ reforms. Third, there was better workforce planning. Fourth, there were better primary care data. |

| | |
|---|---|
| | "If there isn't the workforce to deliver the strategy, then it can create real difficulties in maintaining trust in the process and maintaining morale, because people feel they've been promised something which is then not deliverable."<br><br>"Collect data from the outset so that we know if any of this has made an improvement...we won't have the data that says, actually, this has had an impact on that population group. If you're introducing reforms you need to be capturing the data and consistently reviewing that data. For me it's just been a huge gap in all of this:. |
| From which stakeholders' perspective? (General practitioners, patients, other HCPs) | 12 GP CQLs and 6 PCNs (three of whom were practicing GPs) |
| How is work-related joy constructed? (Diversity of ways individuals may think about joy) | |
| What prevents/removes work-related joy? | The lack of progress on health inequalities was especially emphasised by CQLs in the clusters in deprived areas: "The GP reforms – they are not there to support people who are in deprived areas. I don't think deprivation has been considered at all...the deprived patients are not getting any more help, any more support, any more anything. In fact they are getting less because of COVID". |
| Organisational characteristics (small, large) | However, CQLs from remote and rural areas felt that the contract and wider primary care reforms were too 'city centric' and did not take into account the unique and diverse challenges of providing primary care services in their areas: 'They're only going to make health inequalities worse because everything is getting centralised — patients are going to get less basic and less personalised services ... with the investigation and treatment rooms why should somebody have to do a 50-mile round trip to have three stitches out when there is a GP surgery just around the corner? Not everyone has a car...The cost of travel for our patients, that's a huge thing.'<br><br>"Expanding the MDT workforce sounded great. Then you discovered they'll be allocated according to practice population size … Meaning you'll be eligible for two hours of this (MDT staff) a week. That won't help at all. The reforms and models just don't fit rural areas...what we really need is more doctors. If you can't have MDT staff every day in the practice, the only person that could fulfil all those roles is the GP."<br><br>All interviewees reported poor progress around cluster working. Although good examples of cluster working were reported, most felt that clusters had not reached the expected levels of performance and proficiency. This was ascribed to four main factors: a history of limited collaboration between practices (see below), greater guidance and shared learning required (specifically not enough guidance from Scottish government on the type of quality improvement activities clusters should focus on), lack of infrastructure support for clusters and training for CQLs (e.g. lack of useful primary care data, health intelligence, support with analysis, training in quality improvement, leadership training, and evaluation) and challenges in fulfilling the extrinsic and intrinsic role of clusters (more progress on quality improvement within cluster practices than on developing integrated care in the HSCP/IJB).<br><br>All interviewees reported that the COVID-19 pandemic had had a negative impact on the implementation of the primary care reforms. Although clusters continued to meet (virtually) during the pandemic, the intrinsic and extrinsic functions largely came to a halt as general practice went into 'survival mode' and GP clusters then took on a supportive role to help GPs cope. A ubiquitous concern was the high |

| | |
|---|---|
| | volume of work that primary care is facing as Scotland emerges out of the pandemic, and that tackling this backlog of patient care will further hamper attempts to progress the primary care reforms. |
| Managing ill-defined illnesses and chronic complex multimorbidity | Because of the slow progress of reforms, almost all interviewees stated that GPs were not able to provide longer consultations for patients with complex needs (one of the key aims of the reforms).<br><br>Lack of time was regarded as a major barrier to delivering on some of the key aims of the contract, notably on GPs working as expert medical generalists in extended consultations for complex patients, and CQLs having the time to fulfil the intrinsic and extrinsic roles of clusters. For many interviewees, the shortfall in GP numbers and insufficient numbers of MDT staff were significant factors driving these time pressures. |
| Human connection with patients | Another key area of relationships was between GP practices and patients. Good patient relationships were regarded as central to good general practice, and there were concerns that some of the changes, such as patients being triaged by reception staff to a new MDT member, might damage these relationships: "You've got to invest in taking the public with you if you're going to make big changes in primary care. Otherwise they feel that things are being taken away from them. There is a real need for ongoing reassurance an communication. A lot of work still needs to be done around helping people to navigate these new models of care". |
| Human connection with peers | Expanding the MDT workforce was, in principle, welcomed as a means of improving patient care and reducing GP workload. Despite this widespread support, most interviewees reported that this had not developed at the pace and scale required.<br><br>"We've established a very good APP [advanced physiotherapy practitioner] team across all the practices in our cluster. So reduction in onward referral and in analgesics, with high patient satisfaction, high doctor satisfaction. A generally wonderful service. Similarly, primary care mental health nurses have also been wonderful, right person, right place, and right time. It's all there."<br><br>CQLs in all three sites highlighted disadvantages of new MDT staff being employed and line managed by the HSCPs rather than the actual GP practices they work in. Many also spoke about the significant training and supervision requirements GPs had to provide to new MDT staff, which they felt the new GP contract did not sufficiently address.<br><br>Cluster meetings facilitated shared learning in coping with the challenges of providing GP services during the pandemic. |
| Human connection across other organisations (e.g., secondary care) | A number of interviewees reflected that, historically, there had often been limited collaboration between GP practices in the same location, and that actually getting GPs together and cooperating in a collegiate manner was one distinct positive outcome of clusters.<br><br>Interviewees spoke frequently of the crucial importance of good relationships across a range of fronts: "We've got a CEO of the HSCF who I've never ever met, and I can't believe as a CQL that I've never met that person. I think those sort of people need to meet us. They need to understand what's going on with us"; "It does require quite significant teamwork. It means collaborative working across the whole interface. So that's the relationships with clusters and your local HSCP ... the HSCPs and the cluster leads — they need sound relationships, also the relationships with your PQLs. Building relationships takes time and it is a very iterative process". |
| Learning systems (e.g., competence, capability, mastery, socially situated) | |

| | |
|---|---|
| Additional useful information | |
| Summary of findings | This qualitative study with key national PCSs and CQLs reveals that, although most supported the aims of the new Scottish GP contract, the majority view is that progress has been slow, and this is only partly because of the pandemic. None of the CQLs (and few PCSs) felt that GP workload had reduced significantly, nor that the care of patients with complex needs had improved. Lack of time and poorly developed relationships were key barriers, as was a lack of adequate support across a range of areas including communication between clusters, guidance on areas of clinical care to focus on, primary care data, health intelligence, analysis, training in quality improvement, leadership training, and evaluation. |
| Strengths, limitations, and future research directions | A strength of the current study is that, in addition to gaining the views of senior national PCSs, clusters in three health boards were purposively sampled (in clusters serving urban deprived areas; urban mixed areas; and remote and rural areas) to gain the views of CQLs working in these diverse settings. However, because the first 12 clusters that agreed to participate in the programme of research were recruited there may be a sample bias — those who were keen to take part may have held stronger views, either positive or negative, than CQLs as a whole. A further limitation in this study is that only the views of PCSs and CQLs were sought; however, in the next phase of this work the authors will interview other GPs, MDT staff, and patients |

| | |
|---|---|
| Paper Number | 99 |
| Author | Doran et al |
| Year | 2016 |
| Title | Lost to the NHS: a mixed methods study of why GPs leave practice early in England |
| Journal | BJGP |
| Region (England, Scotland, Wales, and Northern Ireland) | England |
| Study methodology | Survey of 143 early GP leavers and interviews with 21. Survey data were analysed using descriptive statistics, and qualitative data using thematic analysis. |
| Type of document | Primary research |
| Topic or focus of paper | GP retention |
| Research objectives and focus | Explore the reasons why GPs leave general practice early. |
| Theoretical framework/theories/ underpinnings | |
| Description of intervention | |

| | |
|---|---|
| Rationale for paper/review | The loss of GPs in the early stages of their careers is contributing to the GP workforce crisis. |
| What creates meaning that sustains GPs engagement with, and involvement in, work? | Overall, participants felt that their job was not meeting expectations, particularly among GPs who had been in practice for ≥10 years, it was felt that their current job was unrecognisable from the professional role they had initially taken on. |
| What solutions/recommendations are proposed? | To improve retention of GPs in practice, NHS leaders need both to minimise the pace of administrative change and to reduce the amount of time spent by GPs on work that is not face-to-face patient care. |
| From which stakeholders' perspective? (General practitioners, patients, other HCPs) | Early career GPs who have left practice. |
| How is work-related joy constructed? (Diversity of ways individuals may think about joy) | |
| What prevents/removes work-related joy? | Participants described a radically altered working environment caused by an unprecedented increase in organisational changes, many of which were felt to be made without any 'long-term vision' (GP19), and for 'little health gain' (GP15). Unhappiness with day-to-day life as a GP was indicated by 78.9% of survey responders, in particular, changes to the role of the GP (43.7%): "Cases were getting more complicated, more was being transferred from the responsibility of the hospital to the responsibility of GPs...I was spending more and more time doing administrative things and less and less time being able to devote my mental attention to the patients in front of me. I just felt more and more stretched"<br><br>According to participants, continual organisational changes fundamentally altered their professional role to a 'government clerk' or a 'data clerk for public health and for management'(GP15).<br><br>For most participants, the introduction of the Quality and Outcomes Framework (QOF) marked a defining point where 'modern medicine' became a "more target driven culture".<br><br>Participants perceived that management targets, regulations, and guidelines impinged on their day-to-day work in general practice, increasing their workload. Fifty per cent of survey responders thought that having too high a non-clinical workload was a factor in their decision to leave practice early, while 83.8% said that aspects relating to pressure of work featured in their decision.<br><br>Strategies to cope with increasing pressures included staying late at work, taking work home, or changing their appointment times.<br><br>Factors relating to patients and the media were cited by 63.4% of survey responders. Concerns about media attacks on the medical profession were indicated more frequently (57.0%) than fear of litigation (25.4%) or complaints (17.6%): "We were targeted in a completely |

| | |
|---|---|
| | unsympathetic light [...] without any recognition of what as a profession we gave to the public really and it did, over time, become very wearing.'<br><br>Pressures, intensified by a perceived 'blame culture', led to disillusionment and a raised anxiety about the risk of making a mistake.<br><br>Participants described a series of conditions which they felt contributed to an increasingly pressurised working environment. These included organisational changes resulting in a clash of values and diminishing professional autonomy as health care became more centralised, standardised, and depersonalised; an unprecedented increase in administrative workload; and a lack of support not only from government, but also across services and the wider community due to an ongoing negative media campaign.<br><br>One-third of the survey sample experienced ill health, including stress and anxiety disorder. Burnout was cited by 38.0% of the survey responders, although some participants self-diagnosed the early symptoms of burnout.<br><br>Personal factors were cited by 90.8% of survey responders, in particular, feeling overworked (53.5%), a wish to improve their work–life balance (49.3%), the work being too stressful (43.0%), and lack of enjoyment of the work (41.5%). |
| Organisational characteristics (small, large) | |
| Managing ill-defined illnesses and chronic complex multimorbidity | |
| Human connection with patients | The increasing influx of administrative tasks left many feeling professionally compromised as they came to face conflicting priorities in the consulting room. Among survey responders, 55.6% stated that the goalposts were being moved too often and 52.1% disliked the 'target-driven' approach to patient care: "you spent more time ticking boxes than you did talking to the patients sometimes...that put more stress on me and I felt it affected my rapport with the patients".<br><br>Administrative workload has led to a fundamental change in the doctor-patient relationship: "You see it does change the doctor-patient relationship because it changes how you react to people and how you interact with people. I mean it's obvious stuff, but when you're really stressed and you've still got 15 people to see, you don't have the time for people, you don't have the interest".<br><br>Some GPs grew to "hate everything around the job": "passionately adoring my work and my patients, I mean, really I can't tell you how much I miss them. Absolutely loved the actual job, but everything around the job I hated". |
| Human connection with peers | Participants described conflicts within their practices over funding, career progression, flexible hours, and workload distribution. These issues within practices were exacerbated by the lack of time for more informal interactions and support among colleagues. Although all participants felt supported during their training and registrar years, once fully qualified they became increasingly isolated in practice.<br><br>"There is a really aggressive, vicious, bullying culture that permeates management in the NHS. That then flows all the way down to whoever your locality middle managers are. It's a dreadful, awful, bullying culture and to shift from that to a non-overseeing, facilitative, hands-off, |

| | |
|---|---|
| | trusting culture is...I don't know if people are capable of that cultural shift". In light of this reported culture of bullying several interviewees expressed the need for more support, particularly in the form of a more 'robust' occupational health service for doctors. Among survey responders, unhappiness with their professional culture was important for 61%, while 44% highlighted the feeling of a loss of autonomy and professional control. |
| Human connection across other organisations (e.g., secondary care) | As referral systems became more complex and hospitals more specialised, interviewees experienced a more fragmented and depersonalised healthcare system that was increasingly challenging for them to navigate. |
| Learning systems (e.g., competence, capability, mastery, socially situated) | For the majority of participants, attempts to juggle what they saw as 'impossible targets' with 'unrealistic appointment times' (GP12) detracted from delivering good patient care.<br><br>Participants perceived that management targets, regulations, and guidelines detracted from their ability to provide good care: "I felt I was cutting corners, I felt I wasn't offering a good service unfortunately".<br><br>'Patients are dissatisfied … because they're not being given sufficient time to give their history properly and be investigated at the primary care level ... there isn't that reflective quality that allows differential diagnosis, use of time, the use of your personal knowledge of the individual and their social circumstances to be applied.' (GP9) |
| Additional useful information | |
| Summary of findings | Participants had been attracted to GP work in the expectation that it would offer continuity of patient care, professional autonomy, and flexibility in working hours, along with the intellectual challenges inherent in problem solving. However, participants described multiple factors leading to their decision to leave practice early in their careers. The extent and rapidity of organisational changes to the NHS, which had led to an increase in day-to-day administrative tasks and overall workload, were perceived by participants to have fundamentally changed the doctor–patient relationship: the very hallmark of general practice. Lack of time with patients meant the ability to practise patient-centred continuity of care was perceived to be compromised and, with it, the GPs' professional autonomy and values, resulting in diminished job satisfaction. Once their job satisfaction had become negatively impacted, the combined pressures of increased patient demand and the negative media portrayal left many feeling unsupported and vulnerable to burnout and ill health. |
| Strengths, limitations, and future research directions | Participants were self-selecting and therefore may have had particularly strong views. However, interviewees represented a maximum variation sample in terms of age, number of years as practising GPs, and geographical location. |

| | |
|---|---|
| Paper Number | 100 |
| Author | Drennan et al |
| Year | 2014 |
| Title | Investigating the contribution of physician assistants to primary care in England: a mixed-methods study |
| Journal | Health Services and Delivery Research |

| | |
|---|---|
| Region (England, Scotland, Wales, and Northern Ireland) | England |
| Study methodology | Macro- and meso- levels of the healthcare system. A rapid review of empirical evidence of the contribution of PAs to primary care was undertaken. A documentary analysis was conducted of published commentaries and of UK workforce policy. A scoping survey, using semi-structured interviews, was undertaken of key informants in professional bodies, NHS workforce planning organisations, patient organisations, higher education institutions and commissioning bodies in England and Wales.<br><br>Micro-level investigation. Comparative case study design with six general practices employing PAs and six not employing PAs, matched by practice size, sociodemographics and health economy setting. Multiple methods were used to collect data. GPs, PAs, nurses and administrative practice staff were interviewed. Patients were surveyed and interviewed. The patient records of those reconsulting for the same problem within 2 weeks were reviewed. Consultations were videoed and analysed using a validated tool for assessing GP competency. An economic analysis was conducted at two levels: practice team configurations and costs; and patient-level comparison of the contribution and costs of GP and PA consultations.<br><br>Emerging findings were tested with advisory group members, patient and public involvement group, and participants of both phases.<br><br>Did not extract evidence from the scoping review as only 1 study out of 49 was from the UK. Also did not extract evidence from patient surveys or analysis of consultations, as not relevant to our review. |
| Type of document | Primary research |
| Topic or focus of paper | Physician associates |
| Research objectives and focus | To investigate the contribution of PAs to the delivery of patient care in primary care services in England. The specific research questions addressed were:<br>1. How are PAs deployed in general practice and what is the impact of including PAs in general practice teams on the patients' experiences and outcomes?<br>2. What is the impact of including the PAs in general practice teams on the organisation of general practice, the working practices of other professionals, relationships with these professionals and the practice costs?<br>3. What factors support or inhibit the inclusion of PAs as part of English general practice teams at the local and macro level? |
| Theoretical framework/theories/ underpinnings | |
| Description of intervention | |
| Rationale for paper/review | |
| What creates meaning that sustains GPs engagement with, and involvement in, work? | |

| | |
|---|---|
| What solutions/recommendations are proposed? | 1. Physician assistants have the potential to be a competent and flexible addition to current and future primary care staffing teams. Patients, the public and primary care professionals need more information to understand both the role and its potential contribution to the NHS. There may be a role for professional organisations, general practices and local primary care organisations in doing this.<br>2. General practitioners and managers of primary care services should consider the inclusion of PAs within their staffing configurations, and CCGs (and their equivalents outside England) should consider their potential in future service and innovation planning.<br>3. The efficiency of PAs in primary care is likely to be enhanced by the legal authority to prescribe, which can be achieved only by including PAs in the regulatory framework for health-care professionals. This has implications for policy with regard to regulatory frameworks for health professionals in the UK.<br>4. In order for there to be a supply of PAs, there needs to be cognisance of this group in health profession education commissioning at a local and national levels (Health Education for England and its equivalent in other UK countries). Educational commissioners at local and national levels should consider the contribution PAs could make to a future flexible primary care workforce. |
| From which stakeholders' perspective? (General practitioners, patients, other HCPs) | Very wide range of stakeholders including GPs, national policymakers, PAs and patients. |
| How is work-related joy constructed? (Diversity of ways individuals may think about joy) | |
| What prevents/removes work-related joy? | |
| Organisational characteristics (small, large) | |
| Managing ill-defined illnesses and chronic complex multimorbidity | |
| Human connection with patients | |
| Human connection with peers | In the review of commentaries, negative comments from GPs and nurses about PAs included the transferability of a US model to a UK setting; confusion for the public when faced with multiple roles; concern that PAs were not cost-effective in general practice when compared |

| | |
|---|---|
| | with GPs; and a viewpoint that nurses and NPs fulfilled this role in the UK health-care workforce and offered greater value to patients than another professional trained in the medical model. It should be noted that none of these GPs and nurses had any direct experience of PAs.<br><br>For survey respondents who could envisage a role for PAs in primary care, that role tended to be one of triage or dealing with patients with minor, self-limiting illnesses in general practices and other first-contact settings such as walk-in centres. The explanations for this were that this was the area of patient pressure on the practices, but also that the role could free up the doctors' time to deal with patients' more complex problems or with other types of activities such as commissioning.<br><br>Most used the evidence from studies of NPs, reporting that compared with doctors (particularly in general practice) their consultations with patients were longer, resulting in no financial gain to the service. They considered that the evidence about PAs would be similar: "Practices have tried nurse practitioners and generally they take twice as long to see the patients and don't come to the conclusion or the management as quickly. So you actually often end up when you look at the, and there's been published papers to show it, when you look at the cost actually the nurse practitioner doesn't become any cheaper." - GP.<br><br>Survey respondents also expressed concerns that PAs were not a professional group regulated by legislation, although, as one participant from a patients' organisation pointed out, the existence of a regulatory body would not necessarily result in regular checks of individual competence.<br><br>"So you know if you talk about physician assistants I actually don't think many people in this country really understand what they are." - GP<br><br>In some practices, PAs saw mostly the same range of patients as doctors. However, in others, more nuanced systems had been developed so that the GPs and PA were seeing different types of patients. GPs were more likely to see same day/urgent appointments and complex patients.<br><br>It was observed by a number of case study participants that the difference between nurses and PAs in their chronic disease management was often recognisable through the point at which the patient was referred back to the doctor. PAs were described as often managing more problems and transition points, whereas the nurses referred back for any problem or change: "She [the PA] does all the things the nurse practitioner will do on the day but she also does a couple of other things because she's covered the training. So she is happy to see straightforward depressions, which the nurses, none of our nurse are prepared to at present . . . And she is happy to see back pains also which the nurses haven't covered so they don't do that. Another difference is the PA will do some of her own referrals [to secondary care] whereas the nurse practitioner would refer the patient to the doctor who would be responsible for doing the referral." - GP<br><br>The practices varied in the length of the appointment slots they assigned to PAs. Most – but not all – assigned longer slots (15 minutes) to PAs than to GPs (10 minutes), but often shorter appointment times than those allocated for nurses.<br><br>PAs were described as having been given more authority to order tests and make referrals to secondary care (although some needed countersigning in some areas by the GP) than the nurses, who mainly described usual practice as referral back to the GP. |

| | |
|---|---|
| | Physician assistants are not authorised to prescribe medicines in the UK. The lack of ability to prescribe was reported to cause frustration and additional time to the PAs, the GPs and the patients. It was evident that the GPs developed trust in the clinical competency of the PAs over time, which had the effect of reducing the amount of time they spent reviewing consultations before signing prescriptions.<br><br>The GPs, practice managers and PAs all described an induction process of establishing the PAs' competency in clinical work, confidence in working in the practice team and with practice systems through additional training and supervision sessions offered by the GP. There was also agreement from most participants, across all professional roles, as to the importance of ongoing supervision and mentorship: "Dr [name] routinely and regularly looks at my work and patient care notes to see if there is anything he needs to reassess or talk to me about." A number of practice managers described how the appointment slots for the supervising GP were tailored to ensure that there were free slots to compensate for interruptions and discussions.<br><br>Knowledge of PAs among other practice staff, prior to the employment of the first PA, was reported to be negligible.<br><br>In the observed clinical meetings it was evident that the PAs were considered integral to the practice team. They contributed information, opinions, and potential solutions and took proactive roles in administration and patient management.<br><br>GP partners and practice managers reported trying to use staff most efficiently to cover the work required: "There is an active attempt to feed work down to the person most appropriate to do it if you see what I mean, I mean it's pointless paying your mostly highly paid and qualified staff to do tasks that can be done by others" - GP<br><br>"Does [cost] come into it, yes it certainly does. I mean if you could have an experienced PA performing a largely similar role to a salaried doctor then clearly the practice is able to employ more staff to achieve, to provide consultations to patients because the money goes further. However, you have to set against that the fact that there's supervision time, there's education time that's required for PAs, there's the fact that they aren't fully registered here." - GP.<br><br>There was no consensus on the likely replacement of doctors by PAs or NPs in substantial numbers. Some GPs and practice managers suggested that GPs were required to appropriately identify the very small percentage of urgent and complex problems presented which NPs or PA were not trained to identify. Others argued that using the GP as the first contact, particularly in triage systems, was more effective and cost-efficient owing to their expertise levels and ability to take risk. A further argument was that PAs and NPs can be more costly than GPs due to supervision (see quote above). One GP noted that making these types of judgements was based on the individual capabilities of PAs and the requirements of a practice and, consequently, it was hard to generalise to an occupational group. Those who commented that there was a place for replacing doctors with PAs also commented that there were problems to be overcome. Chief among these was the lack of regulation for PAs combined with the consequent lack of authority to prescribe. A second issue was the lack of a 'supply' or 'pool' of UK PAs to recruit from, combined with reported problems in attracting US PAs.<br><br>Analysis of the consultation records of PAs in the case study practices demonstrated that PAs saw significantly younger patients than GPs, but (interestingly) patients with a greater deprivation. Descriptively, GPs appear to be consulted by patients whose average number of QOF registers is higher, who have attended the practice more often in the 3 months preceding the index event and who have had a higher number |

| | |
|---|---|
| | of repeat prescriptions in the year preceding the index event than those patients consulting PAs. PAs tended to see more patients with problems classified as 'minor/symptoms' than GPs, while the GPs saw more patients classified as 'chronic' or 'acute.<br><br>For the 1812 consultations with a time available, PA consultations were seen to have a significantly longer mean consultation time, 5.5 minutes in excess of the GP consultations, in unadjusted comparisons. Physician assistants recorded that advice was taken from a supervising doctor in 53 (35.3%) of their index consultations, whether this was at the time of the consultation or later routine quality assurance by the doctor.<br><br>Comparison of the mean number of reconsultations for the same or linked, same or same at practice problems for PA or GP index consultations shows equivalence against the study's primary outcome for PAs and GPs, with no difference found in the rates of reconsultation for the two groups, unadjusted or adjusted for potential confounding factors. |
| Human connection across other organisations (e.g., secondary care) | |
| Learning systems (e.g., competence, capability, mastery, socially situated) | |
| Additional useful information | Survey respondents were in general agreement as to the direction of workforce travel in primary care. These included (1) greater numbers of staff (medical, nursing, allied health professions, support staff and administrative staff) to provide the range of services moving from hospital to primary care settings and accessible for longer hours, 7 days a week, (2) increased availability in primary care of medical specialists in ambulatory sensitive conditions, (3) increased subspecialism by GPs, and (4) increased stratification in some occupational groups, for example more clinical nurse specialists and advanced NPs, fewer registered nurses and more HCAs.<br><br>Work diaries indicated that the majority of the PAs' working hours were spent in providing clinical consultations. These were mostly the follow-up of patients with long-term conditions. The most frequently reported non-clinical activities by PAs were dealing with test results and referrals to other professionals.<br><br>Controlling for other covariates (set at their mean values), the difference in consultation times for the average patient (age 38.7 years) increased to 5.8 minutes (17.03 minutes vs. 11.23 minutes) minutes. This increase reflects, in part, the fact that, compared with GPs, a higher proportion of PA consultations are for minor symptoms and a lower proportion are for chronic and for acute conditions Although GPs see more patients per hour (three patients for every two seen by PAs), their salary and related costs are more than twice those of PAs, and so PA consultations are still less costly. Using national figures, GP and PA consultations cost £3.08 and £1.67 per minute (based on consultations of 11.7 minutes at £36 and 15 minutes at £25), giving consultation costs within this study (for the adjusted consultation lengths of 17.03 minutes and 11.23 minutes) of £34.36 and £28.14, respectively, a saving of £6.22. Physician assistants thus appear to deliver similar outcomes from same-day consultations as GPs, at lower cost. However, the apparent cost advantage of PAs is eroded when broader factors are taken into consideration. GPs spend time on training and supervising PAs; in addition, GP consultations are interrupted for signing PA prescriptions and when PAs seek advice about patients. |

| | |
|---|---|
| Summary of findings | GPs without any direct experience of PAs reported negative perceptions, including concerns that PAs were not regulated or were not cost effective when compared to GPs. However, in practices included in the comparative case study, PAs were considered integral to the practice team. PAs saw more patients with ‘minor’ symptoms, whereas GPs saw more complex acute or chronic patients (although this triaging of patients was not the case in all practices. Economic analysis suggested that PA consultations are less costly than those of GPs, taking salary, caseload and length of consultation/ productivity into account. However, this cost advantage is likely to be eroded when the training and supervision of PAs by GPs is taken into account. |
| Strengths, limitations, and future research directions | General practices are very busy, as small organisations, and the study was shaped by the extent to which the practice staff were able to participate in the anticipated way, in the face of competing demands and priorities. The variation in the completion of workload diary sheets was an example of this. There are limitations in the data collection tools. Using the anonymised clinical records reduced the reliance on clinical staff having to undertake additional work and ensured that we collected data that were representative of usual practice. However, this could be open to the challenge that there was a lack of study-prescribed uniformity in data collected.<br><br>The case study element was not a trial which randomised patients to PA or GP, but rather was a pragmatic approach that was adopted based on usual systems in place in the participating practices. The sample size was relatively small, although well powered for the analyses undertaken. |

| | |
|---|---|
| Paper Number | 101 |
| Author | Ferguson et al |
| Year | 2020 |
| Title | Exploring ‘work–life balance’ at appraisal and how this links with organisational support |
| Journal | BJGP |
| Region (England, Scotland, Wales, and Northern Ireland) | UK |
| Study methodology | Quantitative and qualitative analysis of WLB questionnaire included on Wessex appraisal platform.<br><br>Commentary on how ‘the Mayo clinic program’ might support UK doctors’ wellbeing. |
| Type of document | Primary & secondary research |
| Topic or focus of paper | Burnout |
| Research objectives and focus | Make the case for inquiring about work-life balance (WLB) during GP appraisal, and suggest that the UK could learn from the experience of ‘the Mayo Clinic program’ in the US, which has improved doctors’ wellbeing with organisational support following nine steps (see below). |

| | |
|---|---|
| Theoretical framework/theories/ underpinnings | |
| Description of intervention | In Wessex an optional WLB questionnaire was added to an online appraisal platform, to encourage doctors to think about their own WLB. |
| Rationale for paper/review | The ‘work–life balance’ (WLB) issues of doctors are often neglected, contributing to the problems of recruitment and retention. The 2017 National GP Worklife Survey shows only 49.9% of GPs are satisfied with work, 92.3% report pressure from ‘increasing workloads’, and 46.0% of GPs plan to leave within the next 5 years. |
| What creates meaning that sustains GPs engagement with, and involvement in, work? | The optional WLB questionnaire was used by 2602 GPs (49% response rate) who undertook online appraisal suggesting it is popular amongst GPs.<br><br>88% of 67 GPs who completed qualitative evaluation of the WLB questionnaire found the WLB questions useful for reflection by raising awareness, prompting reflection, and quantifying WLB: ‘It made me think more about the different elements of WLB’; ‘It allows GPs’ voices to be heard’; ‘feels taboo to discuss this subject...opens it up for discussion’; ‘elephant in the room’; and ‘it would be good for a summary to be sent to the media ... to highlight the crisis in the GP workforce’. 28% were surprised by their results; 22% made changes with lifestyle improvements, exercising and changing their work patterns.<br><br>The WLB questionnaire is a step towards ‘acknowledgement and assessingWLB‘, it is popular, the cost minimal, and it could be extended to all GPs. |
| What solutions/recommendations are proposed? | The Mayo clinic program comprises nine steps:<br>1. Acknowledge and assess the problem (e.g. WLB questionnaire)<br>2. Harness the power of leadership (e.g. presentations at the RCGP conference)<br>3. Develop and implement targeted interventions (e.g. Review roles, delegating work, using clerical staff effectively, using integrated e-records, and devolving work to the healthcare team to allow more time for better patient care)<br>4. Cultivate community at work (e.g. formal meetings and informal coffee breaks)<br>5. Use rewards and incentives wisely (e.g. opportunities for flexible working)<br>6. Align values and strengthen culture (e.g. organisations promote health and wellbeing)<br>7. Promote flexibility and work-life integration<br>8. Provide resources to promote resilience and self-care (e.g. walking to collect patients from the waiting room, practice walks, park runs, and using bicycles for home visits)<br>9. Facilitate research and support for doctors’ wellbeing (e.g. include WLB in appraisal)<br><br>The Mayo Clinic Program has reduced doctor burnout from 48.8% to 32.9%. |

| | |
|---|---|
| From which stakeholders' perspective? (General practitioners, patients, other HCPs) | GPs |
| How is work-related joy constructed? (Diversity of ways individuals may think about joy) | |
| What prevents/removes work-related joy? | |
| Organisational characteristics (small, large) | |
| Managing ill-defined illnesses and chronic complex multimorbidity | |
| Human connection with patients | |
| Human connection with peers | |
| Human connection across other organisations (e.g., secondary care) | |
| Learning systems (e.g., competence, capability, mastery, socially situated) | |
| Additional useful information | |
| Summary of findings | Including the optional WLB questionnaire with online appraisal proved popular amongst GPs. This may form part of a wider strategy mirroring 'the Mayo Clinic program' which has been successful in the USA. |
| Strengths, limitations, and future research directions | The responders to the WLB questionnaire may be self-selecting with poor WLB. |

| | |
|---|---|
| Paper Number | 102 |

| | |
|---|---|
| Author | Richard Fieldhouse |
| Year | 2017 |
| Title | My locum life: 20 years trying to make myself obsolete |
| Journal | BJGP |
| Region (England, Scotland, Wales, and Northern Ireland) | UK |
| Study methodology | Commentary |
| Type of document | Secondary research |
| Topic or focus of paper | Locum GPs |
| Research objectives and focus | Describe some of the problems locum GPs face and how |
| Theoretical framework/theories/ underpinnings | |
| Description of intervention | |
| Rationale for paper/review | The *General Practice Forward View* suggests that locums are part of the problem rather than part of the solution. Reports are coming in that previous hard-won support structures for sessional GPs are being dismantled. |
| What creates meaning that sustains GPs engagement with, and involvement in, work? | The author suggests that both prejudice against locums and patient safety risks associated with locums have decreased over the last 20 years, with the founding of the National Association of Sessional GPs and the proper recognition of GP locums by professional bodies like the BMA. |
| What solutions/recommendations are proposed? | The author explains that we are not currently making the most of practice-based GPs or the estimated 17 000 GPs working as locums. A revised model could see both being properly enabled to be the ‘named GP’ for patients with chronic or complex needs requiring relationship as well as management continuity. Thirty-minute appointments would allow for holistic care, with each named GP at the heart of a comprehensive team, all responsible for the same named patients and all with excellent links with any secondary care teams. And in symbiosis with these practice-based GPs, we’d see small, smart, agile, highly engaged GPs flexibly deployed to manage patients who still need excellent continuity of management, but don’t have ongoing, complex problems requiring a ‘named GP’ approach. GPs with special interests or skills can put these into use across different settings.<br><br>Furthermore, the author argues that we are never going back to 24/7 cradle-to-grave general practice. We must accept as normal that patients will see different GPs at different times — sometimes this will be a locum so partners can go on holiday or be ill. So we must work hard to develop excellent ‘continuity of management’. |
| From which stakeholders’ perspective? (General | (Locum) GP |

| | |
|---|---|
| practitioners, patients, other HCPs) | |
| How is work-related joy constructed? (Diversity of ways individuals may think about joy) | |
| What prevents/removes work-related joy? | Common sentiments expressed towards locum GPs include “no doctors here today – just the locum!” and “when are you going to find a proper job?”.<br><br>The author notes that locums can post a patient safety risk. “I am exposed to risk and frustrating amounts of waste, often related to lack of access to basic information about how each practice worked and interacted with other services. These ‘information gaps’ would pile up, causing delay and, at worst, the potential for serious lapses of care”. |
| Organisational characteristics (small, large) | |
| Managing ill-defined illnesses and chronic complex multimorbidity | |
| Human connection with patients | |
| Human connection with peers | |
| Human connection across other organisations (e.g., secondary care) | |
| Learning systems (e.g., competence, capability, mastery, socially situated) | |
| Additional useful information | |
| Summary of findings | Historically, locum GPs were subject to prejudice and posed patient safety risks. These problems have lessened over the last 20 years, in a large part due to the formation of the National Association of Sessional GPs. Going forward, we need to recognise the important role that flexible, locum GPs play, and empower them to achieve ‘continuity of management’. |
| Strengths, limitations, and future research directions | |

| | |
|---|---|
| Paper Number | 103 |
| Author | Davis et al |
| Year | 2018 |
| Title | Factors affecting decisions to extend access to primary care: results of a qualitative evaluation of general practitioners' views |
| Journal | BMJ Open |
| Region (England, Scotland, Wales, and Northern Ireland) | England |
| Study methodology | Interviews with GPs from practices participating in the Sheffield Enhanced Primary Care programme (EPCP), which was part of the second wave of the Prime Minister's Challenge Fund for England. Responses were analysed using a framework analysis approach. |
| Type of document | Primary research |
| Topic or focus of paper | Enhanced Primary |
| Research objectives and focus | Report general practitioners' (GPs') views and experiences of an Enhanced Primary Care programme (EPCP) funded as part of the Prime Minister's Challenge Fund (second wave) for England which aimed to extend patient access to primary care. |
| Theoretical framework/theories/ underpinnings | |
| Description of intervention | |
| Rationale for paper/review | The first wave of the Prime Minister's Challenge Fund for England launched in 2013, and was a £50 million investment to improve general practice by increasing access for patients. This emphasised the need to improve patient access to GPs—specifically increasing OOHs options. Although evaluation of the scheme demonstrated a reduction in ED attendances, GPs' perspectives were limited to a survey of collective staff satisfaction and GPs' perspectives in relation to the national policy implementation of the EPCP have not been explored. |
| What creates meaning that sustains GPs engagement with, and involvement in, work? | |
| What solutions/recommendations are proposed? | The study makes the case for enhanced planning and organisational development with GPs as stakeholders within individual practices and groups. This would ensure that policy implementation is effective and sustained at local level. A failure to localise implementation may be associated with increased workloading in primary care without the sustained benefits to patients and the public. |
| From which stakeholders' perspective? (General practitioners, patients, other HCPs) | GPs |
| How is work-related joy constructed? (Diversity of | |

| | |
|---|---|
| ways individuals may think about joy) | |
| What prevents/removes work-related joy? | Most GPs regarded the centralisation of the programme as an imposition; upsetting the balance between their public health responsibilities and their patient-centred ethos. Their viewpoints reflected the priorities of their own practice as the primary organisational unit and the specific patient population associated with that practice. |
| Organisational characteristics (small, large) | Some GPs appeared to view the EPCP programme as a short-term centrally driven initiative with little to offer in terms of the core aims of primary care, while others saw it as a pragmatic solution to manage additional demand.<br><br>Overall, GPs were sceptical about the need to offer a 7-day service in general practices. Local (practice-based) capacity to manage patient access was a key factor influencing their views on the investment in OOH appointments: "We manage our services very well (at the practice) ... we don't have access problems and if we don't have a big problem we are reluctant to change practice"<br><br>GPs expressed frustration that the EPCP was not a good use of resource and several suggested it was not the best way to address the needs of their practice population. They proposed that funds could have been better used by devolving them to individual practices. |
| Managing ill-defined illnesses and chronic complex multimorbidity | |
| Human connection with patients | |
| Human connection with peers | There was general enthusiasm for the incorporation of pharmacy specialists in general practice and acknowledgement of the contribution of these additional professional knowledge and skills: "And she's been part of the team, the enormous paper chases ... So loading people's medications onto the computer, all their repeats and then reviewing them. So for our circumstances it's been timely". However, they identified the variation in levels of skills and the need to invest in new practitioners coming into the service. "Pharmacists within other people's practices seem to be doing loads of stuff, you know, adding drugs that have been discharge summaries from hospital, doing medication reviews, that type of thing. We've got quite a junior Pharmacist so I'm not sure at the moment that it massively reduces our workload but I could see how it could." |
| Human connection across other organisations (e.g., secondary care) | |
| Learning systems (e.g., competence, capability, mastery, socially situated) | |
| Additional useful information | Financial imperatives were seen as key drivers for involvement in the programme. GPs saw the opportunity to use incentive payments within the EPCP as additional funding for their practice. The business imperative to cover the cost of their surgeries and balance the books was an important factor in their decision to engage with the programme. |

| | |
|---|---|
| | Some GPs questioned quality of access to the EPCP. Some were doubtful that patients with English as a second language and/or with a lack of personal transport were equally served by the schemes to enhance OOH primary care. Population knowledge was a determining factor for the GP's involvement in using centrally delivered schemes and many elected not to use the new capacity on offer. |
| Summary of findings | This paper mainly focusses on evaluating the success of the EPCP. The most important finding relevant to our review is the importance of co-designing policy interventions with GPs in order to maximise efficiency and sustainability. Similar to other studies, this paper emphasises how ‘new’ roles can be incredibly useful additions to the practice team, however require training and supervision from other practice staff in some cases. |
| Strengths, limitations, and future research directions | The GP perspective on transformational interventions which this paper addresses a significant gap in knowledge. The overall sampling approach ensured geographical spread and coverage of all key characteristics at practice level. We did not purposively sample for practice size and demographic complexity which may have extended the range of viewpoints and experiences. At practice level, those who volunteered to participate were effectively self-selected.<br><br>More research is needed to identify how to improve access without generating unnecessary additional demand or compromising continuity of care. |

| | |
|---|---|
| Paper Number | 104 |
| Author | Freund et al |
| Year | 2014 |
| Title | Skill mix, roles and remuneration in the primary care workforce: Who are the healthcare professionals in the primary care teams across the world? |
| Journal | International Journal of Nursing Studies |
| Region (England, Scotland, Wales, and Northern Ireland) | N/A |
| Study methodology | Skill mix in the primary care workforce of six countries (US, Canada, Australia, England, Germany and the Netherlands) is discussed by a team of national experts (the authors). Characterization of the workforce and issues for each country was informed by scientific publications, policy reports of local authorities (including websites) and supplied by personal communication if further information was needed. |
| Type of document | |
| Topic or focus of paper | ‘New’ roles in general practice |
| Research objectives and focus | Provide an overview of education, tasks and remuneration of nurses and other primary care team members in six OECD countries. |

| | |
|---|---|
| Theoretical framework/theories/ underpinnings | |
| Description of intervention | |
| Rationale for paper/review | World-wide, shortages of primary care physicians and an increased demand for services have provided the impetus for delivering team-based primary care. The diversity of the primary care workforce is increasing to include a wider range of health professionals such as nurse practitioners, registered nurses and other clinical staff members. |
| What creates meaning that sustains GPs engagement with, and involvement in, work? | |
| What solutions/recommendations are proposed? | Important facilitators for expanding the multidisciplinary team in general practice include:<br>1. Clear definitions of each team-member's role may which allows optimally shared responsibility for patient care within primary care teams<br>2. Sufficient training is provided<br>3. Information being shared timely and comprehensively among all team members<br>4. Reimbursement schemes accounting for services delivered by non-physician health professionals. |
| From which stakeholders' perspective? (General practitioners, patients, other HCPs) | 'National experts' (academics) |
| How is work-related joy constructed? (Diversity of ways individuals may think about joy) | |
| What prevents/removes work-related joy? | |
| Organisational characteristics (small, large) | |
| Managing ill-defined illnesses and chronic complex multimorbidity | |
| Human connection with patients | |

| | |
|---|---|
| Human connection with peers | In Western countries, as numbers of primary care physicians decrease and patient demand increases, innovations are required to reduce the workload of primary care physicians. Task shifting from physicians to non-physician health professionals is a key element of these innovations. However, task shifting requires both willingness to give up tasks on physicians' side as well as the ability and capacity to perform these tasks on nonphysician health professionals' side. The former may by hampered established professional roles in developed countries. Non-physician health professionals' ability to perform specific tasks is dependent on education and training.<br><br>Task shifting from doctors to non-physician health professionals has raised two major concerns: patient safety/quality of care and decreasing continuity of care. To date evidence supporting each of these concerns is lacking. In contrast, a number of studies have shown that quality of care delivered by non-physician health professionals like nurse practitioner is not inferior if compared to physicians. Introducing protocol-based care (e.g., standing orders for medication refill) may further help to facilitate task shifting as it may face problems deriving from limited training and lack of legal accountability. Adding members to care teams obviously increases the risk of decreasing continuity of care if defined as 'seeing he-same-health-care-provider-every-time'. However, electronic health records shared across all team members may at least in part overcome the potential harms of increasing numbers of health care providers per case (Green et al., 2013). Finally, the notion of delegating tasks 'downward' to non-physician health professionals has increasingly been replaced by efforts to form 'care teams' in all countries under study. Although further attempts to promote team constitution are greatly needed in all of the countries, awareness and appreciation of each team 'players' role may be the first step and facilitated by this overview.<br><br>Current regulations and reimbursement schemes may also hinder role expansion of non-physician health professionals and paraprofessionals in many countries. Particularly, fee-for-service schemes may hamper role expansion of non-physician health professionals if only services delivered by physicians are reimbursed.<br><br>It has been shown that nurse practitioners and physician assistants are able to deliver at least 60% of office-based primary care (Ginsburg et al., 2009).<br><br>The role of extended practice nurses and practice nurse auxiliary staff is defined unclearly in most countries – as do levels of professional training required. |
| Human connection across other organisations (e.g., secondary care) | |
| Learning systems (e.g., competence, capability, mastery, socially situated) | |
| Additional useful information | |
| Summary of findings | Much of this paper is devoted to describing primary care skill-mix in non-UK countries, therefore is not relevant to our review. Some of the overall, international conclusions are relevant to our review, including how highly established professional roles can be a barrier to diversifying teams; the importance of training, clarity over roles, information sharing and appropriate remuneration schemes; how there is no |

| | |
|---|---|
| | evidence supporting the claims that 'new' roles decrease quality and continuity of care (and, indeed, how these problems can be mitigated); and the importance of forming care 'teams' as opposed to viewing changes as simple downward delegation. |
| Strengths, limitations, and future research directions | |

| | |
|---|---|
| Paper Number | 105 |
| Author | Katherine Garratt |
| Year | 2023 |
| Title | The NHS workforce in England |
| Journal | House of Commons Library: Research Briefing |
| Region (England, Scotland, Wales, and Northern Ireland) | England |
| Study methodology | [Does not say – likely some form of literature review and synthesis] |
| Type of document | Grey literature (secondary research) |
| Topic or focus of paper | NHS England workforce |
| Research objectives and focus | Discuss the NHS workforce in England, focusing on the clinical professions, including doctors and nurses |
| Theoretical framework/theories/ underpinnings | |
| Description of intervention | |
| Rationale for paper/review | [Does not specifically state – in general, these briefings are published to support the work of MPs. Of note, this briefing was released following the publication of the NHS Long Term Workforce Plan] |
| What creates meaning that sustains GPs engagement with, and involvement in, work? | The National GP retention scheme, a package of educational and financial support, aims to support GPs at risk of leaving the profession because they require greater flexibility, including for personal reasons such as caring responsibilities or illness, or are approaching retirement. The Primary care flexible staff pools initiative also aims to help GPs work more flexibly across primary care networks (PCNs). The RCGP has called for the evaluation and expansion of retention initiatives and for more flexibility and control for all GPs, not just those involved in retention schemes |

| | |
|---|---|
| What solutions/recommendations are proposed? | |
| From which stakeholders' perspective? (General practitioners, patients, other HCPs) | N/A |
| How is work-related joy constructed? (Diversity of ways individuals may think about joy) | The House of Commons Health and Social Care Committee noted that many GPs turn to locum work to gain more control over their work-life balance and said NHS England should focus on encouraging this group back into regular employment by offering more flexible working patterns. |
| What prevents/removes work-related joy? | A survey of GP trainees in 2022 found less than a third of respondents said they are planning to work full-time a year after qualifying, with most saying this was due to the "intensity of the working day.<br><br>Survey results in a report by the Royal College of General Practitioners (September 2022) found "23% of GPs across the UK were so stressed that they felt they couldn't cope most days or every day".<br><br>A survey by the Royal College of General Practitioners (RCGP) noted that, despite a trend in GPs reducing their number of contracted hours in clinical practice, many are working more hours than they are contracted for – 84% of UK members said they worked over their contracted hours on most days. The RCGP say "this suggests that efforts to boost the workforce by driving up participation rates alone are unlikely to be effective or sustainable for most GPs."<br><br>The General Medical Council have reported differences across types of doctors, with GPs almost twice as likely to cite burnout (42.8%) compared to specialists (22.2%). |
| Organisational characteristics (small, large) | There has also been concern that in General Practice there has been a shift away from partnership roles towards salaried roles, which is having a "negative impact on the full time equivalent (FTE) GP workforce." To attract more GPs into partnerships, the New to Partnership Payment Scheme gives a GP becoming a partner for the first time a "golden handshake" of up to a £20,000, plus a contribution to on-costs and training. |
| Managing ill-defined illnesses and chronic complex multimorbidity | |
| Human connection with patients | |
| Human connection with peers | The NHS Long Term Workforce Plan promises to allow doctors other than GPs to work in primary care.<br><br>The NHS Long Term Workforce Plan promises to extend the Additional Roles Reimbursement Scheme (ARRS), increasing non-GP direct patient care staff numbers by around 15,000 by 2036/37. |

| | |
|---|---|
| | There has been some criticism of the ARRS. The King's Fund reported there is a lack of understanding about the purpose or potential contribution of the scheme.158 The online general practice magazine Pulse reported that GP leaders have warned extra staff are not enough on their own to reduce the pressure on GPs' own workload – and in some cases add to it in the short-term. Furthermore, A survey by the Royal College of General Practitioners heard that the ARRS and the Investment and Impact Fund need reform as they are "too inflexible" and "have too many requirements attached". |
| Human connection across other organisations (e.g., secondary care) | The delivery plan for recovering access to primary care published in May 2023 by the DHSC and NHS England promises to cut bureaucracy by improving the interface between GPs and NHS trusts, reduce the amount of medical evidence required from GPs and increase the use of self-certification |
| Learning systems (e.g., competence, capability, mastery, socially situated) | |
| Additional useful information | Female GPs work fewer hours on average than male GPs.<br><br>The NHS Long Term Workforce Plan promises to increase GP speciality training places to 5,000 per year by 2027/28 and 6,000 by 2031/32. 500 new places will be available from September 2025.<br><br>In 2020/21, 47% of new GP trainees were international medical graduates. |
| Summary of findings | This paper discusses the NHS workforce in England, including demographics, policy, recruitment, retention, wellbeing and pay. Of particular relevance to our review, it describes a number of recent national policy documents (released since 2019) relating to the GP workforce, including the NHS Long Term Workforce plan, the National GP Retention Scheme, the New to Partnership Payment scheme, and the delivery plan for recovering access to primary care. |
| Strengths, limitations, and future research directions | |

| | |
|---|---|
| Paper Number | 106 – as most of this document is potentially relevant to our review, I have extracted an overview of the key policies. |
| Author | NHS England |
| Year | 2016 |
| Title | General Practice Forward View |
| Journal | N/A (policy document) |
| Region (England, Scotland, Wales, and Northern Ireland) | England |
| Study methodology | [Does not say] |

| | |
|---|---|
| Type of document | Grey literature – policy document |
| Topic or focus of paper | General practice investment and reform |
| Research objectives and focus | Set out a program of national investment and reform to help address the pressures in general practice. |
| Theoretical framework/theories/ underpinnings | |
| Description of intervention | |
| Rationale for paper/review | The "real pressures" facing British general practice, caused by "a rise in patient expectations, a target driven culture and a growing requirement for GPs to accommodate work previously undertaken in hospitals, or in social care". |
| What creates meaning that sustains GPs engagement with, and involvement in, work? | |
| What solutions/recommendations are proposed? | This policy document, published in 2016, set out NHSE's funded plans to respond to the pressures facing general practice across the areas of investment, workforce, workload, infrastructure and care redesign. It constituted the most 'far-reaching' support offered to general practice in over a decade. An overview of the measures is as follows.<br>1. Investing a further £2.4 billion a year by 2020/21 into supporting and growing general practice services. This represents a 14 percent real terms increase.<br>2. Supplementing this with a one-off Sustainability and Transformation package of non-recurrent investments, totalling over half a billion pounds<br><br>This money will pay for the following reforms:<br>1. £40 million for a new practice resilience programme starting in 2016/17, and an extra £16 million to provide services for doctors suffering from burn out<br>2. £206 million for workforce measures to grow the medical and non-medical workforce (including major national and international recruitment campaigns to double the growth rate of doctors working in general practice, a new offer to every practice in the country to access a clinical pharmacist leading to an extra 1,500 pharmacists in general practice, support for every practice to help their reception and clerical staff play a greater role in signposting patients and handling paperwork to free up GP time, investment in practice nurse development and return to work schemes, investment in practice manager development, piloting medical assistant roles, training and investment for 1,000 new physician associates, and 3,000 new mental health workers) |

| | |
|---|---|
| | 3. £246 million to support practices in redesigning services, including a requirement on CCGs to provide around £171 million of Practice Transformational Support and a new national £30 million Releasing Time for Patients development programme for general practice, to help practice release capacity and work together at scale, enable self-care, introduce new technologies, and make best use of the wider workforce, so freeing up GP time and improving access to services.<br>4. Supporting the increased use of technology backed by both increases in recurrent funding for GP IT, and investment to support the take up of online consultation systems in every practice<br>5. Adopting an intelligent approach to introducing extended access through flexibilities in delivery of the Government's access commitment, enabling integration with out of hours provision, the ability for extended access to boost overall capacity and reduce demand in normal working hours, and an understanding that no GP will be forced to open seven days or work seven days<br>6. Supporting new models of care in vanguard sites, to spread innovative solutions, and the development of a voluntary MCP contract for larger GP groups and community health services<br>7. Improving the interface between hospitals and general practice, beginning with changes to the NHS Standard Contract from April 2016<br>8. Continuing to make capital investments, with the estimated likely capital investment over the next five years to reach over £900 million<br>9. Bringing forward proposals to tackle indemnity costs<br>10. Reducing the frequency of CQC inspection for good and outstanding general practices, whilst continuing to protect patients and drive-up quality |
| From which stakeholders' perspective? (General practitioners, patients, other HCPs) | National policymakers at NHSE, alongside the RCGP. |
| How is work-related joy constructed? (Diversity of ways individuals may think about joy) | |
| What prevents/removes work-related joy? | |

| | |
|---|---|
| Organisational characteristics (small, large) | |
| Managing ill-defined illnesses and chronic complex multimorbidity | |
| Human connection with patients | |
| Human connection with peers | |
| Human connection across other organisations (e.g., secondary care) | |
| Learning systems (e.g., competence, capability, mastery, socially situated) | |
| Additional useful information | |
| Summary of findings | This national policy document published by NHSE set out a substantial increase in national funding allocated to general practice, alongside a range of reforms (see overview above). |
| Strengths, limitations, and future research directions | |

| | |
|---|---|
| Paper Number | 107 |
| Author | Gibson et al |
| Year | 2022 |
| Title | Primary care workforce composition and population, professional, and system outcomes: a retrospective cross-sectional analysis |
| Journal | BJGP |
| Region (England, Scotland, Wales, and Northern Ireland) | England |
| Study methodology | A multivariable regression analysis was undertaken, relating numbers of staff in four groups — GPs, nurses, healthcare professionals, and health associate professionals — to patient access and satisfaction, quality of clinical care and prescribing, use of hospital services, GP |

| | |
|---|---|
| | working conditions (subsample of practices), and costs to the NHS. Data were obtained from the GP Patient Survey 2019, Quality and Outcomes Framework, prescribing data, the Hospital Episode Statistics database, the NHS Payments to General Practice 2019/2020, and the Tenth NationalGPWorklifeSurvey2019. |
| Type of document | Primary research |
| Topic or focus of paper | Diversification of the primary care workforce |
| Research objectives and focus | Estimate associations between workforce composition and outcomes. |
| Theoretical framework/theories/ underpinnings | |
| Description of intervention | |
| Rationale for paper/review | The diversification of types of staff delivering primary care may affect professional, population, and system outcomes. |
| What creates meaning that sustains GPs engagement with, and involvement in, work? | |
| What solutions/recommendations are proposed? | Current health policy in England continues to drive changes in workforce composition with the employment of ever-increasing numbers of workers through the Additional Roles Reimbursement Scheme (ARRS). However, the present analysis indicates that deployment of newer types of practitioners cannot be assumed to provide a straightforward, complete, or cost-neutral solution to the GP workforce and workload crisis. |
| From which stakeholders' perspective? (General practitioners, patients, other HCPs) | Patients and GPs. |
| How is work-related joy constructed? (Diversity of ways individuals may think about joy) | |
| What prevents/removes work-related joy? | |
| Organisational characteristics (small, large) | |
| Managing ill-defined illnesses and chronic complex multimorbidity | |

| | |
|---|---|
| Human connection with patients | Having additional GPs (for the average practice) was associated with higher patient satisfaction but a greater number of staff in each of the other three groups was associated with lower patient satisfaction. |
| Human connection with peers | Additional GPs were associated with lower proportions of work that GPs think can be delegated, and higher GP job satisfaction. Additional nurses had the opposite effects on practice functioning and working conditions. Healthcare professionals and health associate professionals had similar effects as nurses on practice functioning and working conditions; however, healthcare professionals had a much larger effect on perceived opportunities for work delegation, and health associate professionals were associated with lower GP job satisfaction. |
| Human connection across other organisations (e.g., secondary care) | |
| Learning systems (e.g., competence, capability, mastery, socially situated) | |
| Additional useful information | |
| Summary of findings | The multivariable regression analysis undertaken by this study relates the number of staff in a given practice to a wide range of variables. Relevant to our study, it found that more GPs and less ‘other’ staff was associated with a greater patient satisfaction. Furthermore, more GPs and less ‘other’ staff were also associated with a greater GP job satisfaction and a lower proportion of work that GPs think can be delegated. |
| Strengths, limitations, and future research directions | May be prone to bias from unmeasured confounding factors. |

| | |
|---|---|
| Paper Number | 108 |
| Author | Gibson et al |
| Year | 2023 |
| Title | General practice managers’ motivations for skill mix change in primary care: Results from a cross-sectional survey in England |
| Journal | Journal of Health Services Research & Policy |
| Region (England, Scotland, Wales, and Northern Ireland) | England |
| Study methodology | Cross-sectional survey of GP practice managers in England focussing on 6 non-GP roles: advanced nurse practitioner, specialist nurse, health care assistant, physician associate, paramedic and pharmacist |
| Type of document | Primary research |
| Topic or focus of paper | ‘New’ roles in general practice |

| | |
|---|---|
| Research objectives and focus | Determine the factors that motivated GP practice managers in England to employ nonmedical roles, and to identify an ideal hypothetical GP practice workforce. |
| Theoretical framework/theories/ underpinnings | |
| Description of intervention | |
| Rationale for paper/review | In England, as a result of workforce shortages and the increasing frailty and complexity of patients, GPs have seen substantial increases in their workload. This has implications for patients needing timely access to primary and secondary care, and exacerbates retention of GPs and wider workforce challenges. Skill mix change has been proposed as a way to address these challenges, that is, expanding the primary care workforce by employing a diverse range of practitioners to free up GP time for more complex patients, offer additional health care services or act as partial or total substitutes for GPs. However, any reconfiguration of the primary care workforce will require active management to ensure it does not undermine continuity of care or lead to reduced productivity. |
| What creates meaning that sustains GPs engagement with, and involvement in, work? | |
| What solutions/recommendations are proposed? | |
| From which stakeholders' perspective? (General practitioners, patients, other HCPs) | GP practice managers |
| How is work-related joy constructed? (Diversity of ways individuals may think about joy) | |
| What prevents/removes work-related joy? | |
| Organisational characteristics (small, large) | |
| Managing ill-defined illnesses and chronic complex multimorbidity | |
| Human connection with patients | |

| | |
|---|---|
| Human connection with peers | 'Desire to release GP time' and 'improve working conditions' were common reasons given for employing 'new' roles. |
| Human connection across other organisations (e.g., secondary care) | |
| Learning systems (e.g., competence, capability, mastery, socially situated) | |
| Additional useful information | There is some evidence that adding nursing staff does not always release doctor time and instead can create a duplication of appointments or workload (10-12). Additional funding to encourage new ways of working can incentivise skill mix change, but there is a risk that if additional funding for new roles is not maintained, then new roles may cease with the funding. |
| Summary of findings | The most commonly selected 'motivating factor' for employing advanced nurse practitioners and physician associates was 'to increase overall appointment availability'; for employing pharmacists and paramedics it was 'desire to release GP time' and for employing specialist nurses and health care assistants it was 'desire to achieve a better match between what patients need and what the practitioner team can deliver'. Supply factors such as cope with recruitment issues – our choices are limited by the availability of suitable practitioners' and 'unable to recruit a GP' were most commonly selected by practices employing advanced nurse practitioners, paramedics and physician associates. Other commonly selected motivating factors across the staff groups were desire to 'improve cost-effectiveness', 'move forward with national policy for skill mix (i.e. different types of practitioners)' and 'to provide additional or improved services to patients such as increased access beyond what is currently available'. |
| Strengths, limitations, and future research directions | This study confirms that more diverse teams of practitioners are playing an increasing role in providing primary care in England. Managers prefer not to employ all new roles directly within the practice. A more detailed investigation of future workforce requirements is necessary to ensure that health policy supports the funding (whether practice or population based), recruitment, training, deployment and workloads associated with the mix of roles needed in an effective primary care workforce. |

| | |
|---|---|
| Paper Number | 109 |
| Author | Lawson & Kumar |
| Year | 2020 |
| Title | The Wass report: moving forward 3 years on |
| Journal | BJGP |
| Region (England, Scotland, Wales, and Northern Ireland) | UK |
| Study methodology | Editorial |

| | |
|---|---|
| Type of document | Secondary research |
| Topic or focus of paper | GP recruitment |
| Research objectives and focus | Highlight 5 key areas from the Wass report, which published a series of recommendations giving medical schools guidance and strategies to enable recruitment of their graduates into general practice. |
| Theoretical framework/theories/ underpinnings | |
| Description of intervention | |
| Rationale for paper/review | The Wass report made a number of important recommendations that offer a sound foundation for medical schools to develop and build on; however, there is an increasing risk that these recommendations are becoming accepted as static articles of faith with little academic discussion to develop them further. |
| What creates meaning that sustains GPs engagement with, and involvement in, work? | |
| What solutions/recommendations are proposed? | Robust, determined, and forceful advocacy is needed from medical schools and politicians to address this scandalous funding gap between primary and secondary care medical education placements.<br><br>Investment in academic general practice that is accessible at all career stages and dovetails with frontline clinical demands.<br><br>Medical students and young doctors are committed to the intellectual demands of generalism and we need systems that support those aspirations and passions, rather than selling general practice as simply offering an easy life. |
| From which stakeholders' perspective? (General practitioners, patients, other HCPs) | Two GPs |
| How is work-related joy constructed? (Diversity of ways individuals may think about joy) | It is true that general practice affords unique opportunities that are more difficult to find in other specialties; these include teaching, research, focusing on population health, varied portfolio and flexible working, and a scholarly and rigorous approach to clinical care that should be celebrated. |
| What prevents/removes work-related joy? | A detailed costing exercise, just published in the BJGP, has shown that GP remuneration for medical student placements is just over half (54%) of the real cost. The current payment to primary care stands in stark contrast to hospitals where, after the agreement of a secondary care national tariff in 2013, payments can be nearly double that currently offered to GPs.<br><br>General practice (and psychiatry) are repeatedly subject to professional denigration more than other medical specialties. |

| | |
|---|---|
| Organisational characteristics (small, large) | |
| Managing ill-defined illnesses and chronic complex multimorbidity | |
| Human connection with patients | |
| Human connection with peers | |
| Human connection across other organisations (e.g., secondary care) | |
| Learning systems (e.g., competence, capability, mastery, socially situated) | Selling general practice as the ‘lifestyle’ option can, when presented out of context, undervalue the specialty and mis-sell what the working life of a GP entails. |
| Additional useful information | 39% of GPs indicate a high likelihood of leaving direct patient care in the next 5 years, with this rising to 62% in GPs aged >50 years. This is on a wider background of retention challenges in the profession generally, with only 47% of newly qualified doctors exiting the Foundation Programme entering any kind of speciality training |
| Summary of findings | This editorial is mainly focussed on recruitment into general practice. However, points relevant to our review include the lack of funding for medical student placements in general practice, which may contribute to the pressures experienced by GPs and prevent GPs from enjoying the human connection associated with teaching. Furthermore, selling GP as a ‘lifestyle’ specialty can risk devaluating the contributions made by GPs . Unique opportunities within general practice which create joy include teaching, research, focusing on population health, varied portfolio and flexible working, and a scholarly and rigorous approach to clinical care. |
| Strengths, limitations, and future research directions | |

| | |
|---|---|
| Paper Number | 110 |
| Author | Gough & Heffer |
| Year | 2016 |
| Title | When two heads are better than one |
| Journal | BMJ |

| | |
|---|---|
| Region (England, Scotland, Wales, and Northern Ireland) | England |
| Study methodology | Commentary |
| Type of document | Primary research |
| Topic or focus of paper | Job sharing |
| Research objectives and focus | Describe and evaluate a job-sharing arrangement between two senior doctors aged 60 |
| Theoretical framework/theories/ underpinnings | |
| Description of intervention | |
| Rationale for paper/review | Considering different models of job share may help the NHS to retain skilled and experienced older staff at a time when there is particular stress on the medical workforce. |
| What creates meaning that sustains GPs engagement with, and involvement in, work? | |
| What solutions/recommendations are proposed? | "We see no reason why other primary care partnerships could not consider this model." |
| From which stakeholders' perspective? (General practitioners, patients, other HCPs) | Two senior GPs. |
| How is work-related joy constructed? (Diversity of ways individuals may think about joy) | A year after starting this job share we believe that it's worked well for everyone. We've both regained a spring in our step professionally as we have time off to recuperate from the workload but not so long that we disengage.<br><br>The authors note that another pair of doctors is due to follow their example, which is an endorsement of the success of the arrangement. They also note that "many of our contemporaries have retired, but some have said they wished that they'd done something similar." |
| What prevents/removes work-related joy? | On the downside we've noted that there's a lot of administration in our working month because one doctor essentially runs two lists. |
| Organisational characteristics (small, large) | The essential prerequisites for job sharing are an enjoyment of general practice, supportive partners in the practice and at home, and trust in each other's clinical ability and values. We are fortunate to tick all those boxes—if not we'd advise taking things no further.<br><br>The doctors work alternate months – because no holiday cover is needed, the practice has the equivalent of 1.2 full time doctors. |

| | |
|---|---|
| Managing ill-defined illnesses and chronic complex multimorbidity | |
| Human connection with patients | We wrote a positive, upbeat letter to patients, emphasising that the two senior doctors in the practice were choosing to carry on working rather than retiring. We informed the patient participation group, wrote a notice on the website and in the practice newsletter, and spoke to patients in consultations. Patients have accepted the arrangement. |
| Human connection with peers | |
| Human connection across other organisations (e.g., secondary care) | |
| Learning systems (e.g., competence, capability, mastery, socially situated) | |
| Additional useful information | |
| Summary of findings | Job sharing may be a valuable method of retaining senior GPs. Prerequisites include an enjoyment of general practice, supportive peers and family, and trust between job sharing partners. The GPs have noticed increased enjoyment of their job alongside time to recuperate from the intense workload. Downsides include the increase in administrative work as they are essentially running two lists. |
| Strengths, limitations, and future research directions | |

| | |
|---|---|
| Paper Number | 111 |
| Author | Gray & Sanders |
| Year | 2020 |
| Title | A reflection on the impact of COVID-19 on primary care in the United Kingdom |
| Journal | Journal of Interprofessional Care |
| Region (England, Scotland, Wales, and Northern Ireland) | UK |
| Study methodology | Commentary |
| Type of document | Primary research |

| | |
|---|---|
| Topic or focus of paper | COVID-19 |
| Research objectives and focus | Describe and analyse the personal experience of one of the authors, who returned to work as a GP after a period of ill health due to burn out and then suspected COVID-19. |
| Theoretical framework/theories/ underpinnings | |
| Description of intervention | |
| Rationale for paper/review | The COVID-19 pandemic has presented both challenges and opportunities for those working in health and social care in the United Kingdom (UK). With much focus on secondary and acute care at this time, there has been less communication and understanding about the impact on primary care. |
| What creates meaning that sustains GPs engagement with, and involvement in, work? | The recent increase in inspection, regulation, and incentives for outcomes such as the Quality and Outcomes Framework (QOF) has had a number of consequences, both intended and unintended. Improvements in accountability and quality measures selected have undoubtedly been made as exemplified by the identification and monitoring of chronic conditions such as hypertension, but there have been questions raised around the longer-term benefits of this approach with unintended consequences including overtreatment (Sheppard et al., 2018). Important elements of care, valued by professionals and patients alike, can be eroded by these approaches, such as continuity and person-centered care (Derksen et al., 2020). |
| What solutions/recommendations are proposed? | Although the range of health-care professionals in general practice is increasing, there is a need to support and train this workforce to work alongside doctors as exemplified by the development of Advanced Clinical Practitioners in the UK. Addressing this need requires interprofessional education in establishing shared frameworks and models for how patients relate to experts. It is crucial that GPs familiar with the principles of IPE share with and support the workforce taking on these responsibilities. The education and support of 'new' roles through regular daily team meetings has been important for addressing important issues and challenges relating to patients, team members, and patient safety. As noted by others, such meetings can be an ideal opportunity to utilize the principles of IPE in a short period of time.<br><br>Addressing the anxiety and fear caused by COVID-19 by facilitating resilience is of utmost importance. This can be addressed through collaborative team working by building on and developing previous work provided to support team members.<br><br>The development of integrated care pathways requires interprofessional relationships with capabilities such as respect for difference, communication with other professionals, and the ability to negotiate collaborative person-centered care. The inclusion of IPE, particularly in the preparation and support of professionals involved in the integrated care of vulnerable groups, can help address the interprofessional challenges that integrated care engenders.<br><br>The COVID-19 pandemic offers an opportunity to reflect on the recent increase in inspection, regulation, and incentives. |
| From which stakeholders' perspective? (General practitioners, patients, other HCPs) | Two GPs. |
| How is work-related joy constructed? (Diversity of | |

| | |
|---|---|
| ways individuals may think about joy) | |
| What prevents/removes work-related joy? | Information changed very rapidly in the context of COVID-19 – meaning that even short periods off-work were challenging to catch up on.<br><br>For primary health-care teams working in areas of social deprivation, in spite of the extra workload due to health and social problems resulting from austerity, minimal additional resources have been made available. This has exacerbated the long-standing problem of the "inverse care law" (Tudor Hart, 1971) that there is an inverse relationship between the need for care, and its accessibility and utilization.<br><br>The reduced number of GPs along with the extra workload required in these areas of social deprivation present a particular challenge to all those involved in reducing health inequities and the health divide. |
| Organisational characteristics (small, large) | Remote contact has become 'the norm' during COVID-19.<br><br>Values-based practice theory as described by Fulford et al. (2012) addresses ways of working with complex and conflicting values in healthcare, and can explain how advances and rapid changes in technology can result in a diversity of options for both healthcare professionals and patients. These new options are closely associated with values and evidence. To be acceptable to all concerned, an effective and balanced approach should be adopted for new technologies that address concerns by acknowledging and working with patients' values.<br><br>When patients are given the option, remote consultations can improve accessibility, reduce consultation length, and tend to be associated with high satisfaction amongst patients and staff (Greenhalgh et al., 2020; The Kings Fund, 2018). An example of the improvement of information flow is the ability of patients to send photographs (eg of rashes) to their clinicians.<br><br>Although the option of remote consultation can be helpful to some, it can be difficult for marginalized groups in society and so, following values-based practice, inclusion must be prioritized and more traditional means of consulting must remain available when needed.<br><br>The reduction in systems administration (eg QOF tasks, CQC inspections and appraisals) during COVID 19 been welcomed: it offers the potential for post-pandemic reviews to include the introduction of seamless interprofessional systems accountability processes, to unburden practitioners. |
| Managing ill-defined illnesses and chronic complex multimorbidity | |
| Human connection with patients | Many patients who previously would have sought health advice have been reluctant to seek help from health-care professionals either because of anxiety about catching COVID-19 or concern about unnecessarily taking up valuable professional time. Whilst this can encourage self-responsibility and self-management of many minor conditions, adverse outcomes may occur if patients with significant symptoms fail to consult at an appropriate early stage. In this context, accurate remote triage assessment of patients presents a window of opportunity. |

| | |
|---|---|
| | It should be emphasized that there have been significant gaps associated with care within the community and many have become more apparent during the pandemic. Examples include safeguarding, domestic violence, homelessness, loneliness, and isolation. |
| Human connection with peers | “Advanced clinical practitioners are working with an altered workload, some of which may be outside their skill sets in an atmosphere of increased fear, stress, uncertainty, and risk”<br><br>There is much anxiety and fear amongst GP team members at the present time not only due to changes in professional practice but also related to concerns about the health of friends and colleagues, and bereavement caused by the virus. Sources of significant anxiety among health and social care staff, particularly those working in residential and nursing homes, have included inadequate personal protective equipment (PPE), the increased risk of severe complications, and increased mortality rate for Black, Asian, and Minority Ethnic groups (BAME).<br><br>Given the pace of change during COVID-19, there is a danger of information overload from multiple sources, resulting in meetings consisting of information being communicated to team members in a didactic way, which can reduce the value and opportunity for interprofessional team learning.<br><br>One of the key themes emerging from the author’s experience is the importance of professionals supporting each other (regular team meetings ‘flourished’ during the pandemic). Matheson et al. (2016) described how the development of professional resilience depends on the concept of personal traits (for example, flexibility, tolerance assertiveness, and being a team worker) linking with workplace features (for example, management support, team work, and supportive colleagues acting as workplace buffers) and social factors (for example, friends, family and leisure activities). Barr and Gray (2013) described how when doctors are faced with the stress of problems beyond their traditional role, they take one of three possible actions: 1) set aside their problems, 2) go beyond their role at risk of increasing stress and overload, and providing less than optimal care; and 3) work in partnership with other professions to spread the load and respond more fully to range of needs. Therefore, working collaboratively in teams not only improves the effective response to new emerging needs, but can reduce stress experienced by team members. This echoes advice by Anderson et al. (2019) who described how health-care professionals, following a patient death, can use team-based reflection to successfully address needs, to enhance emotional wellbeing, and to support practice learning leading to improved patient outcomes. Reflective practice or Balint groups have been found to be useful in facilitating resilience and countering the emotional toll of clinical practice and are now widely recommended by the General Medical Council in the UK (General Medical Council, 2020).<br><br>Team meetings have been essential for providing support, setting common goals, reflecting on performance, and effective team functioning. Lack of such meetings has been reported as reducing job satisfaction, increasing burnout and workload endangering patient safety (Thistlethwaite, 2012). However, up until now, there has been limited time found in some UK general practices to reflect and communicate with other professionals. One of the authors’ experience showed that finding protected time for daily interprofessional team meetings during the present crisis has been possible, essential, and worthwhile. These team meetings, that require effective facilitation and skilled leadership, can provide a forum for a number of functions especially important during the current pandemic, including good communication about planning work, informal handoffs, and debriefing (The Kings Fund, 2018), team support (Anderson et al., 2019) and addressing moral injury and distress (Greenberg et al., 2020). Remote attendance to such meetings enables the involvement of team members otherwise geographically isolated, though poses a challenge to ensure equity of involvement in team learning. |

| | |
|---|---|
| Human connection across other organisations (e.g., secondary care) | COVID19 outbreaks have rapidly increased within care homes and this has necessitated increased working and communication patterns between practice team members and care home staff. This process has included regular proactive and reactive telephone and digital contacts instead of face-to-face visits. Many care home staff describe how their role has significantly changed with easier interaction, improved approachability and communication with GPs.<br><br>Although primary healthcare is expected to play a leading role in the move towards integrated care, the authors of this paper note that there is little evidence that this is effectively occurring nationally at the present time. Hierarchical structures, perverse financial incentives, outdated expectations of roles, and inappropriate styles of leadership have all contributed as stressful barriers to the development of integrated care (Brennan, 2019, Coughlan et al., 2020). |
| Learning systems (e.g., competence, capability, mastery, socially situated) | Practitioner anxiety associated with COVID 19, alongside reduced time for communication and reflection, has the potential to result in defensive decisions and miscommunication. This amalgamation of factors can be a potent cause of error, potentially reducing patient safety. |
| Additional useful information | The Kings Fund (2018) described how 20% of work done by GPs could be effectively performed by nurse practitioners and how 12.5% of work done by nurses could be performed by health-care assistants. The Fund suggests that challenging the boundaries of traditional roles can, by expanding the skill mix, enhance patient safety, improve accessibility and quality of patient care.<br><br>During the past few years, most health and social care professions in UK primary care have experienced a recruitment and retention problem. However, during the COVID-19 pandemic, many professionals who had previously retired or left practice early have returned, highlighting that return to practice can be achieved (also underlines the importance of common purpose). |
| Summary of findings | There were a number of rapid changes to primary care during COVID-19, most notably remote assessment, reduced health seeking behaviours from patients, increased co-worker support, reduced administrative burden, and worsening pressures particularly on practices with high deprivation. The authors feel that these changes present an opportunity for primary care to make permanent changes to ways or working and become more sustainable. They group the implications for future working under four themes: integrated care, team meetings, remote consultations and communication, and supporting the workforce. These themes are all interrelated and inform one another. Organizational changes as a result of the pandemic will impact both on the development of the workforce and integrated care. These require effective interprofessional collaborative working informing holistic person-centered care whilst facilitating resilience of professionals. In order to sustain support for such organizational change, endorsement for this process is required by senior-level management, underscored by inter-institutional agreements. Regular Team meetings can provide a helpful forum for communication and support in the context of change and new ways of working such as for the use of remote consultations and communication. However, to facilitate such change in the workforce requires collaborative leadership and effective interprofessional team working with organizational and administrative support. |
| Strengths, limitations, and future research directions | |

| | |
|---|---|
| Paper Number | 112 |
| Author | Guck & Buck |
| Year | 2021 |

| | |
|---|---|
| Title | Reducing clinician inefficiency and restoring meaning in practice: A professional coaching approach for family medicine residents |
| Journal | The International Journal of Psychiatry in Medicine |
| Region (England, Scotland, Wales, and Northern Ireland) | N/A |
| Study methodology | Non-systematic review of the literature. |
| Type of document | Secondary research |
| Topic or focus of paper | Coaching |
| Research objectives and focus | Provide a summary of evidence for coaching interventions, along with evidence supporting an expansion to these approaches in clinical efficiency in outpatient settings. Outline a prospective coaching approach which targets common sources of clinic inefficiency for resident practitioners. |
| Theoretical framework/theories/ underpinnings | |
| Description of intervention | |
| Rationale for paper/review | "We believe that a viable means to address burnout [in primary care physicians] can be found in utilizing a performance coaching approach to equip resident physicians for the practical and emotional demands of the primary care work environment into which they are entering." |
| What creates meaning that sustains GPs engagement with, and involvement in, work? | Mounting high-quality evidence, including meta-analytic and systematic review research have provided support for use of professional coaching to reduce physician burnout and enhance physician well-being. Specifically, studies demonstrated that coaching approaches can reduce burnout symptoms by promoting resilience and improving stress management. Several of the coaching initiatives implemented in these studies explicitly sought to increase physicians' sense of meaning and reconnect with purpose in clinical work, in addition to focusing on areas such as work-life balance |
| What solutions/recommendations are proposed? | Key factors which are likely to reduce resident physician efficiency in clinic include lack of preparation for clinic, lack of agenda-setting with patients, misallocation of time during the medical encounter, poor coordination with members of the medical team, problematic use of technology and clinical tasks left complete. To address these, areas of clinical development within a coaching framework include: improved pre-visit planning and chart review; clinic observation and real-time feedback on agenda setting and visit structure with behavioral sciences faculty; leading pre-clinic huddles, and/or performing "mini huddles" with relevant team members; enhance proficiency with time-saving technology; shadow efficient faculty and upper-level residents; increased structure and accountability.<br><br>While these represent general strategies for improvement, individual resident physicians may also struggle with specific or unique aspects to their practice setting which may hinder efficient clinical practice (e.g. non-English speaking patients). |
| From which stakeholders' perspective? (General practitioners, patients, other HCPs) | Two 'family medicine' residents from the USA. |

| | |
|---|---|
| How is work-related joy constructed? (Diversity of ways individuals may think about joy) | |
| What prevents/removes work-related joy? | We believe the area of clinic efficiency has significant well-being implications for resident physicians. Inefficient clinicians can experience work-life balance challenges, such as work after clinic time bleeding into family time, time for rest, and hobbies. Poor clinic efficiency has also been seen to contribute to "work-work conflict", meaning that the less desirable and more overwhelming aspects of clinic (charting, etc.) take over clinical time. For clinicians who struggle with efficiency, this means that the aspects of work that are less meaningful e.g., documentation, data entry) edge out the more meaningful aspects of practice (time spent talking with patients, working with the rest of the health care team) and lead to a less joyful practice. The American Medical Association has named joy in medicine as a key component of their physician burnout. |
| Organisational characteristics (small, large) | |
| Managing ill-defined illnesses and chronic complex multimorbidity | |
| Human connection with patients | |
| Human connection with peers | |
| Human connection across other organisations (e.g., secondary care) | |
| Learning systems (e.g., competence, capability, mastery, socially situated) | |
| Additional useful information | |
| Summary of findings | High quality evidence demonstrates that coaching can reduce burnout symptoms by promoting resilience and improving stress management. A key factor that might influence joy and therefore resilience is clinical efficiency. In turn, efficiency is influenced by preparation, agenda setting, misallocation of time, poor team coordination, problematic use of technology and incomplete clinical tasks. The authors propose a theoretical coaching approach to target these areas. |
| Strengths, limitations, and future research directions | Consistent with the approaches noted in previous coaching literature, our methodology seeks to engage residents in meaningful personal change on aspects of the patient encounter over which they have the most agency and control. Nevertheless, we acknowledge the limits of this approach, and that systems-based areas such as organizational dysfunction, poor work culture, or low staffing ratios also contribute to clinic inefficiency. |

| | |
|---|---|
| | The authors concede that the basis for the approach outlined in the paper is conceptual and theoretical rather than empirical.<br><br>Future research on this prospective coaching approach will focus on systematic measurement and examination of their ability to enhance workplace satisfaction and reduce burnout among residents using the RCT methods utilized by previous coaching validation studies literature. Faculty and more senior clinicians who struggle with efficiency as a contributor of professional burnout may similarly benefit from such approaches and present another important avenue for research and development of burnout interventions. |

| | |
|---|---|
| Paper Number | 113 |
| Author | Hardman et al |
| Year | 2020 |
| Title | Subjunctive medicine: Enacting efficacy in general practice |
| Journal | Social Science & Medicine |
| Region (England, Scotland, Wales, and Northern Ireland) | England |
| Study methodology | Conducted an ethnography of a general practice surgery in England, including participant observation, interviews, and focus groups with patients, clinicians, and support staff, from February 2018 to March 2019 |
| Type of document | Primary research |
| Topic or focus of paper | Person-centred care |
| Research objectives and focus | Exploring how person-centred care is achieved regarding long-term conditions. Specifically, aimed to: identify the relevant processes and practices of clinicians; explore the views and experiences of patients and clinicians; and generate theory on how person-centred care is achieved in modern general practice. |
| Theoretical framework/theories/ underpinnings | Person-centredness, which is characterised by a focus on practice, empowerment, the coordination of care, and the centrality of developing and maintaining interpersonal relationships |
| Description of intervention | |
| Rationale for paper/review | Modern general practice is complex. Issues such as multimorbidity, polypharmacy and chronic illness management can make applying myriad single condition evidence-based guidelines increasingly difficult. This is compounded because the problems presented in general practice often require both clinical and social solutions. In response to these issues, generalist clinicians are now expected to practise |

| | |
|---|---|
| | 'person-centred care': enabling and empowering patients by combining the technical rationality of medical science with individual values, needs and preferences. |
| What creates meaning that sustains GPs engagement with, and involvement in, work? | |
| What solutions/recommendations are proposed? | By conducting consultations as much in the subjunctive as the indicative mood, clinicians in our study did not imitate their role but constantly re-made it through collaboratively enacting each consultation anew. We term this mode of clinical practice subjunctive medicine, and suggest it may be a useful way to sustainably develop the multiple, flexible habits necessary to achieve the aims of person-centred care in general practice. To summarise, we propose three principal actions that comprise subjunctive medicine:<br>1. Conceive of each consultation as collaboratively enacted anew.<br>2. Exploit the importance of the imagination in developing interpersonal relationships.<br>3. Explicitly adopt a clinical role to improve resilience. |
| From which stakeholders' perspective? (General practitioners, patients, other HCPs) | GPs, nurses, HCAs, administrative staff and the ethnographers. |
| How is work-related joy constructed? (Diversity of ways individuals may think about joy) | |
| What prevents/removes work-related joy? | |
| Organisational characteristics (small, large) | |
| Managing ill-defined illnesses and chronic complex multimorbidity | |
| Human connection with patients | Findings suggest that clinicians are confronted with considerable constraints to practicing person-centred care in general practice, which we broadly categorise as the limits of biomedicine and the structural constraints of general practice.<br><br>The limits of biomedicine, in turn, manifested in two main ways. First, through the need to consider the irreducible interconnection between a patient's health and their broader life. For example, when GPs deliver the diagnosis of fatal conditions. Second, complex problems such as chronic illness management were major challenges. Both of these issues can create tensions between patient and clinician, undermining the potential for person-centred care. |

| | |
|---|---|
| | The main structural constraint within general practice was clinician workload. The authors quoted recent retrospective analysis of 100 million general practice consultations in England, which showed a substantial increase in consultation rates, consultation duration, and total clinical workload. This increased workload informs a central tension that general practice surgeries contend with: balancing patient access with relational continuity of care. |
| Human connection with peers | One GP described the difference between present and previous general practice, inasmuch as "it used to be the GP for everything, [but] now we've got diabetic nurses and COPD nurses, and other nursing staff who are very expert and very good at doing these different things". This reflects sociological research highlighting the distributed and collective nature of modern clinical decision making, particularly in general practice, which suggests that shared decision making can be conceived of at the team as well as the individual level. |
| Human connection across other organisations (e.g., secondary care) | |
| Learning systems (e.g., competence, capability, mastery, socially situated) | GPs mitigated these constraints by forming good habits. These can be categorised in two broad categories: using expert judgement and taking patients seriously.<br><br>GPs used expert judgement in several ways. They acknowledged the uncertainty of general practice, and this focus is reflected in the findings of an ethnographic study exploring knowledge management in general practice, in which clinicians relied on collective, socially constructed knowledge in practice as much as explicit evidence from research (Gabbay and le May, 2004). For clinicians in our study, a central aspect of dealing with uncertainty was the process of managing expectations. GPs also made a notable effort to communicate not just a diagnosis and treatment plan, but the underlying concept, often using analogy: "Nowadays, your heart beats a little off. [It] jumps like a clock, which is impossible to show by taking your pulse. The ECG shows that it's normal. It's a bit like having a car misfiring". A patient having a workable conceptual understanding of disease reduces paternalism and increases self-management. Lastly, one GP suggested that the content and timing of advice is as important as its content.<br><br>GPs took patients seriously in several ways. They respected patients' intelligence, accepting that patients can accrue considerable knowledge, and giving patients the time to get across their problems in their own words. "I was wondering if it is a combination of any of the tablets?" asked the patient's wife. "You're probably right" said the GP, "for example the beta-blocker, that could make you tired.". They established trust with patients, "[Some nights] I just can't get to sleep until 4 or 5 in the morning" said the patient (P32). The GP (S13) paused. "You won't" he said, "you have to resign yourself". This was a consequence of the drugs the patient was on. "The downside is that you've got two days a week when you, say, have to get the DVDs in. You have to accept that, be pragmatic". They conducted shared decision making.<br><br>These good habits were explain to be 'not easy' and clinicians in the study did not merely will themselves towards these good habits.<br><br>Through interpreting how clinicians in our study made sense of and accounted for their own practice, we suggest they intuitively developed a meta habit of enaction, which informs and guides person-centred clinical practice. This is defined as clinicians and patients actively bringing the consultation into being together. In turn, the habit of enaction is reflected in the finding that clinicians viewed success in two ways: biomedical treatment outcome and developing a shared understanding. "I think a successful [consultation], for me, is when somebody is happy to open up and talk, and feels comfortable coming in". The explicitly creative and participatory nature of the general practice |

| | |
|---|---|
| | consultation was reflected in the views of clinicians: “I felt entirely during medical school, and ever since, that I was learning to act, that I was learning a role. Definitely. A clinical role. I often find I put on my stethoscope around my neck and it's like a priest putting on his robes or something”. This suggests an important feature of the general practice consultation: it is not, as might traditionally be assumed, only conducted in the indicative mood (things ‘as they are’). It is also conducted in the subjunctive mood (things ‘as if they are’). In the context of general practice, subjunctive conditionality means that good interpersonal understanding between patient and clinician (the consequent of a conditional) can be seriously considered even if the clinician being genuinely engaged (the antecedent) does not hold outside the consultation (the particular situation). Although we agree with proponents of person-centred care that the paternalistic model of acting on patients is misguided, our subjunctive interpretation of the consultation suggests that the person centred approach has moved too far towards transcendent sincerity towards searching for unity in a person – which may be restricting clinical opportunities. Furthermore, if we accept the proposition that consultations are collaboratively enacted, then we also don't want the patient to be as they are outside the consultation either. It is beneficial if, in some way, they play their role too. We suggest that by practising subjunctively, clinicians can create the conditions to help patients become what Epstein (2017) terms a “mindful patient”, improving potential for a good treatment outcome: “because, sitting in here, in my uniform, I'm not somebody just wandering down the street or meeting them in the shop or having a cup of coffee. It's like a closed area, and they can sit and talk to me about things they know won't go beyond the door”. In framing interpersonal relationship building as an imaginative and generative process, subjunctivity may also allow clinicians to be more resilient. |
| Additional useful information | |
| Summary of findings | GPs are confronted with considerable constraints to practicing person-centred care in general practice, which are broadly categorised as the limits of biomedicine and the structural constraints of general practice. GPs mitigate these constraints by forming good habits: using expert judgement and taking patients seriously. The authors propose that GPs did not ‘will’ themselves into forming these good habits, but developed and adapted them by intuitively adopting a second-order ‘meta’ habit of enaction – treating each consultation as collaboratively co-created anew. This suggests an important feature of the general practice consultation: it is conducted as much in the subjunctive as the indicative mood. Practicing subjunctive medicine may help clinicians sustainably and resiliently achieve the aims of patient-centred care. |
| Strengths, limitations, and future research directions | The in-depth fieldwork permitted a deep understanding of the specific general practice environment, which aided and informed theory generation. However, the general practice surgery in this study has a slightly lower than average list size, and is located in a small market town with low deprivation and a relatively socio-culturally homogenous population. Further, due to ethical concerns consultations booked on the same day were not included in the study. These issues must be considered when interpreting the findings more widely. |

| | |
|---|---|
| Paper Number | 114 |

| | |
|---|---|
| Author | Hayhoe et al |
| Year | 2018 |
| Title | Integrating a nationally scaled workforce of community health workers in primary care: a modelling study |
| Journal | Journal of the Royal Society of Medicine |
| Region (England, Scotland, Wales, and Northern Ireland) | England |
| Study methodology | Modelling exercise using publicly available data on general practice demographics, population density, household size, salary scales and screening and immunisation uptake. |
| Type of document | Secondary research |
| Topic or focus of paper | Community health workers |
| Research objectives and focus | To model cost and benefit of a national community health worker workforce. |
| Theoretical framework/theories/ underpinnings | In the 1960s, programmes in the US funded members of the community to provide a bridge between patients and healthcare providers. Facilitating appointment-keeping and increasing compliance with medications, community health workers improved access to and quality of healthcare, while reducing costs. Growing evidence now supports building primary care services with community health workers.<br><br>Community health workers in Brazil have basic training in disease identification and monitoring, immunisation, and screening support and health promotion. Their skillset includes supporting patients with medication adherence and healthcare system navigation, monitoring chronic disease and identifying new symptoms. Each is responsible for around 150 households, in a defined catchment area, which they visit at least once per month. Community health workers gain detailed knowledge of all members of these households and liaise proactively with general practitioners and practice nurses to avoid crises and complications. |
| Description of intervention | |
| Rationale for paper/review | Increasing workload, a reduced percentage of the budget and workforce retention and recruitment problems challenge the capacity of available general practitioners in the UK NHS. Community health workers have been shown in other countries to improve access to and quality of healthcare, whilst reducing costs. This study builds the case for a scaled community health worker workforce in England by estimating likely costs and key benefits of their deployment throughout NHS primary care in England, following the Brazilian Family Health Strategy model. |
| What creates meaning that sustains GPs engagement with, and involvement in, work? | |
| What solutions/recommendations are proposed? | |

| | |
|---|---|
| From which stakeholders' perspective? (General practitioners, patients, other HCPs) | [Used publicly available data – the authors are a mixture of GPs, public health doctors and academics] |
| How is work-related joy constructed? (Diversity of ways individuals may think about joy) | In addition to the possibility of improvements in patient engagement in areas such as health promotion and disease prevention, chronic disease management, cancer screening and immunisation (see additional information below for results of modelling) community health workers have the potential to liase with GPs, identify problems early and support chronic disease monitoring – which might reduce unnecessary workload burden on general practitioners, improving access while reducing use of acute and secondary care services. |
| What prevents/removes work-related joy? | |
| Organisational characteristics (small, large) | |
| Managing ill-defined illnesses and chronic complex multimorbidity | |
| Human connection with patients | |
| Human connection with peers | |
| Human connection across other organisations (e.g., secondary care) | |
| Learning systems (e.g., competence, capability, mastery, socially situated) | |
| Additional useful information | If community health workers were each responsible for 137.5 households, 165,878 would be needed to cover the population registered with general practices in England. A total of 110,585 community health workers would be required if there were 206.25 households per community health worker, and 82,939 if 275 households. Assuming a midpoint salary scale of Band 2 Point 5, we estimate annual NHS employment costs of these numbers of community health workers to be £3.32bn, £2.22bn and £1.66bn, respectively,<br><br>Taking the middle scenario, assuming 206.25 households per community health worker, each would regularly support approximately 29 patients with asthma, nine patients with chronic obstructive pulmonary disease, four patients with dementia, 34 patients with diabetes and 69 patients with hypertension.<br><br>If community health workers led to successful screening of 20% of previously missed individuals, this would equate to an additional 753,592 new cervical cancer screenings nationally, with 365,166 new breast cancer screenings and 482,924 bowel cancer screenings, during |

| | |
|---|---|
| | the relevant time periods for each programme. Successful referral of 20% of children who had missed immunisations would mean that each year a further 16,398 children would receive MMR1 at 12 months and 24,716 children would receive MMR2 at five years of age.<br><br>All patients with chronic diseases would have regular health promotion, and individuals would be proactively identified for emerging physical health, mental health or social care issues. |
| Summary of findings | Community health workers improved access to and quality of healthcare, while reducing costs. Relevant to our review, this study suggests that community health workers have the potential to liase with GPs, identify problems early and support chronic disease monitoring – which might reduce unnecessary workload burden on general practitioners. It also models the costs and benefits of a large scale implementation of community health workers in England. The conservative estimate is that they would cost the NHS £2.22bn, whilst significantly improving screening and vaccination uptake in the order of hundreds of thousands (alongside providing home-based health promotion and support to patients with chronic disease, which were not modelled). |
| Strengths, limitations, and future research directions | The Brazilian health system differs from that in the UK, and the impact of community health workers in the UK may be smaller overall, given differences in baseline health provision, health needs, health inequalities and health literacy.<br><br>As in any modelling exercise, this study is limited by assumptions such as average household size and the number of households that community health workers have responsibility for. We took measures to minimise the effect of these by using published data on general practice list size, population characteristics, population density, disease prevalence and screening and immunisation uptake, and by modelling a variety of different scenarios.<br><br>Modelling impact of community health workers on cancer screening and immunisation uptake required assumptions as to possible effect size. Wide variation exists in reported effect size of community health worker interventions, ranging in immunisation uptake from no effect to 36% relative increase in immunisations. We opted therefore to provide alternative models assuming community health workers facilitate uptake by 10%, 20% or 30% of eligible but unscreened or unimmunised individuals.<br><br>Next steps should include pilot studies to explore acceptability and feasibility of introduction of community health workers in NHS primary care following the Brazilian model, allowing a reference case health technology assessment to be carried out. |

| | |
|---|---|
| Paper Number | 115 |
| Author | Heponiemi et al |

| | |
|---|---|
| Year | 2014 |
| Title | General practitioners' psychosocial resources, distress, and sickness absence: a study comparing the UK and Finland |
| Journal | Family Practice |
| Region (England, Scotland, Wales, and Northern Ireland) | UK (with Finland as a comparison) |
| Study methodology | Two independent cross-sectional surveys conducted in general practice in the UK and Finland. Analyses of covariance were used for continuous outcome variables and logistic regression for dichotomized variable (sickness absence) adjusted for gender, qualification year and response format. |
| Type of document | Primary research |
| Topic or focus of paper | GP wellbeing |
| Research objectives and focus | To compare psychosocial resources (job control and participative safety), distress and sickness absences between GPs from the UK and those from Finland. We also examined differences in how psychosocial resources are associated with distress and sickness absence and how distress is associated with sickness absence for both countries.<br><br>Note that participative safety is defined as team participation, which can be measured by interaction frequency and information sharing. |
| Theoretical framework/theories/ underpinnings | |
| Description of intervention | |
| Rationale for paper/review | Many countries, including the UK and Finland, face difficulties in recruiting GPs and one reason for these difficulties may be due to negative psychosocial work environments. |
| What creates meaning that sustains GPs engagement with, and involvement in, work? | GPs from the UK perceived having more job control and higher participative safety levels than Finnish GPs. However, UK GPs reported statistically significantly higher levels of distress than GPs from Finland (though the absolute difference in mean scores of 0.16 of a scale point was not large).<br><br>Job control was associated with distress among Finnish GPs but not with UK GPs. Similarly, as with job control, high participative safety levels were associated with less distress in Finnish GPs but not among UK GPs. Distress was significantly associated with sickness absence spells among UK GPs but not among Finnish GPs.<br><br>The higher distress levels among UK GPs compared with Finnish GPs may reflect higher work load levels. Finnish full-time GPs work on average 42 h a week, whereas GP partners in the UK work on average 44 h per week. In addition, GPs from the UK have contractor status, financial responsibilities and smaller practices (average of 3.8 GPs), whereas Finnish GPs work for municipalities, have no financial responsibilities and larger practices (average of 11.7 GPs); this fact may also influence here. It is possible the UK's contractor system and smaller practices gives more sense of one's control opportunities and participative safety. |

| | |
|---|---|
| What solutions/recommendations are proposed? | |
| From which stakeholders' perspective? (General practitioners, patients, other HCPs) | |
| How is work-related joy constructed? (Diversity of ways individuals may think about joy) | |
| What prevents/removes work-related joy? | |
| Organisational characteristics (small, large) | |
| Managing ill-defined illnesses and chronic complex multimorbidity | |
| Human connection with patients | |
| Human connection with peers | |
| Human connection across other organisations (e.g., secondary care) | |
| Learning systems (e.g., competence, capability, mastery, socially situated) | |
| Additional useful information | |
| Summary of findings | This study found that UK GPs had higher levels of distress than Finnish GPs and this high distress was associated with sickness absences. The authors suggested that this might be due to the greater workload and contractor status unique to UK GPs. The study also found that UK GPs perceived having more job control and higher participative safety levels than Finnish GPs (although this was not linked to distress). The authors highlighted that the UK's contractor system and smaller practices may give more sense of one's control opportunities and participative safety. |
| Strengths, limitations, and future research directions | We were able to use large samples with good response rates. This study relied on self-reported measures, which may lead to problems associated with an inflation of the strengths of relationships due to common method variance. Moreover, although we controlled for gender, |

| | |
|---|---|
| | qualification year and response format, we cannot rule out the possibility of residual confounding. In addition, the Finnish and UK samples were gathered in different ways that may have influenced the results; the Finnish sample was a random sample of all physicians in Finland, whereas the UK sample was recruited from general practices registered with a national research network. Moreover, results regarding higher levels of distress among the UK GPs should be taken cautiously, given that the first answer option among the UK GPs differed from the one that was used among Finnish GPs and generally. Thus, it is possible that the UK GPs are scoring higher on distress by virtue of having the option 'much less than usual' instead of 'not at all' option.<br><br>Future studies should examine the effects of financial issues and other work-related factors, such as organizational justice, social support and work arrangements, for the well-being of UK GPs. |

| | |
|---|---|
| Paper Number | 116 |
| Author | Imtiaz-Umer et al |
| Year | 2021 |
| Title | Helping Us Grow generations of GPs (HUGG): A pilot study evaluating Longitudinal Support Networks (LSNs) on recruitment, retention and resilience of GPs in Derbyshire. |
| Journal | Education for Primary Care |
| Region (England, Scotland, Wales, and Northern Ireland) | England |
| Study methodology | Longitudinal support networks (see below) were piloted in a 6-month study. Data was collected prior to, during and at the end of the pilot period using a mixed methods approach of questionnaires with Likert scales and focus groups. The survey questions were organised into groups for the purpose of paired analyses and focused on questions relating to social connection, support, career intentions and preferred methods of communication. |
| Type of document | Primary research |
| Topic or focus of paper | Cross-generational collegiality |
| Research objectives and focus | Longitudinal support networks (LSNs) were developed and piloted in Derbyshire to promote cross-generational collegiality and provide opportunities for networking and support across the working life of a GP. Linking different GP career stages together aimed to overcome hierarchical boundaries enabling cross-generational conversation between GPs from medical school to retirement. |
| Theoretical framework/theories/ underpinnings | |

| | |
|---|---|
| Description of intervention | 39 participants interested in GP from medical school, through foundation, GP training and all stages of the GP career were joined together to create longitudinal support networks (LSNs). During the 6 month pilot study, LSNs met face to face once and then virtually to create an informal discussion network. |
| Rationale for paper/review | Previous work by the RCP has shown that the breakdown of the medical team was a central factor contributing to low morale and disengagement amongst GPs. It is therefore conceivable that developing a sense of collegiality and togetherness across a transgenerational grouping of GPs may boost morale and engagement, leading to greater resilience within the GP workforce. |
| What creates meaning that sustains GPs engagement with, and involvement in, work? | |
| What solutions/recommendations are proposed? | |
| From which stakeholders' perspective? (General practitioners, patients, other HCPs) | 39 participants from all stages of the GP career: from medical students interested in GP to retired GPs. |
| How is work-related joy constructed? (Diversity of ways individuals may think about joy) | |
| What prevents/removes work-related joy? | |
| Organisational characteristics (small, large) | |
| Managing ill-defined illnesses and chronic complex multimorbidity | |
| Human connection with patients | |
| Human connection with peers | Paired analysis showed a strong sense of professional social connection and good opportunities to network. Participants also felt the LSNs provided opportunity to meet colleagues they would not ordinarily meet and felt a strong sense of 'being inspired' by their peers.<br><br>The focus group and free-text answers to the questionnaire demonstrating that LSNs supported the workforce through increased knowledge, communication, networking, resilience and well-being. An increase in clinical and non-clinical knowledge was noted by all participant groups. |

| | |
|---|---|
| | • With rapid changes in primary care surrounding COVID-19 pandemic, LSN's helped disseminate clinical COVID-19 information across the spectrum of primary care.<br>• Participants recognised the positive impact networking offered by increasing their exposure to other career stages of general practice in the form of longitudinal support<br>• The unprecedented changes in primary care as a result of the COVD-19 pandemic enabled the LSNs to provide a mechanism for peer support and helped increase well-being. Participants from all backgrounds agreed that the LSN gave a forum for them to discuss their experiences during the pandemic and had a positive impact on their mental well-being and resilience. |
| Human connection across other organisations (e.g., secondary care) | |
| Learning systems (e.g., competence, capability, mastery, socially situated) | |
| Additional useful information | |
| Summary of findings | This study created groups comprising GPs at lots of different career stages to foster cross-generational collegiality. Surveys and focus groups with participants demonstrated that this supported the workforce through increased knowledge, communication, networking, resilience and well-being. |
| Strengths, limitations, and future research directions | |

| | |
|---|---|
| Paper Number | 117 |
| Author | NHS England |
| Year | 2019 |
| Title | Interim NHS People Plan |
| Journal | [Policy document] |
| Region (England, Scotland, Wales, and Northern Ireland) | England |

| | |
|---|---|
| Study methodology | [Policy document] |
| Type of document | Grey literature |
| Topic or focus of paper | NHS Workforce |
| Research objectives and focus | Set out NHSE's vision for the NHS workforce to enable the delivery of the NHS Long Term Plan |
| Theoretical framework/theories/ underpinnings | |
| Description of intervention | |
| Rationale for paper/review | Changes are needed in the NHS workforce to deliver the NHS Long Term Plan |
| What creates meaning that sustains GPs engagement with, and involvement in, work? | |
| What solutions/recommendations are proposed? | Recognises the current lack of GPs. Highlights the variety of national schemes already in place to recruit and retain GPs, including the GP Retention Scheme, GP Career Plus, the Local GP Retention Fund, and the GP Health Service. Pledges to take further action moving forward as set out in the General Practice Forward View. Will also implement a new two-year Primary Care Fellowship Programme that offers newly qualified GPs – and nurses entering primary care –a secure contract of employment, working in a role tailored both to their career aspirations and interests and to the needs of local health services.<br><br>"At the same time, we will make general practice a better place to work, providing more opportunities for mentoring and coaching, widening the availability of portfolio roles to offer GPs greater variety in their careers, and addressing burnout through the Practitioner Health Service. We have committed to fund a package of measures to tackle workload pressures and improve retention of GPs and other clinical staff in primary care. This includes practical resources to help establish mentoring/coaching, portfolio roles, and greater flexibilities for GPs and other primary care clinicians at all stages in their careers."<br><br>Highlights the expansion of community multidisciplinary teams aligned with primary care networks, as set out in the NHS Long Term Plan. These are anticipated to reduce GP workload. |
| From which stakeholders' perspective? (General practitioners, patients, other HCPs) | |
| How is work-related joy constructed? (Diversity of ways individuals may think about joy) | |

| | |
|---|---|
| What prevents/removes work-related joy? | |
| Organisational characteristics (small, large) | |
| Managing ill-defined illnesses and chronic complex multimorbidity | |
| Human connection with patients | |
| Human connection with peers | |
| Human connection across other organisations (e.g., secondary care) | |
| Learning systems (e.g., competence, capability, mastery, socially situated) | |
| Additional useful information | |
| Summary of findings | This policy document, published by NHSE, sets out how the NHS workforce will be reformed to deliver the NHS Long Term Plan. Relevant to our review, it recognises the current lack of GPs in the workforce and reaffirms NHSE's commitment to GP recruitment and retention. It explains how general practice will be made a better place to work by the expansion of mentoring/coaching, portfolio roles and schemes to address burnout. The document also reiterates how multidisciplinary teams are being rolled out in primary care, which is anticipated to reduce GP workload. |
| Strengths, limitations, and future research directions | |

| | |
|---|---|
| Paper Number | 118 |
| Author | Sampson et al |
| Year | 2016 |
| Title | The relationship between GPs and hospital consultants and the implications for patient care: a qualitative study |
| Journal | BMC Family Practice |

| | |
|---|---|
| Region (England, Scotland, Wales, and Northern Ireland) | Scotland |
| Study methodology | Interviews with GPS and hospital consultants, followed by thematic analysis of responses |
| Type of document | Primary research |
| Topic or focus of paper | Relationships between GPs and hospital consultants |
| Research objectives and focus | Study how both GPs and hospital specialists regard and behave towards each other and how this may influence patient care |
| Theoretical framework/theories/ underpinnings | |
| Description of intervention | |
| Rationale for paper/review | Research suggests that the nature of clinicians' connection at the interface, and how they regard and behave towards each other, may influence patient care . However, very little qualitative research has been carried out with regard to these relationships |
| What creates meaning that sustains GPs engagement with, and involvement in, work? | |
| What solutions/recommendations are proposed? | Addressing barriers to forming relationships (including greater clinician workloads and less shared meeting time) would seem necessary in order to improve effective care delivery. New methods of sharing information across the interface where a new clinician arrives in an area may be usefully established. National and local health authorities may consider the merits of establishing a "universal pause."<br><br>One specific recommendation that may be considered is the formal promotion of "shadowing days" where GPs and specialists spend time in each others "kingdoms," to help them to better understand each others roles, minimising the generation of "unrealistic expectations."<br><br>Joint working groups (involving clinicians from both sides of the interface) to help formulate shared clinical guidance, analyse where things could have gone better, or work together through dumping/resistance issues, may help foster a sense of "we're in this together."<br><br>Clinicians saw education as a tool for developing relationships, with specialists keen to emphasise a model of shared learning. Aligning primary and secondary care protected learning times may be advantageous, given "out-of-hours" educational meetings may be limited in their ability to engage with clinicians who have worked an intense clinical day.<br><br>Presently, there are no standardised approaches to communication across the interface (or shared knowledge on accessibility and preferred method of communication for individual clinicians); this may be an important area of focus, both for future research, and for high-level policy consideration. Embracing newer technologies to assist in this (e.g., email, video-conferencing, online communication) may be of value. |

| | |
|---|---|
| From which stakeholders' perspective? (General practitioners, patients, other HCPs) | GPs and hospital consultants |
| How is work-related joy constructed? (Diversity of ways individuals may think about joy) | |
| What prevents/removes work-related joy? | |
| Organisational characteristics (small, large) | |
| Managing ill-defined illnesses and chronic complex multimorbidity | |
| Human connection with patients | |
| Human connection with peers | |
| Human connection across other organisations (e.g., secondary care) | Participants' responses can usefully be grouped into four main themes; communication, conduct, relationships, and unrealistic expectations.<br><br>Communication<br>• Accessing and Ping-Pong: Clinicians expressed problems with accessing one another; having to go "back and fore." Contextually, for GPs this may have been in relation to seeking specialist advice regarding a patient; for a specialist, this may have been in relation to trying to contact a GP to communicate a significant new diagnosis in a patient. This "Ping-Pong" type of communication also existed where letters transitioned the interface, as each side tried to set boundary lines of responsibility, and was more apparent where a perceived lack of respect between clinicians, and poor working relationships between individual specialists/specialities and GPs/GP practices. "I think there are certain departments where you just think if you try and get, try and speak to a consultant you either won't manage or it will be difficult or it will take lots of time and lots of kind of telephone ping pong.", "I think they think we are difficult to get hold of, I think they think that we are all part time and stuff and that you know the GP is not there again. The GP is not there again because they are maybe out on visits or they are in surgery [...] then they leave a message and you phone them back or you do this kind of ping pong which is intensely frustrating for everybody."<br>• Listening: A sense, for the GP, of being listened to at the point of urgent hospital referral gave an impression of working together for the patient's benefit. There was a sense for both GPs and specialists that knowing one another led to a "better conversation." Likewise, in complex community-based patient scenarios that did not readily fit secondary care systems/guidelines/protocols, |

| | |
|---|---|
| | listening to one another appeared to be even more important in relation to patient care: "I'm just not trying to tick boxes you know what I mean, I know we could get her scoped you know I know that but listen to me I'm trying to tell you something else, I'm trying to say that you know she is frail, I'm wanting you to think a little bit more holistically than that, I want you to you know, I know you can scope people, and I don't even want you to scope her I just wanted you to think a little bit more kind of, I want you to support me making these very difficult decisions cause sometimes I feel I am on my own the whole time."<br><br>Conduct<br>• Dumping: For GPs dumping was related to perceived work transfer (from secondary to primary care) without discussion or subsequent resource reallocation. This work transfer included, for example, responsibility for following up results of secondary care initiated investigation, and the follow-up of patients in the community (where there was a perception of patients being discharged too soon from hospital). GPs alluded to a lack of control over further elements of workload that were apparently being transferred to them on a regular basis. "I personally feel that we are being dumped upon because other colleagues are not completing their job you know, why are they doing that? Are they doing that because their training wasn't sufficient? Are they doing that because they are lazy? Are they doing that because they are too busy? We could all say we are too busy", "well that's when you know the usual games, patient phones up [hospital], speaks to secretary saying I need to be seen sooner, the secretary speaks to the consultant, consultant says tell them to go through their GP, the patient comes to me, [...] I write to the consultant and the reply comes back "which one of my patients do you want me to budge out the way so that your patient gets in first?" So that's the sort of nonsense that I would like to get rid of and for, it's the hiding behind organisations, hiding behind waiting lists, you know it's the culture, if somebody phones up and says I need a home visit today they get one or they get seen, somebody phones a consultant, I write to the consultant saying this patient needs seen, the consultant has no obligation whatsoever to see the patient. It's that lack of ownership of demand, it's for consultants feeling the pressure and feeling the demand and the clinical need as much as we do. That's what I'd like them to share in."<br>• Resisting: This term refers to the opposition of colleagues to take on work handed over to them. This was described to be more evident where poor relationships between primary and secondary care existed. This "resistance" impacted negatively on shared care, leading to a sense of not "being in it together."<br><br>Relationships<br>• Continuing care: Specialists sensed reduced continuity within primary care leading to a felt need to fill a "continuity gap" previously unoccupied by them.<br>• Implications for patient care: While most clinicians felt professional relationships shouldn't impact on patient care, they gave examples of how relationships did impact upon patient care. "Consultants that I know that I have met face to face tend to give quicker and more comprehensive responses regarding referrals that I make either by [electronic hospital referral software] or by email, so knowing them personally makes a huge difference. They write more pleasant letters and are more helpful. Consultants that I don't know are more distant and tend to give more hand offs and bounce people about rather than give a helpful response." |

| | |
|---|---|
| | • Forming relationships: Clinicians listed a number of facilitators to forming relationships, including the use of shared space, the role of education, and purposefully modifying their tone of communication when liaising with interface colleagues. Exclusive to rural GPs was the experience of being able to work closely alongside visiting specialists, which led to them e.g., being able to "sit in" on specialist clinics involving their own patients. The familiarity of one another, in this shared space, allowed GP managed patients to have better access to specialist input. Medical education was felt to be a method of developing interface relationships. A common awareness for older clinicians was that joint educational events were less evident. Barriers to forming relationships were described, with increasing doctor numbers impacting negatively on the ability to establish connections. GPs underlined a lack of information when a new consultant arrived in the area. Specialists highlighted a greater proportion of part-time working, and locum use, as providing difficulty in keeping track of who was who in primary care. Clinicians on both sides acknowledged excessive workload as a barrier to developing relationships. *Working in each other's kingdom* referred to the perceived benefit of spending time in each other's workplaces, to better understand each other's roles. GPs were keen to spend time in hospitals, in order to improve their knowledge of colleagues, and their work. GPs, however, also wanted specialists to spend part of their training in primary care, to encourage both a greater understanding of the patient in the community, and a greater appreciation of the GP role.<br><br>Unrealistic expectations<br>• For GPs, unrealistic expectations centred on patients being misled as to when secondary care test results would become available (to GP or patient), or in the timing of specialist outpatient follow-up. This led to patients booking appointments in primary care only to be told "the result isn't back yet," which was felt to waste both doctor and patient time. "she saw the consultant last September and was told that she would be reviewed in 3 months time, [...] eventually this month she came back to see me again saying that it's a year since and I was still seeing her every 3 months to make sure there is no obvious clinical recurrence and when we phoned up this time I actually phoned up the secretary rather than my secretary phoning up and said what's going on and they say she is on the list, [...] we are running 11 months behind. I said to them why am I only finding that out now cause actually you would have known, the consultant should have known last year when he said see you in 3 months knowing he wouldn't see her for 11 months and it's to do with the secretary said well the consultants choice is that she is seen in 3 months and it's the hospital who can't fulfil that, it's completely bats, its mad." |
| Learning systems (e.g., competence, capability, mastery, socially situated) | |
| Additional useful information | |
| Summary of findings | This study interviewed GPs and hospital consultants to examine the primary-secondary care interface, and how this might impact patient care. Responses could be grouped into four themes: communication (GPs expressed problems accessing hospital consultants and described how being listened to by hospital consultants gave a sense of working together), conduct (GPs perceived work transfer from secondary to primary care without resource reallocation and hospital consultants being reluctant to take on work), relationships (the primary-secondary |

| | |
|---|---|
| | care relationship was described to impact patient care – and facilitators and barriers to this were discussed) and unrealistic expectations (GPs described patients being misled as to when secondary care test results would become available or the timing of specialist follow up). |
| Strengths, limitations, and future research directions | The research team sought to ensure that a range of demographic and professional perspectives were included within the interview sample, which means that the ensuing sample reflected diversity rather than being representative.<br><br>While the study was located in one health board area, both the broad spectrum of participants and the structural similarity of the primary/secondary care interface to that of other regions provides grounds for transferability of findings.<br><br>Reasons for non-response to study invitation were not sought, and it is not known therefore to what extent differences between those who did/didn't respond would impact upon the generalisability and transferability of study findings. |

| | |
|---|---|
| Paper Number | 119 |
| Author | Brown, Gregory & Gray |
| Year | 2020 |
| Title | The power of personal care: the value of the patient–GP consultation |
| Journal | BJGP |
| Region (England, Scotland, Wales, and Northern Ireland) | N/A |
| Study methodology | Commentary |
| Type of document | Secondary research |
| Topic or focus of paper | Changes to GP work following the pandemic |
| Research objectives and focus | The pandemic has resulted in some radical changes in the ways GPs work, most importantly remote working and shorter consultations. This commentary examines these changes. |
| Theoretical framework/theories/ underpinnings | |
| Description of intervention | |
| Rationale for paper/review | |
| What creates meaning that sustains GPs engagement | |

| | |
|---|---|
| with, and involvement in, work? | |
| What solutions/recommendations are proposed? | The average length of consultations should be set at 15 minutes, as longer consultations are significantly more patient centred and less stressful for GPs.<br><br>To decide the balance between remote and FTF appointments, GP practices need access to good quality data. This will allow practices to fully assess the cost-effectiveness of these new models for different patient populations and plan ahead. How many telephone/video/faceto-face/email consultations in a year? How many of these are for the same medical problem? What is the average consultation duration? What is the level of continuity within the practice? What is the difference between triage and consultation? |
| From which stakeholders' perspective? (General practitioners, patients, other HCPs) | |
| How is work-related joy constructed? (Diversity of ways individuals may think about joy) | |
| What prevents/removes work-related joy? | Shorter consultations are more stressful for GPs. Anecdotal reports show that in some practices GPs are now in contact with patients at only 6-minute intervals. An international consensus exists that 10-minute consultations are too short, and the UK is an outlier with so many short consultations. Additionally, continuity of GP care is lower in the UK than in many other countries from Canada to Germany, so patients, GPs, and the NHS may be missing out.<br><br>Remote consultations, now widespread, can paradoxically increase overall GP workload and a recent study warns of extra GP work by up to a third. |
| Organisational characteristics (small, large) | |
| Managing ill-defined illnesses and chronic complex multimorbidity | |
| Human connection with patients | |
| Human connection with peers | |
| Human connection across other organisations (e.g., secondary care) | |

| | |
|---|---|
| Learning systems (e.g., competence, capability, mastery, socially situated) | |
| Additional useful information | Two recent systematic reviews show that continuity of care with doctors, including within primary care, is associated with reduced mortality in patients. In addition, GP continuity achieves significantly fewer hospital admissions and lower costs across whole healthcare systems. GP empathy has been associated with patient satisfaction, and a sense of security for patients, fewer physical complications in diabetes, and reduced all-cause mortality. However, these studies were based, preCOVID-19, on face-to-face GP consultations, which are now much reduced. Moreover, there is research evidence that GPs are significantly less effective when consulting on the telephone or when triaging requests from patients for new consultations. |
| Summary of findings | This commentary explains that studies demonstrating the benefits of continuity of care were conducted based on FTF GP consultations. As we emerge from the pandemic, we should think carefully about the 'right level' of remote working – and practices need access to good quality data to determine this data. Furthermore, it emphasises that consultation lengths in the NHS are currently too short, which detracts from continuity of care and can lead to burnout on the part of GPs. It recommends that the average length of consultations should be set at 15 minutes. |
| Strengths, limitations, and future research directions | |

| | |
|---|---|
| Paper Number | 120 |
| Author | Johnson et al |
| Year | 2022 |
| Title | New ways of working releasing general practitioner capacity with pharmacy prescribing support: a cost-consequence analysis |
| Journal | Family Practice |
| Region (England, Scotland, Wales, and Northern Ireland) | Scotland |
| Study methodology | Cost-consequences evaluation of a prospective observational cohort study in 15 urban practices involving 69 GPs in 1 locality serving a population of 82,000 people. GPs recorded the time they spent addressing key targeted prescribing activities during 5 distinct 2-week audit periods. Pharmacists performed these key prescribing activities to release GP capacity. An additional 225 h of pharmacists' time per week was committed to the locality. Standardized staff costings were used to estimate the financial impact. Prescribing indicator performance was assessed against the other 7 localities within the health board. |
| Type of document | Primary research |
| Topic or focus of paper | Pharmacists in general practice |

| | |
|---|---|
| Research objectives and focus | Evaluate the cost-consequences of additional pharmacists in releasing GP capacity |
| Theoretical framework/theories/ underpinnings | |
| Description of intervention | |
| Rationale for paper/review | General practice in the United Kingdom is experiencing a workforce crisis. Greater multidisciplinary working, including more general practice pharmacists, is seen as part of the solution. However, it is unknown what impact and cost-consequences that pharmacists may have in freeing general practitioner (GP) capacity. |
| What creates meaning that sustains GPs engagement with, and involvement in, work? | |
| What solutions/recommendations are proposed? | |
| From which stakeholders' perspective? (General practitioners, patients, other HCPs) | GPs and pharmacists recorded time taken to address prescribing activities. Safety, quality and cost-efficiency indicators were also collected from practices. |
| How is work-related joy constructed? (Diversity of ways individuals may think about joy) | |
| What prevents/removes work-related joy? | |
| Organisational characteristics (small, large) | On average 3 h of pharmacist time released 1h of GP capacity, requiring an estimated additional investment of £16.73 (range £5.97-20.87) per h compared to employing extra nonsalaried GPs to address key prescribing activities. There were no detrimental changes in safety and quality prescribing measures, and prescribing cost-efficiencies were achieved.<br><br>The self-reported time taken to complete prescribing activities varied by profession, staff banding, and activities being undertaken. The length of time taken to complete the range of activities reduced in the following order: band 7, band 8a, and GP, such that GPs spent approximately half the time addressing immediate discharge letters, and 3 quarters of the time address other medicines issues that pharmacists spent |
| Managing ill-defined illnesses and chronic complex multimorbidity | |

| | |
|---|---|
| Human connection with patients | |
| Human connection with peers | |
| Human connection across other organisations (e.g., secondary care) | |
| Learning systems (e.g., competence, capability, mastery, socially situated) | |
| Additional useful information | Previous studies have reported that pharmacists take more time to complete prescribing activities due to pharmacists being more accurate at documenting medicines changes and eliciting more medicines information, therefore appropriately taking longer to complete the prescribing activities. |
| Summary of findings | This study found that appropriately funded and resourced general practice pharmacy services can free GP capacity, although requires a greater investment than employing extra GPs (possibly due to pharmacists taking longer than GPs to complete key prescribing tasks). The employment of pharmacists was also not associated with detrimental changes in safety and quality prescribing measures, and prescribing cost-efficiencies were achieved. |
| Strengths, limitations, and future research directions | The main strengths of this study are that it addresses a real-world challenge, demonstrating that changes in routine practice can free GP capacity.<br><br>Another strength was assessing differences in staff costs across a range of grades and salaries, as more general practices struggle to employ nonsalaried partners and salaried GP positions become more common; therefore, this study may be of use when considering future service development and planning.<br><br>Social desirability bias may have influenced the selfreporting of the time taken to complete prescribing activities. In some cases, this may have been under or over estimates, as clinicians were not timed while completing activities; however the reductions achieved were similar from Spring 2017 onwards.<br><br>This study was not designed to identify and evaluate the economic impact associated with freeing GP capacity for other activities, or the possible subsequent impact that pharmacist interventions had on other areas of prescribing.<br><br>Finally, some may consider it a limitation that this study was carried out in 1 region. The findings may not be readily generalizable to other areas with different prescribing support resources and relationships, and rural populations, although they may be of interest to others working in similar urban practices and localities<br><br>Further research is needed to assess a range of staffing and service delivery models effectiveness and costs in freeing general practice capacity and streamlining service delivery, to ensure optimal use of resources. |

| | |
|---|---|
| | Future research should consider identifying and exploring the potential educational needs of general practice pharmacists. By addressing these potential learning gaps it may be possible to free more system capacity, but may also support staff's self-efficacy and resilience that enables more optimal service delivery. |

| | |
|---|---|
| Paper Number | 121 |
| Author | Komwong et al |
| Year | 2018 |
| Title | Clinical pharmacists in primary care: a safe solution to the workforce crisis? |
| Journal | Journal of the Royal Society of Medicine |
| Region (England, Scotland, Wales, and Northern Ireland) | United Kingdom |
| Study methodology | Commentary drawing from previously conducted studies about the pharmacy workforce in primary care |
| Type of document | Secondary research |
| Topic or focus of paper | Clinical pharmacists in primary care |
| Research objectives and focus | Discuss the use of clinical pharmacists in general practices to address the objectives of safe prescribing, health outcomes, access to primary care, cost effectiveness and patient satisfaction, along with challenges faced in their integration. |
| Theoretical framework/theories/ underpinnings | |
| Description of intervention | |
| Rationale for paper/review | Workload and time pressures exacerbate prescribing errors. Concerns about workload and access in primary care have led the UK Government to pledge increases in the general practitioner workforce, but general practitioners take at least 10 years to train and declining numbers of medical graduates internationally suggests a limited pool for recruitment. In this article, we discuss integration of clinical pharmacists in general practices as a potential solution to these problems. |
| What creates meaning that sustains GPs engagement with, and involvement in, work? | |

| | |
|---|---|
| What solutions/recommendations are proposed? | Increase the workforce of pharmacists in primary care to alleviate workload, allow GPs to deal with more complex cases, and reduce errors in prescribing |
| From which stakeholders' perspective? (General practitioners, patients, other HCPs) | 2 authors are practicing GPs in the NHS; 3 authors undeclared. |
| How is work-related joy constructed? (Diversity of ways individuals may think about joy) | |
| What prevents/removes work-related joy? | |
| Organisational characteristics (small, large) | |
| Managing ill-defined illnesses and chronic complex multimorbidity | In an aging population with increasing chronic disease and co-morbidity, another significant demand on general practitioners' time is reviewing and managing patients with long-term conditions. Pharmacists' ability of to perform these activities [repeat prescriptions, medication reviews, and reconciliation of medications on letters and discharge summaries are often squeezed into consultations or carried out between seeing patients and after surgeries] has the potential to address the significant general practitioner workload associated with medicines and chronic disease management. This could free up time for general practitioners to focus on other aspects of care and at the same time improve access to primary care. |
| Human connection with patients | Pharmacist consultations in primary care are positively received by patients, who report high satisfaction with care and appreciation of the time offered by pharmacists as well as recognition of their expert drug knowledge. Patients feel comfortable consulting with pharmacists in general practices and appreciate the privacy offered in consulting rooms. They also show greater acceptance of pharmacists as part of the team in general practices, with more appreciation and respect for their advice. |
| Human connection with peers | |
| Human connection across other organisations (e.g., secondary care) | |

| | |
|---|---|
| Learning systems (e.g., competence, capability, mastery, socially situated) | |
| Additional useful information | |
| Summary of findings | Substantial general practitioner workforce growth is unlikely to be achievable in the near future, making consideration of alternative models of primary care essential. Pharmacists integrated into general practices can be seen to offer a practical solution, with the potential to reduce general practitioner workload and improve access, while at the same time improving quality.<br><br>However, challenges such as cost-effectiveness, acceptance of clinical pharmacists, clarity in their role, with uniformity across areas and practices, and definition of competencies for their new extended activities, still need to be addressed. Promotion of the role of pharmacists will also be important, among other healthcare professionals as well as the public, to increase awareness and understanding of their expertise. |
| Strengths, limitations, and future research directions | |

| | |
|---|---|
| Paper Number | 122 |
| Author | Kordowicz et al |
| Year | 2022 |
| Title | Navigating new organisation forms: a qualitative study of primary care networks |
| Journal | BJGP Open |
| Region (England, Scotland, Wales, and Northern Ireland) | UK |
| Study methodology | |
| Type of document | Primary research |
| Topic or focus of paper | |
| Research objectives and focus | To capture GP PCN leaders’ perceptions of the opportunities and pitfalls of PCNs, as well as points of learning, during their inception and development, in order to guide the future development of PCN form and function |
| Theoretical framework/theories/ underpinnings | |

| | |
|---|---|
| Description of intervention | Nine PCN GP leaders were interviewed in depth to gather their views and experiences of PCNs. In addition, 31 free-text survey responses pertaining to how participants perceived the purpose of PCNs were collated |
| Rationale for paper/review | As PCNs are newly created, there is a gap in knowledge concerning how newly appointed PCN leaders navigate these new organisational forms. This study captures insights regarding how PCNs have been experienced by PCN leaders, their perceived limitations and opportunities, as well as recommendations for future PCN and PCN leader development. |
| What creates meaning that sustains GPs engagement with, and involvement in, work? | |
| What solutions/recommendations are proposed? | |
| From which stakeholders' perspective? (General practitioners, patients, other HCPs) | PCN GP Leaders |
| How is work-related joy constructed? (Diversity of ways individuals may think about joy) | Good working relationships<br>Sense of team<br>Effective decision making<br>Collaborative partnership with leaders |
| What prevents/removes work-related joy? | The potential for contractual obligations to be stifling was raised. One participant expressed that many of their achievements happened outside of contractual obligations – but contracts may now prevent change as people might not see the same shared purpose/goal if it's not part of their contract. |
| Organisational characteristics (small, large) | Large – primary care networks |
| Managing ill-defined illnesses and chronic complex multimorbidity | |
| Human connection with patients | |
| Human connection with peers | Emphasis placed on leadership that promotes inclusive, supportive and collaborative working as opposed to strictly operating in a hierarchical structure. |

| | |
|---|---|
| | Good relationships lead to good teamwork, which leads to better decision-making – therefore PCNs with leaders that promote relationship building and PCNs with good relations in place already enable the PCN to carry out their projects. |
| Human connection across other organisations (e.g., secondary care) | There is sentiment that the NHS is "very top-down – the NHS is very command and control" and that there is a relative rigidity in the integrated care system dictating what PCNs are supposed to do.<br><br>Participants…highlighted the need to establish their own power base within the wider institutional context so that PCNs also tell the integrated care system what is needed.<br><br>Local autonomy to meet the population needs would be greatly appreciated. |
| Learning systems (e.g., competence, capability, mastery, socially situated) | |
| Additional useful information | |
| Summary of findings | Purpose-setting, flexibility to adapt to local needs, good working relations and facilitative leadership that promotes collaborative working are some things identified as crucial to the success of a PCN.<br><br>PCN leaders appear to be walking a tightrope between a traditional hierarchical response to the challenges of contracting, workforce, and variation in quality across general practice, and a networked response to meeting needs, collaborating for acuity, and innovating with local assets. |
| Strengths, limitations, and future research directions | - In-depth interview show reflections across several PCNs<br>- Survey response triangulation brought more insights<br><br>- Transferability may be limited – interviews only with GPs<br>- Self-selected sample<br>- Perhaps a biased view as participants played a significant role in the leadership and organisation of their PCN and therefore arguably displayed greater engagement with the reorganisation initiative |

| | |
|---|---|
| | - Future directions – quantifiable impact of PCNs on quality of clinical care and disease management |

| | |
|---|---|
| Paper Number | 123 |
| Author | Edwards, Coward & Carey |
| Year | 2022 |
| Title | Barriers and facilitators to implementation of non-medical independent prescribing (IP) in primary care in the UK: a qualitative systematic review |
| Journal | BMJ Open |
| Region (England, Scotland, Wales, and Northern Ireland) | UK |
| Study methodology | Systematic review of qualitative studies relating to non-medical IP, followed by thematic analysis of extracted data. |
| Type of document | Primary research |
| Topic or focus of paper | Non-medical independent prescribing |
| Research objectives and focus | Explore stakeholders' views on IP in primary care and identify barriers and facilitators influencing implementation. |
| Theoretical framework/theories/ underpinnings | |
| Description of intervention | |
| Rationale for paper/review | To support workforce deficits and rising demand for medicines, independent prescribing (IP) by nurses, pharmacists and allied health professionals is a key component of workforce transformation in UK healthcare. |
| What creates meaning that sustains GPs engagement with, and involvement in, work? | |
| What solutions/recommendations are proposed? | |
| From which stakeholders' perspective? (General practitioners, patients, other HCPs) | Stakeholders involved in implementing independent prescribing schemes, which included GPs. |

| | |
|---|---|
| How is work-related joy constructed? (Diversity of ways individuals may think about joy) | |
| What prevents/removes work-related joy? | IP was recognised as more efficient than former methods of nurses, pharmacists and other AHPs accessing prescribed medications. These involved request, referral and/or counter-signing of prescriptions by doctors, which could increase GP workload (and also burdened patients). By removing doctor input, IP improved responsiveness with respect to medicines, enhanced care quality and helped prevent adverse outcomes.<br><br>GPs in one study stressed that their acceptance of pharmacist IP rested on whether it increased existing workload.<br><br>"Because I have to write, send it to the GP, it has to land on the GP's desk, then the patient has gotta make an appointment to see that GP, then the prescription comes from the GP, and then they go and fulfil that prescription, and then make an appointment to come back and see me." |
| Organisational characteristics (small, large) | |
| Managing ill-defined illnesses and chronic complex multimorbidity | |
| Human connection with patients | |
| Human connection with peers | Lack of team clarity and transparency on IP role intentions were persistent barriers to implementation. Poor team understanding of IP could limit integration, and promote role ambiguity.<br><br>Doctors, receptionists, dispensing pharmacists and peer colleagues all played supervisory and/or infrastructural roles in IP implementation and understanding the need for this input was essential.<br><br>Respectful, trusting interprofessional relationships promoted an appreciation of different professional skill sets, helped ratify the purpose of IP and built team confidence in the prescribing competence of nurses and pharmacists. Good relationships facilitated information transfer, promoted supervision provision, shared learning and team working. Acceptance and positive attitudes towards IP as a shared skill were facilitative to implementation and mitigated the likelihood of 'turf wars' emerging if IP roles was perceived to encroach on professional territories. While many nurses/pharmacists reported positive relationships with doctors, others described jurisdictional tensions over prescribing authority.<br><br>Given the important supervisory role of doctors in IP, consideration of their strength in adoption planning is pertinent. Good communication networks were more likely where established relationships and positive attitudes towards IP prevailed, and were important for imparting information to teams about IP, for developing supervision and peer support and promoting teamwork. |

| | |
|---|---|
| | One study found that the specific models of employment/funding influenced how well IP roles were integrated, with direct GP practice employment as opposed to commissioned Primary Care Trust (PCT) funded roles creating greater sense of permanence, better role use and enhanced team involvement. This was assumed to result from improved relationships, trust and team building. |
| Human connection across other organisations (e.g., secondary care) | |
| Learning systems (e.g., competence, capability, mastery, socially situated) | |
| Additional useful information | |
| Summary of findings | 'Traditional' methods of AHPs accessing prescribed medications, like countersigning by doctors, are inefficient and can increase GP workload. IP has been recognised as more efficient and can enhance care quality. Facilitators to IP implementation, relevant to our review, include good team relationships, good communication networks and acceptance towards IP. Barriers included poor team understanding of the IP role, and jurisdictional tensions over prescribing authority with GPs. |
| Strengths, limitations, and future research directions | We did not however include grey literature and although qualitative synthesis enabled rich description of elements perceived by stakeholders to influence implementation of IP in the UK, reviews that include quantitative literature in primary care are encouraged. |

| | |
|---|---|
| Paper Number | 124 |
| Author | Lockyer et al |
| Year | 2014 |
| Title | The GP retainer scheme: report of a national survey |
| Journal | Education for Primary Care |
| Region (England, Scotland, Wales, and Northern Ireland) | UK |
| Study methodology | Online survey of GP retainers, followed by interviews with selected respondents. Subsequent thematic analysis of data. |
| Type of document | Primary research |
| Topic or focus of paper | GP retainer scheme |
| Research objectives and focus | Evaluate the experiences and views of current and past GP retainers and provide a rigorous assessment of the retainer scheme |

| | |
|---|---|
| Theoretical framework/theories/ underpinnings | |
| Description of intervention | |
| Rationale for paper/review | The current context of organisational change and new working patterns, together with the high cost of medical training, mean it is of vital importance that the NHS retains its trained workforce. The GP retainer scheme supports doctors who for reasons of personal circumstance are restricted in their ability to compete for employment in medicine, and aims to facilitate the retention of their skills and confidence. However, there is small existing literature on the GP retainer scheme. |
| What creates meaning that sustains GPs engagement with, and involvement in, work? | The predominant reason for GPs to join the retainer scheme was the need to work around child-care responsibilities (92.2%). Health was the reason for 2.6% of respondents; 1% were caring for people other than children.<br><br>There was overwhelming support from participants for the retainer scheme. Over 70% of current and ex-retainers responding to the survey rated the scheme as 'extremely helpful': none opted for scale point 1 (not at all helpful). Many respondents wrote of how the GP retainer scheme had kept them in medicine, at a time when they would otherwise have been forced to consider giving up the career for which they had trained. Specifically, when compared to locum work, respondents liked the opportunity to work in a stable practice environment with a set number of sessions and to receive performance review. They also appreciated the ability to fit child care into the working week and have continuity of patient care. Illustrative quotes include:<br>• “It has been a Godsend for me. Working as a Locum was difficult with erratic support. Getting on the scheme has enabled me to increase my confidence and ability to be a GP as well as renew my ambition to keep working. It opened up a middle road where I could go on caring for my family without losing touch (perhaps forever) with medicine.”<br>• “Able to provide continuity of care with the learning opportunities and satisfaction that comes with following patients up. Being part of a team.”<br>• “A steady job with set hours (very important with two children in nursery). Paid annual leave and study leave.”<br>• “A reliable position in a practice where I feel part of the team and able to ask questions if I need to.” |
| What solutions/recommendations are proposed? | 1. Publicise retainer scheme to increase awareness amongst GPs and draw attention to its applicability to male doctors, and to those using the scheme for reasons other than childcare responsibilities.<br>2. The British Medical Association could consider revising the model contract for GP retainers to provide a more realistic structure which meets the needs of both retainers and practices. |

| | |
|---|---|
| | 3. To improve the implementation of the scheme, Local Education and Training Boards (LETBs) and deaneries need to give more definitive and consistent guidance to practices and to retainers. and ensure robust policing of the scheme in practice.<br>4. Retained doctors would welcome deanery and LETB support for a process in which the retainer and the practice agree a contract, and support for retainers and practices in situations of disagreement over entitlements.<br>5. Consideration should be given to extending the scheme or making more flexible use of extensions in situation in which needs continue beyond five years.<br>6. A review of skills, confidence and competence should take place before the final year of the scheme and suggestions made for widening experiences to eliminate areas where confidence is found to be lacking. |
| From which stakeholders' perspective? (General practitioners, patients, other HCPs) | GP retainers |
| How is work-related joy constructed? (Diversity of ways individuals may think about joy) | |
| What prevents/removes work-related joy? | A minority of respondents spoke of negative experiences and were critical of the way the scheme operates in some practices. Complaints included a feeling of being exploited as a cheap form of labour and related difficulties in accessing the educational entitlement within the scheme. A number of respondents argued the need for increased monitoring of the scheme by deaneries, to regularise employment practices:<br>• "My practice would not give me the BMA recommended contract for retainer GPs and the deanery do not check this. At times I feel overworked and just a cheap pair of hands for practice. Would be useful if deanery checked on workloads and stopped practices doing this."<br><br>The scheme was described by a small number of respondents as 'chaotic' and 'disorganised' and some GP retainers were aware of disparities in the implementation of the scheme across practices.<br><br>Other issues raised included the limitation of the scheme to five years, which is not long enough for women with more than one child, and related concerns about what would happen after the end of the period allowed on the scheme. A lack of guidance on exit plans was mentioned by some.<br><br>There was felt to be a need for flexibility in allowing retainers to also work as locum doctors to widen their experience and network of contacts, to support future employment |

| | |
|---|---|
| Organisational characteristics (small, large) | |
| Managing ill-defined illnesses and chronic complex multimorbidity | |
| Human connection with patients | |
| Human connection with peers | Of those currently on the scheme, 93.8% said they did receive clinical support; 88.2% of those who had left the scheme reported receiving clinical support. |
| Human connection across other organisations (e.g., secondary care) | |
| Learning systems (e.g., competence, capability, mastery, socially situated) | A wide range of opportunities for skills development were highlighted by survey respondents currently working as retainers, and those who had left the scheme: 90.5% of current retainers, and 85.7% of those who had left the scheme, considered they had opportunities to maintain consultation skills; 87.7% of current retainers, and 86.7% of past retainers, had the opportunity to undertake home visits; 80.1% of current retainers, and 68.6% of those who had left the scheme, said they had paid protected time for CPD.<br><br>Interviewees were asked how confident they felt about working in general practice. Confidence levels were generally high: "I work four sessions a week and I do all the things that all the other GPs do .... I feel clinically far more confident .. . when the retainer scheme comes to an end that I will be able to go out there and market myself."<br><br>Areas where respondents commonly lacked confidence included management skills, on-call work and performing minor operations. |
| Additional useful information | The peak age for women doctors to leave general practice of 35 years compared to a peak age of 57 years for men leaving general practice. |
| Summary of findings | The retainer scheme is primarily used by women GPs as a way of continuing to practice whilst caring for young children, although a minority are using it to return to work following illness or disability. GP retainers report overwhelmingly positive experiences, particularly due to the opportunity to work in a stable practice with a set number of sessions and continuity of care. They felt clinically supported and were able to maintain their skills. |
| Strengths, limitations, and future research directions | Future research should follow subsequent career pathways over an extended period and capture the perspectives of practices employing GP retainers. |

| | |
|---|---|
| Paper Number | 125 |
| Author | Sophie Lumley |
| Year | 2020 |

| | |
|---|---|
| Title | Trust me – I'm a millennial GP |
| Journal | BJGP |
| Region (England, Scotland, Wales, and Northern Ireland) | N/A |
| Study methodology | Commentary |
| Type of document | Commentary (academic journal) |
| Topic or focus of paper | Changes to general practice in the coming years |
| Research objectives and focus | Describe changes and innovations the author – a trainee GP – would like to see in general practice |
| Theoretical framework/theories/ underpinnings | |
| Description of intervention | |
| Rationale for paper/review | Little has changed in much of general practice for the last 20 years, despite the evolution of PCTs, CCGs, STPs, and PCNs. However, COVID-19 has shown us innovation can happen quickly with the money and shared agenda to drive it. |
| What creates meaning that sustains GPs engagement with, and involvement in, work? | |
| What solutions/recommendations are proposed? | 1. Conduct partnership management activities at a salaried rate, without the associated risks. This might be achieved via a national salaried contract for GPs – which could include extra pay for extra services (e.g. delivering training) and primary care clinical excellence awards.<br>2. Flexible use of estates: working in a central hub where there is consolidated back office function; large rooms for educational sessions and meetings; and the space to have a multitude of clinicians virtually consulting, with an option to work from home. This allows for socialising with GPs from across the locality; frees up space in dispersed estates for face-to face consulting with complex patients, and those who need review after triage; allows co-location of relevant third-sector organisations, and for some secondary care outpatients to come out in the community; and could facilitate delivery of care outside traditional GP facilities, rotating sites to bring care closer to rural patients or to vulnerable groups.<br>3. Special interests encouraged for all, based on population need and gaps in secondary care provision. |

| | |
|---|---|
| | 4. Shift in service provision from secondary care to primary care, which might comprise extra investigations, monitoring, and prescribing. However, alongside this, there must be: (1) accessible and consistent protocols and advice and (2) clear remuneration for this substantial increase in workload.<br>5. Much more tailored support throughout the career. An economy of scale could facilitate formalising: mentorship, peer support groups, and career breaks for all GPs.<br>6. Utilise retired GPs to support the generations below them. This support is particularly important for those in their first 5 years, where a model to formalise exposure to leadership, teaching, research, or specialist interests would be invaluable for all. |
| From which stakeholders' perspective? (General practitioners, patients, other HCPs) | Trainee GP |
| How is work-related joy constructed? (Diversity of ways individuals may think about joy) | |
| What prevents/removes work-related joy? | GP at the moment can feel like 'being confined to the same building for the next 40 years' |
| Organisational characteristics (small, large) | Partnership described as 'picking 1-7 new best friends and being financially tied to them for the next 30 years'. |
| Managing ill-defined illnesses and chronic complex multimorbidity | |
| Human connection with patients | |
| Human connection with peers | |
| Human connection across other organisations (e.g., secondary care) | |
| Learning systems (e.g., competence, capability, mastery, socially situated) | |

| | |
|---|---|
| Additional useful information | |
| Summary of findings | In this commentary, a trainee GP suggests a number of changes she would like to see in general practice, in the coming years. These include: (1) a national salaried contract for GPs, (2) a hub and spokes model of general practice, (3) encouragement of special interests, (4) shift in services to primary care, alongside increased support and renumeration, (5) more peer-support, particularly from retired GPs and particularly towards GPs in the first 5 years of their career. |
| Strengths, limitations, and future research directions | |

| | |
|---|---|
| Paper Number | 126 |
| Author | Martin Marshall |
| Year | 2022 |
| Title | The workforce crisis in general practice |
| Journal | BJGP |
| Region (England, Scotland, Wales, and Northern Ireland) | N/A |
| Study methodology | Editorial |
| Type of document | Editorial (academic journal) |
| Topic or focus of paper | GP workforce crisis |
| Research objectives and focus | Explain the importance of retention (and recruitment) in addressing the GP workforce crisis |
| Theoretical framework/theories/ underpinnings | |
| Description of intervention | |
| Rationale for paper/review | The volume and complexity of what is done in general practice has increased inexorably over the last two decades. This is because the UK population, is larger, older and more likely to seek professional help for their health concerns. Furthermore, many services have been shifted from secondary to primary care, and advances in technology offer treatments unavailable in the past. Changes to structures and ways of working in general practice have failed to match demand with supply – this must be done by improved retention and recruitment of GPs. |
| What creates meaning that sustains GPs engagement | |

| | |
|---|---|
| with, and involvement in, work? | |
| What solutions/recommendations are proposed? | |
| From which stakeholders' perspective? (General practitioners, patients, other HCPs) | (Former) president of the RCGP |
| How is work-related joy constructed? (Diversity of ways individuals may think about joy) | |
| What prevents/removes work-related joy? | In a recent survey of doctors who had left the workforce, carried out by the General Medical Council, 43% of GPs reported that they resigned because they were at high risk of burnout, more than any other medical specialty. Similarly, a third of practising GPs expect to leave the workforce within the next 5 years. If this intent were realised it would represent a potential loss of over 14 000 GPs. |
| Organisational characteristics (small, large) | |
| Managing ill-defined illnesses and chronic complex multimorbidity | |
| Human connection with patients | Increasing workload has left many clinicians feeling they are unable to provide the quality of care that patients need and expect. Even patient safety is at risk; fears of missing a diagnosis or making a prescribing error haunt a growing number of clinicians on a regular basis. Clinicians often respond to these pressures by leaving or working fewer patient-facing sessions. |
| Human connection with peers | There is doubt as to whether the so-called Additional Roles Reimbursement Scheme in England is making as significant a contribution to the workload crisis as policymakers claim. There are increasing reports of a lack of capacity in many practices to support and mentor new team members, who as a consequence are feeling isolated in an unfamiliar working environment. In the absence of adequate investment in the development of this new workforce their contribution to the crisis may be marginal, at least in the short term. |
| Human connection across other organisations (e.g., secondary care) | |
| Learning systems (e.g., competence, capability, mastery, socially situated) | |
| Additional useful information | |

| | |
|---|---|
| Summary of findings | The increased volume and complexity of general practice must be addressed by improved retention (and recruitment, although this is not relevant to our review). In a survey of doctors who had left the workforce, 43% reported resigning because they were at high risk of burnout. This is often because of workload pressures and doctors being unable to provide the quality of care they expect. Adding 'new' roles into general practice may not help as there are reports of a lack of capacity to support and mentor new team members. |
| Strengths, limitations, and future research directions | |

| | |
|---|---|
| Paper Number | 127 |
| Author | McDermott et al |
| Year | 2019 |
| Title | Achieving integrated care through commissioning of primary care services in the English NHS: a qualitative analysis |
| Journal | BMJ Open |
| Region (England, Scotland, Wales, and Northern Ireland) | England |
| Study methodology | Reviewed CCG application documents, conducted telephone surveys with CCGs, interviewed CCG staff/policymakers who were involved in primary care commissioning and observed primary care commissioning committee meetings. Subsequent thematic analysis of data. |
| Type of document | Primary research |
| Topic or focus of paper | Commissioning of primary care services |
| Research objectives and focus | Analyse how CCGs have responded to the responsibility of commissioning primary care services, and identify challenges and factors that facilitated or inhibited achievement of integrated care systems. |
| Theoretical framework/theories/ underpinnings | |
| Description of intervention | |
| Rationale for paper/review | Since April 2015, Clinical Commissioning Groups (CCGs) have taken on the responsibility to commission primary care services. The quest to achieve 'integrated care' is a priority that has been identified internationally, and primary care has been argued to have a central role in achieving this. |
| What creates meaning that sustains GPs engagement with, and involvement in, work? | |

| | |
|---|---|
| What solutions/recommendations are proposed? | |
| From which stakeholders' perspective? (General practitioners, patients, other HCPs) | Stakeholders involved in primary care commissioning – either GPs or healthcare 'managers'. |
| How is work-related joy constructed? (Diversity of ways individuals may think about joy) | |
| What prevents/removes work-related joy? | Highlights a recent review of the QoF that concludes that most of the indicators are unlikely to promote the aims of the GP FYFV relevant to primary care, including integrated and patient-centred care.<br><br>Some CCGs were highlighted as 'using primary care funds to support secondary care budgets'. |
| Organisational characteristics (small, large) | Our case study CCGs could see the opportunities arising from implementing new models of care with some enthusiastic about the direction of travel. However, others were more pessimistic, largely due to resistance to change or change apathy: "I think there's some hesitation by our GPs. So, we're a member organisation. I think there's some worry over what it means with our GP colleagues across the patch and ...I think there's a cohort of people who see it as an opportunity to shape the future and then there's a cohort of people who think, you know, it's concerning about the future of general practice." – GP |
| Managing ill-defined illnesses and chronic complex multimorbidity | |
| Human connection with patients | |
| Human connection with peers | |
| Human connection across other organisations (e.g., secondary care) | Collaborative working was something that could not be taken for granted but must be worked on and facilitated. Factors with the potential to facilitate collaborative working included having a trusted peer who could convince their GP colleagues of the merits of collaborating and adopting a slow and iterative approach when introducing changes affecting practices. "You can't just throw half a dozen professionals in a room and just expect them to work together, because integration is more than co-location in my view." – GP |
| Learning systems (e.g., competence, capability, mastery, socially situated) | |
| Additional useful information | |

| | |
|---|---|
| Summary of findings | Much of this paper focusses on evaluating the success of CCGs in commissioning primary care to achieve integration, hence is not relevant to our review. However, it does highlight that the QoF is unlikely to align with the aims of the GP FYFV, that some GPs are pessimistic about new models of care due to resistance to change apathy, and that collaborative working must be worked on and facilitated. |
| Strengths, limitations, and future research directions | |

| | |
|---|---|
| Paper Number | 128 |
| Author | Moffatt et al |
| Year | 2018 |
| Title | Physiotherapy-as-first-point-of-contact-service for patients with musculoskeletal complaints: understanding the challenges of implementation |
| Journal | Primary Health Care Research & Development |
| Region (England, Scotland, Wales, and Northern Ireland) | England |
| Study methodology | Interviews and focus groups with stakeholders involved in implementing physiotherapy self-referral schemes. Subsequent thematic analysis of responses. |
| Type of document | Primary research |
| Topic or focus of paper | Physiotherapy self-referral schemes |
| Research objectives and focus | 1. Identify the value that stakeholders attribute to a physiotherapy-as-first-point-of-contact-service (PFPCS) within primary care<br>2. Understand barriers to its effective implementation<br>3. Explore the impact of a PFPCS on the working practice of providers |
| Theoretical framework/theories/ underpinnings | |
| Description of intervention | |
| Rationale for paper/review | Primary care faces unprecedented challenges. A move towards a more comprehensive, multi-disciplinary service delivery model has been proposed as a means with which to secure more sustainable services for the future. One seemingly promising response has been the implementation of physiotherapy self-referral schemes, however there is a significant gap in the literature regarding implementation. |
| What creates meaning that sustains GPs engagement with, and involvement in, work? | |

| | |
|---|---|
| What solutions/recommendations are proposed? | 1. If services are built and promoted purely on the premise that they will reduce GP workload by 30%, they will arguably fail to meet that target<br>2. PFPCS provides an opportunity to develop expertise in MSK across the team – but must be mindful of potential for de-skilling the GP |
| From which stakeholders' perspective? (General practitioners, patients, other HCPs) | Physiotherapists, administration/reception staff, general practitioners (GPs) and one practice nurse |
| How is work-related joy constructed? (Diversity of ways individuals may think about joy) | |
| What prevents/removes work-related joy? | |
| Organisational characteristics (small, large) | |
| Managing ill-defined illnesses and chronic complex multimorbidity | |
| Human connection with patients | Benefits for patient experience and well-being were widely discussed and universally perceived as positive: "If you have an injury and you have to wait 7 weeks [to access traditional out-patient physiotherapy services], that's lost the window of opportunity... the economic and emotional cost to that patient not being able to do stuff for several weeks until they get to the physio isn't great. Whereas, get in early, there's... a whole host of benefits" – GP<br><br>As a primary care service, being able to offer a greater skill mix was viewed as empowering patients to make a decision about their preferred care, and therefore consistent with person-centred values. |
| Human connection with peers | All participants described the benefits of the PFPCS in terms of a strategy to help to manage the unprecedented demand faced by primary care. The justification for this was twofold: redistributing work and 'unburdening' the GP. However, whilst GPs acknowledged this to an extent, they argued that such a simplistic trade off was not feasible: "I think estimates range in the prevalence of musculoskeletal work in general practice... probably twenty, thirty percent. But... quite often that's one of a whole number of problems that they might bring to their GP. So it might be nice to think that seeing a physiotherapist might free the GP up for more time, but... quite often with the patient groups that we're seeing, they may be seeing the GP for other things, and it's an add on, it's an extra thing they want to squeeze in. So it may not have saved much time. It's a hard thing to quantify" – GP.<br><br>The number of patients referred back to the GP, or the creation of 'on-work' was described as minimal. However, this was dependent on having experienced MSK physiotherapists working in an extended scope role: "I worry a little bit if you were putting in more junior, maybe cheaper, physiotherapists in, that they might not be as effective... [the physio] has taken the patient and taken ownership of them... and if it was someone less experienced we might find that we were getting more patients bounced back to us" – GP |

| | |
|---|---|
| | Whilst GPs were satisfied by the service provided by physiotherapists, some were still aware of the potential of something being missed. In one practice, a shared information system allowed the transparency for GPs to 'check' that "[the patient's] history is taken, quite a thorough exam is performed and then there's a plan of action quite clearly documented".<br><br>The high level of MSK expertise offered by the two physiotherapists was widely discussed amongst all the GPs. Indeed, some acknowledged that, 'actually, a lot of GPs do not have great MSK skills' (GP4), and that 'patients... are quite quick to pick up on that side of things and say... "I'd like to see a specialist". But actually, if they're seeing a physio who is very knowledgeable... it ticks that worry in their head' (GP4). |
| Human connection across other organisations (e.g., secondary care) | |
| Learning systems (e.g., competence, capability, mastery, socially situated) | Both the GPs and the practice nurse discussed the opportunities for upskilling. However, alongside this, there was a pragmatic concern amongst some GPs that the 'siphoning off' of MSK cases would ultimately have a deleterious effect for their personal professional skills: "This is a very effective service for patients, but it's also taking it away from the GPs... personally I like doing the musculoskeletal things, so I may be losing out a little bit" |
| Additional useful information | |
| Summary of findings | Physiotherapy self-referral schemes were perceived to improve patient care and empower patients. In turn, this helped reduce pressures on GPs, although not to the extent that some commentators have suggested because patients often present to their GP with multiple problems. Furthermore, the success of the scheme depended on the physiotherapists having appropriate experience and training. The presence of physiotherapists – who often have better MSK knowledge than GPs – created opportunities for the upskilling of GPs, however there is a potential for GP to de-skill if they don't see any MSK patients. |
| Strengths, limitations, and future research directions | It is possible that alternative insights may have been elucidated had a larger and more diverse sample been used; participants were drawn from only two general practices and as such the results are context specific.<br><br>Patients' perspectives were not sought in this qualitative evaluation; an evaluation of the patient perspective is ongoing. Given the key theme concerning socio-cultural expectations, future research should consider elucidating this perspective.<br><br>THis evaluation took place just over 12 months from the PFPCS being initiated. Cultural changes in practice delivery and stakeholder perceptions might be expected to take years to become evident.<br><br>Further quantitative evaluation would also be illuminating particularly with regards to the perceived hierarchy in primary care; data regarding the number of patients who declined PFPCS in favour of the GP would be of significance.<br><br>Finally, whilst GPs' concerns regarding the autonomous work by physiotherapists were unfounded in this study, further large-scale, longitudinal studies should explore patient safety. |

| | |
|---|---|
| Paper Number | 129 |
| Author | Murray, Murray & Donnelly |
| Year | 2016 |
| Title | Systematic review of interventions to improve the psychological well-being of general practitioners |
| Journal | BMC Family Practice |
| Region (England, Scotland, Wales, and Northern Ireland) | |
| Study methodology | Systematic review |
| Type of document | Secondary research |
| Topic or focus of paper | GP psychological well-being |
| Research objectives and focus | 1. Evaluate systematically the research evidence regarding the effectiveness of interventions designed to improve General Practitioner (GP) well-being across two continua; psychopathology (mental ill-health focus) and ‘languishing to flourishing’ (positive mental health focus).<br>2. Explore the extent to which developments in well-being research may be integrated within existing approaches to design an intervention that will promote mental health and prevent mental illness among these doctors. |
| Theoretical framework/theories/ underpinnings | |
| Description of intervention | |
| Rationale for paper/review | The health of doctors who work in primary care is threatened by workforce and workload issues. There is a need to find and appraise ways in which to protect their mental health, including how to achieve the broader, positive outcome of well-being. |
| What creates meaning that sustains GPs engagement with, and involvement in, work? | Two studies that employed cognitive-behavioural interventions reported significant short-term improvement in psychological distress indicated by GHQ12 scores. In one of these studies, stress-management techniques was taught, whereas in the other GPs were group coached in cognitive-behavioural techniques addressing issues such a coping skills and time management.<br><br>One study interpreted GP self-reported GHQ scores and developed a mailed, individually-tailored guide to increase awareness about mental health risk among GP intervention ‘cases’ and provide them with self-help advice. This also reported significant short-term improvement in psychological distress indicated by GHQ12 scores.<br><br>One study employing a mindfulness based intervention reported significant improvement in burnout, mood disturbance, empathy and mindfulness immediately postintervention. Weekly sessions that included didactic presentations on awareness of thoughts, feelings, self-care and setting boundaries; formal mindfulness meditation (facilitating non-judgemental awareness); narrative and appreciative inquiry |

| | |
|---|---|
| | (looking for the positive in organizations by identifying current and potential processes that work well) and group discussion. This intervention also had an 8 hour session of silent mindfulness. |
| What solutions/recommendations are proposed? | |
| From which stakeholders' perspective? (General practitioners, patients, other HCPs) | N/A |
| How is work-related joy constructed? (Diversity of ways individuals may think about joy) | |
| What prevents/removes work-related joy? | |
| Organisational characteristics (small, large) | |
| Managing ill-defined illnesses and chronic complex multimorbidity | |
| Human connection with patients | |
| Human connection with peers | |
| Human connection across other organisations (e.g., secondary care) | |
| Learning systems (e.g., competence, capability, mastery, socially situated) | |
| Additional useful information | Long-term follow-up of mental ill-health was not reported in any of the studies. No measures of positive mental health or flourishing were reported. |
| Summary of findings | The findings reported in the four included studies suggest that cognitive-behavioural-based and mindfulness based programmes delivered in a group format may reduce GP distress at least in the short-term. Increasing awareness generally and with specific regard to thoughts, beliefs, self-care, personal health and setting boundaries appeared to improve GP mental health. |
| Strengths, limitations, and future research directions | This review is limited by an English language restriction applied at protocol stage to address potential heterogeneity in well-being terminology and constructs. However, it was conducted using robust methodology and identified a substantial research gap. It is the first review to evaluate an extensive body of research pertinent to the optimisation of well-being in GPs. |

| | |
|---|---|
| | The evidence base in this area is limited. There is a clear need for pragmatic randomised controlled trials using validated assessments of the positive construct of wellbeing to identify strategies that will help safe-guard the mental health of doctors working in primary care. |

| | |
|---|---|
| Paper Number | 130 |
| Author | Newman & Fieldhouse |
| Year | [Does not specify] |
| Title | GP Locum Chambers |
| Journal | Policy document published by NHS Midlands and East SHA. |
| Region (England, Scotland, Wales, and Northern Ireland) | |
| Study methodology | Policy document |
| Type of document | Policy document |
| Topic or focus of paper | GP Locum Chambers |
| Research objectives and focus | 1. To encourage locum GPs to start a chambers themselves or join one already established<br>2. To enable other local organisations to seed and grow interest<br>3. To provide information on options for development, costs and benefits as a basis for local discussion. |
| Theoretical framework/theories/ underpinnings | |
| Description of intervention | |
| Rationale for paper/review | GPs are under significant pressure due to increasing complexity of patient demand, changes in the GP workforce to more part-time and sessional roles, new commissioning responsibilities, poor access to GP locums and difficulties in recruitment with many GPs nearing retirement. The innovative model of GP locum chambers can help address these challenges, |
| What creates meaning that sustains GPs engagement with, and involvement in, work? | |
| What solutions/recommendations are proposed? | This paper proposes the local establishment of Locum Chambers (defined in the 'additional information' section) for five main reasons:<br>1. To ensure a high quality of care provided by GP locums, who have ongoing support to meet revalidation requirements<br>2. To meet with greater ease fluctuating and increasing patient demand and fill gaps in practice rotas due to holidays and sickness<br>3. To encourage GP recruitment and retention, particularly in rural areas and inner cities where there are difficulties attracting new doctors |

| | |
|---|---|
| | 4. To support commissioning by enabling GPs to be released from their practices and through providing a pool of experienced and flexible locum GPs as a local resource<br>5. As an attractive alternative to a conventional partnership, salaried or independent locum contract, especially for increasing numbers of female GPs with childcare responsibilities and those with portfolio careers who want to work flexibly.<br><br>This document outlines a number of benefits of Locum Chambers for both GP locums (flexible working pattern combined with personal autonomy, a sense of belonging, reduced administrative burden, ease of booking and invoicing, peer support and reduced isolation, ease of revalidation, educational support and development opportunities) and salaried GPs (potential access to educational and career development, information cascades and peer support if isolated.)<br><br>A number of positive initiatives for sessional GPs will be delivered via Locum Chambers, including: the creation of self-directed learning groups; a contact register to improve communication; education; support with project work; formal talent management and a change in mind set amongst GP partner colleagues – all of which can be delivered through the creation of a locum GP chambers for both locum and salaried GPs. |
| From which stakeholders' perspective? (General practitioners, patients, other HCPs) | |
| How is work-related joy constructed? (Diversity of ways individuals may think about joy) | |
| What prevents/removes work-related joy? | The main problem that sessional GPs encounter as providers of care is isolation, which can impact on professional effectiveness in a number of areas:<br>• Lack of information about systems and support structures<br>• Effects on personal self esteem, motivation, confidence and empowerment<br>• Missed opportunities for professional peer interaction<br>• Increased risk of professional difficulties, due to missed opportunities for 'benchmarking' and lack of feedback<br>• Unintended ignorance of protocols ('enforced underperformance').<br><br>Locum GPs are regarded as Doctor's Outside Managed Organisations and so will receive a "heavier touch" in revalidation. GP locums may experience:<br>• Difficulty in recording and learning from significant events, either untoward or beneficial<br>• Lack of access to a unique username/password when signing in to a practice's computer system, so that any problems may be incorrectly assigned<br>• Low status and hence susceptibility to less positive feedback from patients and colleagues<br>• Problems in complying with complaints procedures and being unable to respond and change their practice accordingly<br>• Greater difficulty undertaking clinical audit. |

| | |
|---|---|
| Organisational characteristics (small, large) | Market analysis identified the following issues as placing a strain on GPs and practices:<br>1. Practices often experience difficulty in accessing locum cover, especially at short notice and over holiday periods, to cover gaps in rotas, surges in patient demand and release GPs for commissioning.<br>2. The quality of traditional locum cover may be variable, with high profile cases calling for a review of out of hours care.<br>3. Some areas are experiencing difficulty in GP recruitment and retention with an ageing GP workforce and many GPs nearing retirement.<br>4. All GPs, including locums, are required to revalidate, while some locum agencies fail to comply with necessary governance processes.<br>5. Locum agencies may be variable in quality and costly to practices.<br>6. There are increasing numbers of salaried and locum GPs (sessional GPs) who account for approximately 40% of the workforce, and have been described as demotivated and disenfranchised, principally due to isolation (see above)<br>7. There is a lack of career pathway for sessional GPs, a problem for succession planning and leadership in general practice in future<br>8. CCGs will need to commission more services based in the community and primary care, instead of high cost secondary care, which is likely to require additional GP capacity.<br><br>Locum and salaried GPs choose these roles for a number of reasons: to gain experience; identify a practice to work in long term; as a positive career choice; balance work and family life; as part of a portfolio; after retirement; to fit in with existing roles; and as partnerships are unavailable. |
| Managing ill-defined illnesses and chronic complex multimorbidity | |
| Human connection with patients | |
| Human connection with peers | |
| Human connection across other organisations (e.g., secondary care) | |
| Learning systems (e.g., competence, capability, mastery, socially situated) | |
| Additional useful information | Locum GP chambers are self-funding professional groups of locum GPs working as a ‘virtual practice’, supported by a team of managers and clinical directors. All business, professional and governance issues relating to the day-to-day engagement of all GPs in the chambers takes place within the chambers’ managed environment. |
| Summary of findings | Introducing Locum Chambers will increase the quality of care provided by locum GPs, make it easier for practices to fill gaps in rotas, increase the retention of sessional GPs, support commissioning and offer an alternative to conventional contracts, particularly for GPs with portfolio careers who want to work flexibly.<br><br>GPs may choose sessional roles for a number of reasons, yet often experience professional isolation, which can limit their clinical effectiveness (again via a number of mechanisms). |

| |
|---|
| Strengths, limitations, and future research directions |

| | |
|---|---|
| Paper Number | 131 |
| Author | Nussbaum et al |
| Year | 2021 |
| Title | Inequalities in the distribution of the general practice workforce in England: a practice-level longitudinal analysis |
| Journal | BJGP Open |
| Region (England, Scotland, Wales, and Northern Ireland) | England |
| Study methodology | Longitudinal study using quarterly General Practice Workforce datasets (GPs, total clinical staff, PAs, pharmacists and paramedics) from 2015–2020 in England. |
| Type of document | Primary research |
| Topic or focus of paper | GP workforce inequalities |
| Research objectives and focus | Examine recent trends in general practice workforce inequalities. |
| Theoretical framework/theories/ underpinnings | |
| Description of intervention | |
| Rationale for paper/review | Inequalities in general practice, including additional roles, have not been examined in recent years, which is a meaningful gap in the literature. Previous research has found that workforce inequalities are associated with health outcome inequalities. |
| What creates meaning that sustains GPs engagement with, and involvement in, work? | |
| What solutions/recommendations are proposed? | Policymakers should consider why practices and networks in deprived areas are relatively understaffed, and how this can be reversed.<br><br>Given the significant scale of the proposed expansion of the primary care workforce, policymakers must urgently consider what evidence-based mechanisms can be used to encourage equitable recruitment of additional roles in primary care. This might include but not be limited to incentivisation of DPC posts in understaffed areas, enhanced training offers for these roles, and offering practices and networks in understaffed areas additional recruitment support. |

| | |
|---|---|
| From which stakeholders' perspective? (General practitioners, patients, other HCPs) | |
| How is work-related joy constructed? (Diversity of ways individuals may think about joy) | |
| What prevents/removes work-related joy? | |
| Organisational characteristics (small, large) | Fewer GPs, total DPC staff, and paramedics per 10 000 patients were employed in more deprived areas. Conversely, more PAs and pharmacists per 10 000 patients were employed in more deprived areas. With the exception of total DPC staff, these observed inequalities widened over time. The unadjusted analysis showed more nurses per 10 000 patients employed in more deprived areas. These values were not significant after adjustment but approached a more equal or pro-poor distribution over time.<br><br>These findings suggest that the increasing presence of PAs, pharmacists, and to some extent, nurses, may partially alleviate the undersupply of GPs in more socioeconomically deprived areas. However, the comparison of FTE and hours per week values in highlighted the limits of this substitution effect. The relative oversupply of nurses, PAs, and pharmacists in higher IMD deciles only accounted for a total of 7.2 additional hours per week, compared with the observed undersupply of GPs, total DPC staff, and paramedics, which totals to 118.3 fewer hours per week in staffing. |
| Managing ill-defined illnesses and chronic complex multimorbidity | |
| Human connection with patients | |
| Human connection with peers | |
| Human connection across other organisations (e.g., secondary care) | |
| Learning systems (e.g., competence, capability, mastery, socially situated) | |
| Additional useful information | |
| Summary of findings | GP practices in deprived areas have significantly fewer hours per week in staffing. They also have 'less skilled' staff, with a greater number of PAs and pharmacists compared to GPs. This may contribute to increased pressures for GPs working in deprived areas. |

| | |
|---|---|
| Strengths, limitations, and future research directions | The Carr-Hill formula, which has not been substantively updated since it was first implemented in 2004, has been criticised for not adequately considering the extra pressures that practices in areas of high socioeconomic deprivation face. As a result, the adjusted SIIs reported may underestimate the true adjusted inequality indices, that is, there may be higher levels of pro-rich inequality than this analysis reports. The reliance on a crude regression analysis masks variation across and within IMD deciles. The publicly available general practice workforce datasets are limited, especially for newly introduced additional roles.<br><br>Further research is needed to better understand the effects of skill-mix redesign in the health workforce. |

| | |
|---|---|
| Paper Number | 132 |
| Author | Kathy Oxtoby |
| Year | 2021 |
| Title | GP life now: problems to tackle, opportunities to grasp, and changes to absorb |
| Journal | BMJ |
| Region (England, Scotland, Wales, and Northern Ireland) | N/A |
| Study methodology | Commentary |
| Type of document | Commentary |
| Topic or focus of paper | The changing role of a GP |
| Research objectives and focus | Examines how the role of a GP is developing, what these changes mean to GPs, and what being a GP could soon look like |
| Theoretical framework/theories/ underpinnings | |
| Description of intervention | |
| Rationale for paper/review | The role of a GP changed dramatically in 2020 |
| What creates meaning that sustains GPs engagement with, and involvement in, work? | |
| What solutions/recommendations are proposed? | “Less bureaucracy, less ‘media bashing’ of doctors, and having less of a regulatory burden from bodies such as the Care Quality Commission and NHS England” |

| | |
|---|---|
| | During the pandemic more people have turned to self-care, and more conditions are being managed at home. This might help reduce the workload of GPs: “Making these services more accessible would be a ‘win-win,’ reducing the need to make appointments with a GP and empowering patients to make the right choices.”<br><br>The RCGP’s wish list:<br>• Longer appointments, so that GPs have more time with patients<br>• Higher trust in the profession, with less arduous regulation<br>• Ongoing investment in technology, so that general practice is digitally enabled<br>• Reduced workload by greatly expanding the GP and practice team workforce<br>• Support for practices serving the most deprived populations<br>• Support for GPs in managing the pandemic, with particular focus on delivering the covid vaccination programme, covid testing, and managing “long covid”<br><br>The BMA’s wish list:<br>• Trusting GPs to do their job<br>• Enabling flexibility of work<br>• Reducing bureaucracy and regulation<br>• Supporting practices to prioritise workload<br>• Investing in IT, premises, and occupational health services |
| From which stakeholders’ perspective? (General practitioners, patients, other HCPs) | Data drawn from a variety of surveys of GPs and a number of interviews with GPs |
| How is work-related joy constructed? (Diversity of ways individuals may think about joy) | Part time working is positive for GPs who are doing this of their own volition, such as wanting a career portfolio or time with their family. |
| What prevents/removes work-related joy? | 47% of GP survey respondents said that their levels of stress, anxiety, and emotional distress had got worse since the pandemic began. Compared to the start of the pandemic, 29% of GPs were now more likely to take early retirement, 22% to leave the NHS for another career, and 25% to take a career break. [This is data from the BMA’s COVID tracker]<br><br>Unlike secondary care clinicians, GPs have very little access to occupational health services.<br><br>There are still a lot of GPs retiring early because it’s the only way to cope with general practice pressures.<br><br>Some GPs are reducing their hours due to the risk of burnout. |

| | |
|---|---|
| | "Pre-covid, stress levels in general practice were pretty high," he says. "There was an element of 'change fatigue' and having to deal with an under-resourced NHS. Then came covid-19, and overnight it pivoted how we work. The most enormous changes I've seen in my career happened in just a few days. But still there was enormous good will from already overworked GPs. |
| Organisational characteristics (small, large) | The option to work as a salaried GP reflects the specialty's flexibility, says the BMA's Vautrey. "A GP may start out as salaried, gain confidence to become a partner, then later choose to be salaried again," he says.<br><br>The past five years have seen an ongoing trend towards salaried posts, driven by GPs quitting partnership roles and newly qualified GPs not wanting them. Over the years the number of GP partners has fallen from 85% to below 50% – Martin Marshall.<br><br>Marshall says that PCNs "are what communities want" and that they will "help take general practice to a new level with a broader, skilled, multidisciplinary team, working to a large scale and sharing staff."<br><br>Grouping practices together in PCNs is "artificial" and "forced," says David Turner, GP partner at Chorleywood Health Centre in Hertfordshire, who wants practices to have the freedom to spend money on frontline roles. "What we want are more doctors and nurses."<br><br>PCNs could potentially result in the end of the traditional partnership model. Gerada believes that "the old idea of being a partner within a practice will disappear. Instead we will be equivalent to stakeholders within a PCN, whether that's for profit as in a shareholder agreement or whether it's a partnership as in a sort of consultant body. But that will be determined by the next generation of doctors."<br><br>Vautrey adds that GPs need time to recruit and that NHS England "isn't giving much freedom as to who is recruited. What NHS England really needs to do is to trust us. We need greater flexibility in terms of the utilisation of the workforce so that we can make the best use of PCN funding." |
| Managing ill-defined illnesses and chronic complex multimorbidity | |
| Human connection with patients | Before the pandemic about 90% of GP consultations were face to face, and only a small proportion were done remotely. At the height of the pandemic this reversed, as around 70% of consultations were done remotely. "The pendulum is now swinging back a bit, and practices are doing 55% of consultations face to face,"<br><br>Vautrey notes that "doctors didn't train to work in call centres. They want to have that regular face-to-face interaction with their patients, which is one of the joys of general practice."<br><br>During the covid crisis "many consultations have by necessity become more transactional, and this has reinforced the mistaken view of some people that general practice is largely a transactional medical specialty rather than a relational one," Marshall told the 2020 RCGP conference. But GPs, he said, know that "the trusting relationship between a patient and a doctor—often but not always based on continuity of care—is the most effective intervention that we have at our disposal. It's what helps us to use medications, investigations, and specialist referrals appropriately and to reduce the risks of medicalising non-medical problems. It's what encourages patients to engage with our advice, [and] it's what many patients want from their GP." |

| | |
|---|---|
| | Gupta hopes that his practice will continue to provide a personal service to patients, as "that is the reason why most GPs have chosen to do this job. We are the only people in the healthcare system who have that relationship with patients and their families, and it would be a huge shame if that personal approach was lost." |
| Human connection with peers | |
| Human connection across other organisations (e.g., secondary care) | |
| Learning systems (e.g., competence, capability, mastery, socially situated) | |
| Additional useful information | For Gupta, remote consultations come with "some 'health warnings.'" He says, "There is a risk that some populations, such as older people or those for whom English is not their first language, who are digitally disadvantaged, might get poorer outcomes as a result of there being too much focus on remote consultations." |
| Summary of findings | GP stress was higher during the pandemic and this had a negative effect on retention. Unlike in hospitals, GPs have little access to occupational health to help deal with this. The flexibility of part-time working may be positive for some GPs, but it is often used as a tool to help GPs avoid burnout. Less GPs are opting for partnership. Remote consulting is on the rise, but it is important to remember that GPs do not want to work in 'call centres'. There is a rise in self-care which may decrease GP workload. PCNs might take general practice to a 'new level' however GPs need to be given flexibility and time to organise these. Some GPs believe that PCNs will lead to the end of the traditional partnership model. Relational continuity between GPs and patients is extremely important to both parties, and improves quality of care. |
| Strengths, limitations, and future research directions | |

| | |
|---|---|
| Paper Number | 133 |
| Author | Roos et al |
| Year | 2014 |
| Title | Motivation for career choice and job satisfaction of GP trainees and newly qualified GPs across Europe: a seven countries cross-sectional study |
| Journal | Education for Primary Care |

| | |
|---|---|
| Region (England, Scotland, Wales, and Northern Ireland) | Europe |
| Study methodology | Surveyed GP trainees and recently qualified GPs in the Czech Republic, Denmark, Germany, Italy, Norway, Portugal and the United Kingdom using a web-based questionnaire. |
| Type of document | Primary research |
| Topic or focus of paper | Reasons for choosing general practice |
| Research objectives and focus | Explore motivation for career choice and job satisfaction of GP trainees and newly qualified GPs (NQGP) across seven European countries. |
| Theoretical framework/theories/ underpinnings | |
| Description of intervention | |
| Rationale for paper/review | Cross-national exploration of motivation for career choice and career satisfaction could help inform workforce planning. |
| What creates meaning that sustains GPs engagement with, and involvement in, work? | |
| What solutions/recommendations are proposed? | |
| From which stakeholders' perspective? (General practitioners, patients, other HCPs) | 3722 GP trainees and newly qualified GPs – UK respondents represent 58.5% of the sample. |
| How is work-related joy constructed? (Diversity of ways individuals may think about joy) | Important reasons for career choice in general practice for UK GPs were Compatibility with family life (female – 76.6%, male – 63.2%), Challenging medically broad discipline (female – 59.8%, male – 61.8%), Individual approach to people (female – 40%, male – 41.2%), Holistic approach to the patient (female – 41.4%, male – 30.1%), Autonomy and independence (female – 18%, male – 34.8%), Strong representation of communicational aspects (female – 20.6%, male – 12.2%).<br><br>For UK GPs, 86.6% were satisfied with the daily time they spend at work, 72% are satisfied with the time left for private life, 83% are satisfied with earnings. |
| What prevents/removes work-related joy? | |

| | |
|---|---|
| Organisational characteristics (small, large) | |
| Managing ill-defined illnesses and chronic complex multimorbidity | |
| Human connection with patients | |
| Human connection with peers | |
| Human connection across other organisations (e.g., secondary care) | |
| Learning systems (e.g., competence, capability, mastery, socially situated) | |
| Additional useful information | |
| Summary of findings | UK GPs chose GPs because of compatibility with family life; medically broad discipline; individualised and holistic approach; independence; and communicational aspects. The majority were satisfied with work-life balance and earnings. |
| Strengths, limitations, and future research directions | The main limitation of this project was the difficulty of knowing how representative the sample was of the overall population of trainee and newly qualified doctors, and therefore how generalisable the results are. |

| | |
|---|---|
| Paper Number | 134 |
| Author | Julia Segar |
| Year | 2015 |
| Title | Under-representation of women on governing bodies: women general practitioners on Clinical Commissioning Groups in England |
| Journal | Journal of Health Services Research & Policy |

| | |
|---|---|
| Region (England, Scotland, Wales, and Northern Ireland) | England |
| Study methodology | Commentary |
| Type of document | Secondary research |
| Topic or focus of paper | This essay examines some of the reasons for the under-representation of women in governing bodies including the predominance of women in the salaried and part-time sector of general practice and gendered management styles within the National Health Service. |
| Research objectives and focus | |
| Theoretical framework/theories/ underpinnings | Re reasons and theories why women are under-represented in governing bodies:<br><br>There are structural issues that are relevant here:<br>- the first is that women GPs make up the majority of salaried and part-time positions<br>- the second is that the numbers of salaried and part-time positions are growing in relation to the numbers of partner positions.<br>- Furthermore, salaried and part-time GPs are under-represented on the governing bodies.<br><br>Management styles also may be a factor. In general terms, heroic leadership styles are more often associated with men and post-heroic styles are more commonly employed by women. Women GPs already facing structural disadvantages may also find it difficult to make headway in an environment that favours masculine management styles. |
| Description of intervention | |
| Rationale for paper/review | In the course of attending a number of these meetings as a researcher, I became increasingly aware of the dearth of women GPs seated at the ‘top table’. We know that women are well represented in general practice, so their absence on CCG governing bodies seemed noteworthy. |
| What creates meaning that sustains GPs engagement with, and involvement in, work? | |
| What solutions/recommendations are proposed? | Employ quotas – so that a certain proportion of those who are hired come from a previously disadvantaged or under-represented group. There are examples of this working in other European countries for various industries and in the UK judiciary system, where the proportion of women on boards of directors or among the upper echelons has increased.<br><br>Commit to a quota of 40% women on the governing bodies to be more representative of the general population, fairer to women GP and bring new leadership styles to governing bodies. |

| | |
|---|---|
| From which stakeholders' perspective? (General practitioners, patients, other HCPs) | Women GPs |
| How is work-related joy constructed? (Diversity of ways individuals may think about joy) | |
| What prevents/removes work-related joy? | |
| Organisational characteristics (small, large) | CCGs |
| Managing ill-defined illnesses and chronic complex multimorbidity | |
| Human connection with patients | Many women choose to consult women GPs (particularly on 'women's issues') |
| Human connection with peers | It is suggested that women and men tend towards different management and leadership styles – usefully summarized by Claes[12] who talks about masculine and feminine poles of behaviour. She notes the association of masculine management skills with dominance, assertiveness and decisiveness, and that relationship building, collaboration and supportive behaviour are associated with women. |
| Human connection across other organisations (e.g., secondary care) | Women make up 48.5% of the GP workforce yet only 20.2% of governing body members on CCG boards were women. |
| Learning systems (e.g., competence, capability, mastery, socially situated) | There are few who would argue that women GPs are less talented than their male counterparts, and few who would contend that women are not able to manage and lead others. |
| Additional useful information | Roles of leadership in CCGs are set to expand in future and there is a drive for GPs to take on these roles – it's important for female GPs to also engage with these. Women GP leaders can act as role models for upcoming generations, inspiring and encouraging other women to take on leadership positions. |
| Summary of findings | This paper largely discusses the reasons for few women being on governing bodies and the importance of needing representation of women GPs on CCG boards for the sake of fairness with their male counterparts, being role models for upcoming generations, reflecting the broader community and their needs, and involvement in the ever-expanding leadership roles that the government is keen to employ GPs in. The paper also suggests a solution to implement in the form of employment quotas, citing examples where gender quotas have been successful in recruiting women leaders. The author calls for affirmative action to push more women towards the "top table". |

| |
|---|
| Strengths, limitations, and future research directions |

| | |
|---|---|
| Paper Number | 135 |
| Author | Smith et al |
| Year | 2017 |
| Title | Factors influencing the decisions of senior UK doctors to retire or remain in medicine: national surveys of the UK-trained medical graduates of 1974 and 1977 |
| Journal | BMJ Open |
| Region (England, Scotland, Wales, and Northern Ireland) | UK |
| Study methodology | Postal and web-based questionnaires (identical to each other) sent to UK medical graduates of 1974 and 1977. |
| Type of document | Primary Research |
| Topic or focus of paper | Factors which influenced doctors' decisions to retire and factors which encouraged doctors to remain in work. |
| Research objectives and focus | To report attitudes to retirement of late-career doctors. |
| Theoretical framework/theories/ underpinnings | |
| Description of intervention | The surveys sent to both cohorts were identical and comprised structured, 'closed' questions and statements, with scope and encouragement for additional free-text comment. |
| Rationale for paper/review | In 2015, over 80% of senior hospital doctors in the UK were considering early retirement, with stress identified as the main cause. The early retirement of experienced doctors creates challenges for the medical workforce, places more stress on remaining staff and may adversely affect patient safety.<br><br>Much of the research focus on how to retain doctors nearing retirement has been on GPs. We have studied the careers of the UK-trained medical qualifiers of 1974 and 1977 periodically from the first year after they qualified. (This paper included doctor of various specialities.) |
| What creates meaning that sustains GPs engagement with, and involvement in, work? | Reduced impact of work-related bureaucracy<br>Workload reduction/shorter hours<br>Financial incentivisation<br>Improved working conditions |

| | |
|---|---|
| What solutions/recommendations are proposed? | |
| From which stakeholders' perspective? (General practitioners, patients, other HCPs) | UK medical graduates of 1974 and 1977 in different specialities |
| How is work-related joy constructed? (Diversity of ways individuals may think about joy) | |
| What prevents/removes work-related joy? | Pressure of work<br>Reduced job satisfaction<br>Lack of financial security<br>Possibility of deteriorating skill/competence<br>The prospect of revalidation |
| Organisational characteristics (small, large) | |
| Managing ill-defined illnesses and chronic complex multimorbidity | |
| Human connection with patients | |
| Human connection with peers | |
| Human connection across other organisations (e.g., secondary care) | |
| Learning systems (e.g., competence, capability, mastery, socially situated) | |
| Additional useful information | Women were more likely than men to retire because of the retirement of a spouse or partner. This gender difference was present in all specialities except surgery, O&G, radiology, pathology and psychiatry.<br><br>Women were more likely than men to retire for family reasons. |

| | |
|---|---|
| | Men were more likely than women to retire for financial reasons in GP.<br><br>Of those doctors who had retired, two-thirds had retired when they had planned to retire and a quarter had an unplanned retirement due to either a change in the work environment or a change in personal circumstances. |
| Summary of findings | This paper focused on reasons of retirement in doctors and reasons that may motivate them to engage with work longer. One of the main reasons for retirement was to have more time on leisure/other interests; another key reason was work pressures. There were notable gender differences in retirement reasons, with women citing family factors or the decisions of a spouse/partner more often than men. The main factors that would retain doctors for longer include a reduction in work-related bureaucracy and reduced workload. Financial incentivisation would also be encouraging. |
| Strengths, limitations, and future research directions | • This is a large nationwide study with a very high response rate.<br>• For retired doctors, the data are based on actual retirements and not intentions.<br>• Doctors who have not yet retired were surveyed at a key stage for retirement planning.<br>• Although a large number of retired doctors responded, some may not have done, and there is a possibility of some responder bias.<br>• The small minority of non-contactable doctors largely comprised those who were unregistered, either through retirement or through having left medicine or the UK. It is possible that, if surveyed, their views on the topics of the paper would differ from those of the respondents. |

| | |
|---|---|
| Paper Number | 136 |
| Author | Spiers et al |
| Year | 2016 |
| Title | Who cares for the clinicians? The mental health crisis in the GP workforce |
| Journal | British Journal of General Practice |
| Region (England, Scotland, Wales, and Northern Ireland) | UK |
| Study methodology | Commentary |
| Type of document | Secondary research |
| Topic or focus of paper | The experience of GPs living with mental health problems, with a call for action to improve the provision of mental health services for GPs |
| Research objectives and focus | |

| | |
|---|---|
| Theoretical framework/theories/ underpinnings | |
| Description of intervention | |
| Rationale for paper/review | The fact that a significant proportion of the UK's GPs are living with mental health problems has been known for some time.<br><br>We have been funded by the National Institute for Health Research School for Primary Care Research to conduct a qualitative study of the experiences of GPs living with mental health problems. We aim to identify what helps and hinders GPs when they seek treatment, so that in future their access to appropriate care and support will be greatly improved. |
| What creates meaning that sustains GPs engagement with, and involvement in, work? | |
| What solutions/recommendations are proposed? | |
| From which stakeholders' perspective? (General practitioners, patients, other HCPs) | GPs |
| How is work-related joy constructed? (Diversity of ways individuals may think about joy) | |
| What prevents/removes work-related joy? | Stress<br>Burnout<br>Depression<br>Anxiety<br>Increased workload<br>Negative media climate<br>Sense of isolation<br><br>Just as they would for any other member of the population, mental health difficulties take their toll on all aspects of GPs' lives, including self-esteem, personal relationships, finance, work–life balance, and work performanc |
| Organisational characteristics (small, large) | |

| | |
|---|---|
| Managing ill-defined illnesses and chronic complex multimorbidity | |
| Human connection with patients | Despite the clear and critical effect on GPs themselves, it is striking how frequently existing narratives suggest that physician health only matters because of its potential negative impact on patients. While doctors are encouraged to see their patients holistically, they are often not afforded the same treatment themselves. |
| Human connection with peers | While other emotionally demanding professions, such as psychotherapy, offer regular supportive supervision, GPs are often left to manage their emotional responses to their patients alone.<br><br>On presentee-ism: GPs may continue working because they do not wish to let their colleagues down. Some may feel they have no choice. Faced with workforce shortages and difficulty finding locums, GPs are at times forced to work when they are unwell.<br><br>The emergence of Facebook groups such as Resilient GP, GP Survival, and Tea & Empathy, which indicate a grass roots response to the issues, highlight the critical mental health crisis of GPs and doctors as a whole. |
| Human connection across other organisations (e.g., secondary care) | There is evidence that GPs have difficulty accessing appropriate mental health or support services,[2] for reasons around availability or concerns about confidentiality.<br><br>campaigns such as the Royal Medical Benevolent Fund's (RMBF, chaired by *BJGP*'s Editor Professor Roger Jones) 'What's Up Doc', which has provided a guide called *The Vital Signs*,[9] designed to help doctors recognise and cope with symptoms of stress and burnout.<br><br>The Practitioner Health Programme already runs an excellent local specialist service helping doctors and dentists living in London with mental health concerns; services outside London are limited.<br><br>The Department of Health is investing in an occupational health service for doctors living with mental health problems.<br><br>However, some local services such as the GP and NHS Dentist Physician Health (GPDPH) in Cornwall, which have helped around 1000 GPs and dentists over 20 years, have been threatened with closure due to reduced funding |
| Learning systems (e.g., competence, capability, mastery, socially situated) | |
| Additional useful information | Many proposed interventions emphasise GP time management and the development of 'resilience', and encourage solutions such as mindfulness. In contrast, there is little work on how organisations can better support their workforce and how the systemic factors contributing to mental ill health can be addressed. |
| Summary of findings | It is important to understand the factors that underly mental health problems among GPs; discourage the unhealthy tendency to ignore illness and continue to work; and improve access to appropriate care and support. This article sets the foundation for future research on the mental health of GPs to occur, starting with publication of the authors' own data. |
| Strengths, limitations, and future research directions | |

| | |
|---|---|
| Paper Number | 137 |
| Author | Stavely and Sampson |
| Year | 2020 |
| Title | Will the GP workforce crisis be solved by top-down initiatives? |
| Journal | BJGP |
| Region (England, Scotland, Wales, and Northern Ireland) | UK |
| Study methodology | Commentary |
| Type of document | Secondary Research |
| Topic or focus of paper | Approaches to improve GP retention |
| Research objectives and focus | Sustainable retention initiatives that get to the root of issues causing GPs to leave the profession, work less than full time, and burn out should be prioritised.<br><br>Furthermore, retention needs to be part of a quality improvement approach. Evidence- based initiatives should be implemented and, where more experimental approaches are used, these must be fully evaluated. |
| Theoretical framework/theories/ underpinnings | |
| Description of intervention | |
| Rationale for paper/review | In the next 5 years, 8500 GPs plan to leave the health service;[5] many others are choosing to work less than full time.<br><br>Furthermore, many GPs who remain are burning out, resulting in poor care for patients as well as damage to their own health and wellbeing.<br><br>In 2017, 100 million GBP was committed to recruit 3000 doctors from abroad by NHS England.[8] Despite the investment in international recruitment, few GPs have been recruited; there is a global shortage of healthcare workers.[9] Retention funding per doctor has been proportionally smaller than that spent on international recruitment per doctor. This must change. |

| | |
|---|---|
| What creates meaning that sustains GPs engagement with, and involvement in, work? | Collaborative working at scale, allowing doctors to feel valued and enabled to start quality improvement projects<br>Autonomy<br>Belonging<br>Competence |
| What solutions/recommendations are proposed? | It would be better for regions to have a pot of retention money with a pick-and-mix set of retention options so that they could localise and distribute according to need.<br><br>Influential bodies should emphasise that these retention schemes do not stand alone. They must be part of the Long Term Plan's move to stabilise and strengthen general practice.<br><br>A primary care staff satisfaction survey (PCSSS) would support those implementing retention initiatives to understand and measure the factors that are instrumental to effective workforce retention.<br><br>A survey created for primary care should be solely to support local leaders in designing initiatives to support primary care staff engagement and satisfaction rather than for performance management. Furthermore, the survey should be released as part of a resource pack, which includes a toolkit and set of UK case studies, to support leaders to improve on the results. |
| From which stakeholders' perspective? (General practitioners, patients, other HCPs) | GPs |
| How is work-related joy constructed? (Diversity of ways individuals may think about joy) | |
| What prevents/removes work-related joy? | Lack of clarity regarding funding<br>Lack of sustainable, impactful, localised and flexible retention funding initiatives<br>Disproportionate assurance requirements attached to funding<br>Burnout |
| Organisational characteristics (small, large) | |
| Managing ill-defined illnesses and chronic complex multimorbidity | |

| | |
|---|---|
| Human connection with patients | North Cumbria Primary Care Alliance designed an innovative not-for-profit with key stakeholders to allow their GPs to become salaried. The model has strong support from patient groups. |
| Human connection with peers | For example, in Haringey their federation, Federated4Health, created peer group support for newly qualified GPs with a mentor available by WhatsApp, who made the doctors feel valued and enabled them to start quality improvement projects in their practices.<br><br>the North Cumbria Primary Care Alliance faced a 40% GP vacancy rate. Furthermore, they recognised that their young GPs did not want traditional partnerships and did not want to buy into premises. This was leaving senior GPs trapped, working excessive hours, and unable to sell their premises. |
| Human connection across other organisations (e.g., secondary care) | |
| Learning systems (e.g., competence, capability, mastery, socially situated) | |
| Additional useful information | |
| Summary of findings | The UK has a falling GP-to-patient ratio. Influential bodies must champion and adequately fund locally implemented retention initiatives to ensure that primary care is sustainable for the future and meets the changing needs of our population. Funding needs to be coordinated, recurrent, consistent, and with realistic timelines attached.<br><br>Furthermore, these bodies must coordinate and support systematic measurement of staff engagement and satisfaction in primary care through a survey to improve design of retention initiatives, and to support the development of an evidence base of effective schemes. |
| Strengths, limitations, and future research directions | |

| | |
|---|---|
| Paper Number | 138 |
| Author | Maden et al |
| Year | 2017 |
| Title | General practice: The heart of the NHS |

| | |
|---|---|
| Journal | BJGP |
| Region (England, Scotland, Wales, and Northern Ireland) | England |
| Study methodology | Commentary |
| Type of document | Secondary research |
| Topic or focus of paper | Discussion on delivering the GP Forward View |
| Research objectives and focus | |
| Theoretical framework/theories/ underpinnings | |
| Description of intervention | |
| Rationale for paper/review | Never before has the need for an optimised version of general practice felt more critical to the long-term sustainability of the NHS. |
| What creates meaning that sustains GPs engagement with, and involvement in, work? | Meaningful self care<br>A strengthened primary care team<br>autonomy, sense of purpose, and the opportunity to innovate<br>partners wish to remain autonomous independent contractors<br>salaried GPs looking for a more stable workload and job security<br>increasing number of GPs seeking greater flexibility by carving out a portfolio career<br>options to move between these three roles |
| What solutions/recommendations are proposed? | increased GP training places<br>improvements to both the Induction and Refresher and the Retainer schemes<br>international recruitment pilots<br>post-CCT fellowships<br>salary supplements in hard-to-recruit areas<br>increasing numbers of clinical pharmacists<br>training for new roles such as GP medical assistants, care navigators and physician associates<br>GP Career Plus scheme, designed to offer flexibility and support for GPs considering leaving general practice<br>medical school places are being increased in the future<br>support for increasing indemnity costs that have distorted individuals’ decisions to work<br>streamlining processes of the Performers List<br>a new GP Health Service for GPs and trainees facing burnout |

| | |
|---|---|
| | Overall, nearly 2000 practices will benefit from the support offered through the Vulnerable Practices Fund (£10M) and Practice Resilience Programme funds (£16M in 2016/2017 and £40M total). Furthermore, the £30M General Practice Development Programme will start to support a third of all practices this financial year, and includes 400 free leadership-training places a year for those working in primary care |
| From which stakeholders' perspective? (General practitioners, patients, other HCPs) | NHS England |
| How is work-related joy constructed? (Diversity of ways individuals may think about joy) | |
| What prevents/removes work-related joy? | Burnout<br>external pressures such as disproportionate regulation and avoidable hospital hand-offs<br>not having a manageable working day, caused in part by the burden of low-value activities that cumulatively challenge delivery of care |
| Organisational characteristics (small, large) | |
| Managing ill-defined illnesses and chronic complex multimorbidity | |
| Human connection with patients | |
| Human connection with peers | our success depends on pulling together as a profession to create a sustainable future for the generation coming through the ranks, and for our patients |
| Human connection across other organisations (e.g., secondary care) | Amid these new models (MCPs and PACS) exciting opportunities for career progression should emerge, for example, by influencing local service reform and fostering closer relationships with secondary care colleagues.<br><br>We also need to redefine our role in orchestrating a strengthened primary care team that includes care navigators, medical administrators, clinical pharmacists, physician associates, mental health therapists, community services colleagues, and the voluntary sector. |
| Learning systems (e.g., competence, capability, mastery, socially situated) | |
| Additional useful information | The article comments on aims to improve practices' service provision to include technology that offers a step-change in utility to patients, connecting clinicians, and streamlining back-office processes, as well as integrating with other providers |
| Summary of findings | |
| Strengths, limitations, and future research directions | |

| | |
|---|---|
| Paper Number | 139 |
| Author | Stobbe, Groenewegen and Schafer |
| Year | 2021 |
| Title | Job satisfaction of general practitioners: a cross-sectional survey in 34 countries |
| Journal | Human Resources for Health |
| Region (England, Scotland, Wales, and Northern Ireland) | 34 countries |
| Study methodology | |
| Type of document | Primary research |
| Topic or focus of paper | Comparing job satisfaction across different countries and the reasons for these differences |
| Research objectives and focus | |
| Theoretical framework/theories/ underpinnings | SPF theory is a theory of goal-oriented behaviour, which assumes that people try to optimise the achievement of universal goals, given their resources and constraints, in order to produce their own well-being<br><br>we define GP job satisfaction as 'the production of positive feelings regarding aspects of their work as GP, influenced by the resources that are available and the present circumstances'<br><br>The SPF theory assumes that people act goal-oriented against the background of their ideas and knowledge about the appropriate means and ends to achieve this [17]. It depends on the social and institutional context in what ways, i.e. through which instrumental goals, they can reach the universal goals of physical and social well-being.<br><br>We theorised that GPs 'produce' job-related well-being through doing stimulating work, deriving comfort, social status and behavioural confirmation from their work. |
| Description of intervention | |
| Rationale for paper/review | |
| What creates meaning that sustains GPs engagement with, and involvement in, work? | In this study, job satisfaction is measured using a scale of six statements about satisfaction with different aspects of GPs work. These statements are about whether:<br>- they think their work is useful<br>- it is interesting<br>- they feel like their administrative tasks take up too much time<br>- their work causes too much stress<br>- they regard GPs to be respected |

| | |
|---|---|
| | - there is a good balance between their work and the rewards they receive for it<br><br>GPs who carry out more technical procedures and who are more involved in preventive care and health promotion are more satisfied with their job.<br>GPs who take more weeks of vacation per year are more satisfied.<br>GPs who receive more feedback from their colleagues have higher job satisfaction.<br>GPs with more satisfied patients are more satisfied with their job as well. |
| What solutions/recommendations are proposed? | |
| From which stakeholders' perspective? (General practitioners, patients, other HCPs) | GPs |
| How is work-related joy constructed? (Diversity of ways individuals may think about joy) | |
| What prevents/removes work-related joy? | GPs who work more hours are less satisfied<br><br>In the area of *comfort*, a surprising finding is that there is no significant relationship between administrative work and GP job satisfaction, especially since this was found to be one of the most important factors that decreases job satisfaction [72]. |
| Organisational characteristics (small, large) | |
| Managing ill-defined illnesses and chronic complex multimorbidity | |
| Human connection with patients | GPs with more satisfied patients are more satisfied with their job as well. |
| Human connection with peers | GPs who receive more feedback from their colleagues have higher job satisfaction. |
| Human connection across other organisations (e.g., secondary care) | |

| | |
|---|---|
| Learning systems (e.g., competence, capability, mastery, socially situated) | |
| Additional useful information | of all the country variables that were tested, only GDP per capita is strongly related to GP job satisfaction.<br><br>the control variable age was found to be positively related to job satisfaction; older GPs are more satisfied. This may be partly explained by the 'healthy worker effect'; older GPs who are satisfied may tend to stay in work longer, while less satisfied GPs retire earlier and are therefore not included in this study.<br><br>Another secondary analysis of the QUALICOPC data showed a broader range of tasks, and higher workload for rural GPs, but no difference in job satisfaction between rural and urban GPs |
| Summary of findings | In conclusion, a high level of job satisfaction of GPs is important because it may help to maintain the current workforce and to make general practice a more appealing career option for new doctors [77]. The finding that a broad range of services is related to higher job satisfaction suggests that GP job satisfaction could be increased by stimulating GPs to spend more time on, e.g. technical procedures and health promotion. The negative relationship between work hours and job satisfaction is important in view of the work–life balance which has become increasingly important for GPs, especially for younger and female GPs [79], which suggests that this issue will be even more important in the future. Feedback from colleagues and patients could be strengthened by policies at practice level as well as regional or national level. However, the evidence base for policy development is still rather small. Finally, the strong relationship between GDP per capita and GP job satisfaction should be examined further. Given the clustering of GP job satisfaction within countries, national interventions aimed at increasing job satisfaction can be very useful, but these can only be developed if we have more insight in (health)policy-amenable influences. Therefore, further research into differences in job satisfaction between countries is recommended. |
| Strengths, limitations, and future research directions | Our study has a number of strengths. The first strength is the theoretical approach to GP job satisfaction. The SPF theory is an attractive framework, because it emphasises agency in the form of goal-oriented behaviour. Many of the variables in the hypotheses we tested refer to modifiable variables, either by GPs themselves or at a collective level by professional organisations and policymakers [76]. This means that targeted interventions could result in higher job satisfaction. A further advantage of our approach is that other variables that can be subsumed under the theoretical categories, such as behavioural confirmation where we found, e.g. associations with peer feedback, might also increase job satisfaction. A second strength is the use of a large international data set, although the fact that this study was not designed to study job satisfaction at the same time is a weakness. We used a state-of-the-art statistical approach by performing random effects multilevel analysis, which takes the nested structure of the data into account [65].<br><br>There are also weaknesses. It should be noted that cross-sectional nature of the data only show how variables are related to each other, and cannot indicate causal relationships. This means that it is not possible to say whether statistically significant variables would cause a change in GP job satisfaction. This also means that certain variables, such as patient satisfaction or performing certain services, could be dependent on GP job satisfaction instead of the other way around, or that they influence each other. Secondly, it should be noted that this study could be subject to sample selection bias, as it could be that GPs who are more satisfied are also more willing to participate in a large survey. However, this does not necessarily have to be the case since job satisfaction was not the main aim of the survey and it was only mentioned in |

| | |
|---|---|
| | one out of the 60 questions. The questionnaire of the QUALICOPC study did not include psychological variables, such as coping abilities; hence, there is risk of misspecification in the sense that some of the relations we found, e.g. the relationship with working hours, could actually relate to the psychological abilities of GPs to cope with their workload.<br><br>When designing new, international comparative research specifically into job satisfaction of GPs, several points come up from this secondary analysis. First of all, the measurement of job satisfaction could be improved. The six items to measure job satisfaction that were used in the QUALICOPC study, were taken from a previous study. However, in a new and specific study on GP job satisfaction, a comparison should be made with other existing instruments. Moreover, the measurement of job satisfaction could linked to the SPF theory as has been done for general well-being [78]. A survey among GPs on job satisfaction should also include more attention to psychological determinants of job satisfaction (as mentioned in the previous paragraph). Finally, it would be important both from a scientific and a policy standpoint, to have a wider exploration of the international differences in job satisfaction. This could be done by also studying another population than GPs to find out to what extent international differences in job satisfaction are specific for GPs or more general for large groups of the working population. |

| | |
|---|---|
| Paper Number | 140 |
| Author | Swaithes et al |
| Year | 2020 |
| Title | Experience of implementing and delivering group consultations in UK general practice: a qualitative study |
| Journal | BJGP |
| Region (England, Scotland, Wales, and Northern Ireland) | UK |
| Study methodology | Qualitative telephone interviews of twenty multidisciplinary professionals (eight GPs, eight PNs, one nurse associate, one pharmacist, one deputy practice manager, and one healthcare assistant) |
| Type of document | Primary research |
| Topic or focus of paper | Implementation and delivery of group consultations |
| Research objectives and focus | explore the experience of implementing and delivering group consultations in primary care in the context of the NHS |
| Theoretical framework/theories/ underpinnings | Data analysis was based on the principles of the Framework Method, using Normalisation Process Theory (NPT) as an underpinning theoretical approach.<br><br>An inductive thematic analysis approach was taken before applying theory. |
| Description of intervention | Semi-structured interviews via telephone. A multidisciplinary stakeholder advisory group was conducted with general practice staff, primary care academics, and public contributors. |

| | |
|---|---|
| Rationale for paper/review | To understand how to optimise implementation, it is therefore necessary to engage with stakeholders to identify and address potential implementation challenges [of group consultations] from a staff perspective. |
| What creates meaning that sustains GPs engagement with, and involvement in, work? | Personal interest in a condition being managed by the approach [of group consultations] or in the approach itself<br>Appealing to clinicians' sentiment of care<br>Confidence<br>More time<br>Consultations driven and led by patients<br>Group dynamic<br>Team cohesiveness and enhanced multidisciplinary working<br>Enhanced knowledge and capability<br>Providing more patient-centred care |
| What solutions/recommendations are proposed? | Group consultations |
| From which stakeholders' perspective? (General practitioners, patients, other HCPs) | GPs, PNs, nurse associate, pharmacist, deputy practice manager, HCA |
| How is work-related joy constructed? (Diversity of ways individuals may think about joy) | Reducing workload burden<br>Increasing capacity (freeing up appointment time)<br>Good working structures and relationships<br>Regular meetings<br><br>Support from 'facilitators' (those who do the bulk of work and administrative tasks) and 'champions' (those who lead and influence the development of group consultations)<br>Once challenges were overcome, group consultations were reported to mitigate the problems of individual consultations, because of both actual and perceived extra time, resulting in staff feeling more relaxed and to *'bring the joy back into consultations'*<br><br>Clarity regarding roles and responsibilities |
| What prevents/removes work-related joy? | Burn-out<br>Increasing demands of services<br>Individual beliefs about their capability or capacity - *'You have huge imposter syndrome, I can't do this, I'm not trained to do this.'*<br><br>individual consultations were described as monotonous, isolating, and better suited to acute care, with pressure to get patients *'out of the door'* |

| | |
|---|---|
| | Tick-box mentality<br>Significant administrative and organisational burden<br>Vacancies in the team<br>Practice pressures<br>Insufficient resources for clinical and professional support<br>Issues with payment - participants highlighted a lack of flexibility in terms of CCG support and how tariff and payment systems were not structured to recognise group consultation activity for payment. |
| Organisational characteristics (small, large) | |
| Managing ill-defined illnesses and chronic complex multimorbidity | |
| Human connection with patients | group consultations were perceived as being driven and led by patients. The group dynamic was suggested as the most powerful benefit of group consultations. |
| Human connection with peers | Champions [those who voluntarily adopted leadership] were often supported by 'professional launch pads' (incentives from a range of funding streams and schemes such as fellowships or financial initiatives) to undertake the work associated with initiating group consultations.<br><br>Many spoke of the difficulties in influencing colleagues and key decision makers to 'buy-in' to the approach.<br><br>Engagement ranged from 'passive support', and 'blockers'.<br><br>*'Get the decision makers involved because if they're not involved, they will stop you, you know, passively, if not actively,* because all they *need to do is push it to the curb and people's enthusiasm goes. Because particularly clinicians, you know, life's busy and unless you've got some space to be able to keep pushing, keep pushing, it's difficult to keep things going.'* |
| Human connection across other organisations (e.g., secondary care) | |
| Learning systems (e.g., competence, capability, mastery, socially situated) | Group consultations were described to improve team cohesiveness, enhance knowledge and capability of junior clinicians, and create an opportunity for staff to learn clinical knowledge and skills, facilitation, and presentation skills |
| Additional useful information | |
| Summary of findings | Findings illustrate that clinicians enjoyed group consultations because of enhanced multidisciplinary working and the ability to provide more patient-centred care. |

| | |
|---|---|
| | Factors required to initiate (cognitive participation) and sustain group consultations include clear leadership (from a group consultation champion), an implementation plan at the outset, clarity regarding roles and responsibilities, and wider support from policy mechanisms (tariffs, for example) and CCGs. |
| Strengths, limitations, and future research directions | Strengths:<br>- Multidisciplary mix of general practice staff<br>- Wide geographical spread<br>- Robust approach to data analysis<br>- Involvement of a multidisciplinary stakeholder group<br>- Use of NPT to capture the complexities involved in implementation<br><br>Limitations:<br>- Participants recruited via one group consultation training provider (those trained by other providers might have different experiences)<br>- Sample is predominantly female<br>- Sample may have introduce bias ie encouraging participation from those with more positive views towards group consultaitons<br>- Range of interview duration<br><br>Further research and practice-based evaluation is needed to capture and better understand process outcomes. |

| | |
|---|---|
| Paper Number | 141 |
| Author | Turner et al |
| Year | 2023 |
| Title | Role and response of primary healthcare services in community end-of-life care during COVID-19: Qualitative study and recommendations for primary palliative care delivery |
| Journal | Palliative Medicine |
| Region (England, Scotland, Wales, and Northern Ireland) | UK |
| Study methodology | Descriptive qualitiative study with semi-structured interviews |

| | |
|---|---|
| Type of document | Primary research |
| Topic or focus of paper | the delivery of community end of life care during the COVID-19 pandemic |
| Research objectives and focus | (1) to gather detailed insights from the perspectives of general practitioners and community nurses on factors that enabled the delivery of community end-of-life care during the COVID-19 pandemic, and<br>(2) to develop recommendations to improve primary care delivery of end-of-life care, including during pandemics and other times of increased need. |
| Theoretical framework/theories/ underpinnings | Behaviour change theory[19] provides a framework through which to consider how interventions described as beneficial worked in practice during the pandemic in order to develop recommendations for future practice and policy. The Behaviour Change Wheel is underpinned by behaviour change theory, with capability, opportunity and motivation interacting to generate behaviour (the COM-B system). |
| Description of intervention | Semi-structure interciews with 25 primary healthcare professionals on roles, changes to the delivery of community end-of-life care during the pandemic, opportunities for service innovation, and their concerns for the future delivery of end-of-life care during times of increased need. |
| Rationale for paper/review | there was scarce evidence to inform primary care policy and practice in relation to the delivery and sustainability of community end-of-life care during the COVID-19 pandemic.<br><br>Increased understanding about what works in the delivery of community end-of-life care at times of increased demand is vital, including from the perspective of primary care. |
| What creates meaning that sustains GPs engagement with, and involvement in, work? | Partnership working and group achievement<br>Multidisciplinary online staff education, training and peer support<br>Care planning<br>Physical presence and face-to-face contact |
| What solutions/recommendations are proposed? | - Maintain multi-professional, cross-boundary clinical team meetings, with virtual meetings as an option.<br>- Continue to develop and deliver online collaborative training and education.<br>- Resource time and capability in primary care for effective communication about advance and end-of-life care planning with patients<br>- Maintain effective systems to support care planning, such as electronic prescribing.<br>Provide clear, consistent and unified guidance for end-of-life care in primary care.<br>- Recognise the importance of the physical presence of all members of the primary healthcare team in policy and practice guidance.<br>- Enhance and support effective primary care system leadership at local, regional and national levels. |
| From which stakeholders' perspective? (General | GPs and community nurses |

| | |
|---|---|
| practitioners, patients, other HCPs) | |
| How is work-related joy constructed? (Diversity of ways individuals may think about joy) | Being able to communicate with colleagues online enhancing partnershuo working<br>Increased skill and confidence<br>Increased time and resource allocation to care planning<br>Presence of general practitioners in the home |
| What prevents/removes work-related joy? | Lacking meaningful conversation – important conversations taking place in a hurry, during a crisis and often remotely were a source of practicioner distress<br>Loss of human connection with peers through technology<br>Lacking or inadequate technology eg electronic patient record sharing<br>Filling gaps in service provision where there was a risk to personal health |
| Organisational characteristics (small, large) | |
| Managing ill-defined illnesses and chronic complex multimorbidity | an increase in numbers of community patients needing end-of-life care during COVID-19, and greater complexity of individual care needs.<br><br>Earlier and more consistent conversations were associated with better advance and end-of-life care planning. |
| Human connection with patients | Many of these participants expressed a strong sense of moral and emotional distress resulting from guidance to reduce face-to-face contact with patients dying at home. |
| Human connection with peers | Virtually all the nurses interviewed expressed concerns that some general practitioners were not providing home visits. Community nursing teams therefore provided most hands-on end-of-life care. Where this happened, community nurses described feeling abandoned and unsupported |
| Human connection across other organisations (e.g., secondary care) | Easier to get a hold of specialists in secondary care and district teams as they can dial into a virtual meeting<br>At a healthcare organisation level, the importance of the presence of primary care professionals '*at the strategic side of things'* to contribute to service planning, policy and practice guidance for end-of-life care was specifically highlighted |
| Learning systems (e.g., competence, capability, mastery, socially situated) | |
| Additional useful information | |
| Summary of findings | The study findings provide insights into factors that affected delivery of community end-of-life care during the COVID-19 pandemic, a time of greatly increased demand. Partnership working was valued, with the shift to online meetings facilitating support and role-modelling from specialist palliative care. However, the physical presence of general practitioners was key to the effective partnership delivery of end-of-life care in the community. |
| Strengths, limitations, and future research directions | - Focus on the element of community end-of-life care delivery during a pandemic<br>- Detailed, in-depth insight into the role of primary HCPs delivering end-of-life care in the COVID-19 pandemic |

| | |
|---|---|
| | - Limitations include self-selecting participants whose views might not be representative of the wider workforce, and who are exclusively GPs and community nurses – other primary care professions have not been represented. There is also no patient and family carer perspective collected.<br>- Further research should include a patient-centred approach to increase the understanding of the experiences and nature of services provided. |

| | |
|---|---|
| Paper Number | 142 |
| Author | Maskrey et al. |
| Year | 2018 |
| Title | Releasing GP capacity with pharmacy prescribing support and New Ways of Working: a prospective observational cohort study |
| Journal | BJGP |
| Region (England, Scotland, Wales, and Northern Ireland) | Scotland |
| Study methodology | Audit and survey |
| Type of document | |
| Topic or focus of paper | |
| Research objectives and focus | |
| Theoretical framework/theories/ underpinnings | |
| Description of intervention | Data collection: GPs recorded the time they spent dealing with special requests, immediate discharges, outpatient requests, and other prescribing issues for 2 weeks prior to the study and for two equivalent periods during the study.<br>Intervention: Specialist clinical pharmacists performed these key prescribing activities to release GP time and Read coded their activities.<br>Results: GP and practice staff were surveyed to assess their expectations at baseline and their experiences during the final data-collection period. Prescribing support staff were also surveyed during the study period. (along with recorded data) |
| Rationale for paper/review | it is unknown what impact prescribing support teams may have on freeing up GP capacity and time for clinical activities.<br><br>General practice and primary care are experiencing difficulties coping with demand. There are medical and political expectations that specialist clinical pharmacists and non-medical prescribers can be employed to release GP time and capacity, but information is lacking |

| | |
|---|---|
| | regarding the effect of these health professionals on primary care. This study demonstrates that they can free up direct GP time and GP capacity, and that the effects on staff morale and patient safety can be positive. |
| What creates meaning that sustains GPs engagement with, and involvement in, work? | better work–life balance to allow GPs to fulfil personal responsibilities and need |
| What solutions/recommendations are proposed? | |
| From which stakeholders' perspective? (General practitioners, patients, other HCPs) | GPs and clinical pharmacists |
| How is work-related joy constructed? (Diversity of ways individuals may think about joy) | Some practices reported increasing some consultations to 15 minutes and one GP indicated having had time to take a tea break. [when surveyed during intervention] |
| What prevents/removes work-related joy? | increases in workload and pressures that compromise patient-centred care, diminishing GPs' job satisfaction and health |
| Organisational characteristics (small, large) | |
| Managing ill-defined illnesses and chronic complex multimorbidity | |
| Human connection with patients | All of this [aging population with greater multimorbidity, changes in UK governmental policy to shift more patients to primary care, increases in workload and pressure affecting person-centred care, diminishing GP satisfaction and health] is driving general practice staff away from **the NHS**, away from **their professions**, and away from **patients** |
| Human connection with peers | The survey conducted in spring 2017[during intervention utilising clinical pharmacists] when the initiative was under way, indicated:<br>- an increase in GP capacity<br>- reduced stress<br>- an improvement in morale, practice prescribing processes, and safety |
| Human connection across other organisations (e.g., secondary care) | |

| | |
|---|---|
| Learning systems (e.g., competence, capability, mastery, socially situated) | |
| Additional useful information | The majority (87%) of pre-2016 and all new staff [involved in pharmacist support work/prescribing work] indicated that the changes were positive, enabling them to use more of their skills and expertise, and that they were comfortable and confident carrying out the activities. Most reported workload as feeling sustainable, although some staff were concerned about pressures of GP/practice expectations, staff shortages, and training new team members |
| Summary of findings | The inclusion of additional specialist clinical pharmacists to perform key prescribing activities released an average of 5 hours' direct GP time per practice per week; this is equivalent to one GP practice session per week. The additional pharmacy resource was well received and appreciated by general practice staff. However, as well as freeing up GP capacity, practices and practitioners identified positive effects on patient safety and staff morale, along with reductions in stress during the study period. |
| Strengths, limitations, and future research directions | Strengths:<br>- addressing a real-world challenge in routine practice, freeing up system capacity and GP time, as well as demonstrating that this can be achieved.<br>- this is the first prospective study to demonstrate the impact of specialist clinical pharmacists and prescribing support teams directly freeing up system capacity and GP clinical time in routine practice.<br>- a combination of data — subjective self-reporting by GPs, Read code activity, and survey data<br>- The relative simplicity of GPs' self-recording and reporting of time taken on prescribing tasks was low impact and non-resource intensive<br>- a 'bottom-up', flexible approach was encouraged.<br><br>Limitations: |

| | |
|---|---|
| Paper Number | 143 |
| Author | McCallum & MacDonald |
| Year | 2021 |
| Title | Exploring GP work in areas of high socioeconomic deprivation: a secondary analysis |
| Journal | BJGP Open |
| Region (England, Scotland, Wales, and Northern Ireland) | Scotland |

| | |
|---|---|
| Study methodology | A qualitative secondary supra analysis in-depth interviews (*n* = 10) exploring GP training in areas of high socioeconomic deprivation.<br><br>The study uses Corbin and Strauss's framework on managing chronic illness as an analytical lens.<br>The framework describes that managing chronic illness required three types or 'lines' of work:<br>- illness work (work required to manage illness)<br>- everyday work (organisation and coordination of various types of work necessary to operationalise any plans of action)<br>- and biographical work (the work required in defining and making an identity).<br><br>These three lines of work require to be balanced, defined as 'relative equilibrium'. |
| Type of document | Primary research |
| Topic or focus of paper | The impact of socioeconomic deprivation, complex social needs and multimorbidity on the type of work completed managed by GPs and how workload is managed |
| Research objectives and focus | To explore GP work in areas of high socioeconomic deprivation and the strategies GPs employ, using Corbin and Strauss's framework on managing chronic illness as an analytical lens. |
| Theoretical framework/theories/ underpinnings | Corbin and Strauss's framework on managing chronic illness |
| Description of intervention | Qualitative interviews of GPs working in areas of high levels of socioeconomic deprivation |
| Rationale for paper/review | There is a GP workforce crisis in the UK,**14** with an urgent need to reduce GP burnout and stress.<br><br>Understanding if, and how, socioeconomic deprivation impacts GP work, and what strategies GPs use to manage that work, is critical to ensure primary care services are effective, well equipped, resourced, and supported.<br><br>GPs carry out four specific types of work (everyday, illness, biographical, and emotional). Socioeconomic deprivation influences this work, which is often not recognised by the wider health system, and is under-resourced. Current workforce strategies focus only on illness work. The present article suggests that supporting all types of work may be beneficial, particularly for practices working in the context of high socioeconomic deprivation. |
| What creates meaning that sustains GPs engagement with, and involvement in, work? | Managing social complexity<br>Attendant patient advocacy<br>Alignment with personal values about equality and justice<br><br>The impact of wider values, and the alignment of identity and purpose in GPs' work environments (particularly in the context of reducing inequality and making a difference), provided ongoing motivation to continue their work.<br><br>While recognising that what could be achieved was limited by wider factors, the potential to make a difference in people's lives, provided powerful motivation and resilience. |

| | |
|---|---|
| | Similarly, being involved in additional professional activities, which allowed them time to reflect and remind themselves of their early motivation for opting to work in general practice was important. |
| What solutions/recommendations are proposed? | |
| From which stakeholders' perspective? (General practitioners, patients, other HCPs) | GPs |
| How is work-related joy constructed? (Diversity of ways individuals may think about joy) | All the GPs were involved in external activities (such as undergraduate or postgraduate training, appraising, local medical committees, or part of the Scottish GPs at the Deep End steering group). These roles fostered resilience and reduced burn out. They offered a break from clinical work and provided professional tasks that could be completed in contrast to the uncertainty and unending demand from clinical work.<br><br>the need for mutual sustaining was uniformly recognised as vital. GPs spoke of how a supportive flexible team, with a shared ethos, was essential in managing workloads |
| What prevents/removes work-related joy? | Workload increased:<br>- Health system configurations, with an emphasis on self-management and self-referral, failed to consider the everyday realities of patients' lives, further disadvantaging an already disadvantaged group. The additional GP work required to fill this gap was absorbed into routine everyday work<br><br>Unsustainable workload<br>Fatalistic attitude about whether this could change in future due to GP shortages<br>Biographical work and additional routine work<br>Being emotionally impacted by being unable to offer help in the difficult situations patients experience |
| Organisational characteristics (small, large) | |
| Managing ill-defined illnesses and chronic complex multimorbidity | All GPs interviewed identified key characteristics of the communities they worked in that impacted their work: higher prevalence of early multimorbidity; patients managing the impact of poverty, alcohol, and drug addiction; vulnerable families; migrant populations; and patients for whom life was chaotic.<br><br>Managing this 'multimorbidity plus' was perceived to result in work unlike that undertaken by colleagues who worked in other less deprived areas |

| | |
|---|---|
| Human connection with patients | Constantly asserting professional identity, including refusing requests (for example, letters for housing applications), was an ongoing source of daily biographical work.<br>GPs were having to "fill the gaps" when patients were unable to self-refer |
| Human connection with peers | GPs spoke of how a supportive flexible team, with a shared ethos, was essential in managing workloads |
| Human connection across other organisations (e.g., secondary care) | Routine work was increased, for example, by building links with the third sector in the context of helping patients who struggle to use the self-referral system in context of low literacy |
| Learning systems (e.g., competence, capability, mastery, socially situated) | All the GPs were involved in external activities (such as undergraduate or postgraduate training, appraising, local medical committees, or part of the Scottish GPs at the Deep End steering group). |
| Additional useful information | |
| Summary of findings | The analysis suggests that, in the context of high socioeconomic deprivation, general practice comprises several lines of 'work', stretching far beyond the biomedical management of illness.<br><br>While current strategies for managing workload primarily focus only on reducing illness work, the research suggests that strategies that support and resource the additional and varied nature of work are required.<br><br>While the context of high socioeconomic deprivation brought unique workload issues, much of the work described by GPs was thought to align well with GPs in other communities. |
| Strengths, limitations, and future research directions | - participating GPs, despite coming from a range of practices (in terms of location and size), were consistent in their description of work and the challenges they encountered.<br>- the findings regarding GP work reflect socioeconomic deprivation, not just local geography.<br>- using Corbin and Strauss's well-established *Managing c hronic illness at home : three lines of work* **17** as an analytical lens provided a framework to interpret the findings, which increases and extends its applicability in settings outside of patient experience, and provides a basis for further research exploring health professionals' work in other contexts.<br><br>Limitations:<br>- small sample size<br>- recognition that GPs working in other areas have different challenges<br>- interviews are 4 years old – don't take into account the impact of the pandemic<br>- no information on GP workload eg number of patients<br><br>Future research directions: |

| | |
|---|---|
| | - Further research with GPs in other contexts is required to understand GP workload and wider applicability of this analytical lens. Wider sampling to understand GP work in different contexts (for example, affluent and rural populations) is needed, as it is expected there are different workload implications in different contexts, which may require different resourcing.<br>- Repeating this work in the current context (the ongoing pandemic) would allow deeper understanding of the impact that COVID-19 has had on GP workload. |

| | |
|---|---|
| Paper Number | 144 |
| Author | Smith, Goldacre and Lambert |
| Year | 2017 |
| Title | Adverse effects on health and wellbeing of working as a doctor: views of the UK medical graduates of 1974 and 1977 surveyed in 2014 |
| Journal | JRSM |
| Region (England, Scotland, Wales, and Northern Ireland) | UK |
| Study methodology | Questionnaires |
| Type of document | Primary research |
| Topic or focus of paper | Wellbeing of doctors |
| Research objectives and focus | To report on any adverse effects on health and wellbeing of working as a doctor, as described by senior doctors who graduated in 1974 and 1977 |
| Theoretical framework/theories/ underpinnings | |
| Description of intervention | Questionnaires sent in 2014 to all medical graduates of 1974 and 1977, with a mix of "Yes/No" options and descriptive text questions. |
| Rationale for paper/review | Much of the focus has been upon medical students and junior doctors. However, studies which have included more senior doctors have also found rising levels of stress and ill health. |
| What creates meaning that sustains GPs engagement with, and involvement in, work? | Support from those around them including employers and colleagues |
| What solutions/recommendations are proposed? | |

| | |
|---|---|
| From which stakeholders' perspective? (General practitioners, patients, other HCPs) | Doctors graduated in 1974 and 1977 |
| How is work-related joy constructed? (Diversity of ways individuals may think about joy) | |
| What prevents/removes work-related joy? | Stress and workload<br>Lost time with family<br>Burnout<br>Depression<br>Physical illness caused or aggravated by stress<br>Mental illness due to stress at work<br>Job-related/procedural illness<br>Litigation<br>Unrealistic expectations and demands<br>Political pressure from policy changes and increased bureaucracy<br>Lack of support from employer or colleagues<br>Bullying<br>Amount of activity not reducing with growing older and reducing stamina |
| Organisational characteristics (small, large) | |
| Managing ill-defined illnesses and chronic complex multimorbidity | |
| Human connection with patients | |
| Human connection with peers | The doctors were presented with the statement 'The NHS of today is a good employer when doctors become ill themselves'. Twenty-eight per cent of doctors agreed or strongly agreed with the statement, 29% answered 'neither', and 43% disagreed or strongly disagreed.<br><br>More GPs (49%) than hospital doctors (37%) disagreed with this statement. |
| Human connection across other organisations (e.g., secondary care) | |

| | |
|---|---|
| Learning systems (e.g., competence, capability, mastery, socially situated) | |
| Additional useful information | Several doctors, while agreeing in the previous question that, ‘yes’, they felt that working as a doctor had had adverse effects on their health or wellbeing, went on to play down the negative effects they had experienced |
| Summary of findings | Almost half of senior doctors said that working as a doctor had had adverse effects on their own health or wellbeing. More GPs than hospital doctors believed this to be the case. The most frequently mentioned adverse effect in text comments was ‘Stress/work–life balance/workload’ followed by illness.<br>Almost half of senior doctors did not agree that the NHS is a good employer when doctors become ill themselves. More women than men, and more GPs than hospital doctors disagreed that the NHS is a good employer when doctors become ill themselves. |
| Strengths, limitations, and future research directions | A response rate of 84.6% is very high for a self-completed survey.<br>However, some non-response bias may have been present, though we have no evidence to suggest that responders are different from non-responders in characteristics relevant to this study.<br>We present both numeric and qualitative summaries of the textual data. A purely numeric approach would have been inappropriate where the representativeness of the sample is not known. |

| | |
|---|---|
| Paper Number | 145 |
| Author | Kidd et al |
| Year | 2023 |
| Title | Challenges in implementing GP clusters in Scotland: A comparison of the views of senior primary care stakeholders in 2016 and 2021 |
| Journal | BJGP Open |
| Region (England, Scotland, Wales, and Northern Ireland) | Scotland |
| Study methodology | Qualitative analysis of semi-structured interviews with 12 senior primary care national stakeholders in 2016 (*n* = 6) and 2021 (*n* = 6). |
| Type of document | Primary research |
| Topic or focus of paper | GP cluster working in Scotland |
| Research objectives and focus | To compare predicted challenges of cluster implementation in 2016 with reported challenges in 2021. |

| | |
|---|---|
| Theoretical framework/theories/ underpinnings | Clusters are geographical groupings of 5–8 GP practices, each represented by a nominated practice quality lead (PQL), who works collaboratively to engage in peer-led quality improvement activity relevant to their local population.<br><br>The aspirations and intended functions for cluster working are subdivided into intrinsic (improving quality of care across practices within the cluster) and extrinsic (engaging in the broader integration of health and social care locally) roles. |
| Description of intervention | |
| Rationale for paper/review | While the high-level expectations for GP clusters have been clearly defined, the progress of clusters towards achieving these intended impacts over the past 6 years is less clear. The limited available literature suggests that GP clusters may not have progressed at a pace that was envisaged at the time of their conception.<br><br>As such, this longitudinal qualitative study aimed to explore and compare the expected progress of GP clusters and to gain insight into key challenges and factors that have facilitated or hindered effective implementation to date. |
| What creates meaning that sustains GPs engagement with, and involvement in, work? | |
| What solutions/recommendations are proposed? | |
| From which stakeholders' perspective? (General practitioners, patients, other HCPs) | Senior stakeholders from main national organisations involved in primary care. Organisations and types of stakeholders are not named to maintain the confidentiality/prevent identification of stakeholders. |
| How is work-related joy constructed? (Diversity of ways individuals may think about joy) | Adequate support – eg data analytic support, administrative support, training in quality improvement, and project support |
| What prevents/removes work-related joy? | Deficits in motivation for engagement and understanding of the philosophy of cluster working among GPs working within the clusters<br>Lack of support<br>Problems with access to and quality of data - lack of national systems or 'dashboards' for data extraction, which would provide the capacity to engage meaningfully with quality assurance, improvement, and planning based on robust assessment of local population health needs.<br>Limited resources and workforce shortages<br>Micromanagement/national oversight and tension with local autonomy<br>Inadequate funding of protected time<br>Unrealistic expectations from the Scottish Government and wider NHS<br>Burnout |

| | |
|---|---|
| | Work and time commitment to create meaningful engagement in clusters was substantial<br>The impact of COVID-19 pandemic |
| Organisational characteristics (small, large) | |
| Managing ill-defined illnesses and chronic complex multimorbidity | |
| Human connection with patients | |
| Human connection with peers | |
| Human connection across other organisations (e.g., secondary care) | Unrealistic expectations from the Scottish Government and wider NHS<br>Excess demands placed on clusters by secondary care and other organisations |
| Learning systems (e.g., competence, capability, mastery, socially situated) | |

| | |
|---|---|
| Additional useful information | 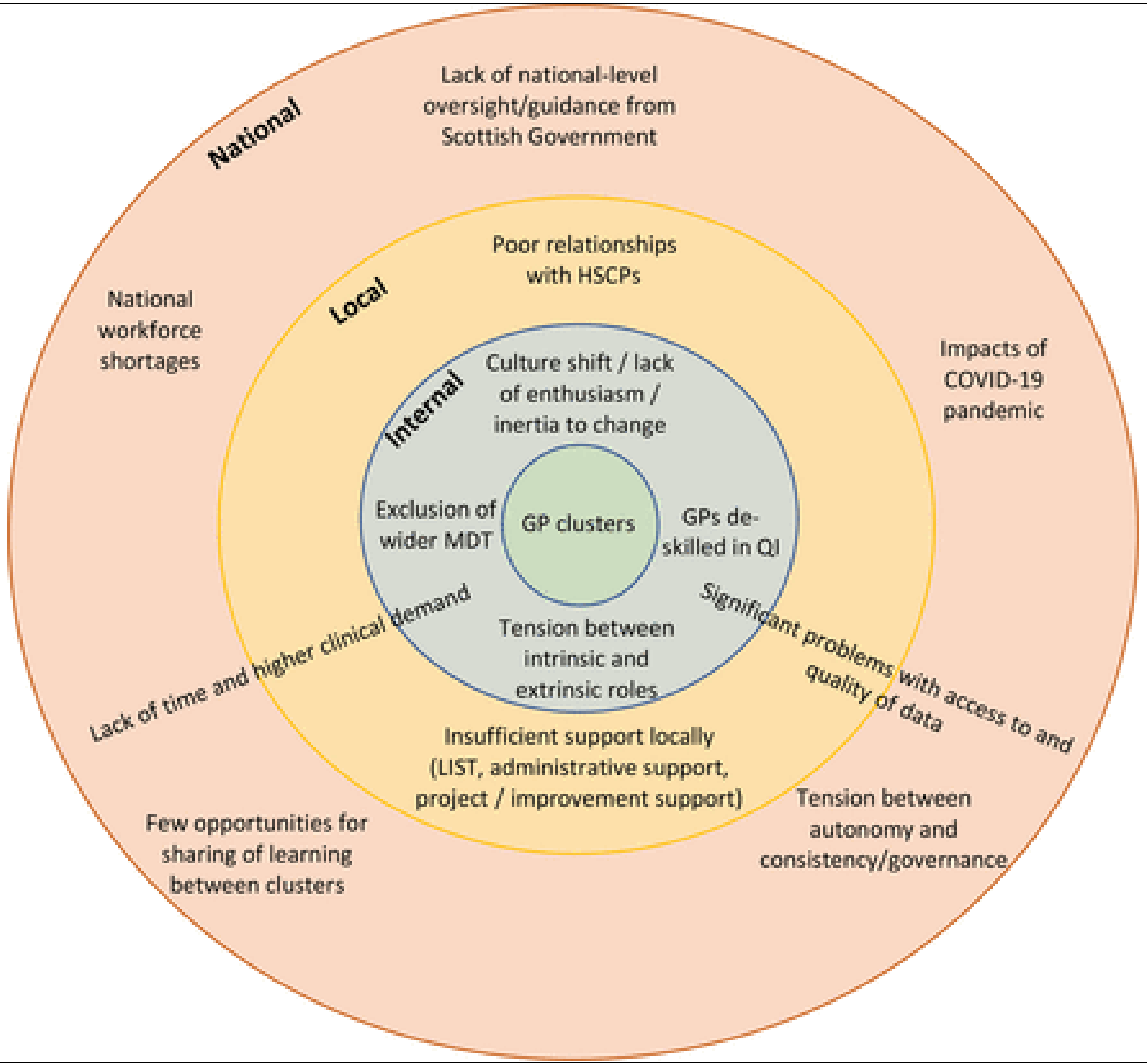 |
| Summary of findings | Progress of clusters in 2021 was perceived as suboptimal, and was reported to vary significantly across the country, reflecting differences in local infrastructure. Practical facilitation (data, administrative support, training, project improvement support, and funded time) and strategic guidance from the Scottish Government was felt to be lacking. GP engagement with clusters was felt to be hindered by the significant time |

| | |
|---|---|
| | and workforce pressures facing primary care. The end result could be viewed as a vicious cycle of non-progression whereby clusters become frustrated and burnout in the face of multiple system barriers impeding their work, leading to disengagement and further stagnation. Considerable investment in addressing these limiting factors will thus be required if clusters are to begin to exert a more meaningful impact on the NHS improvement landscape in Scotland |
| Strengths, limitations, and future research directions | - Use of secondary analysis enabled an in-depth analysis of themes relating specifically to GP clusters<br>- Secondary analysis may be lacking in methodological rigour<br>- Measures taken to ensure the credibility of the current analysis included an emphasis on thorough data familiarisation and use of introspection as well as intersubjective reflection to ensure reflexivity in interpretation.<br>- Different stakeholders were interviewed from all major national organisations involved in primary care which increased the breadth of stakeholder perspectives but also increased influence of individual differences between groups<br>- Only senior stakeholder views were reported<br>- Limitations include small sample size and exclusion of cluster members as participants both of which may potentially serve as sources of sampling bias within the data. |

| | |
|---|---|
| Paper Number | 146 |
| Author | Parkinson, Smith and Sidhu |
| Year | 2021 |
| Title | Early development of primary care networks in the NHS in England: A qualitative mixed-methods evaluation |
| Journal | BMJ Open |
| Region (England, Scotland, Wales, and Northern Ireland) | England |
| Study methodology | A qualitative mixed-methods rapid evaluation, informed by a literature review and stakeholder workshop |
| Type of document | Primary research |
| Topic or focus of paper | The core characteristics of PCNs, how they are managed and the role they play in the healthcare system |
| Research objectives and focus | collect early evidence on their implementation and development, including motivations to participate and what enables or inhibits progress. |

| | |
|---|---|
| Theoretical framework/theories/ underpinnings | ?framework analysis |
| Description of intervention | Semistructured interviews with 25 participants using a topic guide, with both clinical and non-clinical staff – mainly with those in leadership or management positions within the PCN. |
| Rationale for paper/review | This paper contributes to this body of literature, and applies existing theoretical work on healthcare networks management to the early experience of PCNs in the NHS in England. |
| What creates meaning that sustains GPs engagement with, and involvement in, work? | |
| What solutions/recommendations are proposed? | |
| From which stakeholders' perspective? (General practitioners, patients, other HCPs) | |
| How is work-related joy constructed? (Diversity of ways individuals may think about joy) | Some of those involved in the leadership and management of PCNs expressed that they have experienced a sense of empowerment in working on something larger than a single practice, and being involved in strategic planning of local primary and community health services over and above single-practice working.<br><br>The level of local engagement with and ownership of PCNs is connected to how they are structured as networks. Where PCNs are felt locally to be part of existing efforts to improve care, population health management and practice management, more individuals within GP practices appear to have bought into the premise of network working. In turn, the network is perceived to be founded on shared goals and interests, and less on top-down mechanisms that contractually bind network members together. |
| What prevents/removes work-related joy? | Frustration was expressed about frequently changing NHS policy that disrupts extant ways of working.<br><br>Contract issues:<br>There was also some irritation expressed by interviewees and observed in meetings about the prescriptive nature of the services required by the DES contract,[28] which further tempers local buy-in to PCNs, particularly where services specified in the DES contract are perceived as not tailored to the needs and preferences of local populations.<br><br>Where PCNs are thought to be another top-down policy change, fewer individuals buy into the idea of PCNs, and there is likely to be increasing frustration about imposed interruptions to existing ways of working at a local level. |

| | |
|---|---|
| Organisational characteristics (small, large) | |
| Managing ill-defined illnesses and chronic complex multimorbidity | |
| Human connection with patients | |
| Human connection with peers | Bottom-up motivation for joining PCNs:<br>- practices typically collaborate to fill gaps in the services that single practices are able to provide, and to facilitate GPs working with community health teams, social services and the voluntary sector to provide extended care that addresses local population health needs.<br>- A clear desire to improve the sustainability of primary care<br>- the potential for financial efficiency and sustainability by sharing back office functions, reducing duplication of administrative tasks, introducing more robust financial management processes and making it easier to recruit and retain new staff |
| Human connection across other organisations (e.g., secondary care) | Top-down motivation for joining PCNs: PCNs are, in effect, perceived to be mandatory given the sizeable financial incentives associated with PCN membership.<br><br>Networks can be built on the shared interests, goals and motivations of members, and also through formalised structures and top-down regulation that require or incentivise membership.<br><br>National policy has prompted the forming of PCNs, but in the absence of national policy incentives, it would remain in GP practices' best interests to still collaborate with one another to provide services, improve management and realise efficiencies based on their mutual interests.<br><br>Variation in local context was also evident in the relationship between PCNs and pre-existing forms of GP collaboration, including GP federations and super-partnerships. At times, PCNs had been able to build on good working relationships established from previous collaborative working between practices and with other parts of the health and social care system and voluntary sector, which helped establish the position of PCNs locally. |
| Learning systems (e.g., competence, capability, mastery, socially situated) | |

| | |
|---|---|
| Additional useful information | On the engagement with PCNs:<br>Where PCNs are perceived as a continuation of existing efforts to improve GP sustainability and local healthcare, there seems to be a high level of enthusiasm and buy-in. However, where PCNs are perceived as a disruption to previous ways of working and a divergence from the goals of pre-existing forms of GP collaboration, there seem to be tensions and frustrations.<br><br>On the role of PCNs:<br>At times, the PCN is a mechanism for collaboration on certain specified tasks, while at other times, it is a primary unit to deliver critical activities such as primary care's pandemic response, and a focal point for interaction between local primary care and other health and social care organisations. |
| Summary of findings | PCNs demonstrate significant potential to swiftly deliver new services to patients, respond to national priorities, bring together primary care providers with common motivations and interests, and improve financial and workforce sustainability in primary care.<br><br>The task ahead for PCNs will be to ensure that they are able to address national priorities that are centrally defined, as well as adapting to fit local health needs. Focusing on the shared goals that motivate GP practices to want to collaborate with one another, and protecting PCNs from over-regulation, will be especially important as PCNs find their place within the wider NHS as it emerges from the pandemic, and as ICSs are implemented.<br><br>In order to enable these organisations to reach their full potential, the core characteristics of these organisations must be considered in deciding how they should be managed, including the motivations driving individual providers to join these organisations and the policy context that led to their development. |
| Strengths, limitations, and future research directions | • As a rapid evaluation, this study responds to current policy-relevant questions about the early development of primary care networks (PCNs), and developments in how primary care is delivered in the National Health Service (NHS) in England.<br>• The qualitative approach provides insights into why general practices participate in PCNs and the experience of implementation.<br>• The mixed-methods approach to this evaluation allows data to be triangulated between sources and ensures that a broad range of perspectives is captured.<br>• The use of a theoretical framework to interpret the findings from this evaluation helps contextualise them within the wider literature, and understand what this evaluation means for other meso-level primary care organisations internationally.<br>• This evaluation provides an insight into the early development and implementation of PCNs, along with information about their initial response to the COVID-19 pandemic. Although the study reflects on how PCNs will continue to develop, for example, in response to new policies in the English NHS, definitive conclusions about the impact of PCNs were outside the scope of this study. |

| | |
|---|---|
| Paper Number | 147 |
| Author | Smith et al |

| | |
|---|---|
| Year | 2022 |
| Title | Early evidence of the development of primary care networks in England: a rapid evaluation study |
| Journal | Health and Social Care Delivery Research |
| Region (England, Scotland, Wales, and Northern Ireland) | England |
| Study methodology | qualitative cross-comparative case study evaluation |
| Type of document | |
| Topic or focus of paper | Implementation of PCNs and how they operate |
| Research objectives and focus | to investigate<br>(1) the contextual background of primary care networks;<br>(2) the rationale for general practices to enter into collaborations;<br>(3) the early learning from establishing primary care networks;<br>(4) barriers to and facilitators of effective collaboration across general practices;<br>(5) the likely future progress of primary care networks in the English NHS, including in the light of COVID-19. |
| Theoretical framework/theories/ underpinnings | |
| Description of intervention | a workshop with academics, policy experts and patient/public representatives; interviews with stakeholders, observations of meetings, a survey and documentary analysis across four case study sites |
| Rationale for paper/review | This study examined the early implementation of primary care networks, especially what has helped or hindered progress, how they operate in relation to pre-existing collaborations and issues for rural networks. |
| What creates meaning that sustains GPs engagement with, and involvement in, work? | practices collaborating with one another can help to distribute workload and pressures between them, and can enable some tasks and procedures to be shared between practices to reduce the burden on any one practice team. Collaboration between practices can also help recruit and retain staff within primary care, including by enabling the organisation to offer better training resources, flexible work arrangements and better benefits to staff.<br><br>It was noted that social prescribers would be able to look at lifestyle factors and could potentially bring about improvements in patients' health, and that roles such as social prescribers, pharmacists and physiotherapists would be able to provide more community services to support primary care.<br>New roles would help to reduce general practitioner workload by creating efficiencies (e.g. by having a single pharmacist to work across all care homes in an area) and by taking away work from general practitioners.<br>Centralised and standardised processes: |

| | |
|---|---|
| | - Shifting of administrative load to centrally based appropriately skilled staff<br>- Mobility of staff across different practices<br>- Shared training resources and support for QOF requirements<br>- Standardised processes across practices in a PCN eg for triaging<br><br>Some of the organisational impact from collaborating across practices was attributed to other forms of collaboration that preceded, and were continuing alongside, PCNs. For example, the super-partnerships at different sites<br><br>The funding coming directly into primary care had given more financial freedom and security to primary care.<br><br>Some evidence pointed to more efficient spending and achieving economies of scale (Int14, Int15), for example through better financial management centrally within PCNs or the prior collaboration (Int2, Int16), or the ability to purchase medications at wholesale prices (Int11). However, some of these positive impacts in terms of financial efficiency may have been attributable to other forms of GP collaboration rather than the introduction of PCNs<br><br>It was noted that it was important for CCGs to be supportive of PCNs and other forms of pre-existing GP collaboration, while also providing them with room to innovate and make autonomous decisions |
| What solutions/recommendations are proposed? | Developing shared goals and objectives emerged as an enabler of progress, and of positive working relationships within and beyond the PCN. |
| From which stakeholders' perspective? (General practitioners, patients, other HCPs) | |
| How is work-related joy constructed? (Diversity of ways individuals may think about joy) | It was also reported that PCNs had generated enthusiasm in terms of general practitioners becoming involved in local strategic planning for health services and resulted in new training and shared resources. Much of the positive impact on staff described by interviewees was attributed to other prior models of collaboration, rather than the PCN.<br>- improved support and resources to staff, including additional training, flexible working arrangements and more generous benefits<br>- more opportunities for specialisation within the field of primary care, which was described as having the potential to enhance career options |

| | |
|---|---|
| | Many of the facilitators of collaborative working focused on strong leadership and management of the PCN and other forms of pre-existing collaboration…. They noted that having a single person (CDs) responsible for resolving issues, facilitating collaboration and building relationships between members was an important aspect of networks |
| What prevents/removes work-related joy? | This evaluation has revealed a tension between the desire by general practitioners and their teams for local autonomy and influence over PCNs and the perceived top-down nature of PCN policy.<br><br>Purpose:<br>A central tension about the purpose of PCNs arose again and again throughout this evaluation. Some respondents had a view that PCN funding should be spent to help improve the financial sustainability of primary care and reduce general practitioner workload, whereas others thought that the new resource should be allocated to developing new primary care services.<br><br>Organisational:<br>Reasons cited for negative views of the PCN included a lack of long-term vision, an excessive burden for practices (and lead general practitioners in particular) in terms of workload, failure to move quickly to implement the PCN, perceived irrelevance of national PCN policy in rural contexts and a perceived lack of leadership from NHSE&I.<br>By obliging practices to form collaborations, the implementation of PCN policy had in effect forced some practices to work together that had fundamental and sometimes long-standing differences in terms of their operational environment, ways of working and values. Bringing together practices in this top-down fashion had caused tensions within some PCNs<br>There have been challenges that have arisen because of the sometimes ‘imposed’ nature of PCN collaborations. Many expressed an anxiety that general practitioners may have about the loss of independence caused by PCNs and other forms of collaboration<br><br>Work content:<br>The administrative and managerial aspect of PCNs was also time-consuming, particularly for PMs and CDs, despite funding being made available for managerial aspects of networks<br>A shift in the proportion of the responsibility for funding and implementing local primary care service development increased workload, as GPs found it difficult to meet the additional services<br><br>Financial:<br>Despite some examples of success, there was evidence that PCNs could have a negative financial impact on practices. Some people expressed a view that, although new funding was coming into primary care, this money had had to be spent on the up-front additional management and administrative costs associated with the new networks owing to an underestimation of the time and resources needed. There had also been reports that funding from NHSE&I had not been sufficient to cover expenses.<br><br>Career:<br>at least one practice was reported in our fieldwork as not having allowed one of their general practitioners to stand for election for the CD role because GP shifts would be left uncovered<br><br>Teamwork: |

| | |
|---|---|
| | clear communication across the network and a good relationship between practices, and particularly between PMs, was important [in PCN development]<br>Establishing shared goals and aims was also cited as important<br>Staff away days and other forums for staff from across the collaboration to talk to one another were reported to create a sense that staff are working as part of a unified team rather than a collection of practices<br><br>some interviewees expressed a view that too much was being expected too soon from PCNs.<br>Some interviewees also commented on what they considered to be a lack of clarity about how PCNs should be set up and what their longer-term vision was to be<br><br>Distance and a lack of reliable public transport could also make it undesirable or challenging for both patients and staff to travel to health centres other than their home practice |
| Organisational characteristics (small, large) | Small - although they are small organisations, PCNs need a significant range of administrative and management capacity and skills, including finance and accounting, human resources, IT, staff engagement and governance support, which were not present in all case study sites in this evaluation. |
| Managing ill-defined illnesses and chronic complex multimorbidity | Collaborating between practices can help fill gaps in service provision when single practices are not able to provide all services, and help provide patients with better co-ordinated care, particularly for patients with complex health and social care needs |
| Human connection with patients | In response to patient demand and access to primary care, PCNs experienced a significant drop in patients requesting appointments with general practitioners, with the majority of consultations carried out by video or telephone, a transition that the majority of patients were pleased with as they received a quicker than usual response from their practice.<br><br>PCNs had the potential to increase the influence of primary care in the local health system, and act as a mechanism to build relationships with community providers and the voluntary sector<br><br>Some commented that, although new roles might help a small number of patients, the roles would not make a difference for most patients who may not need or have contact with the new PCN-funded roles and would not reduce workload for most staff |
| Human connection with peers | As many PCNs had already been working collaboratively to deliver services and improve patient care prior to becoming a network, some felt that the PCN was a step backwards and that these new networks had started to undo the work of previous GP collaborations. Some reported that the PCNs were smaller in scope in terms of what services would be delivered through the network and that the parties involved in the collaboration were more limited in number and type of profession.<br><br>Where the establishment and implementation of PCNs had built on the strengths of previous GP collaborative working arrangements, this was almost always reported to have been a facilitator of network development. |

| | |
|---|---|
| | Several respondents also reported that they felt that the NHS direction of travel in terms of consolidation of independent practices into larger entities would negatively affect the continuity of care that patients experienced in smaller practices and the ability for services to be planned on a smaller scale in a way that aligns with local population needs.<br><br>differences in ethos, operational environments, organisational culture and operating procedures had made it difficult at times to get activities or services under way across practices in the PCN |
| Human connection across other organisations (e.g., secondary care) | In some instances, CCGs have enabled and supported PCNs, providing resource and expertise to help establish inter-practice working, hire new staff and operate contracts.<br><br>In other areas, however, there is evidence of the CCG attempting to hold on to resources or control that have been delegated to PCNs and are therefore not perceived to be operating within the spirit and expectation of PCN policy.<br><br>Although positive relationships with CCGs were viewed as facilitators of collaborative working, they was not present across all the case study sites. It was also reported that the agenda of PCNs and that of CCGs did not necessarily align, as some general practitioners were more likely to want to spend network budgets on supporting practices and decreasing general practitioner workload than the commissioning group, which would probably want something different to be done with the money (Int2, Int19, Int21), such as an increased community-focused approach to population health<br><br>national PCN service specifications were sometimes still considered too demanding and overly prescriptive considering the (pre-COVID-19 pandemic) financial and workforce challenges in primary care |
| Learning systems (e.g., competence, capability, mastery, socially situated) | |
| Additional useful information | A large proportion of interviewees emphasised that, although PCNs had changed the policy landscape in terms of some of the requirements made of primary care, they had not yet led to any substantial changes to modes of working within practices, services offered to most patients or ways of collaborating between practices<br><br>Some interviewees in this evaluation felt that the overall PCN policy had been designed more for urban areas, and had not been sufficiently well thought out in terms of implications for rural practices. Some of these challenges were related to distance and the time taken to drive across the geographical boundary of a rural PCN which could make it difficult for shared staff. Rural areas face particular workforce challenges that have an impact on their ability to hire shared staff across a PCN. For example, rural areas may be less attractive to locums, who get paid on a fee-for-service basis, as they will necessarily cover fewer patients in a less populated area than in a busy urban setting<br>It is likewise potentially more challenging in rural areas to find geographically contiguous practices that share common outlooks and ways of working than in urban areas |

| | |
|---|---|
| Summary of findings | Key lessons focus on increasing the engagement of general practices and wider primary care teams with networks; building leadership and management capacity to support networks in fulfilling their contractual obligations and meeting local health needs; and clarifying how primary care networks will operate in the post-COVID-19 health and social care system. |
| Strengths, limitations, and future research directions | quantitative tracking of PCNs can then form the basis for a qualitative study of what has enabled or hindered progress, and an exploration of what additional support and investment may be needed to enable effective collaboration and achievement of goals.<br><br>Arranging and carrying out interviews proved difficult given the workload associated with implementing networks and wider time pressures. Following the onset of the COVID-19 pandemic, the team was unable to undertake planned face-to-face workshops to explore findings. Given that primary care networks were in their first year of operation at the time of this evaluation, we were cautious in drawing definitive conclusions.<br>Limited information about PCNs' response to the COVID-19 pandemic was collected through this evaluation, although additional data gathering on how networks formed part of the primary care response to COVID-19 would be useful in understanding this more fully.<br><br>In further research, it will also be important to evaluate how PCNs move from an initial stage characterised by relatively high up-front costs to a more established stage in which the resources put into networks might contribute more meaningfully to sustainability and efficiency in primary care. |

| | |
|---|---|
| Paper Number | 148 |
| Author | Stokes-Lampard |
| Year | 2018 |
| Title | GP Forward View: assessment of progress: year 2 |
| Journal | |
| Region (England, Scotland, Wales, and Northern Ireland) | England |
| Study methodology | |
| Type of document | Report |
| Topic or focus of paper | 2-year progress assessment of GP Forward View |
| Research objectives and focus | |
| Theoretical framework/theories/ underpinnings | |

| |
|---|
| Description of intervention |
| Rationale for paper/review |
| What creates meaning that sustains GPs engagement with, and involvement in, work? |
| What solutions/recommendations are proposed? |
| From which stakeholders' perspective? (General practitioners, patients, other HCPs) |
| How is work-related joy constructed? (Diversity of ways individuals may think about joy) |
| What prevents/removes work-related joy? |
| Organisational characteristics (small, large) |
| Managing ill-defined illnesses and chronic complex multimorbidity |
| Human connection with patients |
| Human connection with peers |
| Human connection across other organisations (e.g., secondary care) |
| Learning systems (e.g., competence, capability, mastery, socially situated) |
| Additional useful information |

| |
|---|
| Summary of findings |
| Strengths, limitations, and future research directions |

| | |
|---|---|
| Paper Number | 149 |
| Author | Napier and Clinch |
| Year | 2019 |
| Title | Job strain and retirement decisions in UK general practice |
| Journal | Occupational Medicine |
| Region (England, Scotland, Wales, and Northern Ireland) | UK (although all participants from London) |
| Study methodology | Biographical narrative interviewing of 12 London GPs aged 55-65 |
| Type of document | Primary research |
| Topic or focus of paper | In this empirical study, we analyse the evolving professional experience during a career spanning several decades, addressing the question ‘How have institutional and social changes affected the morale of older GPs?’. |
| Research objectives and focus | To explore the impact upon morale and retirement decisions of changes in psychosocial aspects of UK general practice over the course of a career. |
| Theoretical framework/theories/ underpinnings | Jobs-Demands-Control-Support model was used to theorize findings |
| Description of intervention | |
| Rationale for paper/review | Retention in UK general practice has been falling for several years. This has usually been ascribed to high workload, in particular administrative burden and a complaints culture. However, few studies have used an empirically based framework to understand these workforce challenges.<br><br>Retrospective sense-making of professional experience affords a broad perspective from which to review an individual’s career, stepping back from day-to-day concerns [9], and has previously been used with senior GPs to explore what made their work satisfying, and how they |

| | |
|---|---|
| | adapted to organizational change. We extend the current literature by exploring career narratives of GPs aged 55–65, in which retirement decisions are seen in the context of a whole career story. |
| What creates meaning that sustains GPs engagement with, and involvement in, work? | Professional autonomy – both clinical and managerial<br><br>therapeutic power and idiosyncrasy of relationship-based care, and the sense of freedom the GPs had enjoyed to respond holistically to the patient, for example laying clinical notes aside to simply listen, or fixing a wheelchair, thus garnering trust.<br><br>Most of the GPs moved stepwise towards retirement, often developing a portfolio career to remain connected with meaningful work, whilst relinquishing the strain of partnership. |
| What solutions/recommendations are proposed? | For the GP Forward View to be successful in supporting the role of the GP whilst also meeting the changing healthcare needs of the UK population, balancing psychosocial demands and resources is important. Drawing upon empirically tested models like the JDCS model could inform job and service redesign to improve staff morale and retention. Support for professional autonomy is key, empowering GPs to be driven by professional values and expertise. Working conditions in which GPs and their teams can thrive would increase opportunities for |

| | |
|---|---|
| | collegial interactions and mentorship. There are demands that can be reduced and resources increased, as demonstrated in the table: |

**Table 2.**
Potential psychosocial demands and resources in general practice

| Demands | Resources |
|---|---|
| Work volume | Meaningfulness of clinical work |
| Work complexity | Ongoing learning and development |
| Working with lots of uncertainty | Appointment availability to review patients |
| Keeping up to date with broad, changing knowledge base | Support for learning |
| Time pressure | Administrative, clinical and practical support |
| Emotional labour of care | Satisfying patient relationships, fostered through continuity of care |
| Stressful/demanding consultations | Opportunities for supervision and mentoring |
| Lack of role clarity | Job security |
| Bureaucracy | Keeping administrative requirements to a minimum |
| Achieving clinical and administrative targets | Sparing use of targets, allowing adequate clinical discretion |
| Organizational change | Clinical autonomy |
| Time-consuming organizational processes | Genuine opportunities to shape GP locally and nationally |
| Organizational changes dictated | Opportunities to shape practice culture and process |
| Appraisal documentation | Supportive administrative and organizational processes |
| Complaints and litigation | Positive feedback from patients and colleagues |
| Regulatory demands | Good income/pension |
| Clinical responsibilities | Career progression opportunities |
| Employment responsibilities | Positive team environment, support for team development |
| Shift patterns/long/irregular hours | Choice over number of sessions |

| | |
|---|---|
| From which stakeholders' perspective? (General practitioners, patients, other HCPs) | GPs |
| How is work-related joy constructed? (Diversity of ways individuals may think about joy) | GPs enjoyed clinical and managerial autonomy, orchestrating the community team within and beyond the practice. These GPs described moving flexibly between thinking about the individual patient, the community context and an appropriate use of NHS resources. |
| What prevents/removes work-related joy? | Feeling less "in control"<br><br>The introduction of performance indicators in GP contracts resulted in growing resentment.<br><br>Target-driven care increased the administrative burden, and several directives encroached upon the discretion of the primary care team in organizing their work. For example, GPs saw their skill in triaging, an area in which clinical acumen comes into its own, being overridden by the national directive around 48-h access.<br><br>Those who took up roles within clinical commissioning groups (borough-level organizations tasked with delivering local healthcare services) described centrally driven priorities, with only lip-service being given to the 'clinical voice'.<br><br>increasing administrative and regulatory burdens, intense time pressures and the evolution of a consumerist patient culture<br><br>disillusionment and burnout, feeling alienated from a system that had changed the role of general practice from a community-based, person-centred profession that offered continuity of care, to a managed service emphasizing targets and budgets.<br><br>some were scared of what was perceived to be the potential void of retirement, the loss of professional identity, and others expressed guilt at making decisions that prioritized their own welfare over that of ongoing service. |
| Organisational characteristics (small, large) | |
| Managing ill-defined illnesses and chronic complex multimorbidity | |
| Human connection with patients | Several GPs gave examples of being so preoccupied with the 'bean-counting' of targets that they were in danger of missing the obvious and serious. |
| Human connection with peers | [practices were being] reshaped by a bureaucratic culture, losing the informal social support that the team previously offered one another. |

| | |
|---|---|
| Human connection across other organisations (e.g., secondary care) | Participants also described an erosion of their autonomy in their interaction with hospital colleagues, as referrals were increasingly processed via anonymous referral management systems. They also described a shift of work between secondary and primary care.<br><br>- “Letters [from secondary care] that demand we do things – blood tests, chase up results. Every letter will say something, as if we are their handmaidens in the community.” |
| Learning systems (e.g., competence, capability, mastery, socially situated) | |
| Additional useful information | |
| Summary of findings | |
| Strengths, limitations, and future research directions | This study offers both depth and historical breadth spanning four decades of general practice. A retrospective account, however, will be affected by subsequent events, whilst recounting and analysis of the interviews was shaped by the fact that the interviewer was a GP herself.<br><br>Whilst there may be sampling bias, with the participants representing a particular subsection of London GPs all of whom had extra-clinical professional interests, GPs across the UK are likely to resonate with the impact of progressive erosion of their autonomy. However, the small sample size of GPs working in an urban environment means these results are not generalizable. There is also possible responder bias with those motivated to discuss their career possibly differing from those who did not participate. |

| | |
|---|---|
| Paper Number | 150 |
| Author | Krachler and Kessler |
| Year | 2023 |
| Title | Ownership power and managing a professional workforce: general practitioners and the employment of physician associates |
| Journal | Human Resource Management Journal |
| Region (England, Scotland, Wales, and Northern Ireland) | UK |
| Study methodology | Review of policy documents covering workforce issues in primary care and focusing on development of the PA role<br>Informal discussion with innovative policymakers in London<br>Semi-structured interviews with GP partners, Pas and relevant stakeholders from London STP’s region. |

| | |
|---|---|
| Type of document | Primary research |
| Topic or focus of paper | This article explores the use of different forms of power by a profession, general practitioners (GPs), in engaging with a new healthcare role, the physician associate (PA). |
| Research objectives and focus | In this article, we examine why and how an established profession, the GP, harnesses ownership with managerial and knowledge-based power, to engage with a nascent professional role, the PA. |
| Theoretical framework/theories/ underpinnings | The three forms of power presented as underpinning the establishment and protection of professional interests are distinct, resting on differences in the scope and nature of decision-making:<br><br>• ***Managerial power***, founded on administrative seniority, with the capacity to direct and dispose of labor in daily work processes.<br>• ***Knowledge-based power***, derived from expertise which ensures occupations' control over the performance of work tasks.<br><br>• ***Ownership power***, allowing for authoritative organizational decision-making on strategic direction, but also for choices on the delegation of operational issues.<br><br>We take the case of the GP to explore how ownership, combined with management and knowledge, have been used to respond to the emergence of a new clinical work role in healthcare. |

| | |
|---|---|
| | 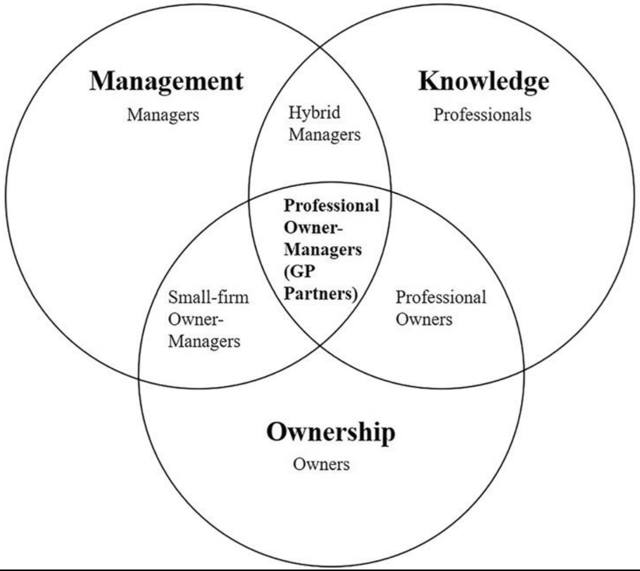 |
| Description of intervention | |
| Rationale for paper/review | |
| What creates meaning that sustains GPs engagement with, and involvement in, work? | |
| What solutions/recommendations are proposed? | |
| From which stakeholders' perspective? (General practitioners, patients, other HCPs) | GPs and PAs |
| How is work-related joy constructed? (Diversity of ways individuals may think about joy) | long-established personal relationships with patients.<br><br>viewing PAs' work as complementary, to meet the physician shortage and reduce GP workloads |

| | |
|---|---|
| What prevents/removes work-related joy? | The threat posed by PAs to GPs' professional values and status<br><br>the introduction of new roles that might disturb the unmediated GP-patient-relationship.<br><br>Fear of substitution by Pas as they are more cost-effective |
| Organisational characteristics (small, large) | |
| Managing ill-defined illnesses and chronic complex multimorbidity | |
| Human connection with patients | |
| Human connection with peers | |
| Human connection across other organisations (e.g., secondary care) | |
| Learning systems (e.g., competence, capability, mastery, socially situated) | |
| Additional useful information | |
| Summary of findings | First, our approach brings in ownership power to analyze the under-examined category of professional owner-manager, presenting the GP as a case of such a position. Second, we have conceptualized how professional owner-managers' three forms of power interact regarding workforce innovation through a new role (Figure 4). In doing so, we make our final contribution of highlighting two new control strategies used by established professions to address a nascent profession: employment denial and employment with subordination. |
| Strengths, limitations, and future research directions | Our research has several practical implications. Policymakers wishing to integrate the PA into the dominant GP- and nurse-led primary care model in England need to devise a business case establishing the new role's quality inputs and cost-benefits. […]work- force innovation in organizations owned and managed by professionals may be difficult to achieve without: a clear cost-benefit; resources to manage the new role; and professional concerns around safety being sufficiently allayed. |

| | |
|---|---|
| | This research has several limitations. As is common in theoretically-driven, qualitative research, our multi-source empirical approach aimed at enhancing the internal validity of the relations between categories in our conceptu- alization. The external validity of our conceptualization and findings could be further tested<br><br>Testing our findings for the case of PAs in primary care more formally might involve developing a database from a survey of practices employing PAs with administrative data about practices from NHS Digital to run regression or econometric analyses. Finally, while we have emphasized the power benefits of ownership, future research could examine any disadvantages of ownership power like large workloads that could make ownership unattractive for partnership professionals. |

| | |
|---|---|
| Paper Number | 151 |
| Author | Woodhams et al |
| Year | 2021 |
| Title | Pay gaps in medicine and the impact of COVID-19 on doctors' careers |
| Journal | The Lancet |
| Region (England, Scotland, Wales, and Northern Ireland) | England |
| Study methodology | |
| Type of document | Commentary |
| Topic or focus of paper | Gender pay gap in medicine and how the COVID-19 pandemic has affected the pay that women receive |
| Research objectives and focus | |
| Theoretical framework/theories/ underpinnings | |
| Description of intervention | |
| Rationale for paper/review | |
| What creates meaning that sustains GPs engagement with, and involvement in, work? | |
| What solutions/recommendations are proposed? | Our report makes recommendations to reduce the pay gap in medicine, which will be beneficial for women and for BAME colleagues. A first step in this process is an analysis of ethnicity and intersectional pay gaps. |

| | |
|---|---|
| From which stakeholders' perspective? (General practitioners, patients, other HCPs) | Doctors across hospitals, GPs and clinical academic roles |
| How is work-related joy constructed? (Diversity of ways individuals may think about joy) | |
| What prevents/removes work-related joy? | Pay, career opportunities, and work content:<br>[Managing] serious overwork and the worsening imbalance of work and life during the pandemic typically results in missed experience and leadership opportunities for women<br><br>the severe reduction in child-care provision will have disproportionately stalled the careers of women in medicine. Inadequate accessible child-care for UK medics has been shown to contribute to limited career progression.<br><br>The assumption that most women will be predisposed to prioritise family life, even if they have no children, is already damaging to their prospects of career progression.<br><br>During the COVID-19 pandemic it is likely that most accommodations to adjust to the shortage of child-care will have been made by female doctors.<br><br>the pay gap is wider for women in minority ethnic groups than for White women; the groups most affected are Pakistani and Bangladeshi women with pay gaps of 30% or more.<br>The reasons for this may include structural inequalities or discrimination. |
| Organisational characteristics (small, large) | |
| Managing ill-defined illnesses and chronic complex multimorbidity | |
| Human connection with patients | |
| Human connection with peers | |
| Human connection across other organisations (e.g., secondary care) | |

| |
|---|
| Learning systems (e.g., competence, capability, mastery, socially situated) |
| Additional useful information |
| Summary of findings |
| Strengths, limitations, and future research directions |

| | |
|---|---|
| Paper Number | 152 |
| Author | Robin Miller |
| Year | 2016 |
| Title | Pioneering outcomes based innovation in general practice |
| Journal | |
| Region (England, Scotland, Wales, and Northern Ireland) | |
| Study methodology | |
| Type of document | Abstract on emerging findings of primary research |
| Topic or focus of paper | |
| Research objectives and focus | how to motivate and enable practices to take on these enhanced responsibilities and be willing to work differently with other professionals and services.<br><br>The ‘pioneer project’ aimed to introduce a new purchasing approach which would motivate and liberate general practices to develop and adopt new integrated models of care through providing a more ‘enabling environment’. |
| Theoretical framework/theories/ underpinnings | |
| Description of intervention | The key elements of this approach were -<br><br>- Requiring practices to work in groupings rather than as single practices<br><br>- Providing funding up front rather than in arrears on completion of required activity or quality |

| | |
|---|---|
| | - Determining success by achievement of outcomes rather than by implementing process<br><br>- Embedding a culture of joint learning and reflection between the practices and with the commissioner |
| Rationale for paper/review | |
| What creates meaning that sustains GPs engagement with, and involvement in, work? | The practice leads confirmed that the previous arrangements had stifled innovation and led to them not feeling able or motivated to suggest or lead more radical improvements. Adjusting to the new expectations and freedoms involved a significant paradigm shift for them (and the commissioner) |
| What solutions/recommendations are proposed? | |
| From which stakeholders' perspective? (General practitioners, patients, other HCPs) | |
| How is work-related joy constructed? (Diversity of ways individuals may think about joy) | |
| What prevents/removes work-related joy? | |
| Organisational characteristics (small, large) | |
| Managing ill-defined illnesses and chronic complex multimorbidity | All groupings of practices have been able to provide enhanced diagnostics and treatment for key conditions such as diabetes. This has enabled diversion of these activities from acute to primary care and provided care closer to patients' homes. |
| Human connection with patients | |
| Human connection with peers | |
| Human connection across other organisations (e.g., secondary care) | The majority of groupings have developed new ways of working which respond to the needs of their local populations – these include the development of innovative model of shared care between acute and primary care regarding people with diabetes from south asian |

| | |
|---|---|
| | communities, the introduction of new joint protocols with accident and emergency, and community link workers to connect people with complex social needs with appropriate support from statutory and voluntary sector services |
| Learning systems (e.g., competence, capability, mastery, socially situated) | |
| Additional useful information | - There was greater success in engaging with key services in the acute sector through engaging with key professionals – community health and adult social care were harder to influence due to lack of personal connection and alternative initiatives being pursued |
| Summary of findings | - The pioneer project facilitated a more collaborative relationship between purchaser and general practice which encouraged and facilitated greater clinical leadership. For these groups of practices funding was an enabler rather than an incentive, providing 'capacity and flexibility' rather than 'motivation and reward' |
| Strengths, limitations, and future research directions | - The final data gathering will seek to establish improvements in outcomes and quality, and the extent to which changes have been sustained when the project phase is completed. |

| | |
|---|---|
| Paper Number | 153 |
| Author | Chris Thomas and Harry Quilter-Pinner |
| Year | 2020 |
| Title | Realising the neighbourhood NHS: A new deal for primary care in England |
| Journal | |
| Region (England, Scotland, Wales, and Northern Ireland) | England |
| Study methodology | Semi-structured interviews with young GPs relatively near the start of their career.<br>Polls to ensure a diverse range of views on the future of General Practice in England could be established.<br>This was complimented by secondary analysis of key NHS datasets |
| Type of document | Primary research |
| Topic or focus of paper | |
| Research objectives and focus | |
| Theoretical framework/theories/ underpinnings | |
| Description of intervention | |
| Rationale for paper/review | primary care in England needs fundamental change |

| | |
|---|---|
| What creates meaning that sustains GPs engagement with, and involvement in, work? | Control of workload<br>Work-life balance<br><br>GPs increasingly want more flexibility eg portfolio career<br><br>Small practices have allowed GPs to build strong relationships with patients and the local community, delivering much valued continuity of care. Furthermore, their independent status has allowed GPs to manage their own work environment and innovate with patient care without having to tackle the NHS bureaucracy.<br><br>GPs are primarily funded through core national contracts called the GMS for delivery of essential services (see table 2.5). GMS funding is made up of the global sum (capitated payments) based on the age and gender of patients and other factors. Partners fund their costs through this and keep any excess as profits. These contracts therefore create what we call the 'inverse pay law': those GPs that are the most efficient and deliver the most basic care keep the most profit. This limits the incentive for general practitioners to innovate and expand provision.<br><br>Even in the short time since the *Five Year Forward View* was published, salaried GPs have increased by 3,000, and now make up nearly a third of all GPs, whilst partners have decreased in numbers by a similar amount (see figure 3.1) though it is worth noting that locum GPs have been growing at an even faster pace (see box 3.2). If these trends continue at their current pace, salaried GPs will become the majority by 2026.[4]<br><br>It was felt that variety was the antidote to ensuring the role of general practitioner remained sustainable in the future.<br><br>Time for career development, respite and developing other specialisms |
| What solutions/recommendations are proposed? | Instead, we will need to shift to primary care at scale. Only across larger populations of patients - as a result of economies of scale - is it cost effective to invest in longer opening ours; diagnostics and treatment in the community, and a wider team of healthcare professionals to deliver integrated care.<br><br>Experts have argued that new 'neighbourhood NHS' organisations should be set up around real places – neighbourhoods – to serve between 50,000 and 100,000 people each<br><br>these new organisations should take a whole-person population health approach. This means segmenting the population into groups with broadly similar needs (eg those with long-term conditions) and then bringing together all parts of the health and care system around them to deliver joined up and preventative care. |

| | |
|---|---|
| | while the GP may remain the focal point of primary care, these new 'neighbourhood NHS' systems should be team based. This will help to relieve pressure on the GP, reduce demand on the acute sector and offer more joined up and accessible care to the patient.<br><br>To this end, we propose that over time PCNs should move away from loose 'federations' or 'networks' towards new neighbourhood care providers (NCPs) across every local area, on the same geographic footings of PCNs – or of several PCNs, where they are smaller. These trusts could either be newly created or could be formed by existing community trusts, more advanced PCNs or multi-speciality community providers (MCPs). Over time these NCPs should take on the contracts for primary, mental health and community care. They could also deliver social care and public health in order to really fulfil the possibility of population health.<br><br>All general practitioners in the NHS in England should be given a right to NHS employment. Senior general practitioners (both partners and salaried employees) should be recruited into management roles within the new NCPs (on similar salary levels to a current GP partner) alongside other non-clinical senior managers. All newly qualified GPs should also be offered a salaried role within the NHS. In the short-term GPs should have the right to refuse this offer (eg to maintain their partnership model) but over time policymakers should consider ending the right of partnerships to create new or replace existing partnership roles.<br><br>We need to move away from a nine-session week (which works out to far more than 37.5 hours indicated by government) - and which is dominated by clinical sessions. Full-time work should be defined as eight sessions, of which a maximum of six sessions should be clinical. This may seem extravagant in the context of GP shortages, but overwork is a false economy: it is driving GPs to reduce hours or leave the profession.<br><br>Automation also has significant potential to ease pressure (see table 3.3) by reducing workload (particularly administration). Elsewhere, we should invest in the wider primary care team (eg social prescribers, nurses, pharmacists etc) and invest in patient empowerment to support people to manage their health better in the community.<br><br>The NHS has rightly proposed that practices be moved to 1,500 purpose built 'super hubs' across the country… We therefore recommend that the government provides funding for at least one new hub per PCN, delivered as a strategic transformation fund and prioritised by levels of need.<br><br>government should commit to a radical upgrade of infrastructure. On the one hand, this must mean much higher quality use of technology. This can help support integration – through achieving interoperability. It can also help to save time – by automating administrative parts of the role and letting staff work on what they joined the career to do. |
| From which stakeholders' perspective? (General practitioners, patients, other HCPs) | GPs, general public and senior community health professionals |

| | |
|---|---|
| How is work-related joy constructed? (Diversity of ways individuals may think about joy) | |
| What prevents/removes work-related joy? | The average GP has dropped their hours from 0.84 FTE to 0.76 FTE (full-time equivalent) – though this is likely as much an indicator of the unfeasible increase in the amount work represented by each GP ‘session’, both clinical and non-clinical, which is making full-time work unsustainable for many.<br><br>The move to part-time work (or to lower levels of clinical work) is likely to represent an attempt by GPs to control their workload and work-life balance (GMC 2019). This is likely driven, at least in part, by the increase in workload, stress and pressure on an individual GP.<br><br>Simultaneously the workload of general practitioners is rising, driven by three key trends.<br><br>- Increased bureaucracy<br>- Growing complexity<br>- Growing knowledge<br><br>GPs shouldn’t be blamed for increasingly choosing to locum instead of taking salaried roles. Instead, this is the result of the inability of policymakers to address the workforce crisis in primary care which means salaried roles remain unattractive in terms of workload, stress and progression.<br><br>The majority of GPs believe workload is risking patient safety<br><br>There are concerns over the health of GPs as a result of stress and overwork: a survey of 1,000 GPs found that two in five have a mental health problem<br><br>Being a partner often includes a significant administrative burden and financial risks which GPs are often neither trained to cope with, nor passionate about.<br><br>Standards set by government for full-time GP work.<br><br>Many do not have the investment in technology that is needed to drive 21st century primary care such as remote care and digital administration limiting the potential of automation<br><br>The partnership model is at the heart of this crisis. This is because partnership in most cases creates a link between the property market and care, with senior GPs owning and managing the primary care estate. This has contributed to underinvestment in the sector with partners |

<table>
<tr>
<td></td>
<td>unable or reluctant to take on the financial risk of investing in the primary care estate and the NHS rightly unwilling to make big investments in what is essentially private property.</td>
</tr>
<tr>
<td>Organisational characteristics (small, large)</td>
<td>
Small scale independent general practice limits the type of care the primary care system is able to offer.
<ul>
<li>often unable to offer quick and seamless access to the wider primary care team</li>
<li>confines care to inflexible and short appointment slots</li>
<li>limits the ability of primary care to invest in the modern buildings and new technologies needed to offer telephone, email, skype based care or diagnostics/treatment in the community.</li>
</ul>
It is also easier for crucial community partnerships to form between larger organisations
Organisation forms of large-scale general practice organisations
<table>
<tr><th>Organisational form</th><th>Characteristics</th><th>Example</th></tr>
<tr><td>Network</td><td>No formal ties; practices maintain GP contracts<br>No executive function<br>Share principally intangible objectives</td><td>Most of country</td></tr>
<tr><td>Federation</td><td>Growing ties; maintain GP contracts but develop legal agreements for shared activity<br>Employ an executive function & some clinical roles<br>Share some organisational goals</td><td>PCNs</td></tr>
<tr><td>Super-partnership</td><td>Close ties; practices merge GP contracts<br>Employ executive function and management team<br>Shared goals across whole organisation (multiple practices)<br>Pool all/most income and risk</td><td>Modality</td></tr>
<tr><td>Multi-site practice organisation</td><td>Tight ties; same as above but directors hold all GP contracts in a single organisation</td><td>Hurley Group</td></tr>
</table>
A ‘network’ or ‘federation’ model locks in lose partnership working and fragmentation in primary care
However, all these arrangements [QoF incentives, PMS contracts and other funding streams] are ultimately imperfect workarounds that fail to address the underlying problem. As long as we have many independent practices built around the GMS contract, the incentive will always be to protect organisational boundaries (to maintain smaller organisations) and therefore form only loose federations.
</td>
</tr>
</table>

| | |
|---|---|
| Managing ill-defined illnesses and chronic complex multimorbidity | Compared to other countries the NHS underperforms in terms of cancer survival rates, partly as a result of a failure to diagnose cancers early. Whilst, for other patients treated by the primary care system, such as those with diabetes or mental ill-health, quality may be similar or better than other countries but still requires improvement<br><br>Notably, people are growing older but are living more of their life in ill health. Their illnesses are increasingly chronic rather than acute, including diabetes, mental illness, frailty and dementia. This shift in the disease burden demands a fundamentally different model of care to that which we have had in the past |
| Human connection with patients | Patients report highly positive experiences of primary care. In 2019, 84 per cent reported a good experience, a small increase on 2018 (83 per cent). These national scores can, however, mask avoidable variation in care quality.<br><br>Primary care is struggling to provide adequate levels of service access. New polling commissioned by IPPR shows that 36 per cent of people in England waited longer than a week for an appointment, the last time they needed one.<br><br>Furthermore, access to a GP is actually slower for those with a pre-existing diagnosis. Of all those living with a long-term condition (n = 1,018), 43 per cent faced a waiting time of more than a week.<br><br>Patients increasingly demand that care is preventative, joined up, accessible and personal<br><br>Over half of people living with a long-term condition received no bespoke support from primary care – despite many of the interventions listing being key and long-standing NHS commitments.<br><br>However, people with long-term conditions do have better access to primary and community professionals who are not GPs |
| Human connection with peers | This {establishing PCNs) is a significant step forward: it formalises partnership working in all areas of the country and ties significant income to participation in this collaboration.<br><br>whilst a recent survey by the Nuffield Trust found that over four-fifths of GPs were working with other GPs, they also found that over half were not formalised, one-fifth had hired no non-clinical staff and the majority were loose ‘networks’ or ‘federations’ of independent practices rather than more integrated ‘super-partnerships’ or single integrated organisations |

| | |
|---|---|
| Human connection across other organisations (e.g., secondary care) | policymakers have in fact bent over backwards to protect the partnership model in primary care at the expense of delivering truly integrated primary care at scale. This must now change. |
| Learning systems (e.g., competence, capability, mastery, socially situated) | |
| Additional useful information | Without primary care at scale, joined-up, accessible, preventative and personal care is simply not obtainable. |
| Summary of findings | Comment: Overall, there seems to be a big push from this document to eliminate the model of independent GP practices and erase the boundaries of keeping practices as small organisations; instead a primary care at scale approach should be used to create a large network of GP practices that are tied to more formal partnerships and shared responsibility. The paper also suggests that all GPs should be employed by the NHS rather than be partners. |
| Strengths, limitations, and future research directions | |

| | |
|---|---|
| Paper Number | 155 |
| Author | Kathy Oxtoby |
| Year | 2022 |
| Title | A true family doctor: the general practitioner |
| Journal | BMJ |
| Region (England, Scotland, Wales, and Northern Ireland) | England |
| Study methodology | Interview with senior GP |
| Type of document | Commentary |
| Topic or focus of paper | Continuity of care |
| Research objectives and focus | Interview a senior GP (Biplab Das) about how his love of being a “cradle to grave” family doctor and how his commitment to serving his community drew him back from retirement |
| Theoretical framework/theories/ underpinnings | |

| | |
|---|---|
| Description of intervention | |
| Rationale for paper/review | |
| What creates meaning that sustains GPs engagement with, and involvement in, work? | |
| What solutions/recommendations are proposed? | |
| From which stakeholders' perspective? (General practitioners, patients, other HCPs) | |
| How is work-related joy constructed? (Diversity of ways individuals may think about joy) | |
| What prevents/removes work-related joy? | |
| Organisational characteristics (small, large) | |
| Managing ill-defined illnesses and chronic complex multimorbidity | |
| Human connection with patients | "I like the continuity of care that general practice brings. It gives me the opportunity to look after patients from birth until their last day," he says.<br><br>Over the years he has enjoyed looking after his patients' children, and their children's children. His patients send him birthday and Christmas cards, and their relatives ask him to give readings at their funerals because he knew them so well. "I see myself as a family GP. I enjoy that caring bond with my patients. So many people have no one they can talk to. My patients can talk to me." |
| Human connection with peers | |
| Human connection across other organisations (e.g., secondary care) | |

| | |
|---|---|
| Learning systems (e.g., competence, capability, mastery, socially situated) | |
| Additional useful information | |
| Summary of findings | Biplab Das explains how being a “cradle to grave” GP is very meaningful. |
| Strengths, limitations, and future research directions | |

| | |
|---|---|
| Paper Number | 156 |
| Author | Adrian O’Dowd |
| Year | 2022 |
| Title | Role model: Venu Prasad |
| Journal | BMJ |
| Region (England, Scotland, Wales, and Northern Ireland) | Wales |
| Study methodology | Interview with a senior GP |
| Type of document | Commentary |
| Topic or focus of paper | Continuity of care & human interaction in general practice |
| Research objectives and focus | Interview a senior GP (Venu Prasad) about why he is happy to continue working as a locum after almost 30 years in full time employment |
| Theoretical framework/theories/ underpinnings | |
| Description of intervention | |
| Rationale for paper/review | |
| What creates meaning that sustains GPs engagement with, and involvement in, work? | |

| | |
|---|---|
| What solutions/recommendations are proposed? | |
| From which stakeholders' perspective? (General practitioners, patients, other HCPs) | |
| How is work-related joy constructed? (Diversity of ways individuals may think about joy) | |
| What prevents/removes work-related joy? | |
| Organisational characteristics (small, large) | |
| Managing ill-defined illnesses and chronic complex multimorbidity | |
| Human connection with patients | |
| Human connection with peers | "It appealed to me, and I decided to give it a go. It was one of the best decisions I ever made. I am a people person and like human interaction and the continuity of care you get in general practice."<br><br>He secured a principal GP position at a small practice in north Wales in 1988, where he continued working for the next 29 years. "It was a small town and a small practice, and it took time for me to get to know the patients and for them to get to know me. But gradually, over a period of time, you establish that wonderful relationship and patients trust you," he says. |
| Human connection across other organisations (e.g., secondary care) | |
| Learning systems (e.g., competence, capability, mastery, socially situated) | |
| Additional useful information | |
| Summary of findings | Venu Prasad explains how he likes the "human interaction and continuity of care" you get in general practice. |

| |
|---|
| Strengths, limitations, and future research directions |

| | |
|---|---|
| Paper Number | 157 |
| Author | Claire Fuller |
| Year | 2022 |
| Title | Next steps for integrating primary care: Fuller Stocktake report |
| Journal | Policy document |
| Region (England, Scotland, Wales, and Northern Ireland) | England |
| Study methodology | Engagement (written submissions, interviews and roundtables with a wide range of primary care staff) |
| Type of document | Policy document |
| Topic or focus of paper | Consider the role of primary care within newly established ICSs |
| Research objectives and focus | |
| Theoretical framework/theories/ underpinnings | |
| Description of intervention | |
| Rationale for paper/review | Patient access and satisfaction with primary care is at an all-time low. At the same time, primary care teams are stretched beyond capacity and staff morale is at a record low. In short, left as it is, primary care as we know it will become unsustainable in a relatively short period of time |
| What creates meaning that sustains GPs engagement with, and involvement in, work? | |
| What solutions/recommendations are proposed? | In order for GPs to provide same day urgent care AND continuity of care, we need to understand that NHS urgent care is what patients access first in their community (e.g. online advice, community pharmacy).<br><br>We need to improve the experience of working in primary care for everyone by making the employment culture more compassionate and inclusive, and listening much more effectively to what primary care staff are telling us. |

| | |
|---|---|
| | The NHS staff survey is already being piloted in some areas of general practice and now needs to be extended nationwide and considered for NHS-funded primary care. Identifying ways to support and listen to staff who are working as carers would also be very welcome, and primary care staff should have access to Freedom to Speak Up guardians, promoting an open and listening culture. Workforce data, staff surveys and other feedback mechanisms for staff, should be used by ICSs and local leaders across primary care to take action to improve equality, diversity and inclusion across the primary care workforce.<br><br>We must tackle racial discrimination and harassment because it is the right thing to do, it is crucial to retain our staff, and to further strengthen how the primary care workforce reflects and strengthens its connection with the diverse communities it serves. We must value the important contribution that individuals with protected characteristics, including age, sex, religion or belief, people with disabilities, those from the LGBTQ+ community, black and minority ethnic backgrounds, and with caring responsibilities, make as part of our workforce. Ensuring flexible working and other forms of support are available to these groups and any others that experience discrimination in the workplace should be central to local, system-level and national workforce strategies.<br><br>Systems should drive a more standardised and improved employment offer for primary care in line with the NHS People Promise: for example, by ensuring parity of access to system staff health and wellbeing hubs and occupational health services, and by encouraging employers to adopt NHS terms and conditions by sharing existing good practice and model contracts.<br><br>Decisions about digital infrastructure in primary care need to be made in partnership with those who will use them – including engagement with both staff and patients. |
| From which stakeholders' perspective? (General practitioners, patients, other HCPs) | Written by a senior GP and ICB CEO on the back of extensive engagement with a range of primary care staff. |
| How is work-related joy constructed? (Diversity of ways individuals may think about joy) | |
| What prevents/removes work-related joy? | Primary care has never been busier, and capacity gaps lie behind most of the challenges that the NHS faces. These gaps – and the increased demand for services – were growing in the decade before COVID-19 due to workforce pressures and reduced staff satisfaction, the increasing number of people living with multiple long-term conditions, and changes in public expectations. |
| Organisational characteristics (small, large) | There are 8,911 premises in England, 22% of which are pre-1948 and 49% of which are owned by GPs, 35% owned by a third party, and 14% owned by NHS Property Services. Around 2,000 premises have been identified by GPs as not being fit for purpose, and there was strong feedback throughout the stocktake that we do not start thinking about estates early enough in our planning and frequently regret it. |
| Managing ill-defined illnesses and chronic complex multimorbidity | |
| Human connection with patients | |

| | |
|---|---|
| Human connection with peers | "Continuity of care, specifically the relationship between a named GP and their patient, is directly linked to improvements in patient experience and lower mortality, especially for more complex patients. This is a core strength of primary care, and we repeatedly heard the fundamental importance of this from staff across primary care and patients alike." – however, not all patients want or need continuity of care. Determining which patients benefit most from more personalised continuity of care can depend on a range of medical, psychological or social reasons and should be determined through conversations with patients and using clinical judgement, as well as supported by risk stratification using the wealth of data increasingly available to primary care teams. |
| Human connection across other organisations (e.g., secondary care) | |
| Learning systems (e.g., competence, capability, mastery, socially situated) | |
| Additional useful information | |
| Summary of findings | Much of this report is not directly relevant to GPs relationship with work. However, it highlights opportunities to improve the experience of primary care staff, explains how we can balance access AND continuity, highlights widespread problems with the primary care estate, and emphasises that – whilst continuity of care is important – it might not be necessary for all patients. |
| Strengths, limitations, and future research directions | |

| | |
|---|---|
| Paper Number | 158 – [extracted an overview of the main points] |
| Author | House of Commons Health and Social Care Committee |
| Year | 2022 |
| Title | The future of general practice |
| Journal | N/A |
| Region (England, Scotland, Wales, and Northern Ireland) | UK |
| Study methodology | Prepared following the submission of extensive written and witness evidence to the House of Commons Health and Social Care Select Committee |
| Type of document | Policy document |
| Topic or focus of paper | The future of general practice |
| Research objectives and focus | [Not stated] |

| | |
|---|---|
| Theoretical framework/theories/ underpinnings | |
| Description of intervention | |
| Rationale for paper/review | [Not stated] |
| What creates meaning that sustains GPs engagement with, and involvement in, work? | |
| What solutions/recommendations are proposed? | We recommend abolishing the Quality and Outcomes Framework (QOF) and Impact and Investment Framework (IIF) which have become tools of micromanagement and risk turning patients into numbers. GPs should be treated like professionals and incentivised to provide relationship-based care for all patients by restoring individual patient lists.<br><br>It is time to recognise the need to make the job not just manageable but once again fulfilling and enjoyable. For doctors it should allow a cradle to grave relationship with patients not possible for other specialties but for many infinitely more rewarding. To do that general practice needs to have its professional status restored with a decisive move away from micromanagement and short staffing to a win-win environment in which investment in general practice reduces pressure on hospitals and saves resources for the NHS. |
| From which stakeholders' perspective? (General practitioners, patients, other HCPs) | Engagement with a very wide range of primary care stakeholders |
| How is work-related joy constructed? (Diversity of ways individuals may think about joy) | |
| What prevents/removes work-related joy? | We know up to 90% of healthcare is delivered by primary care. Yet currently the profession is demoralised, GPs are leaving almost as fast as they can be recruited, and patients are increasingly dissatisfied with the level of access they receive. The root cause of this is straightforward: there are not enough GPs to meet the ever-increasing demands on the service, coupled with increasing complexity of cases from an ageing population.<br><br>GPs are often left to deal with multiple complex cases one after another. This can contribute to clinician burnout and concerns by the clinicians they might make mistakes or not be able to practise safely. This combination of intensely complex cases, done at speed, with fear over reprisals on the individual clinician is driving a systemically toxic environment in primary care.<br><br>For GP partners at the end of their careers, one of the biggest barriers to staying on longer is the huge pensions tax bills that many face. We continue to call for the Government to take specific action to allow senior doctors, including GPs, to carry on working without facing these tax bills. |

| | |
|---|---|
| Organisational characteristics (small, large) | Historically one of the key drivers of innovation and improvement in general practice has been the GP partnership model, which gives GPs the flexibility to innovate with a focus on the needs of their local population. We know there are significant pressures on GP partners at the moment but the evidence we received was clear that the partnership remains an efficient and effective model for general practice if properly funded and supported. It is important that the model of general practice can vary according to local needs, so other models of delivery should also continue to be explored where this works for local communities. |
| Managing ill-defined illnesses and chronic complex multimorbidity | |
| Human connection with patients | Since 2004 the majority of GPs have not had individual lists of patients even though there is clear international and UK research showing that seeing the same GP over a long period of time leads to fewer hospital visits, lower mortality and less cost for the NHS.<br><br>There can sometimes be a trade-off between access and continuity, and we believe that the balance has shifted too far towards access at the expense of continuity. Seeing your GP should not be like phoning a call centre or booking an Uber driver who you will never see again: relationship-based care is essential for patient safety and patient experience. It is also much more motivating for doctors. |
| Human connection with peers | |
| Human connection across other organisations (e.g., secondary care) | |
| Learning systems (e.g., competence, capability, mastery, socially situated) | |
| Additional useful information | Continuity of care is beneficial for all patient interactions even if it cannot always be offered. It should not therefore be available only for patients with complex needs, because part of the purpose of a long-term relationship between a doctor and patient is to prevent chronic or long-term illness before it happens. |
| Summary of findings | Key points relevant to our review include: abolishing the QOF and incentivising relationship-based care; combination of intensely complex cases with fear of reprisals creating toxic environment; GP partnership model as a key driver of innovation; balance between access and continuity shifted too far towards continuity. |
| Strengths, limitations, and future research directions | |

| | |
|---|---|
| Paper Number | 159 – [This is a long and detailed document, so I have extracted an overview of the main points] |
| Author | McDonald et al |
| Year | 2020 |
| Title | The impacts of GP federations in England on practices and on health and social care interfaces: four case studies |

| | |
|---|---|
| Journal | Health Services and Delivery Research |
| Region (England, Scotland, Wales, and Northern Ireland) | England |
| Study methodology | Structured cross-sectional comparison of four case studies of GP federations, using observation of meetings, interviews and analysis of documents. We combined inductive analysis with literature on ‘meta-organisations’ and networks to provide a theoretically informed analysis. |
| Type of document | Primary research |
| Topic or focus of paper | GP federations |
| Research objectives and focus | Assess how federating affects practice processes, workforce, innovations in practices and the interface with health and social care stakeholders. |
| Theoretical framework/theories/ underpinnings | |
| Description of intervention | |
| Rationale for paper/review | |
| What creates meaning that sustains GPs engagement with, and involvement in, work? | |
| What solutions/recommendations are proposed? | |
| From which stakeholders’ perspective? (General practitioners, patients, other HCPs) | Wide range of stakeholders including GPs, |
| How is work-related joy constructed? (Diversity of ways individuals may think about joy) | |
| What prevents/removes work-related joy? | |
| Organisational characteristics (small, large) | We found evidence that federation initiatives were helping with recruitment issues in two sites (authoritative and indulgent). In addition, solutions to avoid reliance on external locums, supporting resilience for practices, as well as consistency for patients, were implemented at these sites. At one site (authoritative), the move to add partners to contracts also provided support for practices, which felt themselves to be |

| | |
|---|---|
| | especially vulnerable to practice closure in the event of GP sickness. Here, non-GP partners were transferred from their employing practice over to the federation's employment and pharmacists were also employed by the federation. Federating meant that, in some sites, there were central federation staff who could provide advice and support, but for many of the senior GPs who actively participated in federation meetings and activities their overall workload increased.<br><br>Federations were also able to provide new services for patients. |
| Managing ill-defined illnesses and chronic complex multimorbidity | |
| Human connection with patients | |
| Human connection with peers | |
| Human connection across other organisations (e.g., secondary care) | |
| Learning systems (e.g., competence, capability, mastery, socially situated) | |
| Additional useful information | |
| Summary of findings | Most of this paper is not relevant to GPs' relationship with work. It notes that federating was useful to increase resilience of practices in response to GP sickness. However, federations could lead to increased GP workload. From the perspective of patients, federations were able to provide new services. |
| Strengths, limitations, and future research directions | The focus was on only four sites; therefore, any federations that were more active than those federations in these four sites will have been excluded |

| | |
|---|---|
| Paper Number | 160 |
| Author | Ward, Cartwright-Terry & Kirby |
| Year | 2015 |
| Title | The need to lead – a newly qualified GP's perspective |
| Journal | Education for Primary Care |
| Region (England, Scotland, Wales, and Northern Ireland) | England |

| | |
|---|---|
| Study methodology | Commentary |
| Type of document | Commentary |
| Topic or focus of paper | GP leadership |
| Research objectives and focus | Highlight the importance of addressing leadership skills in GP training – and offer some suggestions of how to do this. |
| Theoretical framework/theories/ underpinnings | |
| Description of intervention | |
| Rationale for paper/review | Newly qualified GPs do not want to take up leadership roles |
| What creates meaning that sustains GPs engagement with, and involvement in, work? | |
| What solutions/recommendations are proposed? | Suggestions to help GPs develop leadership skills:<br>1. In general, many GPs have leadership skills and implicitly develop these during their training. However, these need to be signposted more. In particular, this would dispel preconceptions about leadership.<br>2. Routine half day release to discuss different leadership experiences with our peers and take inspiration from them. Facilitation of such groups by more experienced tutors who know how to translate the implicit experience of leadership into the explicit language of leadership skills would be ideal<br>3. Possibility of mentorship would be another powerful way of building leadership ability beyond the core skills for First5© GP's.<br>4. Urgent need for organisations such as the CCG, British Medical Association and Local Medical Committees to actively encourage and recruit trainees and newly qualified GPs to their teams and furthermore, support them in developing leadership skills.<br>5. There may be a lack of awareness amongst GPs about leadership learning resources |
| From which stakeholders' perspective? (General practitioners, patients, other HCPs) | Two salaried GPs and a GP partner. |
| How is work-related joy constructed? (Diversity of ways individuals may think about joy) | |
| What prevents/removes work-related joy? | Currently, management battles to preserve the primary care service that we and our patients value are undertaken mainly be experienced, senior GPs. This can lead to burnout. One way to prevent this is to share the burden of leadership responsibility, by promoting leadership skills in GP training. |

| | |
|---|---|
| Organisational characteristics (small, large) | |
| Managing ill-defined illnesses and chronic complex multimorbidity | |
| Human connection with patients | |
| Human connection with peers | |
| Human connection across other organisations (e.g., secondary care) | |
| Learning systems (e.g., competence, capability, mastery, socially situated) | |
| Additional useful information | |
| Summary of findings | Experienced, senior GPs bear the burden of leadership, which can lead to burnout. Can prevent this by sharing the burden and promoting leadership skills in GP training. This may be done by explicitly signposting skills and resources, half day release to discuss leadership with experienced facilitator, mentorship, and involvement of organisations like the CCG, BMA and local medical associations. |
| Strengths, limitations, and future research directions | |

| | |
|---|---|
| Paper Number | 161 |
| Author | RCGP learning |
| Year | 2020 |
| Title | Managing stress and avoiding burnout in primary care |
| Journal | RCGP blog |
| Region (England, Scotland, Wales, and Northern Ireland) | N/A |
| Study methodology | Commentary |
| Type of document | Commentary |

| | |
|---|---|
| Topic or focus of paper | Managing stress |
| Research objectives and focus | Discuss how GPs can recognise and deal with stress |
| Theoretical framework/theories/ underpinnings | |
| Description of intervention | |
| Rationale for paper/review | Nine out of ten GPs reported experiencing considerable or high pressure from increasing workloads, amongst other stressors such as paperwork and having insufficient time in the working day to do their jobs. |
| What creates meaning that sustains GPs engagement with, and involvement in, work? | |
| What solutions/recommendations are proposed? | |
| From which stakeholders' perspective? (General practitioners, patients, other HCPs) | [Does not specify] |
| How is work-related joy constructed? (Diversity of ways individuals may think about joy) | Taking time to improve wellbeing relieves stress. The new economics foundation outlines five ways to wellbeing:<br>**Connect –** Make time for family and friends. Build relationships with colleagues and neighbours. Become part of your local community.<br>**Take notice –** Think about what matters to you most and take time to appreciate it. Be aware of your feelings and emotions. Try meditation and mindfulness.<br>**Keep learning –** Try something new and embrace new experiences. Learning a new skill can boost your confidence and create an escape from stressors.<br>**Be active –** Think about which physical activity you enjoy and that suits you. Regular exercise is associated with lower rates of depression and anxiety and helps to slow down age-related cognitive decline.<br>**Give –** Do something nice for a friend or stranger or volunteer your time to the community. Feeling part of a wider community can be rewarding and build connections with those around you.<br>In order to ensure the wellbeing of doctors in their workplace, the GMC outline an 'ABC of doctors' core needs':<br>**Autonomy or control –** the need to have control over our work lives, and to act consistently with our work and life values. |

| | |
|---|---|
| | **Belonging –** the need to be connected to, cared for, and caring of others around us in the workplace and to feel valued, respected and supported.<br>**Competence –** the need to experience effectiveness and deliver valued outcomes, such as high-quality care |
| What prevents/removes work-related joy? | 72% of younger GPs feel that their work-life balance has deteriorated as a result of their stress at work. |
| Organisational characteristics (small, large) | |
| Managing ill-defined illnesses and chronic complex multimorbidity | |
| Human connection with patients | |
| Human connection with peers | |
| Human connection across other organisations (e.g., secondary care) | |
| Learning systems (e.g., competence, capability, mastery, socially situated) | |
| Additional useful information | The BMA's 2019 report on 'Caring for the mental health of the medical workforce' found that a total of 88% of GP partners and 85% of sessional GPs face a 'high' or 'very high' risk of burnout<br><br>For any GPs who are struggling with the pressures of their role or showing signs of stress or burnout, the following resources may help: NHS GP Health Service, Online CBT, The Balint Society, Mentoring, Mental Health Apps, Big White Wall, RCGP. |
| Summary of findings | General practice is acknowledged as one of the most stressful jobs in medicine. In general, wellbeing is increased through five routes (connect, take notice, keep learning, be active, give). The BMA's ABC of doctors core needs are: Autonomy, Belonging, Competence. |
| Strengths, limitations, and future research directions | |

| | |
|---|---|
| Paper Number | 162 |
| Author | Mark Vorster |
| Year | 2017 |

| | |
|---|---|
| Title | Support for grassroots could rescue general practice |
| Journal | BJGP |
| Region (England, Scotland, Wales, and Northern Ireland) | England |
| Study methodology | Commentary |
| Type of document | Commentary |
| Topic or focus of paper | Grassroots general practice |
| Research objectives and focus | Explain how establishing a robust, local primary care team will help solve some of the problems in general practice |
| Theoretical framework/theories/ underpinnings | |
| Description of intervention | |
| Rationale for paper/review | The current model of general practice is unsustainable: without fundamental change the service will struggle on with an increasingly unhappy workforce and reduction in quality of service with reduced patient satisfaction. |
| What creates meaning that sustains GPs engagement with, and involvement in, work? | |
| What solutions/recommendations are proposed? | Reinvigorate the confidence and trust of the two main parties involved in primary care, that is, the primary care team and the patients. This means moving away from the current vogue of encouraging GPs to become ‘Leader-Managers’ and aiming to refocus on establishing a robust local primary care team. This will probably include a shift towards a salaried GP service, especially as (unsurprisingly) the vast majority of younger GPs are shying away from becoming principals. Once this structure is established, appropriate initiatives can be implemented to correct the problems with the current system, including:<br>• introducing new methods of working such as a separation of acute care (aligned with a quality out-of-hours service) from management of patients with long-term conditions or multiple comorbidities where the latter will truly benefit from continuity of care and longer consultations;<br>• reducing the ‘churn’ among practice staff who move on due to an unhappy working environment;<br>• encouraging improved working conditions and further career development options for GPs; and<br>• thus attracting doctors to work in general practice by witnessing happy GPs working in a pleasant working environment. |
| From which stakeholders’ perspective? (General practitioners, patients, other HCPs) | A locum GP |

| | |
|---|---|
| How is work-related joy constructed? (Diversity of ways individuals may think about joy) | |
| What prevents/removes work-related joy? | "The unsustainability of the current system has become more apparent over the last decade since the New Contract. The principle paradigm is one of data collection. Numerous other management diktats have also been imposed onto the primary care team trying to carry out the already difficult and messy job of frontline medicine. However, the imposed processes do not fit most of the time with the real world of clinical practice. The QOF may have improved evidence-based care standardisation in specific disease areas such as diabetes, but this model is not appropriate for the vast majority of patient contacts where the GP knows that the subtle nuances of 'whole person management' are the principle requirement. In addition, individual practices do not have enough time or a big enough management team to deal with the escalating bureaucratic requirements."<br><br>The vast majority of GPs probably want to focus primarily on the medicine that they were trained to do while allowing managers to manage. |
| Organisational characteristics (small, large) | |
| Managing ill-defined illnesses and chronic complex multimorbidity | |
| Human connection with patients | |
| Human connection with peers | |
| Human connection across other organisations (e.g., secondary care) | |
| Learning systems (e.g., competence, capability, mastery, socially situated) | |
| Additional useful information | |
| Summary of findings | The current model of general practice is unsustainable, principally due to data collection and other management data collection and other management diktats. A solution to this would be to reinvigorate the confidence and trust of the two main parties involved in primary care, that is, the primary care team and the patients |
| Strengths, limitations, and future research directions | |

| | |
|---|---|
| Paper Number | 163 |
| Author | Voorhees et al |
| Year | 2022 |
| Title | Accessing primary care and the importance of 'human fit': a qualitative participatory case study |
| Journal | BJGP |
| Region (England, Scotland, Wales, and Northern Ireland) | England |
| Study methodology | Interviews, focus groups and observation with service users and providers across general practice settings. |
| Type of document | Primary research |
| Topic or focus of paper | Access to primary care |
| Research objectives and focus | To fill the gap between the complex understanding of primary care access in the literature and the narrow definition of access assumed in UK policies. |
| Theoretical framework/theories/ underpinnings | Levesque et al's conceptual framework of patient centred access was applied. This juxtaposes five dimensions of accessibility of the healthcare system (approachability, acceptability, availability and accommodation, affordability, and appropriateness) with the abilities of patients to identify health needs, seek, reach, and utilise care. |
| Description of intervention | |
| Rationale for paper/review | While the academic literature on access to care emphasises its complexity, policies aimed at improving access to general practice in the UK have tended to focus on measurable aspects, such as timeliness or number of appointments. |
| What creates meaning that sustains GPs engagement with, and involvement in, work? | This study advances a modified concept of access as the 'human fit' between the needs and abilities of the population with the capacity and abilities of the healthcare workforce (see model below). This is in contrast to how access is normally framed by policymakers, in terms of whether you can get an appointment in a certain length of time. One GP explained: "I don't think the politicians appreciate any of that…they just see it as: you've got the flu, how quickly can you get to see your GP? Not that you need to see your GP with the flu, but that's their own direct experience of doctors, and they don't give any thought to the opportunistic stuff, the preventative stuff, the vulnerable people who don't actually ask for appointments, but need probably more care than most voting adults. And so yeah, there is just an impoverished debate around it. There's a lack of imagination about the true nature of the problem". |

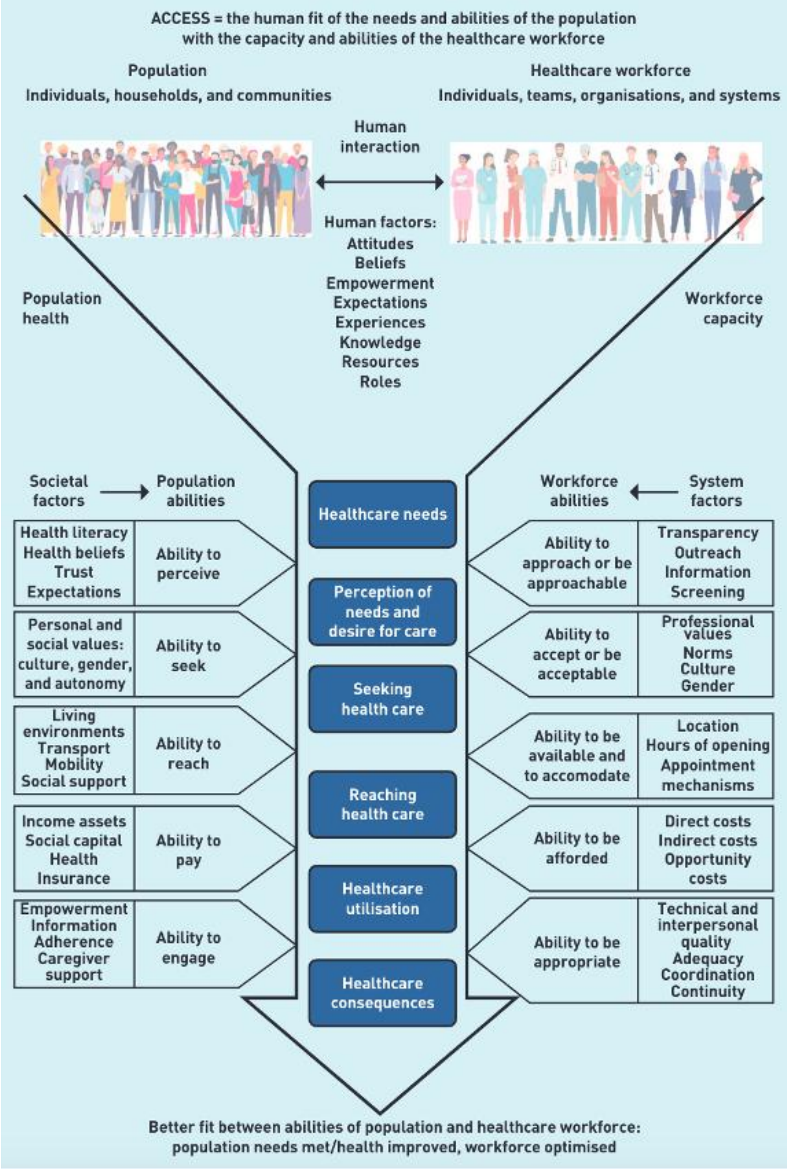
ACCESS = the human fit of the needs and abilities of the population
with the capacity and abilities of the healthcare workforce
Population
Individuals, households, and communities
Healthcare workforce
Individuals, teams, organisations, and systems
Human interaction
Human factors:
Attitudes
Beliefs
Empowerment
Expectations
Experiences
Knowledge
Resources
Roles
Population health
Workforce capacity
Societal factors
Population abilities
Workforce abilities
System factors
Healthcare needs
Perception of needs and desire for care
Seeking health care
Reaching health care
Healthcare utilisation
Healthcare consequences
Health literacy
Health beliefs
Trust
Expectations
Ability to perceive
Personal and social values: culture, gender, and autonomy
Ability to seek
Living environments
Transport
Mobility
Social support
Ability to reach
Income assets
Social capital
Health Insurance
Ability to pay
Empowerment
Information
Adherence
Caregiver support
Ability to engage
Ability to approach or be approachable
Transparency
Outreach
Information
Screening
Ability to accept or be acceptable
Professional values
Norms
Culture
Gender
Ability to be available and to accomodate
Location
Hours of opening
Appointment mechanisms
Ability to be afforded
Direct costs
Indirect costs
Opportunity costs
Ability to be appropriate
Technical and interpersonal quality
Adequacy
Coordination
Continuity
Better fit between abilities of population and healthcare workforce:
population needs met/health improved, workforce optimised

| | |
|---|---|
| What solutions/recommendations are proposed? | Politicians and policymakers can apply this understanding of access to convey a different goal for improving access to care: better human fit. Policies that take account of this conceptualisation would see continuity as an important contributor to better access, with attendant improvements in a wide range of outcomes. |
| From which stakeholders' perspective? (General practitioners, patients, other HCPs) | Service users: patients, carers, voluntary sector<br><br>Service providers: GPs, practice managers, receptionists, commissioners and other NHS roles |
| How is work-related joy constructed? (Diversity of ways individuals may think about joy) | |
| What prevents/removes work-related joy? | |
| Organisational characteristics (small, large) | |
| Managing ill-defined illnesses and chronic complex multimorbidity | |
| Human connection with patients | |
| Human connection with peers | |
| Human connection across other organisations (e.g., secondary care) | |
| Learning systems (e.g., competence, capability, mastery, socially situated) | |
| Additional useful information | |
| Summary of findings | This study advances a modified concept of access as the 'human fit' between the needs and abilities of the population with the capacity and abilities of the healthcare workforce (see model above). This is in contrast to how access is normally framed by policymakers, in terms of whether you can get an appointment in a certain length of time. Going forward, this should be accounted for by policymakers. |
| Strengths, limitations, and future research directions | The study's main strength lies in the application and advancement of a theoretical framework. Triangulation of methods provided a detailed understanding from multiple perspectives, and the participatory approach ensured that the research was grounded in and steered by the lived experiences of the local community. |

| | |
|---|---|
| | A potential limitation is that the study occurred in one area. A further limitation is the potential for social desirability bias, in that participants could respond positively to the access theory in order to please the researcher. |

| | |
|---|---|
| Paper Number | 164 – This paper summarises a paper published in the BJGP |
| Author | Abi Rimmer |
| Year | 2016 |
| Title | Reduce paperwork to retain young GPs, researchers say |
| Journal | BMJ |
| Region (England, Scotland, Wales, and Northern Ireland) | England |
| Study methodology | Survey of 143 GPs under the age of 50 who had left the English medical performers' list between 2009 and 2014. Telephone interviews were conducted with 21 of these GPs. |
| Type of document | Primary research |
| Topic or focus of paper | GP retention |
| Research objectives and focus | Investigate why so many GPs leave the NHS below the age of 50 years |
| Theoretical framework/theories/ underpinnings | |
| Description of intervention | |
| Rationale for paper/review | Many GPs leave the NHS below the age of 50 years |
| What creates meaning that sustains GPs engagement with, and involvement in, work? | |
| What solutions/recommendations are proposed? | To improve retention of young GPs, "the pace of administrative change needs to be minimised and the time spent by GPs on work that is not face to face patient care reduced." |
| From which stakeholders' perspective? (General practitioners, patients, other HCPs) | GPs who have left the NHS under the age of 50 |

| | |
|---|---|
| How is work-related joy constructed? (Diversity of ways individuals may think about joy) | |
| What prevents/removes work-related joy? | The negative portrayal of GPs in the media, a lack of time for interaction with patients and colleagues, and a perceived blame culture also contributed to young GPs leaving the profession |
| Organisational characteristics (small, large) | According to participants, continual organisational changes fundamentally altered their professional role to a 'government clerk' or a 'data clerk for public health and for management |
| Managing ill-defined illnesses and chronic complex multimorbidity | |
| Human connection with patients | Of the GPs that responded to the survey, 50% said that rising non-clinical workload was a factor in their decision to leave general practice early, while 84% said that pressure of work contributed to their decision. The higher administrative workload reduced the time available to spend with their patients, leading to a fundamental change in the doctor-patient relationship.<br><br>Tim Ballard, vice chair of the Royal College of General Practitioners: "With more and more of our working hours being taken up with form filling, ticking boxes, and preparing for CQC [Care Quality Commission] practice inspections, we are drowning in red tape and this only serves to keep us away from delivering frontline patient care, which is why we become doctors in the first place." |
| Human connection with peers | |
| Human connection across other organisations (e.g., secondary care) | |
| Learning systems (e.g., competence, capability, mastery, socially situated) | |
| Additional useful information | |
| Summary of findings | Rising non-clinical workload is leading to GPs leaving the NHS, as it reduces the time available for them to spend with their patients, and also causes increased pressures. Other factors include a perceived blame culture and a negative portrayal of GPs in the media. |
| Strengths, limitations, and future research directions | |

| | |
|---|---|
| Paper Number | 165 |
| Author | Robertson et al |

| | |
|---|---|
| Year | 2013 |
| Title | 'It's coming at things from a very different standpoint': evaluating the 'Supporting Self-Care in General Practice Programme' in NHS East of England |
| Journal | Primary Health Care Research & Development |
| Region (England, Scotland, Wales, and Northern Ireland) | England |
| Study methodology | Interviews with key stakeholders (including patients and GPs). Surveys of clinicians who had and had not completed the SSCiGP programme. Practice-based case studies in a number of practices that had completed the SSCiGP programme. |
| Type of document | Primary research |
| Topic or focus of paper | Self-care interventions in general practice |
| Research objectives and focus | To undertake a service evaluation of the NHS East of England Supporting Self-Care in General Practice programme. |
| Theoretical framework/theories/ underpinnings | |
| Description of intervention | The Supporting Self-care in General Practice (SSCiGP) programme seeks to transform the relationships between people with long-term conditions and health professionals. The programme is delivered by a team of specially trained tutors, actors and patient cofacilitators who lead three, 2-h workshops that are practical, skills-based and use a significant amount of action learning (role play) to focus on building effective clinical/patient partnerships that make the best use of limited consultation times. Participants developed the skills for supporting patient selfcare, based on the seven Common Core Principles of Self-care (Skills for Care: Skills for Health, 2008), in these three sessions and between through:<br>> Goal setting: setting personally relevant goals based on feedback from patients.<br>> Behaviour change in small steps: experimenting with skills in small, achievable steps in a safe environment that lead to successes.<br>> Skill rehearsal with feedback: receiving feedback and integrating that feedback into skill rehearsals with actors.<br>> Observational learning: seeing other clinicians successfully using the skills.<br>> Applying the skills in practice: trying out skills in the workplace and reflecting on those experiences. |
| Rationale for paper/review | The number of people purported to live with long-term conditions continues to rise generating increasing policy emphasis on the importance of self-care. NHS East of England Strategic Health Authority developed and commenced the Supporting Self-care in General Practice (SSCiGP) programme, which seeks to transform the relationships between people with long-term conditions and primary care practitioners. |
| What creates meaning that sustains GPs engagement with, and involvement in, work? | |

| | |
|---|---|
| What solutions/recommendations are proposed? | Practices that had completed the programme had implemented various changes to processes and structures that helped facilitate use of the SSCiGP skills within consultations. For example, longer consultations:<br><br>"You'll have noticed, we've introduced routine 15 minute appointments [in the morning surgery]. That's partly an acknowledgement of the impossibility of doing motivational work with patients within the constraints of conventional 10 minute consultations. I think that's certainly improved my professional quality of life because it's made it much more satisfying for me and I think from the patients' point of view, hopefully, it's better for them" – GP<br><br>Also, long-term disease clinics with longer appointments; telephone triage; virtual clinics for reviewing long-term conditions. |
| From which stakeholders' perspective? (General practitioners, patients, other HCPs) | Key stakeholders involved in the SSCiGP programme (including GPs) |
| How is work-related joy constructed? (Diversity of ways individuals may think about joy) | |
| What prevents/removes work-related joy? | |
| Organisational characteristics (small, large) | |
| Managing ill-defined illnesses and chronic complex multimorbidity | |
| Human connection with patients | Following the SSCiGP programme, clinicians talked about adopting a more empathic, patient-centred approach to consultations, being more mindful of patients' priorities, understanding more about why patients engage in behaviour which is potentially harmful to their health and appreciating this within the wider context of a patient's life. Following interviews with clinicians and patients, authors constructed a schematic diagram of the 'ideal consultation': |

<table>
<tr><td></td><td>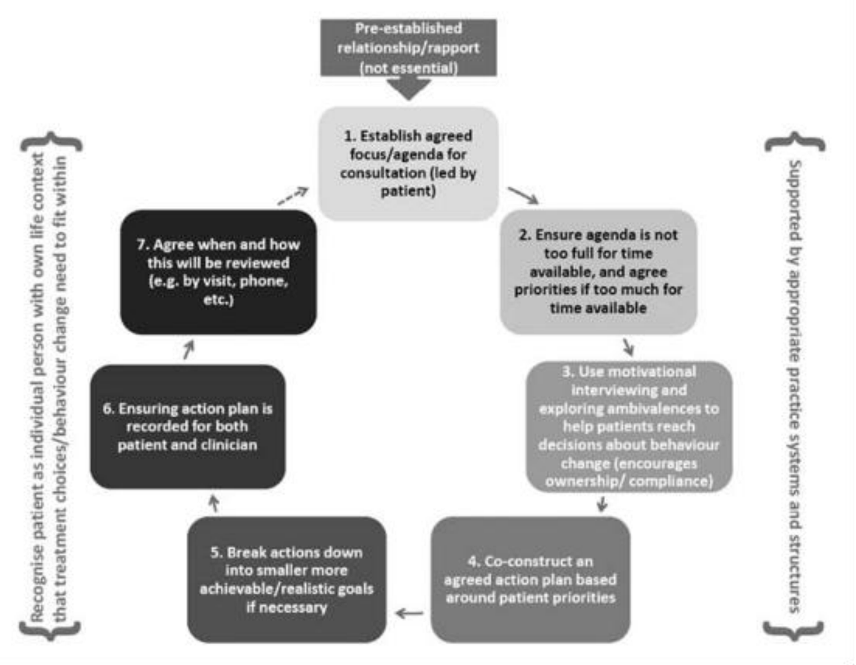

<br>However, clinicians noted barriers to implementing the learning (the ‘ideal’ consultation) from the programme into their consultations. These were: time, clinician reluctance fo alter consultation styles and wider practice systems and structures (2/3 of clinicians surveyed perceived that practice systems did not facilitate self-care).<br><br>“It’s really important for clinicians to have skills of keeping every individual patient at the centre of that consultation. It’s very easy as a clinician to tell the patient what they think they need to do or need to know because it’s in ten minutes – out. I guess it’s very tempting in a busy surgery to deal with the problem by telling the patient what to do, what to decide, what to take, or how to behave, rather than putting the patient at the centre” – Other practice staff</td></tr>
<tr><td>Human connection with peers</td><td></td></tr>
<tr><td>Human connection across other organisations (e.g., secondary care)</td><td></td></tr>
<tr><td>Learning systems (e.g., competence, capability, mastery, socially situated)</td><td></td></tr>
</table>

| | |
|---|---|
| Additional useful information | |
| Summary of findings | Following the SSCiGP programme, clinicians talked about adopting a more empathic, patient-centred approach to consultations (the ‘ideal’ consultation). However, time pressures and wider practice systems pose barriers to the implementation of this consultation style. |
| Strengths, limitations, and future research directions | Caution needs to be applied following this evaluation. The data presented are all self-reported and is not therefore a direct measure of whether changes actually did occur or, importantly, if they did whether they were sustained. |

| | |
|---|---|
| Paper Number | 166 |
| Author | Rosen et al |
| Year | 2022 |
| Title | Clinical risk in remote consultations in general practice: findings from in COVID-19 pandemic qualitative research |
| Journal | BJGP Open |
| Region (England, Scotland, Wales, and Northern Ireland) | UK-wide |
| Study methodology | Semi-structured interviews and focus groups, with a total of 176 clinicians and 43 patients. Data were analysed thematically, taking account of an existing framework of domains of clinical risk. |
| Type of document | Primary research |
| Topic or focus of paper | Remote consulting |
| Research objectives and focus | To develop an empirically based and theory-informed taxonomy of risks associated with remote consultations. |
| Theoretical framework/theories/ underpinnings | Data were organised around a clinically focused framework developed by the Royal Australian College of General Practitioners (RACGP), which defines risk as ‘anything that threatens a practice team’s ability to achieve its clinical objectives’. The RACGP defines the following five domains of general practice to which risks may relate: communication and the patient—doctor relationship; applied professional knowledge and skills (including all aspects of clinical performance); population and public health (including preventive care and societal influences on health); professional and ethical role (including the clinician’s duty of care, their own wellbeing, and the responsibility to keep up to date); and organisational and legal dimensions (including information technology, privacy and confidentiality, and practice management). |
| Description of intervention | |
| Rationale for paper/review | The COVID-19 pandemic-related rise in remote consulting raises questions about the nature and type of risks in remote general practice. |
| What creates meaning that sustains GPs engagement | |

| | |
|---|---|
| with, and involvement in, work? | |
| What solutions/recommendations are proposed? | Design and delivery of services<br>• Provide training for all staff to identify patients' ability to engage with remote consulting and offer appointments according to these abilities • Include patients as 'co-designers' of digital services and/or processes<br>• Maintain varied access routes into general practice, including in-person attendance to book appointments<br>• Support digital inclusion and preserve equity of access through actions such as peer-to-peer teaching provided by patient participation groups or signposting to local training in digital skills<br>• Invest in digital infrastructure to reduce the risk of failed or disrupted consultations<br><br>During consultations<br>• Use remote consultations as one of several different modes to engage with patients in their individual contexts and swap between them when necessary and possible<br>• Pay attention to screening, preventive care, and lifestyle advice while consulting remotely<br>• Use training and guidance to build clinician skills in identifying and managing safeguarding concerns<br>• Develop robust quality and safety assurance processes for remote consulting<br><br>Supporting patients and staff<br>• Improve communications to patients about how to access online services, including what type of consultation works best for different health problems and the principles to be applied when choosing between different types of consultation • Provide training and guidance for all clinicians (see above) on how to use digital modes of access to general practice<br>• Develop the role of care navigators and social prescribers to support highly vulnerable patients and others to access services and to navigate between providers • Work with external organisations to strengthen digital skills in vulnerable groups and to reduce digital exclusion<br><br>Addressing the needs of the wider population<br>• Monitor use of other services and onward referral rates following remote consultations<br>• Ensure that |
| From which stakeholders' perspective? (General practitioners, patients, other HCPs) | Clinicians (including GPs) and patients involved in delivering remote consultations. |
| How is work-related joy constructed? (Diversity of ways individuals may think about joy) | |
| What prevents/removes work-related joy? | |

| | |
|---|---|
| Organisational characteristics (small, large) | Risks associated with remote consultations:<br>1. Practice set-up and organisation: sufficient appointments are available, Care pathways are tortuous and involve double- handling, Patients are unable to access care (including various kinds of digital exclusion), Patients choose not to access care (for example, because they do not think they are a priority), Technology is inadequate or breaks down<br>2. Communication and the clinical relationship: Information exchange is inadequate in both content and tone, Consultations are overly transactional, with important concerns unsurfaced and loss of caring routines, The therapeutic relationship is not established or becomes eroded<br>3. Quality of clinical care: Diagnoses are missed or delayed (for example, because physical examination is limited or impossible), Safeguarding is compromised (for example, through lack of privacy or inadequate information), Patients are over- investigated or over-treated to compensate for information deficits<br>4. Patient's role in own care: Excessive burden is placed on the patient to make judgements, navigate care pathways, convey their symptoms, monitor their own illness, and use equipment, Opportunities for patient education and information sharing are reduced<br>5. Population and public health: Opportunities for screening and lifestyle advice are reduced, Opportunities to understand and engage with the societal and family context of illness are reduced<br>6. Professional development and wellbeing: Clinical staff become stressed, burnt out, and demotivated, Opportunities for learning and development are reduced<br><br>Illustrative quotes include:<br>Domain (1): "There was a patient who phoned with abdominal pain and fever and she couldn't speak English very well. So, we eventually got a ... we got an interpreter to do a three-way conversation who spoke to her over the phone, navigated her how to get into Near Me and then we did a video assessment of her at home. She had a ... I think one of her family members was there who was able to do some self-examination or directed examination for us to feel her stomach . And it was very time consuming, but it ended up ... we ended up admitting her as well through the ... using Near Me." – GP<br>Domain (2): "... And I think it's all a wee bit false and artificial ... on the video ... probably a wee bit more conscious of myself and my body language and all that sort of stuff. And that there's less ... I think there's less informal chats." – GP<br>Domain (3): "[On the text or email] maybe a vague symptom that doesn't really allow you to respond ... and it sort of forces you to phone up and say "well, actually sorry I didn't quite follow what you're saying about that tummy ache", and then you realise that actually there was a bit more going on there and you didn't respond to that with that text about the eczema because that's not really what they wanted to talk about, it's that hidden agenda thing. That is no small part of what we do as GPs and it's all a bit unspoken and difficult to quantify." – GP<br>Domain (3): "You were dealing with so many unknowns. You were having to deal with patients remotely. Patients that you haven't seen face-to-face. The uncertainty, the unknown level, the stress level was quite high. And it did worry me about the medicolegal issues. The medicolegal risks ... I think that if you ask most people, they were prescribing more than they would normally do. Because you haven't seen the patient you are going to prescribe amoxycillin antibiotics for a cough in case it's a chest infection. It might not be a chest infection, but if it is and you don't prescribe, and they go on to have pneumonia then that's not good." – GP<br>Domain (6): "I'm trying to try my best in my consultations, I'm really focusing. I'm really tired from listening on the phone, because it is exhausting. I'm really tired from these Zoom meetings, because it's very different to have this sort of like overload of, you know, concentration and physically trying to do it, overload of information." – Nurse |

| | |
|---|---|
| | Domain (6): "... the partners were saying, you know, "try to stay in your room at work as much as possible" ... So it was, you know, I don't want to spread anything, I don't want to get anything. So I'd come into ... and I just stay in my room ... And then you feel more isolated because you're not getting that general little chat about a case or ... about how things are." – Nurse |
| Managing ill-defined illnesses and chronic complex multimorbidity | |
| Human connection with patients | |
| Human connection with peers | |
| Human connection across other organisations (e.g., secondary care) | |
| Learning systems (e.g., competence, capability, mastery, socially situated) | |
| Additional useful information | |
| Summary of findings | Risks of remote consultations were identified across the following domains: Practice set-up and organisation, Communication and the clinical relationship, Quality of clinical care, Patient's role in own care, Population and public health, Professional development and wellbeing. |
| Strengths, limitations, and future research directions | The main strength of this study is that the combination of the three different primary studies provided a large and diverse dataset of interviews, focus groups, free-text responses and field notes representing a breadth of experience of general practice clinicians, patients, and support staff from across the UK. The main limitation is that because of the COVID-19 pandemic, the authors were unable to observe or record consultations directly, so can only report on risks as recounted by participants. |

| | |
|---|---|
| Paper Number | 167 |
| Author | Spiers et al |
| Year | 2017 |
| Title | Barriers, facilitators, and survival strategies for GPs seeking treatment for distress: a qualitative study |
| Journal | BJGP |
| Region (England, Scotland, Wales, and Northern Ireland) | [Not specified] |

| | |
|---|---|
| Study methodology | Interviews with GPs who self-identified either as had experienced or had not experienced mental distress. Responses analysed thematically. |
| Type of document | Primary research |
| Topic or focus of paper | GPs seeking treatment for distress |
| Research objectives and focus | To establish what might help or hinder GPs experiencing mental distress as they consider seeking help for their symptoms, and to explore potential survival strategies. |
| Theoretical framework/theories/ underpinnings | |
| Description of intervention | |
| Rationale for paper/review | GPs are under increasing pressure due to a lack of resources, a diminishing workforce, and rising patient demand. As a result, they may feel stressed, burnt out, anxious, or depressed. |
| What creates meaning that sustains GPs engagement with, and involvement in, work? | |
| What solutions/recommendations are proposed? | There needs to be a cultural shift within the medical profession and the NHS, so that the ongoing stigma of mental illness is confronted, making it easier for GPs to recognise and speak out about their difficulties before they reach such a crisis.<br><br>The authors recommend that individual practices, the RCGP, deaneries, clinical commissioning groups, LMCs, and the Department of Health do more to advertise specialist mental health services, such as the new NHS GP Health Service. The authors also suggest that GPs are offered specific raining about how to effectively support and treat their GP and other medical colleagues when they attend as patients. Finally, the authors recommend that researchers and policymakers investigate and develop organisational changes that will enable the workload stresses experienced by GPs to become more manageable for individuals.<br><br>One surgery disseminated helpful information about services for GPs: “my practice has just recently produced a document, like an A4 document, with online help for GPs and the local support agencies. So they’ve just laminated that, and they’re just putting it in the doctors’ runs…I think probably having something there and a number makes it a bit more accessible”<br><br>Participants talked enthusiastically about the benefits of going to a specialist, confidential treatment facility: “people who are treating doctors need to treat them like patients. That is a very skilled job. Because it’s so easy to collude, it’s so easy to see your stuff in them. It’s an incredibly hard job to do well. And so it needs to be specialised.” |
| From which stakeholders’ perspective? (General practitioners, patients, other HCPs) | GPs who self-identified either as had experienced or had not experienced mental distress |

| | |
|---|---|
| How is work-related joy constructed? (Diversity of ways individuals may think about joy) | Survival strategies:<br>Cutting down or varying work content: “I’ve actually always worked part time since I’ve been a GP. And I think that is the essence of good mental health. I don’t think people should be allowed to work full time really”<br>Locuming: “You pretty much don’t get any admin, so it’s all just see patients, go home on time”<br>Longer consultations: “Patients booked in at 15-minute intervals. Which is bliss”<br>Portfolio career: “I worked at the Sally Army helping the homeless with drug problems…And that was nice, because a day a week I was doing that…If you still want to work 4 or 5 days a week, you can. But if for one or two of those days you’re not going just GP-ing, it’s a break”<br>Asserting boundaries: “Establishing boundaries, understanding what your limitations are, you know, saying no, just not for the sake of saying no, but learning to say no and don’t feel guilty”<br><br>One surgery disseminated helpful information about services for GPs: “my practice has just recently produced a document, like an A4 document, with online help for GPs and the local support agencies. So they’ve just laminated that, and they’re just putting it in the doctors’ runs…I think probably having something there and a number makes it a bit more accessible” |
| What prevents/removes work-related joy? | Work related barriers and facilitators were as follows:<br>Participants talked about a perceived need to go to work, even when unwell, as a barrier to seeking support. Such ‘presenteeism’ was compounded by workforce shortages and difficulty accessing locum cover, in addition to guilt at letting down patients and colleagues: “I fought and fought and fought not to go off sick, because the guilt associated with going off sick is so huge…everybody’s struggling, so if I go off sick, I will just make everybody else struggle even more. But it got to the point where it was making me quite ill”.<br>Several participants perceived that there was a specific stigma attached to being a GP experiencing such ill health: “there is a sort of an unwritten rule that you don’t admit to mental illness in any way in general practice, I think. Even though everybody knows that everybody has it, it’s just not talked about”.<br>Many feared their patients might find out about their mental health concerns: “If you went to like a counselling group, you’d be worried you’d bump into one of your patients.”<br>Also prevalent were concerns around the impact of relationships with colleagues on confidentiality: “I knew all these GPs in [location], and it was a close-knit community. And, suddenly, all my colleagues that were training to be GPs were training in all the different practices in [location], which potentially had – could have – wherever I was registered would have access to my notes and know exactly what was going on with me”<br>Participants found lack of time to be an impediment to both help-seeking and self-care: “Never mind reflective practice and Balint groups and all of this, people are just, you know, nose to the grindstone absolutely all of the time”<br>GPs talked about the difficulty of moving between doctor and patient identity to accept help: “Whether it’s being a doctor as a patient that’s difficult, or that fact that you’re being a patient and seeing the room from, you known, a very familiar situation from another viewpoint”.<br><br>Access to support:<br>Participants reported a perceived inadequacy of psychological treatments as a difficulty when they were seeking effective help: “I saw her for the first couple of sessions, and then she said: “Oh, I can’t see you for another 2 weeks.” And I was quite suicidal at the time” |

| | |
|---|---|
| | One found that his status as a GP appeared to negatively impact the quality of help he received: "She [GP] said:"Well, what do you do?" "I'm a GP." "Well, we're all stressed, aren't we?" was the answer.'<br>A good relationship with one's own GP helped secure effective treatment<br>Some participants felt unsure of where to turn if they decided to seek help: "I did a burnout score around that time, on the RCGP website, and it said "Oh, you're at risk of burnout." And then it had no suggestions of what to do" |
| Organisational characteristics (small, large) | "My role as senior partner, oddly enough, had been just to sort of keep my head down and keep going. So if anything happened to me, it sort of destabilises them and then they would react very badly to the fact that I was off". |
| Managing ill-defined illnesses and chronic complex multimorbidity | |
| Human connection with patients | |
| Human connection with peers | Participants discussed how openness around distress could ease help seeking: "I have got a colleague here with mental health problems. And I have to say, she's fantastic. She's absolutely brilliant about telling you how she is, how she's feeling, medication she's taking. And that degree of openness is really refreshing."<br><br>"I'd said how stressful I'd been to these two female partners of mine, and we'd had a meeting and talked about it. And they tried to find some ways of helping. I said things like, it would be nice, if I get lots of extras, if someone from the staff could make a cup of tea?" |
| Human connection across other organisations (e.g., secondary care) | |
| Learning systems (e.g., competence, capability, mastery, socially situated) | |
| Additional useful information | |
| Summary of findings | Barriers and facilitators were related to work, stigma, and symptoms. Specifically, GPs discussed feeling a need to attend work, the stigma surrounding mental ill health, and issues around time, confidentiality, and privacy. Participants also reported difficulties accessing good-quality treatment. GPs also talked about cutting down or varying work content, or asserting boundaries to protect themselves. Systemic changes, such as further information about specialist services designed to help GPs, are needed. |
| Strengths, limitations, and future research directions | The authors interviewed a range of GPs from different geographical areas and in different roles (that is, partners, salaried, or locums), offering a balanced view of GPs in need of effective support.<br><br>A potential limitation is that the sample was self-selected, meaning the authors spoke to GPs who felt able to communicate with them. It may be that the most vulnerable GPs were too distressed to respond to advertisements or feel able to talk. |

| | |
|---|---|
| Paper Number | 168 |
| Author | Polly Morland |
| Year | 2022 |
| Title | A fortunate woman: A country doctor's story |
| Journal | N/A (book) |
| Region (England, Scotland, Wales, and Northern Ireland) | Southwest England – Gloucestershire valley. |
| Study methodology | |
| Type of document | Primary research |
| Topic or focus of paper | This book highlights the challenges and the joys of general practice (continuity of care, human connections, systems that enable personalised care). |
| Research objectives and focus | A country doctor's story by Polly Morland. |
| Theoretical framework/theories/ underpinnings | N/A |
| Description of intervention | N/A |
| Rationale for paper/review | When Polly Morland is clearing out her mother's house, she finds a book that will lead her to a remarkable figure living on her own doorstep: the country doctor who works in the same remote, wooded valley she has lived in for many years. This doctor is a rarity in contemporary medicine – she knows her patients inside out, and their stories are deeply entwined with her own. In *A Fortunate Woman*, with its beautiful photographs by Richard Baker, Polly Morland has written a profoundly moving love letter to a landscape, a community and, above all, to what it means to be a good doctor |
| What creates meaning that sustains GPs engagement with, and involvement in, work? | Long-term therapeutic relationships with patients/families:<br><br>"Where the scalpel is the essential instrument of the surgeon, so the relationship is the instrument of the GP."<br><br>"Young doctor relished these longer relationships, pieced together over numerous encounters."<br><br>"She is a doctor who knows her patients. She is the keeper of their stories. Over years and across generations, witness to the infinite variety of their lives. These stories she says are what her job is all about. They are what sustains her even in days as hard as these." |

| | |
|---|---|
| | "My patients trust is more important to me than anything I can think of."<br><br>Accumulating knowledge about patients and communities:<br><br>"Moments of light that flow from knowing your community and knowing your patients, and from connecting the two." |
| What solutions/recommendations are proposed? | |
| From which stakeholders' perspective? (General practitioners, patients, other HCPs) | GP |
| How is work-related joy constructed? (Diversity of ways individuals may think about joy) | (continuity of care, human connections, systems that enable personalised care). |
| What prevents/removes work-related joy? | |
| Organisational characteristics (small, large) | |
| Managing ill-defined illnesses and chronic complex multimorbidity | Intellectual stimulation and variety: "No two patients, no two people are ever the same. Pay attention to this above all and it will not just make you a nicer doctor, but a better doctor." |
| Human connection with patients | Long-term therapeutic relationships with patients/families: "Where the scalpel is the essential instrument of the surgeon, so the relationship is the instrument of the GP."<br><br>"She is a doctor who knows her patients. She is the keeper of their stories. Over years and across generations, witness to the infinite variety of their lives. These stories she says are what her job is all about. They are what sustains her even in days as hard as these."<br><br>"[patient] he'd follow her (GP) to the end of the earth now."<br><br>"For their extends behind every encounter the doctor has, on every day of her working life, not just a medical history, but also a personal history. A winding corridor of experience and emotion, the patient's whole life. In any given consultation, she's afforded but a fragment of that history. It is one of the joys of being a long serving family doctor in a community like this - that she has both time and opportunity to piece the fragments together over years and across families and generations." |

| | |
|---|---|
| | "This is what general practice is all about - talking to people, listening to their stories every bit as much as it's about clinical examination."<br><br>"On days when the administrative paperwork piles high, these stories are what keeps her going. They are the raw material of her relationship with each patient and a source of endless complexity and fascination."<br><br>Reciprocal caring: mutual acts of compassion: "The simple symmetry of help given and help received." "… who bakes sponge cakes for neighbours and give their GP a tin of quality street at Christmas." |
| Human connection with peers | |
| Human connection across other organisations (e.g., secondary care) | |
| Learning systems (e.g., competence, capability, mastery, socially situated) | "That willingness to invest in the moment, that instinctive ability to occupy, reflect, and respond to the here and now, both emotionally and intellectually, sits at the heart of the doctor–patient relationship as it plays out in general practice." |
| Additional useful information | |
| Summary of findings | This book highlights the challenges and the joys of general practice (continuity of care, human connections, systems that enable personalised care). |
| Strengths, limitations, and future research directions | |

The comprehensive report on the ReGROUP study comprises a collection of mostly self-contained workstreams and contains too much information for extraction into a single form. Consequently, separate data extraction forms were generated for each workstream. Please see below.

Paper number: 47

Author: Campbell et al

Year: 2019

Title: Policies and strategies to retain and support the return of experienced GPs in direct patient care: the ReGROUP mixed-methods study

Journal: Health services and delivery research

Region: England

Study methodology: (1) Systematic review of factors affecting GPs' decisions to quit or take career breaks, (2) census survey of GPs in south-west England[1], (3) interviews with GPs identified intending to quit from the census, and subsequent thematic analysis of responses, (4) thematic analysis of Care Quality Commission practice report data to explore examples of good and poor practice, (5) RAND/UCLA Appropriateness Method, with a panel of GP partners and GPs working in national stakeholder organisations, to prioritise emergent policies and strategies, (6) modelling framework to identify those practices at highest risk of facing a workforce supply–demand imbalance[2], (7) potential 'emergent' policies and strategies that aim to support the retention of GPs in direct patient care were road-tested in two stakeholder consultation meetings

[1]I have not extracted the results of the census survey as the results of this were also published in a separate paper, which I have extracted data from (number 82).

[2]I have not extracted the workforce modelling workstream as its findings are not relevant to our review.

Type of document: Primary & secondary research

Topic or focus of paper: GP retention

Research objectives and focus: (1) To identify the key policies and strategies that might (i) facilitate the retention of experienced GPs in direct patient care or (ii) support the return of GPs following a career break. (2) To consider the feasibility of potentially implementing those policies and strategies.

Rationale for paper/review: UK general practice faces a workforce crisis, with general practitioner (GP) shortages, organisational change, substantial pressures across the whole health-care system and an ageing population with increasingly complex health needs. GPs require lengthy training, so retaining the existing workforce is urgent and important.

| | |
|---|---|
| Workstream | Workstream 1: Systematic review of factors affecting GPs' decisions to quit or take career breaks |
| Workstream methodology | Systematic review |
| Research objectives and focus | Investigate the question: 'what are the factors which affect GPs' decisions to quit direct patient care (including reducing their time commitment to it), take career breaks from general practice or return to general practice after a career break?'. |
| What creates meaning that sustains GPs engagement with, and involvement in, work? | Qualitative studies revealed that low levels of morale were attributed by GPs to reductions in the perceived value of GP work (with loss of identity), and changes in professional culture in relation to a range of aspects of work such as a more target- and standards-driven reward system, multidisciplinary team (MDT)-based working (yet, for some, paradoxically, also lone working/isolating culture), a more aggressive top-down managerial culture within the NHS, and more widespread norms and expectations for early retirement. Low levels of morale were also seen to be associated with a perceived lack of support from both government and political parties, and negative portrayals of GPs by news media. Morale was also closely linked with job satisfaction (or dissatisfaction), neglect of personal well-being/health and feelings about work–life balance. |
| What solutions/recommendations are proposed? | [extract appendix 23] |
| From which stakeholders' perspective? (General practitioners, patients, other HCPs) | N/A |
| How is work-related joy constructed? (Diversity of ways individuals may think about joy) | The qualitative studies examined the various ways in which GPs of different personalities either continue and cope. This theme highlighted the major importance of flexible working or working reduced hours (e.g. by becoming a locum) as a method of coping and regaining work–life balance and job satisfaction. For others, the adoption of alternative work roles outside general practice, often part-time, allowed use of and learning of other skills – either as relief and variety from working as a GP or, for some, as a potential alternative career option. |
| What prevents/removes work-related joy? | The 22 UK survey studies that focused on GPs' intentions to quit fully from general practice (e.g. early retirement), their reasons for actually quitting general practice/patient care or intending to reduce hours/ working part-time all revealed a recurrent and linked set of key job-related factors that are associated with leaving, intending to leave or reducing their hours devoted to patient care. They are workload, job (dis)satisfaction, work-related stress and work–life balance. The studies of the self-reported reasons for quitting general practice reveal much more detail within and beyond these reasons; for example, underlying problems of high workload appear to be issues relating to high clinical work hours, more demanding patients and perceptions of excessive paperwork/administration. In addition, job dissatisfaction (and perhaps also work-related stress) is now reported alongside undesirable changes in the NHS, excessive managerial duties and fear of making mistakes.<br><br>The survey studies revealed that, as expected, age was an important determinant of intending to quit or reduce direct patient care, with the most dominant association being that older GPs of both genders were more likely to quit fully or retire early.<br><br>The survey studies highlighted several barriers to GPs returning to work following a career break: negative perceptions about the changing job of being a GP, high workload, low job satisfaction, unsociable hours, excessive administrative work, and recurrent and unwanted changes in the way the NHS and primary care is organised, which now includes revalidation. |

| | |
|---|---|
| | One survey paper highlighted by the systematic review – Doran et al, 2014 – examined the reasons given by GPs for having left general practice: the expectations of my patients were too high, my income was falling, The remuneration was not enough for the amount of work I was doing, Work schedule incompatible with doing other things more important to me, I was getting burnt out, I no longer enjoyed being a GP, I felt a loss of autonomy and professional control, The work was too stressful, I was unhappy with changes to the role of the GP, My working hours were too long, The workload was unreasonably intense, I wanted to improve my work–life balance, The non-clinical workload was too high, I did not like the target-driven approach to patient care, I felt overworked, I did not like the media's attacks on the medical profession, The goalposts were being moved too often.<br><br>The qualitative studies examined by the systematic review revealed that many GPs are experiencing working as a GP as undoable and unmanageable owing to, among other things, high/increasing administrative workloads, high/increasing patient demand (both number of patients and their complexity and higher expectations), and a perceived lack of training and resources to cope with these pressures. |
| Organisational characteristics (small, large) | One survey study examined in the systematic review found that either small practices or larger than average practices are associated with a more common intention to quit.<br><br>The qualitative studies included in the systematic review examined the impact of organisational changes. The perceived key changes or factors under this theme were changes in referrals (both restricted opportunities to refer to secondary care and higher numbers of, and more complex, referrals from secondary care), a greater focus on targets and assessments, and fears about reaccreditation (including evidence that some GPs might retire early in order to avoid reaccreditation). Some of the organisational changes had imposed increased clinical and non-clinical responsibilities and work on GPs. Together, such changes were believed to have undermined some of the basic tenets and traditional expectations of being a GP, such as the doctor–patient relationship and having autonomy and control over one's clinical work. |
| Managing ill-defined illnesses and chronic complex multimorbidity | |
| Human connection with patients | |
| Human connection with peers | Qualitative studies highlighted possible tensions between older and younger GP partners (in the way practices are run, in major investment/refurbishment decisions or in relation to planning for partners retiring and needing new partners to buy out their share of a practice). |
| Human connection across other organisations (e.g., secondary care) | |
| Learning systems (e.g., competence, capability, mastery, socially situated) | |

| | |
|---|---|
| Additional useful information | The survey studies revealed that social deprivation of area or practice population was not associated with intention to quit in any of the three multivariable analyses that included it as a potential factor<br><br>The survey studies revealed that unlike the other ways in which GPs may quit practice, intentions to take a (temporary) 'career break' appear to be influenced more by a specific range of 'pull' factors than by negative 'push' factors to do with the job or workload. The main reasons GPs say they will be taking a career break are to work abroad, to have or look after children, or to engage in research or further study. |
| Summary of findings | For many UK GPs, four closely related job-related factors seemed to play a major part in decision-making about both early retirement and part-time working: workload, job (dis)satisfaction, work-related stress and work–life balance. However, there were clearly many other detailed factors involved for some GPs that either underlie these higher-level factors (e.g. health service reform fatigue or unsupportive practice partner relationships) or may combine to influence an individual GP's decision to quit general practice or reduce their hours devoted to it. |
| Strengths, limitations, and future research directions | In terms of involving relevant stakeholders in the review, several GPs were involved in the development of the review protocol, and two GPs were closely involved in the conduct of the review (a GP trainee, and an experienced GP and Professor of Primary Care).<br><br>Our judgements about the applicability of findings to GPs in the UK NHS in 2016 were based on subjectively weighing up information such as the age of the study data, the geographical scope of the study, whether or not it was limited to certain types of GP (e.g. older/younger, practice principals/non-principals) and whether or not there were differences between respondent characteristics and target GP population characteristics. |

| | |
|---|---|
| Workstream | Workstream 3: Interviews with GPs |
| Workstream methodology | Interviews with GPs and stakeholders, followed by thematic analysis of responses. |
| Research objectives and focus | Identify the potential content of policies and strategies aimed at retaining experienced GPs and/or supporting the return of GPs following a career break. |
| What creates meaning that sustains GPs engagement with, and involvement in, work? | There was a perception of unfairness and a consistent reduction in rewards and rights for GP work. This affected morale deeply. Several GPs noted a relative decrease in their salary in recent years. GPs felt that they were asked to take on more and more responsibilities, but their pay was relatively less. This contributed to feelings of not being valued. "I think most people, if you ask them why they do jobs, it's a complex mixture and a lot of it comes about being valued and appreciated. I mean, people always focus on incomes and things but, the more detailed the analysis is, it always comes back to things like being appreciated, feeling valued".<br><br>Many GPs perceived their role as being less clear than, or as being different from, the one they had envisaged at the start of their career. |

| | |
|---|---|
| | There was an emotive split between GPs: although some were very saddened and upset at the loss of the 'traditional' model of general practice, others saw new models as being the only way forward. GPs were also divided in their opinions regarding new models of care, with some feeling that it was another unwanted result of a lack of government support for the current model of general practice.<br><br>Many GPs felt that their professional identity and ability to practise in their preferred ways were being compromised. This was considered to be partly due to unrealistic demands, concerns about complaints, targets and guidelines, the complexity of cases and lack of time to address them, a lack of continuity and loss of autonomy: "GPs tend to go down one of two routes: they either – to cope with demand – start to cut corners . . . Or . . . you overburden yourself and you won't cut corners . . . and that has its consequences at the end of the day". |
| What solutions/recommendations are proposed? | Long-term commitment and follow-through from the government and NHS to appropriately finance primary care.<br><br>Support to determine optimum practice size so that practices can benefit from economies of scale while retaining a supporting culture. GP support may be compromised above a certain practice size and personal lists (important factor in job satisfaction for some) difficult to maintain.<br><br>Support to determine optimum practice size so that practices can benefit from economies of scale while retaining a supporting culture. GP support may be compromised above a certain practice size and personal lists (important factor in job satisfaction for some) difficult to maintain<br><br>Evidence about how widening the practice team works in practice in the longer term, including the cost implications, and patient satisfaction with seeing non-GPs, could be helpful<br><br>Support and evidence to practices about the costs and benefits of different workload management methods, including shared learning from 'success stories' and good practice, could be helpful<br><br>Administrative support would be widely welcomed but with the recognition that there may be additional training requirements, and funding implications. Additional training for GPs could be helpful (e.g. managing telephone triage, business and HR issues).<br><br>A cap on the number of hours worked each day could help make direct patient care more sustainable. Longer appointment times are required owing to increasing complexity – especially if the more 'straightforward' care is dealt with by a wider workforce<br><br>Support for those who might otherwise leave could encourage experienced GPs to continue to contribute to the workforce, including supporting trainee/less experienced colleagues. Options need to be clearly signposted, making it 'easy' to stay.<br><br>Simplify and streamline return-to-work processes. |

| | |
|---|---|
| | Demonstrate support for primary care (e.g. Crown Indemnity), change in press/media culture towards reporting mistakes, etc. Provide a quick and supportive process for minor complaints. Help GPs to feel that they are valued, their opinions are respected, their views are taken into consideration and that the public, media and government support general practice<br><br>Address funding of primary–secondary care workload. Develop effective and closer communication between GPs and hospitals/consultants, and clarify the boundaries of responsibility. Address the ‘us and them’ feelings<br><br>Reduce the amount of work required by the appraisal and revalidation processes. Make appraisals and revalidation more supportive and less onerous<br><br>Provide occupational health resources, particularly related to mental health. These need to be accessible, confidential, anonymous and available proactively or in the early stages of a problem or crisis Supporting GP well-being could include: time for exercise, ability to take time off sick or for holidays when required, early-stage support for mental health, formal and informal sources of support.<br><br>Ensure government support and commitment to the future of primary care<br><br>Enable GPs to offer longer appointment times when necessary. Help manage patient demand and expectations<br><br>Support provision of continuity of care and support patients to take more ownership of their own health.<br><br>Introducing a fee system may increase patients’ perception of service value<br><br>Support to patients and appropriate signposting to help them access non-medical services (when appropriate) and to address lifestyle and social circumstances [which currently lead to (inappropriate) consultations with GPs]<br><br>Create a shared future strategic vision of general practice. This could help to reduce GP uncertainties and anxieties about the future, help manage patient demand and help GPs to feel more in control. Consultants’ expectations of primary care also need to be managed and the GP role clarified<br><br>Short-term incentives are unlikely to be effective in longer term. GPs need to feel ‘fairness’ in their remuneration<br><br>Ensure more support from GP colleagues within the workforce (e.g. between younger and older GPs; part-time and full-time GPs) |
| From which stakeholders’ perspective? (General practitioners, patients, other HCPs) | GPs and stakeholders (including practice-level roles and area-level roles) |

| | |
|---|---|
| How is work-related joy constructed? (Diversity of ways individuals may think about joy) | Several GPs described part-time and/or portfolio working as 'coping strategies' adopted to help them sustain working in direct patient care. There was a view that fewer GPs are now able and/or willing to work fulltime in a direct patient care role owing to the high demands and personal costs to GP well-being.<br><br>The enjoyable aspects of the job included GP–patient interactions. Enjoyment also came from 'being part of trying to figure out how the practice can deliver the best-quality care'. Being part of a team and having supportive relationships with colleagues added to this.<br><br>Seeing 'people with things you can make better quickly' creates enjoyment for GPs.<br><br>Four key coping strategies were identified by GPs: (1) reducing hours/number of sessions worked, (2) change in role (e.g. from partner or salaried GP to locum), (3) taking a career break, (4) portfolio working. |
| What prevents/removes work-related joy? | Although the potential value and benefits of targets, guidance and CQC inspection were recognised, the additional work created by these was burdensome. Many GPs strongly disliked CQC inspections (which they felt had a negative, punitive slant) and other aspects of their work that they considered to be 'pointless' or 'tick box exercises'.<br><br>There was almost unanimous agreement regarding workload increase, and increased length of working day, in combination with diminishing support services (e.g. mental health and social care for an ageing population).<br><br>Some activities that traditionally would have come under the secondary care remit had been passed to primary care. Some GPs felt that there was a lack of understanding from secondary care on the consequences of their actions/ requests. Transfer of care was not adequately recognised or resourced.<br><br>Participants noted that the 'buck [often] stops' with the GP.<br><br>GPs noted the implications of the current working situation on their own and other GP colleagues' mental and physical health and well-being. Some recognised that they did not have a good work–life balance; stress had affected sleep, and their health was being jeopardised. GPs described colleagues having periods of sick leave, some not returning to work, and being visibly exhausted. Participants recognised that there was then an increased risk of making a mistake. The impact of ageing was also discussed (including the effects of menopause); some GPs felt less able to carry out the demands of their role with increasing age, or expressed a desire to 'quit while ahead', and had noticed a reduction in their stamina.<br><br>It was noted that the cost of indemnity is increasing and becoming prohibitive, especially for part-time workers. Comparisons were made with other parts of the world (e.g. New Zealand where Crown Indemnity exists).<br><br>Those who considered or who had returned after a maternity break noted difficulties relating to childcare flexibility (especially for partners and locums). Those who were considering returning or had returned after a period working abroad noted difficulties with accessing reliable information and support for return, and also issues related to staying up to date and |

| | |
|---|---|
| | meeting the requirements to practise in the UK. Some GPs were reluctant to return 'in the middle of a workforce crisis', and comparisons of their working conditions overseas with returning to the UK were often unfavourable.<br><br>General practice has an inherent level of risk that is different from other aspects of medicine: "I'm the paid risk-taker for the NHS . . . with specialists, everything will be excluded but it will cost a fortune. If you see a GP, it will be cheap, but occasionally things will go wrong. And that's the nature of the risk". Risk was exacerbated by the perceived culture of patients being encouraged, and having easy routes, to complain, contrasted with a lack of support and little recourse for GP.<br><br>The fear of making mistakes led to some GPs practising more defensively (e.g. spending more time writing notes, and choosing face-to-face rather than telephone consultations), thus adding further to their workload.<br><br>Some participants had a pessimistic view of the future: "There seems to be a lack of belief that the NHS will survive, let alone GPs [as] part of it."<br><br>Older GPs who were partners were experiencing stress and anxiety owing to concerns about changes to practice mortgages, the threat of having to make staff redundant or practice closure, and responsibilities arising from joint civil liability for a practice.<br><br>Many GPs described a range of inter-related factors that had contributed to their decision-making to leave practice: factors relating to workload, their practice, their personal circumstances (e.g. menopause) and the wider social context (e.g. 'GP bashing'). GPs described how these factors accumulated and how each had a compounding effect, to create a 'perfect storm' situation over time that ultimately could lead to decisions to leave or to reduce their hours.<br><br>Erosion of resilience was linked to a loss of control: ". . . not feeling in control of where the money's coming from, not feeling in control of your future because if you're going to have contracts imposed on you by the government, you're not in control. So that's where I feel the loss of resilience is coming from, primarily." |
| Organisational characteristics (small, large) | Some GPs noted that 10-minute appointments were adequate if the practice was suitably staffed, and a personal list system employed. Some practices already worked to different (longer) appointment structures.<br><br>Some participants felt that widening the practice team was a reactive rather than proactive change: through necessity, GPs had recruited a wider clinical practice team in response to workload, recruitment and financial pressures. Although a diverse workforce sometimes reduced the workload of the GP, and was considered to be a positive adaptation or response to the changing workload and demands, this was not always the case. Other health professionals were sometimes considered to be an alternative solution to the 'ideal' of recruiting another GP. Some participants felt that additional non-GP staff could be expensive/not cost-efficient.<br><br>Several participants noted that some IT and systems that had been introduced to help manage workload or reduce GP tasks had, conversely, created additional work. They described how IT facilitated counting, measuring and 'double-checking' but that this, in turn, could lead to additional (unnecessary) work |

| | |
|---|---|
| Managing ill-defined illnesses and chronic complex multimorbidity | The wider practice team helping with 'quick win' consultations led to an increasing proportion of complex cases being seen by the GP, further exacerbating workload intensity.<br><br>A high number of patients combined with increasingly complex health needs were reported by almost all participants. The issue of a steep rise in costs, time and complexity associated with an ageing population and multimorbidity was discussed. Increased polypharmacy increased monitoring requirements. Clinical decision-making was more complex<br><br>There was general agreement that 10-minute appointment times were too short and based on outdated calculations. Complex patients and increased additional workload (targets, administration, etc.) meant that many consultations required more than 10 minutes. Reallocation of the 'easy' appointments to other team members also meant less opportunity for the GPs to use minutes saved from easier/ shorter appointments as 'catch up' time. However, there was also some concern that patients may 'fill up' longer appointment times if they were made available, irrespective of clinical need. |
| Human connection with patients | Expectations and demands of patients were identified as a considerable burden and source of stress for many GPs. This was complicated by feelings of diminished GP–patient relationships owing to lack of continuity of care, unrealistic patient demands and fear of litigation.<br><br>Participants talked about 'GP bashing' and the feeling that the media wanted only to report negatively about GPs rather than acknowledge the positive contribution that GPs can make. The media was also identified as having a role in encouraging unrealistic expectations from patients.<br><br>Defensive practice was also seen as a response to patient expectations, with one GP describing the feeling of 'I'd better give this person what they want, or they will complain!'.<br><br>When complaints had been made, or GPs had been sued, the process had been drawn out and stressful for GPs. As well as fear about the outcome of any complaint, it was noted that complaints 'wound the doctor severely . . . When you're kicked in the teeth like that, either by the government or the patient, it really hurts'. |
| Human connection with peers | Participants recognised that the appraisal system was designed to help, but the majority of participants found it to be an unhelpful or time-wasting process; only a minority reported positive experiences. Most felt that appraisal and revalidation were a 'missed opportunity' to provide support and could be more beneficial to the GP.<br><br>The need for opportunities to debrief or have supervision style support (similar to the support that counsellors receive) was identified, to help GPs 'carry the burden' of their role. However, it was noted that many GPs have concerns about confidentiality and stigma attached to accessing mental health support.<br><br>Participants described fewer opportunities for peer support activities (e.g. meeting for coffee during the working day). Colleague support was deemed important by most and vital for some. However, the pressured environment led to instances of bullying, conflict over work patterns and lack of understanding between colleagues (e.g. older and younger GPs). |

| | |
|---|---|
| | Tensions existed between partner, salaried and locum GPs.<br><br>Participants described concerns about the safety of practice and the quality of care being delivered to patients. This arose from having to manage imposed ‘unmanageable’ workloads, and focused primarily on the large number of decisions that had to be made, the complexity of cases and the impact of accumulative decision-making throughout the day.<br><br>Some described leaving practice for the benefit of their colleagues: “The worry is about being miserable around people who don’t need misery . . . Like I say, sever the gangrenous limb and you save the patient! And it’s funny feeling like that, I’ve never thought of myself as a gangrenous limb, but actually perhaps that’s what happens when you get older in a practice. That is coming to terms with the fact that the best thing you could do is leave so that actually you’re not polluting in any way.” |
| Human connection across other organisations (e.g., secondary care) | Many GPs had struggled with maintaining good communication between primary and secondary care and recognised issues relating to the distribution of funding. Communication pathways were seen as complicated and created opportunities for misunderstanding.<br><br>Many GPs had struggled with maintaining good communication with secondary care. Relationships had changed to become less supportive and less accessible. Participants recognised issues relating to the distribution of funding contributing to an ‘us and them’ mentality. |
| Learning systems (e.g., competence, capability, mastery, socially situated) | There was concern that regulations can devalue GP knowledge and experience, and deskill GPs – especially regarding problem solving, creating and testing hypotheses. This could lead to clinical over-investigation.<br><br>There was general agreement that ‘to survive in today’s NHS you have to be comfortable taking risks and cutting corners’ (SH1040 – CCG stakeholder). However, GPs reported tension between ‘cutting corners’ and being thorough and conscientious in their practice. |
| Additional useful information | |
| Summary of findings | This study identified three underlying themes that may influence GPs’ experiences and decisions about remaining in or leaving direct patient care, namely (1) identity and value (subthemes were: the identity of general practice as a profession lacks clarity and boundaries, GP identity, professionalism and morale, being listened to and being valued), (2) fear and risk (subthemes were: risk to patient care and safety, fear of complaints and being sued, risk to professional status and identity, risk to own health and well-being, uncertainty about the future of general practice, financial risk) and (3) choice and volition (subthemes were: accumulation, compounding and combinations of factors, GP resilience, decisions do not happen in isolation, the only route left). A very wide range of potential solutions are also described. |
| Strengths, limitations, and future research directions | Sampling GPs from the survey returns gave a large pool to draw from and also provided the opportunity to ensure a maximum variation sample.<br><br>Patient and public involvement and project team discussion enabled modification of the original sample targets to ensure that the views and experiences of ‘staying’ GPs were also captured. The PPI group and the GP representative supported the analysis process and reflective practice. |

| | |
|---|---|
| | The main limitation was that the GPs were self-selecting: they responded to the original survey and agreed to be contacted about possible interview, and they also consented to interview. It is possible that those GPs who were not available for interview may have had different experiences to report.<br><br>Of the survey returns (workstream 1), 93% of respondents were white and 96% of those eligible for interview were white. GPs from other ethnic groups were approached but none agreed to be interviewed.<br><br>It is also of note that all of those GPs interviewed and who were attached to one practice were from practices rated as ‘good’ or ‘outstanding’ by the CQC. |

| | |
|---|---|
| Workstream | Workstream 4: Thematic analysis of Care Quality Commission practice report data |
| Workstream methodology | Thematic analysis of 8 outstanding practices in the south-west of England. |
| Research objectives and focus | Use CQC reports of general practices to explore current examples of good practice in the south-west of England. |
| What creates meaning that sustains GPs engagement with, and involvement in, work? | |
| What solutions/recommendations are proposed? | |
| From which stakeholders’ perspective? (General practitioners, patients, other HCPs) | GPs |
| How is work-related joy constructed? (Diversity of ways individuals may think about joy) | |
| What prevents/removes work-related joy? | |
| Organisational characteristics (small, large) | Some practices purchased innovative technology to relieve some of the pressures on GPs. For example, one practice installed a health pod in the waiting room, which enabled patients to do blood pressure, weight, height and lifestyle checks. |

| | |
|---|---|
| Managing ill-defined illnesses and chronic complex multimorbidity | One practice reported giving patients with long-term conditions longer appointments of 40 minutes owing to the complexity of their conditions. |
| Human connection with patients | One high performing practice reported good continuity of care: "staff knew patients well and co-ordinated follow-up care and support to meet each person's needs" |
| Human connection with peers | GPs were concerned that they could not give patients with mental illnesses the time in their 10-minute slot, so they recruited a mental health nurse consultant. This had proved to be successful, and the mental health nurse consultant had improved faster patient access for mental health treatment and reduced patient referrals to secondary care. They had reduced GP workload. Patient feedback was positive and had attracted more patients to register at the practice.<br><br>One of the most important factors in supporting GPs with their work-life balance was a stable, supportive staff group. For example, nursing and administrative staff at one practice described being actively supported in their employment and described the practice as having an open, supportive culture and being a good place to work. GPs said that there was support for each other when it was identified as being needed. However, the CQC did not see the same level of proactive support or risk assessment given by GPs to themselves as a staff group |
| Human connection across other organisations (e.g., secondary care) | Practices reported working closely with the local community, such as signposting activities in the community such as singing groups, lunch clubs and well-being services. Practices collaborated with other practices in the area in order to share different ways of working and expertise. They also co-commissioned specialist services to relieve pressure on GPs – eg an over-75-year-olds nurse specifically to visit patients at home to carry out routine health-care checks and care planning. |
| Learning systems (e.g., competence, capability, mastery, socially situated) | "All call handlers and patient-facing staff had knowledge of all the GPs and their specific skills (e.g. dermatology, family planning, steroid injections). This awareness enabled them to allocate appointments or queries to the most appropriate GP."<br><br>All of the GPs had specialist interests and provided leadership and clinical governance for clinics for patients with long-term conditions and chronic diseases. For example, one GP held a master's degree in management of respiratory conditions and patients benefited from their expertise with this. Data for hospital referrals for patients with respiratory diseases was nearly 50% lower than the CCG and national averages. This showed that the practice was effective in managing long-term and chronic conditions |
| Additional useful information | |
| Summary of findings | Examples of good practice from the CQC reports include using technology and collaborative, community working to reduce pressure on GPs; giving patients with complex conditions longer appointment times; recruitment of 'new' staff with specialist skills; and the value of specialist interests in providing good quality care and reducing referrals. |
| Strengths, limitations, and future research directions | The main limitation of the data was that it had been collected and reported to meet the CQC's own requirements (the quality of care being delivered) and, consequently, did not focus sufficiently on our line of enquiry (the impact of care organisation and delivery on GPs). It is also important to note that CQC is a regulatory body, and, thus, the GPs may have been reluctant to share data relating to their own morale, resilience, etc. Finally, in order to deliver this briefing report within the time scales available, several pragmatic decisions had to be made, most notably around sampling and the selection of examples of good |

| | |
|---|---|
| | practice from all those available in the CQC reports. The use of researcher judgement may have affected the rigour and credibility of this process. |

| | |
|---|---|
| Workstream | Workstream 5: RAND/UCLA Appropriateness Method |
| Workstream methodology | RAND/UCLA Appropriateness Method |
| Research objectives and focus | Answer the following questions:<br>1. What are the potential policies and strategies that might facilitate the retention of experienced GPs in direct patient care and support the return of GPs to direct patient care following a career break?<br>2. How appropriate are these policies and strategies and how feasible are they to implement from the perspective of GPs? |
| What creates meaning that sustains GPs engagement with, and involvement in, work? | |
| What solutions/recommendations are proposed? | A total of 16 policies and strategy areas were deemed by consensus as both appropriate and feasible.<br><br>For implementation at a national/regional level:<br>1. General practices identified as being 'at risk' should be managed with an appropriate and sensitive supportive arrangement and this should be optional (uncertain for compulsory).<br>2. General practices identified as being 'at risk' should be allocated a specialist team for managing recruitment and retention, and this should be optional (uncertain for compulsory).<br>3. New incentive and support packages should be available to GPs and other organisations setting up new practices or new ways of working in under-doctored areas.<br>4. There should be a publicity campaign focusing on managing expectations of patients in line with the resources and constraints of GP-based primary care services.<br><br>For implementation at a general practice level:<br>1. GPs who are returning to work after a period of absence or after a career break should have access to 'health and well-being programmes' to help them manage their re-entry into the workforce. Engagement with such a course should be optional.<br>2. GPs who are returning to work after a period of absence or after a career break should have access to schemes that have a range of routes and options that can be combined in a personal package for re-entry.<br>3. GPs who are returning to work after a period of absence or after a career break should have access to schemes that use a mix of online education and face-to-face meetings to ensure timely access to induction and refresher courses.<br><br>For implementation at a GP level: |

| | |
|---|---|
| | 1. Peer support initiatives should be made available to GPs aimed specifically at health and well-being. Shown as appropriate specifically for those not near retirement age.<br>2. GPs should have access to their own specialised health-care service to ensure a quick and confidential occupational health-care service.<br>3. A structured programme of training and support should be made available to all GPs in their first 5 years following qualification as an independent GP to help them establish healthy, productive careers. Engagement with such a course should be optional (uncertain for compulsory).<br>4. GPs should consider portfolio working as part of their career pathway and this should be optional (inappropriate for compulsory).<br>5. Incentives and support packages should be available for those GPs developing portfolio careers who are linking their portfolio activities to specialisms/areas that are directly beneficial to local clinical priorities.<br>6. For GPs who are reaching retirement age and could take their pensions on exit, a comprehensive flexible careers scheme should be introduced with a view to supporting annualised hours, part-time working and/or ad hoc contributions to direct patient care.<br>7. For GPs who are reaching retirement age and could take their pensions on exit, the annual appraisal and revalidation process for such GPs should be reviewed with a view to streamlining and simplifying the process for such GPs who have not encountered any concerns in the previous revalidation/appraisal processes/for such GPs who would like to work with a specified and limited scope of practice.<br>8. For GPs who are reaching retirement age and could take their pensions on exit, the working hours of GPs should routinely include fully funded, dedicated time to accommodate the full range of roles (administrative, clinical, training, management, CPD, business) undertaken as part of care professional activity.<br>9. For GPs who are reaching retirement age and could take their pensions on exit, contracts based on specified programmed activities should be available to GPs to work across several general practices and on other health-related activities.<br><br>Three policies were deemed by consensus as appropriate but of uncertain feasibility:<br>1. Where a strong case can be made that there is a financial risk directly relating to the work of the practice (e.g. ownership of premises), GPs should have access to schemes to reduce financial burden (e.g. buy-back schemes for premises) (uncertain when applied to all GPs).<br>2. There should be an agreed maximum in the number of consultations that a GP should be allowed to conduct in a working day in order to protect patient safety as well as the health of the GP.<br>3. There should be contractual changes to encourage longer consultations when appropriate.<br><br>Five policies were deemed by consensus as appropriate but were not rated for feasibility:<br>1. General practices should be able to self-register their organisation's 'at-risk' status.<br>2. General practices identified as being 'at risk' should be provided with a toolkit to manage recruitment and retention.<br>3. To support flexible working and managed exits, general practices should implement strategically planned exits for retiring GPs. |

| | |
|---|---|
| | 4. Career support should be available to GPs to enable portfolio opportunities to be identified and taken up in a strategic way to inform their future ambitions for all GPs or specifically those who are not reaching retirement age and cannot take their pension on exit.<br>5. For GPs who are reaching retirement age and could take their pensions on exit, there should be financial incentives for such GPs who have maintained a prolonged/sustained period of direct patient care. |
| From which stakeholders' perspective? (General practitioners, patients, other HCPs) | GP partners and GPs working in national stakeholder organisations with a role in workforce planning. |
| How is work-related joy constructed? (Diversity of ways individuals may think about joy) | |
| What prevents/removes work-related joy? | |
| Organisational characteristics (small, large) | |
| Managing ill-defined illnesses and chronic complex multimorbidity | |
| Human connection with patients | |
| Human connection with peers | |
| Human connection across other organisations (e.g., secondary care) | |
| Learning systems (e.g., competence, capability, mastery, socially situated) | |
| Additional useful information | |
| Summary of findings | This consensus study conducted with senior GPs identified a range of policies to support the retention of GPs, at a national, practice and GP level. Many of these policies and strategies related to the provision of personal or professional support, such as 'health and well-being programmes' to support GPs who were returning to work, which aim to manage their re-entry into the workforce; flexible working; and targeting career stages in the first 5 years or when nearing retirement. When considered |

| | |
|---|---|
| | at a national level, there was recognition that early self-reporting of practice at-risk status in respect of retention issues was appropriate and feasible. |
| Strengths, limitations, and future research directions | A key strength of this study was the ability to present a large number of statements with multiple layers to establish a more refined set of policies and strategies for consideration at national and local levels.<br><br>The main limitations include the selection of just one professional group for the panel, the paucity of evidence regarding the effectiveness of policies and strategies aimed at retaining GPs within the workforce, the exclusion of skill mix/multidisciplinary working policies and strategies in the statements, and the use of appropriateness and feasibility as the rating scales. |

| | |
|---|---|
| Workstream | Workstream 7: stakeholder consultation |
| Workstream methodology | Two facilitated stakeholder group consultations which discussed policies recommended by the RAND/ULCA appropriateness method.<br><br>Discussions were structured as follows:<br>l. What factors could have an impact – positively or negatively – on the implementation of the emergent recommendation?<br>2. Please comment on feasibility and acceptability of the emergent recommendation.<br>3. What are the structures or groups that would need to be involved in introducing change to this area?<br>4. What are the three key next steps to move this agenda forward? |
| Research objectives and focus | Provide preliminary evidence on the feasibility and acceptability of implementing the policies and strategies prioritised by the RAND/ULCA appropriateness method. |
| What creates meaning that sustains GPs engagement with, and involvement in, work? | |
| What solutions/recommendations are proposed? | There were 11 discussion topics in three broad categories (derived from recommendations that emerged from the RAND/ULCA appropriateness method).<br><br>'Protection of' GPs and managing patient expectations<br>• Using marketing strategies to influence demand<br>○ Positive factors: Co-ordination of existing publicity campaigns regarding use of general practice and primary care services is needed; Supportive PPGs in general practices and in CCGs could have a potential role in any campaign that is aimed at educating patients or changing their help-seeking behaviour; There is a need for such publicity campaigns to target 'high-user' patient groups.<br>○ Barriers: There is currently no overall media strategy that allows for consistent, multichannelled sustainable messages that can be delivered both at national and local levels, without misinterpretation, which has been informed by and has buy-in of clinicians and staff at practice level; Small campaigns may work, but are likely |

| | |
|---|---|
| | to be unsustainable; GPs can inform/educate patients, but often patients cannot readily change their consulting behaviour owing to broader environmental factors and/or social constraints.<br>o Considerations & next steps: NHS England: A co-ordinated national media strategy is needed to support local GP delivery and should target high-user patient groups and take account of feedback from PPGs; A national brand would enable signposting and delivery of information to targeted patient groups (i.e. where/how to access services)<br>• Maximum number of consultations for GPs<br>o Positive factors: Setting a maximum number of consultations could be achieved (e.g. as per hospital outpatient clinics), potentially positioned as a quality and safety issue; Capping the number of consultations may negate 'supply-induced demand' [sic] (i.e. additional use of primary care services by patients as a result of increasing access); Not all consultations are necessarily for a GP – other health professionals can be used differently such that the overall number of appointments could be increased, which would allow a limit to be set per GP; Both the number and length of appointments need to be calibrated to individual patients' needs; It is a GP's duty to decide when/if he or she is unsafe to see patients.<br>o Barriers: Individual GPs have different consulting styles (longer vs. shorter consulters); achieving consistency of individual ways of working is challenging; Setting a maximum number of consultations per day may be less acceptable to patients; There will inevitably be a need for a 'pressure valve' service for occasions when demand exceeds the maximum number of consultations available. The nature, cost, capacity and quality of such services are of key importance to the success of setting a limit on the number of GP consultations; GPs may fear increased complaints from patients; There is uncertainty over whether or not patient safety is affected by the number or length of consultations, and the point at which care delivery becomes 'unsafe'; The content of consultations is key and can make a consultation more or less stressful for a GP; What would be the reaction of CCGs to setting a maximum number of consultations?; It would be challenging to square the aims of setting a maximum number of consultations with the fact that Sustainability and Transformation Partnerships involve shifting work to primary care.<br>o Considerations & next steps: NHS England/CQC/RCGP, CCGs: Consider a limit on the number of consultations being set at practice level (based on list size), rather than at GP level; Use NICE guidelines as a mechanism for setting safe staffing quotients for GPs as with nursing (on grounds of patient safety);Undertake stakeholder engagement with GPs to see if they would support the idea of setting a maximum number of consultations;Consider a system whereby patients seek reimbursement from state-run health insurance for their consultations<br>• Making consultations longer<br>o Positive factors: Containing a consultation within 10 minutes is a challenge, but longer consultations are likely to increase patient and GP satisfaction; One 15-minute appointment may comprise higher-quality care than two 10-minute appointments, and may prevent the need for follow-up; Certain patient groups need longer than the traditional 10-minute appointment than other patient groups; GPs need to be able to work in different ways to suit individual consulting styles – use of the wider health professional skill mix can help meet overall demand; Research evidence suggests that patients are fairly accurate in assessing how long they require if |

| | |
|---|---|
| | allowed choice of appointment length; Combining longer consultations with telephone triage and patient education may be the most effective strategy; Introducing longer consultations will inevitably have an impact on access and GPs' stress initially, but over time the length of a GP's overall working day is likely to be the same.<br>o Barriers: Offering many longer appointments would potentially affect access and patients' expectations; GPs fear that workload will increase within a longer consultation (supply-induced demand).<br>o Considerations & next steps: Practices – Analyse workload regarding the source/nature of the workload and patients' needs; Allow flexibility within appointment systems and categorise appointments in accordance with complexity (the electronic frailty index may help identify patients who need longer); Improve in-practice conversations about consultation planning. HEE – consider amending the Clinical Skills Assessment to reflect the need for GPs to develop skills to consult within a longer consultation time. Academic partners – participants suggested undertaking a UK study to provide evidence on offering longer appointments/choice of appointment length, including qualitative interviews with GPs about introducing such a system. NICE – guidelines could be a mechanism for bringing about change to the traditional length of consultations.<br><br>Incentives and support mechanisms for GPs<br>• The nature of a practice being 'at risk' and the process of identifying 'at risk' practices<br>o Positive factors: A practice's 'at-risk' status should be on a sliding scale (recognising that particular practices may become 'at risk' unpredictably and at short notice), rather than a binary, objective, stable state; Definition of 'at-risk' status could be considered at the town/regional level (i.e. a shared problem in an area), rather than individual practice level.<br>o Barriers: There were concerns/uncertainty over the implications of being 'at risk' regarding whether this status would be public or anonymous, and how long the status would last (i.e. when would it be reviewed); What data would be used to define 'at-risk' status (i.e. what balance of routine indicators vs. local knowledge/intelligence)?; Who would define 'at-risk' status (and is it most appropriate for a CCG to make the assessment)? Self-identification by a practice was not seen as viable/practical – if a practice would receive greater resources, assessment of 'at-risk' status would need to be backed up by external judgements and/or data.<br>o Considerations & next steps: Fresh approaches to the appraisal/ revalidation process are urgently needed (for GPs of all ages) to reduce burden and make appropriate to a GP's existing role; Introduce (to appraisal/revalidation) a compulsory interview with a trained GP career advisor, with special appraisal at age 50/55 years to give an informed career review of role options<br>• What support could or should be offered to 'at-risk' practices<br>o Positive factors: Support arrangements should tackle the root causes of organisational problems that are beyond issues relating to GP recruitment and retention, to include organisational development, training (e.g. team coaching), recruitment and retention of all practice team staff groups (Leeds) and assessment of clinical resources and the causes of vulnerability (London); Support would need to be tailored, locally delivered and flexible; It would be preferable if support was led by peers (possibly local or regional peers), rather than |

| | |
|---|---|
|  | external parties – it needs people who are knowledgeable about the nature and business of being a GP, including experienced practice managers; The processes and networks involved in identifying 'at-risk' practices should be linked to the provision of support.<br> o Barriers: What would be the scope of support for 'at-risk' practices?; There were concerns regarding whether information about practices receiving support 'stays local' (e.g. within CCG) or if it is shared nationally.<br> o Considerations & next steps:<br>• External HR interventions and monitoring/support<br> o Positive factors: Careful management of a transition to using external HR would be needed for it to be accepted and helpful; External HR arrangements would need to be sustainable, to fit in with current and future service reforms and to be supportive and not proscriptive; Use of external HR systems may be better suited to vulnerable practices.<br> o Barriers: There was hesitancy/reluctance among stakeholders regarding the use of external HR support; Practices (particularly GPs and practice managers) need to be involved and in control of HR processes; GPs and practice staff could be better supported to manage HR processes internally; There was no overall consensus on whether or not the use of external HR is a 'good idea'.<br> o Considerations & next steps: If HR processes are managed externally, a centralised, consistent, sustainable model would need to be available; If HR processes are managed internally, training and support should be available to GPs and practice managers<br>• Supporting successful implementation and uptake of health and well-being interventions for GPs<br> o Positive factors: Priorities in this area are to raise awareness of health and well-being interventions and to ensure their confidentiality; Wider support structures can affect GPs' health (e.g. career advice, RCGP roles).<br> o Barriers: Occupational health can be seen as a 'sticking plaster' solution that does not deal with underlying causes of problems; Bureaucracy around new schemes and initiatives acts as a disincentive; new schemes and initiatives are seen as 'bombs' that are created by one group and that are swiftly passed to GPs to defuse/manage; Informal support networks (such as practice teams/GP colleagues) are being eroded under current working conditions.<br> o Considerations & next steps: Those introducing new schemes and initiatives: Aim to reduce bureaucracy and increase awareness of existing support schemes among doctors and practices; Clarify information/understanding about confidentiality between a doctor and health and well-being intervention to ensure transparency and rules of engagement. Practice teams – should create clear space during the working day for 'headspace' and support (e.g. regular morning coffee gathering). National and professional groups (e.g. the RCGP and RCN) – health and well-being schemes should consider support for other clinicians who are part of the MDT.<br>• Professional support in the first 5 years of career<br> o Positive factors: Professional support (peer to peer) can help with recruitment and reduce isolation; Examples of a model for support include the Practice Based Small Group Learning approach, led by a trained mentor, and the HEE Yorkshire & Humber Nurse Preceptorship Programme; Objectives for professional support need to be set but kept informal; such approaches should not be mandatory but kept as an 'offer'; Use of virtual |

| | |
|---|---|
| | meetings/communication to deliver professional support could be explored in cases in which participation rates among GPs might be lower.<br>o Barriers: Consideration needs to be given to who could deliver various components of professional support and the practicalities of fitting this in with other commitments (e.g. pastoral care does not necessarily require delivery by doctors).<br>o Considerations & next steps: CEPNs/HEE training hubs – should co-ordinate and link with current RCGP First5 activities and responsibilities. RCGP – should embed professional support in the first 5 years and make pastoral/peer-to-peer care an important part of training.<br>• Support and planned exits for GPs nearing retirement age/implications of losing previous pension incentives to stay in practice<br>o Positive factors: To enable GPs nearing retirement to want to continue working, job satisfaction needs to be increased and end-career role options need to be professionally fulfilling; There may be particular role(s) that would persuade a GP to remain in direct patient care and certain elements of work (such as some elements of routine practice administration) from which these experienced GPs might be excused. Alternatively, other support mechanisms might be put in place (such as extended administrative/secretarial support) in order to keep GPs within the workforce; Simplifying the appraisal/revalidation process for this group of GPs is essential; its benefits are far outweighed by the bureaucracy involved.<br>o Barriers: Retaining these GPs through financial incentives was felt to be controversial.<br>o Considerations & next steps: The NHS regional offices were highlighted as the organisations that could take action. The National Performers List could be used to limit the scope of a GP's practice in a positive way (i.e. rather than for disciplinary processes). Fresh approaches to the appraisal/ revalidation process are urgently needed (for GPs of all ages) to reduce burden and make appropriate to a GP's existing role. Introduce (to appraisal/revalidation) a compulsory interview with a trained GP career advisor, with special appraisal at age 50/55 years to give an informed career review of role options<br><br>Portfolio and wider working arrangements<br>• Portfolio working while retaining the core role of the GP/linking portfolio activities to local population priorities and benefits<br>o Positive factors: Portfolio careers are now widely accepted as a legitimate career option; Such an approach may help GPs achieve vital 'headspace' and reflection time; Portfolio working should be seen as a positive mid-career option/reward for experienced GPs, rather than for early-career GPs who were thought to lack the core clinical experience needed for portfolio roles.<br>o Barriers: Portfolio working should be seen as a positive mid-career option/reward for experienced GPs, rather than for early-career GPs who were thought to lack the core clinical experience needed for portfolio roles; Is portfolio working just part of what a GP is/does, or does 'being a GP' fundamentally involve adopting a range of roles?; It is currently hard to define and monitor the scope of portfolio working; Organisational issues arise from the unintended 'knock-on' effects of GPs taking time away from direct patient care for portfolio roles; Overall, there was no clear shared view on how to provide a 'career structure' for portfolio roles. |

| | |
|---|---|
| | ○ Considerations & next steps: none given.<br>• Contractual arrangements for working across general practices<br>○ Positive factors: Introducing contracts for GPs working across practices would be feasible and acceptable to some, but may not be universally acceptable to GPs; Positive examples already exist of this type of arrangement working well (e.g. GP Retention Scheme); Introducing contractual arrangements for this approach reflects the importance of job satisfaction to GPs over simply being paid more to undertake more work (and thus potentially increase workload and concomitant work-related stress); 'Super practices' may be in a good position to offer such approaches to new GPs.<br>○ Barriers: Uncertainties and potential problems were identified regarding who should manage the employment of GPs working across practices and also deal with related legal/liability matters; An associated potential negative impact on continuity of care, team working and long-term commitment to a practice was identified; This style of working may be daunting to older GPs used to a more traditional model; Cross-practice working seems not to last; GPs tend to end up in one practice; Lack of clear career path for a GP working across practices in this way.<br>○ Considerations & next steps: Practice groups/federations, super practices and locum banks should be involved in formalising contractual arrangements; Practice groups/federations, super practices and locum banks should be involved in formalising contractual arrangements; BMA/NHS England – clarify guidance on legal/liability issues and the best arrangement(s) for employment and management by one practice of GPs working across practices; BMA/RCGP/GMC/NHS Digital – agree a definition of FTE for a GP (current definitions differ across NHS Digital, HEE and the BMA differ); RCGP/HEE – long-term commitment to defining a path for this type of career.<br>• Widening MDTs and role substitutions<br>○ Positive factors: Widening the skill base and professional mix of practice teams is seen as an essential way to cope with workload; Patients like having access to a range of health professionals; Other health professionals can indeed help to provide care; GPs are skilled at – and essential for conducting core elements of diagnosis and care; Practice managers are a crucial conduit for multidisciplinary working.<br>○ Barriers: There are limited numbers of health professionals available. Bringing such individuals into primary care may denude other areas; groups recognised a crisis in the nursing workforce of similar magnitude to that being experienced among GPs; Effective multidisciplinary working requires time, capacity and resources; Uncertainty was evident over the governance of extended primary care teams (training, support and 'checking up on' is also needed); There is a need for career planning for other health professionals (their career paths in primary care are largely unknown territory); Costs of employing other health professionals may be almost equivalent to hiring another GP; GPs retain key diagnostic skills and are 'better value for money'; GPs may end up dealing with a highly complex workload, with little respite in intensity of workload; Widening/extending roles across the MDT is affecting GP identity, role and skills; GP trainees are becoming 'de-skilled' in respect of chronic disease management and complex comorbidity, through becoming 'specialist' too soon in their careers and opting to undertake specific activities and not others. |

| | |
|---|---|
| | ○ Considerations & next steps: Practice groups/federations share resources and experiences (e.g. a specialist GP to support multidisciplinary working across practices). Practice teams – structure time for multidisciplinary working and related discussions during the working day. RCGP/HEE – define the required competencies, skills and experience to be a 'consultant GP' and to ensure 'learning on the job' for GP trainees that includes observing other health professionals working in practice. DHSC/NHS England – revive the previous financial incentive to employ a range of health professionals; establish arrangements to anchor such individuals to a practice by passing this money directly to practices, rather than to CCGs/trusts. RCGP – define formal training and career progression for practice managers. |
| From which stakeholders' perspective? (General practitioners, patients, other HCPs) | |
| How is work-related joy constructed? (Diversity of ways individuals may think about joy) | |
| What prevents/removes work-related joy? | |
| Organisational characteristics (small, large) | |
| Managing ill-defined illnesses and chronic complex multimorbidity | |
| Human connection with patients | |
| Human connection with peers | |
| Human connection across other organisations (e.g., secondary care) | |
| Learning systems (e.g., competence, capability, mastery, socially situated) | |
| Additional useful information | |

| | |
|---|---|
| Summary of findings | This stakeholder consultation identified facilitators, barriers and next steps for the recommendations made by the RAND/UCLA appropriateness method. |
| Strengths, limitations, and future research directions | A strength of this work was the successful recruitment and involvement of representatives from key stakeholder organisations, including some in senior and influential roles within medicine and in health more widely, and within major professional, government, policy and related organisations.<br><br>Limitations include the fact that our findings comprise a summary list of the views captured across both meetings; the finer detail of individual comments are not reported. Nor have we systematically attempted to draw out the potential complementarity or inevitable tensions between some of the proposed retention strategies (e.g. between having a daily maximum limit on the number of consultations and increasing the typical length of GP consultations). In addition, in order to make good use of stakeholders' time at the meetings and to present sufficient background material on the findings of the whole of the ReGROUP project, we needed to restrict the number of discussion topics that were possible to cover in detail. |

Supplementary Table S2: CMOCs and supporting evidence

1. Meaningful aspects of clinical practice 2
2. The commodification of GP work 15
3. Reciprocal caring: mutual acts of compassion 21
4. Imbalance between work demands and resources available 24
5. Resource/work mismatch 32
6. The shift to remote consulting 40
7. Cumulative knowledge 43
8. Agency/partnership 47
9. Direct connections 51
10. Depersonalisation 53
11. Paradox of delegation 58
12. Supporting cross-disciplinary learning 64
13. Learning systems 67
14. Informal interaction and peer support (professional and personal) 68
15. Coordination of care: primary, secondary, and community care 74
16. Climate of psychological safety 77
17. Time for care: appropriate consultation time 83
18. Agile and flexible systems 85

***GP = General Practitioner***

**<u>Meaningful work and engagement</u>**

*Contributing and mitigating factors*

1. **Meaningful aspects of clinical practice**

**When GPs can allocate time to aspects of their practice they consider meaningful (C), they are more engaged in their work (O), because they experience a sense of congruence between their core values and the nature of work (M).**

“The joy of being a GP lies in the tapestry of patient interactions. As I embarked on this unexpected journey, I discovered the unparalleled satisfaction of providing continuity of care. Building long-term relationships with patients allowed me to witness their health journey unfold, celebrate their victories, and offer support during setbacks. While the challenges are real and the days can be long, these encounters forge bonds that make GPs a cherished member of their extended family. General Practice isn't just about prescriptions; it's about being constant in their lives, an ally in their journey.” - Pereira, 2023

“Where the scalpel is the essential instrument of the surgeon, so the relationship is the instrument of the GP.” - Morland, 2022

“Moments of light that flow from knowing your community and knowing your patients, and from connecting the two.” - Morland, 2022

“She is a doctor who knows her patients. She is the keeper of their stories. Over years and across generations, witness to the infinite variety of their lives. These stories she says are what her job is all about. They are what sustains her even in days as hard as these.” - Morland, 2022

“And other things have gone, you know. Things that, you know, just doing a postnatal visit on somebody who’s just come back from hospital having their baby, now I haven’t done one of those in years. You know, I would always visit in the past relatives after a bereavement, I don’t, I certainly don’t always manage it now, often it’s unfortunately just a phone call.’ (GP8, female partner, five sessions per week, 16-20 years’ experience, medium-sized suburban practice).” - Croxson et al., 2017

“People are motivated to the tasks of daily work by their perceptions of the meaning behind their work — why it matters. People must be supported to develop and engage with an understanding of why what they do matters. If the positive impact of work done by an individual is emphasised regularly, even mundane tasks can become a means to a larger end and so become accepted into everyday actions." - Kanter's three Ms model in Andah et al., 2021

"They noted that intrinsic factors (for example, career opportunities and job satisfaction) are more important than extrinsic factors (for example, golden handshakes) in influencing retention." - Andah et al., 2021

"The authors described how a low sense of meaning may be contributing to the low morale that leads to burnout and leaving the profession." - Andah et al., 2021

"There's a sense of your own values as well… There's something that keeps some people working in these sorts of places, it's a sense of doing something valuable, or worthwhile.' (F, <5 years)." - Eley et al., 2018

"The medical breadth of the discipline also positively influenced choosing general practice." - Marchand & Peckham, 2017

"There is some evidence that job autonomy, the variety of work, the feeling of doing an important job, social support, and a good practice environment positively affect job satisfaction." - Marchand & Peckham, 2017

"The review examined the evidence on GP recruitment and retention determinants, and found that intrinsic and idiosyncratic factors, such as job satisfaction, were more important than extrinsic factors, such as financial incentives." - Marchand & Peckham, 2017

"I am a people person and like human interaction and the continuity of care you get in general practice." - O'Dowd, 2022

"All the while, we must maintain a relentless focus on protecting the essence of general practice: the deep-seated connection and continuity at the heart of the doctor-patient relationship." - Madan et al., 2017

"Work aligning with personal values was important in sustaining motivation; for example, being part of a strong team and having outside professional interests appeared to build resilience." - McCallum & MacDonald, 2021

"They recommended that, to improve retention of young GPs, "the pace of administrative change needs to be minimised, and the time spent by GPs on work that is not face-to-face patient care reduced." - Rimmer, 2016

"Of the GPs that responded to the survey, 50% said that rising non-clinical workload was a factor in their decision to leave general practice early, while 84% said that pressure of work contributed to their decision. "The higher administrative workload reduced the time available to spend with their patients, leading to a fundamental change in the doctor-patient relationship," the researchers wrote." - Rimmer, 2016

"The negative portrayal of GPs in the media, a lack of time for interaction with patients and colleagues, and a perceived blame culture also contributed to young GPs leaving the profession, the researchers found." - Rimmer, 2016

"With more and more of our working hours being taken up with form filling, ticking boxes, and preparing for CQC [Care Quality Commission] practice inspections, we are drowning in red tape, and this only serves to keep us away from delivering frontline patient care, which is why we become doctors in the first place." - Rimmer, 2016

"The second category, 'low morale', was seen to be associated with reductions in the perceived value of GP work (with loss of identity) and changed professional culture (more target-driven and standards-driven rewards system; multidisciplinary team-based working (yet for some also lone working/isolating culture); a more aggressive top-down managerial culture within the NHS; and more widespread norms and expectations for early retirement)." - Long et al., 2020

"Some of the organisational changes were considered to have imposed increased clinical and non-clinical responsibilities and work on GPs. Together, such changes were believed to have undermined some of the basic tenets and traditional expectations of being a GP, such as the doctor-patient relationship and having autonomy and control over one's clinical work." - Long et al., 2020

"In the contemporary career model, GPs said they are expected to give up autonomy in many areas of their job and are expected to accommodate increasing government regulation and bureaucracy, which increases stress related to workload, particularly 'paperwork'/record-keeping." - Long et al., 2020

"GPs said job satisfaction directly relates to the quality of the doctor-patient relationship, with more time available for GPs to spend with their patients being associated with better job satisfaction." - Long et al., 2020

"Lack of time with patients is perceived to compromise the ability to practise patient-centred care and undermines GPs' professional autonomy and values, resulting in further diminished job satisfaction."- Long et al., 2020

"… Confirmed experiencing greater enjoyment when performing roles affirming their sense of providing valued health care." - Spooner, 2016

"Their accounts confirm that their job satisfaction is greater when they believe they can help achieve positive outcomes for patients and are able to engage in tasks that are well aligned with what they perceive to be good doctoring." - Spooner, 2016

"Satisfying work included the challenge of being the first doctor to assess a new problem, with an additional boost felt on confirmation of a suspected diagnosis:

"I send somebody to hospital, and I phone up and I want to know how they are getting on, that kind of adrenaline rush of YES; I got that diagnosis right.' [Alice]." - Spooner, 2016

"Long-term engagement with patients was highly valued. Because most of these GPs had worked in the same practice for most of their career, they had acquired detailed knowledge of the personal histories of many returning patients and were cognizant of the impact of family relationships and social backgrounds:

‘It’s almost like old-fashioned general practice … if they have got difficulty remembering where their roots are, I can usually fill the gaps in for them. And that’s, that’s a nice feeling.' [Alice].” - Spooner, 2016

“Beyond simple factual knowledge, they spoke of relationships with patients as they negotiated life-altering illnesses together, picking up on previous encounters and continuing to support ill-defined difficulties:

“I saw a patient today who … was already struggling as a teenager when I first met her 22 years ago … psychiatry has bounced around saying she is just personality disorder and so … impossible to treat, and I have seen her through my whole career … she comes back time and time and time and time again. It’s got to be a privilege hasn’t it because you know that’s something that no one else will have with that person, and she obviously respects me.’ [George].” - Spooner, 2016

“In addition to teams within their practices, GPs were members of other teams, networks, and forums through involvement with undergraduate education and postgraduate training roles. These could be particularly stimulating:

“The teaching gives me a high, it’s just being with young students and going back to basics again … teaching examination, looking at interesting patients, so it’s exactly back to basic clinical skills.’ [Jennie].” - Spooner, 2016

“I would really like to always be involved in teaching other doctors … or students, juniors, whatever. As to why? … it’s difficult to say really, just like it gives me a buzz, makes me feel … like a real doctor I suppose, like a real doctor, a real useful person for others, a useful resource, somebody with knowledge, somebody that can help, the helpful guide sort of thing. And an affirmation of my abilities … still love it.’ [Helen].” - Spooner, 2016

“As front-line clinicians, GPs reported fulfilment, excitement, and a sense of purpose, achieved through responding to patients’ needs and offering long-term support.” - Spooner, 2016

“As teachers, they reported great pride in shaping the clinical skills and professional development of medical students and junior colleagues, hoping they would step forwards as future professional leaders.” - Spooner, 2016

“Although neither they nor the conditions in which they worked were perfect, it was still the case that professional pride, ability to make things happen for patients, listening skills, and supportive action were not simply trite and aspirational statements but also remained vital contributors to personal and professional fulfilment.” - Spooner, 2016

“Third, resilience required the integration of personal and professional values rather than keeping the two separate.” - Eley et al., 2018

"…the biographical narratives of well-informed and reflective practitioners to identify characteristics of GP work to which they attach greatest value and to discover what motivates their continuing practice." - Spooner, 2016

"Awareness of doctors' underlying sense of medical identity, and how this appeared to influence their reactions to shifts in expectations, adds to knowledge about how attractive, valuable, and durable roles for GPs may be re-established." - Spooner, 2016

"Positive feedback from patients was a frequently mentioned confirmation that doctors' efforts had been appreciated, with one defining his job as a 'privilege' and another observing that for some patients he could be 'the second most important person in their life'. Some related incidents when they had been active advocates for particular patients for whom they felt the NHS had failed to provide adequate care. In a practice with a high proportion of older patients, a GP's investment of time to 'really listen' allowed her to better understand how to juggle conflicting treatment plans." - Spooner, 2016

"For me, and anecdotally for all the GPs I know who have enjoyed the job, continuity of care, and the complex evolving relationship between doctor and patient, are of central importance. These relationships give a human meaning to the work." - Dr Cym Ryle, House of Commons, 2022 (The future of general practice - Fourth Report).

"Wordsworth et al suggested that enhancing the patient care aspects of a GP's work is more likely to act as a key for retention, and that lack of consultation on changes can lead to dissatisfaction." - Marchand & Peckham, 2017

"Mentorship schemes and opportunities to develop portfolio careers would be welcome at every stage of a GP's career, not just for senior doctors or towards the end of working lives." - Marchand & Peckham, 2017

"Research on GPs as well as physicians in general, shows that those with higher job satisfaction have a lower likelihood of burnout, stay longer in their job and experience less job stress, are more involved in decisions that affect work and perceive their work as interesting." - Stobbe et al., 2021

"Another way for GPs to have more challenging and varied work is to have other paid activities next to working as a GP. GPs who participate in teaching were more satisfied with their work than GPs who did not. Involvement in other professional activities can serve multiple instrumental goals at the same time: more stimulation from a wider variety of tasks; more status from improving their skills; and more behavioural confirmation from feedback and support from, e.g. students." - Stobbe et al., 2021

"The practice location may be related to GP job satisfaction for two reasons. Firstly, GPs in rural areas have a more varied practice because there are less alternatives to their patients, such as hospitals. Secondly, people in rural areas are more attached to and socially integrated in the community than people in urban areas. Thus, GPs in rural areas may be more satisfied due to more opportunities for both stimulation and behavioural confirmation." - Stobbe et al., 2021

“We theorised that GPs ‘produce’ job-related well-being through doing stimulating work, deriving comfort, social status and behavioural confirmation from their work.” - Stobbe et al., 2021

“[...] Table 2 [...] shows that GPs who carry out more technical procedures and who are more involved in preventive care and health promotion are more satisfied with their job.” - Stobbe et al., 2021

“Another possibility is that administrative and organisational tasks increasingly become an accepted part of professionals’ work. GPs might not enjoy these tasks very much, but not resist them either because they acknowledge that it is an important aspect of their work.” - Stobbe et al., 2021

“The most popular reason for career choice varied between countries with ‘compatibility with family life’ most strongly represented in the UK sample; ‘challenging, medically broad discipline’ most important in Norway, Portugal and Germany; ‘autonomy and independence’ in the Czech Republic; and ‘holistic approach to the patient’ in Italy.”- Roos et al., 2015

“Informants thought that for a career in general practice to become attractive again there should be an emphasis on developing portfolio roles and interests (such as education/training, mentoring, special interest, appraising, and research) to sit alongside clinical work. Portfolio roles are particularly attractive to ‘twilight’ GPs as they allow flexibility to reduce workload in specific areas, and to open up roles in others, for instance experienced GPs providing ambulatory or non-ambulatory chronic care could be mentors to newly- qualified GPs in their practice of acute care.” - Lyon-Maris et al., 2015

“I’m interested in the concept of a national salaried contract for GPs, with equal (good) pay per session regardless of region. However, I would want the opportunity to be paid more for delivering extra services, including: extended scope of clinical practice; conducting quality improvement; working outside 8.00 am - 6.00 pm; delivering training for students, trainees, and allied health professionals; leading on research projects; and taking responsibility for local care homes.” - Lumley, 2020

“Able to provide continuity of care with the learning opportunities and satisfaction that comes with following patients up. Being part of a team. Being part of the team of doctors that have provided fantastic long-term peer support and allowing me to provide continuity of care to my patients/case load with the flexibility of altering hours to manage my young children.” - Lockyer et al., 2014

“Being able to fit childcare into the working week was of great importance for many retainers: A steady job with set hours (very important with two children in nursery). Paid annual leave and study leave. Able to organise regular childcare - almost impossible to locum as still have to pay for childcare even if you don't get the sessions.” - Lockyer et al., 2014

“Responding in their own words, the survey participants provided a wealth of detailed data regarding the perceived benefits of the retainer scheme in comparison with locum work. These included the opportunity to work in a stable practice environment with a set number of sessions and to receive performance review: ‘A chance to work in a stable practice environment where I can be confident, I am receiving performance feedback and where I can have a set number of sessions per week.’ ‘A reliable position in a practice where I feel part of the team and able to ask questions if I need to.’” - Lockyer et al., 2014

“Running the practice or having a meaningful conversation with staff is relegated to the edges of the day. Almost every practice is struggling to balance rising workload within tighter financial constraints. Add to this the strain of recruitment issues and it becomes easy to see why morale is so challenged. Clinicians increasingly feel unable to provide the care they want to give, and understandable resentment of working under this pressure is growing.” - GPFV, NHS England, 2016

“GPs were not fully engaged in the programme that sought to increase access and were more motivated by their own particular local delivery and concern for individual patient and population practical needs.” - Davis et al., 2018

“Appraisal commenced in 2002 to promote lifelong learning and continuing professional development (CPD), but it changed with revalidation in 2012. Many doctors felt that appraisal became a summative assessment, losing the pastoral element. Appraisal has evolved, balancing educational development with the requirements of revalidation assessing clinical competence, but there are other core attributes to being a good doctor such as care, compassion, and looking out for vulnerable colleagues.” - Ferguson et al., 2020

“‘Although I enjoyed volunteer work and spending more time with my family and our dog, I still felt that I had that enthusiasm for my job’, he says, explaining his return to general practice. I went through the refresher course and a placement and came back into the workforce in 2020 as a locum. By making myself available regularly on a weekly basis, I am providing continuity of care, even though I’m a locum doctor.” - O’Dowd, 2022

“Over the years he has enjoyed looking after his patients’ children, and their children’s children. His patients send him birthday and Christmas cards, and their relatives ask him to give readings at their funerals because he knew them so well. “I see myself as a family GP. I enjoy that caring bond with my patients. So many people have no one they can talk to. My patients can talk to me.” - Oxtoby, 2022

“Participants had been attracted to GP work in the expectation that it would offer continuity of patient care, professional autonomy, and flexibility in working hours, along with the intellectual challenges inherent in problem solving.” - Doran et al., 2016

“Gupta hopes that his practice will continue to provide a personal service to patients, as that is the reason why most GPs have chosen to do this job. We are the only people in the healthcare system who have that relationship with patients and their families, and it would be a huge shame if that personal approach was lost.” - Oxtoby, 2021

“We don’t really get taught the business side of general practice, and we don’t really get taught leadership skills, and I think that’s so important if you are becoming a partner. I think there’s not anything about the business of general practice in training and I think that’s wrong, and I think if I was changing one thing it would be encompassing a bit of leadership skills.” - Cunningham et al., 2021

“The success of general practice care depends as much on the generalist approach32 as on the ability to establish a therapeutic relationship with one’s patients. Being person-centred is key to the development of relational continuity which, in turn, allows the emergence of a deep contextual understanding about a patient’s personal circumstances and preferences.” - Sturmberg et al., 2023

“The RCGP is launching a major initiative on relationship-based care, which is regarded as being central to effective general practice.” - Jones, 2021

“ …the tens of thousands of life-changing encounters that happen every day in the intimacy of general practice consulting rooms, unseen and often disregarded by politicians and policymakers.” - Jones, 2021

“Core values [of GPs] refer to those that remain unchanged in a changing environment. These would be: colleague-centred problem solving, patient/person-centred healing relationship, context-driven adaptive learning, being agents of progress.” - Sturmberg et al., 2023

“Yeoman (2020) regards meaningful work as an orientation towards taking care of significant things of independent value and that offers autonomy, freedom and dignity. In turn, this work is experienced as worthwhile. Meaningful work relies on mutuality, with shared ethics and appropriate structural enablers needed to allow people to contribute with dignity and autonomy (Laaser and Karlsson, 2022) around a common goal.” - Laverty et al., 2023

“For Yeoman (2020), the traditional social contract reflects what she considers as values, the public promises that set out what parties can expect of each other. Harris (2017) suggests that in healthcare this involves an implicit agreement that doctors will act in the public’s best interest by being devoted, competent and moral physicians in return for social recognition and financial rewards from society (Cruess and Cruess, 2008). In other words, doctors will engage in morally worthy work in exchange for autonomy, freedom and dignity from the state and public.” - Laverty et al., 2023

“On paper then, the work of GPs should be meaningful: they are highly skilled professionals with a large degree of control over their work (Frenkle et al., 1995) recognised as contributing to helping others.” - Laverty et al., 2023

“The loss of control because of time pressures features in the literature as leading to meaninglessness at work.” - Laverty et al., 2023

“Relevantly, studies with GPs have found that commodified time is now a key way in which they talk about their work (MacBride-Stewart, 2013) due to the loss of control over appointment times, the challenges of juggling administrative and caring tasks, and the need to work within decreasing time resources (Horobin and McIntosh, 1983; MacBride-Stewart, 2013). The control of time, therefore, is positioned as both the problem and solution for more meaningfulness at work (Adam, 2003).” - Laverty et al., 2023

“De Boeck et al. (2019) found that employees need to have the opportunity to imagine and realise future selves for work to be meaningful.” - Laverty et al., 2023

“Attachment to the NHS as a morally worthy organisation, however, remained a stable source of value, although concerns about privatisation were adding to uncertainty about the future.” - Laverty et al., 2023

“General practice of the past was seen as desirable and meaningful predominantly because the work environment was perceived as contained, focused and morally significant.” - Laverty et al., 2023

“The participants described feeling like the sacrifices they had had to make were not valued or cared about. Their dignity had been compromised. Yeoman (2020) refers to dignity as a core aspect of meaningfulness - that individuals have to be recognised as people with lives to lead and that to care for others they need to have the capacity to care for themselves. Instead, in these hospital settings, they were treated as a commodity for the current needs of the organisation rather than for their benefit as the next generation of doctors. This had a direct impact on the doctors in the immediate aftermath of foundation training - many described experiencing burnout, and others took a break to reevaluate if they wanted to continue. These episodic moments of meaninglessness give rise to the uncertainty that stretches into present actions and decisions about the future.” - Laverty et al., 2023

“The doctors described a strong commitment to the public values of the NHS as a source of meaningfulness that was a strong motivator for both deciding to become a doctor and to stay. For many participants, continuing in medicine was contingent on the NHS: ‘It sounds, probably, a bit too ideological, but I mean, I do like working in the NHS and if the NHS was to crumble, then I’d have a real good reason to leave’ (Focus Group 4, new GPs).” - Laverty et al., 2023

“For others, their desire to uphold the values of the NHS jarred with their experiences of working within it. Lydia, who has moved away from general practice, for example, states: ‘I feel a loyalty to the NHS . . . I want the NHS to work. And I want to work for the NHS.’ The ideological values of the NHS as an institution were an important source of meaningfulness and buffer to uncertainty for participants, although personal experiences within the structures of the institution differed. In other words, the values of the NHS were a relatively stable source of meaningfulness despite the working environment (the loss of autonomy, dignity and freedom) making meaningful work less possible. The loss of this security, then, could have detrimental impacts on meaningful work now and in the future.” - Laverty et al., 2023

“The novel approach taken here, adopting Adam and Groves’ (2007) theoretical work on futures, shows that there are multiple fluid future orientations that may impact how individuals, and organisations, construct and anticipate meaningful work. This opens up interesting avenues for which to understand meaningful work, particularly under conditions of uncertainty and crisis. In the current study, two conflicting future orientations impact meaningful work during a professional crisis.” - Laverty et al., 2023

“The findings show that there is a mismatch between imagined futures and lived futures as a result of reforms that have led to changes in the workplace environment. In the recent past, general practice was perceived to provide meaningful work through the traditional social contract. Morally significant activities, a work environment that allowed autonomy, freedom and dignity, in a profession that was respected (Laaser and Karlsson, 2022). In other words, components that fulfil Yeoman’s (2014) definitions of meaningful work. These perceptions of general practice shaped the imagined futures of junior doctors choosing to train as GPs. The lived future these GPs were faced with, however, was a work environment characterised by accelerating work pressures and intensity, uncontained and changing work environments, alongside a loss of public respect (Charles-Jones et al., 2003; Fenwick, 2013; Rich et al., 2016).” - Laverty et al., 2023

“Looking at what makes General Practitioners (GPs) happy in their profession, may be important in increasing the GP workforce in the future.” - Le Floch et al., 2016

“Focusing positively on factors of satisfaction keeping General Practitioners (GPs) in their profession, may increase the likelihood of adequate GP provision in the future.” - Le Floch et al., 2016

“Having the opportunity to take responsibility, having a positive self-image and recognition for work undertaken were seen as important prerequisites for being satisfied with the profession.” - Le Floch et al., 2016

“Successful medical management of patients and the subsequent feelings of being competent were associated with a higher degree of satisfaction. In a Scottish qualitative study, GPs derived the greatest satisfaction from consultations where GPs perceived that they personally had contributed to a successful outcome for the patient [27].” - Le Floch et al., 2016

“Freedom to choose the workplace and work organisation was one of the most relevant topics found in the literature, addressed in 13 articles. GPs wanted to have the freedom to choose their work method [16, 19, 22], their payment method [22], and have flexible working hours [18]. GPs need this freedom in order to have job satisfaction [24]. In a qualitative study, an American GP said: “It’s a way of having independence and doing what I want” [26].” - Le Floch et al., 2016

“Vocational choice; being able to be the kind of doctor you want to be: is an important sub-theme in being satisfied with your work as a GP. Six studies stated that the GP’s own personality and personal values played an important role. Values can differ between GPs. If their job were fully compatible with their temperament and personal values, then GPs were more satisfied. Some chose to be a “traditional family doctor” [21]. The mission for most GPs was to help people. “The goal of practice is to meet people’s needs, take care of them and do the best you can” [26].” - Le Floch et al., 2016

“Intellectual stimulation to use abilities; continual professional development: This is one of the most studied aspects of satisfaction. Twelve articles in the review referred to the intellectual aspect of the profession as a positive factor. GPs appreciated the intellectual challenge, and necessary skills that their GP work offered them. “The intellectual and practical challenge to solve and assist with people’s medical problems” [25]. “Doing a wider range of procedures was associated with higher overall job satisfaction” [24].” - Le Floch et al., 2016

“Another group of potentially positive factors was related to the GP and his/her patient contacts and relationships. An efficient doctor-patient relationship was also considered an important factor in GP job satisfaction [17].” - Le Floch et al., 2016

“The opportunity to deal with a variety of medical problems and patients was a positive aspect, even in rural areas.” - Le Floch et al., 2016

“Teaching and academic responsibilities was a final positive sub-theme related to GP practice. Positive GPs wished to communicate their work and skills. Eight articles studied the teaching role of GPs. Academic responsibilities gave GPs positive stimulation. “Teaching, sharing knowledge and experience, and mentoring” [23] “Factors that contribute to career satisfaction for physicians include teaching and research” [22]. As teaching is a factor for GP satisfaction, internships in General Practice were seen as extremely important in attracting students to General Practice [15]. This was an important influence on career choice for young students [31].” - Le Floch et al., 2016

“Of great importance for GP satisfaction is the freedom to organise and manage their own work and to be able to be the kind of doctor they want to be. Providing a context where GPs can build, run and organise their practice in line with their personal values seems a better way of maintaining satisfaction among the GP workforce.” - Le Floch et al., 2016

“Next, the literature analysis highlights the importance of challenges such as intellectual stimulation [25] and being able to practise a wide number of procedures (23). The diversity in practice is the first characteristic of General Practice, according to the WONCA definition. The variety in General Practice is a positive aspect well studied in the literature, found in nine articles in our review. The opportunity to widen activities to teaching and doing academic work, was also found to contribute to a high level of job satisfaction for some GPs, along with feeling clinically competent [27]. With regard to the latter, being able to make a personal contribution to a patient’s health is another very relevant experience contributing to job satisfaction. Our systematic literature review has shown that a satisfied General Practitioner wants to be clinically competent.” - Le Floch et al., 2016

“Mastery and feeling effective and competent, absolutely because if you have the time to address patients’ needs and do it holistically you feel you are in the job. If you are a bit more refreshed, if you have this extra minute to think, quite often you feel so much more competent and you just don't feel rushed, you don't. What I hear from other colleagues is, I hear more and more that they are losing compassion and we basically I think are not only, could work on our competencies but we could also work on our compassion and that's a wonderful combination in what I say the mastery of medicine, the art of medicine." - Dhanani et al., 2022

"GP fellows reported feeling an increased sense of meaning in their work, which they ascribed to having an overarching goal (the Deep End GP vision of improving general practice in deprived areas) and the resources (time and learning) to facilitate efforts towards that goal, including advocacy work. 'In terms of purpose, coming along to the steering group meetings of the Deep End, you see the big picture of what the drive is for the Deep End. How you have to, you need leadership by GPs or primary care to lobby for change which you don't get to see when you are a trainee at all.'" - Dhanani et al., 2022

"The panel deemed at least one statement as appropriate for 24 out of 54 policy and strategy areas, most of which related to the provision of personal or professional support for GPs to potentially protect against burn-out and improve job satisfaction. Examples include policies and strategies to enable flexible working (e.g. portfolio careers, or support for 'programmed health care activities' to allow GPs to work across several GP practices), access to a dedicated occupational healthcare services, and/or peer support schemes." - Chilvers et al., 2019

The importance of honest communication and human connection with marginalised patients was highlighted by professional and lived experience participants and enabled progress of patient and professional agendas. "As a service user you walk in and I know when they're putting on a voice… just be yourself, do you know what I mean? … I think CC1 to me seems, when you meet her in the surgery is how she would talk at home do you know what I mean? It's just her"- BGW2 - Potter et al., 2023

"Professional participants found joy in doing 'a good job' for patients who really need it and that prioritising continuity for patients who really need it may ease pressure on other general practice clinics." - Potter et al., 2023

"[..] Both Shadbolt and Bunker (2009) and Young and Leese (1999) suggest that a wider choice of long-term career paths such as sub-specialisation and portfolio careers (including specialisation related to their practice such as dermatology, paediatrics etc.) are important for both the recruitment and retention of GPs. It is also suggested that increased learning (satisfaction of intellectual and altruistic needs) and functional flexibility (adaptable and transferable within their practice) could improve satisfaction, morale and fulfilment and, as a result, GP retention" - Peckham et al., 2016

"Better quality of life and job satisfaction were identified as positive attributes associated with general practice when compared to other medical specialities; [...] the future of the GP profession (including perceptions of lack of training, poor pay and increased workload), and the lack of respect associated with being a GP were identified as key concerns.” - Mitchell et al., 2018

“Teams and services, and ultimately patients, lose hard-won wisdom and expertise. But, most importantly, doctors don’t all want to retire, many reporting that they still enjoyed the practice of medicine and using their clinical skills to help patients. What they want is a change of pace or role, a move to part-time working, being nearer home, more time with family and friends, and to continue to add value. Yet ‘all or nothing’ is often seen as the only option.” - Bhatti & Viney, 2017

“Successive government ministers and spokespeople either do not see or will not speak of this. Instead, they recurrently talk of ‘fixing’ the problem by training more doctors here or recruiting them from abroad. But such planned staff replenishment is most unlikely to succeed. Apart from the inescapable delay of many years, such measures do nothing to restore what has been lost in the nature and quality of the work — what fuelled and spirited those previous generations of GPs to (usually) very willingly devote a working lifetime to full-time practice, often working longer hours than their much more miserable counterparts now. What has been lost — or jettisoned — in our decades of ‘modernising’ or ‘streamlining’ healthcare reforms are personal experiences of connection, relationships, community, and the rich values and meanings that can grow from these.” - Zigmond, 2023

“The evidence from this review also suggests that selection and educational exposures are important [...]. Students are likely to be retained in a rural or underserved area if they have connections to the area, and are exposed to good educational primary care placements.” - Verma et al., 2016

“My patients' trust is more important to me than anything I can think of.” - Morland, 2022

“No two patients, no two people are ever the same. Pay attention to this above all and it will not just make you a nicer doctor, but a better doctor.” (intellectual stimulation and variety) - Morland, 2022

“On days when the administrative paperwork piles high, these stories are what keeps her going. They are the raw material of her relationship with each patient and a source of endless complexity and fascination.” - Morland, 2022

“Feeling valued on account of their work was important to GPs. However, they reported often feeling undervalued by the general public, the NHS, the media and the government: I think most people, if you ask them why they do jobs, it’s a complex mixture and a lot of it comes about being valued and appreciated. I mean, people always focus on incomes and things but, the more detailed the analysis is, it always comes back to things like being appreciated, feeling valued (GP partner, male, age 60+ years, intending retirement)” - Sansom et al., 2018

“By and large, the GPs reported that seeing patients was the part of the job they enjoyed the most, however, there were several factors that made the job ‘on the cusp of undoable at times’ (GP5, Partner, female, age 55 years), and these factors were all cited as influencing decisions about when to retire/quit direct patient care. Key issues identified by the GPs were: workload; change; NHS structures and systems; concerns about the future of general practice; partnership issues; politics; the impact of ageing and ill health; and reduced morale and emotional resilience." - Sansom et al., 2016

"However, it is important to remember that while face- to-face consultations with patients can be emotionally demanding, many participants indicated that patient contact was also the most enjoyable and satisfying aspect of their work, as these participants highlight: 'My job satisfaction was patients coming in and going out having got their needs attended to and happy.' [...] 'I feel very conflicted, because I love general practice. I've always wanted to be a GP. I love my patients. And on a day when I've got time to spend with them, it's a brilliant job.'" - Riley et al., 2018

"Doctors need to experience effectiveness and feel they are delivering valued outcomes. If doctors are to be attracted into general practice, the professions' contribution must be understood and valued. We have heard successive Prime Ministers refer to general practice as the cornerstone of the NHS, and yet GPs have faced extreme public scrutiny and continue to feel undervalued." - Jefferson et al., 2022

"During the covid crisis 'many consultations have by necessity become more transactional, and this has reinforced the mistaken view of some people that general practice is largely a transactional medical specialty rather than a relational one,' Marshall told the 2020 RCGP conference. But GPs, he said, know that 'the trusting relationship between a patient and a doctor—often but not always based on continuity of care—is the most effective intervention that we have at our disposal. It's what helps us to use medications, investigations, and specialist referrals appropriately and to reduce the risks of medicalising non-medical problems. It's what encourages patients to engage with our advice, [and] it's what many patients want from their GP.' Marshall told conference delegates that the covid experience 'has reinforced our decision to make the promotion of relationship-based care a strategic priority for the [RCGP].'" - Oxtoby, 2021

"For clinicians who struggle with efficiency, this means that the aspects of work that are less meaningful e.g., documentation, data entry) edge out the more meaningful aspects of practice (time spent talking with patients, working with the rest of the health care team) and lead to a less joyful practice." - Guck et al., 2021

### 2. The commodification of GP work

**When policies and guidelines emphasise the compartmentalisation (e.g. single disease management) and commodification (e.g. payment/reward) of GP work (C), this can change the doctor-patient relationship (O), because GPs feel pressurised into reducing opportunities for therapeutic connections, situated knowledge, and continuity of care (M).**

"Successive GP contracts have reshaped services, altered priorities in primary health care, and redefined payment mechanisms." - Spooner, 2016

"Of particular significance was the 2004 contract in which the Quality and Outcomes Framework (QOF) linked payment to targets in the management of certain chronic conditions rather than for provision of services to patients. Studies confirmed that GPs responded to achieve those targets,[8] though possibly with relative inattention to other aspects of medical care.[9-11]." - Spooner, 2016

"That willingness to invest in the moment, that instinctive ability to occupy, reflect, and respond to the here and now, both emotionally and intellectually, sits at the heart of the doctor-patient relationship as it plays out in general practice." - Morland, 2022

"The higher administrative workload reduced the time available to spend with their patients, leading to a fundamental change in the doctor-patient relationship:

"You see it does change the doctor-patient relationship because it changes how you react to people and how you interact with people. I mean it's obvious stuff, but when you're really stressed and you've still got 15 people to see, you don't have the time for people, you don't have the interest.' (GP11)." - Doran et al., 2016

"Most often GPs described a slow process of disillusionment and burnout, feeling alienated from a system that had changed the role of general practice from a community-based, person-centred profession that offered continuity of care, to a managed service emphasizing targets and budgets. Being increasingly squeezed between administrative and patient demands, and having ever less discretion, respect and collegial support afforded to them, sapped their job of pleasure." - Napier et al., 2019

"The introduction of monetized targets led some GPs to lose clarity about whether they were motivated by professional values or by external incentivization."- Napier et al., 2019

"You just put in more effort into the target and you neglect the other things you could or should be doing…'I'm doing this because I'm making money out of this, or I'm doing it because I think it's worthwhile?' (Participant 3)"- Napier et al., 2019

"The introduction of performance indicators in GP contracts resulted in growing resentment. The targets distorted the focus of clinical care onto a small number of conditions, turning clinical practice into a 'treadmill'."- Napier et al., 2019

"PCNs are well suited to deliver on predetermined tasks and respond to direction and guidance from central bodies, and have been effective in quickly making progress towards national strategic goals by establishing new and enhanced services for patients. However, as hierarchical networks they face a risk of over- regulation and excessive performance management that could inhibit motivation and enthusiasm for PCN teams and hamper their ability to innovate locally, which has been an issue for predecessor primary care organisations." - Parkinson et al., 2021

"I had a conversation with a GP last week who described himself as being on the hamster wheel of doom, which I thought was a great phrase, because he is micro-incentivised for so many different things. Paying for another extra thing is probably not the way to [achieve whole-scale transformation], but rather building a vision for the kind of range of functions that have to be delivered[.]" - Dr Rosen (Q122) , House of Commons, 2022

"We recommend abolishing the Quality and Outcomes Framework (QOF) and Impact and Investment Framework (IIF) which have become tools of micromanagement and risk turning patients into numbers. GPs should be treated like professionals and incentivised to provide relationship-based care for all patients by restoring individual patient lists." - Health and Social Care Committee, House of Commons, 2022

"Informants not only described the clinical administrative work extending from face-to face consultations, but also that arising from visits, telephone consultations, and triaging being layered on top. They described a rising tide of 'invisible' paperwork from inside and outside the NHS system, including hospital clinics, out-of-hours, NHS 111, blood tests, radiological investigation results, and insurance reports, all of which is set within a culture of target management and reporting,10 high patient expectation, and increased risk of litigation." - Lyon-Maris et al., 2015

"We are no longer allowed the freedom to make professional decisions in the interest of the community we serve." Lyon-Maris et al., 2015

"It became people doing things in their own time for free because the CCG didn't count for it at the initial stages. Why wait until you've proven a concept and got it just right before you pay staff to do it, why don't you count it for activity, because then it might actually happen. So I think if the CCG were more supportive, and for me, I felt a bit upset by that, I thought well actually, we're doing all this hard work to develop and grow primary care, and to make a real difference and to have impact and to make this change happen, the CCG should be celebrating, they should be joining in and they should be behind it." - Swaithes et al., 2020

"Participants perceived a shift of agenda and power by suggesting that individual appointments have a clinician-led agenda and tick box mentality (driven by policy drivers and financial incentives such as the Quality and Outcomes Framework [QOF]), whereas group consultations were perceived as being driven and led by patients. The group dynamic was suggested as the most powerful benefit of group consultations. For example, by peer support from patients 'challenging each other in a way that staff can't' regarding sensitive issues such as diet and exercise." - Swaithes et al., 2020

"Doctors who were still working were asked what would encourage them to stay in medicine for longer. Factors cited most frequently were 'reduced impact of work-related bureaucracy' (cited by 45%) and 'workload reduction/shorter hours' (42%). Men (30%) were more motivated than women (20%) by 'financial incentivisation'." - Smith et al., 2017

"A recent review of the national quality pay for performance scheme the quality and outcomes framework (QoF) found that most of the indicators were unlikely to promote the aims of the FYFV relevant to primary care, including integrated and patient-centred care leading some CCGs with new models of care to discontinue QoF." - McDermott et al., 2019

"The second category, 'low morale', was seen to be associated with reductions in the perceived value of GP work (with loss of identity) and changed professional culture (more target-driven and standards-driven rewards system; multidisciplinary team- based working (yet for some also lone working/isolating culture); a more aggressive topdown managerial culture within the NHS; and more widespread norms and expectations for early retirement)." - Long et al., 2020

“In the contemporary career model, GPs said they are expected to give up autonomy in many areas of their job and are expected to accommodate increasing government regulation and bureaucracy, which increases stress related to workload, particularly ‘paperwork/record- keeping’.” - Long et al., 2020

“Overall, the rather negative picture portrayed by the four qualitative interview studies was that UK GPs with poor job satisfaction are also those who feel overworked and unsupported. Many feel part of an over-bureaucratised system and describe being at the front end of a service unable to deliver what it promises.” - Long et al., 2020

“Contracts are absolutely pointless. Contracts are often used by people to prevent progress and change and as soon as someone says that’s not in my contract, you know you are onto a loser and know you need to go right back to trying to build the shared purpose and common goal.” - Kordowicz et al., 2022

“Further, some of the participants presented themselves as having to navigate between the transactional and the purposeful: ‘But now actually we are in a position where we are going to have to sub-contract to every single member practice because they are the ones with the DES [Directed Enhanced Services]. So, then you get into implications around things like that, and NHS pensions, that’s the chunky stuff that I think is a threat, because a GP is just going to go ‘do you know what, I just can’t […] and you lose the ownership.” - Kordowicz et al., 2022

“The recent increase in inspection, regulation, and incentives for outcomes such as the Quality and Outcomes Framework (QOF) has had a number of consequences, both intended and unintended. […] Important elements of care, valued by professionals and patients alike, can be eroded by these approaches, such as continuity and person-centred care” - Derksen et al., 2020

“For the majority of participants, attempts to juggle what they saw as ‘impossible targets’ with ‘unrealistic appointment times’ detracted from delivering good patient care:’ The partner would come in before I started surgery and say, ‘Oh don’t forget to do all the QOFs’. And that was more important than actually focusing on the patient. With busier and busier surgeries with more and more extras, something has to go, and I think what ends up going when you’re under pressure to get all the QOFs and the money in, is the actual patient relationship.” - Doran et al., 2016

“For many participants, being portrayed as ‘overpaid and under delivering’ was tantamount to ‘media battering’: ‘One of the frustrations is that I think there was definitely a political spin against general practice/ It doesn’t help when you’ve had a bad day at work and you come home and watch the 10 o’clock news and you see somebody on the telly saying ‘Oh these GPs aren’t working very hard and patients can never get appointments’. Just constant criticism in the press about the fact that GPs were getting paid an awful lot of money and they weren’t having to do the out-of-hours, and they weren’t working nights and weekends.” - Doran et al., 2016

“In both the established and trainee GP groups, a majority stated that they did not find mandatory written reflection helpful or valuable. Respondents reported that written reflection was time consuming, with an adverse impact on other learning opportunities and their work-life balance. Three-quarters of those surveyed stated that they saw written reflection as a ‘box-ticking’ exercise and that writing down their reflections was tedious, using words like ‘meaningless’, ‘wasteful’, ‘counterproductive’, and ‘patronising’.” - Curtis et al., 2018

“At a time when recruitment to medicine is problematic, and burnout and disenchantment worryingly common, the benefits to medical staff of a patient centred, rather than a purely transactional, approach to care should be of particular interest to policymakers and planners.” - Jones, 2021

“General practice has experienced significant reform over the past few decades, with changes to the structure of practices leading to rising patient volume (Forbes et al., 2019), increasing patient complexity, new tasks from secondary care (Charles-Jones et al., 2003) and a greater focus on productivity, efficiency and regulation (Fenwick, 2013).” - Laverty et al., 2023

“Lepisto and Pratt (2016) theorise that professionals may experience ‘anomie’ (feelings of pointlessness or alienation) when there is uncertainty or change in priorities within the working context that conflict with their subjective experience and expectations of their work.” - Laverty et al., 2023

“The previous generation of GPs are blamed by some participants for not adequately future-proofing the profession, while others recognised the fault of policymakers and healthcare leaders who prioritised tackling current needs in the system rather than considering long-term solutions.” - Laverty et al., 2023

“‘The degree of invasive scrutiny by the GMC, CQC, appraisals, and revalidation is completely demeaning to a highly intelligent profession’ [...] ‘Am constantly having to prove that I am not a criminal or an incompetent to the CQC, GMC, NHS England—all treat us as the enemy, or as naughty children who cannot be trusted’” - Dale et al., 2016

“The effects of P4P (pay-for-performance) on practitioners can present in several forms. Practitioners may lose autonomy, performance heterogeneity may lead to wage dispersion and relative income effects, and intrinsic motivation may be eroded by the extrinsic motivation to attain targets. [...] Our findings suggest GP job satisfaction is insensitive to the proportion of income exposure to P4P. Our results suggest, therefore, that policymakers seeking to make changes to the exposure of income to P4P are unlikely to alter GP job satisfaction subject to final income remaining constant.” - Allen et al., 2016

“Spending less time on administrative tasks and more time on their clinical role would allow patient care and job satisfaction to improve. It was felt that this could also be achieved quickly compared with the time needed to train and recruit new GPs. ‘Reduction in administration—we can’t do anything about patient demand, other than train more GPs, which takes a great deal of time. A third of my time is spent on administration, a lot of which is reading through unnecessarily duplicated reports and results, extraneous information from hospitals, or acting as a secretary with a prescribing licence for hospital colleagues.'" - Owen K et al., 2016

"A more realistic approach from the government about what is possible. Instead of constantly beating GP's alongside poor investment in primary care, looking at what we do and how well we do it should be championed. The BMA needs to do more actively to support the profession and the RCGP too.... It is not acceptable to raise expectations of the public saying we can open all hours and weekends. If the money is not there to pay us and to have enough manpower in the practice then they can't have the service" - Dale et al., 2015

"Increasing activity and creating new initiatives to meet targets can lead to poorly designed and overlapping services that fail to improve outcomes (Tan and Mays 2014). Evaluation of the Advanced Access programme used in general practice in the early 2000s found that not only did most practices not meet the target, the over emphasis on rapid access interfered with providing access to appropriate care (Salisbury et al 2007)." - Brooks et al., 2022

"Generalist care is undermined by systemic and bureaucratic devaluing of GP time and expertise, primary care policy that fragments care and encourages short transactional encounters and an excessive focus on disease or procedures. Clinical decision making in the face of uncertainty, undifferentiated symptoms, conflicted goals or chronic disease and grief is also cognitively, emotionally and morally difficult. In order to protect GPs from doubting the value of their work, it is important to understand generalism as so much more than biomedical service provision: it requires philosophical attitudes, overarching priorities and practical skills that are often unnamed and unnoticed." - Lynch et al., 2023

"Dr S quickly cited the longstanding and relentless time-squeezed work pressures, now yoked in an ineptly disincentivising pensions arrangement. Surely adequate money and resources are essential for any competent and compassionate health service: we have seen how they must, again and again, be defined and fought for. So, this is an ongoing battle. Being unavoidable, it will necessarily return to our analysis and debates. Yet, however essential this theme is, it often obscures another, one that is equally essential but more nuanced and less quantifiable, so much less discussed — our loss of community. His otherwise very pertinent observations about money, workload, and resources omitted an equally important and longstanding fact: that GPs generally no longer get the erstwhile deeper, personal work satisfactions that can — and often did — infuse, nourish, and sustain a vocational working lifetime." - Zigmond, 2023

"One GP felt that the Quality Outcomes Framework (QOF) system was not as profitable as some (younger) GPs may believe, and there was a general sense that pursuing QOF points distracted from patient care. 'You've come to me with a bad cough and a bad chest… I'm seeing you thinking: I need to listen to your chest; do you need antibiotics? And then I'm thinking: I'm going to lose my targets—do I know your weight…height… blood pressure…have you had your blood sugar done?

Rather than dealing with you as a person, I'm thinking: heck we've missed a target on this one and it's going to cost the practice.'" - Sansom et al., 2016

"Six GPs described feeling that politicians were using the NHS for their own gain, and felt threatened by Government proposals and changes (eg, 7-day access to primary care). This was experienced as stressful and demoralising. The threats from politicians to change our working hours (are) so stressful and demoralising…we can't see how we can make that work without our work-life balance being fairly comprehensively stuffed…that is an instant turn-off and an instant major demoraliser for GPs who are already overworked and struggling." - Sansom et al., 2016

"Financial incentives are tempting for policy-makers and commissioners to use, particularly for 'contractor' professions such as general practice or dentistry, as even small changes in cash flow can have an impact on small businesses. However, the English NHS is an international outlier in the extent to which it has, over its history, used financial incentives to drive improvements in general practice. The danger of this approach is that there is a significant risk of perverse incentives. By using financial incentives and targets, particularly at national or regional levels, it's possible to distort priorities and hinder practices from addressing the priorities that staff and patients feel are most important." - Brooks et al., 2022

3. **Reciprocal caring: mutual acts of compassion**

**If a patient feels they know or are known by a practice or particular clinicians (C), they are more likely to trust and feel cared for by them (M), maximising collaborative preparation (e.g. pre-empting problems), facilitating patient care for practitioners, and minimising a sense of crisis and perceived urgency of need in relation to their health concerns (O).**

"Patients can be a source of behavioural confirmation when they show their appreciation for the care their GP has provided. There is a positive relationship between patient satisfaction and GP job satisfaction. Furthermore, some of the main sources of lower satisfaction for GPs are increased demands by patients and patients' expectations. GP job satisfaction is higher for GPs who have more satisfied patients." - Stobbe et al., 2021

"GPs in countries where patients are listed with a specific GP or practice, have more opportunities to receive behavioural confirmation from their patients. The list system allows GPs to get to know their patients and the patients' families better, resulting in better continuity of care. In such systems patients are more likely to be familiar with a particular GP which is associated with higher patient satisfaction with care. GP job satisfaction is higher in countries with a patient list system." - Stobbe et al., 2021

"GPs said job satisfaction directly relates to the quality of the doctor-patient relationship, with more time available for GPs to spend with their patients being associated with better job satisfaction. GPs with high job satisfaction describe feeling supported by good practice relationships, while GPs with low job satisfaction describe low morale and feeling unsupported." - Long et al., 2020

"In terms of containment, this was characterised by the size of practices, viewing small or single-handed local practices as enabling a volume of work that could be managed within the confines of a 9-to-5 job. For example, Jenna's (ST3) expectations of general practice were based on work experience in a local 'nice, small practice where it was a single-handed GP' where 'the patients all knew us really well, there was a patient who used to bring us all a bottle of wine every Christmas…and really sort of respected the GP'." - Laverty et al., 2023

"It was the belief of participants that contained and focused work in the past allowed GPs to see the outcome of their clinical work in the relationships and gratitude from patients - they received external recognition that their work was of value. The new GPs, in contrast, reported that the relationship with patients has become less reciprocal. Many participants were concerned about a rising complaint culture and demands outwith traditional doctoring work, as expectations of GPs have shifted. For example, new GP Harry (interview) recounted an incident where he felt he had gone out of his way to help a patient, only to receive a complaint: I brought this patient in at the end of my surgery as an extra, I'd spoken to her on the phone and said, come down and I'll check you out, we'll see if you need antibiotics or not. I saw her, a nice normal conversation in the consultation, examined her. I was like, on balance, I think you've got the flu, I don't think antibiotics are going to help here. Paracetamol, ibuprofen, stay hydrated, sleep when you feel tired. And, she put in a complaint." - Laverty et al., 2023

"Being part of the community was another important satisfaction issue [15, 16, 19, 21, 22, 26]. "GPs in smaller communities were slightly more satisfied with the amount of responsibility they had and recognition they received for good work" [19]. A qualitative study, conducted in Nebraska, highlighted the GPs' relationship with the social community: "You are a very important part of the community and your opinion is listened to" [26]." - Le Floch et al., 2016

"Positive feedback from patients was a frequently mentioned confirmation that doctors' efforts had been appreciated, with one defining his job as a 'privilege' and another observing that for some patients he could be 'the second most important person in their life'. Some related incidents when they had been active advocates for particular patients for whom they felt the NHS had failed to provide adequate care. In a practice with a high proportion of older patients, a GP's investment of time to 'really listen' allowed her to better understand how to juggle conflicting treatment plans." - Spooner, 2016

"Building trust seemed to be key in enabling engagement. Most professional and lived experience participants highly valued continuity as a strategy to achieve this. "Trust is won in small moments, isn't it, over a period of time? In terms of structural things that facilitate that… continuity really matters. Yeah, it does, particularly for this group of patients."- GP5." - Potter et al., 2023

"[W]hen you are ready, you're gonna go to that person 'cause you've built that trust along the way. I trust that they're not gonna railroad me into something"- BGW2 - Potter et al., 2023

"Healthcare professionals who deliver enhanced care to marginalised patients highlighted the importance of proactively contacting marginalised patients. They emphasised not being too pushy, and listening to patients' priorities first before using opportunities to offer assessment or management that might be of the healthcare provider agenda. Proactive, person-centred communication was valued by both professionals and patients, it offered a way to show compassion, and a way to build relationships." - Potter et al., 2023

"I text people a lot… sometimes I'll just send a text saying, 'I hope you're okay'… especially when people aren't engaging, just keeping that relationship going and then when you meet up with them, they will sometimes say, 'oh you know, I like getting your texts, thank you'."-NURSE2 - Potter et al., 2023

"Co-producing service-improvements encouraged empathy, human connection and was rewarding to professionals and marginalised patients. Patients and staff were enthusiastic about the benefits of the changes they had made and outcomes for patients, primary care and collaborating organisations." - Potter et al., 2023

"A couple of lived experience participants reflected that their experience of collaborating with general practice staff helped them empathise with them and feel more comfortable and empowered to engage with general practice." - Potter et al., 2023

"it's actually kind of broken down my own barriers towards GPs… it's made me a little bit more confident to speak up and sort of put my view across and know that I'm able to do that; this is just another human being I'm talking to" -BGW1." - Potter et al., 2023

"General practice participants highlighted the value of meaningful collaboration between those with lived experience and providers in improving contextual understanding. Collaboration improved empathy, challenged assumptions, enabled those with lived experience to feel listened to, and increased the relevance of services to the local population. This new way of working had a different balance of power which general practice participants found insightful and constructive." - Potter et al., 2023

"Over the years he has enjoyed looking after his patients' children, and their children's children. His patients send him birthday and Christmas cards, and their relatives ask him to give readings at their funerals because he knew them so well. "I see myself as a family GP. I enjoy that caring bond with my patients. So many people have no one they can talk to. My patients can talk to me." - Oxtoby, 2022

"I soon realised that I was enfolded into a mindful and concerned network that had anchorage in personal familiarity. The patients, professional, and support staff together functioned largely through shared experiences and individual understandings. From these grew bonds of trust, support, and affection. Such relationships were the threads weaving a nexus of care: how we could both look after, and look out for, one another. Yes, occasionally such benign webs broke — there would be shards of error, misunderstanding, and grievance, even more rarely, bad faith. But mostly the webs held." - Zigmond, 2023

"The simple symmetry of help given and help received." - Morland, 2022

"… who bakes sponge cakes for neighbours and give their GP a tin of quality street at Christmas." - Morland, 2022

4. **Imbalance between work demands and resources available**

**When there is an imbalance between demands (e.g. workload), and resources available (e.g. finance, peer, and organisational support) (C), GPs may be more likely to disengage (O), as they perceive the quality and/or nature of their work as unsustainable (M).**

"Thematic analysis identified four key themes: low morale, disconnect, unmanageable change, and lack of personal and professional support." - Andah et al., 2021

"The impact of discrepancies between need and resources was evident across the dataset. Examples included services being moved into primary care without the equivalent additional supportive resources, and a failure to increase the availability or duration of appointments to meet the needs of an ageing population with complex needs." - Andah et al., 2021

"Lack of support at a personal and professional level for doctors was highlighted across the dataset, with increasing levels of burnout and poor mental health among doctors on the rise." - Andah et al., 2021

"Workload is frequently cited as negatively impacting on commitment to a career in general practice, and many GPs report that their workload is unmanageable." - Croxson et al., 2017

"Most participants described long; intense days spent in general practice. Typical working days ranged from 10-14 hours, often with little or no time for breaks." - Croxson et al., 2017

"It's not just that you're working long hours. The thing which [sic] massively changed is the intensity of when I'm here. What you can't explain is the fact that, within every moment of every day, you know, is that the numbers of tasks, and your multitasking capabilities, have had to increase.' (GP29, male partner, 10 sessions per week, 11-15 years' experience, medium-sized semi-rural practice)." - Croxson et al., 2017

"A full-time career as a GP partner was felt by nearly all participants to be unsustainable." - Croxson et al., 2017

“GPs with good team support and particular consultation styles also felt less negatively.” - Croxson et al., 2017

“Reports of working 12-14-hour days and additional unpaid administration sessions were commonplace. Patient demand for urgent on-the-day appointments was described as unmanageable, and practices also struggled to meet ‘non-urgent’ demand within reasonable timeframes.” - Jefferson et al., 2023

“GPs with poor job satisfaction described feeling overworked and unsupported with negatively impacted doctor-patient relationships.” - Long et al., 2020

“Once job satisfaction has become negatively impacted, the combined pressure of increased patient demand and workload, together with other stress factors, has left many feeling unsupported and vulnerable to burn-out and ill health, and ultimately to the decision to leave general practice.” - Long et al., 2020

“GPs experience working as a GP as ‘undoable and unmanageable’. Many GPs are experiencing working as a GP as undoable and unmanageable due, among other reasons, to high/increasing administrative workload and high/increasing patient demand (both in the number of patients, and their complexity and higher expectations), together with a perceived lack of training and resources to cope with these pressures.” - Long et al., 2020

“'It comes back to the GP numbers again … I know there’s some days I’m going in that I’m doing the job of two people … When I have medical students … I mean I’m 56 years old — I often leave them ten metres behind me because I’m moving, physically moving so quickly … and that sort of pressure, the pressure of time that you’re working under.' (SH03)” - Kidd et al., 2023

“'... we’ve got too much on our plate just now to do [the clusters] justice.' (SH03) '... you can't get the existing workforce to do more than it’s doing now. So, if you want the cluster leads to do more, which I think we need, then you know, we still need other GPs to do the work they're not doing.' (SH01)” - Kidd et al., 2023

“As there are not enough doctors working in general practice, many of the other aims of the GP Forward View are imperilled. Without sufficient GPs, it is impossible for the service to provide extended hours across the board, continuity of care, or support to relieve the pressures of secondary care. The resulting workload for the GPs that remain in practice is not only intense, but also limits capacity to investigate and apply for or initiate potential lifelines offered in the rest of the GP Forward View. Ultimately, this leads to more doctors leaving the profession.” - RCGP, 2018

“UK general practitioners (GPs) are suffering from an unprecedented degree of stress, with 59% describing their work as extremely or very stressful; the highest levels described internationally [1]. Whilst there has been an estimated 16% rise in workload between 2007 and 2014 [2], the workforce has been diminishing [1,3], shrinking by a further 4.5% in the past 3 years [4] despite initiatives to increase the number of doctors in general practice [5].” - Napier, 2019

“The necessity to adjust working hours to manage serious overwork and the worsening imbalance of work and life during the pandemic typically results in missed experience and leadership opportunities for women and pay penalties over and above missed hours.” - Woodhams, 2015

“Job dissatisfaction is most influenced by work-related variables. In particular, these include increased workload intensity and volume to meet the requirements of external agencies, having insufficient time to do the job justice, increased administration and bureaucracy, increased demand and expectation from patients, increasing work complexity, lack of support from colleagues, lack of professional recognition, and long working hours.14,39,40,42.” - Marchand & Peckham, 2017

“More recently, adverse publicity from the media, changes imposed from local primary care organisations, and insufficient resources within the practice have all increased job dissatisfaction.” - Marchand & Peckham, 2017

“There is evidence to show that increased work stress and work intensity leads GPs under ‘high strain’ to report higher levels of anxiety, depression, and dissatisfaction than GPs under ‘low strain’, and that the health impacts of stress continued outside of work, which in turn could increase job dissatisfaction and intention to quit the profession.” - Marchand & Peckham, 2017

“There is some evidence that job autonomy, the variety of work, the feeling of doing an important job, social support, and a good practice environment positively affect job satisfaction.14,39,47 However, GP surveys suggest that a number of these attributes have changed between 2012 and 2015; such as the autonomy in deciding how to do their job and what work to do, the variety of work, and flexibility of working.” - Marchand & Peckham, 2017

“All of the case study CCGs faced increasing demand due to an ageing population and patients with multiple and complex needs, which led to an increased workload. These challenges, along with GP shortages and financial pressures put the local health systems under immense pressure meaning patients did not always receive the quality and standard of care they needed.”- McDermott et al., 2019

“In other surveys conducted between 2014 and 2016, the proportion of GPs in the UK saying they would leave general practice within 5 years varied from 29% to 42% in different regions of England. The most recent (2016) UK survey, of GPs in the South West of England, showed that 70% intend to either quit, reduce their work hours or take a career break in the following 5 years.” - Long et al., 2020

“‘The culture of the NHS is being negatively impacted by the fact that our people are overstretched’. This is evident from the 2018 NHS Staff Survey where more people have reported bullying, harassment and abuse in their workplace in the last 12 months. The theme of staffing pressures causing stress and burnout also runs through the recent Health Education England report on NHS staff and learner wellbeing, which sets out some of the most serious causes of harm to our people's mental health and wellbeing." - Interim NHS People Plan, 2019

"Barr and Gray (2013) previously described how when doctors are faced with the stress of problems beyond their traditional role, they take one of three possible actions: 1) set aside their problems, 2) go beyond their role at risk of increasing stress and overload, and providing less than optimal care; and 3) work in partnership with other professions to spread the load and respond more fully to range of needs." - Gray, 2020

"However, despite this political noise, there is still a recruitment and retention crisis in the profession, with 39% of GPs indicating a high likelihood of leaving direct patient care in the next 5 years, with this rising to 62% in GPs aged >50 years. This is on a wider background of retention challenges in the profession generally, with only 47% of newly qualified doctors exiting the Foundation Programme entering any kind of speciality training." - Lawson, 2020

"Over my 20 years as a GP demand for appointments, and particularly their complexity, has increased beyond recognition. There has been a steady rise in patient expectations, a target driven culture and a growing requirement for GPs to accommodate work previously undertaken in hospitals, or in social care. This has resulted in unprecedented pressure on practices, which impacts on staff and patients." - GPFV, 2016

"So, if anyone ten years ago had said: 'Here's what the NHS should now do - cut the share of funding for primary care and grow the number of hospital specialists three times faster than GPs' they'd have been laughed out of court. But looking back over a decade, that's exactly what's happened. Which is why it's no great surprise that a recent international survey revealed British GPs are under far greater pressure than their counterparts, with rising workload matched by growing patient concerns about convenient access." - GPFV, 2016

"In the General Medical Council's National Training Survey 2023, two thirds of trainees in the UK were found to be at high or moderate risk of burnout, the highest level since tracking began in 2018 A survey of GP trainees in 2023 found many were working beyond their contracted hours to keep on top of their workload. The most common response to the question about career plans one year after qualifying was to work five or six clinical sessions per week out of a possible 10. The report suggested this could be because of the additional unpaid hours of work required to meet the workload." - Garratt, 2023

"49.9% of GPs are satisfied with work, 92.3% report pressure from 'increasing workloads', and 46.0% of GPs plan to leave within the next 5 years. GPs with increased work stress and intensity report higher levels of anxiety, depression, and dissatisfaction, leading to poor WLB, with consequences of deteriorating health and burnout, perfunctory patient care, and increased healthcare system costs." - Ferguson et al., 2020

“In Wessex an optional WLB questionnaire was added to an online appraisal platform, to encourage doctors to think about their own WLB. This commenced in June 2018, and it has proved to be popular, being used in the first year by 2602 GPs (49% response). The quantitative results from the first 6 months (n = 1046) were used for statistical factor analysis, indicating that 54% of GPs responding have significant ‘work-overload risking burnout’.” - Ferguson et al., 2020

“I think it’s so multifactorial, I don’t think there’s any one thing. I think it’s that combination of increased work with decreasing income with high patient expectation with continuous media negativity and no support from the government, just all of those things.” - Doran et al., 2016

“‘Before getting to the point where I really thought I was going to burn out and really hit a very low point mentally and psychologically, I thought actually, I think I recognised those warning signs, and I thought it better to go do something different at this point while I still have the wherewithal to go and do it’. Personal factors were cited by 90.8% of survey respondents, in particular, feeling overworked (53.5%), a wish to improve their work-life balance (49.3%), the work being too stressful (43.0%), and lack of enjoyment of the work (41.5%). - Doran et al., 2016

“In a recent BMA survey, 80% of 1000 responders rated work pressure as ‘high or very high’, with their main workplace stresses being: meeting patients’ demands, lack of time, and excessive bureaucracy.” - Doran et al., 2016

“Doctors, in particular, have seen substantial changes to their work: having to work harder, longer and more intensely with reductions in expected autonomy, deference and respect.” - Laverty et al., 2023

“Participants expressed frustration that having to multi-task administrative work, complex referrals, follow-ups and additional clinical tasks from hospitals (Charles-Jones et al., 2003) took them away from patient-facing work and left them feeling stretched and unable to work in the way they want:
“I just want to feel like when people come to see you, you think if it was you or your relative or anyone else, you want to look after them the best you can and I hate feeling, like, I can’t do that. (Daria (new GP), interview).” - Laverty et al., 2023

“The present experience for trainees and new GPs was one of accelerating pressure with increasing volumes of work, intensification of multiple work tasks and loss of workplace security, resulting in an embodied experience of being harried (Wajcman, 2014). These pressures are perceived to have accelerated over the past decade and did not have an end in sight: A combination of much more work and less staff, so that the balance has tipped - from happy GPs that have gone, and the sort of overworked GPs are left. And I suppose that situation keeps getting worse, doesn’t it, because people try it out and then it’s not a very happy environment. And then it just gets worse and worse. (Lydia (new GP), interview).” - Laverty et al., 2023

“A good income is important for every GP. However, the pitfall is that GPs with higher income rates were less satisfied due to their heavy workload. The challenge is to find the right balance between them [16]. This theme is important for all professions and can be approached from a negative perspective when overwork leads to burn out, or in a positive light when the workload and income are appropriate. A good work-life balance reinforces GPs’ job satisfaction.” - Le Floch et al., 2016

“Almost all respondents (97%) felt the NHS has a culture of viewing excessive stress and workload as the norm. Three themes emerged from qualitative analysis: negative workplace culture; high workload and lack of resources; and generational change.” - Dominic et al., 2021

“There are problems at both ends of the GP career path: older GPs are retiring early or reducing their clinical commitments; newly qualified GPs are choosing locum work or portfolio careers in preference to permanent GP partnership. Working less than full time is common. Various reasons have been given for this: low morale, increased workload and work intensity, and pension changes. The UK has attempted to recruit GPs from abroad to address the deficit, but large international campaigns attracted fewer than forty GPs rather than the target of 500.” - Cunningham et al., 2021

“A high proportion of GPs—around two in every five— currently working within direct patient care in South West England reported an intention to permanently quit direct patient care within the next 5 years, this being one in five within the next 2 years. [...] The majority of responding GPs in our study reported low morale, with less than 15% reporting high morale. GP morale was identified as an important predictor of future career intentions, particularly when morale was very low.” - Fletcher et al., 2017

“Challenges for healthcare teams working in areas of severe socio-economic deprivation in Scotland are well documented and include: premature multi-morbidity [...]; increased GP stress; lower patient enablement; and a relatively older cohort of GPs approaching retirement, such that the GP workforce ‘crisis’- traditionally associated with remote and rural areas - is also felt particularly acutely in urban deprived practices.” - Dhanani et al., 2022

“The overwhelming majority of participants (97%, 701/720) believed that the number of GPs in the UK is much less or somewhat less than patient demand. A similar number- 93% (667/ 720)-perceived the number of GPs in the community in which they practised to be much less or somewhat less than patient demand. Nearly 4 in 10 GPs (38%, 272/720) reported that they were somewhat dissatisfied or very dissatisfied with their career, and 38% (269/720) reported burnout. These responses were similar to other reports on physician burnout in the UK [26]. There was a strong correlation between respondents’ level of reported satisfaction and burn out ($\rho = 0.511$, $p<0.001$)” - Blease et al., 2018

“There is a risk of moral distress in healthcare: the experience of knowing the right thing to do while being in a situation in which it is nearly impossible to do it (33).” - Potter et al., 2023

“The job stressors with the highest scores are: increased workload, changes to meet requirements of external bodies, having insufficient time to do the job justice, paperwork, increased demand from patients and long working hours. Key job stressors that increased between 2012 and 2015 are: adverse publicity by the media, finding a locum, changes imposed from the primary care organisation, and insufficient resources within the practice." - Peckham et al., 2016

"GPs agreed that a means for achieving good resilience was, for many of them, to reduce their clinical working hours. Many had already reduced their clinical work or planned to do so in the future, or reduce clinical hours still further. Participants described the need to reduce clinical hours to be able to make it through their working life to retirement." - Martin et al., 2022

"Another facet was that resilience may be based on surface acting as part of the professional persona or duty of a doctor. GPs felt it important to demonstrate resilience, outwardly disguising their exhaustion and negative emotions. The conclusion is that there is potential shame and stigma about being seen as not resilient or weak in some way. Has having low resilience become synonymous with the failure of the individual GP rather than an acceptance that GP work demands are unmanageable for most?" - Martin et al., 2022

"The increasing pressure arising from the prevalence of comorbiditycomorbidity was a feature of many of our interviews with GPs: "These are people who, as we've controlled all their risk factors over the past 10 or 15 years, they've lived longer and gathered more long-term conditions along the way as well... So, diabetes, hypertension, COPD [chronic obstructive pulmonary disease], asthma - all those major disease groups clustering together in the elderly with less support. Looking at my consultations today, most people had at least two problems to deal with and there's two or three in there who have four or five problems or more." - Baird et al., 2016

"There's definitely a shift towards expecting issues to be dealt with in one meeting as it were. There's little tolerance these days of having three or four visits about the same thing or giving things time to evolve to allow us to, sort of, address how we approach a problem. Many expect all their problems to be solved within a single appointment alongside sometimes unrealistic demands regarding referrals/investigations/treatments, etc." - Baird et al., 2016

"Our study suggests that GPs in England feel additional pressures across all domains of practice compared with their Scottish colleagues. While working together collaboratively as a PCN may increase satisfaction by engendering a sense of local community and support, it seems likely that overall satisfaction will only increase if perceived negative job attributes and job stressors also reduce." - Hayes et al., 2020

"The latest British Medical Association (BMA) tracker survey (BMA, 2016) revealed that 73% of GPs describe their workload as unmanageable (twice the number compared with consultants); 71% of doctors are suffering or at risk of burnout; 45% have considered working part time; 42% retiring early and 25% leaving the profession entirely. The reasons for this are multiple and include increasing volume and complexity of workload, the secondary to primary care shift, recruitment and retention challenges, changes in other services such as community nursing, mental health and care homes creating additional pressure on general practice, increasing non-patient facing administrative duties and a declining trajectory of NHS GP funding." - Hamilton-West et al., 2018

"Studies using the MBI provide further evidence of a rapidly developing crisis in NHS general practice. A study of 564 UK GPs in 2012 found high levels of emotional exhaustion in 46% of respondents, depersonalization in 42% and low levels of personal accomplishment in 34% (Orton et al., 2012). An online survey of 2230 UK GPs in 2015, using the same tool, showed a worsening picture with emotional exhaustion affecting 74% of respondents and a conclusion that 50% of GPs were at risk of burnout (Matthews-King, 2015). Considerable evidence shows that physician burnout can increase medical errors and adversely affect patient care (Prins et al., 2009; Shanafelt and Dyrbye, 2012)." - Hamilton-West et al., 2018

"My wife and I both took early retirement from general practice, in our late 50s. The strain of trying to be 'all things to all people' was just getting too much. We were worried that we would start to make mistakes by just having too many balls to juggle. [...] Many GPs are using their own strategy now to keep their sanity; 'RLE': Retire, Locum, Emigrate. A senior colleague recently said to me that the government has taken general practice back to the 1960s. What is proposed is too little, too late, but may just possibly make the public think something is being done until the election in May." - Glasspool, 2015

"Primary care delivers 90% of NHS activity through GPs. At present, however, GPs describe a highly demanding and stressful work environment. Issues include high workloads, lengthy working hours, and sustained cognitive and emotional challenges. Although many derive joy, meaning, and satisfaction from their work, many also report high levels of stress and job dissatisfaction, and up to 50% experience burnout." - Cheshire et al., 2017

"Only 33% of GP trainees reported that they would become a doctor if they had the chance to go back and 34% stated they would consider specialties other than general practice. Fewer than half reported that they would recommend general practice to junior doctors. Reasons included workload, job-related stress, government policies, long hours, bureaucracy and administrative burden, high patient demand, and threat of litigation." - Simon, 2018

"However, many felt compromised in their ability to practise in their preferred ways (eg, in the length of time they were able to spend with patients) and this impacted negatively on morale, professionalism and identity. This was partly due to: unrealistic demands; concerns about complaints; targets and guidelines; complexity of cases and lack of time to address them; lack of continuity and loss of professional autonomy. GPs felt compromised in striking a balance between delivering high-quality care and doing this within the constraints and burdens they experienced: I like to listen to people, I like to give people time, so I'm a very popular GP, but I'm quite a stressed GP in the NHS setting." - Sansom et al., 2018

"Two GPs noted that the demand for patient care was outstripping supply. Contributing factors included: unrealistic patient expectations; an increase in the number of patient contacts without a corresponding increase in the number of GPs; and additional workload from secondary care. Other demands in addition to contact with patients were felt to compound the problem, including: the need to meet targets, to stay up-to-date with new guidelines, and to prepare for Care Quality Commission (CQC) visits." - Sansom et al., 2016

"One GP felt strongly that managing non-specific and irrelevant emails (sent primarily from CCG colleagues and the health authority) added to workload. Several of the GPs noted that they looked at their work emails from home/in non-work time in order to try and keep up-to-date. Seeing an inbox with 100 new emails…is enough for me to switch it off again and concentrate on patient care for a while." - Sansom et al., 2016

"Participants described concerns about the safety of practice and the quality of care being delivered to patients. This arose from having to manage imposed 'unmanageable' workloads, and focused primarily on the large number of decisions that had to be made, the complexity of cases and the impact of accumulative decision-making throughout the day." - Campbell et al., 2019

5. **Resource/work mismatch**

**When GPs consistently have high-workloads and time pressures with limited resources and respite (C) they are at risk of burnout (O) because they feel out of control and overwhelmed (M).**

**When GPs persistently see patients with complex conditions without adequate time (C), they become dissatisfied with their work (O) because they feel unable to deliver good quality care (M).**

"Dominant messages emerging from contemporary general practice paint a bleak picture of a workforce crisis arising from unsustainable workloads, dissatisfaction, and insufficient recruitment.[1-3]." - Spooner, 2016

"Detailed analyses of workforce demographics and patterns of retention confirm a significant and ongoing shortfall in the number of GPs available to support delivery of accessible and high-quality primary care.[4]." - Spooner, 2016

"Although many GPs list their ability to develop relationships over time with a patient as an attractive aspect of general practice, current workload levels and workforce shortages impede their ability to realise this. In the UK 93% of GPs have stated that heavy workloads negatively affect the patient care they provide.[7]." - Jeffers & Baker, 2016

“Meaningful communication about advance and end-of-life care planning with patients was considered essential but depended on time and resource in primary care that was often in short supply, with staff struggling to create care planning opportunities and manage emotional demands.” - Nicola Turner et al., 2023

“The additional GP work required to fill this gap was absorbed into routine everyday work, unrecognised and under- resourced, by the wider health system: ’I think, as I said earlier, a lot of the increased workload in this kind of context is completely hidden and therefore unmeasurable and therefore unrecognised and I think it’s a dream to think there will ever be any additional funding to recognise that kind of work, it’s never going to happen.’ (GP5, F, aged >50 years)” - McCallum, 2021

“The analysis suggests that, in the context of high socioeconomic deprivation, general practice comprises several lines of ’work’, stretching far beyond the biomedical management of illness. Neither the hidden everyday work, nor the additional emotional work, were perceived to be recognised or resourced within the wider healthcare system. While current strategies for managing workload primarily focus only on reducing illness work, the research suggests that strategies that support and resource the additional and varied nature of work are required. Supporting factors that appear to build resilience are necessary to support GPs, and prevent burnout, particularly in areas of high socioeconomic deprivation.” - McCallum, 2021

“Trying to do more work than could safely be done in the time available was very stressful. The public and government now expect perfection. The threat of litigation was very wearing (male, General Practice).” - Smith et al., 2017

“I found the continuing long hours, despite working part time, caused continual tiredness and exhaustion and I was concerned that this would affect my competence (female, General Practice).” - Smith et al., 2017

“‘I fought and fought and fought not to go off sick, because the guilt associated with going off sick is so huge. […] Everybody’s struggling, so if I go off sick, I will just make everybody else struggle even more. But it got to the point where it was making me quite ill.’ (P24, female, partner)” - Spiers et al., 2017

“The demoralization afflicting these GPs arose in a working context in which there is declining support for an increasingly demanding role, leading to job strain. GPs have continued to gain responsibilities, but with less job control in clinical, leadership and administrative aspects of their role. It is hypothesized that increasing job strain leaves workers with a sense of helplessness and exhaustion in their attempt to manage overwhelming demands [19], and most of the GPs in this study were retiring with a sense of unfinished business.” - Napier, 2019

“’I normally see people who have multiple conditions, huge complexity and who need multiple appointments that I just don’t have. I probably need to increase my appointment to 15 minutes, but that’s all in my own time. It’s impossible to do a 10-minute appointment. I maybe manage that once every 15 patients.’ A portfolio GP working in the Midland” - Thomas. 2020

The workload of GPs has been increasing as a result of increased bureaucracy, growing patient complexity and increases in medical knowledge. Across the sector there is evidence of increased stress, burnout and mental ill-health in the profession. This is leading to a GP shortage and a workforce crisis. - Thomas. 2020

“Many felt very conscious of the threat of burn-out in trying to meet the kind of standards set by government for full-time GP work. Particularly, the idea any GP could sustainably work nine sessions per week was considered unrealistic at best: “Most GPs are really passionate about their job…but I don’t think working in the way government want us to is sustainable.” A young GP in the Midlands.” - Thomas. 2020

“In May this year there were an estimated 27.5 million appointments in general practice, more than two million more than in 2019 Yet over the same period, the number of qualified, full-time equivalent GPs working in the NHS has declined by nearly 500 from 28,094 to 27,627.1 This gap between demand and capacity leaves GPs working harder and facing more burnout as patients find it harder than ever to see them.” - Health and Social Care Committee, House of Commons, 2022

“Part of the reason that patients are receiving poor access at present is that GPs are facing unsustainable workloads, which increase burnout and make GPs more likely to leave the profession. This creates a vicious circle of workforce and workload pressures for the GPs who remain and worsens patient access.” - Health and Social Care Committee, House of Commons, 2022

“The QOF may have improved evidence-based care standardisation in specific disease areas such as diabetes, but this model is not appropriate for the vast majority of patient contacts where the GP knows that the subtle nuances of ‘whole person management’ are the principal requirement. In addition, individual practices do not have enough time or a big enough management team to deal with the escalating bureaucratic requirements.” -Vorster, 2017

The research, published in the British Journal of General Practice, found that many young GPs felt unsupported and vulnerable to burnout as a result of increases in their workload pressures and changes to the nature of their work. - Rimmer, 2016

“Our informants spoke of the workload not only increasing, but being ‘unboundaried:’ ‘I work very long hours, very intense work, not much in the way of breaks or down time. It’s solid seeing patients and masses of paperwork and then outside of work I have to log on to catch up with paperwork.” - Lyon-Maris et al., 2015

“Informants not only described the clinical administrative work extending from face-to face consultations, but also that arising from visits, telephone consultations, and triaging being layered on top. They described a rising tide of ‘invisible’ paperwork from inside and outside the NHS system, including hospital clinics, out-of-hours, NHS 111, blood tests, radiological investigation results, and insurance reports, all of which is set within a culture of target management and reporting,10 high patient expectation, and increased risk of litigation.” - Lyon-Maris et al., 2015

“The reasons for presenteeism may be complex, and more than simply a reflection of GPs’ unwillingness to acknowledge they are unwell or require sick leave. GPs may continue working because they do not wish to let their colleagues down. Some may feel they have no choice. Faced with workforce shortages and difficulty finding locums, GPs are at times forced to work when they are unwell. All these factors can contribute to an unhealthy tendency to ignore illness and continue to work.” - Spiers, 2016

“And as the GP workforce shrinks, the pressure on those remaining builds. Clinicians respond in a predictable fashion, either by deciding that in order to cope they must work fewer patient-facing sessions, though the 6.6 clinical sessions worked on average each week equates in practice to close to a 40-hour week, or they have to leave the workforce prematurely.” - Marshall et al., 2022

“Although most GPs acknowledge that out-of-hours work is an important aspect of their profession, they generally consider it as very demanding and especially night shifts are a major source of stress.” - Stobbe et al., 2021

“Ultimately, if we are to change the perception of general practice for those on the outside looking in, we need to create a more manageable working day for GPs. At the heart of the conundrum lies reducing the burden of low-value activities that cumulatively challenge delivery of care. This will be fundamental in attracting the increase in workforce capacity we need, and in managing the inexorable rise in patient demand.” - Madan et al., 2017

“Doctors have three essential needs: autonomy, belonging, and competence. Staff who show poor engagement and satisfaction offer poor quality of care to patients, are less productive and efficient, and are more likely to suffer from burnout and leave the profession.” - Staveley, 2020

“However as the volume and complexity of clinical work increases, with concomitant rising administrative and bureaucratic burden, there are reports of rising levels of work-related stress and falling job satisfaction that raise concern about the future of primary care.” - Murray, 2016

“Doctor burnout has been described as a global crisis affecting the quality of patient care2-4 and the sustainability of healthcare systems.5 International literature highlights growing problems with chronic stress and burnout that threatened the mental health of doctors working in primary care settings before the COVID-19 pandemic.” - Jefferson et al., 2022

“My practice would not give me the BMA recommended contract for retainer GPs and the deanery do not check this. At times I feel overworked and just a cheap pair of hands for practice.Would be useful if deanery checked on workloads and stopped practices doing this.” - Lockyer et al., 2016

“The combined pressure of increased patient demand and workload, together with other stress factors such as poor IT resources, negative media portrayal, poor practice relationships, and a ‘bullying’ or ‘blame’ culture, has left many feeling unsupported and vulnerable to burn- out and ill health, and ultimately to the decision to leave general practice.” - Long et al., 2020

“Meanwhile, there has been an additional problem emerging in primary care relating to a shortage of GPs exaggerated by older GPs retiring early and younger GPs becoming burnt out. The reduced number of GPs along with the extra workload required in these areas of social deprivation present a particular challenge to all those involved in reducing health inequities and the health divide.” - Gray, 2020

“Health and social care professionals already working under intense stress are now working in a time of high workload, uncertainty, and anxiety. These compounding factors can inform an increased risk of both error and patient safety. In response, the importance of facilitating resilience of and improving communication between primary health-care professionals working in such challenging conditions cannot be overstated.” - Gray, 2020

“A common reason for poor morale is the daily struggle with growing workload. Much of this is generated by a fragmented system, over which practices feel they have little influence. Our first and most pressing priority must be to alleviate this wasteful burden, which takes away from direct patient care. We know we cannot work any harder, so we have to find ways to work differently. A key requirement for wider system change is the urgent need to identify and eliminate needless workload.” - GPFV, 2016

“Participants did think that a persisting and heavy clinical workload would lead to fatigue and burnout.” - Cunningham & Yeoman, 2019

“When GPs were asked what the hardest part of providing care to marginalised patients was, the most frequent answer was ‘not enough time’. GP participants were conscious of the heightened vulnerability of marginalised patients and referenced really wanting to ‘do a good job’ in providing care for them. Participants had some knowledge of how to do this, but described being limited when simultaneously trying to safely manage high volumes of clinical demand.” - Potter et al., 2023

“If you’re trying to deal with someone with really complex needs in the middle of an absolutely overbooked on-call clinic with 50 calls, and you’re just trying to get through the session safely..., you are going to really struggle to provide the empathetic, whole person care that you might want to provide. So structurally, you need to put clinicians in a place where they’ve got the headspace and the opportunity to be kind and trauma-informed and aware of that person's needs. Otherwise, it's just not fair on either person."- GP5 - Potter et al., 2023

Some participants recognised the potential for overwhelm or burnout in professionals who do not have sufficient capacity for the work, how that can detriment the compassion people feel able to offer and risk staff leaving. "I think that staff are really tired… they're very kind of stretched and I think that impacts on people's compassion… staff burnout is I think quite common at the moment"- NURSE1 - Potter et al., 2023

"In our study, GPs were more likely than HDs (hospital doctors) to cite 'pressure of work' as a reason for retiring, while HDs were most likely to cite working hours and out-of-hours work. These results are consistent with data from surveys of UK doctors of around retiral age. For example, Smith et al.'s respondents stated that workload reduction and shorter hours would encourage them to stay in medicine for longer. [...]" - Cleland et al., 2021

"While UK doctors will no doubt welcome less stringent pension taxation measures, which have just been introduced, interventions that ensure sufficient staff to manage workload and good working conditions are likely to be more effective in encouraging skilled doctors to remain in the workforce than those which address pensions taxation only." - Cleland et al., 2021

"GP practices in more socioeconomically deprived areas receive similar funding per registered patient to those serving less deprived areas. But after adjusting for the increased workload associated with greater health needs for people in poorer areas, GP surgeries in deprived areas are worse off. Analysis of data from between 2015/16 and 2018/19 shows that practices serving more deprived areas received around 7% less funding per need- adjusted patient than those in the most affluent areas." - Fisher et al., 2022

"GP turnover - the proportion of staff who leave a practice in a given time period - is also higher in areas of high deprivation. This could affect quality in different ways. High turnover may undermine continuity of care, affect recruitment and retention of GPs, and negatively impact on the ability to deliver care." - Fisher et al., 2022

"At present patient based admin takes several hours a week, practice management takes several hours a week, yearly new contracts and enhanced services take several hours a week and keeping up to date and appraisal take several hours a week. This is impossible' [...] '12-14 hour days is normal with e-mails and e-portfolio work for trainees and revalidation done in what's left of the evenings and at weekends' - Dale et al., 2016

"Respondents were asked to rate on a Likert scale (1=not important, 5=very important) factors that might be contributing to their intention to leave GP (table 4). Intensity of workload had the greatest influence (mean=4.4) followed closely by volume of workload (mean=4.3) and too much time spent on unimportant tasks (mean=4.0). Lack of patient contact, potential introduction of a 7-day working week and reduced job satisfaction also scored a mean >3.” - Owen K et al., 2019

“Comparing current workload with 2 years previously, 51.0% (470) reported working longer hours with almost all (94.4%; 423) giving increased workload as the predominant reason; 26.6% had reduced their hours of work, with most (72.3%; 172) stating this was due to increasing intensity of workload and for many (29.8%; 71) it was related to stress and mental health. This contrasts with the intentions stated in the previous survey where 21.5% of GPs wished to reduce their hours worked and only 2.7% wished to increase.” - Owen K et al., 2019

“The high volume and intensity of work was highlighted by many (32.0%, n=262), and described as ‘ever-increasing’ and ‘unsustainable’ leading to stress and exhaustion. Hugely stressed and exhausted workforce working at or above maximum capacity both individually and as workplace units.” - Owen K et al., 2019

“GPs report reduced work resources, such as insufficient staff recruitment and retention, and time pressures. GP mental ill-health and work-related stress is also on the increase. Proxy measures for these factors include burnout, which remains high in medical professionals in general and GPs in particular.” - Martin et al., 2022

“Crucially, resilience was about a positive sense of thriving at work, and more than just getting by: ‘I think back to when I was sort of stressed and burnt out. I was not enjoying my job, not enjoying life full stop. And I did not feel like I was being successful in anything that I was doing, it was very much about survival rather than thriving. And I think, for me, resilience is being able to thrive.’” - Martin et al., 2022

“...I have recently negotiated with my practice to reduce my hours from 3 to 2 days per week. While I know it sounds pathetic, I really don't feel I can cope working 3 days anymore. The pressure is tremendous. I get home after a 10h day and feel as if I have been hit round the head with a brick...I am only 40 yet feel burnt out. It is a miserable situation.” - Dale et al., 2015

“There are several Drs I know that are burning out, including a colleague who is on long-term sick leave now for this. This is a wonderful experienced GP who we are now likely to lose forever because they feel they cannot take the pressure anymore” - Dale et al., 2015

“Despite loving the fabric of my job, my patients, and celebrating a superb team of staff around me I have never felt demoralised as I do currently. If there were a realistic financial option of retiring in the next 5 years I would take it, not because I want to stop working but because my current conditions are destroying my health and psychological wellbeing...” - Dale et al., 2015

“GPs worked long days, the pace of work was felt to be very difficult to maintain, and there was a general feeling that the work had become more complex in recent years. Looking after patients is okay. It’s the rest of the massive amount of stuff that’s piled upon us that’s driving me out, like it’s driven everybody else out… lifestyle is zilch.” - Sansom et al., 2016

“The additional workload and stress associated with under- taking revalidation, appraisal and care quality commis- sion (CQC) inspections was a dominant theme among the participants, some of whom found the process to be punitive, unhelpful and at times overwhelming: ‘...a lot of younger doctors do get very stressed about appraisals actually. I mean older doctors do as well, because of all the information collecting and it just feels like, ‘Oh bloody hell, I’m doing the job. Can’t you see I’m doing it alright? Why do I have to prove I’m doing it alright?’” - Riley et al., 2018

“Five GPs reported having clinically diagnosed mental health problems; all were female (though with variation in age and job roles). The following quotation displays signs characteristic of burnout: ‘You’re just filling and filling the bucket, and at some point, it will overspill. And you've just got to hope that you don't miss something really important... So, I want to remove myself from that situation for at least a period of time, just while I rebuild my armour I sup- pose and see if I want to do it again.’” - Jefferson et al., 2023

“A recent retrospective study found steady in- crease in GP turnover over the last decade indicating long-term decline in staff retention (Parisi et al., 2021). The UK has one of the lowest health care expenditure per person in the devel- oped economies (Razai & Majeed, 2021). Further, between 2005 and 2006, the pro- portion of money spent in general practice fell from 9.6% to 8.1% (Razai & Majeed, 2021). The underinvestment in general practice is remarkable, as patient consultations are increasing, the size of the population has grown, and people are living longer with complex and multiple health needs (British Medical Association, 2020).” - Razai et al., 2022

“Patients’ expectation of GP care has increased partly due to well- meaning work of patient advocacy groups, single-issue campaigns, and the idealism of regulatory bodies such as the GMC. Pressure from politicians who have made ambitious promises without a matching resolve to provide funding and resources have also contributed to this problem. However, the expectation of a very high standard of care is far removed from reality of working in under- resourced, understaffed environments with often unmanageable workloads.” - Razai et al., 2022

“Workload in general practice saw a reduction during the initial weeks of the pandemic across the UK. RCGP’s Tracking Survey found that 59% of GPs in England worked over their contracted hours every day before the Covid-19 outbreak, which fell to 39% during the outbreak. This is likely to be largely a result of fewer patients seeking appointments, due to the suspension of routine GP services, fear of infection and patients not wanting to trouble their GP. However, members told us that reduced bureaucracy also helped to reduce workload, as one said, “from 140% to 100%”. The number of respondents to our Tracking Survey who said they had enough time to treat patients adequately, went up from 24% to 38% during the pandemic.” - RCGP, 2020

6. **The shift to remote consulting**

**Despite the assumption that remote work is quicker, the transformation from face-to-face to remote consulting (C), can paradoxically increase GP workload (O), due to increased patient access, expectation/demand, and limited time for GPs to schedule breaks and/or time with colleagues (M).**

“The COVID-19 pandemic has resulted in a dramatic shift from face-to-face to remote consulting within general practice. There have been some important gains in terms of travelling for patients and flexible working for general practice teams. However, there have been losses too. Urgent GP cancer referrals were estimated to be at 42% in April 2020 compared with the year before. Digital first approaches, now widespread, can paradoxically increase overall GP workload and a recent study warns of extra GP work by up to a third.” - Brown et al., 2020

“I have a flask in my room and a little fridge to ensure I keep hydrated, otherwise I wouldn’t get a drink because it’s just so, I can’t pop out. I probably have one wee a day! You forget to wee unfortunately, ‘cos you’re so busy, and it’s the non-stop, it’s the non-stop sort of the job I think that’s the hardest.’ (GP28, female partner, seven sessions per week, 6-10 years’ experience, medium-sized urban practice).” - Croxson et al., 2017

Many GPs’ practices had responded to increasing patient demand by offering alternative means of contact, particularly telephone encounters. Although some found this effective, many felt the increase in telephone contact had contributed to the rise in workload, particularly because extra time was not always allocated towards this:

“Patients think it’s much easier to, obviously to reach us now, so perhaps before they would say: “Oh you know, I’ll wait a day or so.” But now: “Oh, I’ll just ring the doctor, they can give advice.” So, I actually think, funny [sic] enough, it has actually increased our workload.’ (GP18, female partner, nine sessions per week, 6-10 years’ experience, medium-sized suburban practice).” - Croxson et al., 2017

“Patient demand and expectations, as well as population change, were cited as factors influencing GP’s ability to engage with new care models due to the greater demands being placed on GPs.” - Humphrey & Cleaver, 2018

“Workload as a resource was cited as a barrier to the implementation of 5YFV, the broad consensus being that the changes were creating more work and, at this moment in time, working life seemed more complex.” - Humphrey & Cleaver, 2018

"In addition to seeing doctors from a team, practices are increasingly adopting telephone triage. The ESTEEM study saw the number of patients in contact with their GP increase, while the number of patients attending the practice fell.[9] This may have resulted in an increase in relational continuity. However, the study also found that telephone triage may lead to workload simply being redistributed from face-to-face to phone consultations." - Jeffers & Baker, 2016

"Everyone seeking care was required to go through triage and pre- assessment, risking inefficiency owing to 'double touches' with patients (risk 1B)." - Rosen et al., 2023

"The crisis has driven a change in current professional roles and specialisms, the development of new professional roles and a proliferation of technological progress such as with the provision of remote consultation and communication. These changes benefit patients and service users but come with a significant risk that care can be fragmented." - Gray, 2020

"In the present climate, with loss of face-to-face human connection, there is increased anxiety, fear, and bereavement being experienced by professionals, patients, and service users alike. An added problem is the challenge to be compassionate when communicating remotely. Resilience of all involved is vital as well as the ability to live with increased uncertainty. Supporting the workforce is important in this context." - Gray, 2020

"Workload difficulties arose because email is not integrated into practice, time is not allowed for 'extra' consultations and, in some cases, GPs reported that email was generating more work and additional consultations. This was supported by patient accounts that they were consulting about concerns for which they would not otherwise have contacted the practice: ' ... because I think, "Oh, God, I have to make an appointment, I have to go there, so let's forget about it".'" - Atherton et al., 2013

"Both GPs and patients managed the impact of using email consultation by placing boundaries around use based on previous experience: while GPs drew on experience with other methods of consultation, patients drew on social norms. There was recognition that long-term concerns about email consultation use would need to be addressed by regulation and guidance." - Atherton et al., 2013

"Some general practitioner participants reflected that the introduction of remote working had made it easier to schedule multidisciplinary team meetings and had reduced the amount of time spent travelling to meetings and events. However, others reflected that the boundaries between their home and professional life had become more blurred with the perception that you are 'on call the whole time'." - Burn et al., 2022

"Participants expressed concerns regarding their ability to develop strong and reassuring doctor-patient relationships on the phone. Trainees described the difficulties they faced trying to 'build up the trust. . . as a new doctor in that surgery' (GPTr6), reporting that they instead felt like a faceless 'call centre' or 'convenience store of general practice' (GPTr7) for their patients." - Grut et al., 2023

"Triaging, used to determine which patients needed to be seen in person, led some GPs to take a first history of patients over the phone and a second history once face-to-face. This could leave them feeling they were 'wasting [their] time double consulting people' (GPTr7). Suggestions for a 'more streamlined' (GPTr10) service were given." - Grut et al., 2023

"Finally, GPs' comments about levels of burnout raise important questions about how AI might mitigate unsafe workloads. Even if new machine learning technologies aim to augment human clinical decision making, it is unclear whether these tools will alleviate current levels of physician burnout. Conceivably, in the short- and medium-term, if such clinical decision support systems are not suitably designed for human clinical workflow, they may result in "alert fatigue," undermining their utility." - Blease et al., 2019

"The increasing prevalence of mental ill health among the medical profession, including GPs, has been attributed to a number of factors, most notably the escalating occupational stresses related to workload, long hours and the emotional demands of the job. In addition, sharp increases in patient demand and transfer of care from specialist to primary care, as well as reductions in resources, have resulted in the pressure on primary care in England being at its highest ever. This pressure is predicted to increase in the future. Findings such as these highlight the potentially stressful context in which GPs are currently working, leaving many vulnerable to distress and/or mental health challenges." - Riley et al., 2018

"Some participants reported feeling a loss of empathy or compassion fatigue, feeling empty, exhaustion, difficulty making decisions, low job satisfaction and loss of enjoyment for their work, using language which highlights the depth of the feelings: 'I'd lost my empathy with my patients. You know, and er I'm just—I was just—you're just like a ketchup bottle in a production line […] But from a personal point of view I'm still feeling a bit washed out by it, you know, I feel a little bit empty. You know, I feel like I'm burnt out really a bit, to be honest.'" - Riley et al., 2018

"The GP participants described numerous troubling psychological and physical symptoms relating to chronic stress, anxiety, depression and/or burn-out with several breaking down at work—crying, feeling despair and some contemplating suicide. Many also talked about their shame, humiliation and embarrassment at their perceived inability to cope with the stresses of their job and/or their symptoms of mental ill health. The experiences and feelings of many GP participants depict a dispiriting emotional landscape, with notable signs of poor mental health and striking levels of distress." - Riley et al., 2018

"GPs described the importance of triage systems for prioritisation and reallocating patients during staff absences. E-consultation systems were perceived to increase demand due to greater accessibility: 'Now eConsults have come in there's no barrier... there'll be 200 eConsults on a Monday that we have to deal with as well as all the other general practice workload and the vaccination programme and PCNs, and it's just really unsustainable and unsafe.'" - Jefferson et al., 2023

**<u>Relationships across individuals, organisations, and communities</u>**

*Knowledge accumulation: long-term patient-GP relationships*

**7. Cumulative knowledge**

**When GP work and services are designed to accumulate knowledge about individuals (people) and communities (place) (C), it enables more appropriate patient care (O), as GPs can draw upon their expanded and deeper understanding in GP-patient and GP-peer encounters (M).**

“Positive feedback from patients was a frequently mentioned confirmation that doctors’ efforts had been appreciated, with one defining his job as a ‘privilege’ and another observing that for some patients he could be ‘the second most important person in their life’. Some related incidents when they had been active advocates for particular patients for whom they felt the NHS had failed to provide adequate care. In a practice with a high proportion of older patients, a GP’s investment of time to ‘really listen’ allowed her to better understand how to juggle conflicting treatment plans.” - Spooner, 2016

“In Hillingdon, new models of care are being piloted for patients with Multimorbidity (multiple long-term conditions) by running multi-morbidity clinics in primary care supported by a pharmacist, nurse, health care assistant and GP using a year of care approach. The model ensures person centred care as opposed to LTC-specific care delivery. Also, various models of care developed for frail elderly patients using care coordinators and community matrons to work closely with GPs to case-manage patients with complexity who are at high risk of an unplanned admission.” - RCGP, 2018

“The enduring connection to people is what makes primary care so valued by the communities it serves: creating the conditions where we can use integrated neighbourhood teams to support practices by providing personalised care to those people with greatest need, and on-the-day urgent care where appropriate, keeps the connection in place for the future.” - Fuller, 2022

“At a place level, we have seen primary care increasingly working in partnership with local authorities (in particular public health and housing teams), local communities and other local system partners, to pool information and population health data. This means sharing expertise to understand what factors lead to poor health and wellbeing in their communities and agreeing how to work together proactively to tackle these. We have seen this type of joint working become commonplace during the pandemic, where a combination of national data tools, collaboration with local authorities and hyper-local engagement were critical success factors. This enabled teams to try different approaches to outreach and communications, get immediate feedback on what is working, and course-correct accordingly.” - Fuller, 2022

“In areas with a significant ethnic minority population, the time needed to organise and utilise translation services, and explain the pathways through and constraints of the NHS, considerably added to workload.” - Croxson et al., 2017

“This evaluation demonstrates that training around clinician change can be effective in shifting service delivery when sat within a cultural framework that genuinely situates patients at the centre of consultations and practice activity.”- Robertson et al., 2013

“Familiarity with community health resources, sociocultural awareness in patient care, community participation and assimilation, and identifying and intervening in the community’s health problems.” - Marchand & Peckham, 2017

“As a consequence of limited knowledge and activity in the programme, GPs saw the EPCP (Enhanced Primary Care Programme) as peripheral to their main task of sustaining their general practice, and this is a risk to programme implementation and systems transformation. Evidence suggests that there is a need for a prepared and proactive primary care team who have the right information, tools and people, with the correct level of systems knowledge, to provide a satisfactory level of the patient provider relationship.” - Davis et al., 2018

“The opportunity to facilitate more integrated care by applying their knowledge about community resources, and encouraging communication and working relationships across organisational and professional boundaries, was viewed as a significant benefit. It was observed on several occasions that medical staff in urgent care environments approached fellows for advice about community and primary care: “I just say ‘pick up the phone’. They say, ‘the named GP is almost never there’ and I was saying ‘don’t worry about their named GP, [the other GPs at the practice] will have access to the same information’.” - Dale et al., 2017

“Trainers often had longstanding service in the practice and knew a lot about the lives of their patients. GPSTs recognised that a considerable number of patients had spent many years in their practice and had an extended family in the area. Trainers had extensive knowledge of the histories of these extended families and acted as oral historians, able to recall what had happened to patients, their families and the locality, through decades of continuing practice in some cases. “He knows exactly who everybody was, and the relationship and you know the granddaughter who took an overdose and died and the nephew that was murdered and how that impacts on every single one of them and that’s invaluable, it’s amazing.” (Participant 2).” - Cunningham & Yeoman, 2019

“They [senior GPs/GP trainers] were regarded as assets to the local area, that they understood life there, and were long-standing champions for patients and the services needed by them.” - Cunningham & Yeoman, 2019

“Some trainers pointed out the social and economic disadvantages that affected patients and their families. This helped to educate GPSTs and influenced their perceptions on the causation of deprivation and the difficulties patients faced. She was very wise, taught me tolerance and patience.

And giving people the benefit of the doubt, always picked up on when I submitted any sort of cynical comments about the patients. She tried to keep my mind open, and you know be optimistic. (Participant 14).” - Cunningham & Yeoman, 2019

“Participants considered that trainers acted as patient advocates, and although some were cynical at times about patients’ behaviours and outcomes, many remained focused and committed to the deprived area.” - Cunningham & Yeoman, 2019

“In terms of containment, this was characterised by the size of practices, viewing small or single-handed local practices as enabling a volume of work that could be managed within the confines of a 9-to-5 job. For example, Jenna’s (ST3) expectations of general practice were based on work experience in a local ‘nice, small practice where it was a single-handed GP’ where ‘the patients all knew us really well, there was a patient who used to bring us all a bottle of wine every Christmas . . . and really sort of respected the GP’.” - Laverty et al., 2023

“Contained work within small practices was perceived to allow a focus on care-related tasks. The new GPs believed that older GPs were familiar with all of their patients and had autonomy and freedom from many of the bureaucratic restraints they now faced.” - Laverty et al., 2023

“Being part of the community was another important satisfaction issue [15, 16, 19, 21, 22, 26]. “GPs in smaller communities were slightly more satisfied with the amount of responsibility they had and recognition they received for good work” [19]. A qualitative study, conducted in Nebraska, highlighted the GPs’ relationship with the social community: “You are a very important part of the community and your opinion is listened to” [26].” - Le Floch et al., 2016

“Being able to successfully apply one’s mix of competencies in the context of varying patient needs and demands results in being capable to practice safely and effectively (ie doing the best one can do in the current context).” - Sturmberg et al., 2023

“Capability entails being able to apply one’s learnings to new situations.55,56 Capabilities emerge from continuing learning and unlearning, modification and integration of new knowledge as time progresses through one’s career.” - Sturmberg et al., 2023

“This process is described as developing mindlines, which are an important collection of complex knowledge sources that are tested and refined within the social, emotional, physical and cultural milieu of both physician and patient.57,58.” - Sturmberg et al., 2023

“Mastery is defined subjectively as attaining comprehensive knowledge and skills in a particular discipline with a feeling of confidence in one’s abilities. It is defined objectively as the ability to deliver an authentic and worthy performance. Those who have achieved mastery are fluent, even creative, in using their knowledge, skills and understanding in key performance challenges and contexts. It usually takes several years after obtaining fellowship qualifications to reach the validated high-level standards of mastery level performance. Confidence in how a person practises includes knowing how to test and refine knowledge in the face of new challenges and unexpected patient presentations." Sturmberg et al., 2023

"It is therefore important to view ongoing professional development in the light of one's practice characteristics (ie geography, social environment and patient profile), which, in turn, determines one's required knowledge, skills and capabilities." - Sturmberg et al., 2023

"Another group of potentially positive factors was related to the GP and his/her patient contacts and relationships. An efficient doctor-patient relationship was also considered an important factor in GP job satisfaction [17]." - Le Floch et al., 2016

"The unique doctor-patient relationship, while feeling useful and being integrated into the community, makes satisfied GPs. Policy makers must keep in mind, when reorganising the professions in primary care, that the most attractive factor in the profession seems to be the unique doctor-patient relationship, along with the longitudinal care and diversity in the work, which are extremely attractive factors in the profession [17, 23, 25]." - Le Floch et al., 2016

"A strong link with the community and social recognition were also important for increasing the GP workforce. Stakeholders and health system should take this aspect into consideration." - Le Floch et al., 2016

"There's so many services that support people but so few of them are actually able to support people in [general practice]… if you've got one good relationship and somebody can just do that with one person then they've got a much higher chance of being able to communicate what they need"-Ptn2." - Potter et al., 2023

"Building trust seemed to be key in enabling engagement. Most professional and lived experience participants highly valued continuity as a strategy to achieve this. "Trust is won in small moments, isn't it, over a period of time? In terms of structural things that facilitate that… continuity really matters. Yeah, it does, particularly for this group of patients."- GP5." - Potter et al., 2023

"[W]hen you are ready, you're gonna go to that person 'cause you've built that trust along the way. I trust that they're not gonna railroad me into something"- BGW2 - Potter et al., 2023

"GPs were then often referred to as family doctors. This was apposite as we very often engaged with, and therefore got to know, patients as members of families. Yet it was also true in another sense: our practices usually functioned much like happier families — with high levels of personal knowledge, understanding, trust, loyalty, and intelligent flexibility. I felt that this family doctor surgery where I now worked provided me with a secondary family, a home from home. This became my professional home for 40 years." - Zigmond, 2023

"The young doctor relished these longer relationships, pieced together over numerous encounters." - Morland, 2022

"For their extends behind every encounter the doctor has, on every day of her working life, not just a medical history, but also a personal history. A winding corridor of experience and emotion, the patient's whole life. In any given consultation, she's afforded but a fragment of that history. It is one of the joys of being a long serving family doctor in a community like this - that she has both time and opportunity to piece the fragments together over years and across families and generations." - Morland, 2022

"This is what general practice is all about - talking to people, listening to their stories every bit as much as it's about clinical examination." - Morland, 2022

"On days when the administrative paperwork piles high, these stories are what keeps her going. They are the raw material of her relationship with each patient and a source of endless complexity and fascination." - Morland, 2022

**8. Agency/partnership**

**When GPs, patients, and peers have opportunities to shape and co-create management plans and potential solutions (C), this may facilitate patient and practitioner enablement and satisfaction (M), leading to higher levels of peer and patient trust and improved self-management capacity (O).**

"Facilitating the involvement of patients in their own care was a particular goal for one GP who enthused about utilising self-management and clinical decision tools, and making his personal evaluation of their effectiveness:

"I have done a little protocol for home blood pressure monitoring for people … it's really empowered people. I have also done a new thing around a self-management plan for people with comorbidity. I have shown ... how that has changed people's health seeking behaviour.' [Mark]." - Spooner, 2016

"In addition to illuminating current access issues as experienced by patients and staff, applying the new conception of access as the human fit between the abilities of both patients and staff can help to define 'better' access, providing specific targets for change. For example, the data suggest that improving access could mean improving continuity, providing more opportunity to achieve that human fit between staff and patients." - Voorhees et al., 2022

"'I think it re-enthused me to try and explore with some patients what the barriers are to behavioural change. My attitude, I'm afraid had become increasingly, ''well if they had been drinking heavily for thirty years or smoking or whatever, then nothing I say or do is going to have much impact''. But now, I'm exploring with patients what their beliefs and expectations are. So, it's coming at things from a very different standpoint'" - Robertson et al., 2012.

"These changes in perception generated adaptations in practitioner-patient dynamics within consultations as clinicians focused more on patient agendas and priorities. Clinicians reported taking more care with language and phrasing, using more open questions and the importance of listening skills and knowing when to listen. They talked about developing additional skills in gaining patient 'buy-in' to behaviour change by introducing tools such as the 'one-to-ten' readiness to change scale to motivate patients to consider what is important to them and why. This facilitated coworking with patients to set achievable action goals appropriate for the individual patient's personal circumstances." - Robertson et al., 2012.

This seems to represent a conceptual difference in how 'communication' is understood and practiced between the two groups. For those that have not completed the programme, the emphasis is on communicating knowledge and information to the patient. For those clinicians that had completed SSCiGP, while transfer of information remained important, the emphasis was very much on communicating with patients, exploring their condition and possible behaviour change within their own context, so they can make informed decisions that are relevant to them and their life." - Robertson et al., 2012.

"Patient-centred care is characterised by information sharing, respect for patient preferences, and shared decision making. It is care that is accessible, comprehensive, and collaborative, which is aligned with patients' goals, and that respects patients' values, cultural traditions, and socioeconomic conditions." - Jones, 2021

"Across a large research literature, patient-centred care has been shown to be associated with greater concordance with medication and treatment plans following shared decision making, increased patient satisfaction with care, increased wellbeing among both patients and clinicians, more appropriate resource allocation, and cost containment." - Jones, 2021

"In some instances, the achievement of autonomous decisions may mean that, to some extent, the ethical principle of beneficence is undermined, since the 'best' and 'most efficacious' treatment option may be side-lined by decisions based on personal preferences. The implications of this evidence based/patient-centred paradox vary across different health care decisions. Indeed, the selection of a less efficient treatment could potentially result in a greater benefit, as a patient may be more likely to adhere to a regimen of care that coheres with their personal beliefs and lifestyle choices." - Clinch et al., 2013

"As Armstrong (2002), has argued, the transformation of knowledge into action through the evaluative practices of GPs could simply be read as means through which the autonomy of individual clinicians is maintained. That is, skilled interpretive judgements made about what is appropriate for a patient, can be construed as a mobilisation of a therapeutic rationality, which enables the maintenance of the autonomy of individual practitioners in the face of evidence-based medicine." - Clinch et al., 2013

"A recurring context necessary to lead on to GP engagement was identifying and agreeing the unique contribution of GPs within the QI initiative, either from the outset or as an integral part of the work. This worked best when the QI initiative focused on what it was that GPs, and GPs alone, could provide, and structured the approach accordingly to facilitate the input of GPs. [...] GPs helped determine how the intervention would interdigitate with general practice, rather than taking a more overarching leadership role. Innovation was required to design ways of making improvement initiatives fit in, or adapt to, existing systems so that what was asked of GPs was recognised as part of their core role and complemented the wider caseload." - Chadborn et al., 2021

"Healthcare professionals who deliver enhanced care to marginalised patients highlighted the importance of proactively contacting marginalised patients. They emphasised not being too pushy, and listening to patients' priorities first before using opportunities to offer assessment or management that might be of the healthcare provider agenda. Proactive, person-centred communication was valued by both professionals and patients, it offered a way to show compassion, and a way to build relationships." - Potter et al., 2023

"I text people a lot… sometimes I'll just send a text saying, 'I hope you're okay'… especially when people aren't engaging, just keeping that relationship going and then when you meet up with them, they will sometimes say, 'oh you know, I like getting your texts, thank you'."-NURSE2 - Potter et al., 2023

"Lived experience participants encouraged general practice staff to remember 'the quiet ones', patients who might not actively ask for or advocate for their needs and are often left behind by general practices that require people to actively seek care and overcome common obstacles like limited appointment slots. This resonated with several practice staff. "don't forget the people who are traumatised and go quiet and disappear, but at the same time, we mustn't hound people… allowing the patient autonomy and control where possible"- GP1." - Potter et al., 2023

"Both the process of co-production and the service-improvements that were developed (see Table 2) initiated or strengthened teamwork and connectedness both within and between general practice teams. Some participants noted the value of relationships and teams within general practice who were focused on delivering better care for marginalised patients. One participant described relief at feeling part of a wider team of practices also motivated to tackle health inequalities. "it's really kind of encouraging for us to know that there are other GP practices that are kind of passionate about health inclusion that's you know, refreshing and a relief!"- NURSE1 - Potter et al., 2023

"Co-producing service-improvements encouraged empathy, human connection and was rewarding to professionals and marginalised patients. Patients and staff were enthusiastic about the benefits of the changes they had made and outcomes for patients, primary care and collaborating organisations." - Potter et al., 2023

"there is people behind these labels and there is humans and human behaviour and I think we have made the difference. I think we made the difference like in Practice2, I think we made a difference in Area8… I love that and I think that's a massive thing"- BGW2 - Potter et al., 2023

"A couple of lived experience participants reflected that their experience of collaborating with general practice staff helped them empathise with them and feel more comfortable and empowered to engage with general practice." - Potter et al., 2023

"it's actually kind of broken down my own barriers towards GPs… it's made me a little bit more confident to speak up and sort of put my view across and know that I'm able to do that; this is just another human being I'm talking to" -BGW1." - Potter et al., 2023

"General practice participants highlighted the value of meaningful collaboration between those with lived experience and providers in improving contextual understanding. Collaboration improved empathy, challenged assumptions, enabled those with lived experience to feel listened to, and increased the relevance of services to the local population. This new way of working had a different balance of power which general practice participants found insightful and constructive." - Potter et al., 2023

"When you change the power dynamic and have a meeting like that, where we're all on an equal footing, I often find that you have unexpected insights… they've challenged the way that we've thought about things and given us a fresh way of thinking… it's improved things for us as doctors as well as for our patients" -GP1 - Potter et al., 2023

"As occurred in our study, co-production can bring fresh thinking to complex problems (25, 26, 27). Some general practices or networks may wish to try similar highly inclusive methods of collaborating with local support organisations and people with lived experience to develop context-specific service improvements." - Potter et al., 2023

"All successful projects needed at least one staff member who 'championed' the approach and supported the coach (person helping to deliver the intervention): You do need champions … you need an enthusiast who will fly your flag and if there's a bit of negativity … if you've got a positive role model that really helps to get everyone on-board" - Abrahamson et al., 2020

"An external coach (person helping to deliver the intervention) was preferable because they brought benefits as an outsider, including the PPI perspective: I think first of all having somebody who's outside of the practice is vital to it because they keep you on track and they're fresh eyes in the whole process, so they ask questions that possibly a patient would ask whereas we just assume that's what it should be. [...] Our findings identified how the interplay between individual attributes, relationships within the practice, with the coach and CCG, and with other organisations all affected successful implementation." - Abrahamson et al., 2020

"[patient] he'd follow her (GP) to the end of the earth now." - Morland, 2022

"Furthermore, if we accept the proposition that consultations are collaboratively enacted, then we also don't want the *patient* to be as they are outside the consultation either. It is beneficial if, in some way, they play their role too. Not the Parsonian role of the docile patient to be 'acted on', but the engaged patient to 'enact with'. We suggest that by practising subjunctively, clinicians can create the conditions to help patients become what Epstein (2017, 289) terms a "mindful patient", improving potential for a good treatment outcome. This suggests an interpretation of person-centred care in which the patient-person distinction is not seen pejoratively. By both clinician and patient 'acting differently' in a consultation they create a social world which is "temporary, fragile to be sure, but not false – a world where differences can be accommodated, tolerance enacted (if not fully understood) and openness to others maintained." - Hardman et al., 2020

*Connection-rich contexts*

9. **Direct connections**

**Direct interactions and connections (e.g. a phone call with a colleague or patient or filing/actioning results of a known patient), nurture opportunities for meaningful practice, development and use of cumulative knowledge, and agile/adaptable approaches to personalise care (M), enabling GPs to flourish and experience psychological safety in their negotiations with risk, uncertainty, and ambiguity in practice (O).**

"GPs said job satisfaction directly relates to the quality of the doctor-patient relationship, with more time available for GPs to spend with their patients being associated with better job satisfaction. GPs with high job satisfaction describe feeling supported by good practice relationships, while GPs with low job satisfaction describe low morale and feeling unsupported." - Long et al., 2020

"William Osler, master of clinical method and, of course, not a GP, was more on target: 'It is much more important to know what sort of patient has a disease than what sort of disease a patient has." - Jones, 2021

"Another group of potentially positive factors was related to the GP and his/her patient contacts and relationships. An efficient doctor-patient relationship was also considered an important factor in GP job satisfaction [17]." - Le Floch et al., 2016

"Mainstream appointment systems are often inaccessible to marginalised patients who often require rapidly responsive, easily accessible routes into care." "Prioritise flexible access and longer appointments to patients in greater need: One practice dovetailed the care coordinator role with a specialist inclusion health clinic for marginalised patients; in the other the care coordinator had a direct telephone line they provided to patients and priority access to appointments when needed. One practice also had protected appointment slots that could be used by drug and alcohol workers who could use this for one of their clients if needed." - Potter et al., 2023

"Several comments linked appraisal and revalidation together as part of an increasingly intrusive, overarching performance management process that included other processes, such as Care Quality Commission (CQC) inspections. These were described as being part of an overall de-professionalisation of general practice that is being driven by political goals and suspicions, as has been discussed elsewhere." - Dale et al., 2016

"I am a people person and like human interaction and the continuity of care you get in general practice." - O'Dowd, 2022

"All the while, we must maintain a relentless focus on protecting the essence of general practice: the deep-seated connection and continuity at the heart of the doctor-patient relationship." - Madan et al., 2017

"They recommended that, to improve retention of young GPs, "the pace of administrative change needs to be minimised, and the time spent by GPs on work that is not face-to-face patient care reduced." - Rimmer, 2016

"The negative portrayal of GPs in the media, a lack of time for interaction with patients and colleagues, and a perceived blame culture also contributed to young GPs leaving the profession, the researchers found." - Rimmer, 2016

"With more and more of our working hours being taken up with form filling, ticking boxes, and preparing for CQC [Care Quality Commission] practice inspections, we are drowning in red tape, and this only serves to keep us away from delivering frontline patient care, which is why we become doctors in the first place." - Rimmer, 2016

"Some of the organisational changes were considered to have imposed increased clinical and non-clinical responsibilities and work on GPs. Together, such changes were believed to have undermined some of the basic tenets and traditional expectations of being a GP, such as the doctor-patient relationship and having autonomy and control over one's clinical work." - Long et al., 2020

"Lack of time with patients is perceived to compromise the ability to practise patient-centred care and undermines GPs' professional autonomy and values, resulting in further diminished job satisfaction."- Long et al., 2020

"Gupta hopes that his practice will continue to provide a personal service to patients, as that is the reason why most GPs have chosen to do this job. We are the only people in the healthcare system who have that relationship with patients and their families, and it would be a huge shame if that personal approach was lost.” - Oxtoby, 2021

“That willingness to invest in the moment, that instinctive ability to occupy, reflect, and respond to the here and now, both emotionally and intellectually, sits at the heart of the doctor-patient relationship as it plays out in general practice.” - Morland, 2022

“Such ‘families’ extended beyond the nuclear; GPs then got to know kindred professionals — hospital specialists and their secretaries; locality district, health visitor, and psychiatric nurses; probation and social workers; housing officers and charity conveners; local pharmacists — these were all people whose faces, voices, and work we got to know. They were local, our jobs were stable, and, before digital technology, our exchanges were mostly personal, conversational, and direct. In such ‘extended families’ we often felt we were helping one another shoulder the burdens of what can be very difficult work.” – Zigmund, 2023.

“This study has demonstrated that compared with GPs in England, GPs in Scotland have lower intentions to reduce work participation, as well as higher levels of job satisfac- tion, lower job stressors and lower negative job attributes. These differences were of a reasonably large magnitude (one-third to one-half of SD), and thus likely to be meaningful in practice. It is possible that these differences relate, at least in part, to the recent changes in primary care in Scotland, including the new GP contract and the abolition (in April 2016) of the increasingly unpopular QOF.” - Hayes et al., 2020

“This is what general practice is all about - talking to people, listening to their stories every bit as much as it’s about clinical examination.” – Morland, 2022.

“The aspects of work that are less meaningful e.g., documentation, data entry) edge out the more meaningful aspects of practice (time spent talking with patients, working with the rest of the health care team) and lead to a less joyful practice.” - Guck et al., 2021

**10. Depersonalisation**

**When GPs experience reduced human connection with colleagues and patients (C), this can contribute to isolation, reduced sensitivity, and (O), because work becomes depersonalised (M).**

“GPs also described feeling isolated, ‘decision fatigue’ and felt that consultations lacked personal contact with patients, which had encouraged their career choice.” - Jefferson et al., 2023

“At a service level, Lester *et al*[32] described continual restructuring and uncertainty about the future, as well as increased administrative work and decreased time with patients.” - Andah et al., 2021

“GP partners tended to describe longer days than salaried or locum GPs. Many GPs reported the need to do administrative work during evenings, weekends, or days off.” - Croxson et al., 2017

“Although many GPs list their ability to develop relationships over time with a patient as an attractive aspect of general practice, current workload levels and workforce shortages impede their ability to realise this. In the UK 93% of GPs have stated that heavy workloads negatively affect the patient care they provide.[7].” - Jeffers & Baker, 2016

“Changing patterns of provision are also impacting on the ability of general practice to provide continuity of care. With more GPs choosing to work as locums, salaried GPs, or part-time, they have less opportunity to provide continuity. Similarly, the transition to new models of care and new working structures are posing new challenges for how continuity is delivered.” - Jeffers & Baker, 2016

“With limitations on the ability to provide traditional, relational continuity, it looks at how greater emphasis can be placed on managerial and informational continuity.” - Jeffers & Baker, 2016

“Also, although traditional relational continuity ideally would be realised for every patient, this is impossible in the current climate, so greater emphasis should be placed on managerial and informational continuity. It explores this at GP level and practice level, and at regional and national level.” - Jeffers & Baker, 2016

“According to participants, continual organisational changes fundamentally altered their professional role to a ‘government clerk’ or a ‘data clerk for public health and for management,’” - Rimmer, 2016

“Of the GPs that responded to the survey, 50% said that rising non-clinical workload was a factor in their decision to leave general practice early, while 84% said that pressure of work contributed to their decision. “The higher administrative workload reduced the time available to spend with their patients, leading to a fundamental change in the doctor-patient relationship,””- Rimmer, 2016

“With more and more of our working hours being taken up with form filling, ticking boxes, and preparing for CQC [Care Quality Commission] practice inspections, we are drowning in red tape, and this only serves to keep us away from delivering frontline patient care, which is why we become doctors in the first place.” - Rimmer, 2016

“GPs talked of the consultation becoming less rich, more transactional, and more awkward in nature, characterised by less active listening and less attention to the emotional dynamics of the interaction (risk 2B) as the following comment illustrates: '… And I think it’s all a wee bit false and artificial … on the video … probably a wee bit more conscious of myself and my body language and all that sort of stuff. And that there’s less … I think there’s less informal chats.' (HFVC06 GP)” - Rosen et al., 2023

“'[On the text or email] maybe a vague symptom that doesn’t really allow you to respond … and it sort of forces you to phone up and say “well, actually sorry I didn’t quite follow what you’re saying about that tummy ache”, and then you realise that actually there was a bit more going on there and you didn’t respond to that with that text about the eczema because that’s not really what they wanted to talk about, it’s that hidden agenda thing. That is no small part of what we do as GPs and it’s all a bit unspoken and difficult to quantify.' (RBD GP CK)”- Rosen et al., 2023

“Low levels of personally reported physician individual accomplishment have correlated with emotional exhaustion and depersonalisation.” - Spiers et al., 2017

“There are increasing numbers of salaried and locum GPs (sessional GPs) who account for approximately 40% of the workforce, and have been described as demotivated and disenfranchised, principally due to isolation.” - Newman et al., 2014

“As referral systems became more complex and hospitals more specialised, interviewees experienced a more fragmented and depersonalised healthcare system that was increasingly challenging for them to navigate: ‘One of the problems with hospital medicine is it’s very fragmented , so if you sent somebody in with one thing, they have that sorted, but they don’t look at the bigger picture, so they’d come back out and there’d be another thing that was developing so you’d have to refer them to somewhere else, so the fragmented nature of hospital medicine makes general practice quite difficult.’” - Doran et al., 2016

“The higher administrative workload reduced the time available to spend with their patients, leading to a fundamental change in the doctor-patient relationship: ‘You see it does change the doctor-patient relationship because it changes how you react to people and how you interact with people. I mean it’s obvious stuff, but when you’re really stressed and you’ve still got 15 people to see, you don’t have the time for people, you don’t have the interest.’” - Doran et al., 2016

“Participants described a series of conditions which they felt contributed to an increasingly pressurised working environment. These included organisational changes resulting in a clash of values and diminishing professional autonomy as health care became more centralised, standardised, and depersonalised; an unprecedented increase in administrative workload; and a lack of support not only from government, but also across services and the wider community due to an ongoing negative media campaign.” - Doran et al., 2016

“Lack of time with patients meant the ability to practise patient-centred continuity of care was perceived to be compromised and, with it, the GPs’ professional autonomy and values, resulting in diminished job satisfaction. Once their job satisfaction had become negatively impacted, the combined pressures of increased patient demand and the negative media portrayal left many feeling unsupported and vulnerable to burnout and ill health, and, ultimately, to the decision to leave general practice.” - Doran et al., 2016

“Vautrey notes that ‘doctors didn’t train to work in call centres. They want to have that regular face-to-face interaction with their patients, which is one of the joys of general practice.’” - Oxtoby, 2021

“Because when you're an [GP]ST, you're studying in a way to pass exams. Your learning is more focused on that. Whereas now, it's actually on what you need to do day-to-day. and what's most useful to you and your patients.” - Cunningham et al., 2021

“At a time when recruitment to medicine is problematic, and burnout and disenchantment worryingly common, the benefits to medical staff of a patient centred, rather than a purely transactional, approach to care should be of particular interest to policymakers and planners.” - Jones, 2021

“Participants considered that their consultations were more doctor-centred, reflecting busy consultations, time constraints and their patients’ reluctance to be involved in shared decision-making.” - Cunningham & Yeoman, 2019

“Contained work within small practices was perceived to allow a focus on care-related tasks. The new GPs believed that older GPs were familiar with all of their patients and had autonomy and freedom from many of the bureaucratic restraints they now faced. Participants expressed frustration that having to multi-task administrative work, complex referrals, follow-ups and additional clinical tasks from hospitals (Charles-Jones et al., 2003) took them away from patient-facing work and left them feeling stretched and unable to work in the way they want: I just want to feel like when people come to see you, you think if it was you or your relative or anyone else, you want to look after them the best you can and I hate feeling, like, I can’t do that. (Daria (new GP), interview).” - Laverty et al., 2023

“From these early-career GPs’ perspectives, the previous generations had it good. Their perception of past work was that the environment was controllable and manageable and the mutual understanding of roles with the public was reciprocal.” - Laverty et al., 2023

“In choosing general practice as a specialism, then, this is what the participants signed up for and expected in their future work. It was their imagined future. Instead, what they are observing is that components of general practice that they imagined would lead to meaningful work, such as autonomy and shared values, are evaporating - the promise cannot be met. Participants were reconciled that these changes would be permanent; for example, a trainee in one of the focus groups recalled a conversation with an older GP: I met one of the GPs who I was working with, he was saying he doesn’t think the sort of general practice that he loved doing, and the sort of general practice that I saw and really liked, is all that common anymore . . . they knew the regulars very, very well, and they knew all the families, locally, as well. And I really loved that, and I think that is just disappearing, and I think that will continue to disappear. (Focus Group 2, trainee GPs).” - Laverty et al., 2023

"Present future solutions (e.g. reducing working days) then, make more sense to individuals than investing in future present solutions (such as taking up partnerships) given the uncertainty. These solutions reflect what Chaudhry et al. (2021) have referred to as depersonalisation and detachment strategies in medical work during crises. It is, currently, unclear whether these individual strategies can support meaningful work and further research exploring these temporal clashes could prove fruitful." - Laverty et al., 2023

"There needs to be system-based responsibility and obligation for the next generations to help reshape and reimagine what the future can and should be. To take a caring approach to futures. For general practice, a new social contract will likely need to be defined to manage the expectations of all parties. This applies to other professions and industries, such as academia (Gill, 2010), experiencing widespread uncertainty and insecurity that result in short-term present future approaches." - Laverty et al., 2023

"Generalists of all types are less engaged in hospital care and, as a result, have much less personal contact with the consultants they use. The times spent with colleagues in labour and delivery, in the ED, or morning rounds are being replaced by e-consults and wordy electronic progress notes more filled with facts than information. Primary care clinicians don't get personal notes or phone calls back—they are told that notes in the EHR (electronic health record) should suffice.[...] Access metrics—getting the patient to a consultant quickly—seem more important to large organizations than relationships." - Frey, 2018

"Beyond a certain number of people in a clinic, familiarity disappears. "Practice teams" are often attempts to break large practices into meaningful working relationships but even team meetings become less spontaneous, are scheduled with agendas, and rarely include "how are we all doing?" We barely know who each other are. Some of the interest in direct primary care practice may be driven by a desire for a practice environment where doctors know something about everyone in the office." - Frey, 2018

"Struggling alone is a reality at times for everyone. However, expecting a colleague to send an email saying "help" assumes they know when and how to ask. More often than not, the "help" is in a colleague's demeanour, voice, eyes, giving us a chance to put a hand on a shoulder and ask, "Are you all right?" Having solitary lunches or eating a yogurt at one's desk doesn't really improve our social intelligence." - Frey, 2018

"The findings show that the traditional public image of the family doctor as a 'virtuous practitioner' (upstanding, honourable, courageous, and selfless) who was personally devoted to their patients and retired only after providing decades of uninterrupted service to a particular community has been replaced in the media by images depicting GPs' moral failings (lazy, greedy, uncommitted, selfish, sexually disinhibited, and even violent). This 'moral decline' of the GP was not, apparently, cited in surveys as a reason for students' and trainees' reluctance to enter general practice (though closeditem questionnaires may not have asked the relevant question), but it may have important subliminal effects on both the public and young clinicians." - Barry et al., 2019

"Wordsworth et al suggested that enhancing the patient care aspects of a GP's work is more likely to act as a key for retention, and that lack of consultation on changes can lead to dissatisfaction." - Marchand & Peckham, 2017

"A number of new services, such as paramedic home visits or 111 triage, are positioned instead or ahead of a GP consultation, alongside 'care navigator' programmes, to support access and navigation of care pathways. These services therefore shift the nature of the GP-patient consultation from expert generalist, to engagement with a more selective patient group with a potentially higher incidence of pathology or complexity. General practice therefore moves from being a 'primary care' to 'secondary care' service, with a range of other healthcare professionals providing the initial patient interface and possibly serving as the 'new' gatekeepers to NHS services." - Park et al., 2019

"GPs now are most unlikely to know or even see their patients — their stories, their homes, their families, their hinterland. They are probably hot-desking, screen-gazing, part-time, on short contracts in large practices where their contact with other professional and support staff is largely business-like, remote, and perfunctory — there is little time for more. The previous hostess function of receptionists (yes, they were mostly women) has been almost entirely automated-out; they, too, are mostly now screen-bound and cyber-bubbled with digital duties." - Zigmond, 2023

*Relationships with allied HCPs*

**11. Paradox of delegation**

**When tasks usually undertaken by the GP are delegated to other healthcare professionals (C), this can allow GPs to focus on more complex patients (O) but can increase 'indirect' care (e.g. supervising or holding responsibility for another's work and management) (M).**

**When patients with less complex problems are seen by practice members other than the GP (C), this can produce high levels of uninterrupted clinical complexity and challenge (M), leading to relentless emotional and cognitive load (O).**

"You rarely see anybody who's just got one problem. You don't see really any just basic chest infections, exacerbation of asthma, sore throats. The nurses see all those things." - Baird et al., 2016

"There was unanimous agreement that primary care would change, with a greater emphasis on the development of multi-professional teams which might include GPs with urgent and emergency care skills, practice nurses/nurse practitioners, physicians' associates, social workers, mental health practitioners, practice/business managers, patient educators and care navigators. There was a consensus that the introduction of new roles would reduce continuity of care, a component of practice highly valued by the interviewees." - Humphrey & Cleaver, 2018

"The financial cost of employing new role professionals against evidence of their value and contribution in general practice needs to be considered, since the very initiatives aimed at alleviating pressure may paradoxically place increased strain on staff, at least initially, and mean that GPs incur extra workload in supervision or mentoring." - Nelson et al., 2019

"While the skill-mix changes introduced locally aimed broadly to provide quality care, increased choice, improved access, and better outcomes for patients, operationalising the measurement of such outcomes was challenging. In particular, as GP workload data were not collected at scale, stakeholders were not able to demonstrate concrete evidence of the roles' effect on releasing GP time." - Nelson et al., 2019

"Although some doctors view additional roles as welcome opportunities to enhance their effectiveness, some have claimed that these changes disrupt the 'proper' work of GPs by reducing time available for patient care.14" - Spooner, 2016

"We came across some great examples of practices and PCNs using additional roles to improve patient care, but we know there is variation across the country, something highlighted in the recent King's Fund report. Some local systems have not yet been able to make best use of the scheme due to a lack of local capacity for clinical and managerial supervision, inadequate space in practices, confusion around the purpose of some roles, administrative complexities, and lack of expertise on organisational development and role redesign to embed new roles." - Fuller, 2022

"Instead, we heard that patients can become confused over who they are signposted to and why, leaving GPs dealing with multiple complex cases one after another and as a result, contributing to clinician burnout and concerns by the clinicians they might make mistakes or not be able to practise safely. This combination of intensely complex cases, done at speed, with fear over reprisals on the individual clinician is driving a systemically toxic environment in primary care." - Health and Social Care Committee, House of Commons, 2022

"We was a bit, at first, unsure. We didn't want to make a mistake… because we was worried that we was booking them in with the physio when it could be something else… So, in the beginning it was a bit hit and miss… and we weren't booking in as many as we should' (Admin 1)" - Moffat, 2018

"There are increasing reports of a lack of capacity in many practices to support and mentor new team members, who as a consequence are feeling isolated in an unfamiliar working environment. In the absence of adequate investment in the development of this new workforce their contribution to the crisis may be marginal, at least in the short term." - Marshall et al., 2022

"Emotional labour and stress is endemic within general practice (Lown et al., 2015) and is not restricted to clinical staff (Ward and McMurray, 2011; Murray, 2014). Our results indicate that the administration team should be acknowledged as integral to managing the process/structure and delivery of the PFPCS service. The provision of formal training for these staff members should be considered in order that they can filter patients appropriately and confidently." - Moffat, 2018

"Data suggests that one in six UK patients has more than one of the conditions listed within the Quality and Outcomes Framework, and that this cohort of complex patients accounts for around one-third of GP consultations (Salisbury et al., 2011). These findings are in keeping with the views expressed by the GP participants in this evaluation, that is that patients commonly present with MSK problems, but as comorbidities to another condition. As such, the GPs perceived that their workload had not necessarily decreased, or at least not to the extent that had potentially been anticipated." - Moffat, 2018

"During the early implementation of these NMP (non-medical practitioner) roles, concerns were raised. Kingsnorth (2005) demonstrated limitations of the surgical care practitioner role, reporting they were more expensive owing to increased operating times, an extended training period and requirement for supervision." - Koworera et al., 2022

"Substantial general practitioner workforce growth is unlikely to be achievable in the near future, making consideration of alternative models of primary care essential. Pharmacists integrated into general practices can be seen to offer a practical solution, with the potential to reduce general practitioner workload and improve access, while at the same time improving quality." - Komwong et al., 2018

"For example, there is some evidence that adding nursing staff does not always release doctor time and instead can create a duplication of appointments or workload." - Gibson et al., 2023
"Very little evidence of direct interactions between the effects of different types of workers on outcomes at the organisation level was found. This may indicate that different staff groups are not coordinating their work and are working quite independently. As such, there may be gains to be made from encouraging more effective team working." - Gibson et al., 2022

"Registered nurses work in a variety of roles in primary care practices. Within the UK career framework for practice nurses there are no formal academic training requirements above those required for registration as a nurse and while many work at advanced levels there is no clear data, as job titles are not applied consistently and there are concerns that training for advanced roles is not always adequate or properly supported by employing practices (Rashid, 2010)." - Freund et al., 2015

"Expanding the MDT workforce sounded great. Then you discovered they'll be allocated according to practice population size. Meaning you'll be eligible for two hours of this (MDT staff) a week. That won't help at all. The reforms and models just don't fit rural areas… what we really need is more doctors. If you can't have MDT staff every day in the practice, the only person that could fulfil all those roles is the GP." - Donaghy et al., 2022

"Recent research in England has identified the complexity of matching patients' needs with practitioners' capabilities when trying to reduce GP workload by redistributing workload to MDTs. A recent King's Fund report on the expansion of MDT staff in primary care networks (PCNs) in England also concluded that while PCNs have swiftly recruited to these roles, they are not being implemented and integrated into primary care teams effectively." - Donaghy et al., 2022

"The lack of a clear, realistic description of the pri- mary care roles that PAs were equipped to undertake caused uncertainty and confusion amongst some prac- tices and patients. Practices often relied on interns to outline their own professional boundaries. Confusion over PAs being independent or dependent practition- ers emerged. Practices often recognised that PAs were dependent on GPs for supervision, but lacked clarity about the wider implications of this (e.g. the process for administration of influenza vaccinations). Some practices described uncertainty among existing staff about the role(s) a PA would occupy." - Cottrell et al., 2021

"Perceptions of acceptable supervision levels appeared to be individual GP and/or practice-specific. A single salaried GP in a host-practice refused to provide clinical supervision for the intern due to perceived risks. Some GPs reviewed all intern patients for an extended period but felt over- burdened by this. Conversely, after a short induction period (a few weeks), other GPs allowed supervision to be determined by the intern and case complexity. Whilst training practices were more likely to find the supervision demands acceptable, they often had multiple dependent practitioners/trainees concurrently requiring GP support." - Cottrell et al., 2021

"The PA intern was the tipping point in some practices, who adapted their working day (e.g. reducing booked appointments) to accommodate this. Notably, multiple people requiring support from one GP created delays for some interns. Smaller practices struggled most to provide 'blocked-out' appointments to accommodate 'just-in-time' supervision, due to the relative impact on appointment capacity. The consequences of this were that PAs sometimes rescheduled patients with another clinician to complete the management plan and/or felt less supported. Practices that had invested a lot of high- level supervision early on appeared more satisfied at internship end." - Cottrell et al., 2021

"If all the clinically straightforward consultations are done by others, GPs are left with wall-to-wall, dawn-to-dusk complex patients, which is a sure recipe for burnout. Sometimes we long for a simple sore throat or infected ingrown toenail to interrupt the flow of patients with multiple symptoms and too many medications, severe depression, or cancer. Diverting simpler cases to other staff also disrupts continuity and our attempts to establish relationships. If I don't see the teenagers when they come about their acne, will they later trust me with their eating disorders, unplanned pregnancies, or substance misuse?" - Salisbury, 2023

"In recent years, several professional groups have taken on new roles in UK primary care. The integration of nurse practitioners into primary care teams has been described as a 'dynamic, complex and messy' process, necessitating careful planning, collaboration and redefinition of professional boundaries." - Barker et al., 2021

“Some GPs described the possibility that the thorough methodical approach of some pharmacists could tip the balance such that they were unable to complete the work at an appropriate pace to save GPs time or that they some- times created additional work for the GPs which they viewed as unnecessary.” - Hampson & Ruane, 2019

“Bradley et al. also noted that pharmacists new to the role require clinical support from GPs and warned against practices expecting immediate gains in terms of a reduction in GP workload. It may be that a lack of experience or an inability to cope with clinical uncertainty con- tributes to some pharmacists increasing workload by seeking reassurance or sticking rigidly to guidance.” - Hampson & Ruane, 2019

“Nuanced histories and examination cannot be reliably achieved through triage, with the result that pre-consultation segregation by presenting problem may put comprehensive and skilled assessment at risk, as well as limiting patients’ choice of available healthcare provider and continuity of care.11,12 This model leads to further questions about responsibility; the report is silent on the question of who bears ultimate responsibility for independent work delivered by the extended primary care team.” - Checkland and Spooner, 2015

“When I look at the home visits … we send the paramedic and then we send the paramedic the second time and then we send the GP the third time. So have we done more than we would've done normally and … I'm not convinced that we've actually saved anything.” - Schofield et al., 2020

“The tendency for paramedics to see low-complexity cases can result in GPs primarily seeing patients with multimorbidity, chronic illness, and, often, social complexities. This has the potential to impact on the job satisfaction and mental wellbeing of GPs: 'But if the paramedics see all of these “easy stuff” … you're left with only the really complicated stuff … I found that emotionally very hard work and draining.'” - Schofield et al., 2020

Effective collaboration in multidisciplinary teams

“All participants described the benefits of the PFPCS in terms of a strategy to help to manage the unprecedented demand faced by primary care. The justification for this was twofold: redistributing work and ‘unburdening’ the GP. The administration staff and nurses highly valued the ability to redistribute work and have the ability to offer alternative appointments: ‘We always need appointments… It was brilliant when you were doing telephone triage because you’d think, “Cor, I’ve got another appointment I can put them in”’ (Nurse 1)” - Moffat, 2018

“On average the additional 3 h of pharmacist time addressing key prescribing activities freed 1 h of GP capacity. This achieved a sustainable 47% (73 h per week, $F(4,56) = 16.05$, $P < 0.001$) increase in freed GP capacity; reducing 155-82 h per week of GP time (Fig. 1). This equated to 4.9 h (95% confidence interval [CI] 3.1-6.7) per practice per week, for the 15 practices. There was a strong correlation between practice size and practice-level GP time released (Pearson's r = 0.622, 95% CI = 0.130-0.854, P = 0.013)." - Johnson et al., 2022

"Widening out the MDT, freeing up more time for GPs to spend with complex multi-morbidity patients, delegating other tasks to MDT staff is a good model. Replacing QOF with GP clusters, driving quality according to the priorities of their local populations, flexibility of practices being able to develop their MDTs according to the needs of their local population - all a really good idea." - Donaghy et al., 2022

"We've established a very good APP [advanced physiotherapy practitioner] team across all the practices in our cluster. So, reduction in onward referral and in analgesics, with high patient satisfaction, high doctor satisfaction. A generally wonderful service. Similarly, primary care mental health nurses have also been wonderful, right person, right place, and right time. It's all there." - Donaghy et al., 2022

"In Brighton and Hove some practices have developed a robust protocol to allow clerical staff to read, code and where appropriate take action on incoming clinical correspondence, rather than the GP having to deal with every letter. Forty-eight practices have now been trained and implemented workflow redirection with substantial changes demonstrated. On average, only 20 percent of letters previously directed to a GP required their direct input. This is saving an average of 40 minutes of each GP's time per day, with no significant events in the first 15,000 letters to be processed. Feedback clearly demonstrates reduced workload pressures and with the time savings generated, increased opportunity for activities related to direct patient care." - GPFV, 2016

"By reviewing roles, delegating work, using clerical staff effectively, using integrated e-records, and devolving work to the healthcare team allows more time for better patient care. The GP practices that have successfully implemented changes are generally stable, well-staffed, and with a good team ethos." - Ferguson et al., 2020

"PAs considered themselves most similar to GPs, although recognised they were not GP replacements. GPs agreed that PAs adopted a similar approach to patient assessment and presentation, but thought PAs had less depth of understanding and reduced ability to diagnose and consistently manage complex conditions. Whilst PAs had some gaps in their capabilities, compared with trained GPs (the lack of prescribing capability was mentioned repeatedly), their flexibility to perform elements of different healthcare professionals' roles was valued by GPs and practice managers. Overall, interns were perceived as a hybrid of multiple primary care roles." - Cottrell et al., 2021

"Over the year, GP face-to-face reviews reduced, and practices and interns developed efficient and sustainable supervision and support methods which, in turn, increased acceptability of supervision. For example, protocolising care for common conditions; utilising electronic prescribing systems (sending electronic pre- scription requests) if no advice was needed; use of medical record screen messaging systems for brief advice; joint debrief sessions with other dependent professionals/

trainees; and catch-up meetings. A mismatch in perceptions of appropriate supervision emerged: some interns felt over-observed and others under-supported." - Cottrell et al., 2021

"The new roles (ARRs) coming into PCN settings provide an opportunity to help existing clinicians meet the demand for mental health care. [...] This important and novel service evaluation explored the efficacy of brief psychological interventions, delivered within General Practice settings, and found both clinically and statistically significant reductions on measures of anxiety and depression and clinical and statistically significant improvements on measures of resilience and wellbeing. Multi-source feedback from individuals who accessed the service, the workforce and General Practice staff was very positive, with benefits for patient care, a positive impact for other practice staff and suitability of service location all cited as benefits." - Budd et al., 2022

" …I suppose we did have a degree of apprehension that they might be a little bit risk averse when dealing with patients because as you say, they're not really used to making that many management decisions - just more stay at home or sending to hospital. But we certainly didn't find that [when the pilot started]. For the most part they were keeping people at home appropriately with appropriate safety net advice… I think that the care they provided was excellent." [Paramedic Practitioners] - Barker et al., 2016

**12. Supporting cross-disciplinary learning**

**When there is a mismatch between service-learning expectations and needs for HCP roles in general practice (C), successful integration can be challenging to establish (O) unless adequate time and resource is allocated for GPs to provide generalist training to colleagues (M).**

**When other members of the clinical team need supervision, 'second opinions', or prescribing by GPs (C) this can increase GP workload (O) because they have responsibility to manage risks and related additional tasks to do (M).**

"There is wide literature on managing uncertainty in healthcare contexts involving different conceptual models, however it is recognised that general practice is characterised by the presentation of undifferentiated and wide-ranging problems, meaning trainee and qualified GPs need to develop strong skills in dealing with uncertainty and risk. The issues raised by stakeholders in this study about adequately preparing new roles professionals to manage risk in general practice speak to the tension between how professionals' previous experience and potentially more protocol-driven training maps on to the often 'unpredictable' setting of general practice. If unaddressed, this may be a significant barrier to the sustainability of new professional roles in general practice." - Nelson et al., 2019

"In particular, there is a crucial need for training leads and general practice staff (with the time and skills for adequate on-the-job supervision) to help role holders develop appropriate risk-management skills over time." - Nelson et al., 2019

“Concerns about safety and supervision were mixed. Some GPs were clearly concerned about the lack of prescribing rights, which was repeatedly mentioned, and led to concern that the level of supervision required, and additional responsibility was unattractive to them.” - Jackson et al., 2017

“If you think … like someone coming out of medical school, they’re no bloody good, are they, to anybody, they’ve got to be trained after that.’ (GP, Group 1).” - Jackson et al., 2017

“You would end up having to actually re-evaluate them in all the areas before you could set them [work].’ (GP, Group 3).” - Jackson et al., 2017

“We are also pleased the Government has committed to increasing the flexibility of staff recruited under the Additional Roles Reimbursement Scheme. However, we are concerned about some gaps within the scheme, particularly the lack of funding for supervision and the absence of any uplift for areas of high need. At present it appears that some staff are not regularly being effectively integrated into general practice and do not receive adequate supervision.” - Health and Social Care Committee, House of Commons, 2022

“However, one GP highlighted an increase in workload when their pharmacist was sick or on annual leave: ‘Unintended consequences, workload increase for GP when pharmacist[s] are away, [on] annual leave/sick.’ (GP, 45)” - Maskrey et al., 2018

“[PAs] are clinically very well-trained individuals…but they can’t prescribe. So, you can’t let them do clinics independently (GP 2, 2019).” - Krachler, 2022

“If there isn’t a whole team approach, I think you can more or less forget it because no clinician will practice like this in isolation. It has to be a philosophy of the practice. It has to eat through everything for us all to continue to be brought into it. It becomes part of the way that the whole practice philosophy is the way we deliver services.” - Robertson et al., 2013

“I guess that there’s still this culture that patients perceive the GP as being at the top of the hierarchy in terms of clinical experience… hopefully with the pilot that we’ve done here… they may start to recognise that, in actual fact, ‘if I’ve hurt my back or hurt my knee, I’d be much better going to see the physio…’” - Moffat, 2018

“Both GPs and physiotherapists believed that whilst the nature of the service ‘expos[ed] physios to a certain degree of clinical risk’ (PT1), its success was dependent upon having experienced MSK physiotherapists working in an extended scope role. Consequently they expressed concerns for the future of the service if this level of seniority was reduced: ‘I worry a little bit if you were putting in more junior, maybe cheaper, physiotherapists in, that they might not be as effective… [the physio] has taken the patient and taken ownership of them… and if it was someone less experienced we might find that we were getting more patients bounced back to us’” - Moffat, 2018

“However, the present analysis indicates that deployment of newer types of practitioners cannot be assumed to provide a straightforward, complete, or cost-neutral solution to the GP workforce and workload crisis. These findings highlight the importance of professional workers in the production of health care. They demonstrate that different types of health professionals are not simple substitutes for one another, and that the quantity and quality of services generated in primary care will depend on who is employed to work in this setting." - Gibson et al., 2022

"However, task shifting requires both willingness to give up tasks on physicians' side as well as the ability and capacity to perform these tasks on non-physician health professionals' side. The first requirement is commonly met in emerging countries (e.g., Brazil, India) where workforce innovations have more easily been implemented (Martiniano et al., 2014) partly because of a lack of established professional roles which in other circumstances may hamper task shifting by expectations and attitudes of each profession (Donelan et al., 2013; Macdonnel and Darzi, 2013)." - Freund et al., 2015

"A commitment to a national scheme was requested, to provide standardised education, practical and financial support to make the integration of PAs into primary care a success. This was felt to be necessary for at least a few years, until a critical mass of experienced primary care PAs is realised. Host-practices wanted a collaborative network to develop a shared understanding and standardised approaches to supporting their interns." - Cottrell et al., 2021

"The GPs valued listening to short talks from colleagues with other roles and, following an afternoon workshop, felt more equipped to work in wide multidisciplinary teams. GPs were enthusiastic about diversification of the workforce, but they cannot successfully lead teams they do not understand. Patient care and satisfaction will remain overly dependent on GPs until leaders are appropriately utilising the skill mix within their teams." - Zigmond, 2022

"General Practice staff identified challenges with integrating the Trainee Associate Psychological Practitioner (T/APP) role into the primary care setting. Some T/APPs expressed feeling isolated and not fully integrated into the General Practice team, even by the end of their training year. Furthermore, both T/ APPs and PCNs acknowledged difficulties with General Practice staff understanding the function and remit of a T/APP and highlighted practical issues such as finding room space for the T/APP. Both T/APPs and General Practice staff suggested increased communication between the T/APP, the PCN and the university may be helpful to resolve these issues." - Budd et al., 2022

"Some respondents reported a lack of engagement by GPs in relation to integrating the CN (care navigator) role into their practice, suggesting that greater and more prolonged engagement work over time was needed to highlight the perceived value of signposting and to support implementation of the role." - Brunton et al., 2022

"Certainly, the biggest frustration…was the lack of GP support, and them not valuing it…if you haven't got the people at the top of the chain engaged, it's not really going to work. So yeah, we still need to work on that, and we keep chiselling away…but it's like an oil tanker, you wouldn't believe that this was actually part of the 10 Point Plan to help you." - Brunton et al., 2022

“Other authors have also suggested that even if patients initially comply with attempts to divert them away from GPs, if they are dissatisfied, this may lead to further efforts to see a GP and a ‘revolving door’ of consultation requests, potentially increasing rather than reducing staff workload” - Brunton et al., 2022

“Previous literature showed that most factors affecting the quality and safety of locum practice related to the organisational context and ways in which organisations deploy and support locums, highlighting that locums ‘can only work as well as their working environment’” - Borek et al., 2022

**Learning and development**

*Enabling cultures and organisations*

**13. Learning systems**

**Where regular connection with colleagues and exchange of knowledge is established (C), GPs feel connected and able to adapt and cope better (M), which can inform clinical expertise and patient care (O).**

“It is therefore important to view ongoing professional development in the light of one’s practice characteristics (ie geography, social environment and patient profile), which, in turn, determines one’s required knowledge, skills and capabilities.[59] The most effective and efficient way for GPs to achieve mastery is a local community-based, peer-supported network (aka community of practice) that provides a safe peer learning environment in which to explore the interdependencies between patients’ health, illness and disease care needs within an inevitably constrained health system.[60] - Sturmberg et al., 2023

“’Working within these primary care networks, you are able to look at things more at scale. Like homelessness, or childhood obesity, or substance abuse. I think the future has to be much larger than your practice.’ A salaried GP working in London” - Thomas, 2020

“Able to provide continuity of care with the learning opportunities and satisfaction that comes with following patients up. Being part of a team. Being part of the team of doctors that have provided fantastic long-term peer support and allowing me to provide continuity of care to my patients/case load with the flexibility of altering hours to manage my young children.” - Lockyer et al., 2014

“GPs need to work with colleagues. This topic was found in seven articles. They emphasised the importance of relationships with other specialities and hospitals [19, 22].” - Le Floch et al., 2016

“General practice staff used Primary Care Network care coordinator and Enhanced Access funding to make service-improvements (outlined in Table 2), which have continued beyond this project. This included two practices recruiting a care coordinator to support marginalised patients who struggle to access mainstream general practice, and one practice co-designing an inclusion health clinic. This clinic included support from the care coordinator with appointment booking, reminders and non-clinical support, and longer appointments with one of two GPs experienced in inclusion health. This additional capacity enabled motivated GPs and care coordinators to collaborate on providing better care for marginalised patients." - Potter et al., 2023

"Professional socialisation of attending courses was viewed positively by R & R participants, with the exchange of ideas valued by those who were relatively isolated. You get the option to meet people outside who you normally work with. You can actually find out what other practices are doing, or what the rest of the world is doing, because it can get quite small otherwise, if you're just in your wee town on the **** coast miles away from everywhere else." - Cunningham et al., 2021

"The nature of the supervised placement in the I&R scheme, which incorporates an induction period and then a gradual increase in patient contact over several weeks, is congruent with the notion of the learner moving from a peripheral to a more established position within the practice over time, in a supported way." - Shah et al., 2021

"Several participants recognised the benefit of building on existing trusted relationships to encourage engagement with other professionals, or help with the practicalities of getting to appointments. "so we actually get that person to feel that it's okay, if we're all singing from the same hymn-sheet and saying, 'You need to go there. Just ask for this person. You'll be absolutely fine'"- CC1 - Potter et al., 2023

"Previous work by the RCP [5] has shown that the breakdown of the medical team was a central factor contributing to low morale and disengagement. It is therefore conceivable that developing a sense of collegiality and togetherness across a transgenerational grouping of GPs may boost morale and engagement, leading to greater resilience within the GP workforce." - Imtiaz-Umer, 2021

**14. Informal interaction and peer support (professional and personal)**

**When there are opportunities for informal interaction and peer support (e.g. group practice meetings, quality circles, mentoring systems, coffee breaks, and dinners) (C), GPs feel part of a community (O), which enables them to learn and flourish (grow, develop, and thrive) (M).**

"Having social support within the practice, peer-to-peer and from both medics and non-medics outside of their practice was found to be useful for preventing burnout. To improve support at the practice level, buddying, and mentoring systems were suggested, along with regular meetings to 'check in' with how team members are doing." - Hall et al., 2018

"F1: But I think also, looking after each other…. I think we're quite good at looking over our shoulder at the other person (…) if you see somebody's got a really full load, getting them a cup of tea, or going and seeing one of their extras, (…) is quite a positive thing about our team that we tend to do. [FG1].” - Hall et al., 2018

“GPs with good team support and particular consultation styles also felt less negatively.” - Croxson et al., 2017

“For people to be motivated to do their job, they must feel part of a community that supports and enables them to flourish both as an individual and collectively. Collective working enables individual strengths and values to be developed and utilised while contributing to broader goals.” - Andah et al., 2021

“The theme of personal and professional support was seen to map to Kanter's third category of membership, which is the importance of building communities of practice, which support and enable individuals to thrive in their role and provides the motivation to stay.” - Andah et al., 2021

“Stress in the profession, I think, is pretty significant I would say. You know, you do hear regularly of sort of GPs, sort of, you know, committing suicide and stuff like that. We need to make sure that we sustain our health and our energy. Surely, we should have probably half an hour downtime to have some food where nobody disturbs us, and we can, and you know, we need to have time when we get home that we don’t feel completely shattered and too tired to go and do some exercise, and stuff like that, and keep ourselves fit.’ (GP13, male partner, nine sessions per week, 16-20 years’ experience, medium-sized suburban practice).” - Croxson et al., 2017

“I felt that this family doctor surgery where I now worked provided me with a secondary family, a home from home.” - Zigmond, 2022

“Having a supportive team was seen as a buffer and support for practitioners, who otherwise might not cope on their own.” - Eley et al., 2018

“If everyone had been just in just their own little worlds and stayed in their rooms, and not wanted to chat about things, or happy to listen to me when I wanted some advice I definitely wouldn’t have stuck around.’ (F, ≥5 years).” - Eley et al., 2018

Routines that brought GPs together were seen as beneficial to resilience because they allowed practitioners to feel like part of a supportive team:

“[We] have lunch together … I think that’s probably a massive, massive contribution to resilience because it’s just a chance to have a chat, have a moan.’ (male GP [M], ≥5 years).” - Eley et al., 2018

“Interviewees sought informal support through family and friends, colleagues and peers. They described the benefits of talking to other medics who could relate to their experiences; this was particularly important to trainees, some of whom were isolated from family and other networks." - Jefferson et al., 2023

"There appeared to be good awareness of the different formal support structures available; ranging from coaching and mentoring support (which several participants had used) to more formal mental health support. Only two male participants discussed using these support services, and, similarly, gender differences were apparent in discussion of approaches to 'self-care'; with comments predominately made by women." - Jefferson et al., 2023

"GPs with high job satisfaction described feeling supported by good practice relationships" - Long et al., 2020

"Those who had experience of a few different teams were particularly vocal on topics related to leadership and team cohesion." - Spooner, 2016

"Team building through finding time for interpersonal communication was highly valued:
'We always stop for coffee, which is one of those things that actually I haven't compromised on.' [Mary]." - Spooner, 2016

"Making sure that all GPs shared a coffee break provided 'very supportive' opportunities to allocate tasks, discuss any problems, and efficiently decide on any necessary action. They could also pass on information, talk through complex cases, or simply consolidate a sense of working together." - Spooner, 2016

"Purposeful teamwork did not happen by accident: maintaining a 'good secretarial team', 'a very strong nursing team ... three nursing prescribers and one nurse who does minor surgery' was built on more than regular coffee meetings: 'Our practice is quite well known locally for … lots of team-building things with the staff.' [Stewart]." - Spooner, 2016

"Resilience was enacted through teams rather than through individual strength. Third, resilience required the integration of personal and professional values rather than keeping the two separate. This dynamic adaptive view, with an emphasis on the importance of individuals within teams rather than in isolation, contrasts with the discourse of resilience as a personal characteristic, which should be strengthened at the individual level." - Eley et al., 2018

"The article proposes that long-term engagement of practitioners may be achieved through provision of adequate supportive resources to allow them to enact a sense of medical identity that matches with their acquired expectations of their role in the NHS." - Spooner, 2016

"From my perspective, the strengths are that I am in a very supportive environment. There has been career progression. I have learned from colleagues. I have been able to commit to a registered list of patients. We have been able to innovate.” - Health and Social Care Committee, House of Commons, 2022

“General practitioners tend to work together most successfully when they have some agency over deciding with whom they collaborate and how they do so. Successful collaborations tend to be in response to or focused on issues of particular salience to GPs and their everyday work. Sufficient trust between parties is of crucial importance to successful collaborations - this can take time and effort to establish or build.” - McDonald et al., 2020

“This means moving away from the current vogue of encouraging GPs to become ‘Leader-Managers’ and aiming to refocus on establishing a robust local primary care team. By this I mean the ‘organic’ team developing together and trusting each other on a personal level, which results in an efficient, self-sustaining, and protective way of working. Nothing should be allowed to disrupt this team as has increasingly happened with impositions by political and management ‘initiatives’ that do not bear any relation to the real work of general practice.” - Vorster, 2017

“Even in those who did not report burnout, clinicians faced reduced opportunities for meeting colleagues face to face and seeking their advice about difficult consultations, limiting opportunities for learning from each other (risk 6B): '… the partners were saying, you know, “try to stay in your room at work as much as possible” … So it was, you know, I don’t want to spread anything, I don’t want to get anything. So I’d come into … and I just stay in my room … And then you feel more isolated because you’re not getting that general little chat about a case or … about how things are.' (RBD practice nurse IM)”. - Rosen et al., 2021

“GPs who receive more feedback from their colleagues have higher job satisfaction.” - Stobbe et al., 2021

“Federated4Health created peer group support for newly qualified GPs with a mentor available by WhatsApp, who made the doctors feel valued and enabled them to start quality improvement projects in their practices.” - Staveley, 2020

“Our in-house pharmacist learnt a lot from listening to the asthma specialist. And then used that information when she does one-to-one consultations and also in the reviews from patients who have been in the group consultations, and I think in a way we’ve all learnt together. It’s been an educational activity for the staff, the professional staff as well.” - Swaithes, 2020

“The importance of ‘good working structures and relationships’ and ‘regular meetings’ (P10) for instigating change within a practice was identified: ‘Get the decision makers involved because if they’re not involved, they will stop you, you know, passively, if not actively, because all they need to do is push it to the curb and people’s enthusiasm goes. Because particularly clinicians, you know, life’s busy and unless you’ve got some space to be able to keep pushing, keep pushing, it’s difficult to keep things going.’” - Swaithes, 2020

“Able to provide continuity of care with the learning opportunities and satisfaction that comes with following patients up. Being part of a team. Being part of the team of doctors that have provided fantastic long-term peer support and allowing me to provide continuity of care to my patients/case load with the flexibility of altering hours to manage my young children.” - Lockyer et al., 2014

“Participants recognised the positive impact networking offered by increasing their exposure to other career stages of general practice in the form of longitudinal support, with the group dynamic, covering the spectrum of primary care from medical students to retirement highly important.” - Imtiaz-Umer, 2021

“Poor team climate has been associated with 1.6 times higher likelihood of depression and 1.5 times higher likelihood of antidepressant use, compared with good team climate. Among hospital employees, poor team climate has been found to predict higher likelihood of actually quitting and among stayers reporting higher levels of intentions to leave in follow-up 2-4 years later.” - Heponiemi et al., 2024

“As mentioned, UK GPs work in smaller practices than their Finnish counterparts, thus they may have more alone working and lower levels of collegial support, which may be a source of distress. Therefore, it would be important to promote collegial support and tutoring for UK GPs.” - Heponiemi et al., 2024

“A practice that flourished during the pandemic and will likely continue to do so is the use of regular team meetings in primary care. These meetings have been essential for providing support, setting common goals, reflecting on performance, and effective team functioning. Lack of such meetings has been reported as reducing job satisfaction, increasing burnout and workload endangering patient safety.” - Gray, 2020

“Peer support is vital for dealing with the pressures, significant events, and complaints that are now commonplace for GPs. Both formal meetings and informal coffee breaks enhance team working and professional relationships.12 When GP practices are under pressure with staff shortages, these meetings are often dropped, with a knock-on decline in team working and morale.” - Ferguson et al., 2020

“Although all participants felt supported during their training and registrar years, once fully qualified they became increasingly isolated in practice: ‘I did sometimes feel quite isolated at the practice […] I think the thing that possibly my training hadn’t prepared me for was sort of feeling like a lone worker in many ways, particularly in comparison to working in a hospital where you were always part of a team.’” - Doran et al., 2016

“I’ve been a GP now for nearly eighteen years and it’s the first time I’ve really talked to other GPs, not in an educational setting or something. Clusters have been positive in that it’s brought the practices together and it’s been very good during COVID supporting each other. That was a huge advantage. Prior to clusters there was a lot of miscommunication, or no communication." - Donaghy et al., 2023

"As a qualified GP, if I'm unsure about something, I probably am more likely to phone secondary care for a discussion with a specialist rather than, as a trainee, I might have spoken to my trainer because you have those sessions. You may even be single-handed in a practice which is such a steep learning curve coming out of ST3. You might be the only person in a practice, doing home visits completely on your own and not really having anyone to talk to, it's like: 'Argh!'" - Cunningham et al., 2021

"Interactive case-based learning was attractive and helped engagement in the topic. It increased under- standing by learning how others pragmatically applied guidelines to real-life scenarios. Everyone loves listening to a story, when someone has been there, and you can identify with them and think: 'Oh! This is what they did, this is how it went wrong, and this is how they dealt with it.' You think: 'Well, I don't have to make their mistakes.'" - Conningham et al., 2021

"The most effective and efficient way for GPs to achieve mastery is a local community-based, peer-supported network (aka community of practice) that provides a safe peer learning environment in which to explore the interdependencies between patients' health, illness and disease care needs within an inevitably constrained health system." - Sturmberg et al., 2023

"We need to support our staff and provide them with an environment that helps to protect them from burnout and isolation. Part of developing resilience is having a good support network both in personal and work settings. Having an MDM approach has been shown to improve resilience in physicians and surely this is something that should be embraced in general practice." - Coventry, 2017

"Previous work by the RCP [5] has shown that the breakdown of the medical team was a central factor contributing to low morale and disengagement. It is therefore conceivable that developing a sense of collegiality and togetherness across a transgenerational grouping of GPs may boost morale and engagement, leading to greater resilience within the GP workforce." - Imtiaz-Umer, 202

"The rise of social media forums for GPs has perhaps provided a virtual replacement of the old social spaces colleagues would inhabit for support, for example, the doctors' mess. Social media aiming at supporting GPs may be counterproductive; many participants identified that social media felt difficult — even hostile — at times, and reduced their resilience at work: 'Sometimes ... it seems a bit bullish and very obstructive and if I am not in a good place and I read through some of [the Facebook group] it does make me feel very negative about the profession.'" - Martin et al., 2022

"Good working relationships with other partners in the practice were cited as helping to mitigate some of the difficulties around managing workload, change, etc. It is certainly busy, and I think the important thing is that we do have a good group here and that I work in a good partnership and a good practice. People are aware of other people's needs, and we work together as a group, and I think it is a very supportive practice… I don't think I'd still be in the NHS if I was working in another practice, I probably would have left years ago actually" - Sansom et al., 2016

**15. Coordination of care: primary, secondary, and community care**

**When exchanges between GPs and secondary/community care colleagues are personal, direct, and frequent (C), this may increase patient safety and reduce fragmentation of care (O), as there are higher levels of personal and local/community knowledge, understanding, and coordination between the two (M).**

"Many conditions are managed exclusively at the first point of contact in primary care, though some require more specialised medical attention necessitating transit across the primary-secondary care interface, usually to hospitals as inpatients or outpatients. For these patients, coordination between the different disciplines is essential for the delivery of quality care [6]. In countries with primary care services, effective communication and functional relationships across the interface are vital for both the delivery of optimal patient care, and the minimisation of risk to patient safety (e.g., in relation to significant changes in patient medications as they transition between primary and secondary care [7-9]) [10-13]." - Sampson et al., 2016

"Almost half of the interviewees felt that the pandemic offered opportunities to foster collaboration across primary care networks (PCNs), hospitals, community, and wider services. A greater sense of camaraderie and improved working across PCNs was reported, with groups of practices 'pulling together' during the vaccine rollout." - Jefferson et al., 2023

"The majority of groupings have developed new ways of working which respond to the needs of their local populations -these include the development of innovative model of shared care between acute and primary care regarding people with diabetes from south Asian communities, the introduction of new joint protocols with accident and emergency, and community link workers to connect people with complex social needs with appropriate support from statutory and voluntary sector services." - Miller, 2016

"LHCREs are set to receive up to £7.5m over two years to put in place an electronic shared local health and care record that makes the relevant information about people available to everyone involved in their care and support. This is important as research undertaken by the RCGP in 2017 revealed that one of the major challenges GPs faced at the interface between primary and secondary care was a lack of shared systems and information."- RCGP, 2018

"Some work will always be required in order to care for patients moving between primary and secondary care, but many of the GPs who gave evidence to our inquiry were clear that the workload transfer from secondary care to general practice is increasingly inappropriate, with many citing long

“GP to do lists” produced by hospital consultants, requesting that GPs conduct tasks on their behalf such as arranging blood tests.” - Health and Social Care Committee, House of Commons, 2022

“In practice we found that, while CCGs and their constituent practices are supportive of the idea of integrated care as evidenced in their strategic plans and were actively using their primary care commissioning power to incentivise and support collaborative working among practices, there was limited evidence of wider moves towards integrated care in the community such as integrated budgets between primary and community services or between health and social care, or shifting of resources away from secondary care to support services outside hospitals.” - McDermott et al., 2019

“Increased complexity in referral pathways, for example, hospitals providing increasingly specialised services (ie, shifting more care to primary care) and delays in communication, contributes to GPs experiencing a depersonalised, fragmented healthcare system.” - Long et al., 2020

“The perceived key factors under this theme were changes in referrals—both restricted opportunities to refer to secondary care, and higher numbers of (and more complex) referrals from secondary care—as well as a greater focus on targets and assessments, and fears about re- accreditation (including evidence that some GPs might retire early in order to avoid re- accreditation). Some of the organisational changes were considered to have imposed increased clinical and non- clinical responsibilities and work on GPs.” - Long et al., 2020

“Contextually, for GPs this may have been in relation to seeking specialist advice regarding a patient; for a specialist, this may have been in relation to trying to contact a GP to communicate a significant new diagnosis in a patient. This “Ping-Pong” type of communication also existed where letters transitioned the interface, as each side tried to set boundary lines of responsibility, and was more apparent where a perceived lack of respect between clinicians, and poor working relationships between individual specialists/specialities and GPs/GP practices.” - Sampson et al., 2016

“‘Some individuals will write factually a very good letter, but they don’t actually give you a bit of the flavour of the patient […] you know that the patient’s husband died last year, and she is really very anxious, speaks volume.’. In some cases, communication of “soft intelligence” had a specific impact on care provision e.g., the patient being allocated a “double” outpatient clinic appointment. Conversely, the lack of “soft intelligence,” could lead to unnecessary patient investigation, and an increase in patient anxiety.” - Sampson et al., 2016

“For GPs, good personal relationships with specific specialists led to a sense of being better supported with patient management, of facilitating direct communication with specialists, and of easing patient transitions (“smoothing the waters” as one clinician described it).” - Sampson et al., 2016

“Specialists described their trust in, and perception of a GP’s competence, as impacting upon patient care. They identified that similar referral paragraphs from different GPs could lead to different responses (and therefore outcomes for the patient) based on their prior knowledge and experience of the referring GP. A good personal relationship facilitated a sense of wanting to help one another (rather than perceiving contact as a burden) and assisted the ease with which specialist advice would be given, and be accepted by GPs. Having personally met their GP colleague was acknowledged as influential in how they may respond to a request." - Sampson et al., 2016

"Horder stipulated a set of conditions that were required to be fulfilled in order to develop such relationships, including meeting with one another; "bad relationships thrive on isolation." Specialists were keen to stress that joint educational events with GPs should reflect "shared learning" and not duplicate the traditional hierarchical process; this contrasts with previous research with specialists feeling they had little to learn from GPs. Joint learning events might also help clinicians to develop networks and share learning as a means of establishing 'Communities of Practice'". - Sampson et al., 2016

"As referral systems became more complex and hospitals more specialised, interviewees experienced a more fragmented and depersonalised healthcare system that was increasingly challenging for them to navigate: 'One of the problems with hospital medicine is it's very fragmented […], so if you sent somebody in with one thing, they have that sorted, but they don't look at the bigger picture, so they'd come back out and there'd be another thing that was developing so you'd have to refer them to somewhere else, so the fragmented nature of hospital medicine makes general practice quite difficult." - Doran et al., 2016

"Those participants in small practices, typically in a small practice-owned building, felt more vulnerable and less supported than their colleagues in larger teams accommodated in purpose-built health centres. A large primary healthcare team, consisting of several professions including attached community teams of district nurses and health visitors, was an asset for GPSTs. They also appreciated being in large health centres which included community care, mental health teams, and social work departments (social services)." - Cunningham & Yeoman, 2019

"A number considered that their learning experiences were better because they were in a large practice and part of a significant community resource with different teams working together." - Cunningham & Yeoman, 2019

"A large primary healthcare team, consisting of several professions including attached community teams of district nurses and health visitors, was an asset for GPSTs." - Cunningham & Yeoman, 2019

"They also appreciated being in large health centres which included community care, mental health teams, and social work departments (social services). We had the CPN in the building, we had substance misuse in the building. We had the social workers upstairs so it, during my first week [induction] when you sit in on all the specialities. I went upstairs and sat with, I went to kind of all parts in the building, the big health centre, and chatted to them during my induction weeks and stuff, so I knew exactly where to send folk to and direct them. (Participant 12)." - Cunningham & Yeoman, 2019

"GPs need to work with colleagues. This topic was found in seven articles. They emphasised the importance of relationships with other specialities and hospitals [19, 22]." - Le Floch et al., 2016''

"That first meeting was a whole team meeting because I do feel like access to healthcare is very much a whole team issue. It's not just about GPs."- GP1 - Potter et al., 2023

"Participants stressed the importance of a whole system, consistent approach to AMS and antibiotics within and between organizations and settings. 'I think it's really important personally to have a multi-pronged approach so that you are all doing the same thing, and you don't have a GP saying 'No, you can't have an antibiotic' in their surgery, and then they walk to the local casualty at 7 pm and come home with Augmentin... It's trying to have consistent messages from all areas of the NHS which is the hardest ask.'" - Borek et al., 2020

"Working collaboratively was viewed as instrumental to the success of the pilot. Both ECPs (Emergency Care Practitioners) and GPs reflected on the importance of positive communication, as well as trust and appreciation of each other's practices, which developed over the course of the pilot." - Barker et al., 2021

"In all three areas we studied, vertical integration [between GP practices and local acute hospitals] has been used to address the difficulty faced in maintaining the traditional model of GP partnership-based primary medical care. Vertical integration in these places has provided a more stable financial platform for primary care than the model based on individual practices run as separate businesses. At the case study sites, GPs no longer risk personal financial loss when the practice suffers from high costs (e.g. due to employing locums). At the same time, the acute trust- or LHB-backed general practices are able to offer staff training and career development opportunities, as well as job security, which increases their chances of recruiting and retaining primary care staff." - Sidhu et al., 2022

**16. Climate of psychological safety**

**When there is a climate of psychological safety (e.g., supportive organisational practices and patient trust) (C), GPs are more able to tolerate risk, ambiguity, and diagnostic uncertainty (O), as they feel supported, trusted, and valued (M).**

"The negative portrayal of GPs in the media, a lack of time for interaction with patients and colleagues, and a perceived blame culture also contributed to young GPs leaving the profession, the researchers found." - Rimmer, 2016

"Professionals described concerns about differences between patient and professional understanding of what was possible, citing high patient expectations; dealing with difficult patients; and fear of complaints or litigation as causes of stress and job dissatisfaction." - Andah et al., 2021

"I have got a colleague here with mental health problems. And I have to say, she's fantastic. She's absolutely brilliant about telling you how she is, how she's feeling, medication she's taking. And that degree of openness is really refreshing" - Spiers et al., 2017

“The findings reported in the four included studies suggest that cognitive-behavioural-based and mindfulness- based programmes delivered in a group format may reduce GP distress at least in the short-term. Increasing awareness generally and with specific regard to thoughts, beliefs, self-care, personal health and setting boundaries appeared to improve GP mental health.” - Murray, 2016

“Whilst there is scope within e.g. mindfulness approaches for mental health promotion the emphasis in healthcare professional well-being interventions appears to be on mental illness prevention (psychopathology continuum).” - Murray, 2016

Organisational interventions tend to develop a ‘psychosocial safety climate’ that comprises clearly communicated managerial participation and commitment to, and prioritisation of, employee psychological health; enhancement of (procedural and relational) organisational justice and team-based interventions to promote mental resources and resilience. Research is sparse regarding an organisational, integrated systems approach to addressing doctors’ potential stressors. Although person-directed resilience training has been recommended to proactively prepare doctors for ‘inevitable’ stressors, a distal-focused approach may be more appropriate.” - Murray, 2016

“Shifting from the deficit approach that underpins stress response amelioration towards a more proactive mental health promotion approach that empowers and enhances work and personal resources may prove to be more effective in and appropriate to our population of interest [GPs].” - Murray, 2016

“Previous work by the RCP [5] has shown that the breakdown of the medical team was a central factor contributing to low morale and disengagement. It is therefore conceivable that developing a sense of collegiality and togetherness across a transgenerational grouping of GPs may boost morale and engagement, leading to greater resilience within the GP workforce.” - Imtiaz-Umer, 2021

“Matheson et al. (2016) described how the development of professional resilience depends on the concept of personal traits (for example, flexibility, tolerance assertiveness, and being a team worker) linking with workplace features (for example, management support, team work, and supportive colleagues acting as workplace buffers) and social factors (for example, friends, family and leisure activities).” - Gray, 2020

“We can end up taking on lots and lots and lots, and that’s often the kind of mentality of people who go into medicine and general practice anyway. So, some- thing to kind of push people away from that. They’re like: ‘We don’t actually have to do that. Just do your job and think about who can do that bit of your job’. To me, that’s what resilience is. To me, that’s really helpful to be reminded of that every so often... I think there is a skill to that as well, isn’t there? You can say yes to everything and burn out by the time you’re 40, or you can start to teach people about where the boundaries are for you in a nice way that doesn’t ruin your relationship with them.” - Cunningham et al., 2021

“It was the belief of participants that contained and focused work in the past allowed GPs to see the outcome of their clinical work in the relationships and gratitude from patients - they received external recognition that their work was of value. The new GPs, in contrast, reported that the relationship with patients has become less reciprocal. Many participants were concerned about a rising complaint culture and demands outwith traditional doctoring work, as expectations of GPs have shifted. For example, new GP Harry (interview) recounted an incident where he felt he had gone out of his way to help a patient, only to receive a complaint." - Laverty et al., 2023

"Unsurprisingly, general practice is the most challenging discipline in medicine as it operates in the realm of complexity and uncertainty.48-51 This requires aptitude and emotional coping skills to become a resilient clinician,48,50." - Sturmberg et al., 2023

"Another group of potentially positive factors was related to the GP and his/her patient contacts and relationships. An efficient doctor-patient relationship was also considered an important factor in GP job satisfaction [17]." - Le Floch et al., 2016

"One in two (53%) respondents felt unable to raise workplace concerns regarding wellbeing, stress or workload." - Dominic et al., 2021

"A chance to work in a stable practice environment where I can be confident, I am receiving performance feedback and where I can have a set number of sessions per week.' 'A reliable position in a practice where I feel part of the team and able to ask questions if I need to." - Lockyer et al., 2014

"Participants noted that resilience meant feeling mentally and physically well and happy at work, which enhanced the experience of caring for patients. This entailed a high level of job satisfaction and enjoyment; a feeling they had done their best, that they were doing a good enough job or performing well in a difficult job, and feeling stable in their working environment: 'Professional resilience is you keep [GPs] happy and engaged which makes them better doctors as well.'" - Martin et al., 2022

"The need for more supportive structures and net- works was also emphasised, together with a more flexible and developmental approach to GP appraisal and revalidation: 'Local support for struggling GPs i.e. access to counselling and CBT locally, face to face and confidential. RCGP should stop adding hoops to GMC revalidation supporting information requirements. All GP's should have access to a network, not just '20 plus' and 'first 5'. Do not forget the 1 in 4 GP's who are locums. Allow GP's to take career breaks and fund/ support their return to general practice'." - Dale et al., 2015

"Practice-level leadership is crucial to the success of any change programme. If staff become stressed or frustrated or feel like they are being bombarded with impossible demands, it will affect their ability to deliver change and improvement. For people to perform at their best they need to feel that they have the safety to test and to fail; to work across traditional boundaries and to build trust." - Brooks et al., 2022

"Discussing patient safety issues with the team at practice meetings was thought to significantly assist in ensuring positive outcomes and reduce the possibility of staff burnout." - Agius et al., 2015

“As well as evidence of developing leadership competencies we also identified perceptions of PED (‘professional education and development course’) as a positive influence on confidence, hardiness and other indicators of resilience. The impact of PED on levels of resilience may lie in providing participants with opportunities for discussion and analysis of potentially stressful situations that occur in general practice combined with peer support and the development of creative problem solving and organizational skills.” - Agius et al., 2015

“This study, using a national cohort involving 67 practices and 348 GPs, found that GPs reporting higher levels of diagnostic uncertainty had two to five times higher risk of emotional exhaustion, job dissatisfaction and turnover intention. Beyond diagnostic uncertainty, GPs reporting multiple episodes of sickness presenteeism were also found to have two times higher risk of EE, job dissatisfaction and turnover intention. GPs reporting poor work-life balance had 1.5 times higher risk of emotional exhaustion, whereas those working longer hours had 2 times higher risk of depersonalisation and job dissatisfaction.” - Zhou et al., 2022

“There is also evidence that burnout may contribute to even greater work demands - a longitudinal study revealed that GPs who attempted to gain emotional distance from their patients as a way of coping with exhaustion, evoked demanding and threatening behaviour from their patients (Bakker et al., 2000). GP commentators have urged that, rather than waiting to address such problems as they arise, it would be wiser to tackle GP burnout before it causes harm to self and others (Jones and Davies, 2016) and that ‘until medical professional bodies recognise the size of the burnout problem and start taking action, physicians can only help themselves’ (Lee et al., 2015, p. 106).” - Hamilton-West et al., 2018

“The changes from baseline scores on the MBI parameters emotional exhaustion and depersonalization were of a much greater magnitude in our study than in the previous GP studies. We found a 38% reduction in emotional exhaustion compared with a 17% reduction in the US study (Fortney et al., 2013) from a similar baseline level, and a reduction in depersonalization of 28% compared with 18%. [...] As noted above, increased compassion for self and others was one of the main themes arising from the qualitative analysis, suggesting that this was an important benefit for GPs. Further work could examine the impacts of increased self-compassion on GP well-being and consider whether such ‘soft’ factors may prove crucial in empowering the GP workforce to successfully rise to the extreme challenges that they currently face in the NHS.” - Hamilton-West et al., 2018

“Internal influencers of well-being comprised Team support, Variety (within their roles, practices, or patients), Control (over their work environment and/or timetable) and an Intense and unmanageable workload. The importance of working within a supportive, interactive team was mentioned by all focus groups as particularly vital for good well-being. Those who felt like they did not receive peer support or have the time to interact with their team described how it could have very negative effects on their well-being.” - Hall et al., 2018

“The GPs also discussed the need for more self-awareness and stress management coaching from their education providers during the early stages of professional training. This could encourage practitioner awareness of burnout while simultaneously encouraging a wider, organizational understanding. Additionally, participants suggested various strategies to foster peer-support. Balint groups (a group of clinicians or doctors who regularly meet to discuss their difficult patient cases in a safe and supportive environment) could be one way of increasing both peer-support while also increasing competence and are used by some physicians as a means to prevent burnout.” - Hall et al., 2018

“Resilience consists of a synergy of personal characteristics (self-worth, flexibility, organisational skills, assertiveness, and humour) and professional and organisational promoters (strong management support, teamwork, workplace buffers, and resources).” - Cheshire et al., 2017

“GPs highlighted that organisational factors also needed to be considered in relation to GP stress. Here, it was considered that there was only so much an individual GP could do to manage stress, given the extent of work pressures they faced: ‘What you’ve got to be careful to do is not ignore the fact that, actually, maybe, for most of us, we are not coping with the stressors because there’s too much stress, not because we’re not resilient enough. And therefore, if you don’t solve the root cause you get nowhere.’” - Cheshire et al., 2017

“[...] These participants preferred approaches such as autonomous resilience groups responsible for their own continuing education, despite challenges involved for GPs in attending regular groups: ‘I think if they’re going to be just one-off activities, that’s pointless, absolutely pointless. And I really think this has to be a continuing thing … So, I would say, if you’re going to do resilience training, it can’t be just one-off events, it’s got to be something that can be continuous and done again and again, and perhaps little groups can be autonomous in training themselves rather than getting people in all the way to provide the training. Fair enough about getting people to start off the training, but certainly to create autonomous groups who could then train themselves.’” - Cheshire et al., 2017

“‘When asked what support would help, both MPs and NMPs highlighted menopause-specific organisational support, in the form of formalised workplace policies (MP10, NMP5): I think workplace policies would help so you don’t feel guilty of taking time off if you had to.’ Furthermore, the need for support from individuals within the organisation, including colleagues and managers was repeatedly highlighted. [...]”. - Adelekan-Kamara et al., 2023

“Our study highlights the inability of doctors to recognise and support menopausal colleagues within the workplace, contributing to a lack of support structures within the NHS. This research suggests that raising awareness, specifically among line managers, is an important first step so that doctors can be confident of strong initial support” - Adelekan-Kamara et al., 2023

“It was noted that stigma exists around GPs accessing help—particularly mental health support—and GPs expressed concerns about confidentiality. Concern was also highlighted over GPs exhausting all alternative routes and coping strategies (such as changing to part time or portfolio working) and feeling that the only option they had left was to 'vote with their feet' and leave direct patient care: I've just become more and more desperate (…) in past years I have just felt terribly angry with the way things are going and now I think, 'I can't actually do anything more about it'. And if I could do anything but vote with my feet, but ultimately, it's the only vote which they're going to listen to." - Sansom et al., 2018

"Access to personal, relational and environmental resources are protective factors for good mental health and well-being and impact on individuals' resilience. Crucially, collegial support is a protective factor for good mental health—support from mentors, supervisors and colleagues is associated with resilience and reduced sickness. Balint groups or similarly structured group work or supervision continue to be employed in general practice and are valued by GPs, yet are not used across all practices. Individual or group supervision aims to provide a safe and supportive space where staff can openly discuss the pressures and emotional challenges of their work and may, as previous evidence suggests, provide GPs with the support they need while offering protection against compassion fatigue and burn-out." - Riley et al., 2018

"The following quote highlights the importance of support and collegiality as a protective factor for good mental health, maintaining morale and feeling less isolated in dealing with the challenges of being a GP" 'The ones that are left of us at the moment are being very supportive of each other and making sure that we're not getting too bogged down with work each and that we are OK. And we keep checking on each other and, you know, 'Are you alright?' And we've had a few little social things to try and get everyone's morale up and support each other and things like that...'" - Riley et al., 2018

"At practice level, poor morale and wellbeing is affecting all staff groups, not just clinicians. Progress has been made during the pandemic to develop a culture that promotes and nurtures staff wellbeing; however, there remains issues of presenteeism and a perceived tokenism to these measures. All staff in general practice need to feel it is ok to not be ok. Support is being offered from multiple avenues and schemes such as 'Looking after you too'10 have been met with demonstrable success. A more overt shift towards a culture that allows time for and normalises the proactive pursuit of wellbeing is perhaps necessary if the profession's sense of belonging is not to be eroded further." - Jefferson et al., 2022

"The majority of GPs cited examples of good team- working and described a sense of pulling together during the pandemic. An increased focus on personal and team mental well-being was reported, though some participants were disenchanted with initiatives that sought to improve 'resilience' as they felt that this placed the onus of responsibility at an individual rather than structural level. Others suggested well-being support was perhaps more easily adopted by larger practices with greater infrastructure. Team 'huddles' were used to debrief on complex cases, provide social support and share anxieties, but small rooms and safe distancing in some prac- tices prohibited in-person staff meetings." - Jefferson et al., 2023

"One of the most important factors in supporting GPs with their work-life balance was a stable, supportive staff group. For example, nursing and administrative staff at one practice described being actively supported in their employment and described the practice as having an open, supportive culture and being a good place to work. GPs said that there was support for each other when it was identified as being needed." - Campbell et al., 2019

**17. Time for care: appropriate consultation time**

**When GPs have longer consultations and/or time with patients (C), this may facilitate patients' active involvement in decision-making (M), leading to improved patient outcomes and more cost-effective care (O).**

**Increasing the duration of expert generalist consultations (C), may avoid unnecessary investigations and reduce downstream referral costs (O), leading to improved patient care and satisfaction (M).**

"Research has illuminated the considerable value of the human side of general practice. Over 20 years ago we learned that longer consultations and the patient knowing the GP are significantly associated with enablement — an important outcome of GP care. GPs are not interchangeable." - Brown et al., 2020

"… and a failure to increase the availability or duration of appointments to meet the needs of an ageing population with complex needs." - Andah et al., 2021

"Adequate resources and support that facilitates professional development would assist in dealing with multimorbidity and complex case management and allow time to include patients in decision making." - Spooner, 2016

"Significant evidence exists that longer consultations enhance relational continuity.[8] Only 8% of GPs feel that the standard consultation is long enough, and 43% of GPs say that insufficient time with patients negatively impacts on their commitment to general practice." - Jeffers & Baker, 2016

"70% state that implementing longer consultation times would resolve the pressures they were under in relation to continuity.[8] - Jeffers & Baker, 2016

"Time to care: The growing complexity of patients - notably, those with multiple long-term conditions - require GPs to have the option of moving to longer appointments for those with multiple long-term conditions. Currently, the average appointment is approximately 10 minutes (Davies 2016), where basing workforce planning off a 15-minute appointment would relieve significant strain. This is the best thing for both patients (who need more support) and for staff (who need time to care)." - Thomas, 2020

"Informants were asked about their views on the present structure of primary care and how they saw it evolving. They acknowledged that a 'one size fits all' or '10 minutes for all' approach was not working, and that there was a need for clinical workload differentiation to take account, not only of the differing needs of patients, but also the differing needs of GPs:

If you have been a GP for 30 years you are able and extremely competent to deal with complicated patients' needs, who need an experienced GP to manage things." - Lyon-Maris et al., 2015

"General/family practice is an international discipline and there is vast experience abroad about generalist practice in the front-line of health systems. An international consensus exists that 10-minute consultations are too short, and the UK is an outlier with so many short consultations. Additionally, continuity of GP care is lower in the UK than in many other countries from Canada to Germany, so patients, GPs, and the NHS may be missing out." - Tzortziou Brown, 2020

"The success of general practice care depends as much on the generalist approach32 as on the ability to establish a therapeutic relationship with one's patient. Being person-centred is key to the development of relational continuity which, in turn, allows the emergence of a deep contextual understanding about a patient's personal circumstances and preferences." - Sturmberg at al., 2023

"When GPs were asked what the hardest part of providing care to marginalised patients was, the most frequent answer was 'not enough time'. GP participants were conscious of the heightened vulnerability of marginalised patients and referenced really wanting to 'do a good job' in providing care for them. Participants had some knowledge of how to do this, but described being limited when simultaneously trying to safely manage high volumes of clinical demand." - Potter et al., 2023

"GPs felt that patient needs have increased due to medical and socioeconomic factors that, combined with increasing levels of expectation, led to concerns about the confines of a 10-minute consultation." - Croxson et al., 2017

"Patients in more deprived areas were less well educated about health, and requested more consultations for minor illness, and patients in more affluent areas had higher expectations, and perhaps higher levels of anxiety, also increasing demand for consultations." - Croxson et al., 2017

"Quite a large cohort of homeless people, street-workers, and drug substance abuse that actually take a long time to obviously sort out and the constraints of the 10 minutes, it's pretty much impossible to sort of sort that out. Whereas my rural practice … the patients around here are a lot more affluent and there's a lot of worried well, so a lot of consultations where perhaps not as much health care has happened that I'd expected to, and the sort of demands are sometimes unreasonable.' (GP27, male locum, eight sessions per week, 1-5 years' experience, regularly works at one inner-city and one rural practice)." - Croxson et al., 2017

"Significant evidence exists that longer consultations enhance relational continuity.[8] Only 8% of GPs feel that the standard consultation is long enough, and 43% of GPs say that insufficient time with patients negatively impacts on their commitment to general practice;[7] 70% state that implementing longer consultation times would resolve the pressures they were under in relation to continuity." - Jeffers & Baker, 2016

"Introducing new methods of working such as a separation of acute care (aligned with a quality out-of-hours service) from management of patients with long-term conditions or multiple comorbidities where the latter will truly benefit from continuity of care and longer consultations." - Vorster, 2017

“Yet, creating longer consultation times in these ways obviously requires other changes to be made to facilitate this. Some practices dispensed with separate disease specific clinics. Recognising that many patients with long-term conditions have multiple pathologies, one way to reduce repeated scheduled visits was to bring such patients into a single long-term condition clinic with longer appointment times.” - Robertson et al., 2012.

“Anecdotal reports show that in some practices GPs are now in contact with patients at only 6-minute intervals. This retrograde step is concerning, as this pace, for either triage or consulting, is simply too fast and probably not safe for patients or GPs.” - Tzortziou Brown, 2020

**18. Agile and flexible systems**

**When GPs can adapt standard systems (C), they can apply their expertise in agile and flexible ways (M) to meet individual circumstances or personal needs (O).**

“Furthermore, it is not just the receptionist or clinician’s innate or learned abilities, but their capacity to use those abilities within the context of available resources, system factors, and workload pressures.” - Voorhees et al., 2022

“A revised model could see both being properly enabled to be the ‘named GP’ for patients with chronic or complex needs requiring relationship as well as management continuity. Thirty-minute appointments would allow for holistic care, with each named GP at the heart of a comprehensive team, all responsible for the same named patients and all with excellent links with any secondary care teams. And in symbiosis with these practice-based GPs, we’d see small, smart, agile, highly engaged GPs flexibly deployed to manage patients who still need excellent continuity of management, but don’t have ongoing, complex problems requiring a ‘named GP’ approach. GPs with special interests or skills can put these into use across different settings.” - Fieldhouse, 2017

“Appraisals immediately following a GPs 5-yearly revalidation could be made more flexible and more relevant to each individual doctor's learning preferences. GPs and their appraisers could make better use of their time together, and the resentment of some towards obligatory written reflection could be avoided. We propose that GPs, and their appraisers should agree and choose an option for the following year that would be integral to the individual doctors personal development plan.” - Curtis et al., 2018

“Mainstream appointment systems are often inaccessible to marginalised patients who often require rapidly responsive, easily accessible routes into care.” - Potter et al., 2023

“Prioritise flexible access and longer appointments to patients in greater need: One practice dovetailed the care coordinator role with a specialist inclusion health clinic for marginalised patients;

in the other the care coordinator had a direct telephone line they provided to patients and priority access to appointments when needed. One practice also had protected appointment slots that could be used by drug and alcohol workers who could use this for one of their clients if needed." - Potter et al., 2023

"I've also got the authority, the clearance, to book appointments in when I need to, whereas the staff, they'd have to say, 'Okay, we haven't got any more for today and the next one's in two weeks,' I can change a slot with the doctor."-CC1." - Potter et al., 2023

"One care coordinator offered patients on her list a direct phone number that would only be answered by her so patients had the security of knowing who they would speak to (see Theme 2- Shifting ways of working- provide proactive continuity of care from trusted, supported professionals)." - Potter et al., 2023

"One care coordinator had a designated phone number which she gave out to patients for direct access to a known contact. Despite fears from colleagues that she would be overwhelmed with calls, this did not happen. The coordinator saw it as a valued tool that helped marginalised patients feel more comfortable to access. "People aren't over-using it, not at all, but I think it's just knowing that it's there, that they feel a lot safer, that they can have that line and it's only on my desk; nobody else will answer it."- CC1." - Potter et al., 2023

"Learning and 'tweaking' - as noted above, introducing flexible working or job crafting is a 'process' not an event and needs constant monitoring and a willingness to make changes if needed. This means being willing to adapt and adjust working arrangements or job roles if they are not working for staff or patients rather than hoping that 'things will work out', when they clearly will not." - Bevan et al., 2022

"Every member of staff has personal circumstances which will influence their need for flexibility at work (whether permanent or temporary). Some will be more open about these with colleagues than others, but it is often the case that the more open and supportive a team is, the more comfortable a member of staff will be to disclose that they need flexibility at work." - Bevan et al., 2022

"It was understood that flexible practices would only be sustainable if they matched patient, practice and individual needs. Co-creation and collaboration with the practice team is important to understand how work practices and job design could be improved to help both working time and work intensity." - Bevan et al., 2022

"Organisation is often preferred through a GP cooperative that combines size of scale advantages with organisational features of strong primary care, such as good accessibility, continuity and coordination of care. We expect GPs to be more satisfied with GP-based models than with hospital- and national-based models, and that large doctor based models are more satisfactory than small GP-based models in terms of workload." - Stobbe et al., 2021

**Flexible employment arrangements**

“Workload should be reduced by moving away from a nine-session week towards seven clinical sessions per week for full time staff. The time freed up by this should be used for career development, respite and developing other specialisms such as management, academia or specific types of care (including end of life care and mental health).” - Thomas, 2020

“70% of our locum workforce are female. They are mainly ethnically diverse and mainly working in carer-responsible roles. We need to bring them back into the workforce as well. We need a workforce model that works flexibly for people who cannot quite work in the way that traditional general practice describes.” - Health and Social Care Committee, House of Commons, 2022

“The findings of this study mirror a similar one undertaken in Ireland by Graham,40 and illustrate the tendency for female GPs to take the ‘path of least resistance’ in their professional lives. The present study shows that meeting childcare needs remains a particular challenge for female GPs, and is a significant factor in their availability and ability to work, as well as to engage in professional development. This is particularly marked for those in both the under 34 and 35-49 age groups, which reported significantly more difficulties than the older age group. younger female GPs with childcare needs were less likely to be involved in teaching and training, and even less likely to be involved in PCT, LMC or deanery work, special interests or clinical assistant work, something also found elsewhere.” - Wedderburn et al., 2013

Short-term labour fluctuations within the workforce will become increasingly significant for service provision, the challenge for educators and those with responsibility for shaping the future workforce is to allow for flexibility in professional practice and educational development without loss of career potential. The needs of this sector of the workforce are clear: female GPs require support in terms of their education, training and childcare needs in order to facilitate increased representation in senior practice, healthcare and education roles in readiness for times ahead when they will be critical to service delivery. - Wedderburn et al., 2013

“In terms of career characteristics, Generation Y [adults under 45] are career 'hoppers’: they are likely to switch jobs frequently and have more jobs over their lifetime than previous generations. They move for increased pay and innovative working environments. They want versatility and flexibility in the workplace, and strive for a strong work–life balance. They are more detached from institutions and networked with friends; they use social media and embrace technology. Generation Y often want to have parallel or ‘portfolio’ careers.” - Simon, 2018

“Transferring their skills (eg, taking on roles as an appraiser, clinical commission lead, advisory committee member, pharmaceutical consultant or working for a medical school) was a further option that GPs identified: either having this role alongside direct patient care or as an alternative to it. A key attraction of transferring skills included feeling financially and professionally rewarded. A medical degree is one of the most wide-ranging degrees there is: it’s about science, research, communication, empathy, organisation, management—we’re pretty skilled people… Other people want me to do other stuff now; they'll pay me good money and treat me very differently to what is currently happening to GPs." - Sansom et al., 2016

"Once challenges were overcome, group consultations were reported to mitigate the problems of individual consultations, because of both actual and perceived extra time, resulting in staff feeling more relaxed and to 'bring the joy back into consultations' (P15). In contrast, individual consultations were described as monotonous, isolating, and better suited to acute care, with pressure to get patients 'out of the door' (P8) due to time and workload demands: 'The time pressure's off, you're not looking at the clock and thinking, oh God I'm running late, you're inevitably running late on one-to-ones. Whereas group consults you're not because you've got your half hour or whatever it is, so it's so relaxing and empowering … it's just a nicer experience.'" - Swaithes et al., 2020

"I'm not keen to be confined to the same building for the next 40 years. With flexible use of estates, I can envision working in a central hub where there is a consolidated back office function; large rooms for educational sessions and meetings; and the space to have a multitude of clinicians virtually consulting, with an option to work from home. This allows for socialising with GPs from across the locality and also frees up space in dispersed estates for face-to-face consulting with complex patients, and those who need review after triage." - Lumley, 2020

"After going back to work following maternity leave it helped me maintain my health as I was already exhausted and hugely benefited from knowing exactly what my working pattern would be without extra patients or visits at the end of the day. . . . think it would be a great shame to lose the retainer scheme for future doctors as it is so important to be able to cut back while the children are young. I think more doctors would leave medicine if it were to be abandoned." - Lockyer et al., 2014

"We will significantly increase flexible working through a combination of technology and a change in people practices, to give people greater choice over their working patterns, help them achieve a better work-life balance, and help the NHS remain an attractive career choice. We will need to advertise more roles as flexible (for example, less than full time, term time only, job shares) and, where possible, enable home working to bring our employment offer into line with other sectors." - NHS People Plan, 2019
